# Pathogenic variants in *TMEM184B* cause a neurodevelopmental syndrome associated with alteration of metabolic signaling

**DOI:** 10.1101/2024.06.27.24309417

**Authors:** Kimberly A Chapman, Farid Ullah, Zachary A Yahiku, Sheraz Khan, Sri Varsha Kodiparthi, Georgios Kellaris, Hazel G. White, Andrew T. Powell, Sandrina P Correia, Tommy Stödberg, Christalena Sofokleous, Nikolaos M Marinakis, Helena Fryssira, Eirini Tsoutsou, Jan Traeger-Synodinos, Andrea Accogli, Vittorio Sciruicchio, Vincenzo Salpietro, Pasquale Striano, Candace Muss, Boris Keren, Delphine Heron, Seth I Berger, Kelvin W Pond, Suman Sirimulla, Erica E Davis, Martha RC Bhattacharya

**Author notes:** Corresponding authors: Erica Davis, Martha Bhattacharya. These authors contributed equally (Kimberly A Chapman, Farid Ullah, Zachary A Yahiku).

## Abstract

Transmembrane protein 184B (TMEM184B) is an endosomal 7-pass transmembrane protein with evolutionarily conserved roles in synaptic structure and axon degeneration. We report six pediatric cases who have *de novo* heterozygous variants in *TMEM184B*; five individuals harbor a rare missense variant and one individual has an mRNA splice site change. This cohort is unified by overlapping neurodevelopmental deficits including developmental delay, corpus callosum hypoplasia, seizures, and/or microcephaly.

TMEM184B is predicted to contain a pore domain wherein four of five human disease-associated missense variants cluster. Structural modeling suggests that all missense variants alter TMEM184B protein stability. To understand the contribution of TMEM184B to neural development *in vivo*, we knocked down the *TMEM184B* ortholog in zebrafish and observed microcephaly and reduced anterior commissural axons, aligning with symptoms of affected individuals. Ectopic expression of *TMEM184B* c.550A>G; p.Lys184Glu and c.484G>A; p.Gly162Arg variants cause reduced head size and body length, indicating dominant effects, while three other variants show haploinsufficiency. None of the variants are able to rescue the knockdown phenotype. Human induced pluripotent stem cells (iPSC) with monoallelic production of p.Lys184Glu show mRNA disruptions in key metabolic pathways including those controlling mechanistic target of rapamycin (mTOR) activity. Expression of p.Lys184Glu and c.863G>C; p.Gly288Ala increased apoptosis in cell lines and p.Lys184Glu increased nuclear localization of transcription factor EB (TFEB), consistent with a cellular starvation state. Together, our data indicate that *TMEM184B* variants cause cellular metabolic disruption and result in abnormal neural development.

## INTRODUCTION

Transmembrane protein 184B (TMEM184B) is an endosomal transmembrane protein that facilitates synaptic maintenance and axon degeneration across species. Loss of TMEM184B in mouse and *Drosophila* models causes synaptic malformations and neuronal hyperactivity.^1–3^ While these studies highlight an important role for TMEM184B in the maintenance of nervous system integrity in adult organisms, its role in the processes required for neural development remains less clear. Primary embryonic sensory neurons from mice with a mutation in *Tmem184b* (gene trap model) show a reduction in transcripts related to Wnt signaling, a classical developmental pathway with substantial nervous system effects.^3–5^ Given the broad expression of *TMEM184B* in the central nervous system^6^ coupled with the effects of its loss on the Wnt signaling pathway, it is hypothesized that TMEM184B function may contribute to neuronal differentiation, migration, and/or survival. However, the neurodevelopmental disruptions that result from *TMEM184B* impairment are unknown.

Few studies have examined the effects of *TMEM184B* alteration in human cells. Knockdown of *TMEM184B* in carcinoma cell lines causes reduced proliferation *in vitro* and in tumor xenografts, suggesting that its function is critical for the growth of malignant cells.^7^ *TMEM184B* transcripts are upregulated in squamous cell carcinoma clinical samples and have been implicated in increased metastatic cell migration via regulation of the cytoskeleton.^8^ Finally, genome-wide association studies have implicated *TMEM184B* in mammographic density-associated breast cancer risk.^9^ Together, these studies suggest a link between *TMEM184B* expression and cellular growth and survival. Recently a large-scale Mendelian disease sequencing effort proposed *TMEM184B* as a candidate gene; a single individual with corpus callosum hypoplasia and seizures was reported to have a *de novo TMEM184B* variant, though the precise DNA change and other clinical information was not given.^10^ Aside from this case, pathogenic variants in this gene have not been described or studied in human populations.

During development, alteration of cellular proliferation and growth signaling pathways can have devastating consequences, especially for the central nervous system which grows dramatically in the final trimester of human gestation.^11,12^ Two of the most common neurological diagnoses in children are corpus callosum dysgenesis and seizures. These diagnoses often co-occur in affected individuals, suggesting that the affected processes may be molecularly or functionally related. Microcephaly is a severe presentation of neurodevelopmental growth disruption that is defined by smaller average head circumference (when controlled for other factors). Recent work has implicated pathogenic variants in a group of human genes in microcephaly whose encoded proteins cluster in pathways known to affect axon outgrowth (e.g. tubulin isoforms TUBA1A, TUBB3) and neuronal differentiation (CTNNB1, FOXG1).^13^

Neurons and neural progenitors require cellular nutrient sensors to make decisions about growth and differentiation. Transcription factor EB (TFEB) is a key regulator of lysosomal and autophagosomal gene transcription, and its localization is acutely sensitive to cellular amino acid levels. In high nutrient conditions, TFEB is phosphorylated by multiple kinases that regulate cell growth including the mechanistic target of rapamycin C1 (mTORC1) complex and mitogen activated protein kinase 4K3 (MAP4K3).^14^ Phosphorylation sequesters TFEB in the cytoplasm,^15^ but upon starvation, TFEB becomes dephosphorylated and it enters the nucleus where it promotes transcription of mRNAs encoding autophagosomal and lysosomal components.^16^ By promoting the formation of additional autophagosomes and lysosomes, TFEB transcriptional activity increases cellular catabolism to promote short-term survival.^17^ TFEB nuclear-cytoplasmic shuttling dynamics are altered in neurological diseases. Aberrant accumulation of TFEB in the nucleus occurs in lysosomal storage disorders, which are childhood-onset diseases caused by improper lysosomal cargo degradation.^16^ On the other hand, TFEB nuclear entry is reduced in the most common genetic type of amyotrophic lateral sclerosis (ALS; MIM: 105400) caused by repeat expansion in the *C9ORF72* locus (MIM:614260).^18^ Therefore, promoting TFEB transcriptional activity is being explored as one avenue to treat neurodegenerative disorders by promoting degradation of misfolded proteins and malfunctioning organelles.

Here we describe the identification of a cohort of affected individuals with *de novo*, heterozygous variants in *TMEM184B*. We use zebrafish to implicate the *TMEM184B* ortholog in animal growth, development of anterior structures and emanation of commissural neurons. We test both dominant and loss of function variant hypotheses *in vivo* and show that disease-associated variants can have diverse molecular effects. To gain insight into the molecular mechanism of disease, we perform RNAseq on induced pluripotent stem cells (iPSC) harboring a variant from an affected individual and find alteration of metabolic pathways and functional groups governing axon guidance. Further, and in agreement with our transcriptomic data, we find variant-dependent effects on TFEB nuclear localization, as well as increased susceptibility to apoptosis, suggesting a pathological explanation for the effects of variants identified in affected individuals on cellular function and neural development. Our work highlights a new cause of neurodevelopmental disorders and provides critical insight into the rare syndrome caused by *TMEM184B* disruption.

## SUBJECTS, MATERIALS, AND METHODS

### Inclusion of Human Participants

All research was performed in accordance with the Declaration of Helsinki. Most participants were identified by clinical sequencing and then unified via GeneMatcher.^19^ Subjects and their families agreed to allow de-identified information to be summarized from medical chart review and reported in this study. Subject 5 was also consented for fibroblast utilization and maintenance to a biorepository, which allows for residual samples from clinical procedures be collected, saved and distributed for research.

### Genome Sequencing and Analysis

Sequencing was performed at each institution using either genome or exome approaches. Allele frequency was estimated using data from the gnomAD database (version 4.1.0 unless otherwise specified) and was last queried in June 2025.

AlphaMissense, CADD v1.7, REVEL and GERP were used to evaluate possible pathogenicity of each missense variant.^20,21^ All analyses of sequence variation used RefSeq transcript NM_012264.5.

### Structure Prediction

The native protein sequence (RefSeq NP_036396.2 isoform a) was extracted from Uniprot, and each variant was inserted accordingly. Alphafold, along with MMSeq2, was employed to generate 3D structures of TMEM184B and its five clinically relevant missense variants. Structures were quality assessed via PLDDT and PAE scores, which provided five models for each protein structure. PLDDT gives the confidence score of the structure predicted (the higher, the better). Some loop regions that did not fit the desired criteria were truncated for cross-validation of the results. To assess the quality and reliability of the predicted protein geometry, the backbone dihedral angles of the predicted protein structures were mapped onto a Ramachandran plot. Similar to the PLDDT plot, we truncated the protein structure at the membrane to remove sequences in loops, a common practice in structural biology.^22^ Visualizations, including overlays of native and variant predicted protein structures, were created in Pymol 2.6. The best structure for native protein was chosen based on the highest PLDDT scores and the most favorable Ramachandran plot. Phosphorylation site prediction was performed using the web tool GPS6.0.^23^

### Zebrafish Husbandry and Embryo Maintenance

All experimental procedures were performed according to protocols approved by the Northwestern University Institutional Animal Care and Use Committee (IACUC; Protocol IS00016405). Wild type (WT) AB adult fish were maintained in standard husbandry conditions on a 14h light and 10h dark cycle. Embryos were obtained from natural matings with a 14-day rest period between each mating. Embryos were grown in E3 medium (0. 581 g/L NaCl, 0.026 g/L KCl, 0.096 g/L CaCl_2_·2H_2_0, 0.163 g/L MgCl_2_·2H_2_0, and 0.0002% methylene blue) at 28.5°C until phenotypic readout or tissue harvest.

### Transient Depletion of *tmem184ba*

We obtained morpholinos (MO) to target the splice donor site of *tmem184ba* exon 3 (e3i3) and exon 6 (e6i6), and a standard control MO (Gene Tools, LLC; **Table S1**). *tmem184ba* was suppressed transiently by injecting 1 nl MO (e3i3 or e6i6) at increasing doses (0.6 ng, 1.2 ng and 1.8 ng) and (1 ng, 2 ng and 3 ng), respectively, into one to four cell staged zebrafish embryos. To determine MO efficiency, we extracted total RNA from pools of 20 larvae/condition (controls and MO-injected) at 2 days post-fertilization (dpf) using Trizol (Thermo Fisher Scientific) according to manufacturer’s instructions. cDNA was synthesized with the QuantiTect Reverse Transcription kit (Qiagen). RT-PCR was performed using primers flanking the MO target locus (**Table S1**), and amplified PCR products were separated on a 1% agarose gel. The resulting PCR bands were gel purified with the QIAquick gel extraction kit (Qiagen) and cloned into TOPO-TA cloning vector (Thermo Fisher Scientific). The purified plasmids from resulting colonies (n=4/PCR product) were sequenced using BigDye 3.1 terminator chemistry on an ABI3730 Genome Analyzer according to standard protocols.

### Molecular Cloning, Site Directed Mutagenesis, and *In Vitro* Transcription of *TMEM184B*

To express human *TMEM184B* WT and variant mRNAs in zebrafish embryos, we obtained a Gateway-compatible WT *TMEM184B* (NM_012264.5; cat# HOC18517) open reading frame (ORF) entry vector (GeneCopoeia). We shuttled the ORF into a pCS2+ Gateway destination vector using LR clonase II (Thermo Fisher Scientific). The five *TMEM184B* missense variants identified in affected individuals (c.262G>A, p.Val88Met; c.380T>G, p.Leu127Arg; c.484G>A, p.Gly162Arg; c.550A>G, p.Lys184Glu, and c.863G>C, Gly288Ala); or six variants from gnomAD v4.1.0 (c.83C>T, p.Pro28Leu; c.160G>A, p.Val54Met; c.251T>C, p.Ile84Thr; c.274G>T, p.Ala92Ser; c.340G>A, p.Val114Ile; and c.845G>A, p.Arg282His) were introduced in the pCS2+ constructs using site-directed mutagenesis as described (**Table S1**).^24^ After sequence verification of WT and variant ORFs (**Table S1**), we linearized each construct with *Not*I and performed *in vitro* transcription with the mMessage mMachine SP6 Transcription kit (Thermo Fisher Scientific) according to manufacturer’s instructions. To test dominant effects, we injected 200 pg of each *TMEM184B* mRNA alone, and for a subset of variant mRNAs, titrated with WT in variable doses of 50 pg, 100 pg, and 150 pg while maintaining a total mRNA concentration of 200 pg. For *in vivo* complementation studies to test loss-of-function effects, we injected 200 pg mRNA with 1.2 ng (e3i3) or 3 ng (e6i6) MO.

### qRT-PCR Analysis of Human *TMEM184B* Transcript in Zebrafish Embryos

To monitor human *TMEM184B* mRNAs in zebrafish embryos, we injected equivalent doses of each mRNA (200 pg) into embryos at the one-to-four cell stage and grew them to 24 hours post-fertilization (hpf). We harvested pools of 30 embryos/condition in Trizol, extracted total RNA, and generated cDNA as described above. To detect human mRNA, we used two pairs of primers targeting *TMEM184B* at the 5’ end (exon 2 to exon 3) and 3’ end (exon 7 to exon 8) and performed qRT-PCR with SYBR Green qPCR Master Mix (Thermo Fisher Scientific) and normalized data to zebrafish *b-actin* (two biological replicates, each in technical triplicate; **Table S1**). Statistical differences were calculated with a one-way ANOVA with Tukey’s post-hoc test using GraphPad prism version 8. P-values <0.05 were considered statistically significant.

### Automated Live Imaging of Zebrafish Larvae

To assess head and body size, tricaine-anesthetized larvae were imaged at 3 dpf or 4 dpf using the Vertebrate Automated Screening Technology (VAST) Bioimager (Union Biometrica) mounted to an AXIO Imager.M2m microscope (Zeiss) with a 10x objective lens. Larvae were passed sequentially through a 600 μm capillary on the stage module. Each larva was detected by VAST software (version 1.2.6.7) with a 40-70% minimum similarity and oriented automatically using preset recognition templates while operating in automatic imaging mode as described.^25^ Bright field lateral images were captured with the VAST onboard camera.

### Whole Mount Immunostaining

Larvae were reared to 3 dpf and fixed overnight in Dent’s solution (40% methanol [MeOH] plus 20% dimethyl sulfoxide [DMSO]). Embryos were then rehydrated gradually in MeOH, Phosphate-Buffered Saline (PBS) and 0.1% Tween (T) [PBST] at room temperature. We washed the embryos with bleach solution (9 mL PBST + 1 mL H_2_O_2_ + 0.05g KOH) for 13 minutes followed by proteinase K treatment for 10 minutes and post-fixation with 4% paraformaldehyde (PFA) for 20 minutes at room temperature. Embryos were washed with PBST 3X (10 min each) and incubated in blocking solution (IF buffer [1% BSA in PBST] +10% fetal bovine serum [FBS]) for one to two hours. Primary detection was performed with anti-acetylated tubulin antibody (Sigma-Aldrich, T7451, 1:1,000 overnight at 4° C) and secondary detection with Alexa Fluor 488 goat anti-mouse IgG (Invitrogen; A11001, 1:1,000). Larvae were imaged by capturing fluorescent signal with an AZ100 microscope (Nikon) with a 10X objective mediated by Digital Sight black and white camera (Nikon) and NIS Elements software (Nikon).

### Zebrafish Image Analysis

We used ImageJ (NIH) software to measure head and body size from lateral bright field images of zebrafish larvae using consistent anatomical landmarks. For fluorescence images, we measured size of the optic tecta and counted the number of commissural neurons that cross the midline on dorsal images.

### Statistical Analysis of Zebrafish Morphometrics

All quantitative data were analyzed with a one-way ANOVA with Tukey’s post-hoc test using GraphPad prism version 8. P-values <0.05 were considered statistically significant. Experiments were performed at least twice with investigator masked to injection conditions.

### Amplicon Sequencing of *TMEM184B*

Fibroblasts were collected from Individual 5 under IRB-approved procedures. Fibroblasts were converted to induced pluripotent stem cells (iPSC) by the Washington University in St. Louis Genome Engineering and iPSC Center (GEiC). Control iPSC from an unaffected, sex-matched and approximately age-matched individual were purchased from Coriell Biobank (line GM23476). Cells were maintained in mTeSR media (StemCell Technologies) with antibiotics. RNA was isolated using the RNeasy Mini kit (Qiagen). Reverse transcription was performed using the Quanta Bio First Strand Synthesis kit, and PCR was done using Pfusion polymerase (New England Biolabs) according to product recommendations (**Table S1**). Following gel isolation of PCR products, sequencing was submitted for Premium PCR Sequencing using Oxford Nanopore long-read technology (Plasmidsaurus, South San Francisco, CA).

### iPSC Culture and Gene Editing

Coriell biobank line GM23476 was used as an isogenic, non-disease control line. Following expansion, this line was provided to the Washington University in St.

Louis Genome Engineering and Stem Cell Center for editing. A gRNA overlapping the desired edit (c.550A>G) was transfected into line GM23476 (**Table S1**). Single clones were cloned, genotyped, and expanded. Clones were evaluated for pluripotency using immunohistochemistry to OCT4, SSEA4, SOX2, and TRA-1-60 by the core. Cells harboring a heterozygous or homozygous modification at the c.550A>G locus were used for RNAseq.

### RNA and Transcriptomic Analysis

To analyze RNA levels of *TMEM184B*, RNA was extracted from iPSC cultures as described above and submitted to Novogene (Davis, CA) for sequencing. Files containing raw reads were examined for quality using FastQC and subsequently trimmed for adapter content (Trimmomatic version 0.39) before proceeding to alignment. These trimmed reads were mapped to a transcriptome index for the human genome (version 38) using Salmon (version 1.8.0). Transcript counts in the form of transcripts per million (TPM) were analyzed by DeSeq2 (Galaxy server) to identify differentially expressed genes. Affected genes passing thresholds for fold change and significance were further examined in pathway enrichment analysis using Panther. Graphs were generated in Galaxy (PCA plots) or R (volcano plots), and heatmaps were generated using the Heatmapper web tool.

### Cell Culture and Reporter Cell Line Creation

HEK293T cell lines were cultured in 1X Dulbecco’s Modified Eagle Medium (Ref#11995-065) supplemented with 10% FBS, L-glutamine, and 1X Penicillin Streptomycin at 37°C and 5% CO2. To create stable cell lines expressing both *TFEB-sfGFP* and *H2B-mRuby*, infection and selection were done sequentially. First, retrovirus expressing *TFEB-sfGFP* (Addgene Plasmid #135402) was produced by transfecting HEK293T cells with *TFEB-GFP* plasmid, pVSV-G, and pMD2.G with GeneJuice (Millipore Sigma) and Optimem (Gibco). HEK293T cells were infected with *TFEB-sfGFP* retrovirus along with polybrene (8ug/mL). A population with high green fluorescence levels was isolated via flow cytometry (BD FACSAria III). *TFEB-sfGFP* expressing cells were then infected with *H2B-mRuby* lentivirus (Addgene Plasmid #90236) prepared as above except that VSV-G was replaced with PSPAX2. A population with high TFEB-sfGFP and H2B-mRuby were isolated using FACS and were further selected via sequential treatment with 8 μg/mL puromycin and 400 μg/mL hygromycin.

### TFEB Translocation Assay

*FL-TMEM184B-miRFP670* plasmid constructs were ordered from Twist Bioscience. Plasmid purity and quantity was assessed via Nanodrop (260/280 ratio > 1.9). On Day 0, TFEB-sfGFP/ H2B-mRuby stable cells were transiently transfected with *TMEM184B* constructs (3.75 μg into ∼700,000 cells) using GeneJuice. On Day 1, each condition was seeded in duplicate into 96-well black bottom plates and transferred to the University of Arizona’s Microscope Shared Resource facility and left overnight in 37°C and 5% CO2. On Day 2, media was changed to phenol red-free imaging media. Dulbecco’s Modified Eagle Medium (Ref#21063-029) was supplemented with 10% FBS, L-glutamine, 1X Penicillin Streptomycin, and 110 ng/L sodium pyruvate and applied to one duplicate (complete media, non-starved), and Dulbecco’s Modified Eagle Medium (Ref#A14430-01) was supplemented with only 1X Penicillin Streptomycin for the other duplicate (starved).

The University of Arizona Microscopy Shared Resource facility’s Nikon SoRa spinning-disk confocal microscope was used for imaging. Cells were imaged at 37°C and 5% CO2 in a 4×4 grid with 8% overlap from the center of the well every hour for 6 hours. 20x magnification, 4×4 binning, and 16-bit parameters were used. sfGFP, mRuby, and miRFP670 fluorescent channels were captured.

### Image Analysis

A rolling ball background subtraction of 29.86 μm was applied to the sfGFP channel in Nikon NIS Elements software post-acquisition. Individual frames were then exported and analyzed using QuPath imaging pipelines. Cell nuclei were defined by the H2B-mRuby channel and were expanded 2 μm outwards to define the perinuclear region. Detections were filtered through thresholding for nuclear circularity, nuclear area, cell area, GFP intensity, cell circularity, and miRFP670 intensity.

Nuclear to perinuclear sfGFP fluorescence ratios were quantified in non-starved cells, and cells starved at 1-hour and 6-hour timepoints. The average nuclear to cytoplasmic ratios of all conditions across 3 trials were calculated and then normalized to non-starved HEK293T cells expressing WT *TMEM184B*. GraphPad Prism 10 was used for statistical analysis. Data normality was assessed using Shapiro-Wilk tests. One-Way Parametric ANOVA tests with adjusted p-values were calculated in Graphpad Prism 10, comparing each condition to WT at each timepoint.

### Zombie-Violet cell death assay

HEK293T cells were transfected with *TMEM184B-miRFP670* containing native sequence or variants identified in affected individuals and cultured for 2 days. Prior to Zombie-Violet assay, cells were starved by incubating with Dulbecco’s Modified Eagle Medium (Ref#A14430-01) at 37°C and 5% CO2 for 6 hours. Cell incubation was according to the manufacturer’s protocol (BioLegend). Zombie-Violet was diluted 1:500 in DPBS and 100 μL of solution were used per one million cells. Cells were washed with 0.5% BSA/PBS, fixed in 2% PFA, washed once with DPBS, and transferred to the University of Arizona’s Flow Cytometry Core to be analyzed (BD FACSCanto II). FlowJo software was used for FACS analysis, and GraphPad Prism 10 was used to create graphs of cell percentages and perform statistical testing (One-Way Parametric ANOVA).

## RESULTS

### *De Novo* Heterozygous Disruptions in Human *TMEM184B* Cause Neurodevelopmental Defects

We leveraged the GeneMatcher platform to identify *TMEM184B* variants that may be disease-causing. We identified six affected individuals in which exome or genome sequencing (ES or GS, respectively) identified a rare (<0.1% minor allele frequency in gnomAD v4.1.0) mRNA splice site or missense disruption to *TMEM184B*. For these affected individuals, we obtained information from physicians and genetic counselors about the alteration in the DNA sequence and the clinical findings observed (**Table 1, Note S1, Figure S1 and Table S2**). In this dataset, all six affected individuals were diagnosed with neurodevelopmental deficits as children and then, by sequencing, were found to carry a variant of unknown significance (VUS) in *TMEM184B*.

**Table 1.** Affected individuals in the current study contain rare sequence variants within *TMEM184B.* All affected individuals whose variants were modeled in this study are shown. Allele frequency was determined by search of the gnomAD database (v4.1.0). AlphaMissense and REVEL scores predict the likelihood of a missense variant to be pathogenic (highest likelihood = 1, lowest likelihood = 0). CADD scores show the deleteriousness of nucleotide substitutions, with higher numbers indicating less likely (more deleterious) substitutions. GERP scores indicate evolutionary constraint (max is 6.18, and above 2 is considered constrained). Individual 5 (Family 5-II-1) could not be calculated for AlphaMissense or REVEL because the sequence change results in a splice acceptor variant, not a missense variant. Abbreviations: m, months; y, years; F, female; M, male; N/A, not applicable; ND, not done. See Figure S1 for pedigrees.

| Patient | Age of diagnosis | Sex (assigned at birth) | Origin | Genotype | Protein prediction | Detection Method | Allele frequency | Alpha- Missense score | REVEL score | CADD v1.7 | GERP score | Developmental delay | Intellectual disability | Microcephaly | Thin or hypo-plastic corpus callosum | Seizures |
| --- | --- | --- | --- | --- | --- | --- | --- | --- | --- | --- | --- | --- | --- | --- | --- | --- |
| 1 | 0-5 y | F | De novo | c.262 G>A | p.Val88Met | Whole exome | 0.000029 | 0.902 | 0.63 | 32 | 4.53 |  |  |  |  |  |
| 2 | unknown | M | De novo | c.380 T>G | p.Leu127Arg | Whole exome | Not in gnomAD | 0.981 | 0.8 | 30 | 5.15 |  |  |  |  |  |
| 3 | 0-5 y | F | De novo | c.484 G>A | p.Gly162Arg | Whole exome | Not in gnomAD | 0.697 | 0.36 | 25.7 | 5.12 |  |  |  |  |  |
| 4 | 0-5 y | M | De novo | c.550 A>G | p.Lys184Glu | Whole genome | Not in gnomAD | 0.984 | 0.81 | 28.9 | 5.4 |  |  |  |  |  |
| 5 | 6-10 y | F | De novo | c.787+1G>A | p.Ser254Hisfs*58 | Whole exome | Not in gnomAD | N/A | N/A | 36 | 5.52 |  |  |  |  |  |
| 6 | 6-10 y | M | De novo | c.863 G>C | p.Gly288Ala | Whole exome | Not in gnomAD | 0.628 | 0.3 | 25.1 | 4.77 |  |  |  |  |  |

All six affected individuals show partial phenotypic overlap. Their features coalesce around a few major clinical findings: global developmental delay (5 of 6), corpus callosum hypoplasia (4 of 6, **Figure 1**) or more severe structural disruption resulting in microcephaly (1 of 6), and seizures (4 of 6). Other non-neurological features are also present in some affected individuals (**Table S2**). Four cases showed rare variants outside *TMEM184B*, but these were either variants of uncertain significance or were present in a gene known to cause non-overlapping phenotypes. ES or GS of the remaining four affected individuals did not identify other possible genetic contributors to their clinical sequelae. Five of six variants were absent in human control populations (gnomAD 4.1.0; **Table 1**). One variant, p.Val88Met was rare but present in gnomAD 4.1.0 (34 non-Finnish Europeans and 1 Finnish European of 807,162 samples), and in gnomAD v3.1.2 non-neuro (3 of 67,442 samples; **Table 1**). All variants occurred at highly constrained residues (GERP scores > 4.5) and had CADD scores between 25 and 36 falling among the highest 0.3 percent of predictions of deleteriousness for all variants.^21^ For the five coding region variants, REVEL scores were above 0.77 (moderate evidence for pathogenicity) for two variants and between 0.3 and 0.63 for the remaining three (neither supportive of being pathogenic nor benign). Using AlphaMissense (structural constraint model based on Alphafold), three variants are highly likely to be pathogenic (score > 0.8) while two variants are in a middle range (not strongly pathogenic nor benign).^20^

**Figure 1:**
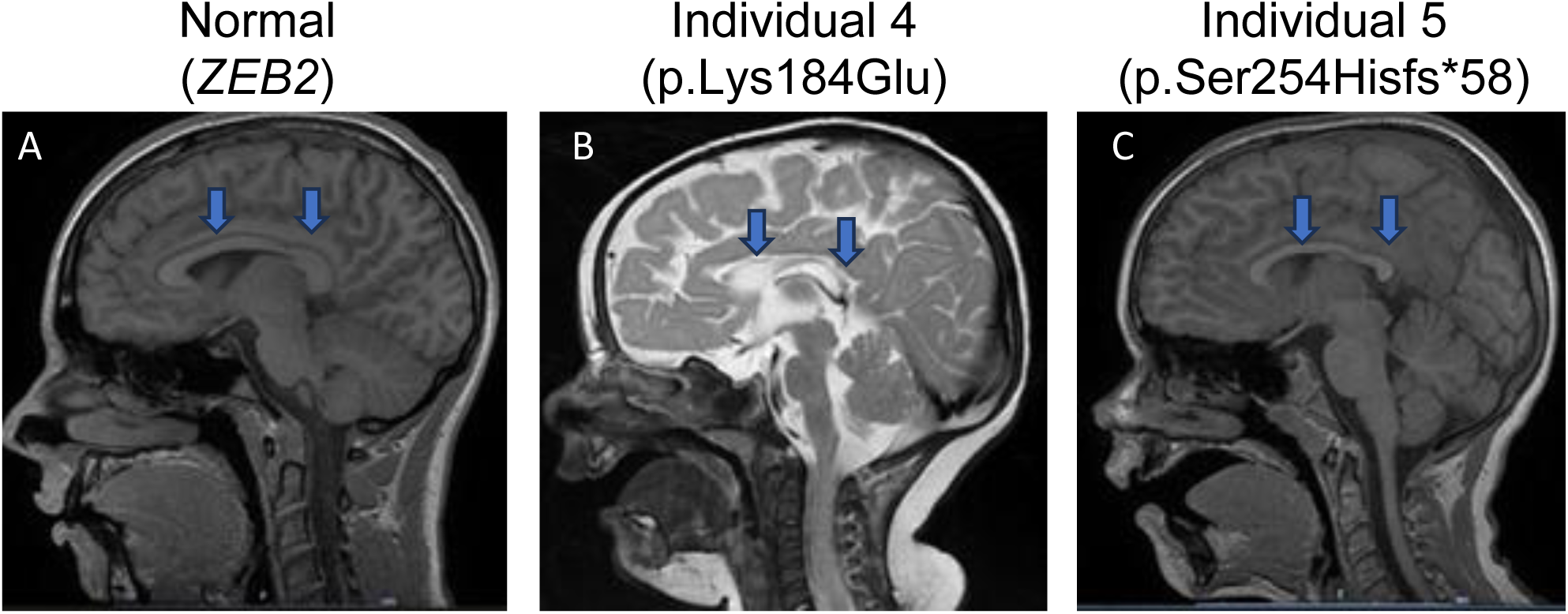
B**r**ain **magnetic resonance imaging (MRI) shows corpus callosum hypoplasia in two affected individuals with *TMEM184B* variants. A** Sagittal plane of spoiled gradient recalled acquisition (SPGR) MRI in an individual of the same age as individual 5 (Family 5-II-1) who has an unrelated pathogenic variant (*ZEB2*) and shows no alterations in corpus callosum thickness. **B** Sagittal T2 MRI in individual 4 (Family 4-II-1; p.Lys184Glu) shows corpus callosum disruption. **C** Sagittal SPGR MRI in individual 5 (Family 5-II-1; c.787+1G>A) shows corpus callosum hypoplasia.

To determine the nature of the variant c.787+1G>A containing a splice donor site mutation, we analyzed *TMEM184B* transcripts from iPSC derived from this individual and from control stem cells from a healthy individual. We consistently observed a 29 bp deletion within exon 7 and the use of a cryptic splice donor site at this position within exon 7 (**Figure S2**). This cryptic splice site is also found in prediction tools (SpliceAI) ^26^, suggesting that this alteration is the most likely result of the splice donor point mutation. The protein predicted from this mRNA contains a frameshift that disrupts the sequence of transmembrane domains 6 and 7 and causes a stop codon shortly thereafter (p.Ser254Hisfs*58).

### Human Missense Variants are Predicted to Alter Structure and Stability of TMEM184B Protein

*TMEM184B* is predicted to be transcribed in two major isoforms, with variant “a” (NM_012264.5) containing all possible coding sequences and producing a 407 amino acid protein. By mapping variants identified in affected individuals onto the protein, we found that variants cluster in portions of the gene that encode the transmembrane (TM) regions or the loop region between TM3 and TM4 (which is predicted to be cytoplasmic) (**Figure 2A**). The 3D structure of the native protein was predicted using AlphaFold, and the best model was selected based on the PLDDT scores and Ramachandran plot (**Figure 2B-D**). The Ramachandran plot shows the backbone dihedral angles in permitted regions, suggesting a high-quality structure. Figure 2D shows the transmembrane portions of TMEM184B predicted by this model, highlighting the 7 transmembrane domains found in the structure. We then built variants into this model and visualized their effects on the predicted structure (**Figure 2E-F**). Variants p.Val88Met and p.Lys184Glu show major disruptions to their transmembrane regions, with large shifts apparent both from top and side views.

**Figure 2.**
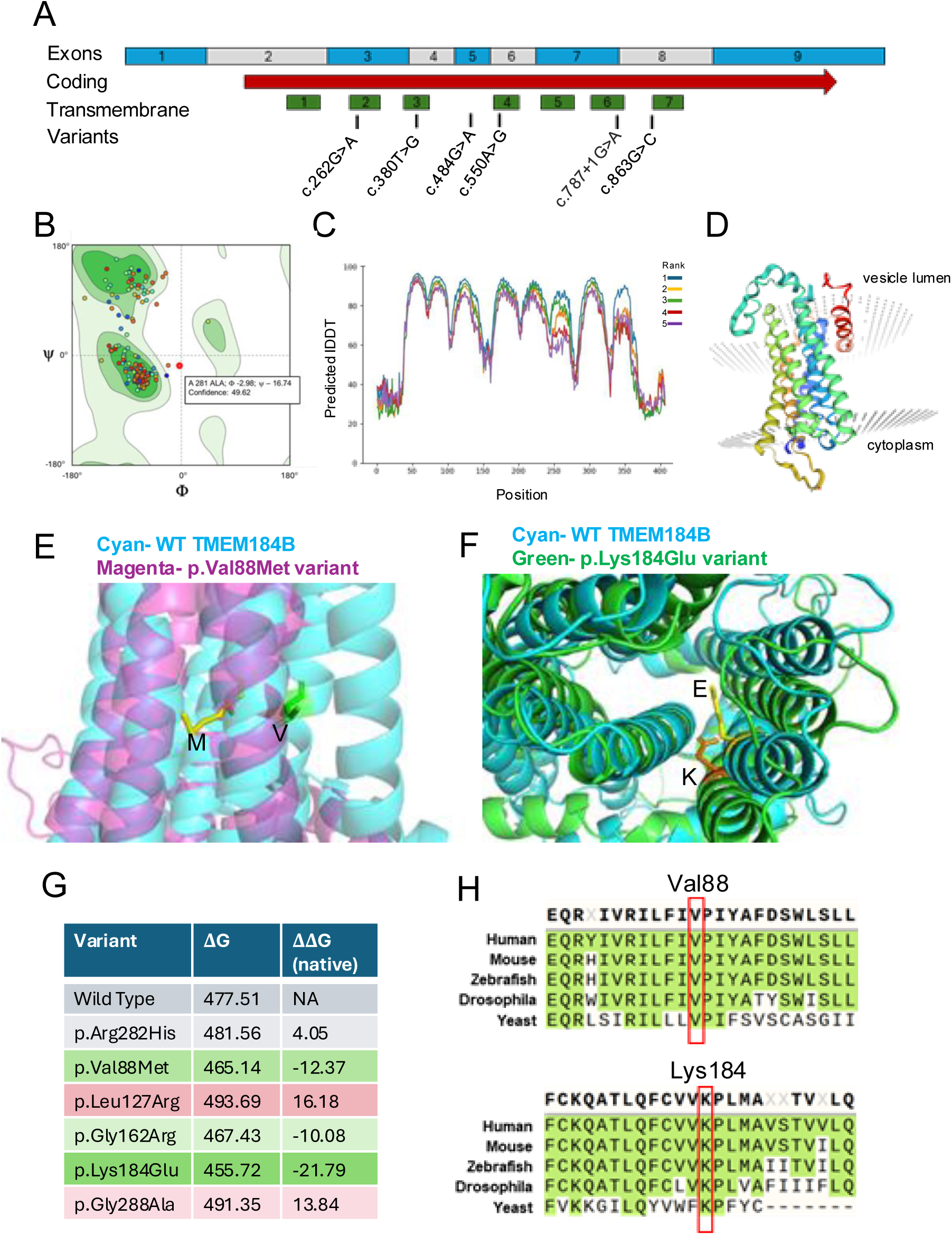
Structural modeling and free energy of TMEM184B variants indicates significant disruptions to protein stability. **A** Structure of the human *TMEM184B* transcript (NM_012264.5). Blue and gray blocks show alternating exons of the spliced mRNA. Red arrow indicates the coding region of isoform a (NP_036396.2). Areas predicted to be included in transmembrane helices (green) and locations of human variants (black) are noted. **B** Ramachandran plot showing that the structure predicted by FoldX analysis shows only allowable angles, providing confidence in the structure predicted. **C** Predicted local-distance difference test (pLDDT) scores per position across the whole protein sequence. Higher scores indicate higher confidence structural predictions in that region of the protein. **D** Native protein structure prediction by FoldX. Transmembrane helices are separately colorized. **E** p.Val88Met variant protein (magenta) overlaid onto the native TMEM184B structure (cyan). **F** p.Lys184Glu variant protein (green) overlaid onto the native TMEM184B structure (cyan). **G** Stability analysis calculations using Gibbs free energy. ΔΔG values that are positive indicate decreased protein stability of variant-containing TMEM184B when compared to the predicted native structure, while negative values indicate abnormally heightened protein structural stability. **H** Amino acid context for the p.Val88Met and p.Lys184Glu variants, both of which are conserved from yeast (*Saccharomyces cerevisiae* Hfl1) to humans. All other variants are conserved from *Drosophila* (Tmep) to human but are not found in yeast.

We undertook a quantitative analysis of protein structural stability of human TMEM184B using FoldX. The 3-D structures were used as input data for the Mac version of FoldX 5.0 to predict their ΔG. The stability of the protein structures was predicted using FoldX, where the native protein and each variant (p.Gly162Arg, p.Gly288Ala, p.Lys184Glu, p.Leu127Arg, p.Val88Met) underwent a stability analysis to calculate their respective total energies. We also performed this analysis for a commonly occurring TMEM184B sequence variant, p.Arg282His (dbSNP: rs370107940; 215 heterozygous and 4 homozygotes are present in 1,609,748 individuals in gnomAD v4.1.0), as a comparison. The change in Gibbs free energy (ΔΔG) was then determined for each variant relative to the native structure to assess the impact of the variants on protein stability. Variants with large positive ΔΔG values were associated with destabilizing variants, potentially leading to loss of function, while negative ΔΔG values suggested stabilizing variants that could result in gain of function or dominant negative effects (**Figure 2G**). The common variant from gnomAD, p.Arg282His, showed a small shift in ΔΔG, consistent with it being presumed benign. The loop region variant, p.Gly162Arg, shows a modest negative free energy change compared to wild type, perhaps due to its location in a more flexible loop region rather than a constrained transmembrane helix. We also analyzed the evolutionary conservation of the specific amino acids altered in affected individuals with neurodevelopmental defects. All amino acid locations show conservation throughout vertebrate and invertebrate animals; remarkably, Val88 and Lys184 are also conserved in the yeast ortholog, Hfl1 (**Figure 2H**). Overall, this analysis provides insight into the potential disruptive effects of the variants identified in affected individuals.

### The Zebrafish Ortholog of *TMEM184B* is Involved in Zebrafish Development

We and others have shown previously that zebrafish are a robust vertebrate developmental model that can inform not only physiological relevance to neurodevelopmental disease, but also variant effect. To test the role of *TMEM184B* in development, first we identified the zebrafish *TMEM184B* ortholog using reciprocal BLAST searches between human TMEM184B, GenBank ID: NP_036396.2, and the translated zebrafish genome. This query identified two zebrafish orthologs: *tmem184ba,* ENSDARG00000045147; 82% similarity, 80% identity and *tmem184bb*, ENSDARG00000090876; 77% similarity, 74% identity, versus human protein, respectively. Next, we used database and literature searches to find that expression of *tmem184ba* is markedly higher in whole larvae during the first five days of development than *tmem184bb* ^27^, and that *tmem184ba* has a ∼23-fold higher expression level in the zebrafish head at 3 dpf than its paralog^28^, and markedly more expression in neural cell groups within the first 5 days of development.^29^ Thus, we reasoned that *tmem184ba* is the most relevant ortholog of *TMEM184B,* and that its spatiotemporal expression in the zebrafish head supported a potential physiological role in neurodevelopment.

To evaluate a possible role for *tmem184ba* in formation and function of neural structures, we transiently suppressed its expression with a morpholino (MO) targeting the exon 6 splice donor site (e6i6; **Figure S3A**). Monitoring of mRNA splicing events in MO-injected larvae with RT-PCR and sequencing of cloned PCR products showed exon 6 exclusion, which removed 92 bp and resulted in a putative premature termination (**Figure S3B, C**). Next, we showed that injection of increasing doses of MO (1 ng, 2 ng, and 3 ng) into WT embryos resulted in a dose-dependent reduction in head size and body length in larvae subjected to automated live bright field imaging at 3 dpf, with the highest dose producing a ∼10-20% reduction in head area compared to either standard control MO or uninjected control (**Figure S3D, E, F, H, I**), supporting a role for *TMEM184B* in overall animal growth. To clarify whether head and body size reductions were proportionate, we calculated a ratio of head to body length in *tmem184ba* e6i6 morphants; we observed persistence of a dose-dependent and significant reduction, suggesting a more predominant defect in anterior structures (**Figure S3G**). Importantly, these observations were replicated with an independent MO targeting the exon 3 splice donor site (e3i3), which also resulted in a dose-dependent reduction in head size and body length at 3 dpf (**Figure S4**).

### *TMEM184B* is Involved in Neurodevelopment and *De Novo* Missense Variants Confer Dominant Toxic or Loss of Function Effects

Given that all variants in our cohort are heterozygous *de novo* changes, we hypothesized possible molecular mechanisms of dominant toxicity or haploinsufficiency. We and others have shown previously that ectopic expression of human mRNA in zebrafish larvae followed by quantitative phenotyping can inform dominant effects such as dominant negative (variant interacts with WT to produce a phenotype)^30^ or dominant toxic (variant gains a new function that is deleterious independent of endogenous WT).^31–33^ We used an established paradigm ^24^ to test for dominant effects by injecting a fixed dose (200 pg) of human *TMEM184B* WT and variant mRNA (p.Val88Met, p.Leu127Arg, p.Gly162Arg, p.Lys184Glu and p.Gly288Ala) in WT zebrafish embryos at the one to four cell stage and quantified the head and body size at 3 dpf as proxies for neurodevelopmental and growth deficits of affected humans. In replicate experiments, WT mRNA injection did not result in any detectable phenotypes compared to controls, but we observed a significant reduction in head size with concomitant reduction in body length for p.Gly162Arg and p.Lys184Glu larvae injected with an equivalent mRNA dose suggesting a dominant effect (**Figure 3A, B, C**). To exclude the possibility that these phenotypes were caused by developmental delay, we repeated microinjection experiments with 200 pg WT, p.Gly162Arg, or p.Lys184Glu-encoding mRNA and aged larvae to 4 dpf. All animals displayed a swim bladder, a developmental hallmark, and head size reduction remained significant for both variants compared to WT mRNA (**Figure 3D, E, F**).

**Figure 3.**
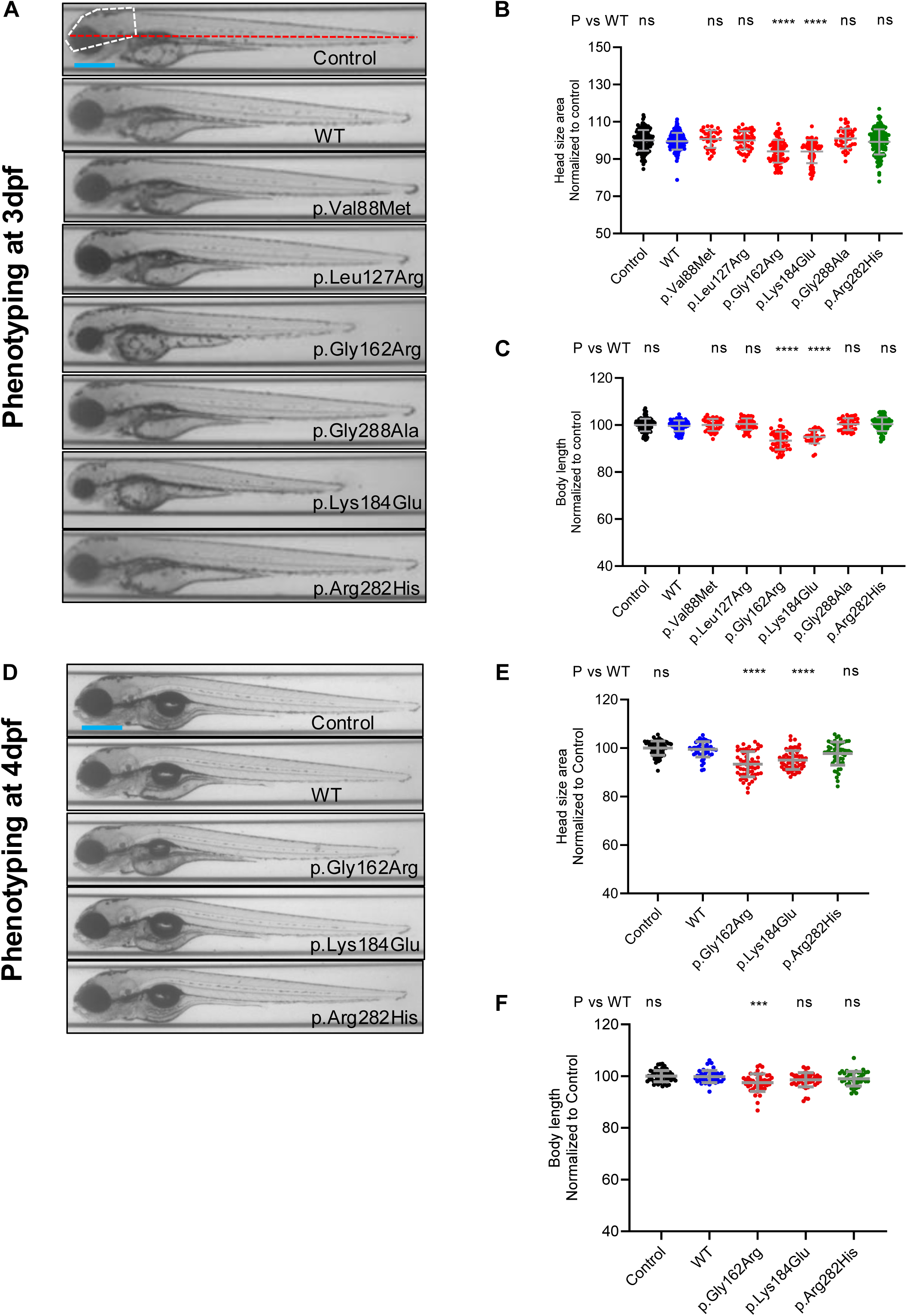
Ectopic expression of *TMEM184B* variants p.Gly162Arg or p.Lys184Glu induces head size and body length phenotypes in zebrafish larvae. **A** Representative bright field lateral images of 3-day post-fertilization (dpf) larvae injected with 200 pg human *TMEM184B* mRNA (WT, p.Val88Met, p.Leu127Arg, p.Gly162Arg, p.Lys184Glu, p.Gly288Ala and p.Arg282His). p.Arg282His is a presumed benign negative control (dbSNP: rs370107940; 215 heterozygotes and 4 homozygotes are present in 1,609,748 individuals in gnomAD v4.1.0). Scale bar, 300 μm. Head area was measured according to white dashed line; body length was measured according to the red dashed line. **B and C** Quantification of the (**B**) lateral head size and (**C**) body length in 3 dpf larvae; n = 35–87/condition. **D** Representative bright field lateral images of 4 dpf larvae injected with 200 pg human *TMEM184B* mRNA (WT, p.Gly162Arg, p.Lys184Glu, and p.Arg282His). Scale bar 300 μm. **E and F** Quantification of the (**E**) lateral head size and (**F**) body length in 4 dpf larvae. Data shown in panels E and F are combined from two biological replicates; n = 54-71/condition. For panels **B, C, E, and F,** statistical differences were calculated using one-way ANOVA with Tukey’s post-hoc test; ∗∗∗p<0.001; ∗∗∗∗p<0.0001 and ns, not significant. Experimental conditions were normalized and presented as a percentage of the control mean. Error bars represent standard deviation of the mean. Data point color: black, uninjected control; blue, WT mRNA; red, variant mRNA identified in affected individuals; green, presumed benign variant mRNA from gnomAD.

By contrast, overexpression of p.Val88Met, p.Leu127Arg, p.Gly288Ala, or p.Arg282His, the presumed benign variant, did not result in any detectable alteration of anterior structures or overall animal size at 3 dpf (**Figure 3A, B, C**). To ensure that all *TMEM184B* variant mRNAs were present in larvae, we used qRT-PCR to monitor the transcript in embryos at 24 hpf; we found a modest reduction from WT but relatively equivalent amount between case-associated variants and p.Arg282His (**Figure S5**). Together, these data indicate divergent variant effects.

To dissect further the possibility of dominant negative or dominant toxic (WT dependent versus independent variant effect, respectively), we co-injected embryos with progressively decreasing WT mRNA doses with progressively increasing variant mRNA doses while maintaining the overall human transcript amount constant (200 pg). Using the same phenotyping measures at 3 dpf, we found that for both p.Gly162Arg and p.Lys184Glu all three conditions injected with WT plus variant mRNA resulted in altered head size phenotype, but their effect was not significantly different from 200 pg variant mRNA alone, suggestive of a dominant toxic effect (**Figure S6A and C**). For body length measurements, we observed significant differences for WT-variant mRNA combinations versus 200 pg variant mRNA alone, but there was no consistent dose-dependent effect that could indicate a dominant negative effect (**Figure S6B and D**).

Although our *in vivo* data cannot resolve the precise molecular mechanism of variant action, they suggest that p.Gly162Arg and p.Lys184Glu confer pathogenic effects through WT-independent biochemical mechanisms.

Next, we pursued a loss of function hypothesis for p.Val88Met, p.Leu127Arg, and p.Gly288Ala by employing a complementation strategy, which compares rescue ability of human WT versus variant *TMEM184B* mRNA in the presence of MO.^24^ First, we established experimental parameters in which co-injection of WT mRNA restored larval head and body length phenotypes to measurements indistinguishable from controls (**Figure 4A, B, C**). Next, we compared the rescue of each variant mRNA to either MO alone or WT mRNA; we observed that each of p.Val88Met, p.Leu127Arg and p.Gly288Ala rescue both head size and body length phenotypes significantly worse than the WT or the negative control variant, p.Arg282His (**Figure 4A, B, C**). These data indicate that p.Val88Met, p.Leu127Arg and p.Gly288Ala confer reduced protein function consistent with a haploinsufficiency basis for disease. As expected, p.Gly162Arg and p.Lys184Glu variants also could not rescue MO effects on head size and body length.

**Figure 4.**
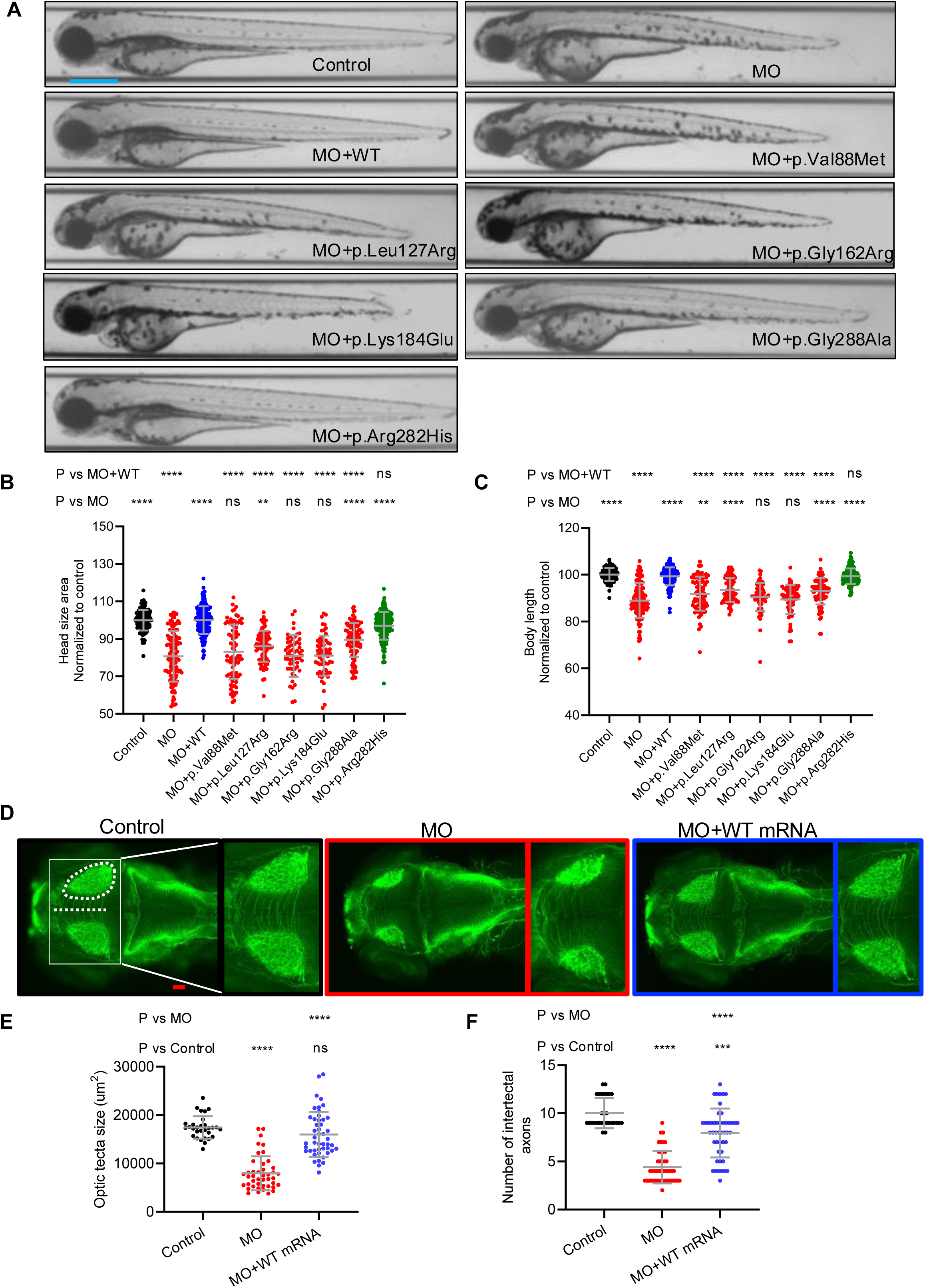
Loss of *tmem184ba* results in neuroanatomical defects and *in vivo* complementation shows that some disease associated variants result in loss of function. **A** Representative bright field lateral images of 3 dpf larvae injected with MO alone or co-injected with human *TMEM184B* mRNA (WT, p.Val88Met, p.Leu127Arg, p.Gly162Arg, p.Lys184Glu, p.Gly288Ala and p.Arg282His). p.Arg282His is a presumed benign negative control (dbSNP: rs370107940; 215 heterozygotes and 4 homozygotes are present in 1,609,748 individuals in gnomAD v4.1.0. Scale bar, 300 μm. **B and C** Quantification of the (**B**) lateral head size and (**C**) body length in 3 dpf larvae as indicated with anatomical landmarks in Figure 3A. Experimental conditions were normalized and presented as a percentage of the control mean. For panels B and C, n = 50–88/condition. **D** Representative dorsal images of 3 dpf larval heads immunostained for with acetylated tubulin. White dotted shape, optic tecta; white dotted line, midline between optic tecta where intertectal commissural axons were counted. Zoomed image at right of each panel corresponds to white box around both optic tecta. Anterior, left; posterior, right; Scale bar, 50 μm. **E** Quantification of optic tecta size as shown in panel D. **F** Quantification of intertectal neurons that cross the dorsal midline as shown in panel D. For panels E and F, n = 30–47/condition. For panels B, C, E and F, statistical analyses were performed with a one-way ANOVA with Tukey’s post-hoc test; ****p<0.0001; ***p<0.001; **p<0.01, and ns, not significant. Error bars represent standard deviation of the mean. Data point color: black, uninjected control; blue, WT mRNA; red, variant mRNA identified in affected individuals; green, presumed benign variant mRNA from gnomAD.

To further demonstrate specificity of the zebrafish rescue assay, we tested five additional variants reported in gnomAD v4.1.0. These variants were selected because they are either proximal to p.Val88Met in the second transmembrane domain (p.Ile84Thr and p.Ala92Ser), have similar frequency to p.Val88Met in gnomAD (p.Val54Met and p.Val114Ile) or are present in homozygosity (p.Pro28Leu), similar to p.Arg282His (**Figure S7A**). Morphometric analysis of larvae at 3 dpf showed four of five variants to be indistinguishable from WT rescue, while the remaining variant (p.Ala92Ser, present in 4 individuals in gnomAD v4.1.0) resulted in complete rescue of body length but modest head size rescue that was still significantly worse than WT mRNA (**Figure S7B, C, D)**. We extended the 3-D protein stability analysis to these five variants using FoldX to predict their ΔΔG vs WT. Whereas the five variants that scored as benign in the zebrafish rescue assay (including p.Arg282His) had ΔΔG values +/-10.95 or less, p.Ala92Ser had the largest positive ΔΔG (18.2) in a range similar to the disease-associated variants (**Figure 2G**). Together, these observations bolster the specificity of the zebrafish complementation test when applied to *TMEM184B*, raise the possibility of individuals in gnomAD who harbor pathogenic *TMEM184B* variants but might not show a phenotype due to incomplete penetrance or genetic background, and support p.Val88Met as a major disease contributor for individual 1 (Family 1-II-1).

To achieve further resolution into the neuroanatomical phenotypes induced by *tmem184ba* impairment, we performed whole mount staining of 3 dpf zebrafish larvae with acetylated tubulin to demarcate optic tecta and intertectal neurons, locations that serve as established proxies for microcephaly and corpus callosum defects in humans.^34,35^ In biological triplicate experiments, we observed a significant reduction in optic tecta size for morphants compared to controls, a phenotype that could be rescued significantly by co-injection of WT human mRNA (**Figure 4D, E**). Additionally, we counted the number of neuronal axon tracts crossing the dorsal midline and found a significant reduction in morphants, which could also be rescued by co-injection of WT human *TMEM184B* (**Figure 4D, F**). These data show discrete neuroanatomical features impaired by *tmem184ba* depletion beyond overall head and animal body growth and further demonstrate reagent specificity with rescue of MO-induced phenotypes with WT human mRNA.

### Transcriptomic Analysis of a Severe Missense Variant Identifies Alterations in Developmental and Metabolic Gene Signatures

To broadly evaluate gene expression changes that could be driving alterations in cells containing monoallelic variants, we created three iPSC lines. The first line was derived directly from fibroblasts from individual 5 (Family 5-II-1; c.787+1G>A, nonsense allele). The remaining two lines were created using CRISPR/Cas9 gene editing on a healthy control line from an iPSC biobank (Coriell).

We modified the genome to create the c.550A>G variant (encoding p.Lys184Glu). While we intended to modify only one allele, we also identified clones containing biallelic c.550A>G modification, which we reasoned could provide additional support for identification of TMEM184B-dependent cellular and molecular pathways. Of note, at this time we do not yet have an isogenically corrected control for the line from individual 5 (Family 5-II-1). We therefore focused our transcriptomic comparisons on the c.550A>G monoallelic and biallelic missense variants compared to their isogenic control. Principal component analysis showed that variants cluster together, suggesting their expression profiles are tightly linked to genotype (**Figure 5A**). We observed both up-and down-regulation of a large number of genes (**Figure 5B**). Using a threshold for significance (adjusted p < 0.05) we detected 496 genes that were over-expressed (fold change (FC) > 1.5) and 668 genes that were under-expressed (FC < 0.75) in the heterozygous variant line (**Table S3**). In the homozygous variant line, additional genes were dysregulated (1478 up, 1663 down) (**Table S4**). Gene Ontology (GO) analysis indicated that among downregulated genes, 12 biological processes were significantly enriched (**Figure 5C-D**). Processes related to transporter activities across membranes and protein and lipid metabolism were decreased. We found significant decreases in axon guidance pathways, which could point to a mechanism for disrupted commissural axon crossing in the zebrafish assays. Interestingly, we found that cellular regulation of pH was the most downregulated biological process. The heatmap (**Figure 5D**) shows genes in this cluster, including four SLC9 (NHE) transporter family members (which function as Na^+^/H^+^ exchangers) ^36^, three SLC4 family members (which function as H+/HCO3-transporters) ^37^, and an accessory subunit of the vesicular ATPase proton pump (ATP6AP1).^38^ Among upregulated processes, the most enriched pathway was negative regulators of TOR signaling (**Figure 5E-F**). Most biological processes were disrupted in both the heterozygous and homozygous c.550A>G cells.

**Figure 5.**
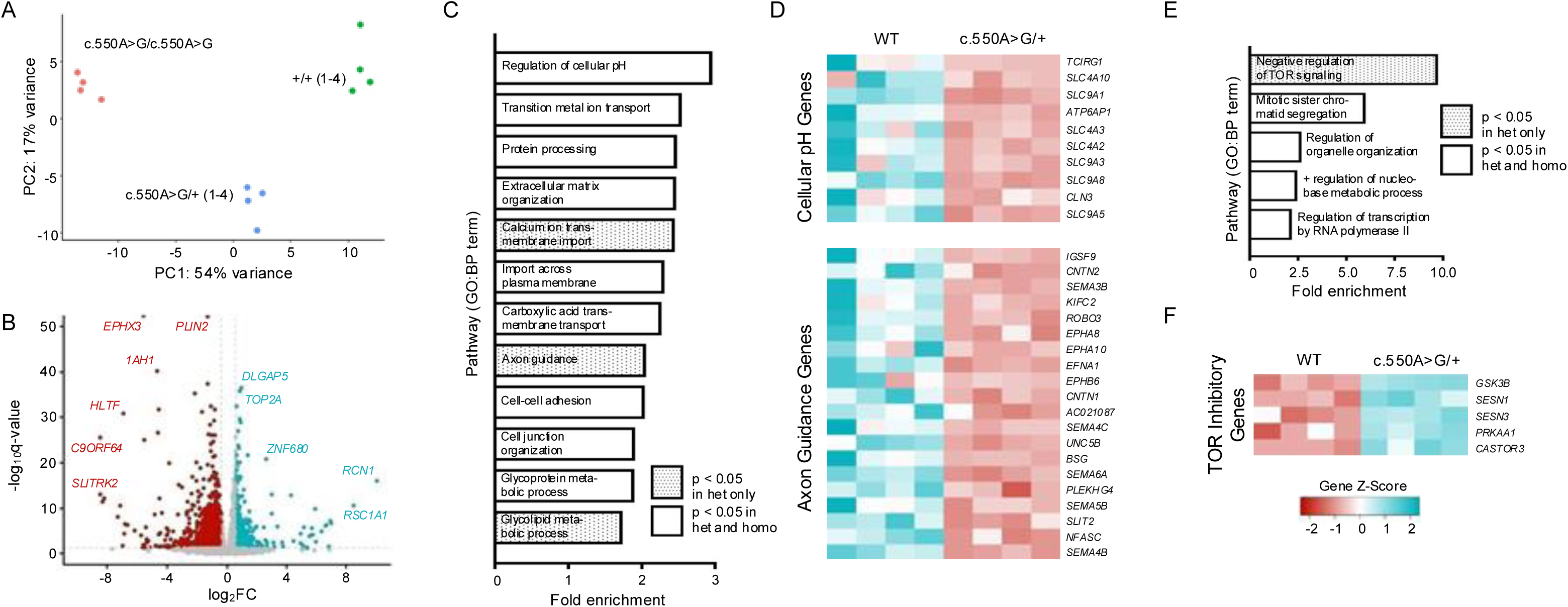
Transcriptomic analysis of variant-containing human iPSC reveals alterations in neurodevelopmental pathways and endolysosomal function. **A** Principal Component Analysis (PCA) plot of RNA-seq data generated using DESeq2. Data were derived from induced pluripotent stem cell (iPSC) lines representing three groups *(N =* 4 biological replicates per group): wild type (WT), *TMEM184B*^c.550A>G/+^, and *TMEM184B*^c.550A>G/c.550A>G^. **B** Volcano plot illustrating the differential expression analysis results for *TMEM184B*^c.550A>G/+^. Genes meeting the significance threshold (p < 0.05) and fold change criteria (log2FC > 0.585 for upregulated genes [=FC>1.5], log2FC <-0.415 for downregulated genes [=FC<0.75]) are highlighted in teal (upregulated) and red (downregulated), respectively. **C** Downregulated GO Biological Processes (Slim) (>1.5x enriched) in *TMEM184B*^c.550A>G/+^. **D** Genes in two processes of likely relevance (regulation of cell pH and axon guidance) are plotted in respective Z-score normalized heatmaps. **E** Upregulated GO Biological Processes (Slim) (>1.5x enriched) in *TMEM184B*^c.550A>G/+^.**F** Heatmap of genes within one process of likely relevance (negative regulation of TOR signaling), scaled identically to D. Abbreviations: het, heterozygous; homo, homozygous; (+), wild type allele.

### TMEM184B Missense Variants Affect Protein Levels and Cell Viability

To further analyze the cellular consequences of disease-linked *TMEM184B* variants, we examined protein stability. Because antibodies to TMEM184B are unreliable in cells (M.R.C.B., personal communication) we modeled variant effects by creating mi670-tagged expression constructs of human *TMEM184B* containing either the WT form, a variant predicted to be benign (p.Arg282His), or a variant matching one of the five disease-associated missense variants. We hypothesized that, given the differences in structural stability between variants in our *in silico* modeling, variant proteins may accumulate at lower levels even when driven by the same promoter. Our analysis showed that, with one exception (p.Gly162Arg), all case-associated variant proteins show reduced overall levels compared to WT (**Figure 6A-C and Figure S8**), even those predicted to be hyper-stabilized. We suspected that this result reflects increased degradation of structurally compromised proteins and/or death of cells containing high levels of variant protein. To examine viability of cells expressing variants of *TMEM184B*, we analyzed apoptosis frequency in human cells transiently transfected with WT, each of the five *TMEM184B* case-associated missense variants, and an additional benign control missense variant (p.Arg282His, as done in zebrafish). We then analyzed apoptosis using a flow cytometry assay based on accumulation of Zombie Violet, an amine-reactive dye, in dead cells. In normal media, cells overexpressing p.Lys184Glu and p.Gly288Ala showed increases in cell death of 2-7x compared to cells producing ectopic WT TMEM184B (**Figure 6D-E**). For p.Lys184Glu, this increase in cell death continued after starvation (**Figure S8B-C**). It is possible that the cells with highest variant levels are those undergoing apoptosis, which could contribute to the reduced protein levels we see in the population-wide analysis. However, it is also plausible that these are independent variant effects. Taken together, our results show that some *TMEM184B* variants have reduced protein stability and can increase apoptosis of human cells.

**Figure 6.**
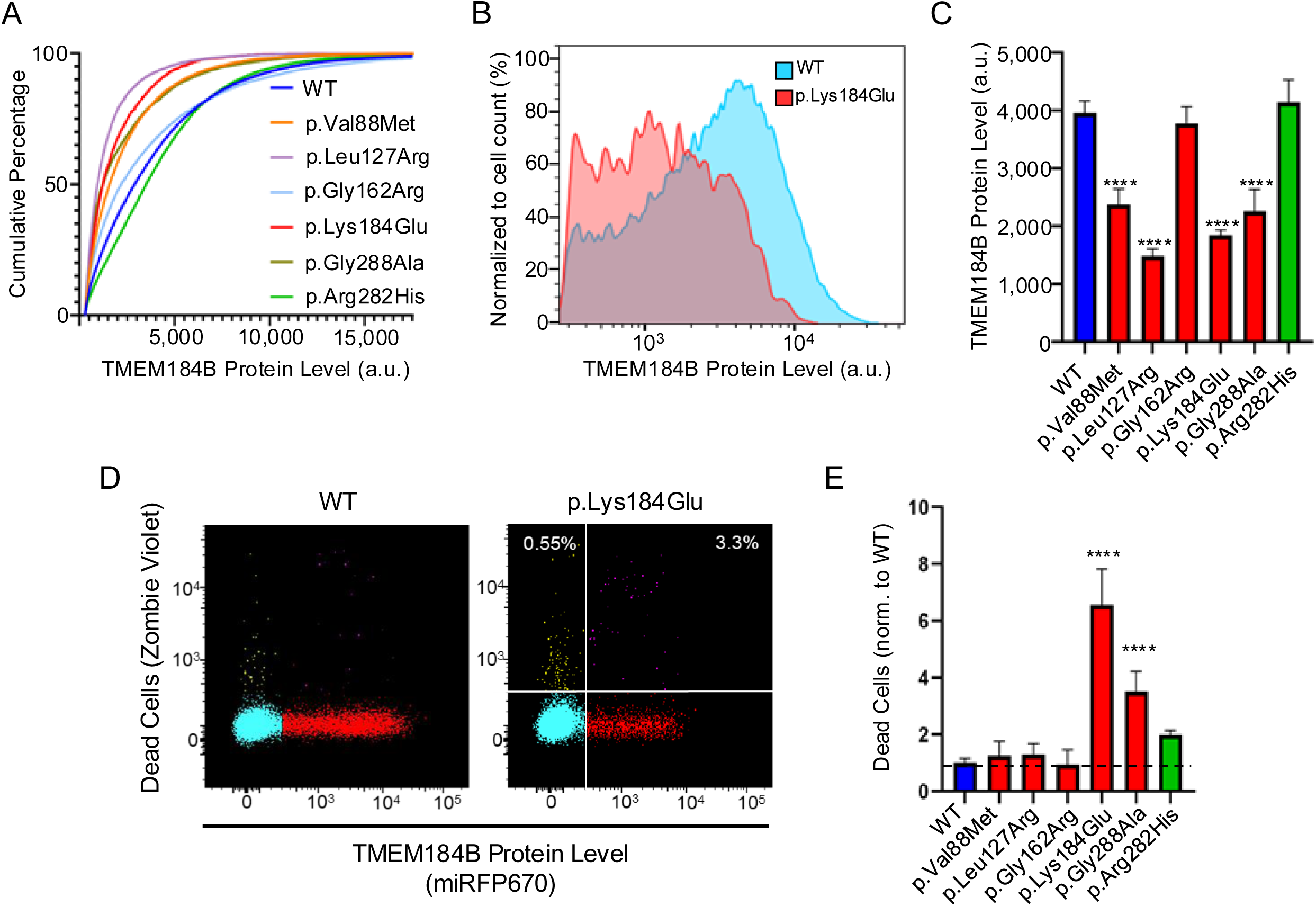
*TMEM184B* variants affect protein levels and cell viability. **A** Cumulative distribution function of cells expressing WT and *TMEM184B* variants tagged with *miRFP670*. **B** Histogram of the fluorescent levels of individual cells expressing p.Lys184Glu tagged with *miRFP670* were grouped into 256 bins and normalized to the percentage mode protein level of each population. **C** Average raw fluorescence of TMEM184B variants compared to WT (N=3 trials). Error bars represent SD. **D** FACS plots of HEK293T cells over-expressing native *TMEM184B* (left) or p.Lys184Glu variant (right) and grown in complete media. White numbers on the plots indicate the percentage of dead cells among untransfected (upper left) or *TMEM184B*-transfected HEK293T cells (upper right). **E** Percentage of dead *TMEM184B* variant expressing cells normalized to WT levels (dashed line). N=3 trials. Error bars represent SD. ****p<0.0001.

### Variant Expression Disrupts Shuttling of the Lysosomal Biogenesis Transcription Factor EB (TFEB)

Signaling pathways that operate downstream of TMEM184B are unknown, but prior work suggests an intersection between these pathways and the processes of synaptic growth, proliferation, and autophagy.^1,2,7^ In our RNAseq analysis, we identified disrupted metabolic processes in variant-containing cells, including disruptions in pH regulation and mTOR signaling. These two processes are linked; for example, if lysosomal pH is too high (alkaline), mTOR complex 1 (mTORC1) is inhibited. mTORC1 inhibition causes transcription factor EB (TFEB) to become dephosphorylated and enter the nucleus to transcribe lysosomal and autophagosomal genes, which promotes recovery from metabolic stress.^16^

Knockdown of *TMEM184B* expression in cell lines alters the starvation-induced distribution of TFEB, causing prolonged nuclear retention.^39^ We therefore postulated that case-associated variants might also alter the nuclear-to-cytoplasmic distribution of TFEB. We generated HEK293T cell lines stably expressing *TFEB-GFP* and *H2B-mRuby* (nuclear marker) and analyzed the nuclear:cytoplasmic ratio of TFEB-GFP in the presence of each TMEM184B case-associated variant. In normal (unstarved) conditions, p.Lys184Glu expression increased the nuclear:cytoplasmic ratio of TFEB (**Figure 7**). Other variants did not significantly alter TFEB, suggesting that their presence does not induce cellular stress. TFEB localization in the presence of p.Lys184Glu continued to be altered under starvation conditions (**Figure 7N, Figure S9**).

**Figure 7.**
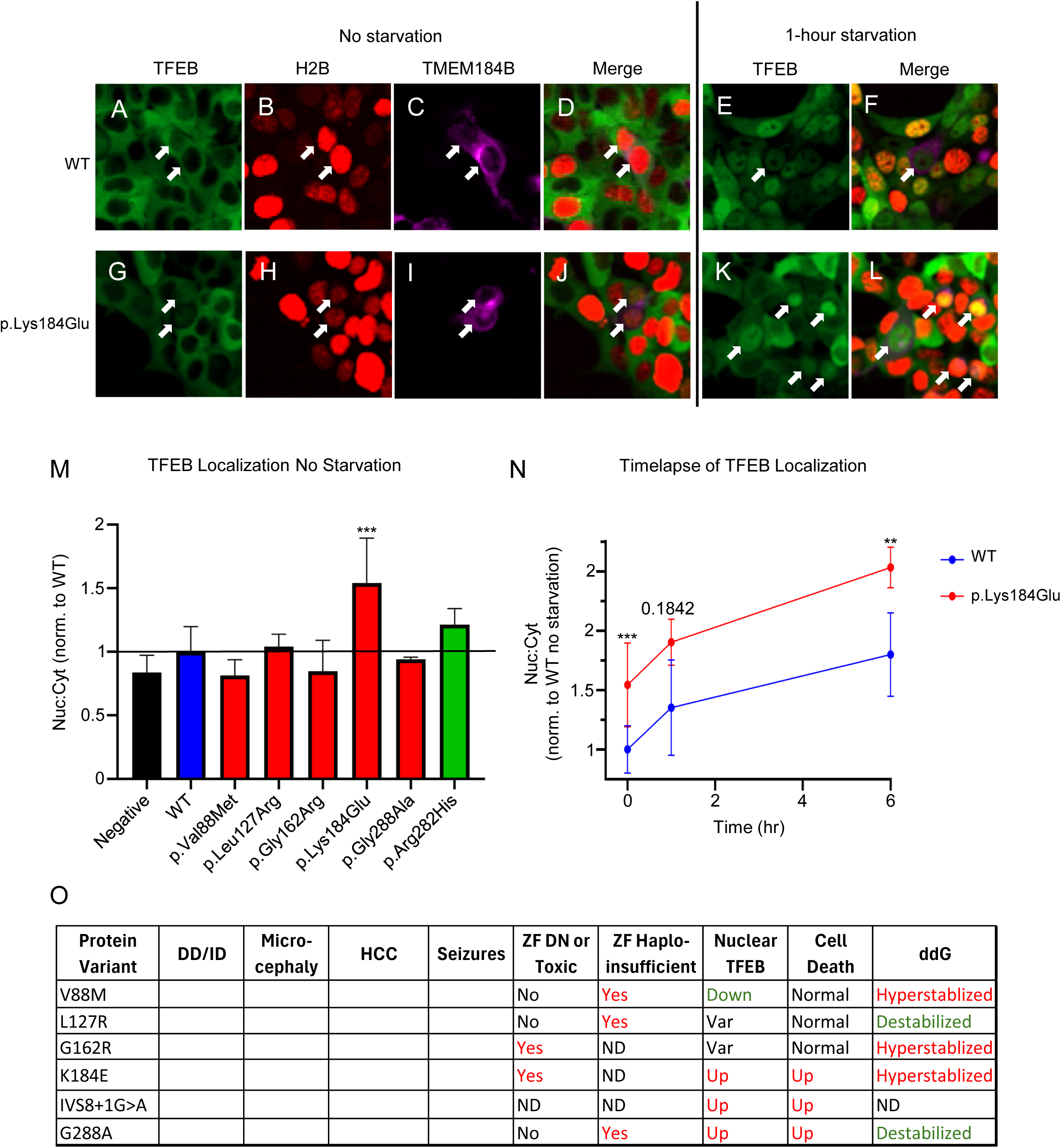
TFEB nuclear-to-cytoplasmic ratio is dominantly altered by expression of *TMEM184B* variants identified in affected individuals. **A-D** Multichannel images of unstarved HEK293T cells overexpressing WT *TMEM184B*. Panels shown are (**A**) TFEB-GFP (green), (**B**) H2B-mRuby (red), and (**C**) TMEM184B-miRFP670 (magenta). **D** is a merged image. **E-F** HEK293T cells expressing WT *TMEM184B* and starved for one hour. Panels show (**E**) TFEB-GFP and (**F**) merge (includes H2B:mRuby and MEM184B-miRFP670). Arrows in all images point to TMEM184B-miRFP670 transfected cells. All H2B-mRuby images had their Lookup Tables adjusted to 4000 for visual clarity. **G-J** Multichannel images of unstarved HEK293T cells over-expressing *TMEM184B* p.Lys184Glu variant. Panels match those of the WT in A-D. K-L *TMEM184B-miRFP670* transfected cells starved for 1 hour. TFEB-GFP and a merged image (TFEB-GFP, H2B:mRuby, and TMEM184B-miRFP670) are shown. **M** Ratio of mean TFEB-GFP fluorescence in the nucleus vs. cytoplasm in *TMEM184B-miRFP670* expressing cells, normalized to WT cells. **N** Time course of TFEB nuclear:cytoplasmic localization of WT and p.Lys184Glu expressing cells over six hours of starvation (N=3 trials). M and N error bars represent SD. ***p<0.001, **p<0.01. 50 cells were quantified per trial, variant, and time point except for p.Lys184Glu at six-hour starvation (42-50 cells). **O** Summary table displaying the clinical, free energy, zebrafish, and cell assay results of each case-associated variant. Red text denotes differences from the WT condition. In the “Free Energy Difference” column, red text indicates destabilization and green text indicates hyperstabilization.

To identify concordance between our zebrafish and cellular findings with phenotypes of affected individuals, we provide a table aligning clinical features and discoveries from our models (**Figure 7O**).

## DISCUSSION

Here we present the consequences of *TMEM184B* rare pathogenic variants on human neural development and signaling. Except for individual 1, who harbors the *de novo* p.Val88Met variant, all other changes identified in our cohort were absent from public databases. We are cautious in the clinical interpretation of p.Val88Met, however the structural modeling and zebrafish variant testing data suggest that this change contributes to this child’s phenotype, possibly in concert with another yet to be characterized pathogenic lesion in individual 1’s genome. While each affected individual’s presentation has some unique aspects, all six variants we characterized result in shared clinical features including developmental delay, structural brain disruptions (hypoplastic corpus callosum and/or microcephaly), and/or seizures, indicating that nervous system disruption is a central result of altered TMEM184B function. In addition to these central nervous system effects, two affected individuals also report gastrointestinal motility disruption, which we suspect could result from enteric innervation pathology. We anticipate that as more *TMEM184B* disorder-linked variants are reported, they will produce scalar modifications to these core nervous system phenotypes.

Our zebrafish data show that currently known *TMEM184B* variants cluster into two functional categories. Variants p.Gly162Arg and p.Lys184Glu appear to work via dominant effects, given their ability to induce microcephaly when over-expressed in zebrafish larvae. Dominant toxic classification for p.Lys184Glu is also supported by cellular results showing that p.Lys184Glu over-expression in HEK293T cells causes increased TFEB nuclear localization both before and during starvation and also increases apoptosis. In contrast, variants p.Val88Met, p.Leu127Arg, and p.Gly288Ala appear to compromise TMEM184B function and thus cannot rescue phenotypes of TMEM184B knockdown. How these divergent effects are produced remain obscure. No variant described here can fully rescue head size and body length. We hypothesize that these failures are due to disruptions to cellular growth pathways across the body, with stronger effects in the head given the enriched expression of *TMEM184B* in this location (GTEx portal). As we gain additional information about the molecular function of TMEM184B in different cell types in the nervous system, we should be better able to understand the sequence requirements for TMEM184B protein function and developmental effects.

Overall, our data align with prior work showing the effects of significant loss of function of TMEM184B in both *Drosophila* and mice. While the overall size of the developing mouse brain has not been evaluated, mice with a genetic disruption of *Tmem184b* (95% reduction in transcript levels) show abnormally swollen intraepidermal nerve fibers (endings of sensory neurons) and swollen motor neuron presynaptic terminals at neuromuscular junctions.^1^ Similarly in flies, mutation of the *TMEM184B* ortholog *tmep* causes abnormal presynaptic sprouting.^2^ This may suggest that, in variants that confer a dominant effect such as p.Lys184Glu, synaptic structure may be compromised. Future work should examine variant effect on synapses *in vivo* using knock-in models of heterozygous variant expression, matching the genotype and zygosity of affected individuals.

While initial studies in rodent and fly models suggested a link to neurodegenerative diseases like ALS, our cohort highlights that the nervous system disruptions caused by *TMEM184B* alteration happen much earlier. TMEM184B-associated syndromes resemble other juvenile-onset disorders of abnormal metabolism including the lysosomal storage disorder Niemann-Pick Type C (NPC; MIM: 257220) caused by pathogenic variation in one of two lipid transporter genes (*NPC1* [MIM: 607623] or *NPC2* [MIM: 601015). Cellular phenotypes of NPC include accumulated lipids in the endolysosomal system, similar to what is seen in mouse models of TMEM184B loss-of-function.^40–42^ In mice containing a disease-associated *NPC1* variant (p.Asp1005Gly)^43^, TFEB is erroneously enriched in the nucleus, similar to what we find in the presence of some *TMEM184B* case-associated variants. These commonalities suggest that the pathways leading to NPC and *TMEM184B*-associated disorders share some downstream effects. Other rare disorders also cause aberrant TFEB nuclear translation, for example Mucolipidosis type IV (MLIV, MIM: 252650). Another similar disorder with corpus callosum hypoplasia, intellectual disability, and seizures is caused by loss of MYCBP2 (MIM: 610392; also called PHR1 (mice), Highwire (*Drosophila*) or Rpm-1 (*C. elegans*)), a RING finger ubiquitin ligase known for roles in synapse development.^44–46^ TMEM184B and MYCBP2/Phr1/Highwire mutants both show excessively branched synapses and delays in injury-induced axonal degeneration, though the effect of TMEM184B loss is more subtle.^1,2,47,48^ It will be of interest to explore the intersections of these proteins in the context of neural development as well as axon degeneration.

In human cells expressing the p.Lys184Glu variant of *TMEM184B*, the increase in TFEB nuclear localization is likely a compensatory response to nutrient deprivation or disrupted autophagosomal or endolysosomal flux (here used to describe the fusion and maturation of these compartments, both of which end up depositing contents into lysosomes). The p.Lys184Glu variant-expressing cells are also more likely to undergo apoptosis, even under nutrient rich conditions. One plausible scenario is that TMEM184B participates in autophagic or endolysosomal flux, ensuring lysosomal cargo degradation and promoting cellular survival. Indeed, in a gene-trap mouse with TMEM184B disruption, cortical neurons show decreased assembly of the vesicular ATPase (v-ATPase) and a corresponding increase in late endosomal and lysosomal pH which would disrupt lysosomal enzyme function.^49^ In this scenario, mTOR activity would be reduced by TMEM184B disruption, and TFEB would show higher nuclear localization, consistent with our results in human cells. Variants like p.Lys184Glu that severely disrupt TMEM184B stability and function would therefore cause excessive cell death during development and could directly lead to microcephaly or other severe disease presentation. Indeed, microcephaly can be caused by reduced proliferation of neural progenitors or by reduced cellular viability.^50,51^ Variants that more subtly alter flux in the cell could impair axon outgrowth pathways necessary to establish the cross-hemispheric connections of the corpus callosum.

Our structural modeling of each of the known case-associated missense variants indicates substantial alterations in the alignment of alpha-helices, and consequent changes in overall protein stability. Here also we find two categories of changes: three variants (p.Lys184Glu, p.Gly162Arg, and p.Val88Met) overly stabilize TMEM184B (indicated by negative values <-10), while two variants (p.Leu127Arg and p.Gly288Ala) destabilize the protein (positive values > 10). With one exception (p.Val88Met), these structural predictions align with the classifications of variants as causing dominant effects (p.Lys184Glu and p.Gly162Arg) or haploinsufficiency (p.Leu127Arg and p.Gly288Ala). TMEM184B is annotated as a putative transporter,^52–54^ though the transported substance is as yet unknown. If we consider this possible function in our working model, over-stability of a transporter would likely be quite deleterious, as it could be locked into a conformation that would prevent the flexibility needed to facilitate movement of its transport substrate. It is also possible that p.Lys184Glu may disrupt cells by creating “leaky” transporters that equilibrate their substrate across endosomal membranes. Gradients of ions (like H^+^) set the proper pH inside endosomal and lysosomal compartments, which is important for proper fusion, flux, and degradation of contents.^55^ If these gradients were disrupted, autophagosomes and lysosomes would not be able to effectively degrade their contents, a result matching what is seen when TMEM184B is disrupted in mice.^1^ With respect to p.Gly162Arg, this change is located in a loop region predicted to face into the cytoplasm and which contains 2 likely phosphorylation sites (Ser152 and Thr164), one of which is only two residues away from the variant location.^23^ We therefore predict that this region is likely to serve an as yet unknown signaling function for the protein, and that the dominant behavior of the p.Gly162Arg variant in zebrafish reflects an increase or decrease in this signaling.

We see substantial disruption of the transcriptome by TMEM184B p.Lys184Glu substitution, even when a WT *TMEM184B* allele is present. In iPSC containing this variant, transporters and regulatory molecules controlling H+ cellular distribution are downregulated, while negative regulators of the mTOR pathway are upregulated. These findings suggest that disrupted compartment pH and metabolic disruption contribute to *TMEM184B* variant-associated clinical sequelae. We also see reduced expression of transcripts in pathways that promote axon outgrowth.

While our RNAseq analysis was performed in undifferentiated stem cells, these results indicate that molecules needed for axon growth are impacted by TMEM184B function and align with the finding of corpus callosum hypoplasia in a group of individuals with *TMEM184B* sequence variation.

Our study demonstrates the connection between *TMEM184B* sequence variation and human neurodevelopmental disease and provides mechanistic information about how variants dominantly disrupt protein structure and metabolic signaling. However, many details remain unknown. For one, our predictions of the molecular function of TMEM184B are based solely on sequence and structural homology. These studies suggest that TMEM184B and its related family members (TMEM184A and TMEM184C) belong in a large and diverse family including transporters, opsins, and G-protein coupled receptors.^54^ We also need to better understand how TMEM184B activity and TFEB signaling are connected. TFEB nuclear translocation is inhibited by phosphorylation, which can occur through at least four different kinase pathways: mTORC1, ERK2, Akt, and GSK3β.^56^

Interestingly, we find that *GSK3*β transcripts are downregulated by the p.Lys184Glu substitution, consistent with our observation of increased TFEB nuclear localization and possibly suggesting a mechanism for this change. Determining which pathways are functionally disrupted by variation in *TMEM184B* and whether these are direct or indirect effects should be a high priority for future study.

A caveat in our modeling of *TMEM184B* associated disorders is that our zebrafish and HEK293T models do not perfectly reproduce the human inheritance pattern. All affected individuals show heterozygous expression of a variant form alongside a normal protein. While the dominant effects of the variants studied here permitted us to investigate their effects via transient expression in WT backgrounds, the overall dosage of TMEM184B is higher than normal in these scenarios. Our initial exploration of transcriptomic changes in heterozygous iPSC lines is an important first step in improved modeling of human *TMEM184B* variants, and indeed this analysis found expression changes that likely influence nervous system growth and development in humans with matching genomic context (monoallelic variation). Future work to build additional case-associated heterozygous variants in model organisms or stem cell lines should expand our understanding of the effects of TMEM184B in key steps of neuronal differentiation and axon guidance.

### Research Contributions

M.R.C.B., E.E.D., S.B. and K.A.C. initiated the project. K.A.C. coordinated the organization of clinical information from the cohort of affected individuals. F.U. and S.K. performed all zebrafish modeling experiments and data analysis. Z.Y. performed all human cell experiments and data analysis. H.G.W. and A.P. performed DNA and RNA analysis from stem cell lines. S.V.K. and S.S. performed structural modeling and analysis. S.P.C., T.S., C.S., N.M., H.F., E.T., J.T.-S., A.A., V.Sciruicchio, V.Salpietro, P.S., C.M., B.K., D.H. and S.C. contributed clinical data for the case cohort. K.P. helped create reporter cell lines and assisted with data analysis. Z.Y., F.U., K.A.C., S.S., E.E.D., and M.R.C.B. wrote the paper.

## Data Availability

All data produced in the present work are contained in the manuscript.

https://www.ncbi.nlm.nih.gov/geo/query/acc.cgi?acc=GSE289220

## Acknowledgments

We thank Dr. Curtis Thorne for access to his widefield microscope, staff of the University of Arizona Flow Cytometry Shared Resource, and staff of the Nikon Imaging Center of Excellence at the University of Arizona for their support. We thank Chloe Reuter for communicating with our team about an additional affected individual. Funding for this work was provided by NIH R01 NS105680 (M.R.C.B.), NIH R01 MH106826 (E.E.D.) and a Core Facility Pilot Award from the University of Arizona (M.R.C.B.). We would also like to thank all the subjects and their families for participating in this research. E.E.D. is the Ann Marie and Francis Klocke, M.D. Research Scholar.

This manuscript is the result of funding in whole or in part by the National Institutes of Health (NIH). It is subject to the NIH Public Access Policy. Through acceptance of this federal funding, NIH has been given a right to make this manuscript publicly available in PubMed Central upon the Official Date of Publication, as defined by NIH.

## Data and Code Availability

Transcriptomic datasets can be found in the NCBI Gene Expression Omnibus (GEO) database under GSE289220.

## Web Resources

BLAST, https://blast.ncbi.nlm.nih.gov/Blast.cgi Ensembl, http://useast.ensembl.org/index.html Galaxy Server, https://usegalaxy.org/ GenBank, https:// https://www.ncbi.nlm.nih.gov/

Gene Expression Omnibus (GEO), https://www.ncbi.nlm.nih.gov/geo/ gnomAD, https://gnomad.broadinstitute.org

GPS6.0, https://gps.biocuckoo.cn

GTEx Portal, https://gtexportal.org/home/gene/TMEM184B#gene-transcript-browser-block

Heatmapper, http://heatmapper.ca/

ImageJ, https://imagej.net/ij/

RefSeq, https://www.ncbi.nlm.nih.gov/refseq/Uniprot, https://www.uniprot.org

## REFERENCES

1. Bhattacharya, M.R.C., Geisler, S., Pittman, S.K., Doan, R.A., Weihl, C.C., Milbrandt, J., and DiAntonio, A. (2016). TMEM184b Promotes Axon Degeneration and Neuromuscular Junction Maintenance. J. Neurosci. 36, 4681–4689. 10.1523/JNEUROSCI.2893-15.2016.

2. Cho, T.S., Beigaitė, E., Klein, N.E., Sweeney, S.T., and Bhattacharya, M.R.C. (2022). The Putative Drosophila TMEM184B Ortholog Tmep Ensures Proper Locomotion by Restraining Ectopic Firing at the Neuromuscular Junction. Mol. Neurobiol. 10.1007/S12035-022-02760-3.

3. Wright, E.B., Larsen, E.G., Coloma-Roessle, C.M., Hart, H.R., and Bhattacharya, M.R.C. (2023). Transmembrane protein 184B (TMEM184B) promotes expression of synaptic gene networks in the mouse hippocampus. BMC Genomics 24, 559. 10.1186/s12864-023-09676-9.

4. Larsen, E.G., Cho, T.S., McBride, M.L., Feng, J., Manivannan, B., Madura, C., Klein, N.E., Wright, E.B., Wickstead, E.S., Garcia-Verdugo, H.D., et al. (2022). Transmembrane protein TMEM184B is necessary for interleukin-31-induced itch. Pain 163, E642–E653. 10.1097/J.PAIN.0000000000002452.

5. Oliva, C.A., Montecinos-Oliva, C., and Inestrosa, N.C. (2018). Chapter Three - Wnt Signaling in the Central Nervous System: New Insights in Health and Disease. In Progress in Molecular Biology and Translational Science WNT Signaling in Health and Disease., J. Larraín and G. Olivares, eds. (Academic Press), pp. 81–130. 10.1016/bs.pmbts.2017.11.018.

6. Yao, Z., van Velthoven, C.T.J., Kunst, M., Zhang, M., McMillen, D., Lee, C., Jung, W., Goldy, J., Abdelhak, A., Aitken, M., et al. (2023). A high-resolution transcriptomic and spatial atlas of cell types in the whole mouse brain. Nature 624, 317–332. 10.1038/s41586-023-06812-z.

7. Lin, Y., Liu, D., Li, X., Ma, Y., and Pan, X. (2022). TMEM184B promotes proliferation, migration and invasion, and inhibits apoptosis in hypopharyngeal squamous cell carcinoma. J. Cell. Mol. Med. 26, 5551–5561. 10.1111/jcmm.17572.

8. Fukumoto, I., Hanazawa, T., Kinoshita, T., Kikkawa, N., Koshizuka, K., Goto, Y., Nishikawa, R., Chiyomaru, T., Enokida, H., Nakagawa, M., et al. (2015). MicroRNA expression signature of oral squamous cell carcinoma: functional role of microRNA-26a/b in the modulation of novel cancer pathways. Br. J. Cancer 112, 891–900. 10.1038/bjc.2015.19.

9. Lindström, S., Thompson, D.J., Paterson, A.D., Li, J., Gierach, G.L., Scott, C., Stone, J., Douglas, J.A., dos-Santos-Silva, I., Fernandez-Navarro, P., et al. (2014). Genome-wide association study identifies multiple loci associated with both mammographic density and breast cancer risk. Nat. Commun. 5, 5303. 10.1038/ncomms6303.

10. Baxter, S.M., Posey, J.E., Lake, N.J., Sobreira, N., Chong, J.X., Buyske, S., Blue, E.E., Chadwick, L.H., Coban-Akdemir, Z.H., Doheny, K.F., et al. (2022). Centers for Mendelian Genomics: A decade of facilitating gene discovery. Genet. Med. 24, 784– 797. 10.1016/j.gim.2021.12.005.

11. Hüppi, P.S., Warfield, S., Kikinis, R., Barnes, P.D., Zientara, G.P., Jolesz, F.A., Tsuji, M.K., and Volpe, J.J. (1998). Quantitative magnetic resonance imaging of brain development in premature and mature newborns. Ann. Neurol. 43, 224–235. 10.1002/ana.410430213.

12. Takahashi, E., Folkerth, R.D., Galaburda, A.M., and Grant, P.E. (2012). Emerging Cerebral Connectivity in the Human Fetal Brain: An MR Tractography Study. Cereb. Cortex 22, 455–464. 10.1093/cercor/bhr126.

13. Dawidziuk, M., Gambin, T., Bukowska-Olech, E., Antczak-Marach, D., Badura-Stronka, M., Buda, P., Budzynska, E., Castaneda, J., Chilarska, T., Czyzyk, E., et al. (2021). Exome Sequencing Reveals Novel Variants and Expands the Genetic Landscape for Congenital Microcephaly. Genes 12, 2014. 10.3390/genes12122014.

14. Hsu, C.L., Lee, E.X., Gordon, K.L., Paz, E.A., Shen, W.-C., Ohnishi, K., Meisenhelder, J., Hunter, T., and La Spada, A.R. (2018). MAP4K3 mediates amino acid-dependent regulation of autophagy via phosphorylation of TFEB. Nat. Commun. 9, 942. 10.1038/s41467-018-03340-7.

15. Martina, J.A., Chen, Y., Gucek, M., and Puertollano, R. (2012). MTORC1 functions as a transcriptional regulator of autophagy by preventing nuclear transport of TFEB. Autophagy 8, 903–914. 10.4161/auto.19653.

16. Sardiello, M., Palmieri, M., di Ronza, A., Medina, D.L., Valenza, M., Gennarino, V.A., Di Malta, C., Donaudy, F., Embrione, V., Polishchuk, R.S., et al. (2009). A gene network regulating lysosomal biogenesis and function. Science 325, 473–477. 10.1126/science.1174447.

17. Napolitano, G., and Ballabio, A. (2016). TFEB at a glance. J. Cell Sci. 129, 2475– 2481. 10.1242/jcs.146365.

18. Cunningham, K.M., Maulding, K., Ruan, K., Senturk, M., Grima, J.C., Sung, H., Zuo, Z., Song, H., Gao, J., Dubey, S., et al. (2020). TFEB/Mitf links impaired nuclear import to autophagolysosomal dysfunction in C9-ALS. eLife 9, e59419. 10.7554/eLife.59419.

19. Sobreira, N., Schiettecatte, F., Valle, D., and Hamosh, A. (2015). GeneMatcher: a matching tool for connecting investigators with an interest in the same gene. Hum. Mutat. 36, 928–930. 10.1002/humu.22844.

20. Cheng, J., Novati, G., Pan, J., Bycroft, C., Žemgulytė, A., Applebaum, T., Pritzel, A., Wong, L.H., Zielinski, M., Sargeant, T., et al. (2023). Accurate proteome-wide missense variant effect prediction with AlphaMissense. Science 381, eadg7492. 10.1126/science.adg7492.

21. Schubach, M., Maass, T., Nazaretyan, L., Röner, S., and Kircher, M. (2024). CADD v1.7: using protein language models, regulatory CNNs and other nucleotide-level scores to improve genome-wide variant predictions. Nucleic Acids Res. 52, D1143– D1154. 10.1093/nar/gkad989.

22. Ruff, K.M., and Pappu, R.V. (2021). AlphaFold and Implications for Intrinsically Disordered Proteins. J. Mol. Biol. 433, 167208. 10.1016/j.jmb.2021.167208.

23. Chen, M., Zhang, W., Gou, Y., Xu, D., Wei, Y., Liu, D., Han, C., Huang, X., Li, C., Ning, W., et al. (2023). GPS 6.0: an updated server for prediction of kinase-specific phosphorylation sites in proteins. Nucleic Acids Res. 51, W243–W250. 10.1093/nar/gkad383.

24. Niederriter, A.R., Davis, E.E., Golzio, C., Oh, E.C., Tsai, I.-C., and Katsanis, N. (2013). In Vivo Modeling of the Morbid Human Genome using Danio rerio. J. Vis. Exp. JoVE. 10.3791/50338.

25. Isrie, M., Breuss, M., Tian, G., Hansen, A.H., Cristofoli, F., Morandell, J., Kupchinsky, Z.A., Sifrim, A., Rodriguez-Rodriguez, C.M., Dapena, E.P., et al. (2015). Mutations in Either TUBB or MAPRE2 Cause Circumferential Skin Creases Kunze Type. Am. J. Hum. Genet. 97, 790. 10.1016/j.ajhg.2015.10.014.

26. Jaganathan, K., Kyriazopoulou Panagiotopoulou, S., McRae, J.F., Darbandi, S.F., Knowles, D., Li, Y.I., Kosmicki, J.A., Arbelaez, J., Cui, W., Schwartz, G.B., et al. (2019). Predicting Splicing from Primary Sequence with Deep Learning. Cell 176, 535–548.e24. 10.1016/j.cell.2018.12.015.

27. White, R.J., Collins, J.E., Sealy, I.M., Wali, N., Dooley, C.M., Digby, Z., Stemple, D.L., Murphy, D.N., Billis, K., Hourlier, T., et al. (2017). A high-resolution mRNA expression time course of embryonic development in zebrafish. eLife 6, e30860. 10.7554/eLife.30860.

28. Lee, Y.-R., Khan, K., Armfield-Uhas, K., Srikanth, S., Thompson, N.A., Pardo, M., Yu, L., Norris, J.W., Peng, Y., Gripp, K.W., et al. (2020). Mutations in FAM50A suggest that Armfield XLID syndrome is a spliceosomopathy. Nat. Commun. 11. 10.1038/s41467-020-17452-6.

29. Sur, A., Wang, Y., Capar, P., Margolin, G., Prochaska, M.K., and Farrell, J.A. (2023). Single-cell analysis of shared signatures and transcriptional diversity during zebrafish development. Dev. Cell 58, 3028–3047.e12. 10.1016/j.devcel.2023.11.001.

30. Sarparanta, J., Jonson, P.H., Golzio, C., Sandell, S., Luque, H., Screen, M., McDonald, K., Stajich, J.M., Mahjneh, I., Vihola, A., et al. (2012). Mutations affecting the cytoplasmic functions of the co-chaperone DNAJB6 cause limb-girdle muscular dystrophy. Nat. Genet. 44, 450–455. 10.1038/ng.1103.

31. Guissart, C., Latypova, X., Rollier, P., Khan, T.N., Stamberger, H., McWalter, K., Cho, M.T., Kjaergaard, S., Weckhuysen, S., Lesca, G., et al. (2018). Dual Molecular Effects of Dominant RORA Mutations Cause Two Variants of Syndromic Intellectual Disability with Either Autism or Cerebellar Ataxia. Am. J. Hum. Genet. 102, 744– 759. 10.1016/j.ajhg.2018.02.021.

32. Gordon, C.T., Weaver, K.N., Zechi-Ceide, R.M., Madsen, E.C., Tavares, A.L.P., Oufadem, M., Kurihara, Y., Adameyko, I., Picard, A., Breton, S., et al. (2015). Mutations in the Endothelin Receptor Type A Cause Mandibulofacial Dysostosis with Alopecia. Am. J. Hum. Genet. 96, 519–531. 10.1016/j.ajhg.2015.01.015.

33. Tan, P.L., Garrett, M.E., Willer, J.R., Campochiaro, P.A., Campochiaro, B., Zack, D.J., Ashley-Koch, A.E., and Katsanis, N. (2017). Systematic Functional Testing of Rare Variants: Contributions of CFI to Age-Related Macular Degeneration. Invest. Ophthalmol. Vis. Sci. 58, 1570–1576. 10.1167/iovs.16-20867.

34. Grange, L.J., Reynolds, J.J., Ullah, F., Isidor, B., Shearer, R.F., Latypova, X., Baxley, R.M., Oliver, A.W., Ganesh, A., Cooke, S.L., et al. (2022). Pathogenic variants in SLF2 and SMC5 cause segmented chromosomes and mosaic variegated hyperploidy. Nat. Commun. 13, 6664. 10.1038/s41467-022-34349-8.

35. Ansar, M., Ullah, F., Paracha, S.A., Adams, D.J., Lai, A., Pais, L., Iwaszkiewicz, J., Millan, F., Sarwar, M.T., Agha, Z., et al. (2019). Bi-allelic Variants in DYNC1I2 Cause Syndromic Microcephaly with Intellectual Disability, Cerebral Malformations, and Dysmorphic Facial Features. Am. J. Hum. Genet. 104, 1073–1087. 10.1016/j.ajhg.2019.04.002.

36. Donowitz, M., Tse, C.M., and Fuster, D. (2013). SLC9/NHE gene family, a plasma membrane and organellar family of Na+/H+ exchangers. Mol. Aspects Med. 34, 236–251. 10.1016/j.mam.2012.05.001.

37. Parker, M.D., and Boron, W.F. (2013). The divergence, actions, roles, and relatives of sodium-coupled bicarbonate transporters. Physiol. Rev. 93, 803–959. 10.1152/physrev.00023.2012.

38. Pareja, F., Brandes, A.H., Basili, T., Selenica, P., Geyer, F.C., Fan, D., Da Cruz Paula, A., Kumar, R., Brown, D.N., Gularte-Mérida, R., et al. (2018). Loss-of-function mutations in ATP6AP1 and ATP6AP2 in granular cell tumors. Nat. Commun. 9, 3533. 10.1038/s41467-018-05886-y.

39. Kanfer, G., Sarraf, S.A., Maman, Y., Baldwin, H., Dominguez-Martin, E., Johnson, K.R., Ward, M.E., Kampmann, M., Lippincott-Schwartz, J., and Youle, R.J. (2021). Image-based pooled whole-genome CRISPRi screening for subcellular phenotypes. J. Cell Biol. 220. 10.1083/JCB.202006180.

40 . Liscum, L., Ruggiero, R.M., and Faust, J.R. (1989). The intracellular transport of low density lipoprotein-derived cholesterol is defective in Niemann-Pick type C fibroblasts. J. Cell Biol. 108, 1625–1636. 10.1083/jcb.108.5.1625.

41 . Lloyd-Evans, E., Morgan, A.J., He, X., Smith, D.A., Elliot-Smith, E., Sillence, D.J., Churchill, G.C., Schuchman, E.H., Galione, A., and Platt, F.M. (2008). Niemann-Pick disease type C1 is a sphingosine storage disease that causes deregulation of lysosomal calcium. Nat. Med. 14, 1247–1255. 10.1038/NM.1876.

42. Maue, R.A., Burgess, R.W., Wang, B., Wooley, C.M., Seburn, K.L., Vanier, M.T., Rogers, M.A., Chang, C.C., Chang, T.-Y., Harris, B.T., et al. (2012). A novel mouse model of Niemann-Pick type C disease carrying a D1005G-Npc1 mutation comparable to commonly observed human mutations. Hum. Mol. Genet. 21, 730– 750. 10.1093/hmg/ddr505.

43. Kim, S., Ochoa, K., Melli, S.E., Yousufzai, F.A.K., Barrera, Z.D., Williams, A.A., McIntyre, G., Delgado, E., Bolish, J.N., Macleod, C.M., et al. (2023). Disruptive lysosomal-metabolic signaling and neurodevelopmental deficits that precede Purkinje cell loss in a mouse model of Niemann-Pick Type-C disease. Sci. Rep. 13, 5665. 10.1038/s41598-023-32971-0.

44. Wan, H.I., DiAntonio, A., Fetter, R.D., Bergstrom, K., Strauss, R., and Goodman, C.S. (2000). Highwire regulates synaptic growth in Drosophila. Neuron 26, 313–329. 10.1016/s0896-6273(00)81166-6.

45. Collins, C.A., Wairkar, Y.P., Johnson, S.L., and DiAntonio, A. (2006). Highwire Restrains Synaptic Growth by Attenuating a MAP Kinase Signal. Neuron 51, 57–69. 10.1016/j.neuron.2006.05.026.

46. AlAbdi, L., Desbois, M., Rusnac, D.-V., Sulaiman, R.A., Rosenfeld, J.A., Lalani, S., Murdock, D.R., Burrage, L.C., Undiagnosed Diseases Network, Billie Au, P.Y., et al. (2023). Loss-of-function variants in MYCBP2 cause neurobehavioural phenotypes and corpus callosum defects. Brain J. Neurol. 146, 1373–1387. 10.1093/brain/awac364.

47. Xiong, X., Wang, X., Ewanek, R., Bhat, P., DiAntonio, A., and Collins, C.A. (2010). Protein turnover of the Wallenda/DLK kinase regulates a retrograde response to axonal injury. J. Cell Biol. 191, 211–223. 10.1083/jcb.201006039.

48. Babetto, E., Beirowski, B., Russler, E.V., Milbrandt, J., and DiAntonio, A. (2013). The Phr1 ubiquitin ligase promotes injury-induced axon self-destruction. Cell Rep. 3, 1422–1429. 10.1016/j.celrep.2013.04.013.

49. Wright, E.B., Larsen, E.G., Padilla-Rodriguez, M., Langlais, P.R., and Bhattacharya, M.R.C. (2025).TMEM184B modulates endolysosomal acidification via the vesicular proton pump. J Cell Sci 2025) 138, jcs263908. doi:10.1242/jcs.263908.

50. Gilmore, E.C., and Walsh, C.A. (2013). Genetic causes of microcephaly and lessons for neuronal development. Wiley Interdiscip. Rev. Dev. Biol. 2, 461–478. 10.1002/wdev.89.

51. Jayaraman, D., Bae, B.-I., and Walsh, C.A. (2018). The Genetics of Primary Microcephaly. Annu. Rev. Genomics Hum. Genet. 19, 177–200. 10.1146/annurev-genom-083117-021441.

52. Dawson, P.A., Hubbert, M., Haywood, J., Craddock, A.L., Zerangue, N., Christian, W.V., and Ballatori, N. (2005). The heteromeric organic solute transporter alpha-beta, Ostalpha-Ostbeta, is an ileal basolateral bile acid transporter. J. Biol. Chem. 280, 6960–6968. 10.1074/jbc.M412752200.

53. Malinovsky, F.G., Brodersen, P., Fiil, B.K., McKinney, L.V., Thorgrimsen, S., Beck, M., Nielsen, H.B., Pietra, S., Zipfel, C., Robatzek, S., et al. (2010). Lazarus1, a DUF300 Protein, Contributes to Programmed Cell Death Associated with Arabidopsis acd11 and the Hypersensitive Response. PLoS ONE 5, e12586. 10.1371/journal.pone.0012586.

54. Yee, D.C., Shlykov, M.A., Västermark, A., Reddy, V.S., Arora, S., Sun, E.I., Saier, M.H., and Jr. (2013). The transporter-opsin-G protein-coupled receptor (TOG) superfamily. FEBS J. 280, 5780–5800. 10.1111/febs.12499.

55. Mauthe, M., Orhon, I., Rocchi, C., Zhou, X., Luhr, M., Hijlkema, K.J., Coppes, R.P., Engedal, N., Mari, M., and Reggiori, F. (2018). Chloroquine inhibits autophagic flux by decreasing autophagosome-lysosome fusion. Autophagy 14, 1435–1455. 10.1080/15548627.2018.1474314.

56. Palmieri, M., Pal, R., Nelvagal, H.R., Lotfi, P., Stinnett, G.R., Seymour, M.L., Chaudhury, A., Bajaj, L., Bondar, V.V., Bremner, L., et al. (2017). mTORC1-independent TFEB activation via Akt inhibition promotes cellular clearance in neurodegenerative storage diseases. Nat. Commun. 8, 14338. 10.1038/ncomms14338.

