## Supplemental Figures and Tables for "Pathogenic variants in *TMEM184B* cause a neurodevelopmental syndrome associated with alteration of metabolic signaling"

### Note S1. Survey for Data Collection from Affected Individuals

We collected information about affected individuals with known variants through an email survey to their providers. The following questions were included in the survey. Variant information was provided separately.

1) How was genetics analysis performed? (i.e., Whole exome, panel on whole exome platform, whole genome, single gene sequencing)?
2) How old was the affected individual when diagnosed (with genetics)?
3) What is the affected individual’s gender?
4) Does the affected individual have intellectual disability or developmental delay? If developmental delay: gross, fine or speech?
5) What growth changes are observed? (Delayed? Normal? Microcephaly? Macrocephaly?)
6) Does the affected individual have seizures? If so, do you know the type?
7) Does the affected individual have congenital heart abnormalities? If yes, what type?
8) Has the affected individual had an MRI/A/S? If so, do you observe any of the following: Corpus collosum hypoplasia? Delayed myelination for age? Any malformations of brain?

### Supplemental Figures

### Figure S1

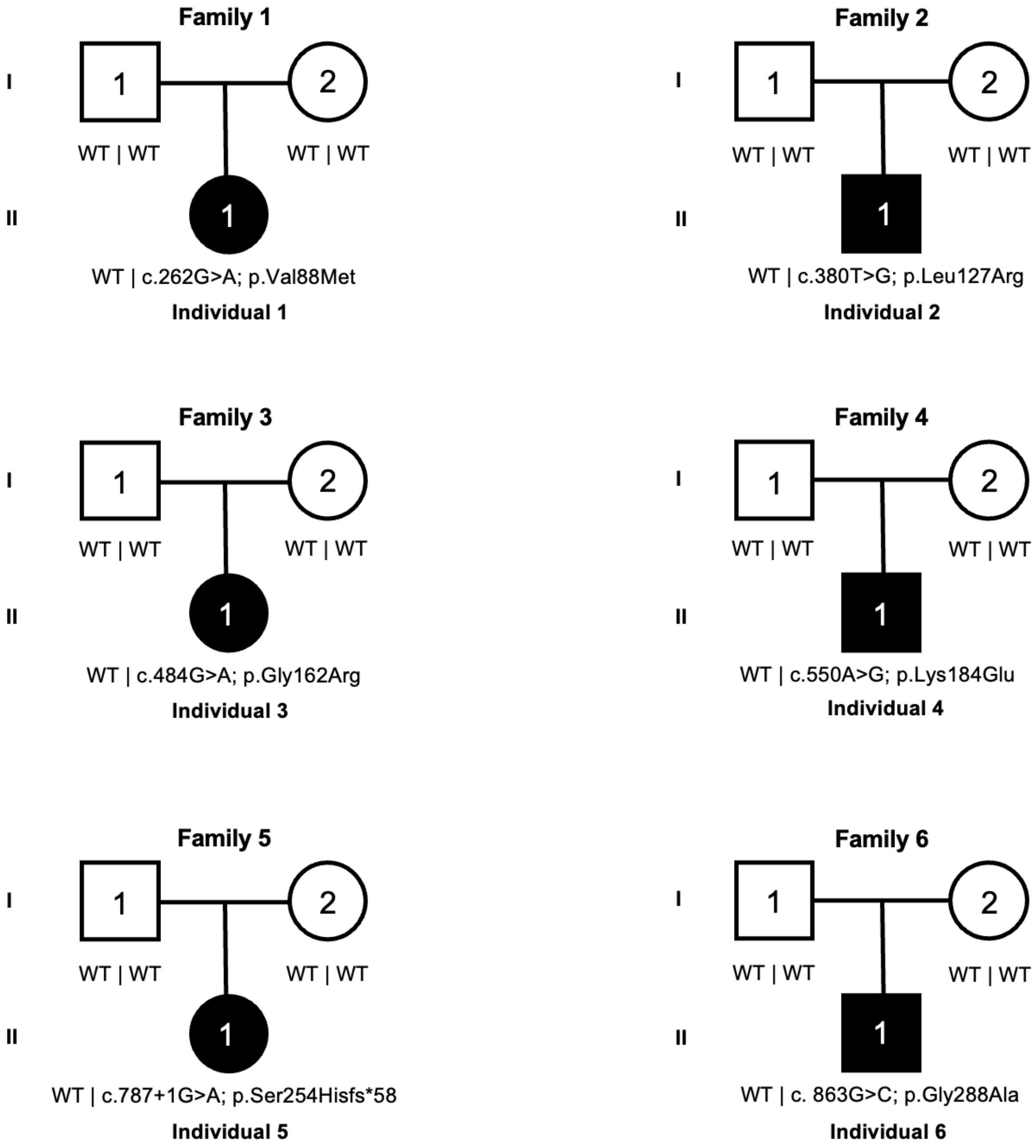


**Figure S1. Pedigrees of affected individuals with *de novo TMEM184B* variants.** Parent-proband trios reported in this study are shown with *TMEM184B* genotypes. Individual numbers correspond to identifiers shown in Table 1 and Table S2. Square, male; circle, female; filled shape, affected; unfilled shape, unaffected; WT, wild type.

**Figure S2**

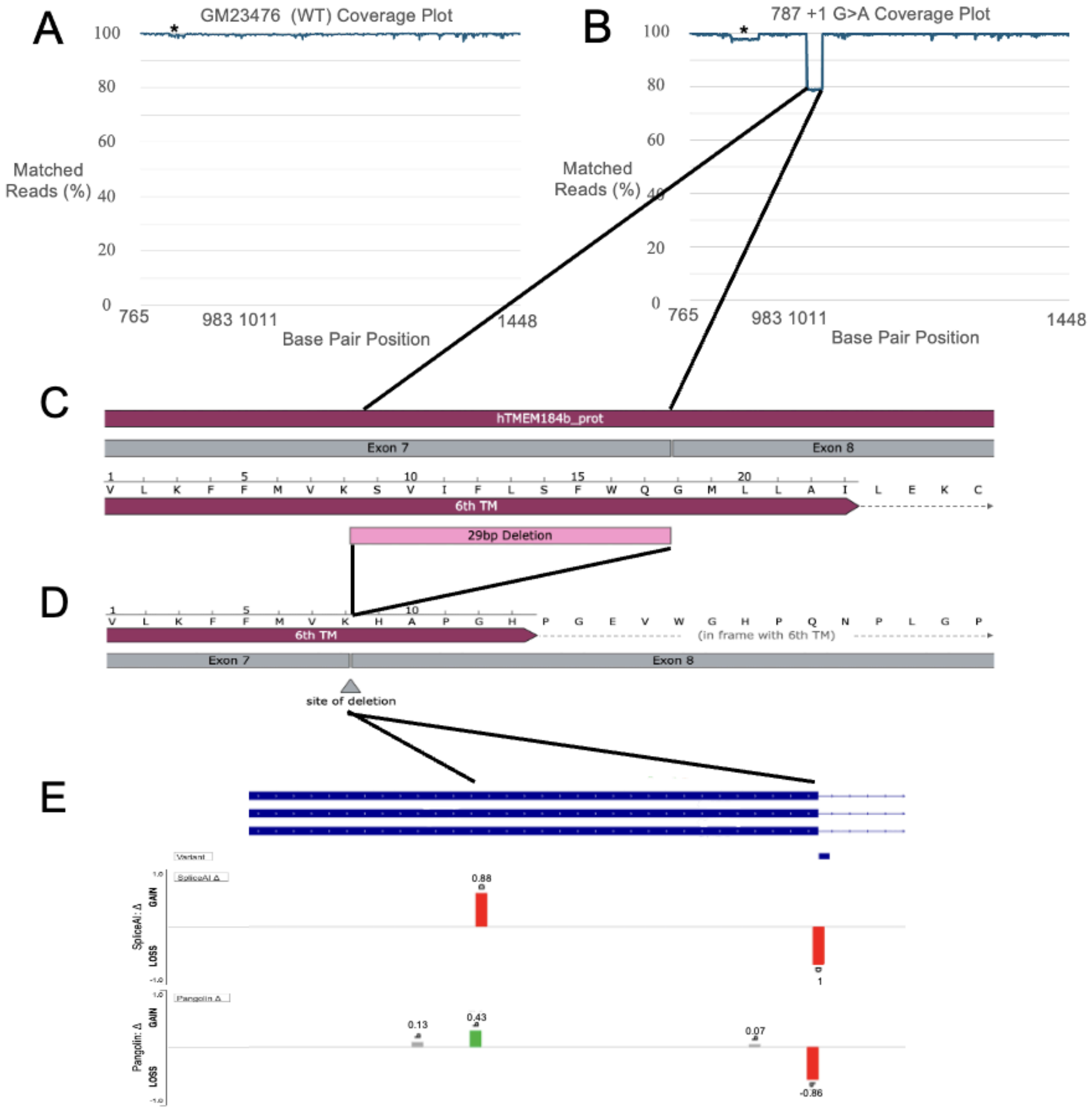


**Figure S2. Individual 5 harbors a splice site variant that causes a predominant 29 base deletion and frameshift in exon 7. A-B** Coverage plots for the control iPSC line (GM23476) (**A**) and the c.787+1 G>A iPSC line (**B**). The number of matched reads at each base pair is divided by the total number of reads at that base pair position. (*) represents a ~2% occurrence seen at a common acceptor site that lasts 51 base pairs. **C** Location of the 29 base pair region shown in B, mapped onto the TMEM184B amino acid sequence. **D** Expected result of the 29 base pair region on the TMEM184B amino acid sequence. Exon 7 is truncated, and Exon 8 is changed completely. 58 amino acids after the deletion, there is a stop codon truncating TMEM184B and removing 88 amino acids from the original sequence (p.Ser254Hisfs*58). **E** Using Splice AI and Pangolin, we found that the 29 base pair deletion region matches the predicted result if an identified cryptic splice site is used.

**Figure S3**

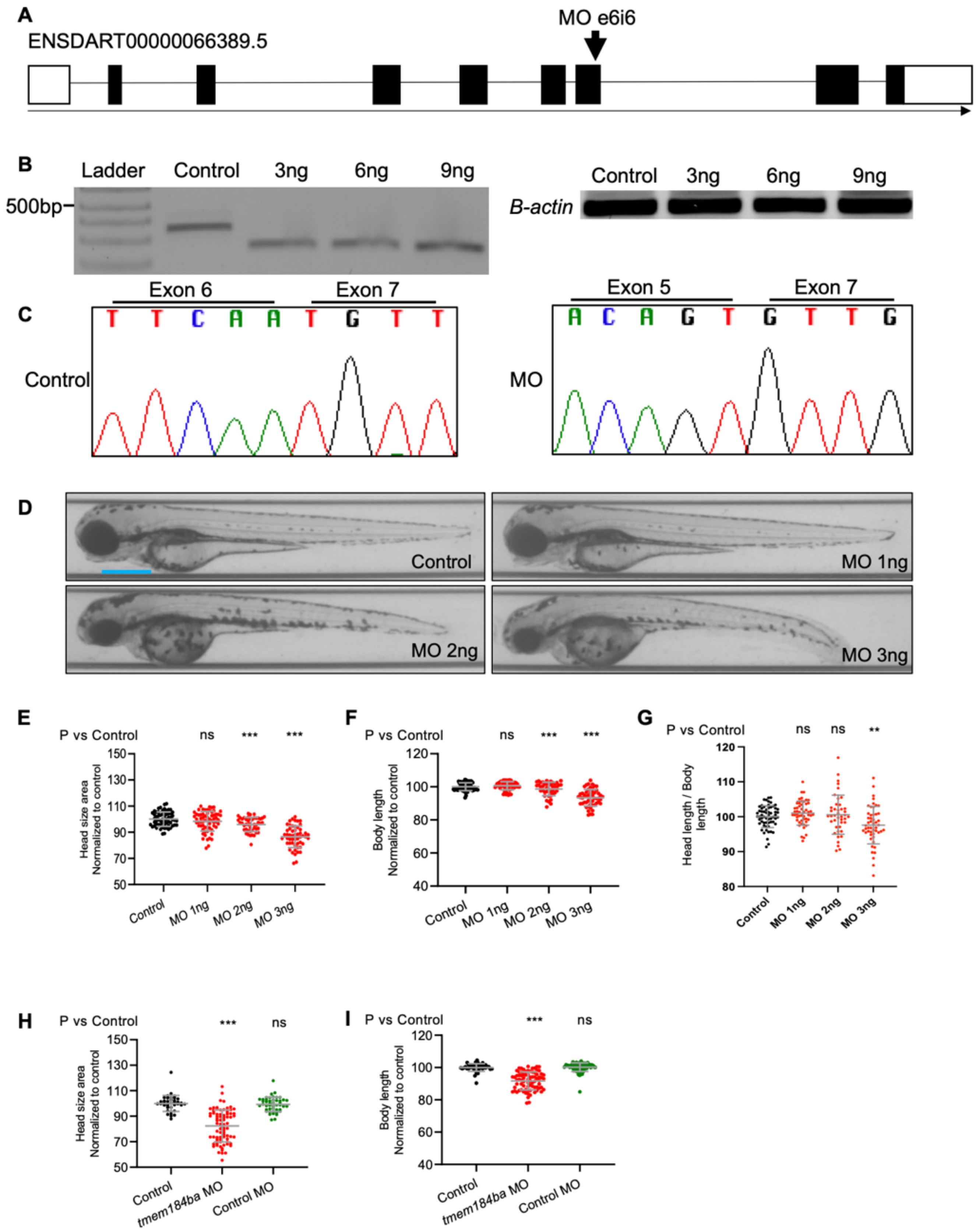
A
ENSDART00000066389.5
MO e6i6
B
Ladder
Control
3ng
6ng
9ng
500bp
Control
3ng
6ng
9ng
B-actin
C
Exon 6
Exon 7
Control
Exon 5
Exon 7
MO
D
Control
MO 1ng
MO 2ng
MO 3ng
E
P vs Control
ns
***
***
Head size area Normalized to control
Control
MO 1ng
MO 2ng
MO 3ng
F
P vs Control
ns
***
***
Body length Normalized to control
Control
MO 1ng
MO 2ng
MO 3ng
G
P vs Control
ns
ns
**
Head length / Body length
Control
MO 1ng
MO 2ng
MO 3ng
H
P vs Control
***
ns
Head size area Normalized to control
Control
tmem184ba MO
Control MO
I
P vs Control
***
ns
Body length Normalized to control
Control
tmem184ba MO
Control MO

**Figure S3. A morpholino targeting exon 6-intron 6 (e6i6) to transiently suppress *tmem184ba* is efficient.**

**A** Schematic of the zebrafish *tmem184ba* at chr12:19,320,657-19,340,220 (GRCz11). White boxes, untranslated regions; black boxes, coding exons; horizontal black line, introns; vertical arrow, morpholino (MO) target site at the exon 6 splice donor site. **B** Inverted agarose gel image showing RT-PCR products amplified with primers flanking the MO target site in controls and *tmem184ba* morphants. The expected WT band size is 375 bp and morphants have undetectable WT with exon 6 exclusion (92 bp) when injected with 3 ng, 6 ng, or 9 ng MO. **C** Sanger sequence chromatograms confirm exon 6 skipping in morphants injected with 3 ng MO. **D** Representative lateral bright field images of 3 dpf larvae injected with increasing doses of *tmem184ba* MO. Scale bar, 300 μm. **E and F** Quantification of (**E**) the lateral head size and (**F**) body length in 3 dpf larvae as indicated with anatomical landmarks in Figure 3A (**G**) Quantification of the lateral head length to body length ratio, showing specificity to anterior structures. Experimental conditions were normalized and presented as a percentage of the control mean. **H and I** Quantification of (**H**) the lateral head size and (**I**) body length in 3 dpf larvae as indicated with anatomical landmarks in Figure 3A; 3 ng *tmem184ba* MO was compared to 3 ng standard control MO. For panels E, F, G, H and I statistical analyses were performed with a one-way ANOVA with Tukey's post-hoc test; ***$p<0.001$; and ns, not significant. n = 40–67/condition. Error bars represent standard deviation of the mean. Data point color: black, uninjected control; red, *tmem184ba* MO-injected; and green, standard control MO-injected.

**Figure S4**

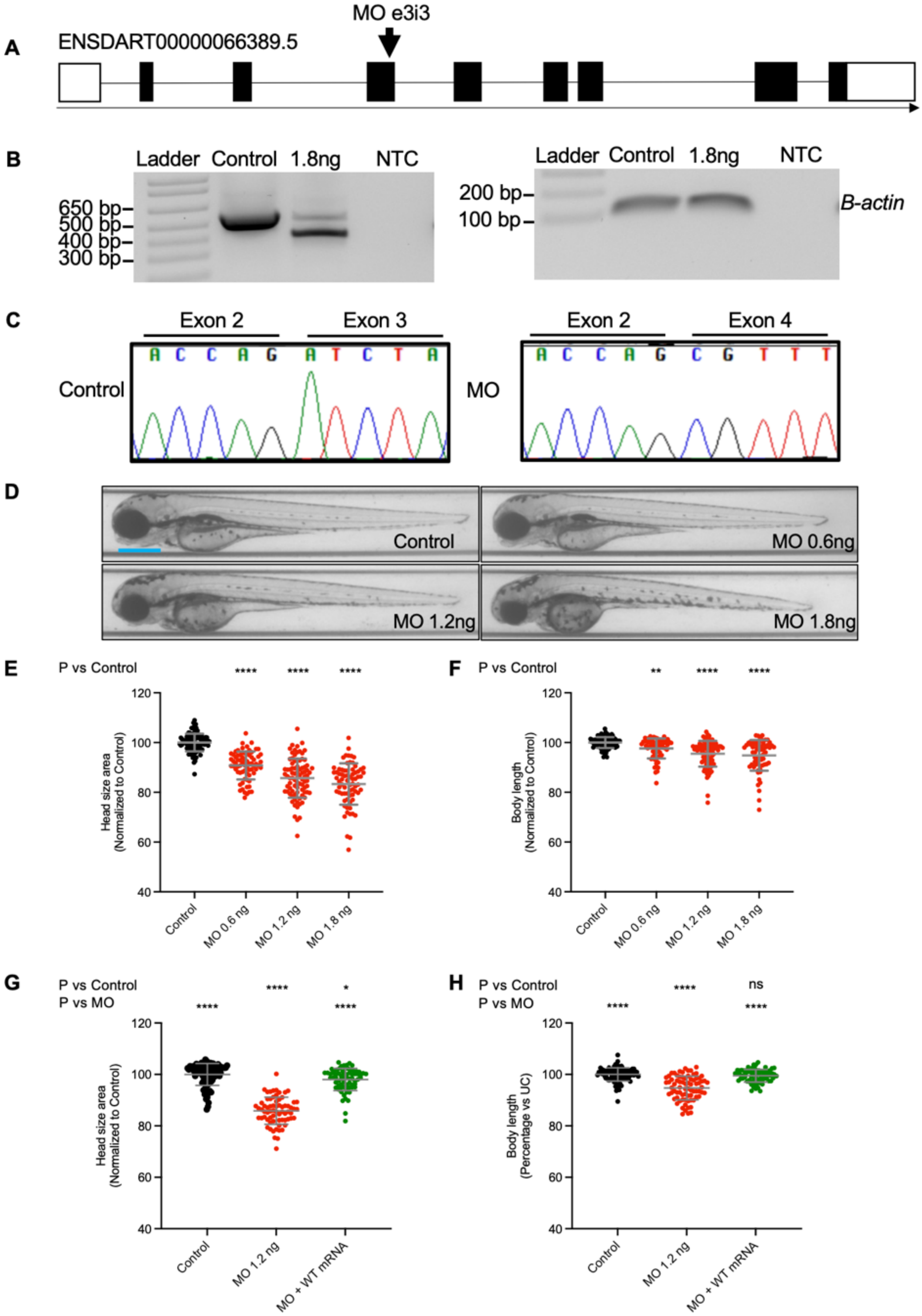
A
ENSDART00000066389.5
MO e3i3
B
Ladder
Control
1.8ng
NTC
650 bp
500 bp
400 bp
300 bp
200 bp
100 bp
B-actin
C
Exon 2
Exon 3
Exon 4
Control
MO
A C C A G A T C T A
A C C A G C G T T T
D
Control
MO 0.6ng
MO 1.2ng
MO 1.8ng
E
P vs Control
****
Head size area (Normalized to Control)
Control
MO 0.6 ng
MO 1.2 ng
MO 1.8 ng
F
P vs Control
**
Body length (Normalized to Control)
G
P vs Control
P vs MO
*
Head size area (Normalized to Control)
MO + WT mRNA
H
ns
Body length (Percentage vs UC)
120
100
80
60
40

**Figure S4. A morpholino targeting exon 3-intron 3 (e3i3) to transiently suppress *tmem184ba* is efficient.**

**A** Schematic representation of the zebrafish *tmem184ba* at chr12:19,320,657-19,340,220 (GRCz11). White boxes, untranslated regions; black vertical boxes, coding exons; horizontal black line, intron; vertical arrow, splice blocking morpholino (MO) at the exon 3 splice donor site. **B** Inverted agarose gel image showing RT-PCR products amplified with primers flanking the MO target site in controls and *tmem184ba* morphants. The expected WT band size is 564 bp and morphants have an aberrantly spliced transcript, resulting in exon 3 exclusion (166 bp) in morphants injected with 1.8 ng MO. The *b-actin* expected fragment, expected size of 120 bp, was amplified to ensure RNA integrity. NTC, non-template control **C** Sanger sequence chromatograms confirm exon 3 skipping in morphants injected with 1.8 ng MO. **D** Representative lateral bright field images of 3 dpf larvae injected with increasing doses of *tmem184ba* e3i3 MO. Scale bar, 300 μm. **E and F;** Quantification of (**E**) the lateral head size and (**F**) body length in 3 dpf larvae as indicated with anatomical landmarks in Figure 3A. **G and H;** Quantification of (**G**) the lateral head size and (**H**) body length in the 3 dpf larvae show significant rescue with 200 pg of WT human *TMEM184B* mRNA. For panels E, F, G, and H statistical analyses were performed with a one-way ANOVA with Tukey's post-hoc test; *p=0.0234; **p=0.0022; ****p<0.0001; and ns, not significant. n = 71-105/condition. Error bars represent standard deviation of the mean. Data point color: black, uninjected control; red, *tmem184ba* e3i3 MO-injected; and green, rescue with WT human *TMEM184B* mRNA.

**Figure S5**

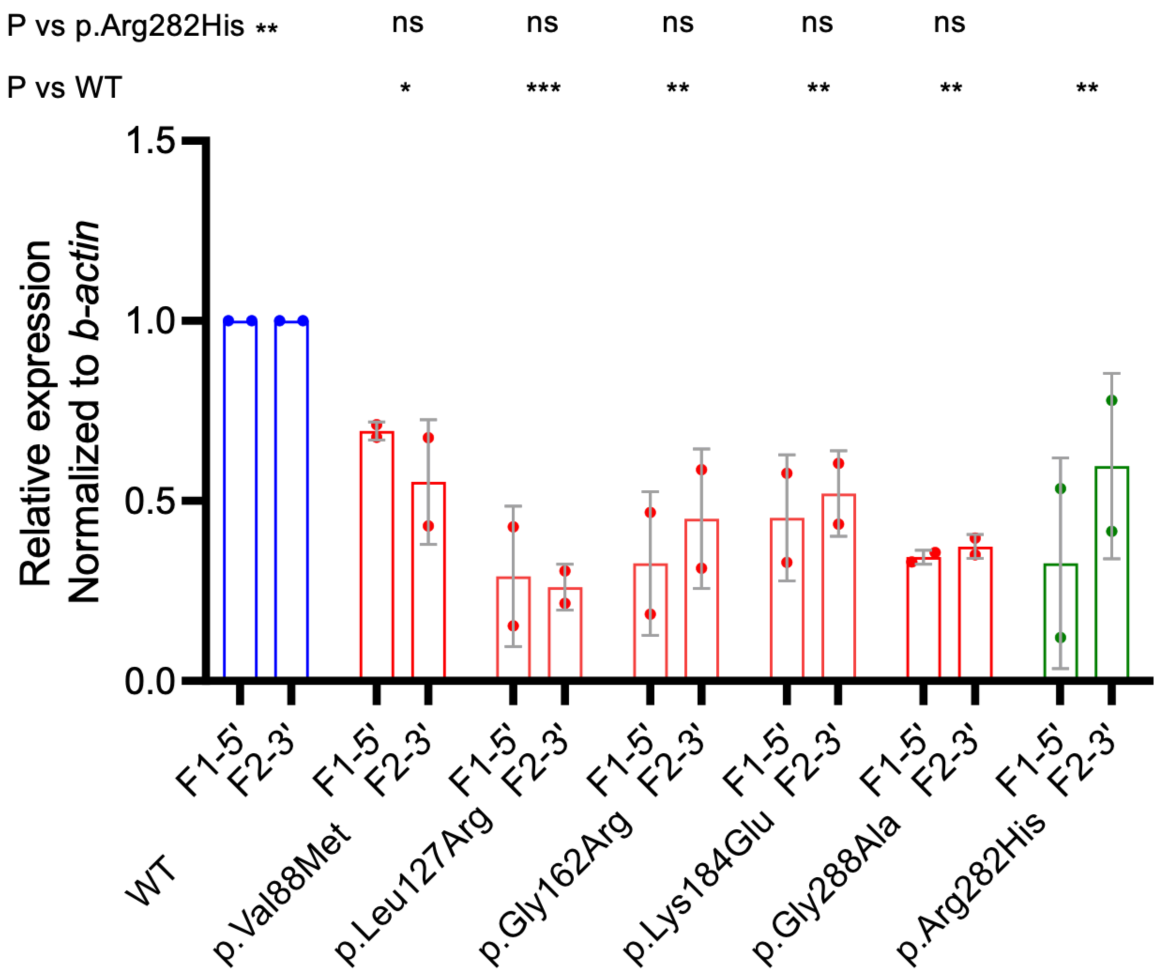


**Figure S5. Expression of human *TMEM184B* variant mRNA in zebrafish embryos 24 hours post-fertilization (hpf).** 200 pg *in vitro* transcribed *TMEM184B* mRNA was injected into embryos at the one to four cell stage and harvested for RNA extraction at 24 hpf. Biological replicate qRT-PCR, each with technical triplicates, was performed with primer pairs amplifying two different regions of *TMEM184B.* F1-5' indicates primers targeting a region spanning exons 2 and 3 and F2-3' indicates primers targeting a region spanning exons 6 and 7. Error bars represent standard deviation of the mean. Statistical analyses were performed with a one-way ANOVA with Tukey's post-hoc test; ***p<0.001; **p<0.01; *p<0.05 and ns, not significant. Data point color: blue, WT human mRNA; red, disease-associated variant mRNA; green, benign control variant mRNA.

**Figure S6**

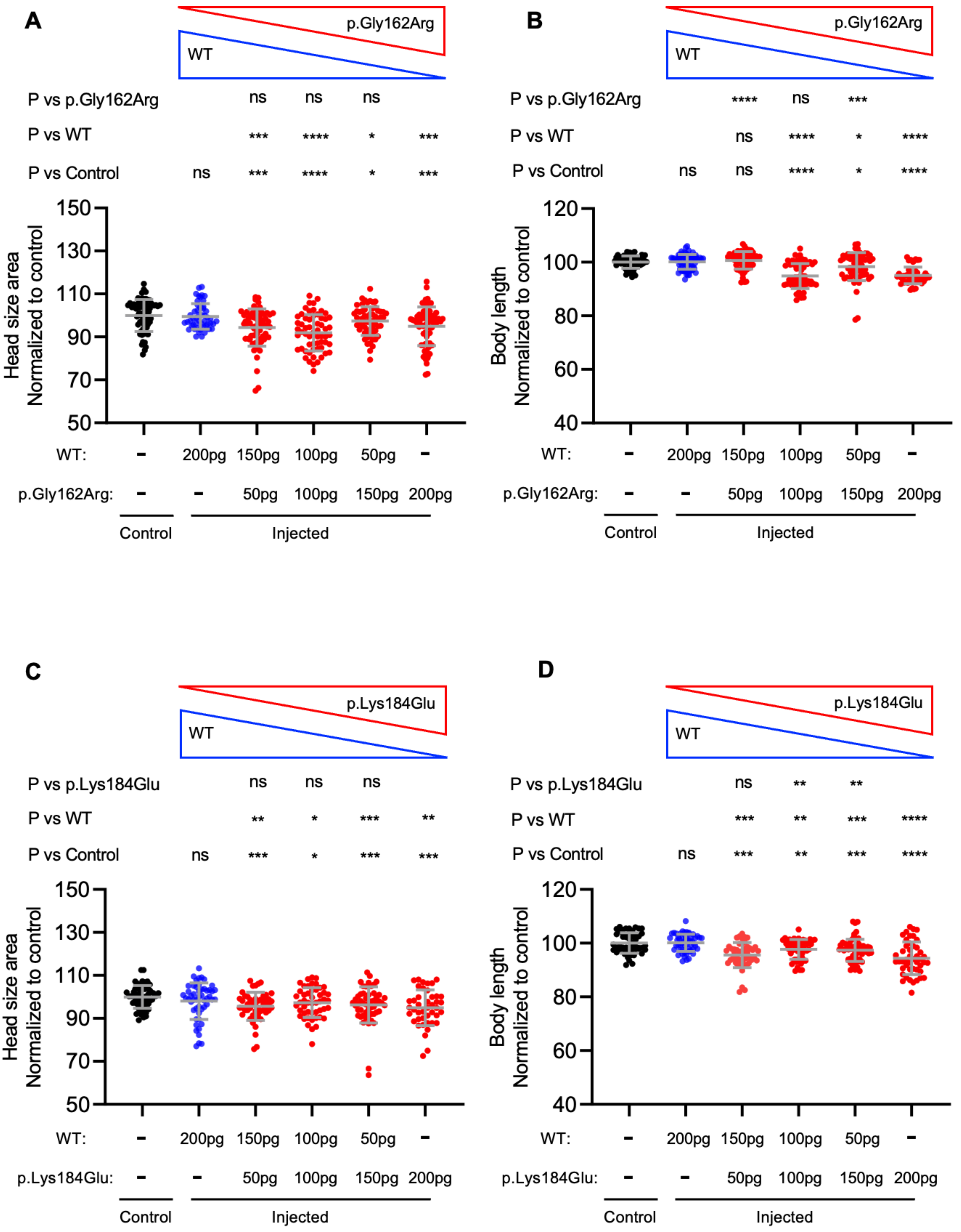
A
p.Gly162Arg
WT
P vs p.Gly162Arg ns ns ns
P vs WT *** **** * ***
P vs Control ns *** **** * ***
Head size area
Normalized to control
150
130
110
90
70
50
WT: - 200pg 150pg 100pg 50pg -
p.Gly162Arg: - - 50pg 100pg 150pg 200pg
Control
Injected
B
p.Gly162Arg
WT
P vs p.Gly162Arg **** ns ***
P vs WT ns **** * ****
P vs Control ns ns **** * ****
Body length
Normalized to control
120
100
80
60
40
WT: - 200pg 150pg 100pg 50pg -
p.Gly162Arg: - - 50pg 100pg 150pg 200pg
Control
Injected
C
p.Lys184Glu
WT
P vs p.Lys184Glu ns ns ns
P vs WT ** * *** **
P vs Control ns *** * *** ***
Head size area
Normalized to control
150
130
110
90
70
50
WT: - 200pg 150pg 100pg 50pg -
p.Lys184Glu: - - 50pg 100pg 150pg 200pg
Control
Injected
D
p.Lys184Glu
WT
P vs p.Lys184Glu ns ** **
P vs WT *** ** *** ****
P vs Control ns *** ** *** ****
Body length
Normalized to control
120
100
80
60
40
WT: - 200pg 150pg 100pg 50pg -
p.Lys184Glu: - - 50pg 100pg 150pg 200pg
Control
Injected

**Figure S6. Titration of p.Gly162Arg or p.Lys184Glu mRNAs with WT *TMEM184B* mRNA to dissect dominant negative or dominant toxic effects.**

**A and B** Quantification of the lateral head size in 3 dpf larvae injected with titrated doses of WT with variant mRNA. Experimental conditions were normalized and presented as a percentage of the control mean. See Figure 3A for anatomical landmarks measured. **C and D** Quantification of the body length in 3 dpf larvae injected with titrated doses of WT with variant mRNA. Experimental conditions were normalized and presented as a percentage of the control mean. See Figure 3A for anatomical landmarks measured. Statistical analyses were performed with a one-way ANOVA with Tukey's post-hoc test; ****$p<0.0001$; ***$p<0.001$; **$p<0.01$; *$p<0.05$; and ns, not significant; n = 49–72/condition. Error bars represent standard deviation of the mean. Data point color: black, uninjected control; blue, WT mRNA; red, variants identified in affected individuals. Large triangles indicate decreasing or increasing amounts of WT (blue) mRNA or variant (red) mRNA.

**Figure S7**

**A**

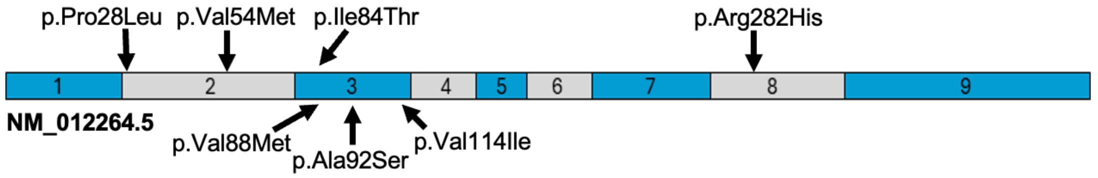


| Variant | Trans-membrane domain | ΔG | ΔΔG | gnomAD alleles v3.1.2 (non-neurological) | gnomAD alleles v4.1.0 |
|---|---|---|---|---|---|
| Wild Type | NA | 477.51 | NA | 134,884 | 1,614,324 |
| p.Pro28Leu | TM1 | 474.75 | -2.76 | 46/134,778 | 688/1,612,142 |
| p.Val54Met | TM1 | 487.33 | 9.82 | 3/134,730 | 35/1,613,790 |
| p.Ile84Thr | TM2 | 466.56 | -10.95 | Not present | 23/1,613,502 |
| p.Val88Met | TM2 | 465.14 | -12.37 | 3/134,780 | 35/1,613,538 |
| p.Ala92Ser | TM2 | 495.71 | 18.2 | 1/134,734 | 4/1,613,542 |
| p.Val114Ile | No domain | 469.21 | -8.3 | 3/134,562 | 37/1,611,328 |
| p.Arg282His | No domain | 481.56 | 4.05 | 13/134,782 | 215/1,609,748 |

**B**

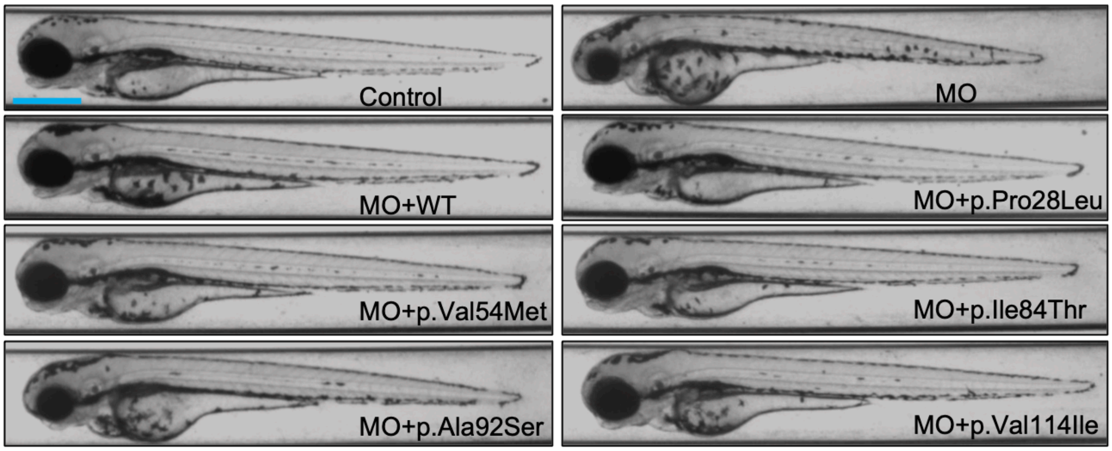


**C**

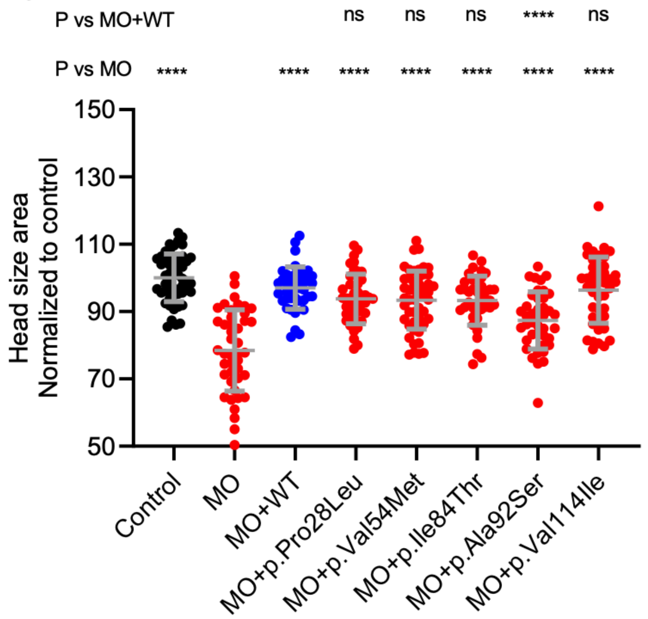


**D**

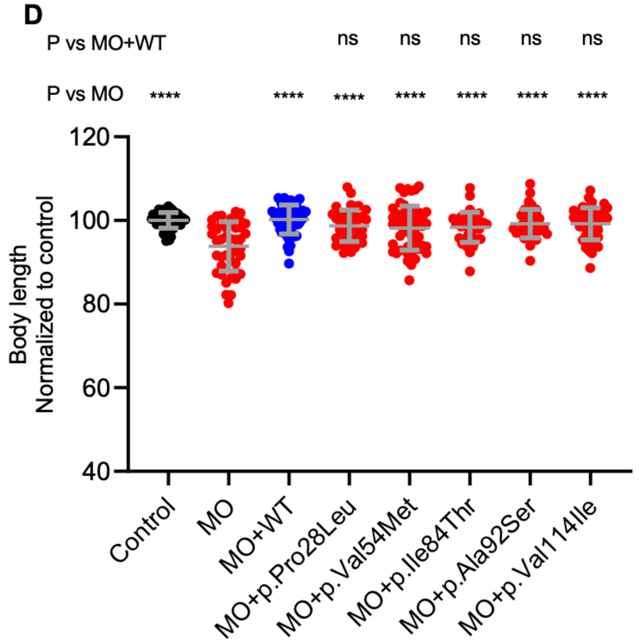

**Figure S7.** ***In vivo*** **complementation testing of variants from gnomAD to demonstrate assay specificity.**

**A** Top: Schematic of the human *TMEM184B* transcript; exons are indicated with alternating blue and gray. Localization of variants tested are shown with black arrows. Bottom: Variant table with gnomAD frequency and protein stability calculations using Gibbs free energy. ΔΔG values that are positive indicate decreased protein stability of variant-containing TMEM184B when compared to the predicted native structure, while negative values indicate abnormally heightened protein structural stability. NA, not applicable; TM, transmembrane domain. **B** Representative lateral bright field images of 3 dpf larvae injected with *tmem184ba* MO alone or coinjected with 200 pg human *TMEM184B* mRNA (WT, p.Pro28Leu, p.Val54Met, p.Ile84Thr, p.Ala92Ser, and p.Val114Ile). Scale bar, 300 μm. **C and D** Quantification of the (**C**) lateral head size and (**D**) body length in 3 dpf larvae as indicated with anatomical landmarks in Figure 3A. Experimental conditions were normalized and presented as a percentage of the control mean. Statistical analyses were performed with a one-way ANOVA with Tukey's post-hoc test; ****$p<0.0001$; and ns, not significant; n = 38–81/condition. Error bars represent standard deviation of the mean. Data point color: black, uninjected control; blue, WT mRNA; red, heterozygous variant mRNA present in gnomAD population.

**Figure S8**

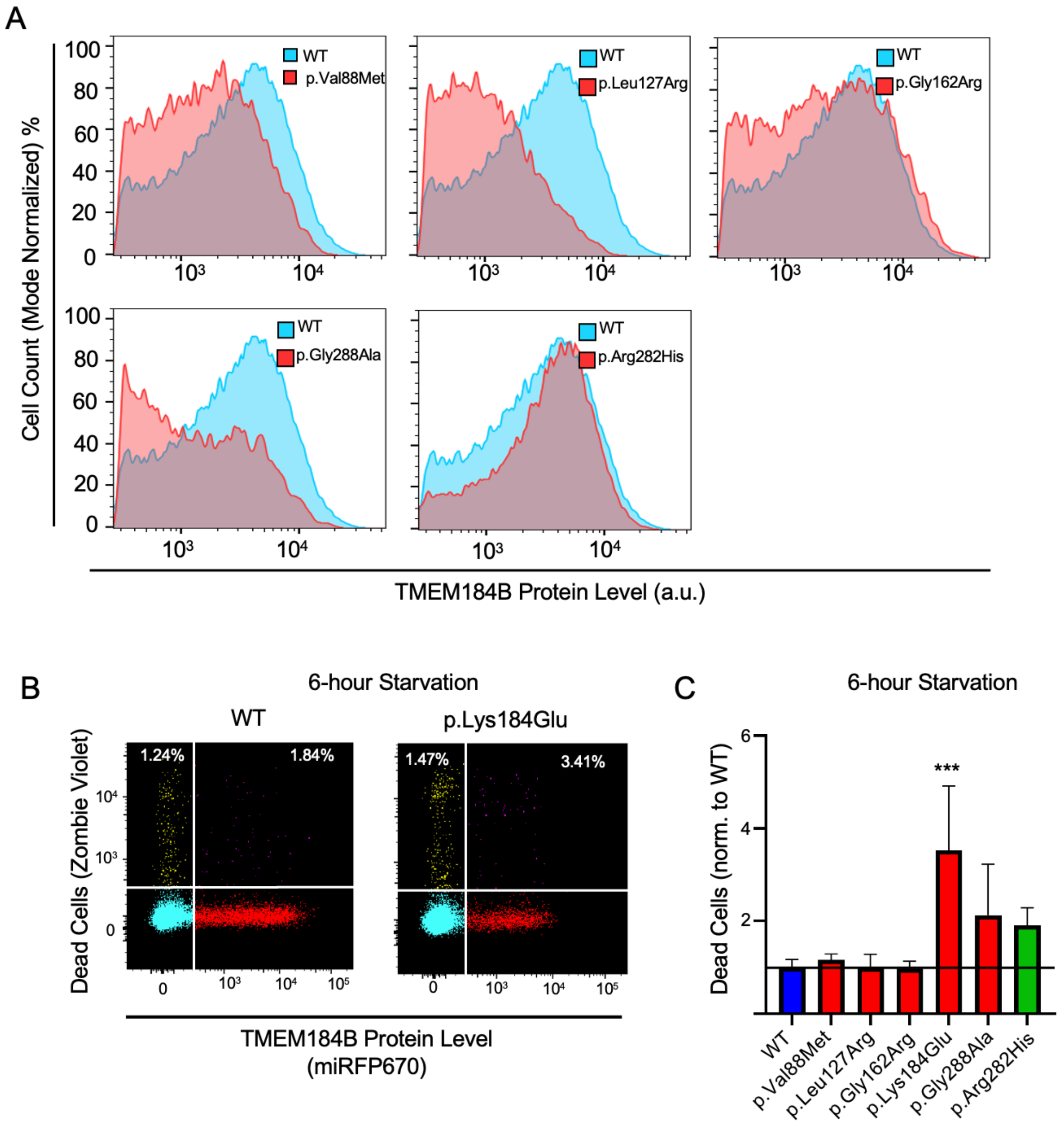


**Figure S8. TMEM184B variants exhibit altered protein levels and differential apoptosis susceptibility during starvation.**

**A** Fluorescence levels of HEK293T cells expressing *TMEM184B* variants tagged with miRFP670 were grouped into 256 bins and normalized to the cell count of each population. All graphs show cells under baseline (unstarved) conditions. **B** Zombie Violet apoptosis assay of HEK293T cells over-expressing WT *TMEM184B* (left) or p.Lys184Glu variant (right) that were nutrient starved for six hours. **C** Percentage of dead *TMEM184B* variant expressing cells that were starved for six hours, normalized to WT (N=3 trials). Error bars represent SD. ***p<0.001.

**Figure S9**

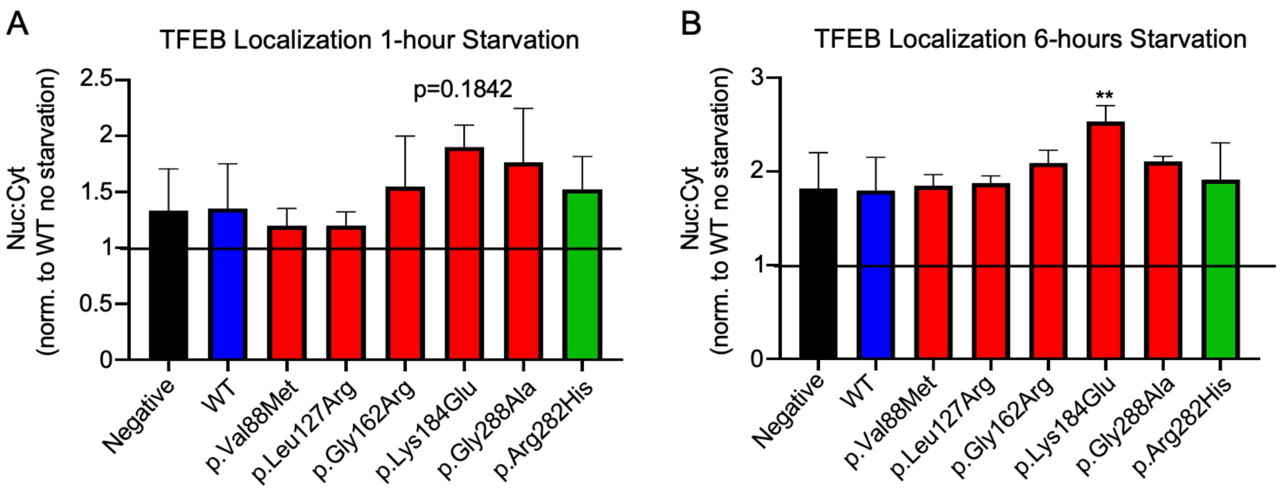


**Figure S9. TMEM184B p.Lys184Glu variant protein perturbs TFEB nuclear:cytoplasmic shuttling during starvation.**

**A** Ratio of mean TFEB-GFP fluorescence in the nucleus vs. cytoplasm in *TMEM184B-miRFP670* expressing cells that were nutrient starved for one hour. **B** TFEB distribution in cells starved for six hours. Both A and B are normalized to non-starved WT cells. **p<0.01.

### Supplemental Tables

### Table S1. Oligonucleotides used in this study.

| | | |
|---|---|---|
| tmem184ba sgRNA2 CRISPR/Cas9 | tmem184ba_G2_ex6_F | TAATACGACTCACTATAGACTGTAATTATGGCCATCAG |
| tmem184ba sgRNA2 CRISPR/Cas9 | tmem184ba_G2_ex6_R | TTCTAGCTCTAAAACCTGATGGCCATAATTACAGT |
| tmem184ba sgRNA2 CRISPR/Cas9 efficiency | tmem184ba_G2_gDNA_F-PCR | AAGTTTGTGGTCCAGGGTCA |
| tmem184ba sgRNA2 CRISPR/Cas9 efficiency | tmem184ba_G2_gDNA_R-PCR | GGCAACACTGTGGGATCAAT |
| Transient knockdown | Control MO | CCTCTTACCTCAGTTACAATTTATA |
| Transient knockdown | tmem184ba MO_e6i6 | ATGCACTTTTGGAGAAGCACATACT |
| Transient knockdown | tmem184ba MO_e3i3 | AGAAGGAGAACCTACCCTCATAGCA |
| MO efficiency | tmem184ba_e3i3_F | TTTGGGGAGGAAATTGTGAG |
| MO efficiency | tmem184ba_e3i3_R | TATGGCCATCAGGGGTTTTA |
| MO efficiency | tmem184ba_e6i6_F | CAACTTCTTGAGCCTTTGTTATGA |
| MO efficiency | tmem184ba_e6i6_R | GCAGGTCTCGAGTGGAGAAG |
| Site directed mutagenesis | TMEM184B_G162R_F | GCACCTGCTGCCTCTGGAGAAAGACTTATTCCATC |
| Site directed mutagenesis | TMEM184B_G162R_R | GATGGAATAAGTCTTTCTCCAGAGGCAGCAGGTGC |
| Site directed mutagenesis | TMEM184B_G288A_F | GTGTCGGTGGGCGAGGCCACCGTGGCTGCCGGC |
| Site directed mutagenesis | TMEM184B_G288A_R | GCCGGCAGCCACGGTGGCCTCGCCCACCGACAC |
| Site directed mutagenesis | TMEM184B_K184E_F | AGTTCTGTGTGGTGGAGCCACTCATGGCGGTC |
| Site directed mutagenesis | TMEM184B_K184E_R | GACCGCCATGAGTGGCTCCACCACACAGAACT |
| Site directed mutagenesis | TMEM184B_L127R_F | GTCATCTATAATTTCCGGAGCCTGTGCTATGAG |
| Site directed mutagenesis | TMEM184B_L127R_R | CTCATAGCACAGGCTCCGGAAATTATAGATGAC |
| Site directed mutagenesis | TMEM184B_V88M_F | CGCATCCTCTTCATCATGCCCATCTACGCCTTT |
| Site directed mutagenesis | TMEM184B_V88M_F | AAAGGCGTAGATGGGCATGATGAAGAGGATGCG |
| Site directed mutagenesis | TMEM184B_P28L_F | AGCGTCTCCGTGATCCTCGAGGGCAGCCCCACT |
| Site directed mutagenesis | TMEM184B_P28L_R | AGTGGGGCTGCCCTCGAGGATCACGGAGACGCT |
| Site directed mutagenesis | TMEM184B_I84T_F | CGCTACATCGTGCGCACCCTCTTCATCGTGCCC |
| Site directed mutagenesis | TMEM184B_I84T_R | GGGCACGATGAAGAGGGTGCGCACGATGTAGCG |
| Site directed mutagenesis | TMEM184B_A92S_F | ATCGTGCCCATCTATCCTTTGACTCCTGGCTC |
| Site directed mutagenesis | TMEM184B_A92S_R | GAGCCAGGAGTCAAAGGATAGATGGGCACGAT |
| Site directed mutagenesis | TMEM184B_V114I_F | GTGTACTTCGGCACCATCCGCGACTGCTATGAGG |
| Site directed mutagenesis | TMEM184B_V114I_R | CCTCATAGCAGTCGCGGATGGTGCCGAAGTACAC |
| Site directed mutagenesis | TMEM184B_V54M_F | ATCTCTGGCTTCTTCATGTGGACGGCCCTGCTCAT |
| qRT-PCR | TMEM184BqPCR_ex2_F | ATGGAGCAGCCTGTGTTCCT |
| qRT-PCR | TMEM184BPCR_ex3_F | AGGATGCGCACGATGTAGC |
| qRT-PCR | TMEM184BqPCR_ex7_F | CCCGTCCTCAAGTTCTTCAT |
| qRT-PCR | TMEM184BqPCR_ex8_F | TCCAGCCTCTTGTCAGCATA |
| Sequencing | TMEM184B_ORF_F | GGCACCATGACAGTGAGGGG |
| Sequencing | TMEM184B_ORF_R | CTAGAATTCATCATCAGAGCTGAGC |
| Sequencing | TMEM184B_SEQ1 | CCTCCTCTTCTTCACCAACG |
| Sequencing | TMEM184B_SEQ2 | AGAGGTAGCCACTGGTGACG |
| Sequencing | TMEM184B_SEQ3 | GGCCATCCTGGAGAAGTGT |
| Sequencing | TMEM184B_SEQ4 | GCTGGTAGGCAGGTGAGAAG |

| | | |
|---|---|---|
| Amplicon sequencing of TMEM184B | Forward_ c.787+1G>A | TGTGTGGTGAAGCCACTCAT |
| Amplicon sequencing of TMEM184B | Reverse_ c.787+1G>A | ATAAGCCTTGCTCCCAGTGT |
| gRNA to introduce c.550A>G into iPSC | TMEM184B gRNA | CTGTGTGGTGAAGCCACTCANGG |

**Table S2. Detailed clinical information table (redacted).**

**Table S3. Transcriptional changes between *TMEM184B*$^{\text{c.550A>G/+}}$ and its isogenic control.** Shown are genes that reached statistical significance (adjusted p< 0.05, DeSeq2).

**Table S4. Transcriptional changes between *TMEM184B*$^{\text{c.550A>G/c.550A>G}}$ and its isogenic control** Shown are genes that reached statistical significance (adjusted p< 0.05, DeSeq2; same control as heterozygous line.

**Supplementary Table S3. K184E heterozygous vs WT**

**UP - p<.05, FC>1.5**

| GeneID | Base mean | log2(FC) | StdErr | Wald-Stats | P-value | P-adj | FC |
|---|---|---|---|---|---|---|---|
| SMS | 8241.6326 | 0.960108 | 0.04586 | 20.9358591 | 2.52E-97 | 8.70E-94 | 1.945456 |
| TOP2A | 9719.4374 | 0.94168 | 0.070636 | 13.3313719 | 1.52E-40 | 2.62E-37 | 1.920764 |
| DLGAP5 | 3981.0515 | 0.833837 | 0.063081 | 13.2185055 | 6.86E-40 | 1.08E-36 | 1.78242 |
| PLS3 | 17219.441 | 0.629672 | 0.050007 | 12.5917593 | 2.34E-36 | 3.11E-33 | 1.547213 |
| KIF20A | 1898.2373 | 0.832877 | 0.069101 | 12.0529746 | 1.87E-33 | 1.70E-30 | 1.781234 |
| BUB1 | 5740.5982 | 0.894568 | 0.075218 | 11.8930101 | 1.29E-32 | 1.06E-29 | 1.859053 |
| USP9X | 41410.907 | 0.721133 | 0.062711 | 11.4992408 | 1.33E-30 | 9.97E-28 | 1.648476 |
| STMN1 | 9687.0842 | 0.592027 | 0.052836 | 11.2050788 | 3.85E-29 | 2.21E-26 | 1.507363 |
| FAM83D | 1329.853 | 0.737348 | 0.068121 | 10.8241118 | 2.65E-27 | 1.34E-24 | 1.667108 |
| CCNB1 | 3878.6213 | 0.805959 | 0.075164 | 10.7226527 | 7.97E-27 | 3.92E-24 | 1.748308 |
| MAP2 | 870.16482 | 1.402984 | 0.135279 | 10.3710222 | 3.36E-25 | 1.52E-22 | 2.644479 |
| PHF6 | 4280.9653 | 0.646467 | 0.062432 | 10.3547379 | 3.98E-25 | 1.76E-22 | 1.56533 |
| ZNF680 | 127.86577 | 2.623642 | 0.258863 | 10.1352439 | 3.85E-24 | 1.54E-21 | 6.16304 |
| KPNA2 | 10844.188 | 0.862913 | 0.08622 | 10.0082438 | 1.40E-23 | 5.49E-21 | 1.818706 |
| RBM12 | 3587.9182 | 0.666614 | 0.066731 | 9.9896267 | 1.69E-23 | 6.48E-21 | 1.587343 |
| KIF11 | 2450.3794 | 0.684424 | 0.070057 | 9.76951095 | 1.52E-22 | 5.14E-20 | 1.607061 |
| MSH6 | 6563.3219 | 0.599214 | 0.061611 | 9.72578885 | 2.34E-22 | 7.76E-20 | 1.514891 |
| IPO7 | 7686.2741 | 0.618831 | 0.064755 | 9.55651518 | 1.22E-21 | 3.89E-19 | 1.53563 |
| STAT1 | 1818.0868 | 0.620534 | 0.067014 | 9.25981065 | 2.05E-20 | 6.09E-18 | 1.537444 |
| G2E3 | 1819.2743 | 0.895066 | 0.097426 | 9.18717531 | 4.03E-20 | 1.16E-17 | 1.859694 |
| PLCL2 | 1075.5977 | 0.832817 | 0.09166 | 9.08597792 | 1.03E-19 | 2.76E-17 | 1.781159 |
| STARD4 | 910.126 | 0.847587 | 0.09336 | 9.07865193 | 1.10E-19 | 2.87E-17 | 1.799489 |
| LTA4H | 4558.1823 | 0.608085 | 0.067281 | 9.03802924 | 1.60E-19 | 3.93E-17 | 1.524235 |
| AL035078. | 106.39536 | 10.05817 | 1.119984 | 8.98063556 | 2.69E-19 | 6.44E-17 | 1066.131 |
| MLLT11 | 944.6367 | 0.667821 | 0.075061 | 8.89705436 | 5.73E-19 | 1.34E-16 | 1.588672 |
| ECT2 | 2809.0344 | 0.621779 | 0.071128 | 8.74164289 | 2.30E-18 | 4.89E-16 | 1.538771 |
| ST13 | 8553.0291 | 0.592898 | 0.067822 | 8.74195791 | 2.29E-18 | 4.89E-16 | 1.508273 |
| NT5DC1 | 1643.5311 | 0.59564 | 0.069498 | 8.57056072 | 1.03E-17 | 2.02E-15 | 1.511143 |
| PCDHGC4 | 305.65517 | 1.096906 | 0.130232 | 8.42271512 | 3.68E-17 | 6.74E-15 | 2.138954 |
| PATL1 | 1074.5725 | 0.609188 | 0.07259 | 8.39221171 | 4.77E-17 | 8.66E-15 | 1.5254 |
| AIFM1 | 2321.1246 | 0.618974 | 0.073978 | 8.36695048 | 5.91E-17 | 1.06E-14 | 1.535782 |
| KYAT3 | 978.92346 | 0.615006 | 0.07562 | 8.13281326 | 4.19E-16 | 6.63E-14 | 1.531564 |
| ANKRD1 | 39.68167 | 3.018813 | 0.372911 | 8.09527141 | 5.71E-16 | 8.71E-14 | 8.105007 |
| GTF2IP4 | 3455.0556 | 0.662049 | 0.081991 | 8.07468882 | 6.76E-16 | 1.01E-13 | 1.582329 |
| DEPDC1B | 1545.1206 | 0.599227 | 0.074309 | 8.06401685 | 7.38E-16 | 1.10E-13 | 1.514905 |
| FOXG1 | 34.459482 | 4.284863 | 0.532934 | 8.04013416 | 8.97E-16 | 1.31E-13 | 19.49271 |
| STON1 | 717.61416 | 0.748702 | 0.093464 | 8.01054739 | 1.14E-15 | 1.60E-13 | 1.68028 |
| VPS41 | 1220.008 | 0.705445 | 0.088112 | 8.00618177 | 1.18E-15 | 1.64E-13 | 1.630647 |
| ARHGAP11 | 2598.2411 | 0.928741 | 0.116374 | 7.98062697 | 1.46E-15 | 1.98E-13 | 1.903614 |
| CKAP2L | 724.88125 | 0.982171 | 0.12314 | 7.97605181 | 1.51E-15 | 2.02E-13 | 1.975436 |
| CCDC117 | 1465.4301 | 0.631539 | 0.079314 | 7.96251035 | 1.69E-15 | 2.24E-13 | 1.549216 |

| | | | | | | | |
|---|---|---|---|---|---|---|---|
| DIP2B | 1204.1075 | 0.605387 | 0.076234 | 7.94117471 | 2.00E-15 | 2.63E-13 | 1.521387 |
| KIFC1 | 1816.4549 | 0.640454 | 0.080917 | 7.91493511 | 2.47E-15 | 3.18E-13 | 1.558819 |
| FILIP1 | 530.29419 | 0.781018 | 0.099199 | 7.8732787 | 3.45E-15 | 4.22E-13 | 1.718343 |
| NCOA6 | 624.75591 | 0.780042 | 0.099093 | 7.87181668 | 3.50E-15 | 4.24E-13 | 1.717181 |
| BNIP3L | 2079.6304 | 0.692518 | 0.088055 | 7.8646163 | 3.70E-15 | 4.46E-13 | 1.616102 |
| EEF1AKNM | 1360.5771 | 0.643546 | 0.081908 | 7.85691071 | 3.94E-15 | 4.71E-13 | 1.562164 |
| UBE2T | 792.93821 | 0.717969 | 0.091971 | 7.80645315 | 5.88E-15 | 6.85E-13 | 1.644865 |
| NASP | 15421.352 | 0.687221 | 0.088152 | 7.7958781 | 6.40E-15 | 7.35E-13 | 1.610179 |
| TPX2 | 4252.5099 | 0.59881 | 0.077065 | 7.7702347 | 7.83E-15 | 8.88E-13 | 1.514467 |
| CCNG1 | 9303.5359 | 0.671635 | 0.087722 | 7.65638784 | 1.91E-14 | 2.11E-12 | 1.592878 |
| LANCL1 | 2206.5436 | 0.666332 | 0.087112 | 7.64918127 | 2.02E-14 | 2.22E-12 | 1.587033 |
| MIB1 | 3094.5143 | 0.597085 | 0.078317 | 7.62398934 | 2.46E-14 | 2.63E-12 | 1.512657 |
| CASTOR3 | 285.21905 | 1.169689 | 0.153706 | 7.60991462 | 2.74E-14 | 2.90E-12 | 2.249632 |
| WDR36 | 1340.8222 | 0.659671 | 0.086875 | 7.5932884 | 3.12E-14 | 3.28E-12 | 1.579722 |
| GSK3B | 1859.0496 | 0.617567 | 0.081464 | 7.58084161 | 3.43E-14 | 3.59E-12 | 1.534285 |
| GATM | 434.21196 | 0.674097 | 0.091231 | 7.38887315 | 1.48E-13 | 1.42E-11 | 1.595598 |
| PTTG1 | 1941.1404 | 0.651451 | 0.088217 | 7.38468753 | 1.53E-13 | 1.44E-11 | 1.570748 |
| SF3A3 | 3292.7116 | 0.676559 | 0.091603 | 7.38575436 | 1.52E-13 | 1.44E-11 | 1.598323 |
| RSC1A1 | 36.227914 | 8.497854 | 1.15993 | 7.32617703 | 2.37E-13 | 2.17E-11 | 361.5004 |
| ARNT | 1025.7415 | 0.592401 | 0.081112 | 7.30348753 | 2.80E-13 | 2.49E-11 | 1.507754 |
| COIL | 569.98481 | 0.629846 | 0.086327 | 7.2960351 | 2.96E-13 | 2.61E-11 | 1.5474 |
| CAMK2D | 641.90531 | 0.759094 | 0.104077 | 7.29357766 | 3.02E-13 | 2.63E-11 | 1.692428 |
| PDE12 | 531.42146 | 0.645289 | 0.088495 | 7.2918369 | 3.06E-13 | 2.65E-11 | 1.564052 |
| TTK | 1384.4838 | 0.733548 | 0.10065 | 7.28809066 | 3.14E-13 | 2.71E-11 | 1.662723 |
| SOX11 | 2930.6753 | 0.824985 | 0.113373 | 7.27674662 | 3.42E-13 | 2.93E-11 | 1.771517 |
| HMGB1P6 | 1794.7248 | 0.679929 | 0.093546 | 7.26838286 | 3.64E-13 | 3.10E-11 | 1.602061 |
| MOB1A | 2428.8619 | 0.624693 | 0.08684 | 7.19364111 | 6.31E-13 | 5.18E-11 | 1.541883 |
| TUBA1C | 11160.442 | 0.60517 | 0.085252 | 7.09862172 | 1.26E-12 | 9.65E-11 | 1.521158 |
| NDUFA6 | 31.783199 | 5.921666 | 0.838066 | 7.0658718 | 1.60E-12 | 1.20E-10 | 60.61766 |
| SLC39A7 | 235.71928 | 23.89999 | 3.383527 | 7.06363108 | 1.62E-12 | 1.22E-10 | 15653548 |
| CBX1 | 4191.6812 | 0.638542 | 0.091223 | 6.99978525 | 2.56E-12 | 1.83E-10 | 1.556755 |
| C4orf46 | 1255.7662 | 0.602885 | 0.086149 | 6.99818839 | 2.59E-12 | 1.85E-10 | 1.518751 |
| CDK1 | 1992.8808 | 0.785199 | 0.112496 | 6.97979015 | 2.96E-12 | 2.10E-10 | 1.72333 |
| PDK2 | 435.7159 | 0.695463 | 0.100164 | 6.94325147 | 3.83E-12 | 2.65E-10 | 1.619404 |
| NBEA | 1447.9917 | 0.609424 | 0.088005 | 6.92488685 | 4.36E-12 | 2.98E-10 | 1.52565 |
| ERCC6L | 759.28032 | 0.592012 | 0.085806 | 6.89943111 | 5.22E-12 | 3.53E-10 | 1.507347 |
| EYA3 | 726.07786 | 0.67092 | 0.09727 | 6.89753813 | 5.29E-12 | 3.55E-10 | 1.592088 |
| CKS2 | 1811.137 | 0.680272 | 0.09882 | 6.88394232 | 5.82E-12 | 3.89E-10 | 1.602442 |
| TRIM61 | 132.05857 | 1.48655 | 0.216875 | 6.8544078 | 7.16E-12 | 4.59E-10 | 2.802182 |
| PUM2 | 3455.6653 | 0.604429 | 0.088212 | 6.85196955 | 7.28E-12 | 4.65E-10 | 1.520377 |
| ATP6V1C1 | 1607.5871 | 0.747598 | 0.110054 | 6.79299685 | 1.10E-11 | 6.63E-10 | 1.678995 |
| KIF14 | 1098.2408 | 0.742903 | 0.109569 | 6.78021079 | 1.20E-11 | 7.16E-10 | 1.67354 |
| ARFIP1 | 958.38211 | 0.731848 | 0.108254 | 6.76045708 | 1.38E-11 | 8.04E-10 | 1.660765 |
| SOX1 | 33.522018 | 3.00159 | 0.449007 | 6.68494619 | 2.31E-11 | 1.26E-09 | 8.00882 |

| | | | | | | | |
|---|---|---|---|---|---|---|---|
| RNGTT | 873.47234 | 0.661501 | 0.0995 | 6.64826605 | 2.97E-11 | 1.57E-09 | 1.581727 |
| ARHGEF26 | 337.69679 | 0.702534 | 0.106691 | 6.58476395 | 4.56E-11 | 2.30E-09 | 1.62736 |
| CCNG2 | 450.44863 | 0.852955 | 0.129662 | 6.57828295 | 4.76E-11 | 2.36E-09 | 1.806196 |
| CHUK | 591.77152 | 0.611035 | 0.093233 | 6.55386551 | 5.61E-11 | 2.75E-09 | 1.527355 |
| TRIM24 | 6710.3383 | 0.587765 | 0.089733 | 6.55012085 | 5.75E-11 | 2.79E-09 | 1.502916 |
| ANKRD50 | 1382.2049 | 0.671857 | 0.102932 | 6.52719976 | 6.70E-11 | 3.23E-09 | 1.593123 |
| ARHGAP32 | 973.85816 | 0.641066 | 0.098337 | 6.51905336 | 7.08E-11 | 3.36E-09 | 1.559481 |
| NCAPG | 2219.6388 | 0.60585 | 0.092953 | 6.51779415 | 7.13E-11 | 3.38E-09 | 1.521875 |
| KIF20B | 1765.6057 | 0.634983 | 0.09846 | 6.44914579 | 1.12E-10 | 5.09E-09 | 1.55292 |
| BACH1 | 1092.166 | 0.591749 | 0.09235 | 6.40764617 | 1.48E-10 | 6.48E-09 | 1.507073 |
| STAU2 | 1500.9449 | 0.652575 | 0.102349 | 6.37601013 | 1.82E-10 | 7.73E-09 | 1.571972 |
| SP8 | 237.62466 | 0.861886 | 0.135208 | 6.37451367 | 1.84E-10 | 7.79E-09 | 1.817412 |
| TNRC6B | 1307.4118 | 0.844086 | 0.13325 | 6.33458386 | 2.38E-10 | 9.77E-09 | 1.795127 |
| RRM2B | 1443.4152 | 0.623368 | 0.098435 | 6.3327614 | 2.41E-10 | 9.86E-09 | 1.540467 |
| HELZ | 1580.0603 | 0.618358 | 0.097947 | 6.31317875 | 2.73E-10 | 1.10E-08 | 1.535127 |
| CEP55 | 553.94467 | 0.631823 | 0.10012 | 6.31063382 | 2.78E-10 | 1.12E-08 | 1.549522 |
| COPS2 | 1900.3156 | 0.624408 | 0.099126 | 6.29910462 | 2.99E-10 | 1.19E-08 | 1.541578 |
| RACGAP1 | 1417.6414 | 0.71857 | 0.114095 | 6.29799489 | 3.02E-10 | 1.20E-08 | 1.64555 |
| FBXO3 | 812.61525 | 0.668426 | 0.106501 | 6.27622003 | 3.47E-10 | 1.36E-08 | 1.589338 |
| KAT6A | 1271.1328 | 0.607733 | 0.097368 | 6.24158201 | 4.33E-10 | 1.68E-08 | 1.523863 |
| SMAD4 | 2747.258 | 0.672526 | 0.108433 | 6.20224871 | 5.57E-10 | 2.09E-08 | 1.593861 |
| HMGCS1 | 3460.1494 | 1.292895 | 0.208697 | 6.19508313 | 5.83E-10 | 2.17E-08 | 2.450192 |
| SESN1 | 675.72859 | 0.676747 | 0.109271 | 6.19330518 | 5.89E-10 | 2.19E-08 | 1.598531 |
| RPL36A | 7410.4744 | 0.600612 | 0.097819 | 6.14003391 | 8.25E-10 | 2.99E-08 | 1.516359 |
| FAM98B | 1838.7374 | 0.714964 | 0.116609 | 6.13130581 | 8.72E-10 | 3.15E-08 | 1.641442 |
| APPBP2 | 525.214 | 1.016395 | 0.165785 | 6.1307842 | 8.74E-10 | 3.15E-08 | 2.022857 |
| LDB2 | 814.46727 | 0.788333 | 0.128817 | 6.11978325 | 9.37E-10 | 3.36E-08 | 1.727077 |
| ARRB1 | 1708.1059 | 0.613035 | 0.100247 | 6.11525702 | 9.64E-10 | 3.43E-08 | 1.529473 |
| IGFBP5 | 427.05617 | 1.472909 | 0.240988 | 6.11196504 | 9.84E-10 | 3.50E-08 | 2.77581 |
| TFDP2 | 1950.4877 | 0.626221 | 0.103327 | 6.06055524 | 1.36E-09 | 4.68E-08 | 1.543516 |
| MLLT3 | 234.40518 | 1.417715 | 0.233948 | 6.05995034 | 1.36E-09 | 4.68E-08 | 2.67162 |
| MMRN1 | 12.317318 | 6.944889 | 1.146737 | 6.05621937 | 1.39E-09 | 4.78E-08 | 123.2026 |
| USP3 | 369.07037 | 0.725091 | 0.119842 | 6.05040131 | 1.44E-09 | 4.94E-08 | 1.653005 |
| PPP1R3B | 3657.6142 | 0.659536 | 0.109373 | 6.03015533 | 1.64E-09 | 5.54E-08 | 1.579575 |
| TFCP2 | 1317.1645 | 0.87537 | 0.145313 | 6.02403044 | 1.70E-09 | 5.69E-08 | 1.834478 |
| CADM2 | 281.74036 | 0.811581 | 0.134798 | 6.02070937 | 1.74E-09 | 5.77E-08 | 1.755134 |
| NFYA | 924.16441 | 0.606362 | 0.10078 | 6.01668664 | 1.78E-09 | 5.90E-08 | 1.522415 |
| RPS10-NU | 328.21972 | 1.200061 | 0.200163 | 5.99540567 | 2.03E-09 | 6.58E-08 | 2.297493 |
| NUSAP1 | 2142.6383 | 0.732466 | 0.122582 | 5.97532246 | 2.30E-09 | 7.29E-08 | 1.661477 |
| HECA | 303.76756 | 0.678034 | 0.114097 | 5.94259207 | 2.81E-09 | 8.73E-08 | 1.599958 |
| RLN2 | 65.147481 | 1.522798 | 0.256748 | 5.93110194 | 3.01E-09 | 9.17E-08 | 2.873478 |
| 5-Mar | 594.4838 | 0.971155 | 0.163832 | 5.92775199 | 3.07E-09 | 9.32E-08 | 1.960409 |
| TTF2 | 2292.3783 | 0.634517 | 0.107313 | 5.91279018 | 3.36E-09 | 1.01E-07 | 1.552417 |
| DNAJA1 | 3141.7138 | 0.633603 | 0.107333 | 5.90314197 | 3.57E-09 | 1.06E-07 | 1.551435 |

| | | | | | | | |
|---|---|---|---|---|---|---|---|
| DDIT4L | 151.94926 | 0.864017 | 0.14876 | 5.80812568 | 6.32E-09 | 1.77E-07 | 1.820099 |
| CAMKK2 | 1183.6371 | 0.65178 | 0.112273 | 5.80532745 | 6.42E-09 | 1.79E-07 | 1.571106 |
| GTF3C3 | 1506.2908 | 0.612725 | 0.105752 | 5.79398898 | 6.87E-09 | 1.90E-07 | 1.529144 |
| ASDURF | 12.604553 | 6.978023 | 1.205026 | 5.79076446 | 7.01E-09 | 1.94E-07 | 126.0649 |
| MAPK8 | 1458.4922 | 0.603108 | 0.104194 | 5.78831483 | 7.11E-09 | 1.96E-07 | 1.518985 |
| SP4 | 655.77652 | 0.764239 | 0.13203 | 5.78838594 | 7.11E-09 | 1.96E-07 | 1.698474 |
| ACACA | 2591.3802 | 0.795082 | 0.137474 | 5.78351589 | 7.32E-09 | 2.00E-07 | 1.735176 |
| TCAF1 | 2113.9042 | 0.705579 | 0.122184 | 5.77473043 | 7.71E-09 | 2.10E-07 | 1.630799 |
| AMPH | 322.64146 | 0.646688 | 0.112558 | 5.74536833 | 9.17E-09 | 2.44E-07 | 1.56557 |
| TMSB15A | 610.36138 | 0.645241 | 0.11235 | 5.74314812 | 9.29E-09 | 2.47E-07 | 1.564001 |
| COL2A1 | 275.59681 | 1.37501 | 0.240543 | 5.71626328 | 1.09E-08 | 2.85E-07 | 2.593697 |
| TIAM1 | 966.84225 | 0.685258 | 0.120201 | 5.70093644 | 1.19E-08 | 3.07E-07 | 1.607989 |
| RBM12B | 1450.443 | 0.872784 | 0.153192 | 5.6973173 | 1.22E-08 | 3.12E-07 | 1.831194 |
| HSPA1L | 10.018414 | 6.657596 | 1.17003 | 5.69010583 | 1.27E-08 | 3.24E-07 | 100.9569 |
| ENC1 | 560.74638 | 1.140811 | 0.200735 | 5.68316185 | 1.32E-08 | 3.35E-07 | 2.20505 |
| AASDH | 449.18607 | 0.615003 | 0.108556 | 5.66533074 | 1.47E-08 | 3.68E-07 | 1.531561 |
| UBE2C | 1965.7942 | 0.610424 | 0.107978 | 5.65321684 | 1.57E-08 | 3.93E-07 | 1.526708 |
| SGO2 | 1073.8163 | 0.669709 | 0.119002 | 5.62772272 | 1.83E-08 | 4.48E-07 | 1.590752 |
| PLAA | 1714.3762 | 0.636728 | 0.113436 | 5.61309915 | 1.99E-08 | 4.84E-07 | 1.554799 |
| TCAF1P1 | 203.37052 | 0.875706 | 0.156269 | 5.60384327 | 2.10E-08 | 5.08E-07 | 1.834906 |
| LAMTOR3 | 608.13367 | 0.612262 | 0.109281 | 5.60261325 | 2.11E-08 | 5.11E-07 | 1.528654 |
| 8-Mar | 464.19286 | 0.899027 | 0.160781 | 5.5916218 | 2.25E-08 | 5.35E-07 | 1.864808 |
| KIAA1586 | 446.6495 | 0.67732 | 0.121175 | 5.58960581 | 2.28E-08 | 5.40E-07 | 1.599166 |
| HES5 | 9.6327084 | 6.586702 | 1.180092 | 5.58151791 | 2.38E-08 | 5.58E-07 | 96.11583 |
| TNPO1 | 7968.357 | 0.657231 | 0.117968 | 5.5712836 | 2.53E-08 | 5.87E-07 | 1.577053 |
| SLC38A4 | 548.99264 | 0.633049 | 0.114143 | 5.54609589 | 2.92E-08 | 6.62E-07 | 1.550839 |
| ZNF85 | 43.95054 | 1.956411 | 0.355058 | 5.51011498 | 3.59E-08 | 7.99E-07 | 3.880952 |
| TP53INP1 | 389.40741 | 0.741788 | 0.134683 | 5.50765809 | 3.64E-08 | 8.08E-07 | 1.672247 |
| LHX2 | 26.734405 | 3.355425 | 0.614106 | 5.46392219 | 4.66E-08 | 9.96E-07 | 10.2349 |
| BCL9 | 2030.5144 | 0.587783 | 0.107852 | 5.44992316 | 5.04E-08 | 1.07E-06 | 1.502935 |
| ZFP82 | 279.71761 | 0.714855 | 0.131327 | 5.44332041 | 5.23E-08 | 1.10E-06 | 1.641319 |
| NSMCE2 | 333.18337 | 0.67204 | 0.123499 | 5.44165675 | 5.28E-08 | 1.11E-06 | 1.593324 |
| PIFO | 212.6137 | 0.800908 | 0.147181 | 5.44164162 | 5.28E-08 | 1.11E-06 | 1.742197 |
| ALX1 | 43.192696 | 1.568072 | 0.288449 | 5.43621204 | 5.44E-08 | 1.14E-06 | 2.965082 |
| CENPI | 420.66046 | 0.846111 | 0.156324 | 5.41256267 | 6.21E-08 | 1.28E-06 | 1.797649 |
| ZIC1 | 40.086314 | 2.480806 | 0.459349 | 5.40069871 | 6.64E-08 | 1.36E-06 | 5.582092 |
| SIX6 | 14.979773 | 5.140841 | 0.952188 | 5.39897859 | 6.70E-08 | 1.37E-06 | 35.28152 |
| PAQR5 | 514.50487 | 0.612384 | 0.113493 | 5.39578967 | 6.82E-08 | 1.39E-06 | 1.528784 |
| GAS2L3 | 394.97729 | 0.651939 | 0.121664 | 5.35853022 | 8.39E-08 | 1.67E-06 | 1.571279 |
| DCTN6 | 496.72549 | 0.662968 | 0.123977 | 5.34750423 | 8.92E-08 | 1.75E-06 | 1.583336 |
| RSBN1 | 582.92231 | 0.710928 | 0.132951 | 5.34729642 | 8.93E-08 | 1.75E-06 | 1.636856 |
| FBXO33 | 342.83661 | 0.647169 | 0.12105 | 5.34627583 | 8.98E-08 | 1.76E-06 | 1.566092 |
| RTL1 | 14.137441 | 5.694015 | 1.066647 | 5.33823921 | 9.39E-08 | 1.82E-06 | 51.76896 |
| PHACTR1 | 236.03441 | 0.710964 | 0.133247 | 5.33569109 | 9.52E-08 | 1.84E-06 | 1.636898 |

| | | | | | | | |
|---|---|---|---|---|---|---|---|
| ZBTB40 | 764.33009 | 0.614739 | 0.115277 | 5.33268978 | 9.68E-08 | 1.86E-06 | 1.531281 |
| ZNF429 | 78.264261 | 1.861428 | 0.349212 | 5.33036067 | 9.80E-08 | 1.88E-06 | 3.633672 |
| PRSS23 | 300.21571 | 0.826442 | 0.155204 | 5.32486195 | 1.01E-07 | 1.93E-06 | 1.773306 |
| LDAH | 682.65796 | 0.649786 | 0.122931 | 5.28577993 | 1.25E-07 | 2.32E-06 | 1.568935 |
| ATF7IP | 3260.7211 | 0.690568 | 0.130702 | 5.28352123 | 1.27E-07 | 2.34E-06 | 1.613918 |
| OSGEPL1 | 298.12794 | 0.679826 | 0.129463 | 5.25112426 | 1.51E-07 | 2.75E-06 | 1.601946 |
| CTDSPL2 | 1883.7567 | 0.656747 | 0.125477 | 5.23402098 | 1.66E-07 | 2.96E-06 | 1.576524 |
| MAP2K6 | 1142.2527 | 0.933425 | 0.178392 | 5.23244865 | 1.67E-07 | 2.98E-06 | 1.909805 |
| PCLAF | 1325.6893 | 0.624848 | 0.120068 | 5.2041242 | 1.95E-07 | 3.40E-06 | 1.542048 |
| ZNF350 | 105.53896 | 1.133963 | 0.218411 | 5.19187061 | 2.08E-07 | 3.60E-06 | 2.194608 |
| KATNAL1 | 421.99546 | 0.731029 | 0.140961 | 5.18603577 | 2.15E-07 | 3.70E-06 | 1.659823 |
| HSPA2 | 438.04217 | 0.616056 | 0.119048 | 5.17485763 | 2.28E-07 | 3.91E-06 | 1.532679 |
| KCNAB1 | 77.020153 | 1.096711 | 0.212424 | 5.16284871 | 2.43E-07 | 4.12E-06 | 2.138666 |
| DMXL2 | 1141.113 | 0.828966 | 0.160854 | 5.15353164 | 2.56E-07 | 4.31E-06 | 1.776411 |
| DNAJC6 | 209.05142 | 0.72036 | 0.139997 | 5.14555001 | 2.67E-07 | 4.47E-06 | 1.647593 |
| C15orf41 | 339.43537 | 0.808343 | 0.157764 | 5.12374344 | 3.00E-07 | 4.97E-06 | 1.751199 |
| SESN3 | 3537.3636 | 0.676935 | 0.132635 | 5.10374634 | 3.33E-07 | 5.46E-06 | 1.598739 |
| SHROOM2 | 1212.7664 | 0.633203 | 0.124275 | 5.09519779 | 3.48E-07 | 5.68E-06 | 1.551005 |
| PBK | 234.92102 | 0.741996 | 0.146342 | 5.07028877 | 3.97E-07 | 6.37E-06 | 1.672488 |
| ZNF287 | 154.52587 | 1.33728 | 0.264463 | 5.05657806 | 4.27E-07 | 6.81E-06 | 2.526745 |
| AC068547 | 143.83045 | 0.86333 | 0.170897 | 5.05174575 | 4.38E-07 | 6.96E-06 | 1.819233 |
| MN1 | 440.64215 | 0.628159 | 0.125136 | 5.01982361 | 5.17E-07 | 8.02E-06 | 1.545592 |
| SNRPD3 | 2002.5554 | 0.703872 | 0.140489 | 5.01014149 | 5.44E-07 | 8.38E-06 | 1.62887 |
| ZIC4 | 8.0763135 | 6.351914 | 1.269882 | 5.00197244 | 5.67E-07 | 8.66E-06 | 81.68015 |
| ZNF512 | 1232.6714 | 0.669282 | 0.133854 | 5.00008613 | 5.73E-07 | 8.73E-06 | 1.590281 |
| TC2N | 540.44241 | 0.769641 | 0.153951 | 4.99924715 | 5.76E-07 | 8.76E-06 | 1.704846 |
| HNRNPA1I | 358.25715 | 0.587309 | 0.117884 | 4.98209737 | 6.29E-07 | 9.45E-06 | 1.502442 |
| KLHL24 | 902.44997 | 0.725483 | 0.145642 | 4.981272 | 6.32E-07 | 9.48E-06 | 1.653455 |
| PDCD10 | 1047.5562 | 0.61457 | 0.123401 | 4.98027665 | 6.35E-07 | 9.52E-06 | 1.531102 |
| CDON | 906.69006 | 0.655931 | 0.132804 | 4.93908074 | 7.85E-07 | 1.14E-05 | 1.575632 |
| LIX1L | 157.78315 | 0.794868 | 0.160933 | 4.93912933 | 7.85E-07 | 1.14E-05 | 1.734919 |
| SEMA3C | 42.353303 | 1.681458 | 0.340864 | 4.93293381 | 8.10E-07 | 1.17E-05 | 3.20752 |
| RADX | 331.74913 | 0.696684 | 0.141312 | 4.93010276 | 8.22E-07 | 1.18E-05 | 1.620775 |
| AC091959 | 184.56339 | 1.453681 | 0.29518 | 4.92472056 | 8.45E-07 | 1.21E-05 | 2.739061 |
| RNASE1 | 83.726945 | 1.00698 | 0.205651 | 4.89655829 | 9.75E-07 | 1.38E-05 | 2.0097 |
| ZNF521 | 659.60179 | 0.900231 | 0.184247 | 4.88599098 | 1.03E-06 | 1.44E-05 | 1.866365 |
| GTDC1 | 379.70647 | 0.772588 | 0.158274 | 4.88134242 | 1.05E-06 | 1.47E-05 | 1.708331 |
| RTF1 | 2479.3791 | 0.585407 | 0.120363 | 4.8636774 | 1.15E-06 | 1.58E-05 | 1.500462 |
| MID1 | 764.73169 | 0.741207 | 0.152856 | 4.84905223 | 1.24E-06 | 1.69E-05 | 1.671573 |
| GSTM3 | 93.546963 | 1.247038 | 0.259089 | 4.81316933 | 1.49E-06 | 1.97E-05 | 2.373537 |
| BRWD3 | 1545.6852 | 0.654482 | 0.136068 | 4.80996162 | 1.51E-06 | 2.00E-05 | 1.57405 |
| MBNL3 | 472.00184 | 0.676282 | 0.140637 | 4.80869453 | 1.52E-06 | 2.01E-05 | 1.598016 |
| IDI1 | 1079.5399 | 0.981678 | 0.204737 | 4.7948303 | 1.63E-06 | 2.14E-05 | 1.974761 |
| CFL2 | 482.83455 | 0.772579 | 0.161288 | 4.79004868 | 1.67E-06 | 2.18E-05 | 1.708321 |

| | | | | | | | |
|---|---|---|---|---|---|---|---|
| GLYATL1 | 206.31315 | 0.728649 | 0.152642 | 4.77359171 | 1.81E-06 | 2.34E-05 | 1.657086 |
| TUBD1 | 316.03332 | 0.674576 | 0.141782 | 4.75784393 | 1.96E-06 | 2.51E-05 | 1.596128 |
| HSPA8 | 22045.483 | 0.955887 | 0.201104 | 4.75320467 | 2.00E-06 | 2.56E-05 | 1.939771 |
| RASL12 | 223.60936 | 0.613831 | 0.129137 | 4.75332386 | 2.00E-06 | 2.56E-05 | 1.530317 |
| ATG4A | 174.79719 | 0.720441 | 0.151646 | 4.75081844 | 2.03E-06 | 2.58E-05 | 1.647686 |
| FEZF2 | 11.652692 | 4.909381 | 1.040912 | 4.71642389 | 2.40E-06 | 3.00E-05 | 30.05184 |
| ZNF730 | 180.54807 | 0.855156 | 0.182621 | 4.68269444 | 2.83E-06 | 3.47E-05 | 1.808955 |
| PPP2R3A | 613.25329 | 0.709155 | 0.151714 | 4.67430025 | 2.95E-06 | 3.60E-05 | 1.634846 |
| EIF2AK2 | 2413.2073 | 0.596536 | 0.128116 | 4.65620808 | 3.22E-06 | 3.88E-05 | 1.512082 |
| LCORL | 553.98839 | 0.740732 | 0.159583 | 4.6416817 | 3.46E-06 | 4.10E-05 | 1.671023 |
| ZNF260 | 992.38849 | 0.657694 | 0.141765 | 4.63931796 | 3.50E-06 | 4.14E-05 | 1.577559 |
| VGLL3 | 1177.5756 | 0.695857 | 0.150693 | 4.61772775 | 3.88E-06 | 4.55E-05 | 1.619847 |
| DLK1 | 649.12316 | 1.32471 | 0.287004 | 4.61564263 | 3.92E-06 | 4.58E-05 | 2.504826 |
| SRSF12 | 308.69281 | 0.659113 | 0.143662 | 4.58793236 | 4.48E-06 | 5.14E-05 | 1.579111 |
| DDHD2 | 735.6743 | 0.842995 | 0.183757 | 4.58755747 | 4.48E-06 | 5.14E-05 | 1.79377 |
| VAX1 | 5.2796352 | 5.731893 | 1.250207 | 4.58475427 | 4.55E-06 | 5.19E-05 | 53.14613 |
| PSD3 | 666.5647 | 0.700846 | 0.15374 | 4.5586396 | 5.15E-06 | 5.77E-05 | 1.625458 |
| CREB1 | 1653.9077 | 0.66497 | 0.145954 | 4.55602401 | 5.21E-06 | 5.81E-05 | 1.585535 |
| SOX9 | 475.33585 | 0.601225 | 0.131946 | 4.55660906 | 5.20E-06 | 5.81E-05 | 1.517004 |
| KLHL8 | 285.10718 | 0.672055 | 0.147798 | 4.547126 | 5.44E-06 | 6.01E-05 | 1.593341 |
| ZNF138 | 269.43506 | 0.603257 | 0.13322 | 4.52828698 | 5.95E-06 | 6.49E-05 | 1.519143 |
| RRM1 | 2937.4127 | 0.694276 | 0.153409 | 4.52566205 | 6.02E-06 | 6.56E-05 | 1.618072 |
| SULT1C4 | 83.533878 | 1.006251 | 0.222423 | 4.52404338 | 6.07E-06 | 6.61E-05 | 2.008685 |
| LYRM4 | 851.8445 | 0.593705 | 0.131937 | 4.49989329 | 6.80E-06 | 7.30E-05 | 1.509117 |
| FAM72D | 224.56016 | 0.79414 | 0.176635 | 4.49592651 | 6.93E-06 | 7.41E-05 | 1.734043 |
| NR2F1 | 7.7925516 | 5.644153 | 1.261283 | 4.47493014 | 7.64E-06 | 8.07E-05 | 50.01028 |
| FRG1 | 279.09796 | 0.599684 | 0.13468 | 4.45267017 | 8.48E-06 | 8.81E-05 | 1.515384 |
| DYM | 1408.9272 | 0.669224 | 0.150909 | 4.43460939 | 9.22E-06 | 9.46E-05 | 1.590217 |
| ZNF184 | 215.05551 | 0.653996 | 0.147636 | 4.42977722 | 9.43E-06 | 9.64E-05 | 1.573521 |
| VAMP4 | 244.15647 | 0.717509 | 0.162053 | 4.42762508 | 9.53E-06 | 9.72E-05 | 1.644341 |
| RSF1 | 2157.0817 | 0.596291 | 0.134756 | 4.42497435 | 9.65E-06 | 9.81E-05 | 1.511825 |
| NEIL3 | 253.81597 | 0.595569 | 0.13467 | 4.42243393 | 9.76E-06 | 9.91E-05 | 1.511068 |
| AURKC | 90.971088 | 1.026259 | 0.232866 | 4.40707799 | 1.05E-05 | 0.000105 | 2.036736 |
| METTL14 | 824.90539 | 0.801095 | 0.183125 | 4.37457527 | 1.22E-05 | 0.00012 | 1.742423 |
| C12orf29 | 307.22507 | 0.588111 | 0.134491 | 4.37285325 | 1.23E-05 | 0.00012 | 1.503277 |
| WNT7B | 38.479568 | 1.664362 | 0.382054 | 4.35635182 | 1.32E-05 | 0.000128 | 3.169734 |
| GAD2 | 6.2252267 | 5.524736 | 1.268274 | 4.35610736 | 1.32E-05 | 0.000128 | 46.03745 |
| SIX3 | 123.48417 | 1.449046 | 0.335995 | 4.31270523 | 1.61E-05 | 0.000153 | 2.730274 |
| SELENOP | 626.73359 | 0.762458 | 0.177612 | 4.29281659 | 1.76E-05 | 0.000165 | 1.696378 |
| EMSY | 793.46965 | 0.693565 | 0.161807 | 4.28636469 | 1.82E-05 | 0.00017 | 1.617275 |
| ZBTB41 | 251.19619 | 0.585366 | 0.136578 | 4.28593695 | 1.82E-05 | 0.00017 | 1.500419 |
| ERCC4 | 315.97634 | 1.094826 | 0.255621 | 4.28299976 | 1.84E-05 | 0.000172 | 2.135873 |
| PAPOLG | 803.11112 | 0.651072 | 0.152155 | 4.27900301 | 1.88E-05 | 0.000175 | 1.570334 |
| AC008012 | 320.27515 | 0.618088 | 0.145261 | 4.25501237 | 2.09E-05 | 0.000192 | 1.53484 |

| | | | | | | | |
|---|---|---|---|---|---|---|---|
| SMIM24 | 98.501333 | 1.123638 | 0.264228 | 4.25252711 | 2.11E-05 | 0.000194 | 2.178958 |
| HOMER1 | 1261.233 | 0.621923 | 0.146468 | 4.2461358 | 2.17E-05 | 0.000199 | 1.538925 |
| PRDM10 | 468.99457 | 0.623018 | 0.147438 | 4.22563769 | 2.38E-05 | 0.000215 | 1.540094 |
| DMAC1 | 853.33836 | 0.649639 | 0.154296 | 4.21034573 | 2.55E-05 | 0.000227 | 1.568776 |
| TUBB4B | 5727.2992 | 0.613751 | 0.146463 | 4.19047423 | 2.78E-05 | 0.000246 | 1.530232 |
| JCAD | 152.30957 | 0.779854 | 0.1863 | 4.18601036 | 2.84E-05 | 0.00025 | 1.716957 |
| RGL1 | 64.645358 | 0.927115 | 0.221754 | 4.18083752 | 2.90E-05 | 0.000255 | 1.90147 |
| C14orf119 | 1021.8513 | 0.600008 | 0.143605 | 4.17819267 | 2.94E-05 | 0.000258 | 1.515725 |
| HSDL2 | 566.35498 | 0.618298 | 0.14814 | 4.1737276 | 3.00E-05 | 0.000262 | 1.535063 |
| EXOC6 | 260.16679 | 0.614099 | 0.147644 | 4.15932 | 3.19E-05 | 0.000277 | 1.530602 |
| TRPM3 | 275.18859 | 0.656364 | 0.157833 | 4.15860237 | 3.20E-05 | 0.000277 | 1.576105 |
| NF1 | 1670.2274 | 0.637392 | 0.153711 | 4.14668933 | 3.37E-05 | 0.000291 | 1.555515 |
| ZNF567 | 317.21232 | 0.665857 | 0.161253 | 4.12927957 | 3.64E-05 | 0.000311 | 1.586511 |
| CEP57L1 | 281.94957 | 0.787041 | 0.192803 | 4.08210295 | 4.46E-05 | 0.000371 | 1.725531 |
| PRKAA1 | 761.07434 | 0.652948 | 0.160309 | 4.07305874 | 4.64E-05 | 0.000384 | 1.572378 |
| FANK1 | 25.320747 | 1.781277 | 0.437375 | 4.07265618 | 4.65E-05 | 0.000384 | 3.437303 |
| CENPL | 247.87631 | 0.604624 | 0.148941 | 4.05948176 | 4.92E-05 | 0.000404 | 1.520582 |
| CCDC177 | 187.14228 | 0.7572 | 0.187338 | 4.04189967 | 5.30E-05 | 0.000432 | 1.690207 |
| SLFN11 | 27.885384 | 1.577092 | 0.390762 | 4.03593939 | 5.44E-05 | 0.000441 | 2.983678 |
| HYLS1 | 158.2116 | 0.645362 | 0.16062 | 4.01793335 | 5.87E-05 | 0.000472 | 1.564131 |
| RNASE4 | 97.782981 | 1.221814 | 0.304112 | 4.01764978 | 5.88E-05 | 0.000472 | 2.332397 |
| CDC27 | 1395.9472 | 0.690277 | 0.171942 | 4.01459281 | 5.95E-05 | 0.000477 | 1.613593 |
| RAB3C | 414.82264 | 0.684777 | 0.171154 | 4.00095165 | 6.31E-05 | 0.0005 | 1.607454 |
| LCOR | 1072.9004 | 0.651366 | 0.163036 | 3.99522159 | 6.46E-05 | 0.000511 | 1.570654 |
| C14orf132 | 1209.5184 | 0.596045 | 0.149388 | 3.98991888 | 6.61E-05 | 0.00052 | 1.511567 |
| CCNY | 1783.082 | 0.668265 | 0.167719 | 3.98443287 | 6.76E-05 | 0.000532 | 1.589161 |
| GEM | 97.216475 | 0.89749 | 0.226488 | 3.96264235 | 7.41E-05 | 0.000576 | 1.862822 |
| ZBTB10 | 921.69014 | 0.61398 | 0.155273 | 3.95421054 | 7.68E-05 | 0.000592 | 1.530476 |
| MGAT5 | 1402.3872 | 0.600672 | 0.15205 | 3.95050389 | 7.80E-05 | 0.0006 | 1.516423 |
| EDNRB | 794.64071 | 0.632714 | 0.160558 | 3.9407104 | 8.12E-05 | 0.000621 | 1.550479 |
| ZNF658 | 179.95225 | 0.806646 | 0.206463 | 3.90697363 | 9.35E-05 | 0.0007 | 1.74914 |
| GNAI1 | 560.6823 | 0.58848 | 0.151024 | 3.896602 | 9.76E-05 | 0.000727 | 1.503662 |
| TRMT5 | 779.99166 | 0.651665 | 0.168205 | 3.87422086 | 0.000107 | 0.000786 | 1.57098 |
| STMN2 | 65.846286 | 1.550403 | 0.40043 | 3.87184812 | 0.000108 | 0.000793 | 2.92899 |
| CAB39L | 177.93625 | 0.639104 | 0.165222 | 3.86814464 | 0.00011 | 0.000803 | 1.557362 |
| PDCD4 | 779.55934 | 0.639508 | 0.165479 | 3.86458437 | 0.000111 | 0.000813 | 1.557798 |
| SIK1 | 151.26659 | 0.744792 | 0.192764 | 3.86375654 | 0.000112 | 0.000815 | 1.675732 |
| ERCC6 | 313.51421 | 0.662736 | 0.172048 | 3.85203391 | 0.000117 | 0.000847 | 1.583082 |
| SRR | 179.40199 | 0.835595 | 0.217608 | 3.83991393 | 0.000123 | 0.000883 | 1.784593 |
| RBM24 | 67.649244 | 0.898573 | 0.234155 | 3.83752171 | 0.000124 | 0.00089 | 1.864222 |
| PPM1K | 436.12864 | 0.613484 | 0.160461 | 3.82326669 | 0.000132 | 0.000936 | 1.52995 |
| AC005524 | 13.701163 | 2.544794 | 0.666238 | 3.81964724 | 0.000134 | 0.000948 | 5.83525 |
| KLF13 | 791.98012 | 0.619568 | 0.164568 | 3.76482205 | 0.000167 | 0.001148 | 1.536415 |
| FMN1 | 153.9822 | 0.899104 | 0.238855 | 3.7642275 | 0.000167 | 0.00115 | 1.864908 |

| | | | | | | | |
|---|---|---|---|---|---|---|---|
| MEX3B | 358.40252 | 0.651297 | 0.17332 | 3.75776594 | 0.000171 | 0.001174 | 1.57058 |
| SOAT1 | 413.68834 | 0.667777 | 0.177974 | 3.75210398 | 0.000175 | 0.001198 | 1.588623 |
| SWT1 | 229.01477 | 0.600239 | 0.160967 | 3.72895625 | 0.000192 | 0.001292 | 1.515967 |
| CXXC4 | 119.46299 | 0.714402 | 0.191768 | 3.72535524 | 0.000195 | 0.001308 | 1.640804 |
| DACH1 | 34.116308 | 1.663523 | 0.447765 | 3.71516908 | 0.000203 | 0.001354 | 3.167891 |
| CMBL | 539.9489 | 0.596327 | 0.160855 | 3.70722626 | 0.00021 | 0.00139 | 1.511863 |
| PLCXD2 | 101.35843 | 0.654442 | 0.176531 | 3.70723 | 0.00021 | 0.00139 | 1.574007 |
| PABPC5 | 99.559832 | 0.872742 | 0.235601 | 3.70431369 | 0.000212 | 0.001404 | 1.83114 |
| ZFHX4 | 139.03303 | 0.857715 | 0.231631 | 3.70294533 | 0.000213 | 0.001409 | 1.812166 |
| LEF1 | 49.86739 | 1.626976 | 0.439705 | 3.70015164 | 0.000215 | 0.001424 | 3.088648 |
| OTUB2 | 165.46614 | 0.818594 | 0.222099 | 3.68571492 | 0.000228 | 0.001496 | 1.763686 |
| EMX2 | 5.8937136 | 4.475171 | 1.218119 | 3.67383621 | 0.000239 | 0.001558 | 22.24132 |
| ZNF649 | 804.61073 | 0.696917 | 0.190056 | 3.66689758 | 0.000246 | 0.001598 | 1.621037 |
| AMER2 | 18.75702 | 1.999333 | 0.546334 | 3.6595473 | 0.000253 | 0.001634 | 3.998152 |
| TDRD7 | 198.78215 | 0.661583 | 0.18147 | 3.64569521 | 0.000267 | 0.001713 | 1.581817 |
| LEMD1 | 30.705345 | 1.298983 | 0.357062 | 3.63797688 | 0.000275 | 0.001756 | 2.460554 |
| LYPD6 | 188.8159 | 0.599494 | 0.1648 | 3.63771129 | 0.000275 | 0.001757 | 1.515185 |
| CCDC141 | 58.116963 | 1.024134 | 0.281851 | 3.63360292 | 0.000279 | 0.001781 | 2.033739 |
| GBX2 | 51.113108 | 1.135292 | 0.313175 | 3.62510855 | 0.000289 | 0.001831 | 2.19663 |
| TPTEP2-CS | 141.28938 | 0.712288 | 0.196978 | 3.61608246 | 0.000299 | 0.001887 | 1.6384 |
| PRDM16 | 22.622173 | 2.063049 | 0.571562 | 3.60949313 | 0.000307 | 0.001927 | 4.178684 |
| MSANTD3- | 925.63657 | 0.808078 | 0.223982 | 3.60777151 | 0.000309 | 0.001937 | 1.750877 |
| ZNF483 | 152.2454 | 0.641232 | 0.178137 | 3.59965107 | 0.000319 | 0.001992 | 1.559661 |
| TNFSF12-T | 6.0973863 | 5.932804 | 1.664646 | 3.56400364 | 0.000365 | 0.002236 | 61.08746 |
| AC114490 | 52.781725 | 1.436394 | 0.405518 | 3.54212258 | 0.000397 | 0.002405 | 2.706436 |
| PAFAH2 | 411.32956 | 0.646019 | 0.182602 | 3.53785747 | 0.000403 | 0.002439 | 1.564844 |
| ZNF624 | 59.06958 | 1.183573 | 0.334745 | 3.53574485 | 0.000407 | 0.002457 | 2.271387 |
| SDAD1P1 | 70.614044 | 0.796293 | 0.225548 | 3.53047674 | 0.000415 | 0.002495 | 1.736633 |
| IL1RAPL1 | 7.1576222 | 3.122935 | 0.885347 | 3.52735671 | 0.00042 | 0.002523 | 8.711586 |
| ZEB1 | 28.469031 | 1.614459 | 0.459823 | 3.51104242 | 0.000446 | 0.002662 | 3.061968 |
| GPSM2 | 437.85224 | 0.690767 | 0.196869 | 3.50876789 | 0.00045 | 0.002682 | 1.614141 |
| GPRIN1 | 333.37679 | 0.590676 | 0.168415 | 3.50725353 | 0.000453 | 0.002695 | 1.505952 |
| TBC1D9 | 68.02731 | 0.772398 | 0.221405 | 3.48861884 | 0.000486 | 0.002866 | 1.708107 |
| RSPO3 | 8.3723973 | 3.675229 | 1.054672 | 3.48471363 | 0.000493 | 0.002898 | 12.7748 |
| FCHSD2 | 408.90889 | 0.600382 | 0.17287 | 3.47301877 | 0.000515 | 0.003009 | 1.516118 |
| DLL1 | 179.95828 | 0.800669 | 0.230629 | 3.47166897 | 0.000517 | 0.00302 | 1.741908 |
| UBE2W | 641.53381 | 0.627462 | 0.181062 | 3.46545943 | 0.000529 | 0.003079 | 1.544845 |
| ZNF699 | 65.619552 | 0.850348 | 0.246044 | 3.45608659 | 0.000548 | 0.003168 | 1.802935 |
| AL121594. | 9.5979574 | 3.720672 | 1.080895 | 3.44221297 | 0.000577 | 0.00331 | 13.1836 |
| ZBTB16 | 12.598149 | 2.657469 | 0.772585 | 3.43971155 | 0.000582 | 0.003336 | 6.309253 |
| PXMP4 | 120.22786 | 0.655954 | 0.191943 | 3.41745201 | 0.000632 | 0.003572 | 1.575658 |
| RPS6KA5 | 155.43333 | 0.599208 | 0.175388 | 3.41647159 | 0.000634 | 0.003583 | 1.514885 |
| ZSCAN20 | 98.825281 | 0.737629 | 0.215949 | 3.41574876 | 0.000636 | 0.00359 | 1.667433 |
| ZNF518B | 376.32344 | 0.625501 | 0.183387 | 3.41083133 | 0.000648 | 0.003645 | 1.542746 |

| | | | | | | | |
|---|---|---|---|---|---|---|---|
| LRRC55 | 107.98325 | 0.679143 | 0.199562 | 3.40316487 | 0.000666 | 0.003732 | 1.601188 |
| FGF3 | 3.1160292 | 4.946061 | 1.454875 | 3.39964731 | 0.000675 | 0.003776 | 30.82568 |
| ESCO2 | 419.04077 | 0.979191 | 0.288677 | 3.3919914 | 0.000694 | 0.003864 | 1.97136 |
| KIAA0513 | 245.5094 | 0.637232 | 0.188192 | 3.38606571 | 0.000709 | 0.003939 | 1.555342 |
| HHEX | 48.213084 | 0.918095 | 0.274386 | 3.34599925 | 0.00082 | 0.004437 | 1.889618 |
| ST3GAL6 | 171.64709 | 0.605393 | 0.181264 | 3.33984554 | 0.000838 | 0.004516 | 1.521393 |
| TTC33 | 445.92546 | 0.603399 | 0.180832 | 3.33679984 | 0.000847 | 0.004555 | 1.519292 |
| CASP6 | 471.47178 | 0.587804 | 0.176281 | 3.33446373 | 0.000855 | 0.004583 | 1.502958 |
| SCML1 | 39.955895 | 0.961085 | 0.289241 | 3.32278653 | 0.000891 | 0.004747 | 1.946773 |
| BCL2L11 | 582.42182 | 0.623011 | 0.187881 | 3.31598717 | 0.000913 | 0.00484 | 1.540086 |
| GRASP | 101.38559 | 0.771033 | 0.232741 | 3.31283791 | 0.000924 | 0.004884 | 1.706491 |
| LZIC | 598.7195 | 0.61889 | 0.186911 | 3.31114927 | 0.000929 | 0.004908 | 1.535693 |
| HSP90AB2 | 46.770695 | 0.845782 | 0.255858 | 3.30567421 | 0.000947 | 0.004989 | 1.797239 |
| WDR49 | 13.549765 | 2.20735 | 0.668437 | 3.30225704 | 0.000959 | 0.005041 | 4.61826 |
| AC092835 | 70.6307 | 0.671585 | 0.203409 | 3.30165719 | 0.000961 | 0.00505 | 1.592822 |
| FANCB | 131.1885 | 0.848776 | 0.257145 | 3.300764 | 0.000964 | 0.00506 | 1.800972 |
| ANKRD12 | 675.52586 | 0.761285 | 0.230722 | 3.29957803 | 0.000968 | 0.005078 | 1.694999 |
| RBIS | 658.0744 | 0.587241 | 0.178065 | 3.29789454 | 0.000974 | 0.005103 | 1.50237 |
| AC092329 | 100.7431 | 0.94101 | 0.285625 | 3.29456361 | 0.000986 | 0.005153 | 1.919871 |
| ID2 | 986.01913 | 0.592229 | 0.179821 | 3.29343653 | 0.00099 | 0.005171 | 1.507575 |
| BEX5 | 256.39521 | 0.646345 | 0.196275 | 3.29304969 | 0.000991 | 0.005176 | 1.565198 |
| LYPD3 | 64.25323 | 0.758581 | 0.231062 | 3.28301498 | 0.001027 | 0.005333 | 1.691825 |
| ZNF225 | 109.56887 | 0.738641 | 0.225549 | 3.27485157 | 0.001057 | 0.005468 | 1.668603 |
| AL033529. | 64.759341 | 1.00392 | 0.306931 | 3.27083603 | 0.001072 | 0.00554 | 2.005442 |
| NCOA7 | 429.02607 | 0.652008 | 0.199867 | 3.26221965 | 0.001105 | 0.005686 | 1.571354 |
| KLHL4 | 1232.1525 | 0.684105 | 0.210591 | 3.2484969 | 0.00116 | 0.005914 | 1.606705 |
| BAHCC1 | 67.589656 | 1.406374 | 0.434492 | 3.23682172 | 0.001209 | 0.006131 | 2.650702 |
| STXBP4 | 419.93364 | 0.749512 | 0.231866 | 3.23252282 | 0.001227 | 0.006196 | 1.681224 |
| CLVS2 | 53.276852 | 1.498132 | 0.463852 | 3.22976251 | 0.001239 | 0.006247 | 2.824767 |
| GUCY1B1 | 31.069334 | 1.225929 | 0.380495 | 3.22192957 | 0.001273 | 0.006381 | 2.33906 |
| CYP2B6 | 90.209484 | 0.713431 | 0.221696 | 3.21805331 | 0.001291 | 0.006457 | 1.639699 |
| DCDC1 | 11.31494 | 2.248735 | 0.699729 | 3.21372471 | 0.00131 | 0.006547 | 4.75266 |
| MEGF9 | 68.416982 | 0.783548 | 0.244542 | 3.20414765 | 0.001355 | 0.006711 | 1.721359 |
| OSBPL3 | 422.67304 | 0.654512 | 0.204507 | 3.20044434 | 0.001372 | 0.00678 | 1.574084 |
| F8 | 130.20937 | 0.680001 | 0.212756 | 3.19615484 | 0.001393 | 0.00686 | 1.602141 |
| DHFRP1 | 218.16473 | 0.678039 | 0.212469 | 3.19123758 | 0.001417 | 0.006954 | 1.599963 |
| ZNF143 | 509.43622 | 0.652725 | 0.205546 | 3.1755618 | 0.001495 | 0.007267 | 1.572135 |
| ZNF436 | 57.187337 | 0.842301 | 0.267393 | 3.1500413 | 0.001632 | 0.0078 | 1.792907 |
| HES2 | 19.782544 | 1.329718 | 0.423738 | 3.13806717 | 0.001701 | 0.008084 | 2.513536 |
| EVI5 | 292.17697 | 0.788659 | 0.25168 | 3.13357542 | 0.001727 | 0.008177 | 1.727468 |
| YPEL2 | 133.91361 | 0.612584 | 0.197171 | 3.10687001 | 0.001891 | 0.008778 | 1.528995 |
| PALM2 | 76.030245 | 1.63515 | 0.527317 | 3.10088483 | 0.001929 | 0.008921 | 3.106198 |
| PMF1-BGL | 83.130237 | 0.746572 | 0.240857 | 3.09965115 | 0.001937 | 0.008948 | 1.677802 |
| NOS3 | 190.78668 | 0.778737 | 0.251543 | 3.09583801 | 0.001963 | 0.009039 | 1.715629 |

| | | | | | | | |
|---|---|---|---|---|---|---|---|
| CYB5R4 | 403.89336 | 0.827495 | 0.267922 | 3.08856381 | 0.002011 | 0.009229 | 1.774602 |
| APBA1 | 49.892723 | 1.644885 | 0.53271 | 3.08776709 | 0.002017 | 0.009249 | 3.127229 |
| DCX | 182.53187 | 0.697467 | 0.227035 | 3.072075 | 0.002126 | 0.009677 | 1.621655 |
| FABP7 | 100.72886 | 0.899416 | 0.293416 | 3.06533162 | 0.002174 | 0.009875 | 1.865311 |
| FBXO16 | 115.50917 | 0.743076 | 0.242676 | 3.06200609 | 0.002199 | 0.009971 | 1.673741 |
| SORCS2 | 84.227246 | 0.970025 | 0.317578 | 3.05444715 | 0.002255 | 0.010154 | 1.958875 |
| SHC3 | 183.01363 | 0.832411 | 0.272695 | 3.05252822 | 0.002269 | 0.010212 | 1.780658 |
| HPSE2 | 24.069127 | 1.287461 | 0.424303 | 3.03429329 | 0.002411 | 0.010737 | 2.440981 |
| CRISPLD2 | 105.45441 | 0.657532 | 0.217035 | 3.02961801 | 0.002449 | 0.010868 | 1.577382 |
| ZNF148 | 518.36458 | 0.627687 | 0.207363 | 3.02699245 | 0.00247 | 0.010946 | 1.545086 |
| ARRDC3 | 484.16309 | 0.626968 | 0.208013 | 3.01407821 | 0.002578 | 0.01135 | 1.544317 |
| UNC5C | 39.482359 | 1.586565 | 0.526458 | 3.01366174 | 0.002581 | 0.011363 | 3.003334 |
| KLHL14 | 141.20943 | 0.625571 | 0.207949 | 3.00829911 | 0.002627 | 0.011533 | 1.542822 |
| IRF8 | 14.57129 | 2.012976 | 0.671955 | 2.99569956 | 0.002738 | 0.011963 | 4.036138 |
| ERLEC1P1 | 37.130764 | 1.030855 | 0.344352 | 2.99360555 | 0.002757 | 0.01203 | 2.043235 |
| ZEB2 | 209.74227 | 1.082253 | 0.361727 | 2.99190365 | 0.002772 | 0.012091 | 2.117339 |
| RPL32P29 | 44.189031 | 0.870759 | 0.291915 | 2.98292365 | 0.002855 | 0.012379 | 1.828625 |
| FRG1BP | 89.936168 | 0.737682 | 0.247355 | 2.98228456 | 0.002861 | 0.012399 | 1.667494 |
| BZW1P2 | 292.65626 | 0.756762 | 0.255685 | 2.95973944 | 0.003079 | 0.013237 | 1.689694 |
| FSTL5 | 8.5300281 | 2.186537 | 0.741551 | 2.94860085 | 0.003192 | 0.013595 | 4.552115 |
| APLNR | 23.514534 | 1.146887 | 0.38899 | 2.94836891 | 0.003195 | 0.013601 | 2.214355 |
| RAI1 | 155.56579 | 0.656746 | 0.224269 | 2.92838329 | 0.003407 | 0.014355 | 1.576523 |
| CNTFR | 182.30125 | 0.655171 | 0.224402 | 2.91963459 | 0.003504 | 0.014681 | 1.574803 |
| AC010616 | 3.3329683 | 5.067525 | 1.750709 | 2.89455551 | 0.003797 | 0.015693 | 33.53336 |
| MYT1 | 18.200621 | 2.678343 | 0.925346 | 2.89442307 | 0.003799 | 0.015696 | 6.401202 |
| CDKL2 | 94.539437 | 0.625721 | 0.216401 | 2.89149452 | 0.003834 | 0.015813 | 1.542982 |
| POLH | 181.2969 | 0.640321 | 0.221833 | 2.8864963 | 0.003896 | 0.016005 | 1.558676 |
| ZNF887P | 46.904551 | 0.827264 | 0.28733 | 2.87913842 | 0.003988 | 0.016329 | 1.774318 |
| PYGO1 | 77.520006 | 0.698092 | 0.242489 | 2.87886503 | 0.003991 | 0.016339 | 1.622358 |
| AC027559 | 23.737232 | 1.06426 | 0.369909 | 2.87708514 | 0.004014 | 0.016412 | 2.091097 |
| ESRRG | 47.366801 | 1.051748 | 0.366468 | 2.86995778 | 0.004105 | 0.016707 | 2.073041 |
| LMO3 | 31.321626 | 1.132115 | 0.396692 | 2.85388985 | 0.004319 | 0.01744 | 2.191798 |
| DAB2 | 101.85023 | 0.774239 | 0.271738 | 2.8492136 | 0.004383 | 0.017674 | 1.710287 |
| AC005943 | 99.751218 | 1.070447 | 0.375913 | 2.84759502 | 0.004405 | 0.017751 | 2.100085 |
| ARMC2 | 47.755901 | 0.781022 | 0.275447 | 2.83547567 | 0.004576 | 0.01833 | 1.718348 |
| PKNOX2 | 169.98885 | 0.817528 | 0.288439 | 2.83432339 | 0.004592 | 0.018381 | 1.762384 |
| ANKRD36C | 72.778044 | 0.855039 | 0.302236 | 2.82904849 | 0.004669 | 0.018622 | 1.808808 |
| HCFC2 | 52.026413 | 0.769946 | 0.272696 | 2.82346072 | 0.004751 | 0.01888 | 1.705206 |
| IFIT1 | 178.23915 | 0.63138 | 0.223916 | 2.81972087 | 0.004807 | 0.019048 | 1.549046 |
| CDC42EP5 | 11.267507 | 6.820388 | 2.429025 | 2.8078705 | 0.004987 | 0.019624 | 113.0164 |
| ARHGAP18 | 131.61013 | 0.653233 | 0.233098 | 2.80239446 | 0.005072 | 0.019895 | 1.572688 |
| SEC23IP | 1661.1871 | 0.713522 | 0.255012 | 2.79799166 | 0.005142 | 0.02011 | 1.639802 |
| SAMD5 | 5.7498659 | 3.773612 | 1.352501 | 2.79009889 | 0.005269 | 0.020523 | 13.67635 |
| ANXA1 | 25.680773 | 1.183695 | 0.424446 | 2.78879922 | 0.00529 | 0.020586 | 2.271579 |

| | | | | | | | |
|---|---|---|---|---|---|---|---|
| AC012184 | 73.879285 | 0.728246 | 0.261789 | 2.78180631 | 0.005406 | 0.020904 | 1.656624 |
| L3MBTL3 | 429.37318 | 0.630611 | 0.227758 | 2.76877287 | 0.005627 | 0.02156 | 1.548221 |
| AC079594 | 31.951129 | 0.907264 | 0.327749 | 2.76816203 | 0.005637 | 0.021577 | 1.875485 |
| RPL17-C18 | 170.81044 | 0.71899 | 0.25988 | 2.76661957 | 0.005664 | 0.021645 | 1.646029 |
| ASXL3 | 5.5268102 | 2.382129 | 0.865359 | 2.75276257 | 0.005909 | 0.022416 | 5.213054 |
| ADGRL4 | 50.055675 | 0.783488 | 0.285152 | 2.74761732 | 0.006003 | 0.022714 | 1.721287 |
| NPTX1 | 839.90789 | 0.752668 | 0.275974 | 2.72731237 | 0.006385 | 0.023877 | 1.684906 |
| TBC1D19 | 85.810152 | 0.605026 | 0.222376 | 2.72073964 | 0.006514 | 0.024226 | 1.521007 |
| MESP1 | 3.8201786 | 3.184535 | 1.172591 | 2.71581058 | 0.006611 | 0.024536 | 9.091604 |
| POU3F2 | 7.5026461 | 1.935975 | 0.713663 | 2.71273019 | 0.006673 | 0.024702 | 3.826366 |
| FBLN7 | 31.5619 | 1.114134 | 0.411335 | 2.7085773 | 0.006757 | 0.024956 | 2.16465 |
| AP003108. | 165.97325 | 0.608256 | 0.225147 | 2.7015952 | 0.006901 | 0.025343 | 1.524415 |
| MAOA | 226.9834 | 0.692288 | 0.257404 | 2.68949445 | 0.007156 | 0.026111 | 1.615844 |
| CREBRF | 190.39323 | 0.627785 | 0.233683 | 2.68647724 | 0.007221 | 0.026312 | 1.545191 |
| NDRG1 | 115.76775 | 1.012269 | 0.376857 | 2.68608229 | 0.00723 | 0.026333 | 2.017081 |
| TRIM15 | 4.5760328 | 2.582847 | 0.962384 | 2.68380026 | 0.007279 | 0.026473 | 5.991208 |
| ERV3-1 | 153.94262 | 1.065527 | 0.399334 | 2.66826023 | 0.007625 | 0.027371 | 2.092934 |
| GLDN | 25.992967 | 0.964197 | 0.361488 | 2.6673001 | 0.007646 | 0.027432 | 1.950977 |
| PAX6 | 19.267331 | 2.2983 | 0.866931 | 2.65107649 | 0.008024 | 0.02848 | 4.918777 |
| GAREM1 | 272.23228 | 0.612954 | 0.231484 | 2.64793005 | 0.008099 | 0.028667 | 1.529388 |
| ST8SIA4 | 13.238749 | 1.313211 | 0.501771 | 2.61715179 | 0.008867 | 0.030928 | 2.484939 |
| LRRN3 | 85.366157 | 1.201227 | 0.459581 | 2.61374053 | 0.008956 | 0.031195 | 2.299351 |
| JAKMIP1 | 88.245505 | 0.622713 | 0.238612 | 2.60972956 | 0.009061 | 0.031486 | 1.539768 |
| MICAL2 | 301.72507 | 0.689763 | 0.264359 | 2.60918448 | 0.009076 | 0.031524 | 1.613018 |
| EIF3CL | 43.785728 | 1.735119 | 0.666746 | 2.60237053 | 0.009258 | 0.032067 | 3.32907 |
| CANX | 1452.3638 | 0.909041 | 0.349475 | 2.60116501 | 0.009291 | 0.032173 | 1.877797 |
| TMEM35A | 258.3348 | 0.613552 | 0.237266 | 2.58592673 | 0.009712 | 0.03333 | 1.530022 |
| GREB1L | 1270.7828 | 0.985568 | 0.381293 | 2.5848066 | 0.009743 | 0.033422 | 1.980092 |
| HMGB1P5 | 88.097399 | 0.749876 | 0.292225 | 2.56608645 | 0.010285 | 0.034936 | 1.681648 |
| MED31 | 54.010943 | 0.70508 | 0.274976 | 2.56414633 | 0.010343 | 0.035106 | 1.630235 |
| MAP1LC3C | 54.091965 | 0.619581 | 0.242439 | 2.55561873 | 0.0106 | 0.035815 | 1.536428 |
| ZNF222 | 58.793682 | 0.598861 | 0.235069 | 2.54759269 | 0.010847 | 0.036456 | 1.51452 |
| CD37 | 35.014446 | 1.54426 | 0.606229 | 2.54732241 | 0.010855 | 0.03647 | 2.916545 |
| KIAA1755 | 5.030026 | 2.362589 | 0.927749 | 2.54658312 | 0.010878 | 0.036519 | 5.142925 |
| IP6K3 | 62.3859 | 0.675325 | 0.266037 | 2.53846715 | 0.011134 | 0.037189 | 1.596957 |
| AC010186 | 53.593398 | 0.735105 | 0.289892 | 2.53579295 | 0.011219 | 0.037394 | 1.664519 |
| SLC12A1 | 24.993508 | 1.121368 | 0.442337 | 2.53509838 | 0.011242 | 0.03744 | 2.175532 |
| TACR1 | 37.416554 | 1.426689 | 0.565022 | 2.5250124 | 0.011569 | 0.038346 | 2.68829 |
| ARHGEF28 | 24.049757 | 1.392659 | 0.55283 | 2.51914372 | 0.011764 | 0.038901 | 2.625622 |
| DGKK | 31.692058 | 0.901723 | 0.358206 | 2.51733244 | 0.011825 | 0.039038 | 1.868296 |
| NSUN3 | 224.71387 | 0.616768 | 0.245005 | 2.51736705 | 0.011824 | 0.039038 | 1.533436 |
| LY75-CD30 | 4.1075269 | 5.377125 | 2.149473 | 2.50160171 | 0.012363 | 0.040399 | 41.56004 |
| NKX3-1 | 25.424222 | 0.970418 | 0.388234 | 2.4995678 | 0.012434 | 0.040589 | 1.959408 |
| EPHA3 | 30.582323 | 1.107146 | 0.443092 | 2.49867965 | 0.012466 | 0.040652 | 2.15419 |

| | | | | | | | |
|---|---|---|---|---|---|---|---|
| BX679664. | 24.381244 | 0.968171 | 0.388393 | 2.49276142 | 0.012675 | 0.04118 | 1.956359 |
| AC005832 | 4.1320237 | 5.388876 | 2.168464 | 2.48511218 | 0.012951 | 0.041949 | 41.89995 |
| PLAGL1 | 38.449314 | 0.763525 | 0.307695 | 2.48143463 | 0.013085 | 0.042305 | 1.697633 |
| ZFP2 | 43.490684 | 0.706151 | 0.285172 | 2.47622509 | 0.013278 | 0.042783 | 1.631445 |
| KANK4 | 21.194878 | 1.058431 | 0.427894 | 2.47357987 | 0.013377 | 0.043021 | 2.082665 |
| SPATA31A1 | 3.4013074 | 5.057975 | 2.047003 | 2.47091779 | 0.013477 | 0.043261 | 33.31212 |
| FRMPD3 | 69.105541 | 0.644918 | 0.261711 | 2.46423368 | 0.013731 | 0.043938 | 1.56365 |
| PECAM1 | 11.990685 | 1.986923 | 0.810253 | 2.45222484 | 0.014198 | 0.045163 | 3.963907 |
| ADAMTS6 | 47.560183 | 0.955193 | 0.389841 | 2.45020973 | 0.014277 | 0.045333 | 1.938839 |
| C7orf25 | 51.706056 | 0.71923 | 0.294233 | 2.44442795 | 0.014508 | 0.045897 | 1.646303 |
| NOS2 | 50.601685 | 0.805914 | 0.330906 | 2.4354774 | 0.014872 | 0.046808 | 1.748253 |
| COL3A1 | 18.326185 | 1.392815 | 0.573419 | 2.42896409 | 0.015142 | 0.047549 | 2.625906 |
| LRTOMT | 111.66717 | 0.672806 | 0.27752 | 2.4243495 | 0.015336 | 0.048017 | 1.594171 |
| SLA | 11.637958 | 1.699642 | 0.702102 | 2.42079069 | 0.015487 | 0.048389 | 3.248203 |
| BCL2 | 132.68224 | 0.811251 | 0.336074 | 2.41390367 | 0.015783 | 0.049215 | 1.754732 |

**DOWN - p<.05, FC<.75**

| GeneID | Base mear | log2(FC) | StdErr | Wald-Stats | P-value | P-adj | FC |
|---|---|---|---|---|---|---|---|
| HLA-A | 3620.36 | -5.63173 | 0.137738 | -40.8872 | 0 | 0 | 0.020169 |
| ZNF544 | 718.7831 | -5.37807 | 0.18176 | -29.5889 | 2.08E-192 | 1.79E-188 | 0.024046 |
| ZNF558 | 252.4732 | -7.14167 | 0.310724 | -22.984 | 6.75E-117 | 3.88E-113 | 0.007082 |
| CEBPZOS | 303.3298 | -3.52277 | 0.161619 | -21.7967 | 2.49E-105 | 1.07E-101 | 0.087004 |
| EPHX3 | 215.6943 | -4.18665 | 0.222857 | -18.7862 | 9.79E-79 | 2.81E-75 | 0.054915 |
| PLIN2 | 648.0368 | -1.30446 | 0.082453 | -15.8206 | 2.24E-56 | 5.53E-53 | 0.404874 |
| IAH1 | 135.129 | -4.70788 | 0.336806 | -13.978 | 2.12E-44 | 4.57E-41 | 0.038264 |
| PABPC1L | 1307.018 | -1.32371 | 0.098058 | -13.4993 | 1.58E-41 | 3.03E-38 | 0.399507 |
| GPR143 | 778.5648 | -2.18637 | 0.166654 | -13.1192 | 2.56E-39 | 3.67E-36 | 0.219703 |
| TNFRSF12/ | 1369.388 | -1.09049 | 0.08665 | -12.5849 | 2.56E-36 | 3.15E-33 | 0.469602 |
| GNA14 | 121.7243 | -4.59368 | 0.36916 | -12.4436 | 1.51E-35 | 1.74E-32 | 0.041416 |
| SLC7A3 | 5117.868 | -0.84269 | 0.067755 | -12.4373 | 1.64E-35 | 1.77E-32 | 0.557604 |
| INPP5F | 1560.199 | -1.67488 | 0.13553 | -12.3579 | 4.41E-35 | 4.47E-32 | 0.313193 |
| HLTF | 582.697 | -6.93137 | 0.564565 | -12.2774 | 1.20E-34 | 1.15E-31 | 0.008193 |
| SLC1A5 | 13419.25 | -0.86629 | 0.072463 | -11.9549 | 6.12E-33 | 5.28E-30 | 0.548557 |
| AMT | 1019.806 | -1.12116 | 0.095082 | -11.7914 | 4.32E-32 | 3.39E-29 | 0.459726 |
| ZNF671 | 68.00682 | -4.63899 | 0.405666 | -11.4355 | 2.78E-30 | 2.00E-27 | 0.040135 |
| BCAN | 450.1266 | -1.23125 | 0.108262 | -11.3728 | 5.71E-30 | 3.94E-27 | 0.425949 |
| SLC3A2 | 12073.47 | -0.81477 | 0.071963 | -11.3219 | 1.02E-29 | 6.77E-27 | 0.568501 |
| RBCK1 | 1191.549 | -1.02494 | 0.090632 | -11.3088 | 1.19E-29 | 7.58E-27 | 0.491431 |
| C9orf64 | 167.8904 | -8.48679 | 0.756821 | -11.2137 | 3.49E-29 | 2.07E-26 | 0.002788 |
| ZNF677 | 74.45155 | -5.53643 | 0.499592 | -11.0819 | 1.54E-28 | 8.54E-26 | 0.021546 |
| CCN1 | 1340.683 | -0.94659 | 0.086044 | -11.0013 | 3.77E-28 | 2.03E-25 | 0.518857 |
| BICDL2 | 597.0003 | -1.11594 | 0.102283 | -10.9103 | 1.03E-27 | 5.38E-25 | 0.461391 |
| MMP25 | 1498.752 | -1.06464 | 0.100879 | -10.5536 | 4.89E-26 | 2.28E-23 | 0.478092 |
| CYP2S1 | 7716.21 | -0.82376 | 0.079733 | -10.3315 | 5.07E-25 | 2.19E-22 | 0.564967 |
| AP1M2 | 2120.616 | -0.93216 | 0.090647 | -10.2835 | 8.37E-25 | 3.52E-22 | 0.524072 |
| MYO15B | 350.2267 | -2.04617 | 0.205142 | -9.9744 | 1.97E-23 | 7.39E-21 | 0.242127 |
| CHP2 | 40.68417 | -3.87079 | 0.389903 | -9.92757 | 3.16E-23 | 1.16E-20 | 0.068356 |
| TMBIM4 | 329.7683 | -1.32491 | 0.134385 | -9.85905 | 6.26E-23 | 2.25E-20 | 0.399175 |
| SLC7A5 | 14729.68 | -1.00617 | 0.102857 | -9.78214 | 1.34E-22 | 4.63E-20 | 0.497867 |
| EME2 | 1122.572 | -1.77795 | 0.183478 | -9.69023 | 3.32E-22 | 1.08E-19 | 0.291598 |
| HAPLN3 | 2137.956 | -0.86782 | 0.091722 | -9.46135 | 3.04E-21 | 9.53E-19 | 0.547975 |
| SLC9A1 | 944.3101 | -0.7283 | 0.0775 | -9.39744 | 5.59E-21 | 1.72E-18 | 0.603613 |
| SLC38A5 | 828.1157 | -0.99655 | 0.107954 | -9.23124 | 2.68E-20 | 7.81E-18 | 0.501198 |
| PCDHGB4 | 72.47075 | -2.33212 | 0.253968 | -9.18274 | 4.20E-20 | 1.19E-17 | 0.198592 |
| AP002956. | 822.4305 | -0.74751 | 0.081813 | -9.13687 | 6.43E-20 | 1.79E-17 | 0.595629 |
| SLC12A9 | 1131.89 | -0.83916 | 0.091938 | -9.12737 | 7.02E-20 | 1.92E-17 | 0.558971 |
| CBSL | 459.1768 | -1.59862 | 0.17597 | -9.08461 | 1.04E-19 | 2.76E-17 | 0.330194 |
| C16orf58 | 1334.622 | -0.64627 | 0.071238 | -9.07187 | 1.17E-19 | 2.97E-17 | 0.638932 |
| RARRES2 | 3903.144 | -1.13696 | 0.125791 | -9.03846 | 1.59E-19 | 3.93E-17 | 0.454717 |
| HPDL | 300.6779 | -1.17568 | 0.130372 | -9.01789 | 1.92E-19 | 4.65E-17 | 0.442674 |

| | | | | | | | |
|---|---|---|---|---|---|---|---|
| PCDHA10 | 259.5309 | -2.05984 | 0.229554 | -8.97324 | 2.88E-19 | 6.80E-17 | 0.239842 |
| RBM14 | 1618.44 | -0.61056 | 0.068734 | -8.88295 | 6.51E-19 | 1.50E-16 | 0.654941 |
| EMC7 | 933.5347 | -0.7827 | 0.089459 | -8.7492 | 2.15E-18 | 4.69E-16 | 0.581278 |
| MST1L | 112.0845 | -3.24766 | 0.371964 | -8.73111 | 2.52E-18 | 5.30E-16 | 0.105283 |
| ITM2C | 10638.69 | -0.56718 | 0.06532 | -8.68308 | 3.85E-18 | 7.90E-16 | 0.674934 |
| CDH3 | 4541.168 | -0.741 | 0.08553 | -8.66359 | 4.57E-18 | 9.27E-16 | 0.598325 |
| STC2 | 6591.615 | -0.64169 | 0.074133 | -8.65593 | 4.89E-18 | 9.80E-16 | 0.64096 |
| C19orf33 | 55.44088 | -3.01539 | 0.350788 | -8.59605 | 8.25E-18 | 1.63E-15 | 0.123674 |
| L3MBTL1 | 578.4402 | -1.59382 | 0.186793 | -8.53256 | 1.43E-17 | 2.77E-15 | 0.331293 |
| SPINT1 | 3967.201 | -0.75388 | 0.090544 | -8.32616 | 8.35E-17 | 1.48E-14 | 0.593007 |
| RHBDL1 | 187.5936 | -1.95347 | 0.235274 | -8.30296 | 1.02E-16 | 1.75E-14 | 0.258195 |
| TMEM35B | 325.2351 | -0.9846 | 0.119741 | -8.22271 | 1.99E-16 | 3.33E-14 | 0.505366 |
| FAM189A2 | 409.9253 | -0.99921 | 0.122203 | -8.17662 | 2.92E-16 | 4.75E-14 | 0.500275 |
| PIGO | 1065.878 | -0.65633 | 0.08029 | -8.17448 | 2.97E-16 | 4.79E-14 | 0.634491 |
| COL7A1 | 670.4036 | -2.38688 | 0.292319 | -8.16533 | 3.21E-16 | 5.12E-14 | 0.191195 |
| SLITRK2 | 66.99834 | -8.52894 | 1.051624 | -8.11026 | 5.05E-16 | 7.84E-14 | 0.002707 |
| PCDHGC3 | 506.7373 | -1.73276 | 0.213852 | -8.10264 | 5.38E-16 | 8.28E-14 | 0.300875 |
| VCAN | 8741.174 | -0.50997 | 0.063365 | -8.0482 | 8.40E-16 | 1.24E-13 | 0.702237 |
| CBARP | 641.7049 | -1.02892 | 0.128147 | -8.02923 | 9.81E-16 | 1.40E-13 | 0.490076 |
| KLHDC7A | 1188.655 | -0.81163 | 0.101086 | -8.02915 | 9.81E-16 | 1.40E-13 | 0.569736 |
| PRSS8 | 3718.591 | -0.70286 | 0.087585 | -8.02493 | 1.02E-15 | 1.44E-13 | 0.614353 |
| P2RY11 | 188.1521 | -1.20022 | 0.150203 | -7.99062 | 1.34E-15 | 1.85E-13 | 0.43521 |
| DDRGK1 | 854.8474 | -0.69383 | 0.086842 | -7.98961 | 1.35E-15 | 1.85E-13 | 0.618208 |
| ATXN2L | 6817.888 | -0.59842 | 0.075418 | -7.93471 | 2.11E-15 | 2.75E-13 | 0.660477 |
| ANO9 | 991.501 | -1.90367 | 0.240023 | -7.93118 | 2.17E-15 | 2.81E-13 | 0.267263 |
| GTF2IP13 | 310.094 | -1.08354 | 0.137104 | -7.90303 | 2.72E-15 | 3.45E-13 | 0.47187 |
| LIMD2 | 1337.098 | -0.61648 | 0.078073 | -7.89619 | 2.88E-15 | 3.59E-13 | 0.652261 |
| PPP1R3C | 29.66145 | -4.52445 | 0.578744 | -7.81769 | 5.38E-15 | 6.35E-13 | 0.043452 |
| AC139769 | 41.76336 | -8.22313 | 1.052052 | -7.81628 | 5.44E-15 | 6.38E-13 | 0.003346 |
| THBS4 | 1311.66 | -0.78706 | 0.101909 | -7.72312 | 1.14E-14 | 1.28E-12 | 0.579525 |
| CH25H | 108.4657 | -2.67105 | 0.350095 | -7.62951 | 2.36E-14 | 2.55E-12 | 0.157012 |
| MAMDC4 | 390.8632 | -1.95318 | 0.255994 | -7.62981 | 2.35E-14 | 2.55E-12 | 0.258246 |
| ZNF700 | 27.76974 | -8.35686 | 1.09728 | -7.61598 | 2.62E-14 | 2.78E-12 | 0.00305 |
| TAZ | 764.0762 | -0.78708 | 0.104135 | -7.55824 | 4.09E-14 | 4.24E-12 | 0.579516 |
| FUT1 | 708.2673 | -0.65717 | 0.087474 | -7.51273 | 5.79E-14 | 5.87E-12 | 0.634121 |
| P4HTM | 583.5223 | -0.73739 | 0.098511 | -7.48536 | 7.13E-14 | 7.19E-12 | 0.599822 |
| CYBA | 2606.341 | -1.1171 | 0.149992 | -7.4477 | 9.50E-14 | 9.41E-12 | 0.461021 |
| PCDH1 | 6098.403 | -0.80476 | 0.108214 | -7.43674 | 1.03E-13 | 1.01E-11 | 0.572459 |
| LAMC3 | 556.4564 | -1.22486 | 0.165359 | -7.40726 | 1.29E-13 | 1.26E-11 | 0.427839 |
| AC010507 | 59.70877 | -2.32738 | 0.314266 | -7.40576 | 1.30E-13 | 1.26E-11 | 0.199246 |
| SYTL1 | 480.1374 | -1.0629 | 0.143931 | -7.38482 | 1.53E-13 | 1.44E-11 | 0.478667 |
| PLPP2 | 781.9249 | -0.71443 | 0.096799 | -7.38059 | 1.58E-13 | 1.48E-11 | 0.609445 |
| GTF2IP20 | 344.3667 | -1.14859 | 0.155974 | -7.36399 | 1.78E-13 | 1.65E-11 | 0.451066 |
| AMH | 485.9003 | -2.21033 | 0.30177 | -7.32457 | 2.40E-13 | 2.19E-11 | 0.216084 |

| | | | | | | | |
|---|---|---|---|---|---|---|---|
| CCN2 | 1344.6 | -0.71125 | 0.097218 | -7.31612 | 2.55E-13 | 2.32E-11 | 0.610789 |
| GRK2 | 2023.578 | -0.57647 | 0.078888 | -7.30738 | 2.72E-13 | 2.43E-11 | 0.670605 |
| GRM4 | 1170.744 | -0.61311 | 0.083953 | -7.30309 | 2.81E-13 | 2.49E-11 | 0.653784 |
| DOCK2 | 41.39779 | -7.14 | 0.978836 | -7.29438 | 3.00E-13 | 2.63E-11 | 0.00709 |
| NFKB2 | 1658.738 | -0.76352 | 0.105135 | -7.2623 | 3.81E-13 | 3.22E-11 | 0.589057 |
| IL27RA | 676.777 | -0.71359 | 0.09867 | -7.23212 | 4.76E-13 | 3.98E-11 | 0.609799 |
| LRP1 | 10105.83 | -0.71614 | 0.09902 | -7.23229 | 4.75E-13 | 3.98E-11 | 0.608722 |
| STRA6 | 380.5426 | -0.89188 | 0.123509 | -7.22117 | 5.15E-13 | 4.29E-11 | 0.53891 |
| PROCA1 | 401.7755 | -0.96285 | 0.133787 | -7.1969 | 6.16E-13 | 5.08E-11 | 0.513042 |
| AL732372. | 1023.4 | -0.66396 | 0.092446 | -7.18217 | 6.86E-13 | 5.58E-11 | 0.631143 |
| RCOR2 | 2889.739 | -0.52603 | 0.073324 | -7.17403 | 7.28E-13 | 5.89E-11 | 0.694462 |
| APBB3 | 331.2714 | -1.24258 | 0.173263 | -7.17165 | 7.41E-13 | 5.97E-11 | 0.422617 |
| LSR | 4432.377 | -0.5844 | 0.081571 | -7.16431 | 7.82E-13 | 6.27E-11 | 0.666927 |
| BCL11B | 54.22784 | -1.82924 | 0.255432 | -7.16136 | 7.99E-13 | 6.34E-11 | 0.281413 |
| TIMP4 | 157.7513 | -1.28143 | 0.178935 | -7.16139 | 7.99E-13 | 6.34E-11 | 0.411389 |
| ZNF512B | 1511.84 | -0.62595 | 0.087665 | -7.14017 | 9.32E-13 | 7.37E-11 | 0.647995 |
| PILRB | 1410.585 | -1.37673 | 0.193139 | -7.12818 | 1.02E-12 | 7.97E-11 | 0.385091 |
| PCSK4 | 239.6412 | -1.4454 | 0.203552 | -7.1009 | 1.24E-12 | 9.54E-11 | 0.367189 |
| COL23A1 | 341.5833 | -0.77637 | 0.109507 | -7.08968 | 1.34E-12 | 1.03E-10 | 0.583834 |
| FAM241A | 1522.722 | -0.69232 | 0.097687 | -7.08713 | 1.37E-12 | 1.04E-10 | 0.618859 |
| AL669831. | 2524.51 | -1.04339 | 0.14761 | -7.06857 | 1.57E-12 | 1.18E-10 | 0.485186 |
| NEDD4 | 988.1871 | -0.56849 | 0.080493 | -7.06258 | 1.63E-12 | 1.22E-10 | 0.674324 |
| COL6A2 | 2528.398 | -0.75948 | 0.107551 | -7.06158 | 1.65E-12 | 1.22E-10 | 0.59071 |
| YJEFN3 | 503.1137 | -1.82386 | 0.25876 | -7.04846 | 1.81E-12 | 1.34E-10 | 0.282464 |
| NPIPB11 | 165.8975 | -1.50825 | 0.214068 | -7.04566 | 1.85E-12 | 1.36E-10 | 0.351537 |
| LRRC61 | 534.0667 | -0.73309 | 0.104356 | -7.02489 | 2.14E-12 | 1.56E-10 | 0.601613 |
| ZNF276 | 309.0067 | -1.15169 | 0.16399 | -7.02295 | 2.17E-12 | 1.57E-10 | 0.450096 |
| ZNF665 | 120.5496 | -2.36093 | 0.336346 | -7.01936 | 2.23E-12 | 1.60E-10 | 0.194665 |
| ATP13A1 | 2024.394 | -0.9964 | 0.143134 | -6.96131 | 3.37E-12 | 2.35E-10 | 0.501249 |
| GPC4 | 15692.02 | -0.55084 | 0.079125 | -6.96171 | 3.36E-12 | 2.35E-10 | 0.682621 |
| ACE | 150.5315 | -1.49965 | 0.215553 | -6.95721 | 3.47E-12 | 2.41E-10 | 0.35364 |
| FMNL1 | 372.6612 | -0.71172 | 0.102689 | -6.93083 | 4.18E-12 | 2.87E-10 | 0.610592 |
| INO80E | 1406.983 | -0.44244 | 0.06407 | -6.90564 | 5.00E-12 | 3.40E-10 | 0.735888 |
| CDX4 | 31.34518 | -2.64244 | 0.383944 | -6.88237 | 5.89E-12 | 3.92E-10 | 0.160157 |
| SLC6A9 | 1113.49 | -0.74019 | 0.107611 | -6.87841 | 6.05E-12 | 4.00E-10 | 0.598659 |
| TNFSF11 | 242.807 | -0.84144 | 0.122398 | -6.87459 | 6.22E-12 | 4.09E-10 | 0.558088 |
| HSPG2 | 5361.828 | -1.21069 | 0.176214 | -6.87055 | 6.40E-12 | 4.18E-10 | 0.432063 |
| CD74 | 1357.957 | -0.76717 | 0.111716 | -6.86717 | 6.55E-12 | 4.26E-10 | 0.587568 |
| VEGFA | 3295.482 | -0.5809 | 0.084687 | -6.85943 | 6.91E-12 | 4.45E-10 | 0.668545 |
| MORC4 | 615.6941 | -0.88795 | 0.13003 | -6.82882 | 8.56E-12 | 5.37E-10 | 0.540382 |
| FAM95C | 339.2086 | -1.29354 | 0.189464 | -6.82739 | 8.65E-12 | 5.40E-10 | 0.407948 |
| WDR90 | 1358.614 | -1.61018 | 0.235883 | -6.82621 | 8.72E-12 | 5.42E-10 | 0.327556 |
| INHBE | 2332.451 | -1.11687 | 0.163922 | -6.81341 | 9.53E-12 | 5.87E-10 | 0.461093 |
| PRR22 | 50.78829 | -2.7051 | 0.397328 | -6.80823 | 9.88E-12 | 6.04E-10 | 0.15335 |

| | | | | | | | |
|---|---|---|---|---|---|---|---|
| ARHGEF1 | 1318.435 | -0.6635 | 0.097498 | -6.80525 | 1.01E-11 | 6.14E-10 | 0.631345 |
| GAL | 933.6753 | -0.70715 | 0.104023 | -6.798 | 1.06E-11 | 6.44E-10 | 0.61253 |
| NPEPL1 | 1349.981 | -1.14385 | 0.168391 | -6.79283 | 1.10E-11 | 6.63E-10 | 0.452549 |
| SLC2A8 | 519.2703 | -0.81198 | 0.119913 | -6.77142 | 1.28E-11 | 7.53E-10 | 0.5696 |
| KMT5A | 867.6487 | -0.73601 | 0.108768 | -6.76684 | 1.32E-11 | 7.74E-10 | 0.600397 |
| FLNA | 55640.95 | -1.19723 | 0.177092 | -6.76051 | 1.38E-11 | 8.04E-10 | 0.436111 |
| COL27A1 | 942.2054 | -1.03359 | 0.153251 | -6.74444 | 1.54E-11 | 8.94E-10 | 0.488492 |
| L1CAM | 1304.075 | -0.53247 | 0.078955 | -6.74403 | 1.54E-11 | 8.94E-10 | 0.691368 |
| NAT8L | 1339.93 | -0.63353 | 0.09398 | -6.74115 | 1.57E-11 | 9.09E-10 | 0.644596 |
| IFRD2 | 1279.796 | -0.51222 | 0.076029 | -6.7371 | 1.62E-11 | 9.31E-10 | 0.701145 |
| SYP | 362.6258 | -0.9657 | 0.143528 | -6.72835 | 1.72E-11 | 9.86E-10 | 0.512028 |
| RPS9 | 11273.51 | -1.53517 | 0.228397 | -6.72151 | 1.80E-11 | 1.03E-09 | 0.345039 |
| PHETA2 | 28.44049 | -4.34345 | 0.646468 | -6.71874 | 1.83E-11 | 1.04E-09 | 0.04926 |
| USP32P1 | 283.3364 | -1.28652 | 0.191997 | -6.70071 | 2.07E-11 | 1.16E-09 | 0.40994 |
| LTB4R | 558.465 | -1.73319 | 0.258822 | -6.69646 | 2.14E-11 | 1.19E-09 | 0.300786 |
| TAC1 | 205.537 | -1.25484 | 0.187577 | -6.68975 | 2.24E-11 | 1.23E-09 | 0.419039 |
| AC005726 | 369.8807 | -1.30201 | 0.194729 | -6.68627 | 2.29E-11 | 1.25E-09 | 0.40556 |
| SLC15A3 | 506.8123 | -0.78868 | 0.118246 | -6.66978 | 2.56E-11 | 1.38E-09 | 0.578875 |
| PKP3 | 272.6898 | -1.32458 | 0.198739 | -6.66494 | 2.65E-11 | 1.42E-09 | 0.399265 |
| SEMA6A | 11094.63 | -0.44087 | 0.066148 | -6.6649 | 2.65E-11 | 1.42E-09 | 0.736689 |
| PHF21B | 814.4226 | -0.80652 | 0.121126 | -6.65855 | 2.77E-11 | 1.47E-09 | 0.571758 |
| PIEZO1 | 2884.345 | -1.49091 | 0.22446 | -6.64223 | 3.09E-11 | 1.62E-09 | 0.355788 |
| PPP1R16A | 646.6675 | -1.31223 | 0.197564 | -6.64206 | 3.09E-11 | 1.62E-09 | 0.402697 |
| PIGQ | 1259.773 | -1.20313 | 0.181234 | -6.6385 | 3.17E-11 | 1.66E-09 | 0.434333 |
| SLC9A5 | 377.5507 | -1.17593 | 0.177206 | -6.63599 | 3.22E-11 | 1.68E-09 | 0.442597 |
| SEMA4B | 1840.356 | -0.60675 | 0.091532 | -6.62882 | 3.38E-11 | 1.75E-09 | 0.656674 |
| SEC14L4 | 36.7939 | -2.67736 | 0.403974 | -6.62756 | 3.41E-11 | 1.76E-09 | 0.156327 |
| TIMM44 | 2890.911 | -0.45273 | 0.068417 | -6.6173 | 3.66E-11 | 1.88E-09 | 0.730656 |
| LAT | 101.4264 | -2.13317 | 0.322415 | -6.61622 | 3.69E-11 | 1.88E-09 | 0.227957 |
| ZNF44 | 20.9285 | -4.84135 | 0.731713 | -6.61647 | 3.68E-11 | 1.88E-09 | 0.034883 |
| SLC52A2 | 1430.363 | -0.77716 | 0.117825 | -6.59593 | 4.23E-11 | 2.15E-09 | 0.583513 |
| SLC2A4RG | 912.481 | -0.89301 | 0.135488 | -6.5911 | 4.37E-11 | 2.21E-09 | 0.538488 |
| AC010507 | 43.61837 | -3.26967 | 0.496634 | -6.58366 | 4.59E-11 | 2.31E-09 | 0.103688 |
| COL6A3 | 287.8704 | -1.0878 | 0.165269 | -6.58199 | 4.64E-11 | 2.33E-09 | 0.470479 |
| SIGMAR1 | 2328.864 | -0.5199 | 0.079001 | -6.5809 | 4.68E-11 | 2.33E-09 | 0.697423 |
| WDR74 | 1401.262 | -0.67548 | 0.102644 | -6.58081 | 4.68E-11 | 2.33E-09 | 0.626125 |
| MTRNR2L8 | 34776.15 | -1.05067 | 0.160073 | -6.56366 | 5.25E-11 | 2.58E-09 | 0.482745 |
| TXNRD2 | 459.9631 | -0.70029 | 0.106881 | -6.55207 | 5.67E-11 | 2.77E-09 | 0.615447 |
| UBR4 | 12056.02 | -0.77081 | 0.11765 | -6.55171 | 5.69E-11 | 2.77E-09 | 0.586088 |
| NPIPB3 | 1530.884 | -1.51978 | 0.232199 | -6.54515 | 5.94E-11 | 2.88E-09 | 0.348739 |
| MYBBP1A | 2990.437 | -0.62463 | 0.095727 | -6.52511 | 6.79E-11 | 3.26E-09 | 0.648585 |
| MAN2C1 | 1393.702 | -0.97164 | 0.14915 | -6.51453 | 7.29E-11 | 3.44E-09 | 0.509925 |
| CCDC84 | 366.8485 | -0.8803 | 0.135286 | -6.50694 | 7.67E-11 | 3.60E-09 | 0.543255 |
| FXYD5 | 1790.666 | -0.54002 | 0.083191 | -6.49133 | 8.51E-11 | 3.95E-09 | 0.687762 |

| | | | | | | | |
|---|---|---|---|---|---|---|---|
| RTEL1-TNF | 354.8797 | -1.14916 | 0.177017 | -6.49183 | 8.48E-11 | 3.95E-09 | 0.450887 |
| COL18A1 | 10294.76 | -0.95239 | 0.147223 | -6.469 | 9.87E-11 | 4.52E-09 | 0.516776 |
| ITGA3 | 1025.69 | -0.78218 | 0.121006 | -6.46395 | 1.02E-10 | 4.65E-09 | 0.581488 |
| SPINK5 | 236.6655 | -1.83822 | 0.284901 | -6.45212 | 1.10E-10 | 5.00E-09 | 0.279667 |
| SPPL2B | 1798.484 | -0.88285 | 0.136919 | -6.44801 | 1.13E-10 | 5.11E-09 | 0.542293 |
| BCAM | 2619.424 | -0.92649 | 0.143781 | -6.44376 | 1.17E-10 | 5.22E-09 | 0.526136 |
| WASH6P | 936.3782 | -0.91963 | 0.142757 | -6.44194 | 1.18E-10 | 5.27E-09 | 0.528644 |
| ZDHHC4 | 789.0146 | -0.88903 | 0.138076 | -6.43869 | 1.21E-10 | 5.37E-09 | 0.539977 |
| SCT | 13.59494 | -7.32494 | 1.139403 | -6.42875 | 1.29E-10 | 5.70E-09 | 0.006237 |
| AL133216. | 223.6782 | -0.93547 | 0.145726 | -6.41937 | 1.37E-10 | 6.05E-09 | 0.522873 |
| ABCB6 | 987.7254 | -1.0673 | 0.166602 | -6.40631 | 1.49E-10 | 6.52E-09 | 0.47721 |
| PGGHG | 2994.114 | -1.36353 | 0.213101 | -6.39851 | 1.57E-10 | 6.81E-09 | 0.388632 |
| NKAP | 921.0641 | -0.66691 | 0.104444 | -6.3854 | 1.71E-10 | 7.35E-09 | 0.629853 |
| SUMF2 | 2520.305 | -0.64607 | 0.101272 | -6.37955 | 1.78E-10 | 7.60E-09 | 0.639019 |
| SCNN1A | 3129.029 | -0.51757 | 0.081162 | -6.37699 | 1.81E-10 | 7.70E-09 | 0.698547 |
| GPR108 | 1044.422 | -0.78514 | 0.123249 | -6.37031 | 1.89E-10 | 7.93E-09 | 0.580297 |
| TSPAN7 | 447.1135 | -0.77081 | 0.120999 | -6.37038 | 1.89E-10 | 7.93E-09 | 0.58609 |
| AL732372. | 356.1759 | -1.2824 | 0.201567 | -6.36213 | 1.99E-10 | 8.30E-09 | 0.411112 |
| PCK2 | 7518.775 | -0.61186 | 0.096171 | -6.36224 | 1.99E-10 | 8.30E-09 | 0.654352 |
| FLNB | 8955.609 | -1.19811 | 0.188432 | -6.35829 | 2.04E-10 | 8.49E-09 | 0.435847 |
| HAGHL | 469.1174 | -1.06709 | 0.167973 | -6.35275 | 2.11E-10 | 8.78E-09 | 0.477281 |
| CHPF | 1699.899 | -0.64957 | 0.102257 | -6.35233 | 2.12E-10 | 8.79E-09 | 0.63747 |
| FASTK | 1228.149 | -1.02296 | 0.16106 | -6.35143 | 2.13E-10 | 8.82E-09 | 0.492106 |
| FLT4 | 362.507 | -1.13672 | 0.179117 | -6.34625 | 2.21E-10 | 9.10E-09 | 0.454793 |
| NME3 | 1028.6 | -0.9343 | 0.147581 | -6.33076 | 2.44E-10 | 9.96E-09 | 0.523296 |
| DUS3L | 519.2659 | -1.05641 | 0.168005 | -6.288 | 3.22E-10 | 1.27E-08 | 0.480826 |
| LARGE2 | 2871.328 | -0.67376 | 0.10721 | -6.28453 | 3.29E-10 | 1.30E-08 | 0.626869 |
| CC2D1A | 892.2724 | -0.75592 | 0.120383 | -6.27932 | 3.40E-10 | 1.34E-08 | 0.592168 |
| SLC25A43 | 284.2136 | -1.06943 | 0.17062 | -6.26789 | 3.66E-10 | 1.43E-08 | 0.476509 |
| KREMEN2 | 630.5875 | -1.28094 | 0.204553 | -6.26216 | 3.80E-10 | 1.48E-08 | 0.411526 |
| COL9A3 | 1812.743 | -1.54058 | 0.246148 | -6.25875 | 3.88E-10 | 1.51E-08 | 0.343747 |
| MTCO1P12 | 1333.433 | -1.40903 | 0.225987 | -6.23503 | 4.52E-10 | 1.75E-08 | 0.376564 |
| AC139256 | 219.6269 | -0.92867 | 0.148959 | -6.23444 | 4.53E-10 | 1.75E-08 | 0.525341 |
| BNC1 | 98.32577 | -2.53473 | 0.407127 | -6.22588 | 4.79E-10 | 1.83E-08 | 0.172573 |
| PRSS12 | 208.7508 | -0.8747 | 0.140615 | -6.22057 | 4.95E-10 | 1.89E-08 | 0.545365 |
| PKD1 | 3106.045 | -1.2725 | 0.204635 | -6.2184 | 5.02E-10 | 1.92E-08 | 0.413942 |
| KIAA0319L | 2975.884 | -0.43628 | 0.070185 | -6.21613 | 5.10E-10 | 1.93E-08 | 0.739037 |
| RHBDD3 | 1121.295 | -0.91586 | 0.147444 | -6.21157 | 5.25E-10 | 1.98E-08 | 0.530028 |
| PRKCG | 256.2817 | -0.84057 | 0.135424 | -6.20695 | 5.40E-10 | 2.03E-08 | 0.558423 |
| FSIP2 | 460.7695 | -0.80871 | 0.130464 | -6.19871 | 5.69E-10 | 2.13E-08 | 0.570894 |
| SEMA3B | 385.8427 | -1.41139 | 0.22771 | -6.19816 | 5.71E-10 | 2.13E-08 | 0.37595 |
| PKMYT1 | 1312.485 | -0.95311 | 0.154102 | -6.18492 | 6.21E-10 | 2.30E-08 | 0.516519 |
| GABBR1 | 1548.122 | -0.64289 | 0.103994 | -6.18199 | 6.33E-10 | 2.34E-08 | 0.640429 |
| NPIPB12 | 1397.667 | -1.36833 | 0.222388 | -6.15287 | 7.61E-10 | 2.78E-08 | 0.387341 |

| | | | | | | | |
|---|---|---|---|---|---|---|---|
| SPATA20 | 1156.832 | -0.54068 | 0.087933 | -6.14872 | 7.81E-10 | 2.85E-08 | 0.687448 |
| ADGRB2 | 1133.671 | -0.93055 | 0.151493 | -6.14256 | 8.12E-10 | 2.96E-08 | 0.524657 |
| WASH5P | 1597.682 | -0.59079 | 0.096254 | -6.13788 | 8.36E-10 | 3.03E-08 | 0.663978 |
| LAMB2 | 3230.254 | -1.31658 | 0.21508 | -6.12137 | 9.28E-10 | 3.34E-08 | 0.401484 |
| MAP3K12 | 684.8327 | -1.14896 | 0.187756 | -6.11939 | 9.39E-10 | 3.36E-08 | 0.450952 |
| RFNG | 895.3944 | -0.84381 | 0.138323 | -6.10026 | 1.06E-09 | 3.75E-08 | 0.557171 |
| SLC44A2 | 5509.217 | -0.5263 | 0.086308 | -6.09799 | 1.07E-09 | 3.79E-08 | 0.694332 |
| ACADVL | 4090.842 | -1.19538 | 0.196248 | -6.09116 | 1.12E-09 | 3.93E-08 | 0.436672 |
| MTRNR2L1 | 8669.879 | -1.19153 | 0.195619 | -6.09106 | 1.12E-09 | 3.93E-08 | 0.437839 |
| MIB2 | 1207.07 | -1.26859 | 0.208314 | -6.08981 | 1.13E-09 | 3.95E-08 | 0.415064 |
| COL16A1 | 527.892 | -1.65044 | 0.271052 | -6.08901 | 1.14E-09 | 3.96E-08 | 0.318543 |
| FAM157A | 1022.94 | -1.05498 | 0.173516 | -6.08004 | 1.20E-09 | 4.18E-08 | 0.481302 |
| FKBP10 | 6283.175 | -0.80199 | 0.132294 | -6.06215 | 1.34E-09 | 4.64E-08 | 0.573559 |
| EPHA1 | 3979.449 | -0.61007 | 0.100816 | -6.05139 | 1.44E-09 | 4.92E-08 | 0.655163 |
| UNC5B | 2273.878 | -0.60386 | 0.100126 | -6.03098 | 1.63E-09 | 5.53E-08 | 0.657992 |
| ZNF154 | 437.742 | -0.8488 | 0.140852 | -6.02615 | 1.68E-09 | 5.63E-08 | 0.555247 |
| KCNG1 | 664.1642 | -0.59952 | 0.099529 | -6.02354 | 1.71E-09 | 5.70E-08 | 0.659974 |
| ITGB5 | 3635.945 | -0.68261 | 0.113494 | -6.01454 | 1.80E-09 | 5.97E-08 | 0.623036 |
| EPHA2 | 920.3524 | -0.62937 | 0.104653 | -6.01388 | 1.81E-09 | 5.98E-08 | 0.646459 |
| FBLL1 | 291.5367 | -0.87886 | 0.146194 | -6.01159 | 1.84E-09 | 6.05E-08 | 0.543798 |
| RCN3 | 136.9291 | -2.00362 | 0.33377 | -6.003 | 1.94E-09 | 6.36E-08 | 0.249374 |
| SLC16A2 | 3121.051 | -0.42004 | 0.069987 | -6.00179 | 1.95E-09 | 6.39E-08 | 0.747402 |
| CEP170B | 1719.805 | -0.53505 | 0.089197 | -5.99853 | 1.99E-09 | 6.51E-08 | 0.690135 |
| ZNF296 | 192.6354 | -0.78612 | 0.13106 | -5.99816 | 2.00E-09 | 6.51E-08 | 0.579902 |
| CHGA | 1963.725 | -0.86388 | 0.144047 | -5.99721 | 2.01E-09 | 6.54E-08 | 0.549472 |
| ITGA7 | 2932.356 | -1.32002 | 0.220178 | -5.99525 | 2.03E-09 | 6.58E-08 | 0.400528 |
| PLD3 | 6713.723 | -0.56854 | 0.09483 | -5.99532 | 2.03E-09 | 6.58E-08 | 0.674301 |
| ZNF132 | 85.284 | -1.23588 | 0.206155 | -5.99492 | 2.04E-09 | 6.58E-08 | 0.424583 |
| CYB5R2 | 442.0346 | -0.67034 | 0.111878 | -5.99171 | 2.08E-09 | 6.70E-08 | 0.628358 |
| SULT4A1 | 409.9936 | -0.69853 | 0.116631 | -5.98925 | 2.11E-09 | 6.79E-08 | 0.616199 |
| DLG5 | 1358.467 | -0.97185 | 0.162368 | -5.98547 | 2.16E-09 | 6.91E-08 | 0.509851 |
| NPIPP1 | 461.3475 | -1.14812 | 0.191809 | -5.98576 | 2.15E-09 | 6.91E-08 | 0.451213 |
| AC007326 | 3567.685 | -0.88401 | 0.147795 | -5.9813 | 2.21E-09 | 7.04E-08 | 0.54186 |
| SSBP4 | 655.0279 | -0.71357 | 0.119505 | -5.97101 | 2.36E-09 | 7.46E-08 | 0.60981 |
| LINC0114! | 282.8421 | -0.98966 | 0.165883 | -5.966 | 2.43E-09 | 7.67E-08 | 0.503598 |
| ARHGAP9 | 120.1524 | -1.54445 | 0.258894 | -5.96556 | 2.44E-09 | 7.68E-08 | 0.342827 |
| ADAP1 | 189.3056 | -0.92113 | 0.154421 | -5.96503 | 2.45E-09 | 7.69E-08 | 0.528097 |
| EPDR1 | 359.2474 | -0.64352 | 0.107946 | -5.9615 | 2.50E-09 | 7.85E-08 | 0.640148 |
| G6PC3 | 2847.136 | -0.65768 | 0.110456 | -5.95422 | 2.61E-09 | 8.17E-08 | 0.633898 |
| TRMT61A | 461.3501 | -0.5544 | 0.09312 | -5.95355 | 2.62E-09 | 8.19E-08 | 0.680942 |
| SLC2A10 | 521.3891 | -0.6612 | 0.111272 | -5.9422 | 2.81E-09 | 8.73E-08 | 0.632352 |
| EPHX1 | 1202.513 | -0.57739 | 0.097174 | -5.94183 | 2.82E-09 | 8.74E-08 | 0.670175 |
| NOMO2 | 2778.534 | -0.7083 | 0.119354 | -5.9344 | 2.95E-09 | 9.07E-08 | 0.612042 |
| SLC29A1 | 5553.903 | -0.50536 | 0.085164 | -5.93393 | 2.96E-09 | 9.07E-08 | 0.704487 |

| | | | | | | | |
|---|---|---|---|---|---|---|---|
| TMEM205 | 1036.523 | -0.66383 | 0.111855 | -5.93477 | 2.94E-09 | 9.07E-08 | 0.6312 |
| WDR76 | 345.7952 | -0.79413 | 0.133872 | -5.93204 | 2.99E-09 | 9.14E-08 | 0.576689 |
| E2F1 | 978.3687 | -0.45172 | 0.076186 | -5.92914 | 3.05E-09 | 9.26E-08 | 0.731172 |
| AC073111 | 602.1781 | -0.61087 | 0.103076 | -5.9264 | 3.10E-09 | 9.38E-08 | 0.654802 |
| PCSK1N | 157.0977 | -1.244 | 0.210039 | -5.92269 | 3.17E-09 | 9.58E-08 | 0.422201 |
| NECTIN2 | 4911.707 | -0.83616 | 0.141253 | -5.91956 | 3.23E-09 | 9.74E-08 | 0.560134 |
| PCDHGA1( | 104.3702 | -1.17238 | 0.198509 | -5.90592 | 3.51E-09 | 1.05E-07 | 0.443689 |
| RGL3 | 1031.38 | -1.19427 | 0.202211 | -5.90605 | 3.50E-09 | 1.05E-07 | 0.437007 |
| AC003112 | 198.6496 | -1.07877 | 0.182704 | -5.90443 | 3.54E-09 | 1.06E-07 | 0.473434 |
| PLOD3 | 1345.624 | -0.77135 | 0.130956 | -5.89019 | 3.86E-09 | 1.14E-07 | 0.585867 |
| CPNE7 | 467.6622 | -1.43396 | 0.243495 | -5.88906 | 3.88E-09 | 1.15E-07 | 0.370115 |
| CRACR2B | 359.2272 | -0.83487 | 0.141898 | -5.88361 | 4.01E-09 | 1.18E-07 | 0.560633 |
| ECEL1P2 | 345.9342 | -1.10995 | 0.188651 | -5.88361 | 4.01E-09 | 1.18E-07 | 0.463311 |
| FURIN | 1397.917 | -0.44996 | 0.076509 | -5.8812 | 4.07E-09 | 1.20E-07 | 0.732061 |
| AC080038 | 1461.742 | -0.91272 | 0.155364 | -5.87475 | 4.23E-09 | 1.24E-07 | 0.531181 |
| ECI1 | 1859.29 | -0.44576 | 0.075948 | -5.86935 | 4.38E-09 | 1.28E-07 | 0.734196 |
| CLDN7 | 1799.575 | -0.71569 | 0.122077 | -5.86259 | 4.56E-09 | 1.33E-07 | 0.608914 |
| OBSL1 | 3649.38 | -0.68735 | 0.117294 | -5.86006 | 4.63E-09 | 1.34E-07 | 0.620994 |
| CCDC88B | 605.4055 | -1.32598 | 0.226434 | -5.85592 | 4.74E-09 | 1.38E-07 | 0.398879 |
| FBXW5 | 1370.859 | -0.42741 | 0.072992 | -5.85559 | 4.75E-09 | 1.38E-07 | 0.743596 |
| ADCK2 | 432.511 | -0.64713 | 0.110581 | -5.85211 | 4.85E-09 | 1.40E-07 | 0.638549 |
| PVR | 749.184 | -0.72361 | 0.123736 | -5.84802 | 4.97E-09 | 1.44E-07 | 0.605581 |
| CLDN1 | 238.8731 | -0.86413 | 0.14792 | -5.84189 | 5.16E-09 | 1.49E-07 | 0.549377 |
| TMEM200E | 285.7455 | -0.80651 | 0.138087 | -5.84056 | 5.20E-09 | 1.50E-07 | 0.571765 |
| DENND4B | 2309.774 | -0.82489 | 0.141334 | -5.83647 | 5.33E-09 | 1.53E-07 | 0.564524 |
| SCARB1 | 1035.994 | -0.50963 | 0.087348 | -5.83446 | 5.40E-09 | 1.54E-07 | 0.702404 |
| PCNX3 | 1625.345 | -0.81058 | 0.139196 | -5.82326 | 5.77E-09 | 1.64E-07 | 0.570154 |
| RHPN1 | 914.0313 | -1.56604 | 0.269036 | -5.82093 | 5.85E-09 | 1.66E-07 | 0.337734 |
| SCARF2 | 156.0511 | -0.94782 | 0.163176 | -5.80854 | 6.30E-09 | 1.77E-07 | 0.518417 |
| P2RX4 | 324.099 | -0.89009 | 0.153267 | -5.80748 | 6.34E-09 | 1.77E-07 | 0.539579 |
| CCDC78 | 249.9897 | -1.72462 | 0.297151 | -5.80383 | 6.48E-09 | 1.80E-07 | 0.302579 |
| TNFRSF8 | 540.2097 | -0.63831 | 0.110018 | -5.80184 | 6.56E-09 | 1.82E-07 | 0.642466 |
| INSYN1 | 198.4468 | -0.81515 | 0.140833 | -5.78803 | 7.12E-09 | 1.96E-07 | 0.56835 |
| TRPM4 | 341.5365 | -0.86885 | 0.150123 | -5.78759 | 7.14E-09 | 1.96E-07 | 0.547583 |
| TLE2 | 270.5727 | -0.92447 | 0.159743 | -5.78722 | 7.16E-09 | 1.96E-07 | 0.526875 |
| ACHE | 95.02029 | -1.28374 | 0.221885 | -5.78558 | 7.23E-09 | 1.98E-07 | 0.410731 |
| RPS6KL1 | 365.3459 | -1.0503 | 0.181744 | -5.77898 | 7.52E-09 | 2.05E-07 | 0.482869 |
| PARP15 | 401.8909 | -0.65468 | 0.113312 | -5.77766 | 7.57E-09 | 2.06E-07 | 0.635218 |
| EIF4HP2 | 22.23837 | -2.54283 | 0.440403 | -5.77387 | 7.75E-09 | 2.10E-07 | 0.171606 |
| TMEM114 | 50.37084 | -1.81136 | 0.313734 | -5.77356 | 7.76E-09 | 2.10E-07 | 0.284922 |
| RHBDF1 | 794.8669 | -1.00424 | 0.174026 | -5.77065 | 7.90E-09 | 2.13E-07 | 0.498532 |
| TPCN1 | 1011.809 | -0.79239 | 0.13744 | -5.76538 | 8.15E-09 | 2.19E-07 | 0.577386 |
| SLC43A1 | 1387.513 | -0.72745 | 0.12622 | -5.76336 | 8.25E-09 | 2.21E-07 | 0.603969 |
| CHST11 | 469.2323 | -0.96206 | 0.166936 | -5.76301 | 8.26E-09 | 2.21E-07 | 0.513325 |

| | | | | | | | |
|---|---|---|---|---|---|---|---|
| SLC22A17 | 1295.618 | -0.88317 | 0.153514 | -5.75302 | 8.77E-09 | 2.34E-07 | 0.542174 |
| MUTYH | 1060.361 | -0.60458 | 0.105321 | -5.74041 | 9.44E-09 | 2.50E-07 | 0.657661 |
| SERPINH1 | 12886.65 | -0.54003 | 0.094114 | -5.73799 | 9.58E-09 | 2.54E-07 | 0.687758 |
| LAMA5 | 9816.628 | -1.97716 | 0.344712 | -5.7357 | 9.71E-09 | 2.56E-07 | 0.253989 |
| TNFRSF1A | 1752.052 | -0.496 | 0.086632 | -5.72536 | 1.03E-08 | 2.72E-07 | 0.70907 |
| SHARPIN | 662.5237 | -0.50754 | 0.088664 | -5.72436 | 1.04E-08 | 2.73E-07 | 0.703419 |
| CLASRP | 1124.523 | -1.08072 | 0.188878 | -5.72179 | 1.05E-08 | 2.77E-07 | 0.472793 |
| SLC27A2 | 244.9859 | -0.97336 | 0.170131 | -5.72123 | 1.06E-08 | 2.77E-07 | 0.50932 |
| MIGA2 | 1051.255 | -0.9172 | 0.160508 | -5.71439 | 1.10E-08 | 2.88E-07 | 0.529534 |
| SLC38A9 | 1034.379 | -0.61541 | 0.107835 | -5.70693 | 1.15E-08 | 2.98E-07 | 0.652745 |
| REEP4 | 780.4175 | -0.75273 | 0.131935 | -5.7053 | 1.16E-08 | 3.01E-07 | 0.593479 |
| MUSTN1 | 9.863232 | -6.8467 | 1.200514 | -5.70314 | 1.18E-08 | 3.04E-07 | 0.008688 |
| MFSD10 | 2734.855 | -1.10562 | 0.19389 | -5.70234 | 1.18E-08 | 3.05E-07 | 0.464701 |
| ILVBL | 1735.168 | -0.7522 | 0.132018 | -5.69773 | 1.21E-08 | 3.12E-07 | 0.593696 |
| NDUFAF1 | 251.9623 | -0.68791 | 0.120967 | -5.68672 | 1.30E-08 | 3.30E-07 | 0.620754 |
| AGBL2 | 58.9411 | -1.98976 | 0.350055 | -5.68413 | 1.31E-08 | 3.34E-07 | 0.251781 |
| SSC5D | 179.1494 | -1.28283 | 0.226035 | -5.67536 | 1.38E-08 | 3.49E-07 | 0.410988 |
| KLHL34 | 8.759184 | -6.6797 | 1.177267 | -5.67391 | 1.40E-08 | 3.52E-07 | 0.009755 |
| CPXM2 | 542.606 | -0.58962 | 0.103981 | -5.67051 | 1.42E-08 | 3.58E-07 | 0.664517 |
| PRNP | 996.396 | -0.51594 | 0.091073 | -5.66519 | 1.47E-08 | 3.68E-07 | 0.699335 |
| CHST7 | 330.5968 | -0.58586 | 0.103548 | -5.6579 | 1.53E-08 | 3.83E-07 | 0.666251 |
| TMEM200( | 341.2493 | -0.60251 | 0.106847 | -5.63899 | 1.71E-08 | 4.24E-07 | 0.658607 |
| PNMA3 | 311.3853 | -0.63699 | 0.113034 | -5.63544 | 1.75E-08 | 4.32E-07 | 0.643051 |
| ADAM15 | 1201.044 | -0.69873 | 0.124059 | -5.63222 | 1.78E-08 | 4.40E-07 | 0.616116 |
| SPNS1 | 1985.513 | -0.72085 | 0.128062 | -5.62892 | 1.81E-08 | 4.47E-07 | 0.60674 |
| WDR27 | 1075.664 | -1.13023 | 0.20096 | -5.62414 | 1.86E-08 | 4.57E-07 | 0.456844 |
| C1QL4 | 49.65165 | -1.6651 | 0.296216 | -5.62123 | 1.90E-08 | 4.64E-07 | 0.315323 |
| UCKL1 | 777.6044 | -0.90868 | 0.16167 | -5.6206 | 1.90E-08 | 4.65E-07 | 0.532671 |
| TP53I13 | 836.0855 | -0.95897 | 0.170676 | -5.61865 | 1.92E-08 | 4.70E-07 | 0.514424 |
| SLC4A11 | 558.0579 | -0.76182 | 0.135887 | -5.60623 | 2.07E-08 | 5.02E-07 | 0.589753 |
| FBF1 | 399.498 | -0.92494 | 0.165229 | -5.5979 | 2.17E-08 | 5.22E-07 | 0.526704 |
| MEGF6 | 1013.024 | -1.35144 | 0.241535 | -5.5952 | 2.20E-08 | 5.27E-07 | 0.391901 |
| RELL2 | 532.2866 | -0.70853 | 0.126709 | -5.59174 | 2.25E-08 | 5.35E-07 | 0.611945 |
| FHOD1 | 1145.289 | -0.79587 | 0.142536 | -5.58363 | 2.36E-08 | 5.56E-07 | 0.575998 |
| FAM220CF | 156.9906 | -1.03391 | 0.185196 | -5.58279 | 2.37E-08 | 5.57E-07 | 0.488385 |
| GALT | 520.7242 | -0.5369 | 0.096186 | -5.58193 | 2.38E-08 | 5.58E-07 | 0.68925 |
| SLC49A3 | 124.5717 | -0.99542 | 0.178557 | -5.5748 | 2.48E-08 | 5.78E-07 | 0.50159 |
| NCLN | 3146.88 | -0.63034 | 0.113101 | -5.57324 | 2.50E-08 | 5.82E-07 | 0.646025 |
| SLC5A6 | 2502.592 | -0.57894 | 0.104028 | -5.56517 | 2.62E-08 | 6.04E-07 | 0.669458 |
| DNASE2 | 1613.241 | -0.55789 | 0.100376 | -5.55807 | 2.73E-08 | 6.28E-07 | 0.679293 |
| SLC1A4 | 365.8394 | -1.241 | 0.223669 | -5.5484 | 2.88E-08 | 6.56E-07 | 0.423078 |
| PLEC | 6396.668 | -0.91663 | 0.165267 | -5.54634 | 2.92E-08 | 6.62E-07 | 0.529747 |
| DAGLB | 845.9968 | -0.62234 | 0.112224 | -5.5455 | 2.93E-08 | 6.64E-07 | 0.649618 |
| AEBP1 | 1357.191 | -0.72083 | 0.130018 | -5.54405 | 2.96E-08 | 6.68E-07 | 0.606749 |

| | | | | | | | |
|---|---|---|---|---|---|---|---|
| TELO2 | 1780.121 | -0.97735 | 0.176366 | -5.54161 | 3.00E-08 | 6.77E-07 | 0.507911 |
| BX255925. | 609.8696 | -1.09203 | 0.197403 | -5.53199 | 3.17E-08 | 7.13E-07 | 0.4691 |
| SLC2A11 | 333.5266 | -0.69261 | 0.1252 | -5.53205 | 3.17E-08 | 7.13E-07 | 0.618732 |
| RNF166 | 664.1307 | -0.59848 | 0.108229 | -5.52981 | 3.21E-08 | 7.21E-07 | 0.660448 |
| ATP1A3 | 849.8605 | -0.48301 | 0.08738 | -5.52765 | 3.25E-08 | 7.29E-07 | 0.715485 |
| JPH4 | 1945.268 | -0.50914 | 0.092116 | -5.52717 | 3.25E-08 | 7.30E-07 | 0.702641 |
| SHC2 | 567.7357 | -1.10594 | 0.200329 | -5.52059 | 3.38E-08 | 7.55E-07 | 0.464601 |
| GDF15 | 184.9092 | -1.5404 | 0.279136 | -5.51846 | 3.42E-08 | 7.63E-07 | 0.34379 |
| SREBF1 | 2006.584 | -1.01959 | 0.184784 | -5.51774 | 3.43E-08 | 7.66E-07 | 0.493257 |
| ARHGAP4 | 1025.192 | -0.92497 | 0.168017 | -5.5052 | 3.69E-08 | 8.14E-07 | 0.526692 |
| GRIN3B | 76.48582 | -1.63541 | 0.297255 | -5.50173 | 3.76E-08 | 8.28E-07 | 0.321878 |
| BCAP31 | 2851.587 | -0.59744 | 0.108616 | -5.50046 | 3.79E-08 | 8.33E-07 | 0.660926 |
| DOK3 | 100.8655 | -1.19503 | 0.217391 | -5.49714 | 3.86E-08 | 8.47E-07 | 0.436777 |
| MTA1 | 5029.256 | -0.61371 | 0.111698 | -5.49436 | 3.92E-08 | 8.58E-07 | 0.653515 |
| TPRA1 | 516.4686 | -0.59857 | 0.109067 | -5.48813 | 4.06E-08 | 8.84E-07 | 0.660408 |
| ULK1 | 2214.577 | -0.60146 | 0.109598 | -5.48787 | 4.07E-08 | 8.84E-07 | 0.659087 |
| TSPAN4 | 1598.207 | -0.71159 | 0.129755 | -5.4841 | 4.16E-08 | 9.00E-07 | 0.610647 |
| TMEM265 | 95.50583 | -1.17598 | 0.214674 | -5.47797 | 4.30E-08 | 9.29E-07 | 0.442583 |
| EGFL7 | 1308.216 | -0.93981 | 0.17172 | -5.47289 | 4.43E-08 | 9.51E-07 | 0.521303 |
| SYT4 | 244.9681 | -1.17419 | 0.214761 | -5.46744 | 4.57E-08 | 9.80E-07 | 0.443132 |
| TRNP1 | 816.0716 | -0.58625 | 0.107296 | -5.46389 | 4.66E-08 | 9.96E-07 | 0.666071 |
| EMC3 | 586.3077 | -0.6062 | 0.111068 | -5.4579 | 4.82E-08 | 1.03E-06 | 0.656924 |
| ADAMTSL4 | 284.9191 | -1.0698 | 0.196146 | -5.45409 | 4.92E-08 | 1.05E-06 | 0.476385 |
| ST14 | 1076.199 | -0.65888 | 0.120871 | -5.45112 | 5.01E-08 | 1.06E-06 | 0.633369 |
| GPSM1 | 546.1019 | -0.53719 | 0.098564 | -5.45014 | 5.03E-08 | 1.07E-06 | 0.689114 |
| SPARC | 8217.475 | -0.46673 | 0.085731 | -5.44407 | 5.21E-08 | 1.10E-06 | 0.723604 |
| DOCK6 | 1335.171 | -0.80642 | 0.1483 | -5.43776 | 5.40E-08 | 1.13E-06 | 0.571799 |
| MNS1 | 18.8756 | -5.4519 | 1.004402 | -5.42801 | 5.70E-08 | 1.19E-06 | 0.022846 |
| MTRNR2L1 | 178470.5 | -0.85719 | 0.158129 | -5.42079 | 5.93E-08 | 1.23E-06 | 0.552029 |
| PPIC | 268.1262 | -0.69745 | 0.128694 | -5.41946 | 5.98E-08 | 1.24E-06 | 0.61666 |
| UVSSA | 1444.807 | -1.19804 | 0.221428 | -5.41052 | 6.28E-08 | 1.30E-06 | 0.435866 |
| MYL5 | 204.0077 | -1.14556 | 0.211859 | -5.40717 | 6.40E-08 | 1.32E-06 | 0.452014 |
| PRODH | 1347.078 | -0.83933 | 0.155255 | -5.40618 | 6.44E-08 | 1.32E-06 | 0.558902 |
| SCNN1D | 179.3122 | -1.51426 | 0.280805 | -5.39258 | 6.95E-08 | 1.41E-06 | 0.350076 |
| GPAA1 | 1691.428 | -0.64306 | 0.119345 | -5.38819 | 7.12E-08 | 1.44E-06 | 0.640355 |
| C16orf54 | 109.3467 | -1.70084 | 0.315797 | -5.38587 | 7.21E-08 | 1.45E-06 | 0.307606 |
| IL17RC | 362.9429 | -0.54733 | 0.101686 | -5.38254 | 7.34E-08 | 1.48E-06 | 0.684285 |
| SLC9A8 | 489.6292 | -0.59107 | 0.109884 | -5.37905 | 7.49E-08 | 1.51E-06 | 0.66385 |
| NLGN3 | 651.9847 | -0.54304 | 0.101048 | -5.37404 | 7.70E-08 | 1.55E-06 | 0.686325 |
| FOXH1 | 2416.214 | -0.62517 | 0.116386 | -5.37152 | 7.81E-08 | 1.57E-06 | 0.648343 |
| SAP25 | 100.8912 | -1.99805 | 0.372115 | -5.36945 | 7.90E-08 | 1.58E-06 | 0.250338 |
| TSTA3 | 1046.766 | -0.51905 | 0.096719 | -5.36661 | 8.02E-08 | 1.60E-06 | 0.697829 |
| COL26A1 | 2803.203 | -0.47756 | 0.089029 | -5.36406 | 8.14E-08 | 1.62E-06 | 0.718192 |
| B3GNT9 | 243.7696 | -0.61219 | 0.114283 | -5.35676 | 8.47E-08 | 1.68E-06 | 0.654204 |

| | | | | | | | |
|---|---|---|---|---|---|---|---|
| EVC2 | 324.6052 | -0.69411 | 0.129643 | -5.35403 | 8.60E-08 | 1.70E-06 | 0.618089 |
| CDC42BP( | 851.5932 | -0.99071 | 0.185102 | -5.35223 | 8.69E-08 | 1.71E-06 | 0.503231 |
| CAPN12 | 121.2774 | -1.18987 | 0.222593 | -5.34552 | 9.02E-08 | 1.76E-06 | 0.438341 |
| CPZ | 946.3954 | -0.81801 | 0.153275 | -5.33692 | 9.45E-08 | 1.83E-06 | 0.567222 |
| TRIM46 | 605.0093 | -0.87187 | 0.163387 | -5.33622 | 9.49E-08 | 1.83E-06 | 0.546439 |
| MMP19 | 156.3464 | -1.08772 | 0.203896 | -5.33468 | 9.57E-08 | 1.85E-06 | 0.470504 |
| SCAP | 3821.941 | -0.61807 | 0.115891 | -5.33324 | 9.65E-08 | 1.86E-06 | 0.651541 |
| ACAP3 | 1332.388 | -1.34579 | 0.252466 | -5.33057 | 9.79E-08 | 1.88E-06 | 0.393439 |
| QTRT1 | 1175.036 | -0.65484 | 0.12286 | -5.32994 | 9.82E-08 | 1.88E-06 | 0.635148 |
| RFT1 | 1020.815 | -0.52444 | 0.098404 | -5.32942 | 9.85E-08 | 1.88E-06 | 0.695231 |
| LMF2 | 1321.207 | -0.8871 | 0.1666 | -5.32473 | 1.01E-07 | 1.93E-06 | 0.5407 |
| SAMD4A | 402.6379 | -0.61696 | 0.115933 | -5.32169 | 1.03E-07 | 1.96E-06 | 0.652045 |
| TAP1 | 296.4655 | -1.0875 | 0.204475 | -5.31848 | 1.05E-07 | 1.99E-06 | 0.470577 |
| MST1P2 | 36.21775 | -2.66059 | 0.500356 | -5.31739 | 1.05E-07 | 2.00E-06 | 0.158155 |
| FKRP | 625.7501 | -0.73556 | 0.138353 | -5.31654 | 1.06E-07 | 2.01E-06 | 0.600586 |
| NOXO1 | 15.4731 | -4.05035 | 0.762466 | -5.31216 | 1.08E-07 | 2.05E-06 | 0.060357 |
| ZIK1 | 586.6686 | -0.90906 | 0.171135 | -5.31194 | 1.08E-07 | 2.05E-06 | 0.532534 |
| CENPT | 1325.698 | -0.85708 | 0.161487 | -5.30742 | 1.11E-07 | 2.09E-06 | 0.552069 |
| MALL | 357.8346 | -0.80433 | 0.151571 | -5.3066 | 1.12E-07 | 2.10E-06 | 0.572628 |
| OGFOD2 | 402.9648 | -0.67843 | 0.127904 | -5.30422 | 1.13E-07 | 2.12E-06 | 0.624844 |
| BAIAP2L2 | 120.142 | -1.17982 | 0.222557 | -5.3012 | 1.15E-07 | 2.15E-06 | 0.441407 |
| YPEL4 | 119.4619 | -1.04615 | 0.197578 | -5.29488 | 1.19E-07 | 2.21E-06 | 0.484258 |
| STK11IP | 587.7959 | -0.95206 | 0.179865 | -5.29319 | 1.20E-07 | 2.23E-06 | 0.516894 |
| CARMIL2 | 1051.434 | -0.91354 | 0.173012 | -5.28019 | 1.29E-07 | 2.38E-06 | 0.530882 |
| SIGIRR | 791.9565 | -1.20827 | 0.228869 | -5.2793 | 1.30E-07 | 2.39E-06 | 0.432787 |
| PPP1R12C | 1382.283 | -0.88401 | 0.167541 | -5.27636 | 1.32E-07 | 2.42E-06 | 0.54186 |
| KRTCAP3 | 421.0768 | -0.60065 | 0.114095 | -5.26443 | 1.41E-07 | 2.58E-06 | 0.659459 |
| TSPAN9 | 880.768 | -0.58867 | 0.111875 | -5.26183 | 1.43E-07 | 2.61E-06 | 0.664957 |
| TEX264 | 338.3206 | -0.7729 | 0.14695 | -5.25963 | 1.44E-07 | 2.64E-06 | 0.585238 |
| CNTNAP1 | 1096.523 | -0.72195 | 0.137306 | -5.25795 | 1.46E-07 | 2.66E-06 | 0.606279 |
| KLRG2 | 429.8224 | -0.56923 | 0.108326 | -5.25479 | 1.48E-07 | 2.70E-06 | 0.673977 |
| DNAJA3 | 1960.744 | -0.46131 | 0.087799 | -5.25414 | 1.49E-07 | 2.71E-06 | 0.726328 |
| MYO15A | 82.06004 | -1.18507 | 0.225845 | -5.24725 | 1.54E-07 | 2.80E-06 | 0.439805 |
| LIME1 | 193.7627 | -1.41727 | 0.270146 | -5.24632 | 1.55E-07 | 2.81E-06 | 0.374419 |
| FKBP11 | 541.304 | -0.76017 | 0.144969 | -5.24363 | 1.57E-07 | 2.84E-06 | 0.590428 |
| MAD2L2 | 4302.483 | -0.43704 | 0.083351 | -5.24342 | 1.58E-07 | 2.84E-06 | 0.738647 |
| NAGLU | 959.274 | -0.60066 | 0.114608 | -5.24101 | 1.60E-07 | 2.87E-06 | 0.659453 |
| LOXL3 | 587.6751 | -0.76873 | 0.146732 | -5.239 | 1.61E-07 | 2.90E-06 | 0.586934 |
| GADD45G | 285.2382 | -0.89504 | 0.170949 | -5.23569 | 1.64E-07 | 2.94E-06 | 0.537734 |
| MAGED4B | 3894.408 | -1.11002 | 0.212137 | -5.23258 | 1.67E-07 | 2.98E-06 | 0.463286 |
| DVL3 | 2241.32 | -0.42293 | 0.080863 | -5.2302 | 1.69E-07 | 3.01E-06 | 0.745909 |
| PNPO | 298.6414 | -0.94472 | 0.180641 | -5.22985 | 1.70E-07 | 3.01E-06 | 0.519529 |
| DMPK | 1065.451 | -1.29303 | 0.247315 | -5.22827 | 1.71E-07 | 3.03E-06 | 0.408093 |
| ENO3 | 345.2254 | -0.96083 | 0.183792 | -5.22777 | 1.72E-07 | 3.04E-06 | 0.513763 |

| | | | | | | | |
|---|---|---|---|---|---|---|---|
| NAT14 | 1012.141 | -0.7074 | 0.135317 | -5.2277 | 1.72E-07 | 3.04E-06 | 0.612424 |
| SH2B1 | 1104.679 | -0.92743 | 0.177482 | -5.22551 | 1.74E-07 | 3.07E-06 | 0.525792 |
| CLN6 | 2050.516 | -0.47791 | 0.0915 | -5.22307 | 1.76E-07 | 3.10E-06 | 0.718016 |
| MAFK | 350.5638 | -0.5918 | 0.113311 | -5.2228 | 1.76E-07 | 3.11E-06 | 0.663515 |
| KIF1A | 18257.36 | -0.52415 | 0.100406 | -5.22035 | 1.79E-07 | 3.14E-06 | 0.695366 |
| RELB | 408.4127 | -0.5839 | 0.111852 | -5.22031 | 1.79E-07 | 3.14E-06 | 0.667158 |
| TSC2 | 2558.555 | -0.98954 | 0.18955 | -5.22044 | 1.78E-07 | 3.14E-06 | 0.50364 |
| ATF5 | 2260.432 | -1.19722 | 0.229438 | -5.21808 | 1.81E-07 | 3.17E-06 | 0.436114 |
| SYNGR1 | 582.7922 | -0.71969 | 0.137973 | -5.21618 | 1.83E-07 | 3.20E-06 | 0.607227 |
| SLC43A3 | 749.8921 | -0.46273 | 0.08876 | -5.21328 | 1.86E-07 | 3.24E-06 | 0.725612 |
| AGRN | 7442.319 | -1.01192 | 0.194205 | -5.21059 | 1.88E-07 | 3.29E-06 | 0.495885 |
| SGSM2 | 1979.92 | -1.05434 | 0.202727 | -5.20079 | 1.98E-07 | 3.45E-06 | 0.481517 |
| MTRNR2L6 | 11630.92 | -0.85756 | 0.16491 | -5.20017 | 1.99E-07 | 3.46E-06 | 0.551884 |
| LAMC2 | 588.4625 | -0.61503 | 0.11832 | -5.19803 | 2.01E-07 | 3.50E-06 | 0.652917 |
| AP5Z1 | 744.6337 | -0.78395 | 0.150899 | -5.1952 | 2.05E-07 | 3.54E-06 | 0.580775 |
| ATP2A3 | 1018.864 | -0.50999 | 0.098169 | -5.19506 | 2.05E-07 | 3.54E-06 | 0.702227 |
| MFGE8 | 25881.99 | -0.57868 | 0.111475 | -5.19113 | 2.09E-07 | 3.61E-06 | 0.669576 |
| LTBP3 | 333.8838 | -1.04751 | 0.201929 | -5.18751 | 2.13E-07 | 3.68E-06 | 0.483803 |
| TMEM63A | 925.5291 | -0.79613 | 0.153854 | -5.17458 | 2.28E-07 | 3.91E-06 | 0.575892 |
| HYI | 221.3063 | -0.73432 | 0.141995 | -5.17144 | 2.32E-07 | 3.96E-06 | 0.601102 |
| CHGB | 148.4427 | -1.0566 | 0.204401 | -5.16922 | 2.35E-07 | 4.00E-06 | 0.480765 |
| C17orf107 | 19.98465 | -2.51136 | 0.486706 | -5.15992 | 2.47E-07 | 4.18E-06 | 0.17539 |
| CRYBG2 | 500.8932 | -0.87119 | 0.169004 | -5.15483 | 2.54E-07 | 4.29E-06 | 0.546696 |
| ZNF865 | 524.9812 | -0.45696 | 0.08867 | -5.15345 | 2.56E-07 | 4.31E-06 | 0.728521 |
| GPR137 | 566.1759 | -1.0089 | 0.195893 | -5.15026 | 2.60E-07 | 4.38E-06 | 0.496924 |
| PAQR6 | 446.6923 | -1.43969 | 0.279556 | -5.14993 | 2.61E-07 | 4.38E-06 | 0.368646 |
| CNTN1 | 477.0946 | -0.84008 | 0.163147 | -5.14922 | 2.62E-07 | 4.39E-06 | 0.558613 |
| ITGA5 | 1637.824 | -0.69275 | 0.134819 | -5.13834 | 2.77E-07 | 4.64E-06 | 0.618674 |
| CHRNB1 | 566.278 | -0.43837 | 0.085425 | -5.13164 | 2.87E-07 | 4.79E-06 | 0.737968 |
| RAB11FIP3 | 773.8903 | -0.6385 | 0.124503 | -5.12837 | 2.92E-07 | 4.87E-06 | 0.642382 |
| DPH7 | 780.6743 | -0.61759 | 0.120494 | -5.12547 | 2.97E-07 | 4.93E-06 | 0.651759 |
| TNKS1BP1 | 4958.193 | -0.66382 | 0.129513 | -5.12545 | 2.97E-07 | 4.93E-06 | 0.631207 |
| GRIN2D | 535.0754 | -0.68045 | 0.132781 | -5.12461 | 2.98E-07 | 4.95E-06 | 0.623969 |
| AIFM3 | 47.3952 | -1.74214 | 0.340389 | -5.11807 | 3.09E-07 | 5.10E-06 | 0.298927 |
| UNC79 | 194.7808 | -0.75351 | 0.147317 | -5.11492 | 3.14E-07 | 5.18E-06 | 0.593157 |
| TMEM25 | 334.3048 | -0.80383 | 0.157293 | -5.11039 | 3.21E-07 | 5.29E-06 | 0.572826 |
| CNNM3 | 1199.557 | -0.53518 | 0.104767 | -5.10829 | 3.25E-07 | 5.35E-06 | 0.690074 |
| BORCS8 | 228.8357 | -0.94679 | 0.185417 | -5.1063 | 3.29E-07 | 5.40E-06 | 0.518785 |
| AC007325 | 3458.815 | -0.68902 | 0.135098 | -5.10014 | 3.39E-07 | 5.55E-06 | 0.620276 |
| SLC35F6 | 463.9609 | -0.50053 | 0.098191 | -5.09758 | 3.44E-07 | 5.62E-06 | 0.706845 |
| CPT1B | 871.8689 | -1.3064 | 0.256782 | -5.08759 | 3.63E-07 | 5.88E-06 | 0.404329 |
| CPT1C | 1281.631 | -0.82878 | 0.163043 | -5.0832 | 3.71E-07 | 6.01E-06 | 0.563006 |
| SFRP5 | 225.0493 | -1.08566 | 0.213769 | -5.07866 | 3.80E-07 | 6.14E-06 | 0.471176 |
| CDC42EP1 | 670.1524 | -0.93924 | 0.185136 | -5.07324 | 3.91E-07 | 6.29E-06 | 0.521508 |

| | | | | | | | |
|---|---|---|---|---|---|---|---|
| TM7SF2 | 970.6798 | -0.67384 | 0.132862 | -5.07171 | 3.94E-07 | 6.33E-06 | 0.626836 |
| AC243919 | 734.3772 | -3.92968 | 0.775195 | -5.06927 | 3.99E-07 | 6.40E-06 | 0.065622 |
| XYLT2 | 1235.723 | -0.6689 | 0.132012 | -5.06694 | 4.04E-07 | 6.46E-06 | 0.628986 |
| SULT1B1 | 584.8758 | -0.5406 | 0.106781 | -5.06269 | 4.13E-07 | 6.60E-06 | 0.687486 |
| NPIPB5 | 2760.266 | -0.73082 | 0.144409 | -5.06078 | 4.18E-07 | 6.66E-06 | 0.60256 |
| CAMTA2 | 387.1619 | -0.92215 | 0.182511 | -5.0526 | 4.36E-07 | 6.94E-06 | 0.527721 |
| SSH3 | 696.4403 | -0.45475 | 0.090057 | -5.04958 | 4.43E-07 | 7.03E-06 | 0.729636 |
| IGSF8 | 1781.342 | -0.64603 | 0.127956 | -5.04881 | 4.45E-07 | 7.06E-06 | 0.639038 |
| ECHDC2 | 327.3935 | -0.77308 | 0.153243 | -5.04476 | 4.54E-07 | 7.19E-06 | 0.585169 |
| HMCN2 | 1028.679 | -1.16502 | 0.231 | -5.04338 | 4.57E-07 | 7.24E-06 | 0.445958 |
| SULT1A3 | 336.965 | -1.59668 | 0.316648 | -5.04245 | 4.60E-07 | 7.27E-06 | 0.330636 |
| RAD52 | 235.5567 | -0.71275 | 0.141364 | -5.04199 | 4.61E-07 | 7.28E-06 | 0.610154 |
| PLIN4 | 112.8452 | -0.90372 | 0.179285 | -5.0407 | 4.64E-07 | 7.31E-06 | 0.534506 |
| TNFRSF25 | 359.2668 | -1.41189 | 0.280184 | -5.03916 | 4.68E-07 | 7.37E-06 | 0.375819 |
| HOOK2 | 1384.659 | -0.96329 | 0.191197 | -5.03818 | 4.70E-07 | 7.40E-06 | 0.512887 |
| CREB3L1 | 618.055 | -0.6144 | 0.122025 | -5.03501 | 4.78E-07 | 7.49E-06 | 0.653203 |
| SLC4A2 | 2419.055 | -0.96814 | 0.192283 | -5.03498 | 4.78E-07 | 7.49E-06 | 0.511163 |
| PLTP | 4306.611 | -0.5017 | 0.09972 | -5.03112 | 4.88E-07 | 7.63E-06 | 0.706272 |
| TMEM8A | 3283.726 | -0.4876 | 0.096986 | -5.02754 | 4.97E-07 | 7.76E-06 | 0.713209 |
| GAA | 2897.525 | -0.87384 | 0.17382 | -5.02729 | 4.97E-07 | 7.77E-06 | 0.545691 |
| FGF21 | 62.39239 | -1.39422 | 0.277457 | -5.02502 | 5.03E-07 | 7.85E-06 | 0.380449 |
| CBS | 11462.14 | -0.50813 | 0.101239 | -5.01915 | 5.19E-07 | 8.04E-06 | 0.703132 |
| CYCSP6 | 6.286125 | -6.21974 | 1.239895 | -5.01634 | 5.27E-07 | 8.13E-06 | 0.013417 |
| ADAMTSL2 | 111.3317 | -0.89624 | 0.178923 | -5.00908 | 5.47E-07 | 8.40E-06 | 0.537285 |
| ORAI1 | 552.7244 | -0.45371 | 0.090639 | -5.00568 | 5.57E-07 | 8.52E-06 | 0.730161 |
| CPT1A | 265.213 | -1.06166 | 0.212183 | -5.00351 | 5.63E-07 | 8.60E-06 | 0.47908 |
| ABCC9 | 295.6929 | -0.62647 | 0.125321 | -4.99896 | 5.76E-07 | 8.77E-06 | 0.647758 |
| RASSF7 | 547.4967 | -0.84806 | 0.169741 | -4.99621 | 5.85E-07 | 8.89E-06 | 0.55553 |
| MTRNR2L1 | 471.1877 | -1.0971 | 0.219616 | -4.99554 | 5.87E-07 | 8.90E-06 | 0.467455 |
| COMT | 819.3391 | -0.59131 | 0.11838 | -4.99498 | 5.88E-07 | 8.92E-06 | 0.663741 |
| MTRNR2L4 | 57.60283 | -1.52411 | 0.305143 | -4.99474 | 5.89E-07 | 8.92E-06 | 0.347694 |
| CELSR3 | 2089.598 | -0.69985 | 0.140139 | -4.99397 | 5.91E-07 | 8.93E-06 | 0.615636 |
| ZMIZ2 | 2160.751 | -0.71488 | 0.143339 | -4.98733 | 6.12E-07 | 9.23E-06 | 0.609256 |
| PKLR | 204.0341 | -0.94042 | 0.188743 | -4.98251 | 6.28E-07 | 9.45E-06 | 0.521083 |
| KIRREL2 | 133.4331 | -0.90562 | 0.181767 | -4.98231 | 6.28E-07 | 9.45E-06 | 0.533804 |
| MATK | 440.2477 | -0.74893 | 0.150376 | -4.98036 | 6.35E-07 | 9.52E-06 | 0.595045 |
| TMC6 | 1597.91 | -0.82737 | 0.166173 | -4.97894 | 6.39E-07 | 9.57E-06 | 0.563557 |
| FADS3 | 1345.008 | -0.84563 | 0.16986 | -4.9784 | 6.41E-07 | 9.58E-06 | 0.556468 |
| KCNK6 | 879.3487 | -0.67595 | 0.135776 | -4.97841 | 6.41E-07 | 9.58E-06 | 0.62592 |
| TPCN2 | 586.3694 | -0.57742 | 0.116051 | -4.9756 | 6.50E-07 | 9.71E-06 | 0.67016 |
| CDCP1 | 707.3078 | -0.54838 | 0.110271 | -4.97303 | 6.59E-07 | 9.83E-06 | 0.683786 |
| CILP2 | 253.0622 | -0.79269 | 0.159435 | -4.9719 | 6.63E-07 | 9.88E-06 | 0.577265 |
| ERFE | 94.39642 | -2.11548 | 0.425657 | -4.96991 | 6.70E-07 | 9.95E-06 | 0.230769 |
| SLC17A9 | 509.8869 | -0.92227 | 0.185676 | -4.96708 | 6.80E-07 | 1.01E-05 | 0.527678 |

| | | | | | | | |
|---|---|---|---|---|---|---|---|
| DPAGT1 | 1096.365 | -0.50016 | 0.100699 | -4.96685 | 6.80E-07 | 1.01E-05 | 0.70703 |
| ACRBP | 47.00867 | -1.31512 | 0.264968 | -4.9633 | 6.93E-07 | 1.03E-05 | 0.401893 |
| CDK11A | 769.0117 | -1.05094 | 0.211765 | -4.96277 | 6.95E-07 | 1.03E-05 | 0.482653 |
| SLC15A1 | 63.14838 | -1.21549 | 0.244985 | -4.9615 | 6.99E-07 | 1.03E-05 | 0.430626 |
| SPTBN1 | 8431.835 | -0.70456 | 0.142007 | -4.96143 | 7.00E-07 | 1.03E-05 | 0.613631 |
| RASGRP2 | 499.9158 | -0.86967 | 0.175378 | -4.95883 | 7.09E-07 | 1.04E-05 | 0.547272 |
| TMEM259 | 4724.066 | -0.90583 | 0.182752 | -4.9566 | 7.17E-07 | 1.05E-05 | 0.533726 |
| SLC39A14 | 7921.786 | -0.53624 | 0.10825 | -4.95369 | 7.28E-07 | 1.07E-05 | 0.689566 |
| LRCH4 | 889.2781 | -0.4817 | 0.097298 | -4.95083 | 7.39E-07 | 1.08E-05 | 0.716132 |
| TMEM201 | 804.5714 | -0.56664 | 0.114477 | -4.94982 | 7.43E-07 | 1.09E-05 | 0.675187 |
| MTND1P23 | 73.61244 | -2.8865 | 0.583286 | -4.94869 | 7.47E-07 | 1.09E-05 | 0.135231 |
| SYNGR2 | 1120.721 | -0.56865 | 0.115013 | -4.9442 | 7.65E-07 | 1.11E-05 | 0.674249 |
| AC005954 | 425.6192 | -0.67155 | 0.135954 | -4.93953 | 7.83E-07 | 1.14E-05 | 0.627833 |
| WDR18 | 1212.552 | -0.59388 | 0.120265 | -4.93806 | 7.89E-07 | 1.14E-05 | 0.66256 |
| AC093668 | 45.44222 | -2.66199 | 0.539286 | -4.93614 | 7.97E-07 | 1.15E-05 | 0.158001 |
| CBWD5 | 668.242 | -0.5553 | 0.112585 | -4.93229 | 8.13E-07 | 1.17E-05 | 0.680516 |
| ADAMTS14 | 115.0713 | -1.02815 | 0.208532 | -4.93044 | 8.20E-07 | 1.18E-05 | 0.490338 |
| EXOC3L4 | 8.541474 | -3.60813 | 0.73245 | -4.92611 | 8.39E-07 | 1.20E-05 | 0.082006 |
| LPCAT4 | 1026.866 | -0.73769 | 0.150023 | -4.91719 | 8.78E-07 | 1.25E-05 | 0.599699 |
| BEST1 | 183.5361 | -1.32767 | 0.270143 | -4.9147 | 8.89E-07 | 1.27E-05 | 0.398411 |
| TOR1A | 1057.943 | -0.41507 | 0.084479 | -4.91324 | 8.96E-07 | 1.27E-05 | 0.749985 |
| AC010761 | 463.8947 | -0.55573 | 0.113135 | -4.91209 | 9.01E-07 | 1.28E-05 | 0.680313 |
| ZNF76 | 620.1909 | -0.48687 | 0.099235 | -4.90627 | 9.28E-07 | 1.32E-05 | 0.71357 |
| SYNGR3 | 425.5032 | -0.65937 | 0.134585 | -4.89929 | 9.62E-07 | 1.36E-05 | 0.633155 |
| PACS2 | 2102.393 | -0.56637 | 0.115654 | -4.8971 | 9.73E-07 | 1.38E-05 | 0.675314 |
| ADM | 1419.517 | -0.53653 | 0.109647 | -4.89329 | 9.92E-07 | 1.40E-05 | 0.689425 |
| ROBO3 | 430.7065 | -1.05395 | 0.215527 | -4.89011 | 1.01E-06 | 1.42E-05 | 0.481647 |
| AC004151 | 1464.008 | -0.57547 | 0.117712 | -4.88881 | 1.01E-06 | 1.42E-05 | 0.671066 |
| RAB13 | 4385.247 | -0.80396 | 0.164448 | -4.88884 | 1.01E-06 | 1.42E-05 | 0.572775 |
| RPS6KA4 | 391.4228 | -0.56593 | 0.11579 | -4.88761 | 1.02E-06 | 1.43E-05 | 0.675518 |
| COL6A1 | 4224.587 | -0.68408 | 0.14003 | -4.88525 | 1.03E-06 | 1.45E-05 | 0.622402 |
| ADAM8 | 131.4218 | -1.07554 | 0.220275 | -4.88273 | 1.05E-06 | 1.46E-05 | 0.474493 |
| HM13 | 2060.514 | -0.53437 | 0.109466 | -4.88158 | 1.05E-06 | 1.47E-05 | 0.690462 |
| CCDC24 | 266.4268 | -0.68415 | 0.140191 | -4.88013 | 1.06E-06 | 1.47E-05 | 0.622373 |
| ALG1 | 1182.109 | -0.54187 | 0.111048 | -4.8796 | 1.06E-06 | 1.48E-05 | 0.68688 |
| COL1A1 | 8768.554 | -0.65232 | 0.133956 | -4.86966 | 1.12E-06 | 1.54E-05 | 0.636256 |
| PDZD4 | 2457.152 | -0.62085 | 0.127527 | -4.86836 | 1.13E-06 | 1.55E-05 | 0.650289 |
| SERPINI1 | 247.8671 | -0.64331 | 0.132165 | -4.86749 | 1.13E-06 | 1.56E-05 | 0.640241 |
| SMPDL3B | 1456.452 | -0.76608 | 0.15764 | -4.8597 | 1.18E-06 | 1.61E-05 | 0.588012 |
| GPR50 | 165.8702 | -0.81726 | 0.168247 | -4.85753 | 1.19E-06 | 1.63E-05 | 0.567517 |
| HLA-DOA | 618.8025 | -0.58758 | 0.121125 | -4.85099 | 1.23E-06 | 1.68E-05 | 0.665461 |
| TMEM43 | 1546.803 | -0.4295 | 0.088548 | -4.85051 | 1.23E-06 | 1.68E-05 | 0.742517 |
| HLA-V | 23.2589 | -2.32301 | 0.47961 | -4.84354 | 1.28E-06 | 1.73E-05 | 0.19985 |
| AC116562 | 246.4042 | -0.74416 | 0.153724 | -4.8409 | 1.29E-06 | 1.75E-05 | 0.597015 |

| | | | | | | | |
|---|---|---|---|---|---|---|---|
| FBXO41 | 799.7966 | -0.41775 | 0.08631 | -4.84016 | 1.30E-06 | 1.76E-05 | 0.748589 |
| KCTD15 | 1254.153 | -0.44846 | 0.092706 | -4.83741 | 1.32E-06 | 1.78E-05 | 0.732825 |
| TMEM82 | 67.69018 | -1.20616 | 0.249386 | -4.8365 | 1.32E-06 | 1.78E-05 | 0.433421 |
| CCDC154 | 28.87308 | -2.41678 | 0.499861 | -4.8349 | 1.33E-06 | 1.79E-05 | 0.187274 |
| FAT1 | 6177.242 | -0.78999 | 0.163394 | -4.83486 | 1.33E-06 | 1.79E-05 | 0.578348 |
| FBXO46 | 826.2078 | -0.50451 | 0.104353 | -4.8346 | 1.33E-06 | 1.79E-05 | 0.704901 |
| MTRNR2L1 | 56.1488 | -1.60229 | 0.331545 | -4.83281 | 1.35E-06 | 1.81E-05 | 0.329353 |
| HDAC10 | 461.5271 | -1.27628 | 0.264203 | -4.83068 | 1.36E-06 | 1.82E-05 | 0.412859 |
| STOM | 735.2932 | -0.93708 | 0.194052 | -4.82899 | 1.37E-06 | 1.84E-05 | 0.52229 |
| YIF1A | 1144.816 | -0.45669 | 0.094645 | -4.82531 | 1.40E-06 | 1.87E-05 | 0.728656 |
| ADM2 | 1101.453 | -0.57173 | 0.11855 | -4.82274 | 1.42E-06 | 1.89E-05 | 0.672808 |
| CPSF1 | 3869.965 | -0.82415 | 0.170886 | -4.82282 | 1.42E-06 | 1.89E-05 | 0.564815 |
| AC242376 | 137.8499 | -1.20153 | 0.249171 | -4.8221 | 1.42E-06 | 1.89E-05 | 0.434815 |
| TYSND1 | 572.298 | -0.47917 | 0.099379 | -4.82165 | 1.42E-06 | 1.90E-05 | 0.717391 |
| SLC27A4 | 792.3604 | -0.71624 | 0.148664 | -4.81782 | 1.45E-06 | 1.93E-05 | 0.608683 |
| MAPK8IP2 | 467.5694 | -0.95687 | 0.198746 | -4.81453 | 1.48E-06 | 1.96E-05 | 0.515174 |
| PWWP2B | 355.1753 | -0.56157 | 0.116692 | -4.81243 | 1.49E-06 | 1.98E-05 | 0.677564 |
| FLII | 5808.58 | -0.47631 | 0.098986 | -4.81183 | 1.50E-06 | 1.98E-05 | 0.718816 |
| FAR2P1 | 82.90888 | -1.99951 | 0.416211 | -4.80407 | 1.55E-06 | 2.05E-05 | 0.250085 |
| ZDHHC15 | 167.1519 | -0.74488 | 0.155128 | -4.80172 | 1.57E-06 | 2.07E-05 | 0.596716 |
| SHISA9 | 803.8838 | -0.57742 | 0.120303 | -4.79975 | 1.59E-06 | 2.09E-05 | 0.670159 |
| UBXN8 | 200.1621 | -0.69053 | 0.143896 | -4.79879 | 1.60E-06 | 2.10E-05 | 0.619627 |
| CYP4V2 | 170.3364 | -0.84575 | 0.176517 | -4.79131 | 1.66E-06 | 2.17E-05 | 0.556423 |
| RNA5-8SP( | 69.54133 | -1.96359 | 0.409911 | -4.79029 | 1.67E-06 | 2.18E-05 | 0.256389 |
| TRIM41 | 899.1613 | -0.67158 | 0.140204 | -4.79004 | 1.67E-06 | 2.18E-05 | 0.627818 |
| AC018638 | 144.5468 | -1.03847 | 0.21682 | -4.78954 | 1.67E-06 | 2.18E-05 | 0.486843 |
| NSMCE1 | 302.0217 | -0.61438 | 0.128274 | -4.78959 | 1.67E-06 | 2.18E-05 | 0.653211 |
| AC138969 | 2170.557 | -1.03717 | 0.216666 | -4.78697 | 1.69E-06 | 2.21E-05 | 0.487282 |
| EMC1 | 2334.856 | -0.4912 | 0.10262 | -4.78663 | 1.70E-06 | 2.21E-05 | 0.711432 |
| CGN | 2194.935 | -0.64944 | 0.135719 | -4.78517 | 1.71E-06 | 2.22E-05 | 0.637528 |
| REX1BD | 827.3976 | -0.66523 | 0.139163 | -4.78024 | 1.75E-06 | 2.27E-05 | 0.630586 |
| AL627309. | 180.2022 | -1.87195 | 0.391893 | -4.77669 | 1.78E-06 | 2.31E-05 | 0.273204 |
| TM6SF2 | 127.6458 | -0.92905 | 0.194529 | -4.77588 | 1.79E-06 | 2.32E-05 | 0.525205 |
| DOC2GP | 91.15645 | -1.28452 | 0.269354 | -4.7689 | 1.85E-06 | 2.39E-05 | 0.410507 |
| HIF3A | 1102.913 | -0.72007 | 0.151036 | -4.76751 | 1.87E-06 | 2.40E-05 | 0.607069 |
| EDEM2 | 712.6929 | -0.50219 | 0.105363 | -4.76626 | 1.88E-06 | 2.42E-05 | 0.706036 |
| SCAMP4 | 1439.045 | -0.69941 | 0.146936 | -4.75995 | 1.94E-06 | 2.49E-05 | 0.615825 |
| SLC7A10 | 91.90792 | -1.29308 | 0.271705 | -4.75911 | 1.94E-06 | 2.49E-05 | 0.40808 |
| SMG9 | 737.194 | -0.52366 | 0.11003 | -4.75925 | 1.94E-06 | 2.49E-05 | 0.695605 |
| KRI1 | 1027.04 | -0.59717 | 0.125706 | -4.75055 | 2.03E-06 | 2.58E-05 | 0.661049 |
| AIRE | 13.20502 | -4.45432 | 0.937977 | -4.74885 | 2.05E-06 | 2.60E-05 | 0.045616 |
| ADGRB1 | 400.6124 | -1.00335 | 0.211332 | -4.74772 | 2.06E-06 | 2.61E-05 | 0.498842 |
| CACNA1H | 2275.326 | -0.70738 | 0.149004 | -4.7474 | 2.06E-06 | 2.61E-05 | 0.612431 |
| PLA2G6 | 429.6838 | -0.65739 | 0.13857 | -4.74411 | 2.09E-06 | 2.65E-05 | 0.634024 |

| | | | | | | | |
|---|---|---|---|---|---|---|---|
| TMEM222 | 620.3895 | -0.56061 | 0.118232 | -4.74164 | 2.12E-06 | 2.68E-05 | 0.678013 |
| ZNF229 | 10.2886 | -6.90316 | 1.457739 | -4.73553 | 2.18E-06 | 2.76E-05 | 0.008355 |
| GSAP | 111.618 | -1.20365 | 0.254199 | -4.73507 | 2.19E-06 | 2.76E-05 | 0.434175 |
| MACF1 | 8116.825 | -0.76344 | 0.161232 | -4.73501 | 2.19E-06 | 2.76E-05 | 0.589091 |
| AC124319 | 2623.157 | -0.54182 | 0.114468 | -4.73337 | 2.21E-06 | 2.78E-05 | 0.686904 |
| E4F1 | 808.661 | -0.80827 | 0.170836 | -4.73124 | 2.23E-06 | 2.80E-05 | 0.571068 |
| WIPF3 | 563.5608 | -0.51481 | 0.108952 | -4.72509 | 2.30E-06 | 2.89E-05 | 0.699887 |
| DOCK9 | 581.9987 | -0.60749 | 0.128638 | -4.72248 | 2.33E-06 | 2.92E-05 | 0.656338 |
| CNPY3 | 1405.206 | -0.5385 | 0.114246 | -4.71351 | 2.43E-06 | 3.05E-05 | 0.688486 |
| LAMP1 | 6545.616 | -0.44935 | 0.0954 | -4.71016 | 2.48E-06 | 3.09E-05 | 0.732372 |
| TMEM86B | 100.7861 | -1.27943 | 0.271732 | -4.70843 | 2.50E-06 | 3.11E-05 | 0.411958 |
| FAM155B | 512.1153 | -0.52905 | 0.112438 | -4.70528 | 2.54E-06 | 3.15E-05 | 0.69301 |
| CEND1 | 114.1406 | -1.36223 | 0.289858 | -4.69963 | 2.61E-06 | 3.22E-05 | 0.388981 |
| MEF2B | 319.1139 | -1.31591 | 0.280013 | -4.69944 | 2.61E-06 | 3.22E-05 | 0.401673 |
| TF | 83.57894 | -1.03469 | 0.220313 | -4.69646 | 2.65E-06 | 3.27E-05 | 0.48812 |
| RHOT2 | 3685.909 | -0.87525 | 0.186373 | -4.6962 | 2.65E-06 | 3.27E-05 | 0.545161 |
| NPHS1 | 43.33835 | -1.43188 | 0.305153 | -4.69234 | 2.70E-06 | 3.32E-05 | 0.370647 |
| ADAMTS13 | 407.8706 | -1.45873 | 0.311193 | -4.68756 | 2.76E-06 | 3.40E-05 | 0.363812 |
| ATP13A2 | 3204.763 | -0.63756 | 0.136029 | -4.68696 | 2.77E-06 | 3.40E-05 | 0.642799 |
| MMP23B | 231.1751 | -1.13972 | 0.243533 | -4.67996 | 2.87E-06 | 3.51E-05 | 0.453847 |
| JMJD7-PLA | 455.5124 | -0.88045 | 0.188157 | -4.67934 | 2.88E-06 | 3.52E-05 | 0.543198 |
| RSPO4 | 427.1315 | -0.71986 | 0.153985 | -4.67485 | 2.94E-06 | 3.59E-05 | 0.607158 |
| TOR4A | 352.981 | -0.70162 | 0.150134 | -4.6733 | 2.96E-06 | 3.61E-05 | 0.61488 |
| PKD1P6 | 417.1539 | -0.74793 | 0.160181 | -4.66927 | 3.02E-06 | 3.67E-05 | 0.595457 |
| DNLZ | 177.5769 | -0.63538 | 0.136121 | -4.66774 | 3.05E-06 | 3.70E-05 | 0.643771 |
| TEDC1 | 484.5066 | -0.58323 | 0.125054 | -4.66383 | 3.10E-06 | 3.76E-05 | 0.667467 |
| NPIPB13 | 200.5024 | -2.00878 | 0.431193 | -4.65866 | 3.18E-06 | 3.85E-05 | 0.248482 |
| NDUFS7 | 1263.55 | -0.47277 | 0.101535 | -4.65623 | 3.22E-06 | 3.88E-05 | 0.720579 |
| TMEM161A | 1226.215 | -0.54892 | 0.117935 | -4.65443 | 3.25E-06 | 3.90E-05 | 0.68353 |
| GSTT2B | 106.0178 | -1.15682 | 0.248667 | -4.65208 | 3.29E-06 | 3.94E-05 | 0.448501 |
| ZSCAN10 | 4228.017 | -0.47742 | 0.10264 | -4.65138 | 3.30E-06 | 3.95E-05 | 0.718261 |
| AGAP9 | 180.4484 | -1.03962 | 0.223544 | -4.65062 | 3.31E-06 | 3.96E-05 | 0.486457 |
| RYR1 | 1280.43 | -0.87189 | 0.18749 | -4.65032 | 3.31E-06 | 3.96E-05 | 0.54643 |
| NEURL4 | 1356.125 | -0.94682 | 0.203747 | -4.64705 | 3.37E-06 | 4.02E-05 | 0.518774 |
| TMUB2 | 559.6313 | -0.52648 | 0.113293 | -4.64706 | 3.37E-06 | 4.02E-05 | 0.694247 |
| MYC | 882.3581 | -0.82694 | 0.17806 | -4.64414 | 3.41E-06 | 4.07E-05 | 0.563724 |
| COQ4 | 689.1118 | -0.47079 | 0.101402 | -4.64276 | 3.44E-06 | 4.09E-05 | 0.721571 |
| ODF3L2 | 12.46172 | -2.85869 | 0.615808 | -4.64217 | 3.45E-06 | 4.10E-05 | 0.137864 |
| NEIL1 | 228.6277 | -0.91289 | 0.196688 | -4.64132 | 3.46E-06 | 4.11E-05 | 0.531119 |
| DDX49 | 843.036 | -0.48745 | 0.105028 | -4.64115 | 3.46E-06 | 4.11E-05 | 0.713285 |
| WNT10B | 90.34829 | -1.10946 | 0.23928 | -4.63665 | 3.54E-06 | 4.19E-05 | 0.463468 |
| NPIPB4 | 343.2448 | -0.76813 | 0.16579 | -4.63315 | 3.60E-06 | 4.26E-05 | 0.587178 |
| CAPS | 117.9629 | -1.24267 | 0.268411 | -4.62972 | 3.66E-06 | 4.32E-05 | 0.422591 |
| MUS81 | 885.6465 | -0.87932 | 0.189938 | -4.62951 | 3.67E-06 | 4.32E-05 | 0.543624 |

| | | | | | | | |
|---|---|---|---|---|---|---|---|
| AGPAT3 | 415.7712 | -0.84946 | 0.183679 | -4.62469 | 3.75E-06 | 4.42E-05 | 0.554993 |
| PIANP | 346.7161 | -0.55869 | 0.120899 | -4.62113 | 3.82E-06 | 4.49E-05 | 0.678918 |
| CCDC130 | 400.28 | -0.47467 | 0.102776 | -4.61846 | 3.87E-06 | 4.54E-05 | 0.719632 |
| BMP7 | 469.6464 | -0.66325 | 0.143716 | -4.61504 | 3.93E-06 | 4.59E-05 | 0.631453 |
| APOE | 25758.28 | -0.78162 | 0.169477 | -4.61196 | 3.99E-06 | 4.65E-05 | 0.581712 |
| INTS11 | 2952.693 | -0.59481 | 0.129007 | -4.61071 | 4.01E-06 | 4.67E-05 | 0.66213 |
| RNASEH1F | 120.8949 | -1.02794 | 0.222966 | -4.6103 | 4.02E-06 | 4.68E-05 | 0.490411 |
| ZFP36 | 149.3161 | -0.81537 | 0.176875 | -4.60988 | 4.03E-06 | 4.69E-05 | 0.568261 |
| AUP1 | 1804.17 | -0.70056 | 0.152002 | -4.60885 | 4.05E-06 | 4.71E-05 | 0.615335 |
| CHD5 | 142.984 | -1.4691 | 0.31879 | -4.60837 | 4.06E-06 | 4.71E-05 | 0.361207 |
| CCDC106 | 429.4216 | -0.509 | 0.110477 | -4.60728 | 4.08E-06 | 4.73E-05 | 0.702709 |
| PSD | 237.3429 | -0.8752 | 0.190091 | -4.60411 | 4.14E-06 | 4.80E-05 | 0.545178 |
| PRRG3 | 200.9655 | -1.56235 | 0.339407 | -4.60317 | 4.16E-06 | 4.82E-05 | 0.3386 |
| MDN1 | 5298.87 | -0.73417 | 0.159542 | -4.60172 | 4.19E-06 | 4.84E-05 | 0.601164 |
| SLC38A10 | 1367.862 | -0.87171 | 0.189431 | -4.60171 | 4.19E-06 | 4.84E-05 | 0.5465 |
| FDX2 | 336.9861 | -0.6145 | 0.133741 | -4.59471 | 4.33E-06 | 5.00E-05 | 0.653157 |
| ROGDI | 719.8208 | -0.42101 | 0.091684 | -4.59197 | 4.39E-06 | 5.06E-05 | 0.746901 |
| IGDCC4 | 1209.094 | -0.64781 | 0.141091 | -4.59144 | 4.40E-06 | 5.06E-05 | 0.638248 |
| EXOC3L1 | 41.10687 | -1.38976 | 0.302726 | -4.59082 | 4.42E-06 | 5.07E-05 | 0.381628 |
| DPF1 | 218.8461 | -0.76485 | 0.166788 | -4.58575 | 4.52E-06 | 5.18E-05 | 0.588515 |
| AC015813 | 389.3178 | -1.32536 | 0.289049 | -4.58526 | 4.53E-06 | 5.19E-05 | 0.399048 |
| MAPK8IP3 | 1807.465 | -0.63742 | 0.139061 | -4.58376 | 4.57E-06 | 5.21E-05 | 0.64286 |
| DNM1 | 1436.289 | -0.82262 | 0.17947 | -4.58359 | 4.57E-06 | 5.21E-05 | 0.565415 |
| P2RX1 | 27.23467 | -1.729 | 0.377482 | -4.58034 | 4.64E-06 | 5.29E-05 | 0.301662 |
| CHTF18 | 2817.864 | -1.10706 | 0.24172 | -4.57994 | 4.65E-06 | 5.29E-05 | 0.464239 |
| ARFRP1 | 604.459 | -0.53023 | 0.115885 | -4.57547 | 4.75E-06 | 5.41E-05 | 0.692446 |
| STIM2 | 536.1112 | -0.61488 | 0.134501 | -4.57159 | 4.84E-06 | 5.50E-05 | 0.652983 |
| ATAD3C | 143.3302 | -1.44011 | 0.315065 | -4.57084 | 4.86E-06 | 5.52E-05 | 0.368539 |
| HTR2C | 194.5567 | -0.59449 | 0.130102 | -4.56939 | 4.89E-06 | 5.55E-05 | 0.662279 |
| TRPV6 | 67.82181 | -1.45793 | 0.319107 | -4.56878 | 4.91E-06 | 5.56E-05 | 0.364015 |
| ELOVL1 | 575.5158 | -0.54284 | 0.118823 | -4.5685 | 4.91E-06 | 5.56E-05 | 0.686417 |
| SEZ6 | 495.6243 | -1.09993 | 0.240763 | -4.56849 | 4.91E-06 | 5.56E-05 | 0.466541 |
| CGAS | 11.47544 | -3.97707 | 0.870613 | -4.56812 | 4.92E-06 | 5.56E-05 | 0.063501 |
| MEGF8 | 3430.548 | -0.43655 | 0.095596 | -4.56661 | 4.96E-06 | 5.59E-05 | 0.7389 |
| CFAP54 | 151.7667 | -0.74868 | 0.164036 | -4.5641 | 5.02E-06 | 5.65E-05 | 0.595149 |
| RDH16 | 53.39535 | -1.46896 | 0.321871 | -4.56383 | 5.02E-06 | 5.66E-05 | 0.361242 |
| LDHD | 274.2877 | -0.72333 | 0.158522 | -4.56299 | 5.04E-06 | 5.68E-05 | 0.605696 |
| MLPH | 207.3002 | -0.74011 | 0.162207 | -4.56277 | 5.05E-06 | 5.68E-05 | 0.598693 |
| RCE1 | 677.0276 | -0.67395 | 0.147721 | -4.56234 | 5.06E-06 | 5.68E-05 | 0.626787 |
| CORO1A | 194.8951 | -0.69226 | 0.151834 | -4.55933 | 5.13E-06 | 5.76E-05 | 0.618882 |
| PXDNL | 34.57327 | -1.68528 | 0.369749 | -4.5579 | 5.17E-06 | 5.79E-05 | 0.310943 |
| ZDHHC11 | 446.4019 | -0.98453 | 0.21603 | -4.5574 | 5.18E-06 | 5.80E-05 | 0.50539 |
| DLG4 | 1438.331 | -0.46516 | 0.102099 | -4.55597 | 5.21E-06 | 5.81E-05 | 0.724391 |
| DST | 4751.377 | -0.82956 | 0.182064 | -4.55643 | 5.20E-06 | 5.81E-05 | 0.5627 |

| | | | | | | | |
|---|---|---|---|---|---|---|---|
| GRIK5 | 1857.756 | -0.46961 | 0.103069 | -4.55623 | 5.21E-06 | 5.81E-05 | 0.722161 |
| SLC22A31 | 175.3434 | -1.0226 | 0.224468 | -4.55566 | 5.22E-06 | 5.81E-05 | 0.492229 |
| SYCP3 | 6.696915 | -4.6099 | 1.011912 | -4.55564 | 5.22E-06 | 5.81E-05 | 0.040953 |
| CRELD1 | 321.6585 | -0.7638 | 0.167666 | -4.55546 | 5.23E-06 | 5.82E-05 | 0.588945 |
| SLC39A1 | 6711.336 | -0.46038 | 0.101086 | -4.55434 | 5.25E-06 | 5.84E-05 | 0.726795 |
| COL4A5 | 2119.304 | -0.88176 | 0.193634 | -4.55373 | 5.27E-06 | 5.86E-05 | 0.542706 |
| FAM20A | 1438.951 | -0.83072 | 0.182626 | -4.54876 | 5.40E-06 | 5.97E-05 | 0.562248 |
| MTRNR2L3 | 53.40952 | -1.45662 | 0.320472 | -4.54522 | 5.49E-06 | 6.05E-05 | 0.364347 |
| CPAMD8 | 360.2947 | -0.50143 | 0.110425 | -4.5409 | 5.60E-06 | 6.16E-05 | 0.706406 |
| PI4KAP2 | 519.2351 | -0.76144 | 0.167746 | -4.53921 | 5.65E-06 | 6.21E-05 | 0.589909 |
| PLPPR4 | 131.8075 | -1.34229 | 0.295728 | -4.53891 | 5.65E-06 | 6.21E-05 | 0.394395 |
| CROCCP3 | 228.8703 | -0.94632 | 0.208696 | -4.53445 | 5.78E-06 | 6.33E-05 | 0.518955 |
| PCSK7 | 1459.799 | -0.58807 | 0.129728 | -4.53306 | 5.81E-06 | 6.37E-05 | 0.665234 |
| SLC30A1 | 989.2748 | -0.80635 | 0.178193 | -4.52516 | 6.03E-06 | 6.57E-05 | 0.571826 |
| HINFP | 673.0515 | -0.47447 | 0.104918 | -4.52231 | 6.12E-06 | 6.64E-05 | 0.71973 |
| ZNF135 | 9.659743 | -4.03413 | 0.892315 | -4.52097 | 6.16E-06 | 6.68E-05 | 0.061039 |
| PQLC1 | 411.5303 | -0.48324 | 0.106932 | -4.51907 | 6.21E-06 | 6.73E-05 | 0.715371 |
| ARHGEF16 | 670.185 | -0.5492 | 0.121586 | -4.51693 | 6.27E-06 | 6.79E-05 | 0.6834 |
| TENM2 | 88.19658 | -2.00958 | 0.444956 | -4.51636 | 6.29E-06 | 6.81E-05 | 0.248345 |
| PARP12 | 364.2627 | -0.56344 | 0.124783 | -4.51535 | 6.32E-06 | 6.83E-05 | 0.676687 |
| REG1A | 16.25416 | -2.49993 | 0.55369 | -4.51503 | 6.33E-06 | 6.83E-05 | 0.176786 |
| ATXN7L2 | 222.6041 | -0.7299 | 0.161723 | -4.51325 | 6.38E-06 | 6.89E-05 | 0.602947 |
| MSANTD2 | 373.7864 | -0.57902 | 0.128422 | -4.50872 | 6.52E-06 | 7.03E-05 | 0.669419 |
| HRH3 | 136.381 | -0.76901 | 0.170748 | -4.50375 | 6.68E-06 | 7.17E-05 | 0.586821 |
| TMEM198 | 235.7465 | -0.53476 | 0.118734 | -4.50387 | 6.67E-06 | 7.17E-05 | 0.690273 |
| P3H3 | 1115.695 | -0.6791 | 0.150943 | -4.49904 | 6.83E-06 | 7.32E-05 | 0.624555 |
| REEP2 | 1163.156 | -0.53595 | 0.119167 | -4.49742 | 6.88E-06 | 7.37E-05 | 0.689707 |
| ABCD1 | 145.6007 | -0.82097 | 0.182551 | -4.49722 | 6.88E-06 | 7.37E-05 | 0.56606 |
| CACNB1 | 333.7272 | -0.76971 | 0.171283 | -4.49378 | 7.00E-06 | 7.48E-05 | 0.586537 |
| RNF128 | 83.76641 | -0.92726 | 0.20638 | -4.49298 | 7.02E-06 | 7.50E-05 | 0.525855 |
| ATP2C2 | 175.1486 | -1.11131 | 0.247473 | -4.49065 | 7.10E-06 | 7.57E-05 | 0.462873 |
| ATP6V0B | 1732.125 | -0.68775 | 0.153208 | -4.48901 | 7.16E-06 | 7.62E-05 | 0.620819 |
| FAM24B | 85.9262 | -1.22887 | 0.273843 | -4.4875 | 7.21E-06 | 7.66E-05 | 0.426652 |
| FOXD3 | 332.4962 | -0.84615 | 0.188823 | -4.48116 | 7.42E-06 | 7.88E-05 | 0.556269 |
| PUS1 | 1083.053 | -0.45956 | 0.102614 | -4.47856 | 7.51E-06 | 7.96E-05 | 0.727206 |
| GGT7 | 911.3551 | -0.66304 | 0.148139 | -4.47584 | 7.61E-06 | 8.04E-05 | 0.631544 |
| DDR2 | 240.9946 | -1.41461 | 0.316514 | -4.46932 | 7.85E-06 | 8.27E-05 | 0.375112 |
| DTX3 | 1336.529 | -0.45997 | 0.102927 | -4.46891 | 7.86E-06 | 8.28E-05 | 0.727 |
| SPTAN1 | 9804.529 | -1.36104 | 0.304787 | -4.46555 | 7.99E-06 | 8.40E-05 | 0.3893 |
| MAPK12 | 628.9527 | -0.70345 | 0.157543 | -4.46511 | 8.00E-06 | 8.41E-05 | 0.614103 |
| P2RX2 | 42.96539 | -1.69786 | 0.380497 | -4.46221 | 8.11E-06 | 8.49E-05 | 0.308244 |
| S100A6 | 131.4825 | -0.94248 | 0.211213 | -4.46223 | 8.11E-06 | 8.49E-05 | 0.520338 |
| DOLK | 562.3092 | -0.58778 | 0.13173 | -4.46196 | 8.12E-06 | 8.49E-05 | 0.665368 |
| FAM156B | 401.5931 | -0.71697 | 0.16075 | -4.46014 | 8.19E-06 | 8.54E-05 | 0.608374 |

| | | | | | | | |
|---|---|---|---|---|---|---|---|
| MATN2 | 451.295 | -0.49879 | 0.111854 | -4.45928 | 8.22E-06 | 8.57E-05 | 0.707702 |
| ADAMTS8 | 352.225 | -0.56105 | 0.12588 | -4.45702 | 8.31E-06 | 8.66E-05 | 0.67781 |
| PCOLCE | 887.4724 | -0.57342 | 0.128761 | -4.45337 | 8.45E-06 | 8.79E-05 | 0.672021 |
| PILRA | 65.84959 | -1.34754 | 0.302667 | -4.45224 | 8.50E-06 | 8.82E-05 | 0.39296 |
| BCS1L | 455.7481 | -0.54572 | 0.122584 | -4.45179 | 8.52E-06 | 8.83E-05 | 0.685051 |
| PON3 | 4.614923 | -5.75688 | 1.29422 | -4.44814 | 8.66E-06 | 8.96E-05 | 0.018493 |
| PRKCSH | 7500.353 | -0.58885 | 0.132454 | -4.44569 | 8.76E-06 | 9.04E-05 | 0.664872 |
| NSUN5P2 | 668.7236 | -1.05489 | 0.237298 | -4.44543 | 8.77E-06 | 9.04E-05 | 0.481333 |
| PDGFRB | 291.7479 | -0.78389 | 0.176333 | -4.44553 | 8.77E-06 | 9.04E-05 | 0.580797 |
| DNAJB12 | 657.183 | -0.58301 | 0.131361 | -4.43824 | 9.07E-06 | 9.33E-05 | 0.667568 |
| NT5C | 845.1216 | -0.54792 | 0.123548 | -4.43489 | 9.21E-06 | 9.46E-05 | 0.684004 |
| CYP2U1 | 645.9443 | -0.42804 | 0.096662 | -4.42819 | 9.50E-06 | 9.70E-05 | 0.743273 |
| TBC1D24 | 636.7578 | -0.45415 | 0.102616 | -4.4257 | 9.61E-06 | 9.80E-05 | 0.729942 |
| SLC48A1 | 343.2393 | -0.62462 | 0.141243 | -4.4223 | 9.77E-06 | 9.91E-05 | 0.648591 |
| PHKG2 | 831.9665 | -0.43655 | 0.098744 | -4.42107 | 9.82E-06 | 9.96E-05 | 0.738897 |
| TESK1 | 331.0029 | -0.59072 | 0.133637 | -4.42035 | 9.85E-06 | 9.99E-05 | 0.66401 |
| PCDHB4 | 48.85705 | -1.52405 | 0.344831 | -4.41969 | 9.88E-06 | 0.0001 | 0.347709 |
| ASIC3 | 91.25661 | -2.0557 | 0.465207 | -4.41889 | 9.92E-06 | 0.0001 | 0.240532 |
| ERP29 | 6167.643 | -0.52573 | 0.119077 | -4.41502 | 1.01E-05 | 0.000102 | 0.69461 |
| TTC25 | 130.8706 | -0.88165 | 0.199721 | -4.41441 | 1.01E-05 | 0.000102 | 0.542746 |
| DDX51 | 860.3729 | -1.09893 | 0.249213 | -4.40961 | 1.04E-05 | 0.000104 | 0.466862 |
| TMBIM1 | 1003.852 | -0.49183 | 0.111553 | -4.40895 | 1.04E-05 | 0.000104 | 0.711121 |
| NAPRT | 911.2714 | -0.85034 | 0.192876 | -4.40874 | 1.04E-05 | 0.000104 | 0.554654 |
| QRICH2 | 112.2197 | -1.17236 | 0.266056 | -4.40642 | 1.05E-05 | 0.000105 | 0.443696 |
| CRYBB2P1 | 328.531 | -0.5059 | 0.114987 | -4.39963 | 1.08E-05 | 0.000108 | 0.704222 |
| BTBD6 | 305.5378 | -0.468 | 0.106455 | -4.39625 | 1.10E-05 | 0.00011 | 0.722965 |
| THNSL2 | 195.3021 | -0.70452 | 0.160411 | -4.39195 | 1.12E-05 | 0.000112 | 0.613647 |
| TLCD1 | 425.9687 | -0.65627 | 0.149446 | -4.39132 | 1.13E-05 | 0.000112 | 0.634518 |
| CYP26C1 | 26.20047 | -2.44969 | 0.558144 | -4.389 | 1.14E-05 | 0.000113 | 0.18305 |
| AC006254 | 55.10514 | -1.42378 | 0.324462 | -4.3881 | 1.14E-05 | 0.000113 | 0.372736 |
| TGFB1 | 232.5829 | -0.57724 | 0.131572 | -4.38723 | 1.15E-05 | 0.000114 | 0.670245 |
| SOCS3 | 316.2985 | -0.64712 | 0.147543 | -4.38595 | 1.15E-05 | 0.000114 | 0.638555 |
| SGCE | 620.7719 | -0.44429 | 0.101316 | -4.38522 | 1.16E-05 | 0.000115 | 0.734945 |
| MCOLN1 | 409.8103 | -0.91038 | 0.207654 | -4.38412 | 1.16E-05 | 0.000115 | 0.532045 |
| WASHC1 | 264.9089 | -0.83555 | 0.190887 | -4.37723 | 1.20E-05 | 0.000118 | 0.560368 |
| AC003665 | 214.3542 | -0.87512 | 0.199966 | -4.37633 | 1.21E-05 | 0.000119 | 0.545209 |
| KIFC2 | 723.0324 | -1.09371 | 0.249957 | -4.3756 | 1.21E-05 | 0.000119 | 0.468554 |
| CHMP1B2I | 154.1738 | -0.65939 | 0.150807 | -4.37242 | 1.23E-05 | 0.00012 | 0.633145 |
| FAM234A | 3547.571 | -0.4429 | 0.101306 | -4.37193 | 1.23E-05 | 0.000121 | 0.735652 |
| PIGB | 176.9189 | -0.70659 | 0.161637 | -4.37149 | 1.23E-05 | 0.000121 | 0.612765 |
| TOGARAM: | 120.9379 | -1.20105 | 0.274812 | -4.37046 | 1.24E-05 | 0.000121 | 0.434958 |
| SLC19A1 | 903.3067 | -0.59338 | 0.135776 | -4.37029 | 1.24E-05 | 0.000121 | 0.662789 |
| PHYHD1 | 271.2029 | -0.6002 | 0.137349 | -4.36991 | 1.24E-05 | 0.000122 | 0.659661 |
| ABCD4 | 1719.406 | -0.55557 | 0.127141 | -4.36977 | 1.24E-05 | 0.000122 | 0.680386 |

| | | | | | | | |
|---|---|---|---|---|---|---|---|
| SEC16A | 2500.595 | -0.47122 | 0.107847 | -4.36937 | 1.25E-05 | 0.000122 | 0.721352 |
| GIGYF1 | 4110.34 | -0.75634 | 0.173118 | -4.36893 | 1.25E-05 | 0.000122 | 0.591997 |
| SELENOO | 504.4195 | -0.79975 | 0.18311 | -4.36759 | 1.26E-05 | 0.000122 | 0.574448 |
| UNKL | 641.0489 | -0.61973 | 0.14192 | -4.36674 | 1.26E-05 | 0.000123 | 0.650793 |
| IL34 | 97.61416 | -1.20611 | 0.276366 | -4.36417 | 1.28E-05 | 0.000124 | 0.433437 |
| RASEF | 748.0693 | -0.46905 | 0.10749 | -4.36367 | 1.28E-05 | 0.000124 | 0.72244 |
| PI4KAP1 | 699.1808 | -0.84886 | 0.194559 | -4.36302 | 1.28E-05 | 0.000125 | 0.555221 |
| FAM86B1 | 1143.491 | -0.43335 | 0.099337 | -4.36244 | 1.29E-05 | 0.000125 | 0.740539 |
| KMT2B | 2106.55 | -0.7244 | 0.166241 | -4.35753 | 1.32E-05 | 0.000127 | 0.605249 |
| PLXNB2 | 3880.305 | -0.72116 | 0.165625 | -4.35418 | 1.34E-05 | 0.000129 | 0.60661 |
| TMEM179E | 402.4369 | -0.68452 | 0.157296 | -4.35182 | 1.35E-05 | 0.00013 | 0.622211 |
| ICAM3 | 217.6108 | -0.62966 | 0.144879 | -4.34611 | 1.39E-05 | 0.000134 | 0.64633 |
| FCGRT | 705.0034 | -0.71674 | 0.164975 | -4.34455 | 1.40E-05 | 0.000135 | 0.60847 |
| AARSD1 | 767.6026 | -0.44006 | 0.101409 | -4.33946 | 1.43E-05 | 0.000137 | 0.737104 |
| POFUT2 | 1198.787 | -0.54797 | 0.126327 | -4.33767 | 1.44E-05 | 0.000138 | 0.683983 |
| DPP7 | 702.6697 | -0.76364 | 0.176138 | -4.33546 | 1.45E-05 | 0.00014 | 0.58901 |
| COMTD1 | 343.6705 | -0.71759 | 0.165594 | -4.33341 | 1.47E-05 | 0.00014 | 0.608114 |
| NDOR1 | 631.4366 | -0.51758 | 0.119466 | -4.33245 | 1.47E-05 | 0.000141 | 0.698542 |
| EXOC3L2 | 68.5212 | -0.98002 | 0.226337 | -4.32992 | 1.49E-05 | 0.000142 | 0.506972 |
| PCDHB5 | 18.95103 | -1.99672 | 0.46131 | -4.32836 | 1.50E-05 | 0.000143 | 0.250569 |
| MYBPC2 | 156.2428 | -0.90361 | 0.208805 | -4.32752 | 1.51E-05 | 0.000144 | 0.534548 |
| KCP | 197.7929 | -0.77886 | 0.180019 | -4.32652 | 1.51E-05 | 0.000144 | 0.582829 |
| CAMSAP3 | 1041.604 | -0.89351 | 0.206604 | -4.32477 | 1.53E-05 | 0.000145 | 0.538302 |
| LCAT | 114.6179 | -1.15962 | 0.268491 | -4.31902 | 1.57E-05 | 0.000149 | 0.447632 |
| ACVR2A | 476.3427 | -0.68275 | 0.158117 | -4.31798 | 1.57E-05 | 0.00015 | 0.622978 |
| PRKAR1B | 997.3311 | -0.50563 | 0.117231 | -4.31308 | 1.61E-05 | 0.000153 | 0.704354 |
| JDP2 | 703.7127 | -0.42225 | 0.097912 | -4.31256 | 1.61E-05 | 0.000153 | 0.746258 |
| CALCA | 85.72562 | -1.32827 | 0.30814 | -4.3106 | 1.63E-05 | 0.000154 | 0.398246 |
| OS9 | 3233.25 | -0.55147 | 0.128066 | -4.30617 | 1.66E-05 | 0.000157 | 0.682323 |
| CD151 | 2101.628 | -0.63834 | 0.148264 | -4.30545 | 1.67E-05 | 0.000157 | 0.64245 |
| BCAR1 | 1017.743 | -0.48891 | 0.113854 | -4.29419 | 1.75E-05 | 0.000165 | 0.712562 |
| RNASE10 | 40.19349 | -1.53192 | 0.35712 | -4.28965 | 1.79E-05 | 0.000168 | 0.345816 |
| GHDC | 252.1321 | -0.7418 | 0.172972 | -4.28854 | 1.80E-05 | 0.000168 | 0.597993 |
| P4HB | 11122.98 | -0.50271 | 0.11727 | -4.28676 | 1.81E-05 | 0.000169 | 0.705781 |
| LGALS3BP | 2022.874 | -0.65648 | 0.153164 | -4.28613 | 1.82E-05 | 0.00017 | 0.634423 |
| SORBS3 | 1295.648 | -0.58195 | 0.135921 | -4.28151 | 1.86E-05 | 0.000173 | 0.668061 |
| OR7E122P | 57.39718 | -1.18052 | 0.276067 | -4.2762 | 1.90E-05 | 0.000177 | 0.441193 |
| PRORSD1F | 62.97926 | -1.1619 | 0.271722 | -4.27606 | 1.90E-05 | 0.000177 | 0.446925 |
| EMC10 | 3281.903 | -0.45059 | 0.105445 | -4.27319 | 1.93E-05 | 0.000178 | 0.731745 |
| MOV10 | 1113.927 | -0.64912 | 0.151954 | -4.27181 | 1.94E-05 | 0.000179 | 0.63767 |
| DVL1 | 2093.375 | -0.54387 | 0.127324 | -4.2715 | 1.94E-05 | 0.00018 | 0.685931 |
| ARHGEF10 | 1301.726 | -0.44921 | 0.1052 | -4.27006 | 1.95E-05 | 0.00018 | 0.732443 |
| MOXD1 | 260.5791 | -0.64744 | 0.151735 | -4.26695 | 1.98E-05 | 0.000183 | 0.638411 |
| SLC22A20 | 223.2945 | -0.83688 | 0.196519 | -4.2585 | 2.06E-05 | 0.000189 | 0.559854 |

| | | | | | | | |
|---|---|---|---|---|---|---|---|
| B3GNT7 | 2257.442 | -0.41903 | 0.098428 | -4.25727 | 2.07E-05 | 0.00019 | 0.747926 |
| FAM83E | 70.93607 | -1.04636 | 0.245969 | -4.25405 | 2.10E-05 | 0.000193 | 0.484187 |
| PNMA6A | 181.0006 | -0.56198 | 0.13215 | -4.2526 | 2.11E-05 | 0.000194 | 0.67737 |
| CFAP410 | 527.5084 | -0.56131 | 0.132024 | -4.25157 | 2.12E-05 | 0.000195 | 0.677686 |
| OBSCN | 2463.636 | -0.93263 | 0.219632 | -4.24632 | 2.17E-05 | 0.000199 | 0.523903 |
| FSD1 | 1696.363 | -0.50611 | 0.119213 | -4.24538 | 2.18E-05 | 0.000199 | 0.704121 |
| IFT172 | 511.61 | -0.99622 | 0.234808 | -4.24272 | 2.21E-05 | 0.000201 | 0.50131 |
| ZNF787 | 1263.536 | -0.44347 | 0.104561 | -4.24132 | 2.22E-05 | 0.000203 | 0.735362 |
| ECEL1P1 | 706.9806 | -0.7363 | 0.173624 | -4.24076 | 2.23E-05 | 0.000203 | 0.600277 |
| CYB5R1 | 366.7917 | -0.62825 | 0.148216 | -4.23873 | 2.25E-05 | 0.000204 | 0.646962 |
| YBX2 | 802.6128 | -0.66786 | 0.15756 | -4.2388 | 2.25E-05 | 0.000204 | 0.629438 |
| CFLAR | 387.74 | -1.18599 | 0.279888 | -4.23738 | 2.26E-05 | 0.000205 | 0.439523 |
| FAM160B2 | 500.9227 | -0.75438 | 0.178046 | -4.23701 | 2.27E-05 | 0.000206 | 0.5928 |
| TMED3 | 1253.888 | -0.41847 | 0.099028 | -4.22576 | 2.38E-05 | 0.000215 | 0.748218 |
| CD40 | 177.331 | -1.03136 | 0.244087 | -4.22536 | 2.39E-05 | 0.000215 | 0.489249 |
| CYB561 | 998.6185 | -0.49556 | 0.117314 | -4.22424 | 2.40E-05 | 0.000216 | 0.709286 |
| LHX4 | 227.1087 | -0.83315 | 0.197222 | -4.22443 | 2.40E-05 | 0.000216 | 0.561302 |
| RAB40A | 169.115 | -0.67108 | 0.158859 | -4.22439 | 2.40E-05 | 0.000216 | 0.628036 |
| KIAA0895L | 727.6651 | -1.11961 | 0.265127 | -4.22292 | 2.41E-05 | 0.000217 | 0.460218 |
| PTPRU | 2071.173 | -0.49502 | 0.117246 | -4.22203 | 2.42E-05 | 0.000217 | 0.709554 |
| ARHGAP33 | 527.232 | -0.78545 | 0.186067 | -4.2213 | 2.43E-05 | 0.000218 | 0.580173 |
| PIGT | 2157.627 | -0.53089 | 0.125846 | -4.21859 | 2.46E-05 | 0.00022 | 0.692127 |
| PRPF40B | 808.0072 | -0.65355 | 0.154918 | -4.21867 | 2.46E-05 | 0.00022 | 0.635714 |
| ADCY3 | 1041.999 | -0.61496 | 0.145958 | -4.21329 | 2.52E-05 | 0.000225 | 0.652946 |
| GOLGA6L9 | 500.0619 | -0.87094 | 0.206758 | -4.21235 | 2.53E-05 | 0.000226 | 0.546791 |
| SAMD10 | 107.2417 | -0.93902 | 0.222963 | -4.21155 | 2.54E-05 | 0.000226 | 0.521588 |
| TFPI2 | 118.9357 | -0.72941 | 0.173207 | -4.2112 | 2.54E-05 | 0.000227 | 0.60315 |
| AC011448 | 74.73239 | -1.46914 | 0.34891 | -4.21065 | 2.55E-05 | 0.000227 | 0.361198 |
| ICOSLG | 313.2395 | -0.57629 | 0.136949 | -4.20805 | 2.58E-05 | 0.000229 | 0.670686 |
| RPN1 | 5893.132 | -0.41939 | 0.099676 | -4.20758 | 2.58E-05 | 0.00023 | 0.747739 |
| TBC1D3L | 512.2051 | -0.98464 | 0.23407 | -4.20661 | 2.59E-05 | 0.000231 | 0.505351 |
| MST1 | 194.304 | -1.1708 | 0.278691 | -4.20106 | 2.66E-05 | 0.000236 | 0.444176 |
| SIT1 | 77.07207 | -1.04349 | 0.248427 | -4.2004 | 2.66E-05 | 0.000236 | 0.485152 |
| MGAT4B | 2450.476 | -0.44392 | 0.105691 | -4.20014 | 2.67E-05 | 0.000236 | 0.735135 |
| PIDD1 | 874.9148 | -1.09324 | 0.260382 | -4.19858 | 2.69E-05 | 0.000238 | 0.468709 |
| KCNMA1 | 201.0054 | -1.00178 | 0.238912 | -4.1931 | 2.75E-05 | 0.000243 | 0.499382 |
| PALM3 | 801.2565 | -0.46305 | 0.1105 | -4.19054 | 2.78E-05 | 0.000246 | 0.725449 |
| CASR | 278.122 | -0.6689 | 0.159652 | -4.18975 | 2.79E-05 | 0.000246 | 0.628985 |
| PLA2G4B | 237.6697 | -1.25219 | 0.299346 | -4.18309 | 2.88E-05 | 0.000253 | 0.41981 |
| BDP1 | 302.194 | -0.78967 | 0.188855 | -4.18137 | 2.90E-05 | 0.000255 | 0.578476 |
| CCS | 821.8221 | -0.50537 | 0.120868 | -4.18115 | 2.90E-05 | 0.000255 | 0.704482 |
| TUBG2 | 466.0868 | -0.53222 | 0.127316 | -4.18033 | 2.91E-05 | 0.000256 | 0.691489 |
| ADGRD1 | 112.1511 | -1.41059 | 0.337528 | -4.17919 | 2.93E-05 | 0.000257 | 0.376157 |
| C1orf159 | 366.7571 | -0.74283 | 0.177897 | -4.17563 | 2.97E-05 | 0.00026 | 0.597565 |

| | | | | | | | |
|---|---|---|---|---|---|---|---|
| AC022400 | 202.2068 | -1.44311 | 0.345808 | -4.17314 | 3.00E-05 | 0.000262 | 0.367774 |
| DPEP1 | 127.0126 | -0.69498 | 0.166586 | -4.17189 | 3.02E-05 | 0.000263 | 0.617717 |
| DIAPH1 | 4266.386 | -0.7058 | 0.16922 | -4.17093 | 3.03E-05 | 0.000264 | 0.613101 |
| TMEM129 | 1316.481 | -0.48073 | 0.115361 | -4.16719 | 3.08E-05 | 0.000268 | 0.716614 |
| INTS1 | 4855.9 | -0.61468 | 0.147581 | -4.16504 | 3.11E-05 | 0.00027 | 0.653073 |
| NOMO3 | 65.13841 | -1.59662 | 0.383363 | -4.16476 | 3.12E-05 | 0.000271 | 0.330652 |
| AC099489 | 343.596 | -0.8525 | 0.205041 | -4.15772 | 3.21E-05 | 0.000278 | 0.553824 |
| AC018638 | 165.4221 | -0.62264 | 0.149902 | -4.15363 | 3.27E-05 | 0.000283 | 0.649483 |
| GNB3 | 201.7605 | -1.30773 | 0.314996 | -4.15159 | 3.30E-05 | 0.000285 | 0.403955 |
| HEXD | 619.0897 | -0.87474 | 0.210809 | -4.14946 | 3.33E-05 | 0.000288 | 0.545351 |
| DOLPP1 | 966.7297 | -0.53339 | 0.128591 | -4.14798 | 3.35E-05 | 0.000289 | 0.690927 |
| SUPT5H | 3731.843 | -0.45981 | 0.110931 | -4.14497 | 3.40E-05 | 0.000293 | 0.727084 |
| CTSH | 558.222 | -0.68409 | 0.165167 | -4.14179 | 3.45E-05 | 0.000296 | 0.6224 |
| CHST13 | 86.005 | -0.8089 | 0.195345 | -4.14088 | 3.46E-05 | 0.000297 | 0.570817 |
| GNPTG | 699.4764 | -0.6683 | 0.161443 | -4.13955 | 3.48E-05 | 0.000299 | 0.629247 |
| SDHAP3 | 262.5739 | -1.02039 | 0.246577 | -4.13821 | 3.50E-05 | 0.0003 | 0.492984 |
| CD320 | 1363.518 | -0.72585 | 0.175426 | -4.13764 | 3.51E-05 | 0.000301 | 0.604641 |
| VSIG10 | 1502.512 | -0.56984 | 0.137903 | -4.13219 | 3.59E-05 | 0.000307 | 0.67369 |
| AGPAT2 | 591.2158 | -0.55096 | 0.133383 | -4.13065 | 3.62E-05 | 0.000309 | 0.682566 |
| PGAP3 | 594.3613 | -0.58034 | 0.140622 | -4.12694 | 3.68E-05 | 0.000314 | 0.668807 |
| ADRA2C | 512.969 | -0.52442 | 0.127179 | -4.12348 | 3.73E-05 | 0.000318 | 0.695239 |
| ERVK3-1 | 854.7769 | -0.42677 | 0.1035 | -4.12337 | 3.73E-05 | 0.000318 | 0.743927 |
| INPPL1 | 3294.622 | -0.64225 | 0.15577 | -4.12305 | 3.74E-05 | 0.000318 | 0.640715 |
| GALNT6 | 416.6781 | -0.6946 | 0.168487 | -4.1226 | 3.75E-05 | 0.000319 | 0.617879 |
| AGAP6 | 410.1954 | -0.75448 | 0.183039 | -4.12196 | 3.76E-05 | 0.000319 | 0.59276 |
| IL9RP3 | 14.18132 | -3.01515 | 0.731837 | -4.11997 | 3.79E-05 | 0.000322 | 0.123695 |
| CPXM1 | 1639.638 | -0.63459 | 0.154128 | -4.11729 | 3.83E-05 | 0.000325 | 0.644124 |
| ATP6V0C | 3916.164 | -0.53976 | 0.131119 | -4.11654 | 3.85E-05 | 0.000326 | 0.687887 |
| GALNT3 | 645.1282 | -0.58392 | 0.141971 | -4.11293 | 3.91E-05 | 0.00033 | 0.667151 |
| C19orf24 | 463.7803 | -0.48474 | 0.117872 | -4.11245 | 3.91E-05 | 0.000331 | 0.714625 |
| CYP3A5 | 172.7823 | -0.65938 | 0.160397 | -4.11092 | 3.94E-05 | 0.000333 | 0.633151 |
| BSG | 20211.44 | -0.64346 | 0.156588 | -4.10927 | 3.97E-05 | 0.000334 | 0.640175 |
| ELMO3 | 339.9172 | -0.89758 | 0.218448 | -4.1089 | 3.98E-05 | 0.000335 | 0.536786 |
| CEMP1 | 129.1407 | -1.9209 | 0.467751 | -4.10666 | 4.01E-05 | 0.000337 | 0.264091 |
| COL20A1 | 41.14337 | -2.63062 | 0.640606 | -4.10646 | 4.02E-05 | 0.000337 | 0.161474 |
| WASH2P | 372.5894 | -0.72175 | 0.17583 | -4.10484 | 4.05E-05 | 0.000339 | 0.60636 |
| OPLAH | 1003.935 | -0.73788 | 0.179848 | -4.10279 | 4.08E-05 | 0.000342 | 0.599621 |
| MBD6 | 913.3563 | -0.66125 | 0.16125 | -4.10076 | 4.12E-05 | 0.000345 | 0.63233 |
| HTRA3 | 165.6028 | -0.7228 | 0.176311 | -4.09958 | 4.14E-05 | 0.000346 | 0.60592 |
| GGA1 | 1230.771 | -0.46361 | 0.113142 | -4.09758 | 4.17E-05 | 0.000349 | 0.725171 |
| PDLIM7 | 2149.424 | -0.81403 | 0.198753 | -4.0957 | 4.21E-05 | 0.000352 | 0.568789 |
| ZNF22 | 956.9502 | -0.44959 | 0.109924 | -4.08998 | 4.31E-05 | 0.00036 | 0.732253 |
| STK36 | 854.8722 | -0.75715 | 0.185149 | -4.08939 | 4.33E-05 | 0.00036 | 0.591665 |
| NFASC | 410.4703 | -0.5559 | 0.135952 | -4.08896 | 4.33E-05 | 0.000361 | 0.680232 |

| | | | | | | | |
|---|---|---|---|---|---|---|---|
| TECTA | 50.40645 | -1.23122 | 0.301659 | -4.0815 | 4.47E-05 | 0.000372 | 0.425957 |
| HCN4 | 976.325 | -0.47467 | 0.116373 | -4.07888 | 4.53E-05 | 0.000376 | 0.71963 |
| ASPSCR1 | 901.4942 | -0.69969 | 0.17158 | -4.07793 | 4.54E-05 | 0.000377 | 0.615704 |
| NRBP2 | 505.2999 | -0.92363 | 0.226521 | -4.07748 | 4.55E-05 | 0.000378 | 0.52718 |
| CAPN15 | 2017.005 | -0.6447 | 0.158168 | -4.07605 | 4.58E-05 | 0.00038 | 0.639626 |
| NPDC1 | 1105.651 | -0.98669 | 0.24221 | -4.07371 | 4.63E-05 | 0.000383 | 0.504633 |
| CTSD | 7183.966 | -0.54543 | 0.133936 | -4.07234 | 4.65E-05 | 0.000385 | 0.685186 |
| ALDH1A3 | 187.5977 | -0.68536 | 0.16841 | -4.0696 | 4.71E-05 | 0.000389 | 0.621849 |
| ADGRF1 | 11.07863 | -3.09099 | 0.759929 | -4.06747 | 4.75E-05 | 0.000392 | 0.11736 |
| C19orf47 | 366.644 | -0.70341 | 0.17324 | -4.06032 | 4.90E-05 | 0.000403 | 0.614119 |
| ARHGAP4( | 94.44146 | -1.10176 | 0.271608 | -4.05642 | 4.98E-05 | 0.000408 | 0.465948 |
| TMEM130 | 220.2746 | -0.9064 | 0.223488 | -4.0557 | 5.00E-05 | 0.000409 | 0.533515 |
| PARP6 | 698.6541 | -0.55342 | 0.136592 | -4.05158 | 5.09E-05 | 0.000416 | 0.681405 |
| LRP2BP | 60.22555 | -1.44215 | 0.356428 | -4.04613 | 5.21E-05 | 0.000425 | 0.368018 |
| C3orf80 | 34.2552 | -1.36833 | 0.338223 | -4.04564 | 5.22E-05 | 0.000426 | 0.38734 |
| CERS4 | 1768.709 | -0.55464 | 0.137201 | -4.04255 | 5.29E-05 | 0.000431 | 0.680826 |
| CIDECP1 | 153.9536 | -0.85021 | 0.210325 | -4.04236 | 5.29E-05 | 0.000431 | 0.554705 |
| UTF1 | 105.1144 | -0.89688 | 0.221958 | -4.04075 | 5.33E-05 | 0.000434 | 0.537048 |
| C1QTNF12 | 62.77647 | -1.86701 | 0.462217 | -4.03925 | 5.36E-05 | 0.000436 | 0.274141 |
| SFI1 | 662.8263 | -0.73191 | 0.18121 | -4.039 | 5.37E-05 | 0.000436 | 0.602107 |
| PLEKHG4 | 464.9707 | -0.57702 | 0.142942 | -4.03673 | 5.42E-05 | 0.00044 | 0.670348 |
| TSPOAP1 | 310.7927 | -1.02398 | 0.253748 | -4.03541 | 5.45E-05 | 0.000442 | 0.491759 |
| ELN | 293.091 | -0.52407 | 0.12989 | -4.03476 | 5.47E-05 | 0.000443 | 0.695405 |
| KLHL17 | 667.9826 | -1.01402 | 0.251402 | -4.03344 | 5.50E-05 | 0.000445 | 0.495166 |
| ARMC5 | 313.2587 | -0.5692 | 0.141163 | -4.03226 | 5.52E-05 | 0.000447 | 0.673988 |
| TMUB1 | 561.3879 | -0.54356 | 0.134854 | -4.03071 | 5.56E-05 | 0.00045 | 0.686076 |
| NTRK3 | 209.2286 | -0.82349 | 0.204351 | -4.02979 | 5.58E-05 | 0.000451 | 0.565073 |
| GRIP2 | 194.3127 | -0.76055 | 0.188772 | -4.02892 | 5.60E-05 | 0.000452 | 0.590272 |
| CDK5RAP3 | 2436.941 | -0.70483 | 0.175141 | -4.02438 | 5.71E-05 | 0.00046 | 0.613513 |
| 9-Mar | 233.4605 | -0.71094 | 0.176845 | -4.02011 | 5.82E-05 | 0.000468 | 0.610924 |
| LAMB3 | 84.46691 | -0.874 | 0.217512 | -4.01817 | 5.87E-05 | 0.000471 | 0.545632 |
| MLXIPL | 626.0243 | -0.93907 | 0.233873 | -4.0153 | 5.94E-05 | 0.000476 | 0.52157 |
| MXRA8 | 266.4158 | -0.63593 | 0.158454 | -4.01332 | 5.99E-05 | 0.000478 | 0.643527 |
| TMEM74B | 183.3366 | -1.11288 | 0.27737 | -4.01225 | 6.01E-05 | 0.00048 | 0.462371 |
| WASH3P | 729.5483 | -0.63013 | 0.1572 | -4.00844 | 6.11E-05 | 0.000487 | 0.646119 |
| SPTBN5 | 86.21153 | -1.70109 | 0.424622 | -4.00613 | 6.17E-05 | 0.000491 | 0.307554 |
| PSEN2 | 473.2705 | -0.66552 | 0.166217 | -4.00391 | 6.23E-05 | 0.000495 | 0.630463 |
| POLRMT | 2797.489 | -0.52155 | 0.130266 | -4.00372 | 6.24E-05 | 0.000495 | 0.696623 |
| DNER | 213.6928 | -0.57198 | 0.142914 | -4.00222 | 6.27E-05 | 0.000498 | 0.672695 |
| CHRD | 57.68908 | -1.05522 | 0.263671 | -4.00202 | 6.28E-05 | 0.000498 | 0.481225 |
| SLC39A11 | 344.0601 | -0.77384 | 0.193515 | -3.99887 | 6.36E-05 | 0.000504 | 0.584858 |
| CFP | 33.25299 | -2.07738 | 0.520156 | -3.99377 | 6.50E-05 | 0.000514 | 0.236944 |
| MFSD6 | 162.0783 | -0.69215 | 0.173412 | -3.99136 | 6.57E-05 | 0.000518 | 0.618931 |
| MIIP | 1267.16 | -0.56185 | 0.140776 | -3.99109 | 6.58E-05 | 0.000519 | 0.677434 |

| | | | | | | | |
|---|---|---|---|---|---|---|---|
| RGS11 | 82.60931 | -1.19744 | 0.300085 | -3.99034 | 6.60E-05 | 0.00052 | 0.436047 |
| TCIRG1 | 528.052 | -1.24043 | 0.311374 | -3.98373 | 6.78E-05 | 0.000533 | 0.423247 |
| MICALL2 | 331.2776 | -1.38568 | 0.347884 | -3.98316 | 6.80E-05 | 0.000534 | 0.38271 |
| AP3B2 | 549.5973 | -0.46614 | 0.117174 | -3.97824 | 6.94E-05 | 0.000544 | 0.723897 |
| PDGFRL | 115.2444 | -0.75797 | 0.190564 | -3.9775 | 6.96E-05 | 0.000546 | 0.591329 |
| ZGLP1 | 101.6362 | -1.50162 | 0.377722 | -3.97548 | 7.02E-05 | 0.00055 | 0.353155 |
| ICAM1 | 354.9086 | -0.55967 | 0.140826 | -3.9742 | 7.06E-05 | 0.000552 | 0.678457 |
| AL445238. | 3.553825 | -5.38734 | 1.356018 | -3.97291 | 7.10E-05 | 0.000555 | 0.023892 |
| LTBP2 | 94.52661 | -0.93259 | 0.234908 | -3.97002 | 7.19E-05 | 0.00056 | 0.523918 |
| EGFL6 | 150.2008 | -0.90568 | 0.228207 | -3.96866 | 7.23E-05 | 0.000563 | 0.533783 |
| SRRM2 | 20496.57 | -0.71286 | 0.179619 | -3.96873 | 7.23E-05 | 0.000563 | 0.61011 |
| PJVK | 49.04548 | -1.17248 | 0.295501 | -3.96778 | 7.25E-05 | 0.000565 | 0.443657 |
| UPP1 | 1349.274 | -0.62662 | 0.158154 | -3.96208 | 7.43E-05 | 0.000577 | 0.647693 |
| CD34 | 45.70441 | -1.33364 | 0.336655 | -3.96143 | 7.45E-05 | 0.000578 | 0.396767 |
| SIRT7 | 352.0243 | -0.76088 | 0.19212 | -3.96045 | 7.48E-05 | 0.00058 | 0.590137 |
| PYCARD | 470.0212 | -0.48243 | 0.12193 | -3.9566 | 7.60E-05 | 0.000588 | 0.715771 |
| NLRP10 | 18.02652 | -2.78583 | 0.704349 | -3.95519 | 7.65E-05 | 0.000591 | 0.145005 |
| SLC49A4 | 377.648 | -0.48527 | 0.122717 | -3.9544 | 7.67E-05 | 0.000592 | 0.714363 |
| CALCB | 144.4879 | -0.71594 | 0.181076 | -3.9538 | 7.69E-05 | 0.000593 | 0.60881 |
| MTATP6P1 | 12182.01 | -0.83172 | 0.210432 | -3.95243 | 7.74E-05 | 0.000596 | 0.561861 |
| CXCL16 | 87.98293 | -0.95174 | 0.240955 | -3.94986 | 7.82E-05 | 0.000601 | 0.517009 |
| SLC24A3 | 194.2864 | -0.66255 | 0.167776 | -3.949 | 7.85E-05 | 0.000602 | 0.631762 |
| ITGA10 | 16.53021 | -1.91376 | 0.48519 | -3.94436 | 8.00E-05 | 0.000613 | 0.265399 |
| TSNARE1 | 375.3416 | -0.88255 | 0.223963 | -3.94064 | 8.13E-05 | 0.000621 | 0.542406 |
| AC139768 | 275.4179 | -0.77051 | 0.195708 | -3.93702 | 8.25E-05 | 0.000629 | 0.586211 |
| TUBB8 | 18.30371 | -1.8083 | 0.459373 | -3.93646 | 8.27E-05 | 0.00063 | 0.285527 |
| ZNF652 | 842.042 | -0.43651 | 0.110893 | -3.93635 | 8.27E-05 | 0.00063 | 0.738919 |
| ATAD3B | 1377.019 | -0.75871 | 0.192804 | -3.93515 | 8.31E-05 | 0.000632 | 0.591023 |
| TLL2 | 115.8868 | -0.76798 | 0.195226 | -3.93383 | 8.36E-05 | 0.000636 | 0.587238 |
| PCDHB15 | 171.092 | -0.61589 | 0.156676 | -3.931 | 8.46E-05 | 0.000642 | 0.652526 |
| ZDHHC24 | 534.9725 | -0.66212 | 0.168475 | -3.93006 | 8.49E-05 | 0.000644 | 0.63195 |
| GOLGA2P5 | 385.4524 | -0.67884 | 0.172769 | -3.92915 | 8.52E-05 | 0.000647 | 0.624669 |
| MMP17 | 763.1892 | -0.47201 | 0.120198 | -3.92694 | 8.60E-05 | 0.000652 | 0.720959 |
| VN1R51P | 36.72568 | -1.41303 | 0.359866 | -3.92655 | 8.62E-05 | 0.000653 | 0.375522 |
| RASSF1 | 328.8479 | -0.49234 | 0.125397 | -3.92626 | 8.63E-05 | 0.000653 | 0.71087 |
| P2RX5 | 169.583 | -0.70379 | 0.179258 | -3.92615 | 8.63E-05 | 0.000653 | 0.613956 |
| PQLC2L | 6.710669 | -3.78545 | 0.964266 | -3.92573 | 8.65E-05 | 0.000654 | 0.072521 |
| ALG9 | 695.0126 | -0.47545 | 0.121118 | -3.9255 | 8.66E-05 | 0.000654 | 0.719243 |
| CXCR2 | 121.2286 | -1.08525 | 0.276505 | -3.92487 | 8.68E-05 | 0.000655 | 0.471312 |
| MAPK15 | 154.1779 | -1.44995 | 0.369428 | -3.92485 | 8.68E-05 | 0.000655 | 0.366034 |
| THSD4 | 215.2868 | -1.07622 | 0.274237 | -3.92442 | 8.69E-05 | 0.000656 | 0.47427 |
| DBN1 | 10822.98 | -0.67459 | 0.172045 | -3.92098 | 8.82E-05 | 0.000665 | 0.626512 |
| CARNS1 | 73.31505 | -1.20946 | 0.308653 | -3.91853 | 8.91E-05 | 0.000671 | 0.432429 |
| CA5BP1 | 435.0101 | -0.51969 | 0.132755 | -3.91467 | 9.05E-05 | 0.00068 | 0.697521 |

| | | | | | | | |
|---|---|---|---|---|---|---|---|
| ALG3 | 1231.271 | -0.44678 | 0.114237 | -3.91095 | 9.19E-05 | 0.000691 | 0.733681 |
| KCNK17 | 76.48899 | -0.91505 | 0.234065 | -3.90939 | 9.25E-05 | 0.000694 | 0.530325 |
| STK33 | 497.6566 | -0.6472 | 0.165655 | -3.90693 | 9.35E-05 | 0.0007 | 0.638517 |
| MLKL | 43.38136 | -1.17379 | 0.300879 | -3.90121 | 9.57E-05 | 0.000715 | 0.443255 |
| EML3 | 742.8681 | -1.10068 | 0.282248 | -3.8997 | 9.63E-05 | 0.000719 | 0.466296 |
| ARFGAP1 | 2687.319 | -0.74605 | 0.191506 | -3.89571 | 9.79E-05 | 0.000729 | 0.596232 |
| YIPF2 | 1053.04 | -0.41505 | 0.10666 | -3.89137 | 9.97E-05 | 0.000741 | 0.749991 |
| MAP3K10 | 524.3054 | -0.6185 | 0.158967 | -3.89077 | 9.99E-05 | 0.000743 | 0.651346 |
| EBF4 | 116.1267 | -1.14768 | 0.295174 | -3.88815 | 0.000101 | 0.000749 | 0.45135 |
| ARHGEF17 | 467.5543 | -0.56915 | 0.146453 | -3.88625 | 0.000102 | 0.000754 | 0.674012 |
| AC140479 | 39.65505 | -1.11511 | 0.287113 | -3.88387 | 0.000103 | 0.000761 | 0.461656 |
| C16orf89 | 59.67836 | -1.03399 | 0.266232 | -3.88381 | 0.000103 | 0.000761 | 0.488357 |
| UQCC3 | 501.7438 | -0.41938 | 0.107978 | -3.88393 | 0.000103 | 0.000761 | 0.747747 |
| CACNA2D3 | 77.8383 | -0.92143 | 0.237279 | -3.88333 | 0.000103 | 0.000762 | 0.527985 |
| HS3ST3B1 | 281.5722 | -0.47124 | 0.121382 | -3.88231 | 0.000103 | 0.000764 | 0.721343 |
| ANKRD11 | 7106.396 | -0.42534 | 0.109594 | -3.88104 | 0.000104 | 0.000767 | 0.744665 |
| USP43 | 236.8956 | -0.4573 | 0.117891 | -3.87903 | 0.000105 | 0.000773 | 0.728347 |
| ACP2 | 630.1153 | -0.44476 | 0.114701 | -3.87756 | 0.000106 | 0.000777 | 0.734706 |
| P3H1 | 1511.903 | -0.55484 | 0.143102 | -3.8772 | 0.000106 | 0.000778 | 0.680735 |
| SLC37A2 | 162.4425 | -0.83802 | 0.216234 | -3.87555 | 0.000106 | 0.000783 | 0.559409 |
| CDK10 | 1255.043 | -0.66881 | 0.172617 | -3.87451 | 0.000107 | 0.000785 | 0.629027 |
| LMAN2 | 2990.519 | -0.44354 | 0.114578 | -3.87104 | 0.000108 | 0.000795 | 0.73533 |
| B4GALT7 | 405.143 | -0.52005 | 0.134356 | -3.87074 | 0.000109 | 0.000795 | 0.697345 |
| ACAP1 | 478.0807 | -0.74686 | 0.193088 | -3.86798 | 0.00011 | 0.000803 | 0.595898 |
| SERPINE1 | 216.818 | -0.56148 | 0.145339 | -3.86329 | 0.000112 | 0.000816 | 0.677604 |
| HAUS5 | 888.4 | -0.59355 | 0.153662 | -3.8627 | 0.000112 | 0.000817 | 0.662709 |
| VAV1 | 57.31268 | -1.21099 | 0.313657 | -3.86088 | 0.000113 | 0.000823 | 0.431971 |
| CNTN2 | 130.7581 | -1.0744 | 0.278524 | -3.85748 | 0.000115 | 0.000833 | 0.474868 |
| RAB11FIP1 | 96.61524 | -1.08631 | 0.281765 | -3.85537 | 0.000116 | 0.000839 | 0.470965 |
| CLCN7 | 2948.757 | -0.72033 | 0.186878 | -3.85458 | 0.000116 | 0.000841 | 0.606957 |
| CFAP69 | 102.9407 | -1.21536 | 0.315433 | -3.85299 | 0.000117 | 0.000845 | 0.430666 |
| F7 | 34.34634 | -1.53743 | 0.39909 | -3.85233 | 0.000117 | 0.000847 | 0.3445 |
| C1orf122 | 678.0931 | -0.48155 | 0.125043 | -3.85112 | 0.000118 | 0.000849 | 0.716205 |
| CLGN | 573.0351 | -0.55116 | 0.143275 | -3.84685 | 0.00012 | 0.000863 | 0.682473 |
| PLCB2 | 217.6017 | -0.63066 | 0.163967 | -3.84625 | 0.00012 | 0.000864 | 0.645882 |
| TMEM121E | 380.2378 | -0.50805 | 0.132096 | -3.84605 | 0.00012 | 0.000865 | 0.703174 |
| SCRIB | 3434.777 | -0.7563 | 0.196728 | -3.84441 | 0.000121 | 0.00087 | 0.592011 |
| LGMN | 1953.753 | -0.43831 | 0.114043 | -3.84335 | 0.000121 | 0.000873 | 0.738 |
| LY6G5B | 80.32771 | -1.20285 | 0.31305 | -3.84236 | 0.000122 | 0.000875 | 0.434416 |
| MANEAL | 205.108 | -0.68051 | 0.177175 | -3.84092 | 0.000123 | 0.00088 | 0.623943 |
| CEBPD | 92.04374 | -0.93221 | 0.242796 | -3.83946 | 0.000123 | 0.000884 | 0.524056 |
| C2orf40 | 37.69029 | -1.11179 | 0.28959 | -3.8392 | 0.000123 | 0.000885 | 0.462719 |
| SLC1A3 | 273.6674 | -0.66382 | 0.173024 | -3.83656 | 0.000125 | 0.000893 | 0.631206 |
| VILL | 168.4379 | -0.64296 | 0.167657 | -3.83497 | 0.000126 | 0.000898 | 0.640398 |

| | | | | | | | |
|---|---|---|---|---|---|---|---|
| PRSS50 | 76.24578 | -1.29232 | 0.337392 | -3.83033 | 0.000128 | 0.000914 | 0.408293 |
| ACP5 | 131.6601 | -0.84899 | 0.22173 | -3.82895 | 0.000129 | 0.000918 | 0.555172 |
| ATP6AP1 | 2161.342 | -0.53681 | 0.140233 | -3.82802 | 0.000129 | 0.000921 | 0.689291 |
| SH2D3A | 384.2724 | -0.76122 | 0.198914 | -3.82689 | 0.00013 | 0.000925 | 0.589996 |
| MAP11 | 335.3323 | -0.61882 | 0.161742 | -3.82595 | 0.00013 | 0.000927 | 0.651204 |
| PIGV | 372.5032 | -0.48383 | 0.1265 | -3.8248 | 0.000131 | 0.000931 | 0.715074 |
| AC092070 | 37.69503 | -1.63239 | 0.427185 | -3.82126 | 0.000133 | 0.000943 | 0.322554 |
| ARSD | 352.9151 | -0.71073 | 0.186048 | -3.82015 | 0.000133 | 0.000946 | 0.611011 |
| PKN3 | 631.4736 | -0.50933 | 0.133378 | -3.8187 | 0.000134 | 0.000951 | 0.702549 |
| CERS5 | 1130.883 | -0.68024 | 0.17817 | -3.81793 | 0.000135 | 0.000953 | 0.624061 |
| FAM186A | 5.355911 | -4.12841 | 1.081767 | -3.81636 | 0.000135 | 0.000959 | 0.057177 |
| PERM1 | 10.53385 | -2.57777 | 0.675568 | -3.81571 | 0.000136 | 0.00096 | 0.167499 |
| CNKSR1 | 443.796 | -0.7991 | 0.209702 | -3.81063 | 0.000139 | 0.000978 | 0.574709 |
| MPPE1 | 243.4185 | -0.73462 | 0.192792 | -3.81041 | 0.000139 | 0.000978 | 0.600977 |
| ZNF839 | 477.8872 | -0.42639 | 0.111899 | -3.8105 | 0.000139 | 0.000978 | 0.744122 |
| YIF1B | 1556.257 | -0.42801 | 0.112462 | -3.80582 | 0.000141 | 0.000995 | 0.743287 |
| STK10 | 577.102 | -0.59735 | 0.157128 | -3.80172 | 0.000144 | 0.00101 | 0.660965 |
| NPIPA1 | 1920.54 | -0.70703 | 0.186038 | -3.80043 | 0.000144 | 0.001014 | 0.612582 |
| NPR1 | 489.4132 | -0.4576 | 0.120639 | -3.79316 | 0.000149 | 0.001041 | 0.728195 |
| CACNA1F | 74.98987 | -1.05581 | 0.278465 | -3.79154 | 0.00015 | 0.001046 | 0.481026 |
| ATP6V1FN | 37.85581 | -1.52503 | 0.40233 | -3.79049 | 0.00015 | 0.00105 | 0.347473 |
| B3GAT3 | 588.6039 | -0.74742 | 0.197191 | -3.79031 | 0.00015 | 0.001051 | 0.59567 |
| ADM5 | 115.8334 | -1.15675 | 0.305503 | -3.78639 | 0.000153 | 0.001065 | 0.448521 |
| PDLIM2 | 467.1897 | -0.54959 | 0.145186 | -3.7854 | 0.000153 | 0.001068 | 0.683216 |
| MRPL55 | 478.8832 | -0.45844 | 0.121129 | -3.78471 | 0.000154 | 0.00107 | 0.727774 |
| AF228730. | 209.8617 | -0.6017 | 0.159 | -3.78431 | 0.000154 | 0.001071 | 0.658975 |
| FBXL6 | 702.2163 | -0.64494 | 0.170538 | -3.78178 | 0.000156 | 0.001081 | 0.63952 |
| RTEL1 | 842.8107 | -0.74707 | 0.197587 | -3.78097 | 0.000156 | 0.001084 | 0.595813 |
| ICAM5 | 125.0303 | -1.19255 | 0.315443 | -3.78056 | 0.000156 | 0.001085 | 0.437528 |
| KIF1C | 5084.796 | -0.7041 | 0.186271 | -3.77997 | 0.000157 | 0.001087 | 0.613826 |
| CNMD | 3108.851 | -0.56923 | 0.150613 | -3.77941 | 0.000157 | 0.001089 | 0.673977 |
| SLC39A4 | 28.88948 | -2.58668 | 0.684626 | -3.77823 | 0.000158 | 0.001094 | 0.166469 |
| BSCL2 | 710.2961 | -0.73912 | 0.195706 | -3.7767 | 0.000159 | 0.0011 | 0.599103 |
| SNPH | 373.4932 | -0.5298 | 0.140342 | -3.77504 | 0.00016 | 0.001107 | 0.692652 |
| KATNB1 | 1212.591 | -0.47781 | 0.126688 | -3.77157 | 0.000162 | 0.001121 | 0.718066 |
| AC009533 | 367.7823 | -0.60158 | 0.159587 | -3.76961 | 0.000164 | 0.001129 | 0.659031 |
| ACP7 | 20.31328 | -1.48537 | 0.39408 | -3.7692 | 0.000164 | 0.00113 | 0.357158 |
| CTSA | 1240.045 | -0.56081 | 0.148956 | -3.76498 | 0.000167 | 0.001147 | 0.677919 |
| SH3D21 | 392.0296 | -0.61777 | 0.164145 | -3.76357 | 0.000168 | 0.001152 | 0.651676 |
| MTND2P28 | 908.4288 | -0.84083 | 0.223435 | -3.76319 | 0.000168 | 0.001154 | 0.558323 |
| SIN3B | 2464.382 | -0.51186 | 0.136079 | -3.7615 | 0.000169 | 0.00116 | 0.701316 |
| ILDR1 | 83.81455 | -0.93474 | 0.248636 | -3.75949 | 0.00017 | 0.001168 | 0.523136 |
| TMEM151A | 57.64406 | -0.93371 | 0.248796 | -3.75291 | 0.000175 | 0.001196 | 0.523511 |
| INHA | 15.96185 | -2.16777 | 0.577658 | -3.75269 | 0.000175 | 0.001196 | 0.222554 |

| | | | | | | | |
|---|---|---|---|---|---|---|---|
| ALG12 | 733.8541 | -0.66345 | 0.176993 | -3.74846 | 0.000178 | 0.001214 | 0.631367 |
| CHKB-CPT | 258.8419 | -1.2657 | 0.337668 | -3.74836 | 0.000178 | 0.001214 | 0.415897 |
| ORAI3 | 326.9597 | -0.44184 | 0.117889 | -3.74796 | 0.000178 | 0.001216 | 0.736193 |
| TRPV1 | 190.3162 | -0.87974 | 0.234798 | -3.74678 | 0.000179 | 0.001221 | 0.543467 |
| GRN | 4706.36 | -0.5397 | 0.144054 | -3.7465 | 0.000179 | 0.001222 | 0.687915 |
| EPHA8 | 238.9647 | -0.64167 | 0.171458 | -3.74243 | 0.000182 | 0.00124 | 0.640972 |
| FAM3A | 528.6319 | -0.48992 | 0.130918 | -3.74217 | 0.000182 | 0.00124 | 0.712067 |
| BCAT2 | 794.7659 | -0.4307 | 0.115202 | -3.73865 | 0.000185 | 0.001257 | 0.741901 |
| CUL9 | 539.7116 | -0.92402 | 0.247203 | -3.73791 | 0.000186 | 0.001259 | 0.527038 |
| MTHFS | 305.3843 | -0.47808 | 0.127902 | -3.73789 | 0.000186 | 0.001259 | 0.71793 |
| GLI4 | 236.288 | -0.86722 | 0.232015 | -3.73776 | 0.000186 | 0.001259 | 0.548204 |
| ATP10A | 103.7252 | -1.18094 | 0.316199 | -3.73479 | 0.000188 | 0.001271 | 0.441065 |
| FIBIN | 36.02184 | -1.5095 | 0.404163 | -3.73488 | 0.000188 | 0.001271 | 0.351233 |
| THY1 | 6112.683 | -0.51141 | 0.136927 | -3.73494 | 0.000188 | 0.001271 | 0.701535 |
| TMEM238 | 51.3975 | -1.02916 | 0.275589 | -3.73441 | 0.000188 | 0.001271 | 0.489994 |
| PLXNB1 | 4154.163 | -1.21867 | 0.326624 | -3.73112 | 0.000191 | 0.001284 | 0.429678 |
| UGT3A2 | 862.0443 | -0.45966 | 0.123204 | -3.73091 | 0.000191 | 0.001285 | 0.727156 |
| ZSWIM9 | 660.2407 | -0.70587 | 0.189212 | -3.73061 | 0.000191 | 0.001286 | 0.613071 |
| HSPB6 | 16.07986 | -1.82392 | 0.488932 | -3.73042 | 0.000191 | 0.001286 | 0.282452 |
| AL731733. | 7.454037 | -3.13505 | 0.841044 | -3.72757 | 0.000193 | 0.001299 | 0.11383 |
| TMEM132A | 2470.034 | -0.7498 | 0.201172 | -3.72716 | 0.000194 | 0.001301 | 0.594685 |
| LRP5L | 126.6516 | -0.83604 | 0.224466 | -3.72456 | 0.000196 | 0.001312 | 0.56018 |
| PNPLA2 | 1317.812 | -0.62309 | 0.167364 | -3.72297 | 0.000197 | 0.001319 | 0.649279 |
| CLDN15 | 106.3383 | -0.9007 | 0.24234 | -3.71666 | 0.000202 | 0.001349 | 0.535629 |
| PTPRCAP | 19.19002 | -3.4727 | 0.93447 | -3.71622 | 0.000202 | 0.00135 | 0.090077 |
| EFNA1 | 136.3322 | -0.59355 | 0.159802 | -3.7143 | 0.000204 | 0.001359 | 0.662709 |
| CYB561A3 | 526.7832 | -0.49894 | 0.134364 | -3.71331 | 0.000205 | 0.001363 | 0.707628 |
| U2AF1L4 | 199.8002 | -0.47556 | 0.1281 | -3.71241 | 0.000205 | 0.001368 | 0.719189 |
| STAG3L5P | 321.3017 | -0.43481 | 0.117236 | -3.7088 | 0.000208 | 0.001384 | 0.739793 |
| RPL13P12 | 115.777 | -0.70803 | 0.190982 | -3.70732 | 0.000209 | 0.00139 | 0.612155 |
| EMILIN2 | 248.3876 | -0.49756 | 0.134253 | -3.70612 | 0.00021 | 0.001396 | 0.708305 |
| FBRS | 464.1857 | -0.45092 | 0.121678 | -3.70582 | 0.000211 | 0.001397 | 0.731577 |
| OR5H6 | 3.335463 | -5.28725 | 1.426984 | -3.70519 | 0.000211 | 0.0014 | 0.025608 |
| EBI3 | 33.33466 | -1.39028 | 0.375395 | -3.70352 | 0.000213 | 0.001407 | 0.38149 |
| PRSS27 | 90.23896 | -0.96152 | 0.259614 | -3.70367 | 0.000213 | 0.001407 | 0.513514 |
| DNHD1 | 439.223 | -0.81584 | 0.22049 | -3.70012 | 0.000216 | 0.001424 | 0.568078 |
| G0S2 | 213.2113 | -1.23431 | 0.333669 | -3.69921 | 0.000216 | 0.001428 | 0.425045 |
| SLC4A3 | 350.253 | -0.68188 | 0.184642 | -3.69297 | 0.000222 | 0.001461 | 0.623354 |
| TMEM212 | 175.9501 | -0.52588 | 0.142474 | -3.69101 | 0.000223 | 0.00147 | 0.694538 |
| MYDGF | 903.2129 | -0.49959 | 0.135358 | -3.69089 | 0.000223 | 0.001471 | 0.707307 |
| NPTXR | 548.7087 | -0.4258 | 0.11539 | -3.69012 | 0.000224 | 0.001474 | 0.744424 |
| KAT2A | 1375.76 | -0.77182 | 0.20923 | -3.68889 | 0.000225 | 0.001481 | 0.585676 |
| PRR7 | 182.9874 | -0.53009 | 0.143734 | -3.68795 | 0.000226 | 0.001484 | 0.692513 |
| SPOCK2 | 144.9335 | -1.06689 | 0.289295 | -3.68792 | 0.000226 | 0.001484 | 0.477345 |

| | | | | | | | |
|---|---|---|---|---|---|---|---|
| AC022415 | 134.2268 | -0.85034 | 0.23086 | -3.68337 | 0.00023 | 0.001508 | 0.554653 |
| LIMS2 | 96.00091 | -0.69379 | 0.188471 | -3.68114 | 0.000232 | 0.00152 | 0.618229 |
| TMEM125 | 472.0207 | -0.57272 | 0.155589 | -3.68101 | 0.000232 | 0.001521 | 0.672346 |
| PLK3 | 157.9256 | -0.84683 | 0.230064 | -3.68084 | 0.000232 | 0.001521 | 0.556005 |
| IGSF21 | 232.2427 | -0.74451 | 0.202399 | -3.67843 | 0.000235 | 0.001534 | 0.59687 |
| ADCY4 | 31.0784 | -1.29728 | 0.352979 | -3.67524 | 0.000238 | 0.001552 | 0.406891 |
| CRELD2 | 513.9078 | -0.66152 | 0.180001 | -3.67508 | 0.000238 | 0.001552 | 0.632212 |
| EOGT | 427.4182 | -0.59749 | 0.162578 | -3.67508 | 0.000238 | 0.001552 | 0.660904 |
| CCDC18 | 440.3445 | -0.47341 | 0.128977 | -3.67048 | 0.000242 | 0.001578 | 0.720262 |
| IKBKG | 406.2171 | -0.64081 | 0.174668 | -3.66873 | 0.000244 | 0.001588 | 0.641353 |
| ANO8 | 919.383 | -0.88918 | 0.242634 | -3.66471 | 0.000248 | 0.001607 | 0.53992 |
| ENTPD8 | 36.67537 | -1.4308 | 0.390441 | -3.66458 | 0.000248 | 0.001607 | 0.370924 |
| AC073585 | 79.99517 | -1.13418 | 0.309825 | -3.66071 | 0.000252 | 0.001627 | 0.455593 |
| CSF2RA | 57.23105 | -1.32309 | 0.361426 | -3.66076 | 0.000251 | 0.001627 | 0.399677 |
| TNK2 | 967.3387 | -0.63159 | 0.172701 | -3.65716 | 0.000255 | 0.001646 | 0.645463 |
| MIA2 | 673.7135 | -0.56271 | 0.154109 | -3.65136 | 0.000261 | 0.001679 | 0.677031 |
| TRAF1 | 116.0498 | -0.81214 | 0.222606 | -3.64831 | 0.000264 | 0.001698 | 0.569538 |
| SLC9A3 | 353.8854 | -0.91158 | 0.249945 | -3.64713 | 0.000265 | 0.001704 | 0.531602 |
| LGALS4 | 7.169165 | -2.75262 | 0.755526 | -3.64331 | 0.000269 | 0.001726 | 0.148382 |
| MUC1 | 395.5855 | -1.08771 | 0.298686 | -3.64166 | 0.000271 | 0.001736 | 0.470507 |
| TCTA | 343.4705 | -0.59395 | 0.163237 | -3.63856 | 0.000274 | 0.001754 | 0.662528 |
| C15orf53 | 17.80402 | -1.74777 | 0.480433 | -3.63791 | 0.000275 | 0.001756 | 0.297762 |
| MICAL1 | 3019.383 | -0.69929 | 0.192219 | -3.63799 | 0.000275 | 0.001756 | 0.615875 |
| SARS2 | 972.5098 | -0.53112 | 0.146004 | -3.63774 | 0.000275 | 0.001757 | 0.692015 |
| AC126755 | 1140.132 | -0.75988 | 0.209025 | -3.63538 | 0.000278 | 0.001771 | 0.590544 |
| TMC8 | 36.9857 | -1.1107 | 0.305736 | -3.63287 | 0.00028 | 0.001785 | 0.463069 |
| NAP1L6 | 14.01896 | -2.00054 | 0.551083 | -3.63019 | 0.000283 | 0.0018 | 0.249907 |
| QSOX1 | 2560.616 | -0.48287 | 0.133029 | -3.62984 | 0.000284 | 0.001802 | 0.715552 |
| SLC26A11 | 580.0384 | -0.53364 | 0.147049 | -3.62896 | 0.000285 | 0.001807 | 0.690812 |
| GAS6 | 1848.443 | -0.68144 | 0.187788 | -3.62878 | 0.000285 | 0.001808 | 0.623542 |
| AGAP7P | 185.3159 | -0.80734 | 0.222513 | -3.62828 | 0.000285 | 0.001811 | 0.571434 |
| COL12A1 | 185.5813 | -1.05827 | 0.291828 | -3.62633 | 0.000287 | 0.001823 | 0.480209 |
| STOML1 | 113.2414 | -0.84166 | 0.232273 | -3.62358 | 0.000291 | 0.00184 | 0.558002 |
| CALML6 | 22.34591 | -1.79336 | 0.495076 | -3.6224 | 0.000292 | 0.001848 | 0.288498 |
| CEBPB | 1394.991 | -0.62425 | 0.172408 | -3.62074 | 0.000294 | 0.001857 | 0.648759 |
| CES3 | 210.3862 | -0.64828 | 0.179042 | -3.6208 | 0.000294 | 0.001857 | 0.638042 |
| MGAT5B | 340.1167 | -0.50412 | 0.139235 | -3.62063 | 0.000294 | 0.001857 | 0.705091 |
| TFEB | 135.6078 | -0.90685 | 0.25048 | -3.62045 | 0.000294 | 0.001858 | 0.533348 |
| DOC2A | 200.1449 | -1.03666 | 0.286346 | -3.62032 | 0.000294 | 0.001858 | 0.487453 |
| LRRC75B | 347.1429 | -0.65906 | 0.182156 | -3.61814 | 0.000297 | 0.001873 | 0.633289 |
| LRFN1 | 205.1546 | -0.55845 | 0.154438 | -3.61602 | 0.000299 | 0.001887 | 0.679032 |
| KIF12 | 65.3486 | -1.17712 | 0.325545 | -3.61586 | 0.000299 | 0.001887 | 0.442232 |
| RNF31 | 1169.549 | -0.51375 | 0.142131 | -3.61461 | 0.000301 | 0.001895 | 0.700401 |
| RRN3P3 | 92.53885 | -0.9821 | 0.27178 | -3.6136 | 0.000302 | 0.001901 | 0.506241 |

| | | | | | | | |
|---|---|---|---|---|---|---|---|
| PPP1R37 | 835.7358 | -0.49974 | 0.138336 | -3.6125 | 0.000303 | 0.001907 | 0.707234 |
| TMEM87B | 666.9091 | -0.61443 | 0.170081 | -3.61257 | 0.000303 | 0.001907 | 0.653189 |
| ANAPC7 | 1657.454 | -0.46939 | 0.130128 | -3.60714 | 0.00031 | 0.001941 | 0.722269 |
| PIGL | 360.8865 | -0.49816 | 0.13818 | -3.60512 | 0.000312 | 0.001953 | 0.708011 |
| PLXNA3 | 904.5596 | -0.74959 | 0.208395 | -3.597 | 0.000322 | 0.002012 | 0.594771 |
| DENND6B | 139.8532 | -1.13523 | 0.315882 | -3.59385 | 0.000326 | 0.00203 | 0.455262 |
| ZNF283 | 46.92621 | -1.27274 | 0.354395 | -3.59131 | 0.000329 | 0.002048 | 0.413872 |
| ARVCF | 1476.346 | -0.62793 | 0.174963 | -3.58893 | 0.000332 | 0.002065 | 0.647104 |
| CALHM2 | 118.3405 | -0.71472 | 0.199183 | -3.58825 | 0.000333 | 0.002068 | 0.609324 |
| JAG2 | 457.1252 | -0.91667 | 0.255565 | -3.58684 | 0.000335 | 0.002077 | 0.529731 |
| PDCL3P4 | 34.28533 | -1.58673 | 0.442557 | -3.58538 | 0.000337 | 0.002087 | 0.332924 |
| SNHG32 | 2987.64 | -0.6125 | 0.170867 | -3.58466 | 0.000338 | 0.002093 | 0.654062 |
| CORO6 | 144.6431 | -1.06135 | 0.296097 | -3.58448 | 0.000338 | 0.002093 | 0.479183 |
| SELENOS | 690.9811 | -0.42628 | 0.118937 | -3.58407 | 0.000338 | 0.002096 | 0.74418 |
| ZNF598 | 1373.494 | -0.62997 | 0.175833 | -3.58276 | 0.00034 | 0.002104 | 0.646192 |
| MCUB | 295.9653 | -0.49947 | 0.139434 | -3.58213 | 0.000341 | 0.002108 | 0.707367 |
| AMN | 19.44829 | -2.98782 | 0.834412 | -3.58074 | 0.000343 | 0.002119 | 0.12606 |
| JMJD8 | 1195.238 | -0.47755 | 0.133432 | -3.57895 | 0.000345 | 0.002133 | 0.718198 |
| WASH7P | 420.4729 | -0.48251 | 0.13485 | -3.57812 | 0.000346 | 0.002138 | 0.715731 |
| SPINT2 | 6632.996 | -0.50222 | 0.140441 | -3.57602 | 0.000349 | 0.002152 | 0.706021 |
| TMEM234 | 163.8531 | -0.8401 | 0.234976 | -3.57525 | 0.00035 | 0.002156 | 0.558606 |
| TPRN | 527.0397 | -0.64114 | 0.179515 | -3.57152 | 0.000355 | 0.002184 | 0.641205 |
| IGSF9 | 560.2394 | -0.67314 | 0.188515 | -3.57075 | 0.000356 | 0.002188 | 0.627141 |
| SLC25A28 | 212.3029 | -0.55072 | 0.154243 | -3.57047 | 0.000356 | 0.00219 | 0.68268 |
| ST20 | 98.77672 | -0.69195 | 0.193835 | -3.5698 | 0.000357 | 0.002194 | 0.619016 |
| GRIN2C | 13.49143 | -1.81094 | 0.507605 | -3.56761 | 0.00036 | 0.002211 | 0.285006 |
| LTBP4 | 2244.863 | -0.49071 | 0.137547 | -3.56759 | 0.00036 | 0.002211 | 0.711675 |
| CLCNKB | 95.69788 | -0.71606 | 0.200903 | -3.5642 | 0.000365 | 0.002235 | 0.608758 |
| EML2 | 528.6653 | -0.44805 | 0.125773 | -3.5624 | 0.000367 | 0.002248 | 0.733031 |
| CHST14 | 604.4245 | -0.59778 | 0.167826 | -3.56191 | 0.000368 | 0.002252 | 0.660769 |
| UBAP1L | 86.5811 | -1.15575 | 0.324513 | -3.56149 | 0.000369 | 0.002254 | 0.448833 |
| CYHR1 | 1299.525 | -0.44975 | 0.126356 | -3.55942 | 0.000372 | 0.002271 | 0.732168 |
| GALE | 415.3308 | -0.46709 | 0.131332 | -3.55658 | 0.000376 | 0.002295 | 0.72342 |
| ZDHHC21 | 459.8737 | -0.56543 | 0.159047 | -3.55512 | 0.000378 | 0.002306 | 0.675754 |
| CD177 | 91.14903 | -0.70102 | 0.197231 | -3.55432 | 0.000379 | 0.00231 | 0.615137 |
| GTF2IRD2E | 151.568 | -0.61119 | 0.171976 | -3.55392 | 0.00038 | 0.002313 | 0.654657 |
| DOT1L | 2912.679 | -0.47175 | 0.13284 | -3.55126 | 0.000383 | 0.002336 | 0.721089 |
| DDX11 | 2882.644 | -0.63575 | 0.179102 | -3.54967 | 0.000386 | 0.002349 | 0.643605 |
| TMEM221 | 136.0159 | -0.69937 | 0.197125 | -3.54782 | 0.000388 | 0.002362 | 0.615843 |
| ASB16 | 55.06428 | -1.06084 | 0.299024 | -3.54767 | 0.000389 | 0.002363 | 0.479353 |
| FAM153CF | 47.1173 | -1.0169 | 0.286663 | -3.54738 | 0.000389 | 0.002365 | 0.494176 |
| GAS8 | 307.5562 | -0.64636 | 0.182345 | -3.54472 | 0.000393 | 0.002386 | 0.638889 |
| IQGAP2 | 2167.46 | -0.6669 | 0.1882 | -3.5436 | 0.000395 | 0.002394 | 0.629857 |
| TTBK1 | 69.36163 | -0.95357 | 0.269188 | -3.54241 | 0.000396 | 0.002403 | 0.516352 |

| | | | | | | | |
|---|---|---|---|---|---|---|---|
| NBEAL2 | 2081.176 | -1.23961 | 0.350122 | -3.54051 | 0.000399 | 0.002417 | 0.423487 |
| ZBTB7B | 1016.808 | -0.58454 | 0.165301 | -3.53618 | 0.000406 | 0.002454 | 0.666864 |
| IFI30 | 778.2122 | -0.48028 | 0.135896 | -3.53416 | 0.000409 | 0.002469 | 0.716839 |
| DCAF15 | 1814.972 | -0.43919 | 0.124335 | -3.53227 | 0.000412 | 0.002483 | 0.737551 |
| TCF7 | 273.0035 | -0.55609 | 0.157671 | -3.52691 | 0.00042 | 0.002525 | 0.680142 |
| TRIM53AP | 3.537384 | -5.3756 | 1.524671 | -3.52574 | 0.000422 | 0.002535 | 0.024087 |
| PIPOX | 1355.129 | -0.42487 | 0.120646 | -3.52165 | 0.000429 | 0.002572 | 0.744904 |
| RRP7BP | 375.5225 | -0.57965 | 0.164597 | -3.5216 | 0.000429 | 0.002572 | 0.669128 |
| DEPTOR | 107.1834 | -0.7964 | 0.22645 | -3.51689 | 0.000437 | 0.002615 | 0.575784 |
| INPP5J | 215.4775 | -0.69151 | 0.196676 | -3.51599 | 0.000438 | 0.002623 | 0.619205 |
| FAM238C | 13.71935 | -2.13816 | 0.608443 | -3.51415 | 0.000441 | 0.002638 | 0.227169 |
| C8orf82 | 427.9381 | -0.46801 | 0.133263 | -3.51197 | 0.000445 | 0.002656 | 0.722959 |
| LRRC34 | 31.2052 | -1.25322 | 0.356931 | -3.51111 | 0.000446 | 0.002662 | 0.41951 |
| MRI1 | 586.504 | -0.59284 | 0.168849 | -3.51108 | 0.000446 | 0.002662 | 0.663036 |
| BRF1 | 594.3324 | -0.4847 | 0.138135 | -3.5089 | 0.00045 | 0.002682 | 0.714644 |
| SGPP2 | 58.38257 | -0.82294 | 0.234577 | -3.5082 | 0.000451 | 0.002687 | 0.565288 |
| UCMA | 37.75732 | -1.24276 | 0.354296 | -3.50768 | 0.000452 | 0.002691 | 0.422565 |
| APCDD1 | 132.0442 | -0.7299 | 0.208346 | -3.50332 | 0.000459 | 0.00273 | 0.602944 |
| AKNA | 2522.283 | -0.73696 | 0.210426 | -3.50222 | 0.000461 | 0.00274 | 0.600003 |
| ABCC10 | 719.9208 | -0.71969 | 0.205733 | -3.49819 | 0.000468 | 0.002779 | 0.607226 |
| MBOAT7 | 1120.763 | -0.63847 | 0.182524 | -3.49803 | 0.000469 | 0.002779 | 0.642392 |
| TBC1D29P | 9.162162 | -2.66214 | 0.76168 | -3.4951 | 0.000474 | 0.002805 | 0.157985 |
| LAMP3 | 34.87032 | -1.86076 | 0.532429 | -3.49484 | 0.000474 | 0.002806 | 0.275332 |
| RAD9A | 437.2227 | -0.41714 | 0.119444 | -3.49237 | 0.000479 | 0.002831 | 0.748906 |
| BAMBI | 253.8662 | -0.47584 | 0.13639 | -3.48881 | 0.000485 | 0.002866 | 0.719049 |
| PDE8A | 287.4629 | -0.76109 | 0.218175 | -3.48844 | 0.000486 | 0.002867 | 0.590049 |
| SYBU | 25.01525 | -1.64043 | 0.470392 | -3.48736 | 0.000488 | 0.002876 | 0.320762 |
| PRELID3A | 156.3403 | -0.59516 | 0.170728 | -3.48603 | 0.00049 | 0.002889 | 0.661969 |
| SBSPON | 127.8792 | -0.8663 | 0.248578 | -3.48504 | 0.000492 | 0.002897 | 0.54855 |
| VCX | 5.561095 | -3.24056 | 0.930223 | -3.48364 | 0.000495 | 0.002908 | 0.105802 |
| C1orf35 | 622.3239 | -0.42303 | 0.121441 | -3.4834 | 0.000495 | 0.002908 | 0.745858 |
| ZNF789 | 268.4779 | -0.55746 | 0.160169 | -3.48047 | 0.000501 | 0.002935 | 0.679496 |
| THEM6 | 156.7819 | -0.52017 | 0.14949 | -3.47963 | 0.000502 | 0.002944 | 0.69729 |
| MYLK3 | 55.08149 | -0.83546 | 0.240227 | -3.47778 | 0.000506 | 0.002962 | 0.560405 |
| RANBP17 | 285.0855 | -0.76395 | 0.219698 | -3.47727 | 0.000507 | 0.002967 | 0.588882 |
| MAN2A2 | 903.0266 | -0.48303 | 0.139193 | -3.47024 | 0.00052 | 0.003034 | 0.715471 |
| POMT2 | 900.7984 | -0.65719 | 0.189391 | -3.47002 | 0.00052 | 0.003035 | 0.634111 |
| CST3 | 608.0806 | -0.49375 | 0.142327 | -3.46913 | 0.000522 | 0.003043 | 0.710175 |
| FCHSD1 | 682.9542 | -0.60984 | 0.175804 | -3.46888 | 0.000523 | 0.003045 | 0.655268 |
| PPIB | 5625.197 | -0.45542 | 0.131299 | -3.46859 | 0.000523 | 0.003047 | 0.729297 |
| ABCB10 | 535.8317 | -0.43877 | 0.126524 | -3.46788 | 0.000525 | 0.003053 | 0.737763 |
| TIAF1 | 71.77348 | -1.2726 | 0.367286 | -3.46486 | 0.00053 | 0.003084 | 0.413914 |
| CARMIL3 | 441.3961 | -0.9143 | 0.263991 | -3.46337 | 0.000533 | 0.003098 | 0.530601 |
| BRSK2 | 629.6563 | -0.48304 | 0.139504 | -3.46255 | 0.000535 | 0.003105 | 0.715468 |

| | | | | | | | |
|---|---|---|---|---|---|---|---|
| SLC26A6 | 1303.086 | -0.77846 | 0.224828 | -3.46246 | 0.000535 | 0.003105 | 0.582989 |
| AL353625. | 69.32797 | -0.95561 | 0.276024 | -3.46206 | 0.000536 | 0.003109 | 0.515623 |
| SPOCK3 | 51.81938 | -1.13192 | 0.326973 | -3.46181 | 0.000537 | 0.003111 | 0.456309 |
| C2CD2L | 123.2234 | -0.53847 | 0.155694 | -3.45848 | 0.000543 | 0.003144 | 0.688503 |
| HSD3B7 | 119.1826 | -0.74705 | 0.216054 | -3.45772 | 0.000545 | 0.00315 | 0.59582 |
| AJAP1 | 31.4099 | -1.23593 | 0.357652 | -3.45568 | 0.000549 | 0.003171 | 0.424569 |
| CLDN3 | 768.5396 | -0.6605 | 0.19122 | -3.45411 | 0.000552 | 0.003189 | 0.632661 |
| CDRT4 | 288.5954 | -1.09752 | 0.317769 | -3.45383 | 0.000553 | 0.00319 | 0.467319 |
| GLB1L3 | 885.9054 | -0.43575 | 0.12621 | -3.4526 | 0.000555 | 0.003203 | 0.739308 |
| CLEC2D | 156.4548 | -1.18133 | 0.342232 | -3.45184 | 0.000557 | 0.003208 | 0.440945 |
| FAM98C | 187.6025 | -0.70451 | 0.204125 | -3.45137 | 0.000558 | 0.003213 | 0.613651 |
| SEMA5B | 1627.388 | -0.5532 | 0.160383 | -3.44921 | 0.000562 | 0.003237 | 0.681509 |
| GTF2H2 | 45.10471 | -1.82962 | 0.530696 | -3.44759 | 0.000566 | 0.003253 | 0.281338 |
| SPON2 | 82.20334 | -0.98561 | 0.285885 | -3.44756 | 0.000566 | 0.003253 | 0.505013 |
| ABCA5 | 76.65166 | -1.42844 | 0.414698 | -3.44454 | 0.000572 | 0.003285 | 0.371531 |
| MFSD3 | 1246.826 | -0.45828 | 0.13306 | -3.44416 | 0.000573 | 0.003289 | 0.727853 |
| GATD1 | 1439.172 | -0.52167 | 0.151722 | -3.43834 | 0.000585 | 0.003351 | 0.696564 |
| HIP1R | 495.3332 | -0.82519 | 0.240099 | -3.43685 | 0.000589 | 0.003365 | 0.56441 |
| ZNF692 | 689.9053 | -0.72201 | 0.210106 | -3.4364 | 0.000589 | 0.003369 | 0.606253 |
| HDAC7 | 2238.573 | -0.48119 | 0.140044 | -3.43602 | 0.00059 | 0.003371 | 0.716384 |
| ELL2 | 743.4951 | -0.48833 | 0.142192 | -3.43429 | 0.000594 | 0.003391 | 0.712851 |
| SHOX | 48.82957 | -1.11012 | 0.323293 | -3.4338 | 0.000595 | 0.003394 | 0.463254 |
| DHRS7B | 463.1162 | -0.47099 | 0.137219 | -3.43238 | 0.000598 | 0.003411 | 0.721471 |
| ASPG | 36.91801 | -1.64905 | 0.481285 | -3.42634 | 0.000612 | 0.003472 | 0.318851 |
| AP000346. | 88.70452 | -1.18071 | 0.34463 | -3.42603 | 0.000612 | 0.003475 | 0.441134 |
| MOGS | 1822.869 | -0.55203 | 0.161166 | -3.4252 | 0.000614 | 0.003483 | 0.682062 |
| CRYZL2P | 10.91234 | -2.39868 | 0.700334 | -3.42505 | 0.000615 | 0.003484 | 0.189638 |
| EHBP1L1 | 366.2222 | -1.05846 | 0.309079 | -3.42454 | 0.000616 | 0.003489 | 0.480146 |
| MID2 | 70.38222 | -0.7782 | 0.2273 | -3.42366 | 0.000618 | 0.003499 | 0.583095 |
| CEL | 74.83979 | -1.10771 | 0.32373 | -3.42171 | 0.000622 | 0.003523 | 0.46403 |
| DPY19L2P | 71.43927 | -0.86366 | 0.252898 | -3.41506 | 0.000638 | 0.003596 | 0.549556 |
| LTB4R2 | 156.2248 | -0.67179 | 0.19687 | -3.41237 | 0.000644 | 0.00363 | 0.627726 |
| CDC45 | 637.8851 | -0.4286 | 0.125659 | -3.41084 | 0.000648 | 0.003645 | 0.742981 |
| ARHGAP22 | 243.9401 | -0.5865 | 0.172098 | -3.40793 | 0.000655 | 0.003681 | 0.665957 |
| CTSF | 97.26166 | -1.07375 | 0.315231 | -3.40625 | 0.000659 | 0.0037 | 0.475081 |
| MT-ND1 | 58280.37 | -1.05874 | 0.310861 | -3.40583 | 0.00066 | 0.003703 | 0.480052 |
| PLOD1 | 2044.938 | -0.43002 | 0.126321 | -3.40417 | 0.000664 | 0.003721 | 0.742253 |
| TRMT2A | 753.8853 | -0.62136 | 0.182749 | -3.40005 | 0.000674 | 0.003771 | 0.650059 |
| BMP8A | 84.86119 | -0.75206 | 0.221261 | -3.39898 | 0.000676 | 0.003781 | 0.593754 |
| S1PR2 | 334.8795 | -0.47907 | 0.141038 | -3.39677 | 0.000682 | 0.003808 | 0.717439 |
| HPSE | 100.6828 | -0.96685 | 0.284748 | -3.39548 | 0.000685 | 0.003824 | 0.511621 |
| SBF1 | 3635.145 | -0.65364 | 0.19263 | -3.39326 | 0.000691 | 0.00385 | 0.635674 |
| C1orf94 | 315.9594 | -0.57455 | 0.16933 | -3.39307 | 0.000691 | 0.003851 | 0.671497 |
| TYK2 | 602.8858 | -0.84287 | 0.248464 | -3.39231 | 0.000693 | 0.003861 | 0.557534 |

| | | | | | | | |
|---|---|---|---|---|---|---|---|
| SGSM3 | 1682.652 | -0.76263 | 0.225164 | -3.38698 | 0.000707 | 0.003929 | 0.589422 |
| ZNF767P | 300.9422 | -0.72428 | 0.214069 | -3.38341 | 0.000716 | 0.003969 | 0.605298 |
| GJB2 | 7.640782 | -2.69844 | 0.797971 | -3.38163 | 0.000721 | 0.003992 | 0.154059 |
| MSLN | 11.36189 | -1.98824 | 0.588198 | -3.38022 | 0.000724 | 0.004011 | 0.252047 |
| AC068946 | 46.78394 | -1.4312 | 0.42352 | -3.37929 | 0.000727 | 0.004021 | 0.370823 |
| CSPG4 | 422.607 | -0.45949 | 0.135971 | -3.37934 | 0.000727 | 0.004021 | 0.727243 |
| AC092821 | 169.9705 | -0.92254 | 0.273156 | -3.37734 | 0.000732 | 0.004041 | 0.527579 |
| FZD9 | 105.1319 | -0.76022 | 0.225386 | -3.37298 | 0.000744 | 0.0041 | 0.590404 |
| RNF187 | 3056.671 | -0.59825 | 0.177405 | -3.37221 | 0.000746 | 0.004107 | 0.660556 |
| MZF1 | 507.6657 | -0.68461 | 0.203099 | -3.37081 | 0.000749 | 0.004123 | 0.622174 |
| LETM2 | 83.41928 | -0.90963 | 0.26987 | -3.37062 | 0.00075 | 0.004123 | 0.532322 |
| NPL | 70.90886 | -0.89663 | 0.266099 | -3.36953 | 0.000753 | 0.004138 | 0.53714 |
| SIDT2 | 685.5065 | -0.53943 | 0.160231 | -3.36657 | 0.000761 | 0.004175 | 0.688044 |
| SYVN1 | 863.4565 | -0.43321 | 0.128783 | -3.36384 | 0.000769 | 0.004212 | 0.740614 |
| ANKRD13D | 468.3522 | -0.74443 | 0.221362 | -3.36298 | 0.000771 | 0.004223 | 0.596902 |
| SSPO | 82.61382 | -1.80076 | 0.535718 | -3.36139 | 0.000776 | 0.004243 | 0.287024 |
| CTIF | 608.3127 | -0.69504 | 0.206875 | -3.35972 | 0.00078 | 0.004266 | 0.617692 |
| AL137013. | 10.16659 | -2.68001 | 0.797751 | -3.35945 | 0.000781 | 0.004269 | 0.15604 |
| TMSB4XP4 | 22.57153 | -1.49722 | 0.44575 | -3.35888 | 0.000783 | 0.004274 | 0.354236 |
| ZNF781 | 15.91857 | -1.94776 | 0.579973 | -3.35836 | 0.000784 | 0.004279 | 0.259218 |
| STXBP2 | 1355.42 | -0.50873 | 0.151505 | -3.35784 | 0.000786 | 0.004284 | 0.70284 |
| CROCC | 2137.112 | -0.67938 | 0.202394 | -3.35671 | 0.000789 | 0.004299 | 0.624434 |
| SRCIN1 | 44.99891 | -1.50739 | 0.449345 | -3.35463 | 0.000795 | 0.004321 | 0.351748 |
| CEACAM19 | 108.9949 | -1.09017 | 0.325279 | -3.35149 | 0.000804 | 0.004366 | 0.469706 |
| AL445423. | 131.7005 | -0.74375 | 0.222218 | -3.34692 | 0.000817 | 0.004426 | 0.597186 |
| SLC16A7 | 145.0128 | -0.49096 | 0.146694 | -3.34682 | 0.000817 | 0.004426 | 0.711551 |
| P3H4 | 937.5748 | -0.44734 | 0.133679 | -3.34634 | 0.000819 | 0.004433 | 0.733396 |
| CLPTM1 | 2228.971 | -0.44664 | 0.133526 | -3.34493 | 0.000823 | 0.004451 | 0.733751 |
| AOC2 | 76.28426 | -0.7314 | 0.218746 | -3.34361 | 0.000827 | 0.004471 | 0.602319 |
| NKAIN4 | 207.725 | -0.67722 | 0.202652 | -3.34176 | 0.000832 | 0.004494 | 0.625371 |
| NFKBIE | 418.106 | -0.45781 | 0.137004 | -3.3416 | 0.000833 | 0.004495 | 0.728089 |
| RHOXF1P1 | 6.926317 | -2.69693 | 0.807197 | -3.34111 | 0.000834 | 0.0045 | 0.154221 |
| SCAND2P | 217.9225 | -0.79258 | 0.2375 | -3.3372 | 0.000846 | 0.004549 | 0.577309 |
| SH3GL1P1 | 34.47319 | -1.19526 | 0.358273 | -3.33617 | 0.000849 | 0.004563 | 0.436708 |
| NSUN5P1 | 1937.439 | -0.70612 | 0.21185 | -3.3331 | 0.000859 | 0.004603 | 0.612967 |
| CD81 | 5230.429 | -0.53635 | 0.161079 | -3.32974 | 0.000869 | 0.004656 | 0.689512 |
| TAF1C | 2045.312 | -0.52926 | 0.158968 | -3.32936 | 0.00087 | 0.00466 | 0.692908 |
| TMEM260 | 453.8173 | -0.49455 | 0.148586 | -3.32838 | 0.000874 | 0.004676 | 0.709782 |
| AL669831. | 49.86965 | -0.99126 | 0.297867 | -3.32784 | 0.000875 | 0.004683 | 0.50304 |
| PAPLN | 1980.963 | -0.7804 | 0.234519 | -3.32767 | 0.000876 | 0.004685 | 0.582205 |
| EMILIN1 | 172.1233 | -0.82052 | 0.246589 | -3.3275 | 0.000876 | 0.004686 | 0.566236 |
| AC073869 | 491.7833 | -0.42816 | 0.128686 | -3.32719 | 0.000877 | 0.004688 | 0.743209 |
| CCDC17 | 38.44279 | -1.15689 | 0.347791 | -3.32639 | 0.00088 | 0.004699 | 0.448479 |
| DOC2B | 105.6973 | -1.05621 | 0.317683 | -3.32474 | 0.000885 | 0.004723 | 0.480893 |

| | | | | | | | |
|---|---|---|---|---|---|---|---|
| TMEM56 | 132.8444 | -1.19233 | 0.358654 | -3.32445 | 0.000886 | 0.004723 | 0.437596 |
| TACR3 | 242.4653 | -0.52026 | 0.15676 | -3.31883 | 0.000904 | 0.004806 | 0.697246 |
| NFATC4 | 1686.609 | -0.69922 | 0.210712 | -3.31835 | 0.000906 | 0.004811 | 0.615906 |
| CNTD1 | 15.38558 | -1.91028 | 0.575983 | -3.31655 | 0.000911 | 0.004835 | 0.266042 |
| ABCB11 | 38.26315 | -0.95744 | 0.288785 | -3.31542 | 0.000915 | 0.004847 | 0.514969 |
| LYPD1 | 171.3684 | -0.65868 | 0.198741 | -3.31427 | 0.000919 | 0.004862 | 0.633457 |
| MACO1 | 514.4237 | -0.41929 | 0.12655 | -3.31319 | 0.000922 | 0.004879 | 0.747795 |
| CCKBR | 282.143 | -0.73051 | 0.220615 | -3.31124 | 0.000929 | 0.004908 | 0.602691 |
| ABCA11P | 141.8285 | -0.58511 | 0.176781 | -3.3098 | 0.000934 | 0.004927 | 0.6666 |
| GALR2 | 9.855301 | -2.23341 | 0.67505 | -3.30851 | 0.000938 | 0.004945 | 0.212656 |
| AHNAK | 1467.373 | -0.82764 | 0.250178 | -3.30821 | 0.000939 | 0.004947 | 0.563449 |
| MAN1C1 | 421.2334 | -0.5938 | 0.17986 | -3.30145 | 0.000962 | 0.00505 | 0.662596 |
| CSF1 | 252.0106 | -0.42306 | 0.128213 | -3.29968 | 0.000968 | 0.005078 | 0.74584 |
| GTPBP3 | 891.6449 | -0.46765 | 0.141734 | -3.29951 | 0.000969 | 0.005078 | 0.723141 |
| POLD1 | 3099.124 | -0.55111 | 0.167168 | -3.29676 | 0.000978 | 0.005119 | 0.682494 |
| MT-ATP8 | 15615.35 | -0.49649 | 0.150688 | -3.29481 | 0.000985 | 0.00515 | 0.708831 |
| AC016586 | 387.1923 | -0.43879 | 0.133279 | -3.29224 | 0.000994 | 0.005186 | 0.737754 |
| ZNF506 | 46.77716 | -1.00025 | 0.30385 | -3.29193 | 0.000995 | 0.00519 | 0.499913 |
| DGAT1 | 640.0645 | -0.64933 | 0.197285 | -3.29133 | 0.000997 | 0.005198 | 0.637576 |
| NUDT14 | 385.4126 | -0.55064 | 0.167348 | -3.29037 | 0.001001 | 0.005213 | 0.682719 |
| PDE6B | 254.8327 | -0.63111 | 0.191996 | -3.28708 | 0.001012 | 0.00527 | 0.645681 |
| ACTN2 | 59.95443 | -1.60218 | 0.487465 | -3.28677 | 0.001013 | 0.005274 | 0.329378 |
| INHBC | 33.78707 | -1.09739 | 0.3341 | -3.2846 | 0.001021 | 0.005306 | 0.467363 |
| VPS13C | 1162.431 | -0.41652 | 0.126874 | -3.28294 | 0.001027 | 0.005333 | 0.749229 |
| AL096814. | 21.50135 | -1.73444 | 0.528439 | -3.2822 | 0.00103 | 0.005345 | 0.300526 |
| TPSD1 | 7.245702 | -3.56129 | 1.085565 | -3.28059 | 0.001036 | 0.005374 | 0.084712 |
| MATN4 | 29.6681 | -1.23356 | 0.376134 | -3.27958 | 0.00104 | 0.00539 | 0.425267 |
| IGFBP6 | 105.3729 | -0.72372 | 0.220769 | -3.27817 | 0.001045 | 0.005416 | 0.605535 |
| SNTB1 | 928.3124 | -0.42707 | 0.130314 | -3.27727 | 0.001048 | 0.005431 | 0.743769 |
| SLC16A13 | 112.8966 | -0.60344 | 0.184176 | -3.27641 | 0.001051 | 0.005446 | 0.658184 |
| ANGPTL4 | 93.41524 | -0.6837 | 0.208681 | -3.27628 | 0.001052 | 0.005447 | 0.622568 |
| SIAE | 151.2228 | -0.64017 | 0.195434 | -3.27565 | 0.001054 | 0.005456 | 0.641636 |
| ZDHHC12 | 550.1638 | -0.41762 | 0.127517 | -3.27499 | 0.001057 | 0.005467 | 0.74866 |
| OR14J1 | 92.64546 | -0.62449 | 0.191088 | -3.26806 | 0.001083 | 0.005586 | 0.64865 |
| ITGAM | 60.63837 | -1.46522 | 0.448513 | -3.26684 | 0.001088 | 0.005607 | 0.36218 |
| PAXX | 521.2036 | -0.445 | 0.136453 | -3.26119 | 0.001109 | 0.005698 | 0.734584 |
| TOR2A | 411.4458 | -0.47507 | 0.145672 | -3.26121 | 0.001109 | 0.005698 | 0.719433 |
| C22orf46 | 478.3325 | -0.5037 | 0.154478 | -3.26066 | 0.001112 | 0.005705 | 0.705296 |
| MT-ND3 | 14593.3 | -0.63372 | 0.194372 | -3.26035 | 0.001113 | 0.00571 | 0.644512 |
| NAGPA | 198.1092 | -0.64369 | 0.19744 | -3.26018 | 0.001113 | 0.005711 | 0.640074 |
| ADGRV1 | 1671.528 | -0.46695 | 0.143294 | -3.25868 | 0.001119 | 0.005738 | 0.723493 |
| ULK3 | 726.1184 | -0.52568 | 0.161339 | -3.2582 | 0.001121 | 0.005744 | 0.694633 |
| GFRA2 | 84.8046 | -0.86112 | 0.264307 | -3.25802 | 0.001122 | 0.005746 | 0.550527 |
| DHRS3 | 558.8056 | -0.60386 | 0.185353 | -3.2579 | 0.001122 | 0.005747 | 0.657991 |

| | | | | | | | |
|---|---|---|---|---|---|---|---|
| NUCB1 | 3575.999 | -0.51764 | 0.158903 | -3.25758 | 0.001124 | 0.005752 | 0.698514 |
| DNAJC4 | 373.351 | -0.53437 | 0.164113 | -3.25612 | 0.001129 | 0.005778 | 0.690459 |
| KCNN4 | 27.00711 | -1.17808 | 0.36215 | -3.25301 | 0.001142 | 0.005835 | 0.44194 |
| CRTC2 | 564.1429 | -0.68429 | 0.210514 | -3.25056 | 0.001152 | 0.00588 | 0.622313 |
| SH2D6 | 15.42092 | -2.02826 | 0.624194 | -3.2494 | 0.001156 | 0.005897 | 0.245151 |
| PITPNM1 | 1056.604 | -0.641 | 0.197365 | -3.24779 | 0.001163 | 0.005925 | 0.641268 |
| PARM1 | 118.3651 | -0.76589 | 0.235977 | -3.24559 | 0.001172 | 0.005966 | 0.588092 |
| NAGK | 668.0685 | -0.50204 | 0.15475 | -3.24421 | 0.001178 | 0.005991 | 0.706107 |
| GRINA | 2319.738 | -0.44296 | 0.13681 | -3.23777 | 0.001205 | 0.006114 | 0.735624 |
| CACNG8 | 293.5646 | -0.60126 | 0.185756 | -3.23683 | 0.001209 | 0.006131 | 0.659178 |
| CDK3 | 130.2008 | -0.57919 | 0.179009 | -3.23553 | 0.001214 | 0.006153 | 0.669339 |
| CFAP74 | 18.81957 | -1.33293 | 0.412067 | -3.23474 | 0.001218 | 0.006164 | 0.396961 |
| DGCR2 | 3132.64 | -0.41884 | 0.129547 | -3.23309 | 0.001225 | 0.006193 | 0.748026 |
| NPR2 | 434.1916 | -0.67678 | 0.20934 | -3.23294 | 0.001225 | 0.006194 | 0.625558 |
| LYSMD4 | 292.736 | -0.44571 | 0.137901 | -3.23213 | 0.001229 | 0.006203 | 0.734221 |
| VAMP5 | 12.92965 | -1.65385 | 0.511748 | -3.23176 | 0.00123 | 0.006209 | 0.317792 |
| ADRA2B | 258.4547 | -0.46206 | 0.143032 | -3.23045 | 0.001236 | 0.006234 | 0.72595 |
| NTN5 | 25.1131 | -2.22341 | 0.688562 | -3.22906 | 0.001242 | 0.006259 | 0.214135 |
| LINC0051 | 24.63592 | -1.39743 | 0.432834 | -3.22856 | 0.001244 | 0.006266 | 0.379605 |
| GPR78 | 64.19781 | -1.00482 | 0.311331 | -3.22748 | 0.001249 | 0.006283 | 0.498334 |
| SOD3 | 173.0692 | -0.58454 | 0.181121 | -3.22733 | 0.00125 | 0.006284 | 0.666864 |
| AL390726. | 41.2288 | -1.24844 | 0.386962 | -3.22625 | 0.001254 | 0.006306 | 0.420904 |
| PPT2-EGFL | 19.23671 | -3.00283 | 0.930835 | -3.22595 | 0.001256 | 0.006309 | 0.124755 |
| UCP3 | 14.24224 | -2.00381 | 0.621541 | -3.22394 | 0.001264 | 0.006346 | 0.249341 |
| ANKS3 | 549.6398 | -0.52703 | 0.163535 | -3.22274 | 0.00127 | 0.006365 | 0.693982 |
| RIC3 | 294.9733 | -0.54943 | 0.170872 | -3.21545 | 0.001302 | 0.006512 | 0.68329 |
| LPCAT2 | 105.6356 | -1.06468 | 0.331183 | -3.21476 | 0.001306 | 0.006526 | 0.47808 |
| SLC26A1 | 31.32354 | -2.81382 | 0.875804 | -3.21284 | 0.001314 | 0.00656 | 0.142219 |
| COL9A2 | 188.8093 | -0.88401 | 0.275267 | -3.21147 | 0.001321 | 0.006582 | 0.541858 |
| FBLN1 | 5639.662 | -0.43652 | 0.135924 | -3.21149 | 0.00132 | 0.006582 | 0.738916 |
| AC008507 | 7.823744 | -2.83991 | 0.884636 | -3.21026 | 0.001326 | 0.0066 | 0.139669 |
| ABCB9 | 158.2187 | -0.80368 | 0.250408 | -3.20949 | 0.00133 | 0.006611 | 0.572884 |
| TFR2 | 122.0723 | -0.65326 | 0.203543 | -3.20944 | 0.00133 | 0.006611 | 0.635842 |
| LEAP2 | 40.09704 | -1.07222 | 0.33419 | -3.20842 | 0.001335 | 0.006625 | 0.475586 |
| CLPTM1L | 1697.531 | -0.54769 | 0.170753 | -3.20748 | 0.001339 | 0.006643 | 0.684116 |
| NRADDP | 15.27298 | -1.88355 | 0.587281 | -3.20724 | 0.00134 | 0.006645 | 0.271016 |
| DND1 | 11.43112 | -2.24308 | 0.699403 | -3.20714 | 0.001341 | 0.006645 | 0.211235 |
| VWCE | 196.6649 | -0.81829 | 0.255486 | -3.20289 | 0.001361 | 0.006733 | 0.567112 |
| ALDH16A1 | 1248.299 | -0.42488 | 0.132659 | -3.20277 | 0.001361 | 0.006733 | 0.744902 |
| PLPPR3 | 1790.164 | -0.60372 | 0.188642 | -3.20033 | 0.001373 | 0.006781 | 0.658057 |
| BRSK1 | 893.7411 | -0.68115 | 0.212853 | -3.2001 | 0.001374 | 0.006784 | 0.623667 |
| RRNAD1 | 512.7119 | -0.60553 | 0.189315 | -3.19853 | 0.001381 | 0.006817 | 0.657231 |
| ACTA1 | 95.27376 | -0.86376 | 0.270131 | -3.19757 | 0.001386 | 0.006838 | 0.549518 |
| NFKBID | 115.7629 | -1.15384 | 0.360891 | -3.1972 | 0.001388 | 0.006843 | 0.449427 |

| | | | | | | | |
|---|---|---|---|---|---|---|---|
| TRABD2A | 243.2652 | -0.45949 | 0.143741 | -3.19667 | 0.00139 | 0.00685 | 0.727243 |
| ADCK5 | 129.3501 | -0.68116 | 0.213149 | -3.1957 | 0.001395 | 0.006869 | 0.623663 |
| ACOT11 | 229.9205 | -0.6015 | 0.188302 | -3.19435 | 0.001401 | 0.006895 | 0.659067 |
| KMT5C | 982.6428 | -0.62575 | 0.196053 | -3.19174 | 0.001414 | 0.006946 | 0.648083 |
| BBS5 | 163.9883 | -0.55222 | 0.173059 | -3.19094 | 0.001418 | 0.006959 | 0.68197 |
| MAP3K6 | 349.1595 | -0.57034 | 0.178795 | -3.18988 | 0.001423 | 0.006983 | 0.67346 |
| GPR37 | 193.1688 | -0.57313 | 0.179738 | -3.18869 | 0.001429 | 0.007006 | 0.672158 |
| FANCA | 1758.913 | -0.77876 | 0.244253 | -3.18834 | 0.001431 | 0.007008 | 0.582867 |
| FAM193B | 1412.421 | -0.7341 | 0.230307 | -3.18747 | 0.001435 | 0.007025 | 0.601195 |
| ATF7-NPFF | 25.86604 | -1.32452 | 0.415564 | -3.18727 | 0.001436 | 0.007026 | 0.399283 |
| HGS | 1914.744 | -0.5279 | 0.165681 | -3.18627 | 0.001441 | 0.007045 | 0.693562 |
| SYCP2L | 185.6008 | -0.64413 | 0.202225 | -3.18524 | 0.001446 | 0.007058 | 0.639876 |
| BTBD17 | 288.821 | -0.69378 | 0.218021 | -3.1822 | 0.001462 | 0.007126 | 0.61823 |
| PPP6R2 | 1537.721 | -0.43773 | 0.137569 | -3.18187 | 0.001463 | 0.007132 | 0.738297 |
| SUSD5 | 161.5177 | -0.74484 | 0.234231 | -3.17992 | 0.001473 | 0.007176 | 0.596736 |
| TMEM206 | 490.7899 | -0.46353 | 0.145789 | -3.17948 | 0.001475 | 0.007184 | 0.725208 |
| FBXO24 | 47.02713 | -0.84675 | 0.266395 | -3.17853 | 0.00148 | 0.007202 | 0.556038 |
| HIRIP3 | 536.2698 | -0.45058 | 0.141759 | -3.17847 | 0.001481 | 0.007202 | 0.731751 |
| BDH1 | 383.4451 | -0.43599 | 0.137202 | -3.17776 | 0.001484 | 0.007218 | 0.739184 |
| NUDT22 | 423.4677 | -0.60088 | 0.189249 | -3.17506 | 0.001498 | 0.007277 | 0.659354 |
| OR1A1 | 82.66594 | -0.85934 | 0.270736 | -3.17408 | 0.001503 | 0.0073 | 0.551206 |
| TRPV2 | 47.67423 | -1.17652 | 0.370762 | -3.17326 | 0.001507 | 0.007316 | 0.442416 |
| LEFTY1 | 106.0652 | -0.76053 | 0.239795 | -3.17157 | 0.001516 | 0.00735 | 0.590281 |
| DGAT2 | 358.7993 | -0.61337 | 0.193409 | -3.17135 | 0.001517 | 0.007354 | 0.653668 |
| GFRA3 | 53.92844 | -0.95223 | 0.300415 | -3.16972 | 0.001526 | 0.007391 | 0.516832 |
| LAYN | 154.5553 | -0.55958 | 0.176798 | -3.16509 | 0.00155 | 0.007491 | 0.678499 |
| NFE2L3P1 | 15.81183 | -1.81755 | 0.574294 | -3.16484 | 0.001552 | 0.007495 | 0.283702 |
| FER1L5 | 22.57331 | -1.45102 | 0.458804 | -3.16261 | 0.001564 | 0.007549 | 0.365763 |
| C2orf92 | 36.63176 | -1.28152 | 0.405356 | -3.16146 | 0.00157 | 0.00757 | 0.411363 |
| HTR7 | 1152.311 | -0.44524 | 0.14083 | -3.16159 | 0.001569 | 0.00757 | 0.73446 |
| FLRT2 | 345.4604 | -0.88507 | 0.279988 | -3.1611 | 0.001572 | 0.007575 | 0.541461 |
| MICAL3 | 1204.257 | -0.45575 | 0.144189 | -3.16081 | 0.001573 | 0.007581 | 0.72913 |
| GPR68 | 35.18221 | -1.07608 | 0.340463 | -3.16063 | 0.001574 | 0.007582 | 0.474316 |
| LOXL1 | 369.7609 | -0.44059 | 0.139401 | -3.1606 | 0.001574 | 0.007582 | 0.736833 |
| MFAP4 | 220.5475 | -0.50521 | 0.159901 | -3.15954 | 0.00158 | 0.007603 | 0.704556 |
| NAALADL1 | 98.01435 | -1.01774 | 0.322198 | -3.15875 | 0.001584 | 0.007622 | 0.493889 |
| PTGFRN | 1104.001 | -0.61632 | 0.195179 | -3.1577 | 0.00159 | 0.007641 | 0.652334 |
| INF2 | 1515.613 | -0.53361 | 0.169031 | -3.15688 | 0.001595 | 0.00766 | 0.690824 |
| PRDX4 | 4689.313 | -0.4356 | 0.138066 | -3.15503 | 0.001605 | 0.007698 | 0.739385 |
| PIK3R6 | 68.62706 | -0.81897 | 0.259588 | -3.15488 | 0.001606 | 0.0077 | 0.566846 |
| DCST2 | 28.74507 | -2.23411 | 0.708583 | -3.15292 | 0.001616 | 0.007743 | 0.212552 |
| DLK2 | 22.05585 | -1.44067 | 0.457114 | -3.15166 | 0.001623 | 0.00777 | 0.368397 |
| PMEPA1 | 315.6805 | -0.61515 | 0.195205 | -3.15131 | 0.001625 | 0.007776 | 0.652861 |
| CECR7 | 212.4597 | -0.50502 | 0.160293 | -3.15061 | 0.001629 | 0.00779 | 0.70465 |

| | | | | | | | |
|---|---|---|---|---|---|---|---|
| UACA | 1328.523 | -0.42927 | 0.13635 | -3.14833 | 0.001642 | 0.007838 | 0.742636 |
| MAN2B1 | 2383.483 | -0.41791 | 0.132787 | -3.14722 | 0.001648 | 0.007863 | 0.748509 |
| BX284632. | 15.73287 | -1.51257 | 0.48119 | -3.1434 | 0.00167 | 0.007955 | 0.350486 |
| ACTN3 | 3017.721 | -0.57255 | 0.18219 | -3.1426 | 0.001675 | 0.007975 | 0.672428 |
| EVA1B | 230.6096 | -0.48569 | 0.154606 | -3.1415 | 0.001681 | 0.008002 | 0.714153 |
| DRAM1 | 269.6761 | -0.51778 | 0.164854 | -3.14085 | 0.001685 | 0.008018 | 0.698445 |
| C1R | 376.4589 | -0.47365 | 0.150954 | -3.13772 | 0.001703 | 0.008091 | 0.720139 |
| MUC19 | 23.43395 | -2.70078 | 0.861169 | -3.13618 | 0.001712 | 0.008125 | 0.15381 |
| AC027763 | 16.88121 | -1.53021 | 0.488143 | -3.13476 | 0.00172 | 0.008155 | 0.346227 |
| RBFOX3 | 332.6986 | -0.70755 | 0.225855 | -3.13279 | 0.001732 | 0.00819 | 0.612357 |
| C1orf210 | 213.5767 | -0.5984 | 0.191021 | -3.13262 | 0.001733 | 0.008192 | 0.660487 |
| NXPH4 | 57.99418 | -0.79398 | 0.25352 | -3.13183 | 0.001737 | 0.008204 | 0.576749 |
| FBXO44 | 778.2007 | -0.53848 | 0.172051 | -3.12976 | 0.001749 | 0.008254 | 0.688496 |
| XKR7 | 335.0744 | -0.7249 | 0.231693 | -3.12869 | 0.001756 | 0.008282 | 0.605041 |
| AL031315. | 31.24112 | -1.06044 | 0.338992 | -3.12821 | 0.001759 | 0.008291 | 0.479486 |
| SLC46A3 | 28.14773 | -1.24994 | 0.399562 | -3.12828 | 0.001758 | 0.008291 | 0.420464 |
| COX6B2 | 124.5183 | -0.87572 | 0.280046 | -3.12705 | 0.001766 | 0.008315 | 0.544983 |
| AL121845. | 159.1027 | -0.77885 | 0.249186 | -3.12556 | 0.001775 | 0.008352 | 0.582833 |
| LINC0134 | 123.9174 | -0.59678 | 0.190972 | -3.12496 | 0.001778 | 0.008367 | 0.661229 |
| ALPG | 72.47239 | -0.94906 | 0.303725 | -3.12474 | 0.00178 | 0.008371 | 0.517969 |
| PKD1P5 | 954.5584 | -0.62648 | 0.200548 | -3.12383 | 0.001785 | 0.008379 | 0.647756 |
| ST8SIA3 | 240.9836 | -0.8746 | 0.279969 | -3.12391 | 0.001785 | 0.008379 | 0.545406 |
| LSMEM1 | 24.96094 | -1.52558 | 0.488404 | -3.12361 | 0.001786 | 0.008383 | 0.34734 |
| TECPR1 | 494.8626 | -0.7638 | 0.24465 | -3.12201 | 0.001796 | 0.008417 | 0.588943 |
| PNCK | 280.6963 | -0.87091 | 0.279186 | -3.11944 | 0.001812 | 0.008486 | 0.546803 |
| TAS2R5 | 33.74652 | -1.23389 | 0.395704 | -3.11822 | 0.001819 | 0.008514 | 0.425169 |
| CD38 | 18.50486 | -2.53542 | 0.813464 | -3.11682 | 0.001828 | 0.008548 | 0.17249 |
| REXO1 | 1211.761 | -0.41746 | 0.133941 | -3.11673 | 0.001829 | 0.008548 | 0.748742 |
| LAMA3 | 55.50106 | -1.81944 | 0.583806 | -3.11652 | 0.00183 | 0.008552 | 0.28333 |
| DDIT3 | 536.4343 | -0.60671 | 0.194786 | -3.11474 | 0.001841 | 0.008601 | 0.656694 |
| TMEM191E | 24.81465 | -1.7168 | 0.551489 | -3.11302 | 0.001852 | 0.008647 | 0.304224 |
| DHX34 | 828.5417 | -0.49982 | 0.160616 | -3.11187 | 0.001859 | 0.008674 | 0.707196 |
| MPDU1 | 1466.424 | -0.42918 | 0.13799 | -3.11025 | 0.001869 | 0.008705 | 0.742682 |
| GPC5 | 7.438844 | -2.65714 | 0.854501 | -3.10958 | 0.001874 | 0.008722 | 0.158534 |
| TRIB3 | 1382.963 | -1.2602 | 0.405378 | -3.1087 | 0.001879 | 0.008744 | 0.417487 |
| SDF4 | 2130.092 | -0.46978 | 0.151141 | -3.10824 | 0.001882 | 0.008755 | 0.722073 |
| GAS2L1 | 131.585 | -0.54611 | 0.175729 | -3.1077 | 0.001886 | 0.008766 | 0.684863 |
| LY6E | 722.0089 | -0.85558 | 0.275355 | -3.10718 | 0.001889 | 0.008775 | 0.552644 |
| SEC31B | 199.595 | -0.73615 | 0.236962 | -3.10662 | 0.001892 | 0.008782 | 0.600339 |
| HIST3H2A | 147.6735 | -0.51895 | 0.16727 | -3.10244 | 0.001919 | 0.00889 | 0.697881 |
| IL20RB | 136.0431 | -1.09892 | 0.35435 | -3.10122 | 0.001927 | 0.008919 | 0.466867 |
| SLC30A2 | 16.53904 | -1.96803 | 0.634642 | -3.10101 | 0.001929 | 0.008921 | 0.255602 |
| RUFY4 | 26.75821 | -1.84273 | 0.594369 | -3.10031 | 0.001933 | 0.008935 | 0.278795 |
| MEGF11 | 141.9891 | -0.99877 | 0.322215 | -3.09972 | 0.001937 | 0.008948 | 0.500425 |

| | | | | | | | |
|---|---|---|---|---|---|---|---|
| MIA3 | 1777.375 | -0.50733 | 0.163733 | -3.09854 | 0.001945 | 0.008972 | 0.703522 |
| ENKD1 | 250.7624 | -0.87703 | 0.283167 | -3.09722 | 0.001953 | 0.009003 | 0.544487 |
| VPS9D1 | 186.6098 | -0.74372 | 0.240228 | -3.09589 | 0.001962 | 0.009039 | 0.597198 |
| HS3ST4 | 70.7507 | -0.66573 | 0.2151 | -3.09499 | 0.001968 | 0.009063 | 0.630369 |
| SEMA4C | 3242.475 | -0.45691 | 0.14771 | -3.09328 | 0.00198 | 0.00911 | 0.728545 |
| STAG3 | 266.5006 | -0.58435 | 0.18904 | -3.09117 | 0.001994 | 0.009166 | 0.666948 |
| B4GALNT2 | 155.4581 | -0.49275 | 0.15944 | -3.09052 | 0.001998 | 0.009183 | 0.710667 |
| SPTBN2 | 3297.306 | -1.1044 | 0.357401 | -3.09008 | 0.002001 | 0.009194 | 0.465097 |
| IGSF22 | 18.46206 | -2.81956 | 0.912694 | -3.08927 | 0.002007 | 0.009212 | 0.141654 |
| RNF224 | 11.01164 | -1.81688 | 0.58812 | -3.0893 | 0.002006 | 0.009212 | 0.283835 |
| TSPAN32 | 16.77932 | -2.75262 | 0.891932 | -3.08614 | 0.002028 | 0.009297 | 0.148381 |
| NLRP3P1 | 15.09877 | -2.06953 | 0.671075 | -3.0839 | 0.002043 | 0.00936 | 0.238237 |
| TNFRSF13( | 81.29577 | -0.88311 | 0.286359 | -3.08391 | 0.002043 | 0.00936 | 0.542198 |
| OXCT2 | 48.60781 | -0.98283 | 0.318752 | -3.08337 | 0.002047 | 0.009374 | 0.505986 |
| ISM1 | 25.95477 | -1.06907 | 0.34686 | -3.08213 | 0.002055 | 0.009411 | 0.476626 |
| ITGA2B | 327.9802 | -0.86167 | 0.2796 | -3.08178 | 0.002058 | 0.009417 | 0.550317 |
| SLC15A2 | 173.2417 | -1.12621 | 0.365986 | -3.07719 | 0.00209 | 0.009538 | 0.458118 |
| TCHH | 21.20962 | -1.17838 | 0.383096 | -3.07593 | 0.002098 | 0.009573 | 0.441849 |
| CIC | 1581.58 | -0.48039 | 0.15622 | -3.0751 | 0.002104 | 0.009592 | 0.716782 |
| AJM1 | 65.8022 | -0.97779 | 0.318285 | -3.07207 | 0.002126 | 0.009677 | 0.507756 |
| DRD5P2 | 9.283365 | -2.40484 | 0.782925 | -3.07161 | 0.002129 | 0.00969 | 0.18883 |
| FOXI2 | 62.3 | -0.76805 | 0.250157 | -3.07029 | 0.002139 | 0.009727 | 0.587209 |
| ST7L | 277.3004 | -0.51892 | 0.169069 | -3.06925 | 0.002146 | 0.009756 | 0.697896 |
| MAN1B1 | 1425.4 | -0.44892 | 0.146355 | -3.06735 | 0.00216 | 0.009813 | 0.732591 |
| SMARCA2 | 497.008 | -0.6025 | 0.196641 | -3.06398 | 0.002184 | 0.009911 | 0.65861 |
| FKBP2 | 658.9901 | -0.53465 | 0.17454 | -3.06322 | 0.00219 | 0.009934 | 0.690324 |
| RSPO2 | 20.90831 | -1.63646 | 0.534642 | -3.06085 | 0.002207 | 0.010002 | 0.321645 |
| PDIA2 | 28.11246 | -1.68507 | 0.550615 | -3.06035 | 0.002211 | 0.010015 | 0.310987 |
| VSIR | 5.614682 | -2.68698 | 0.87801 | -3.06031 | 0.002211 | 0.010015 | 0.155288 |
| RGS16 | 493.3819 | -0.78097 | 0.255215 | -3.06004 | 0.002213 | 0.010022 | 0.581976 |
| CYP46A1 | 74.18755 | -1.04025 | 0.339984 | -3.05972 | 0.002215 | 0.010026 | 0.486242 |
| LPAR2 | 800.0535 | -0.59129 | 0.193251 | -3.05969 | 0.002216 | 0.010026 | 0.66375 |
| MS4A4E | 179.8671 | -0.56391 | 0.184332 | -3.05919 | 0.002219 | 0.010034 | 0.676468 |
| GPR137C | 246.7479 | -0.66387 | 0.217066 | -3.05836 | 0.002226 | 0.010057 | 0.631185 |
| NOXA1 | 77.63774 | -0.88115 | 0.288244 | -3.05695 | 0.002236 | 0.010096 | 0.542935 |
| ANKRD20A | 4.13808 | -5.59874 | 1.831635 | -3.05669 | 0.002238 | 0.010101 | 0.020635 |
| CCDC189 | 64.34998 | -1.27516 | 0.417177 | -3.05665 | 0.002238 | 0.010101 | 0.413179 |
| NPIPB14P | 127.5916 | -0.7485 | 0.244947 | -3.05575 | 0.002245 | 0.010126 | 0.595224 |
| MST1R | 158.8239 | -0.85762 | 0.280692 | -3.05536 | 0.002248 | 0.010132 | 0.551864 |
| MXRA5 | 227.0787 | -0.53441 | 0.175038 | -3.05314 | 0.002265 | 0.010196 | 0.690439 |
| CIITA | 75.94385 | -1.23218 | 0.403595 | -3.053 | 0.002266 | 0.010198 | 0.425674 |
| NEGR1 | 145.4235 | -1.13099 | 0.370909 | -3.04923 | 0.002294 | 0.0103 | 0.456604 |
| OR6J1 | 31.65064 | -1.22615 | 0.402338 | -3.04756 | 0.002307 | 0.010352 | 0.427457 |
| SYPL2 | 77.8606 | -0.93724 | 0.307673 | -3.04621 | 0.002317 | 0.01039 | 0.522232 |

| | | | | | | | |
|---|---|---|---|---|---|---|---|
| SLC25A22 | 971.9616 | -0.43777 | 0.143742 | -3.0455 | 0.002323 | 0.01041 | 0.738277 |
| EDC4 | 1895.411 | -0.56995 | 0.187247 | -3.04381 | 0.002336 | 0.010463 | 0.673642 |
| CATSPER2 | 36.77898 | -1.28188 | 0.421329 | -3.04246 | 0.002347 | 0.010499 | 0.41126 |
| CHST12 | 489.2388 | -0.52828 | 0.173644 | -3.04231 | 0.002348 | 0.010501 | 0.693382 |
| ABCA2 | 1051.87 | -0.66209 | 0.217687 | -3.04145 | 0.002354 | 0.010526 | 0.631964 |
| SMIM5 | 12.50388 | -2.0368 | 0.669938 | -3.04028 | 0.002364 | 0.010562 | 0.243704 |
| NKPD1 | 19.29583 | -1.28871 | 0.424285 | -3.03737 | 0.002387 | 0.010651 | 0.409316 |
| IL17RB | 221.6434 | -0.45026 | 0.148336 | -3.03542 | 0.002402 | 0.010703 | 0.731909 |
| SYDE1 | 452.6754 | -0.41909 | 0.138086 | -3.03503 | 0.002405 | 0.010714 | 0.747894 |
| MFAP2 | 3082.923 | -0.48387 | 0.15956 | -3.03252 | 0.002425 | 0.010789 | 0.715056 |
| SYN1 | 238.2454 | -0.59667 | 0.196787 | -3.03205 | 0.002429 | 0.010798 | 0.66128 |
| TES | 861.6277 | -0.51983 | 0.171542 | -3.03034 | 0.002443 | 0.010851 | 0.697454 |
| TTLL2 | 15.51183 | -2.05183 | 0.677683 | -3.02771 | 0.002464 | 0.010926 | 0.241178 |
| SLC45A3 | 553.0995 | -0.4285 | 0.141717 | -3.02365 | 0.002497 | 0.011054 | 0.743033 |
| CAPN10 | 820.0359 | -0.67286 | 0.222578 | -3.02301 | 0.002503 | 0.011071 | 0.627264 |
| FBN3 | 2239.227 | -0.70149 | 0.23207 | -3.02274 | 0.002505 | 0.011078 | 0.614938 |
| TSGA10IP | 115.1563 | -0.58867 | 0.194831 | -3.02143 | 0.002516 | 0.011115 | 0.664957 |
| RAPGEF3 | 129.5387 | -0.51222 | 0.169808 | -3.01645 | 0.002558 | 0.011283 | 0.701144 |
| ICA1L | 211.8654 | -0.55688 | 0.184642 | -3.01598 | 0.002561 | 0.011293 | 0.679772 |
| SOX15 | 63.70182 | -0.72664 | 0.240934 | -3.01593 | 0.002562 | 0.011293 | 0.60431 |
| PPOX | 612.825 | -0.53667 | 0.178106 | -3.01319 | 0.002585 | 0.011375 | 0.689362 |
| SSR4 | 1651.94 | -0.42512 | 0.141099 | -3.01292 | 0.002588 | 0.011382 | 0.744776 |
| ANKZF1 | 1308.842 | -0.44847 | 0.148891 | -3.01209 | 0.002595 | 0.01141 | 0.732818 |
| RRN3P1 | 231.9659 | -0.54248 | 0.180163 | -3.01107 | 0.002603 | 0.011446 | 0.686589 |
| LRRC56 | 47.93945 | -0.85216 | 0.283126 | -3.00983 | 0.002614 | 0.011484 | 0.553954 |
| PNPLA7 | 58.73316 | -1.00702 | 0.334605 | -3.00957 | 0.002616 | 0.011491 | 0.497573 |
| SLC44A3 | 67.25133 | -0.85579 | 0.284499 | -3.00806 | 0.002629 | 0.011539 | 0.552562 |
| TUBA3D | 4.912071 | -3.11437 | 1.03559 | -3.00734 | 0.002635 | 0.011564 | 0.115473 |
| RNF215 | 507.7192 | -0.55627 | 0.185088 | -3.00544 | 0.002652 | 0.011621 | 0.680057 |
| TDGF1P3 | 128.1271 | -0.63465 | 0.211168 | -3.00544 | 0.002652 | 0.011621 | 0.644096 |
| ENG | 123.6137 | -0.86286 | 0.287277 | -3.00359 | 0.002668 | 0.011683 | 0.54986 |
| TP53TG5 | 14.08316 | -2.46847 | 0.822699 | -3.00045 | 0.002696 | 0.011792 | 0.180683 |
| AC074141 | 44.6352 | -0.98929 | 0.330036 | -2.99754 | 0.002722 | 0.011894 | 0.503724 |
| SGCG | 13.00686 | -1.65713 | 0.553206 | -2.9955 | 0.00274 | 0.011967 | 0.31707 |
| C11orf80 | 718.0742 | -0.48404 | 0.161705 | -2.99336 | 0.002759 | 0.012036 | 0.714972 |
| TBX6 | 94.9192 | -0.60559 | 0.202415 | -2.99181 | 0.002773 | 0.012091 | 0.657204 |
| IRAK4 | 211.4971 | -0.50696 | 0.16947 | -2.99146 | 0.002776 | 0.012102 | 0.703701 |
| VWA1 | 1396.438 | -0.46784 | 0.156411 | -2.99108 | 0.00278 | 0.012113 | 0.723048 |
| CHIC2 | 138.34 | -0.53745 | 0.179781 | -2.98949 | 0.002794 | 0.012163 | 0.688986 |
| ZMYND15 | 15.91479 | -1.36657 | 0.457303 | -2.98832 | 0.002805 | 0.012203 | 0.387812 |
| TRPM5 | 5.267316 | -2.76395 | 0.925398 | -2.98676 | 0.002819 | 0.012262 | 0.147221 |
| FCER2 | 42.19232 | -0.91723 | 0.307118 | -2.98657 | 0.002821 | 0.012266 | 0.529525 |
| MFSD6L | 76.54548 | -0.66737 | 0.223571 | -2.98507 | 0.002835 | 0.012314 | 0.629652 |
| AL513477. | 26.60799 | -1.10682 | 0.370799 | -2.98495 | 0.002836 | 0.012316 | 0.464317 |

| | | | | | | | |
|---|---|---|---|---|---|---|---|
| IL17RA | 429.2839 | -0.41622 | 0.139479 | -2.98413 | 0.002844 | 0.012346 | 0.749384 |
| IL18BP | 157.3262 | -0.60585 | 0.203033 | -2.98398 | 0.002845 | 0.012349 | 0.657086 |
| AC243772 | 23.83619 | -1.23777 | 0.414909 | -2.98323 | 0.002852 | 0.01237 | 0.424027 |
| PCNT | 1462.358 | -0.73281 | 0.245703 | -2.98251 | 0.002859 | 0.012393 | 0.601729 |
| POMT1 | 1191.612 | -1.09831 | 0.368318 | -2.98196 | 0.002864 | 0.012406 | 0.467064 |
| ADAM33 | 313.7443 | -0.82074 | 0.275363 | -2.98059 | 0.002877 | 0.012452 | 0.56615 |
| CCDC194 | 6.938617 | -2.55568 | 0.858241 | -2.97781 | 0.002903 | 0.01254 | 0.170084 |
| SMARCC2 | 2404.035 | -0.45728 | 0.153579 | -2.9775 | 0.002906 | 0.012547 | 0.728358 |
| REC8 | 46.83866 | -1.61039 | 0.540991 | -2.97674 | 0.002913 | 0.012575 | 0.327511 |
| NPIPB7 | 61.07196 | -0.93065 | 0.312953 | -2.97377 | 0.002942 | 0.012694 | 0.524622 |
| CELSR1 | 1807.577 | -0.49206 | 0.165919 | -2.96564 | 0.003021 | 0.013002 | 0.711011 |
| SCN11A | 36.40453 | -0.88276 | 0.297694 | -2.96531 | 0.003024 | 0.013009 | 0.54233 |
| ZNF585A | 161.7084 | -0.6125 | 0.206669 | -2.9637 | 0.00304 | 0.013074 | 0.654061 |
| TSEN54 | 745.3921 | -0.43068 | 0.145584 | -2.95828 | 0.003094 | 0.013273 | 0.741914 |
| CYP2C8 | 32.49866 | -1.2637 | 0.427504 | -2.956 | 0.003117 | 0.013352 | 0.416474 |
| GSTM3P1 | 15.87985 | -1.59209 | 0.538591 | -2.95603 | 0.003116 | 0.013352 | 0.33169 |
| PCDHA7 | 20.85553 | -1.55252 | 0.525229 | -2.95588 | 0.003118 | 0.013353 | 0.340915 |
| AACSP1 | 16.25707 | -1.43056 | 0.484025 | -2.95555 | 0.003121 | 0.013364 | 0.370986 |
| ADCY6 | 1668.063 | -0.5245 | 0.17754 | -2.95424 | 0.003134 | 0.013413 | 0.695202 |
| SDCCAG8 | 109.6175 | -0.8556 | 0.289737 | -2.95303 | 0.003147 | 0.013451 | 0.552635 |
| PAQR7 | 215.8259 | -0.51147 | 0.173282 | -2.95167 | 0.003161 | 0.013503 | 0.701506 |
| AP1G2 | 1354.85 | -1.05575 | 0.357769 | -2.95093 | 0.003168 | 0.013524 | 0.481047 |
| FXYD6-FXY | 4.164939 | -4.15269 | 1.40722 | -2.95099 | 0.003168 | 0.013524 | 0.056223 |
| TARBP1 | 1135.507 | -0.51046 | 0.172985 | -2.9509 | 0.003168 | 0.013524 | 0.701997 |
| HTRA1 | 497.2713 | -0.48711 | 0.165085 | -2.95067 | 0.003171 | 0.01353 | 0.713452 |
| AL590426. | 6.698641 | -2.36882 | 0.80311 | -2.94956 | 0.003182 | 0.013572 | 0.193604 |
| LAG3 | 69.95689 | -0.83792 | 0.284118 | -2.94921 | 0.003186 | 0.013581 | 0.559449 |
| LDHA | 10365.09 | -0.52922 | 0.179467 | -2.94882 | 0.00319 | 0.013595 | 0.692931 |
| AC018638 | 57.58023 | -0.97295 | 0.329966 | -2.94865 | 0.003192 | 0.013595 | 0.509462 |
| CCDC114 | 87.91805 | -0.79835 | 0.270847 | -2.94762 | 0.003202 | 0.013626 | 0.575005 |
| KLK14 | 20.61713 | -1.60199 | 0.543491 | -2.94759 | 0.003203 | 0.013626 | 0.329423 |
| AC025283 | 10.94401 | -7.01265 | 2.384193 | -2.94131 | 0.003268 | 0.013884 | 0.007744 |
| GPR82 | 14.45825 | -1.48983 | 0.506537 | -2.9412 | 0.003269 | 0.013886 | 0.356056 |
| UPK3BL1 | 387.417 | -0.63245 | 0.215043 | -2.94102 | 0.003271 | 0.01389 | 0.645082 |
| ACSL6 | 116.0032 | -0.75312 | 0.256208 | -2.93947 | 0.003288 | 0.01395 | 0.593321 |
| TTC9B | 5.644843 | -3.4844 | 1.185577 | -2.93899 | 0.003293 | 0.013968 | 0.089349 |
| MT-CYB | 85759.75 | -0.95807 | 0.326241 | -2.93669 | 0.003317 | 0.014048 | 0.514746 |
| PPP1R36 | 21.50363 | -1.37255 | 0.467457 | -2.93621 | 0.003322 | 0.014058 | 0.386207 |
| AGA | 383.5134 | -0.661 | 0.225164 | -2.93563 | 0.003329 | 0.014075 | 0.63244 |
| PITPNM3 | 397.2539 | -0.63566 | 0.216542 | -2.93548 | 0.00333 | 0.014078 | 0.643648 |
| GTF3C1 | 4870.535 | -0.48345 | 0.16478 | -2.93388 | 0.003348 | 0.014141 | 0.715268 |
| LRRC37A2 | 306.996 | -0.45242 | 0.154257 | -2.93288 | 0.003358 | 0.014176 | 0.730818 |
| ANKRD54 | 306.2643 | -0.42876 | 0.146226 | -2.93221 | 0.003366 | 0.0142 | 0.742898 |
| AC009133 | 611.2194 | -0.64151 | 0.219299 | -2.92527 | 0.003442 | 0.014481 | 0.641042 |

| | | | | | | | |
|---|---|---|---|---|---|---|---|
| ENPP2 | 59.36055 | -0.85863 | 0.293571 | -2.92476 | 0.003447 | 0.014501 | 0.551477 |
| RNFT2 | 435.3032 | -0.47108 | 0.161116 | -2.92382 | 0.003458 | 0.014528 | 0.721427 |
| GAK | 1796.832 | -0.41981 | 0.143603 | -2.92339 | 0.003462 | 0.014544 | 0.747524 |
| CACNA1D | 52.0561 | -0.9156 | 0.313218 | -2.9232 | 0.003465 | 0.014546 | 0.530124 |
| LRFN4 | 1126.141 | -0.49473 | 0.169309 | -2.92207 | 0.003477 | 0.014592 | 0.709693 |
| LRG1 | 27.15466 | -0.97896 | 0.33514 | -2.92105 | 0.003489 | 0.014629 | 0.507345 |
| CCDC162I | 118.3173 | -0.80087 | 0.274317 | -2.91951 | 0.003506 | 0.014683 | 0.574002 |
| FRK | 197.4796 | -0.50853 | 0.174224 | -2.9188 | 0.003514 | 0.014706 | 0.702941 |
| SLIT2 | 143.2091 | -0.46325 | 0.158772 | -2.9177 | 0.003526 | 0.014751 | 0.725351 |
| ADCY9 | 405.3646 | -0.60114 | 0.206129 | -2.91633 | 0.003542 | 0.014809 | 0.659233 |
| COL6A4P2 | 21.76907 | -1.75937 | 0.603697 | -2.91432 | 0.003565 | 0.014883 | 0.295378 |
| GALNS | 428.8546 | -0.55495 | 0.190423 | -2.91429 | 0.003565 | 0.014883 | 0.680681 |
| ZACN | 17.15938 | -2.70301 | 0.927516 | -2.91425 | 0.003565 | 0.014883 | 0.153572 |
| AIFM2 | 197.8089 | -0.44307 | 0.152058 | -2.9138 | 0.003571 | 0.0149 | 0.73557 |
| BRICD5 | 118.3465 | -1.30654 | 0.448674 | -2.91201 | 0.003591 | 0.014975 | 0.404289 |
| CCT6P3 | 215.0878 | -0.67138 | 0.230656 | -2.91076 | 0.003605 | 0.015027 | 0.627904 |
| REEP6 | 402.3098 | -0.45602 | 0.156669 | -2.91071 | 0.003606 | 0.015027 | 0.728996 |
| GALNT18 | 235.4989 | -0.4408 | 0.151653 | -2.90662 | 0.003654 | 0.015206 | 0.736727 |
| LIF | 131.403 | -0.67104 | 0.230909 | -2.9061 | 0.00366 | 0.015228 | 0.628052 |
| ZDHHC22 | 713.6577 | -0.43631 | 0.150173 | -2.90542 | 0.003668 | 0.015257 | 0.73902 |
| NME8 | 4.942743 | -2.73461 | 0.941451 | -2.90468 | 0.003676 | 0.015283 | 0.150245 |
| TPM2 | 2331.205 | -0.4419 | 0.152214 | -2.90314 | 0.003694 | 0.015346 | 0.736165 |
| CCDC144/ | 143.7286 | -0.5953 | 0.205102 | -2.90248 | 0.003702 | 0.015372 | 0.661905 |
| ZCWPW1 | 58.34734 | -0.86925 | 0.299604 | -2.90132 | 0.003716 | 0.01541 | 0.547433 |
| PQLC2 | 486.6294 | -0.51412 | 0.177242 | -2.90067 | 0.003724 | 0.015431 | 0.70022 |
| C19orf71 | 25.55647 | -1.46101 | 0.504134 | -2.89805 | 0.003755 | 0.015545 | 0.36324 |
| FAM174A | 106.5757 | -0.58779 | 0.202961 | -2.89606 | 0.003779 | 0.015633 | 0.665362 |
| CSF3R | 4.980513 | -4.79171 | 1.656519 | -2.89264 | 0.00382 | 0.015774 | 0.036104 |
| ALDH3B2 | 6.269654 | -3.5317 | 1.221531 | -2.89121 | 0.003838 | 0.01582 | 0.086467 |
| CEP131 | 767.3817 | -0.64974 | 0.224731 | -2.89119 | 0.003838 | 0.01582 | 0.637395 |
| TMEM184/ | 63.8501 | -1.52314 | 0.527026 | -2.89006 | 0.003852 | 0.015862 | 0.347928 |
| ANKRD30E | 5.240298 | -2.96913 | 1.027389 | -2.88998 | 0.003853 | 0.015863 | 0.127703 |
| MSANTD1 | 12.08418 | -1.83929 | 0.637388 | -2.88567 | 0.003906 | 0.016039 | 0.279459 |
| SPIB | 206.6571 | -0.89253 | 0.309767 | -2.88129 | 0.003961 | 0.016241 | 0.53867 |
| TMEM208 | 527.2591 | -0.42927 | 0.149067 | -2.87971 | 0.00398 | 0.016307 | 0.742637 |
| TMEM39B | 457.0989 | -0.48267 | 0.167617 | -2.87962 | 0.003982 | 0.016307 | 0.71565 |
| TCL1A | 3.778264 | -3.96377 | 1.376999 | -2.87856 | 0.003995 | 0.016351 | 0.064089 |
| KLF2P1 | 13.26457 | -1.40776 | 0.489435 | -2.8763 | 0.004024 | 0.016448 | 0.376896 |
| CLU | 14962.35 | -0.49022 | 0.170488 | -2.87538 | 0.004035 | 0.016481 | 0.711917 |
| PID1 | 194.7826 | -0.52758 | 0.18358 | -2.87384 | 0.004055 | 0.016542 | 0.693718 |
| COCH | 566.0369 | -0.45196 | 0.157345 | -2.87241 | 0.004074 | 0.016605 | 0.731049 |
| ZNF26 | 333.8581 | -0.42258 | 0.147121 | -2.87232 | 0.004075 | 0.016606 | 0.74609 |
| POMC | 56.32016 | -0.78189 | 0.272446 | -2.86989 | 0.004106 | 0.016707 | 0.581605 |
| NPIPB15 | 111.1722 | -0.66462 | 0.231679 | -2.86872 | 0.004121 | 0.016757 | 0.630854 |

| | | | | | | | |
|---|---|---|---|---|---|---|---|
| CFAP300 | 7.124446 | -3.25353 | 1.134183 | -2.86861 | 0.004123 | 0.016759 | 0.104855 |
| OR9H1P | 22.58411 | -1.04304 | 0.363777 | -2.86725 | 0.004141 | 0.016827 | 0.485304 |
| CCDC74B | 295.136 | -0.48974 | 0.170887 | -2.86587 | 0.004159 | 0.016889 | 0.712154 |
| ZNF471 | 68.07879 | -0.76502 | 0.267031 | -2.86491 | 0.004171 | 0.016931 | 0.588445 |
| SP140 | 8.727742 | -2.32815 | 0.813138 | -2.86317 | 0.004194 | 0.017009 | 0.199139 |
| MAB21L3 | 91.62272 | -0.58736 | 0.205209 | -2.86228 | 0.004206 | 0.017041 | 0.665558 |
| CCER2 | 13.81897 | -1.6116 | 0.563392 | -2.86054 | 0.004229 | 0.017123 | 0.327234 |
| ZNF93 | 5.567078 | -2.87068 | 1.004191 | -2.8587 | 0.004254 | 0.017211 | 0.136722 |
| SLC16A6 | 21.84168 | -1.1531 | 0.403395 | -2.85848 | 0.004257 | 0.017218 | 0.449659 |
| CLEC4D | 7.287876 | -2.20201 | 0.770751 | -2.85697 | 0.004277 | 0.017292 | 0.217334 |
| SLC25A29 | 3214.692 | -0.45116 | 0.15805 | -2.85456 | 0.00431 | 0.017411 | 0.731453 |
| DGKA | 439.2824 | -0.41513 | 0.145435 | -2.85439 | 0.004312 | 0.017417 | 0.749952 |
| CICP14 | 154.4216 | -0.74873 | 0.262513 | -2.85216 | 0.004342 | 0.017524 | 0.595127 |
| MIOX | 13.4635 | -1.54743 | 0.542548 | -2.85216 | 0.004342 | 0.017524 | 0.342119 |
| AC114267 | 91.50536 | -0.64319 | 0.225666 | -2.85017 | 0.00437 | 0.017626 | 0.640297 |
| CLTCL1 | 708.9699 | -0.42224 | 0.148411 | -2.84508 | 0.00444 | 0.017867 | 0.746265 |
| TCTN1 | 359.0093 | -0.46412 | 0.163143 | -2.84485 | 0.004443 | 0.017876 | 0.724915 |
| PATL2 | 18.14259 | -1.26975 | 0.446426 | -2.84425 | 0.004452 | 0.017893 | 0.414732 |
| NPIPB2 | 40.45469 | -1.12334 | 0.395004 | -2.84387 | 0.004457 | 0.01791 | 0.459029 |
| TEDC2 | 940.4013 | -0.53041 | 0.186709 | -2.84086 | 0.004499 | 0.018063 | 0.692357 |
| NLRC5 | 103.4678 | -0.93836 | 0.330431 | -2.83982 | 0.004514 | 0.018117 | 0.521824 |
| RNPEPL1 | 467.3901 | -0.45028 | 0.158713 | -2.83706 | 0.004553 | 0.01825 | 0.731902 |
| SDSL | 40.7772 | -1.01721 | 0.358541 | -2.83709 | 0.004553 | 0.01825 | 0.49407 |
| FAM228B | 206.8834 | -0.42284 | 0.149168 | -2.83463 | 0.004588 | 0.018367 | 0.745957 |
| IL2RB | 52.25317 | -0.9559 | 0.337534 | -2.83201 | 0.004626 | 0.018487 | 0.51552 |
| TCAP | 37.36208 | -1.49577 | 0.528305 | -2.83126 | 0.004637 | 0.018519 | 0.354592 |
| ZFYVE28 | 89.96684 | -0.68849 | 0.243358 | -2.82914 | 0.004667 | 0.018621 | 0.620501 |
| RASIP1 | 213.7081 | -0.56484 | 0.199764 | -2.82754 | 0.004691 | 0.018692 | 0.67603 |
| ADAM20P1 | 14.13986 | -1.72866 | 0.611887 | -2.82512 | 0.004726 | 0.018816 | 0.301733 |
| FAM156A | 259.589 | -0.52079 | 0.184369 | -2.82474 | 0.004732 | 0.018826 | 0.696989 |
| AC092865 | 21.10504 | -1.12629 | 0.398736 | -2.82466 | 0.004733 | 0.018826 | 0.458091 |
| TVP23A | 176.3322 | -0.50583 | 0.179233 | -2.8222 | 0.00477 | 0.018941 | 0.704255 |
| TMEM175 | 127.7469 | -0.47876 | 0.169663 | -2.82182 | 0.004775 | 0.018959 | 0.717596 |
| RDH5 | 37.93853 | -0.92294 | 0.327202 | -2.82069 | 0.004792 | 0.019013 | 0.527435 |
| USP6 | 56.07709 | -0.91512 | 0.324464 | -2.82041 | 0.004796 | 0.019021 | 0.5303 |
| PLEKHG5 | 738.7766 | -0.45674 | 0.161944 | -2.82033 | 0.004797 | 0.019021 | 0.728633 |
| FSD2 | 42.76545 | -0.90963 | 0.322545 | -2.82017 | 0.0048 | 0.019026 | 0.532321 |
| GIPR | 26.53891 | -1.31428 | 0.466236 | -2.81892 | 0.004819 | 0.019087 | 0.402126 |
| FOXJ1 | 73.84804 | -0.7799 | 0.276854 | -2.81701 | 0.004847 | 0.019188 | 0.582407 |
| SLC29A2 | 689.0568 | -0.5263 | 0.186871 | -2.81638 | 0.004857 | 0.019212 | 0.694333 |
| FOXL2NB | 433.4449 | -0.72692 | 0.258213 | -2.8152 | 0.004875 | 0.01927 | 0.604192 |
| GOLGA6L4 | 79.31778 | -0.85661 | 0.304373 | -2.81434 | 0.004888 | 0.019312 | 0.552249 |
| RECQL4 | 2167.979 | -0.97468 | 0.346783 | -2.81063 | 0.004944 | 0.019492 | 0.508853 |
| HLA-K | 6.413486 | -4.17122 | 1.48415 | -2.81051 | 0.004946 | 0.019495 | 0.055506 |

| | | | | | | | |
|---|---|---|---|---|---|---|---|
| FAIM2 | 61.00198 | -1.45556 | 0.518086 | -2.8095 | 0.004962 | 0.019538 | 0.364613 |
| GDPD3 | 23.18849 | -1.30199 | 0.463889 | -2.80669 | 0.005005 | 0.019682 | 0.405566 |
| PNPLA5 | 119.2 | -0.52114 | 0.185763 | -2.8054 | 0.005025 | 0.019747 | 0.696821 |
| CCDC184 | 58.60362 | -0.96252 | 0.343321 | -2.80355 | 0.005054 | 0.019848 | 0.51316 |
| SVOPL | 96.12344 | -0.71382 | 0.254628 | -2.80338 | 0.005057 | 0.019854 | 0.609704 |
| UNC93B1 | 502.28 | -0.44796 | 0.159828 | -2.80277 | 0.005067 | 0.019877 | 0.733077 |
| TRPM2 | 7.131343 | -2.32426 | 0.830529 | -2.79853 | 0.005134 | 0.020086 | 0.199677 |
| ADAMTS15 | 112.8346 | -0.59286 | 0.212059 | -2.79575 | 0.005178 | 0.020246 | 0.663026 |
| GALNT4 | 143.928 | -0.78174 | 0.279678 | -2.79515 | 0.005188 | 0.020274 | 0.581664 |
| ABCA7 | 1055.302 | -1.15291 | 0.412723 | -2.79342 | 0.005215 | 0.020359 | 0.449717 |
| PRSS53 | 168.4429 | -1.69368 | 0.60648 | -2.79264 | 0.005228 | 0.020404 | 0.309136 |
| PAN2 | 1021.969 | -0.57697 | 0.206716 | -2.79111 | 0.005253 | 0.020477 | 0.670372 |
| PDZD3 | 7.46578 | -2.71217 | 0.971883 | -2.79063 | 0.005261 | 0.020498 | 0.1526 |
| AC124944 | 162.3111 | -0.57344 | 0.205608 | -2.78901 | 0.005287 | 0.020583 | 0.672011 |
| ST5 | 1215.521 | -0.42448 | 0.152264 | -2.78778 | 0.005307 | 0.020633 | 0.745107 |
| DGCR8 | 1492.184 | -0.43287 | 0.15529 | -2.78751 | 0.005311 | 0.020645 | 0.740785 |
| CFTR | 87.96067 | -0.7901 | 0.283571 | -2.78623 | 0.005332 | 0.020713 | 0.578306 |
| HGSNAT | 420.6215 | -0.50997 | 0.18319 | -2.78382 | 0.005372 | 0.020828 | 0.702238 |
| HIVEP2 | 399.761 | -0.86858 | 0.312023 | -2.7837 | 0.005374 | 0.020828 | 0.547686 |
| MCOLN2 | 85.30264 | -0.98639 | 0.354343 | -2.78371 | 0.005374 | 0.020828 | 0.50474 |
| AC092299 | 137.1778 | -2.86296 | 1.030396 | -2.7785 | 0.005461 | 0.021089 | 0.137456 |
| ARHGDIG | 40.91578 | -0.87196 | 0.313923 | -2.77763 | 0.005476 | 0.021113 | 0.546403 |
| E2F3P2 | 29.84057 | -1.05984 | 0.38152 | -2.77795 | 0.00547 | 0.021113 | 0.479684 |
| USP32P3 | 129.7325 | -0.61732 | 0.222248 | -2.77764 | 0.005475 | 0.021113 | 0.651879 |
| SPDYE3 | 76.59383 | -0.64328 | 0.231607 | -2.77746 | 0.005478 | 0.021119 | 0.640255 |
| NPIPA7 | 116.3377 | -0.9711 | 0.349657 | -2.77728 | 0.005482 | 0.021126 | 0.510119 |
| ZNF542P | 447.9909 | -0.52537 | 0.189214 | -2.77657 | 0.005494 | 0.021153 | 0.694782 |
| DLL4 | 19.67953 | -1.3095 | 0.47187 | -2.77513 | 0.005518 | 0.021233 | 0.40346 |
| NLGN2 | 3964.915 | -0.55113 | 0.198633 | -2.7746 | 0.005527 | 0.021258 | 0.682487 |
| TRMU | 1129.176 | -0.42547 | 0.153409 | -2.77341 | 0.005547 | 0.021326 | 0.744598 |
| IGSF10 | 60.20047 | -1.12764 | 0.406618 | -2.77322 | 0.005551 | 0.02133 | 0.457664 |
| SLCO4A1 | 294.8757 | -0.50444 | 0.181985 | -2.77189 | 0.005573 | 0.021393 | 0.704933 |
| AMY2B | 49.79527 | -0.86043 | 0.310501 | -2.77109 | 0.005587 | 0.021441 | 0.55079 |
| CYP24A1 | 10.62379 | -4.40653 | 1.590779 | -2.77005 | 0.005605 | 0.021486 | 0.047152 |
| AGAP2 | 109.3861 | -1.14629 | 0.414078 | -2.76831 | 0.005635 | 0.021573 | 0.451784 |
| CDH24 | 652.6415 | -0.48956 | 0.17686 | -2.76805 | 0.005639 | 0.02158 | 0.712243 |
| SHOX2 | 12.67527 | -2.31349 | 0.836509 | -2.76565 | 0.005681 | 0.021705 | 0.201173 |
| TMEM80 | 207.5919 | -0.42572 | 0.154004 | -2.76436 | 0.005703 | 0.021781 | 0.744466 |
| FLRT1 | 34.361 | -0.87319 | 0.315903 | -2.76409 | 0.005708 | 0.021794 | 0.54594 |
| MT-CO1 | 457914.4 | -0.87187 | 0.315576 | -2.7628 | 0.005731 | 0.021872 | 0.546437 |
| PLXNB3 | 81.62144 | -1.27698 | 0.462207 | -2.76279 | 0.005731 | 0.021872 | 0.412658 |
| NLRP6 | 9.683264 | -1.96154 | 0.710288 | -2.76162 | 0.005752 | 0.021926 | 0.256754 |
| SATL1 | 77.41857 | -0.57641 | 0.208845 | -2.76001 | 0.00578 | 0.022005 | 0.670628 |
| OXER1 | 50.11692 | -0.78659 | 0.285107 | -2.75892 | 0.005799 | 0.02206 | 0.579714 |

| | | | | | | | |
|---|---|---|---|---|---|---|---|
| MMP9 | 101.7158 | -0.56642 | 0.205491 | -2.75642 | 0.005844 | 0.022219 | 0.675291 |
| GSDMD | 684.9968 | -0.73738 | 0.267632 | -2.75521 | 0.005866 | 0.022287 | 0.599827 |
| GOLGA6L5 | 82.56767 | -0.79394 | 0.288216 | -2.75466 | 0.005875 | 0.022319 | 0.576768 |
| ANKDD1A | 205.8397 | -0.66105 | 0.240009 | -2.75429 | 0.005882 | 0.022336 | 0.632416 |
| SLC25A45 | 28.77838 | -1.14859 | 0.417324 | -2.75228 | 0.005918 | 0.022442 | 0.451065 |
| CT62 | 9.908351 | -2.2892 | 0.832214 | -2.75073 | 0.005946 | 0.022539 | 0.204589 |
| AMDHD2 | 1090.944 | -0.56229 | 0.204519 | -2.74934 | 0.005972 | 0.022615 | 0.677225 |
| IDUA | 102.1564 | -0.9392 | 0.34162 | -2.74925 | 0.005973 | 0.022615 | 0.521522 |
| PDGFB | 151.9463 | -0.80411 | 0.292466 | -2.7494 | 0.00597 | 0.022615 | 0.572716 |
| TMEM37 | 253.1713 | -0.47129 | 0.171538 | -2.74742 | 0.006007 | 0.022723 | 0.721321 |
| FGD5 | 766.9986 | -0.49387 | 0.179787 | -2.74696 | 0.006015 | 0.022744 | 0.710118 |
| AC010422 | 140.5886 | -0.75721 | 0.2757 | -2.74651 | 0.006023 | 0.022756 | 0.591638 |
| LIMCH1 | 633.1037 | -0.55264 | 0.201228 | -2.74633 | 0.006027 | 0.022762 | 0.681773 |
| ZNF815P | 72.42057 | -0.67395 | 0.245412 | -2.74618 | 0.006029 | 0.022764 | 0.62679 |
| SLC38A6 | 167.055 | -0.56617 | 0.206283 | -2.74462 | 0.006058 | 0.022867 | 0.675408 |
| REEP1 | 45.8219 | -0.79356 | 0.289185 | -2.74412 | 0.006067 | 0.022892 | 0.576919 |
| AC112206 | 6.800407 | -2.16664 | 0.790764 | -2.73993 | 0.006145 | 0.02314 | 0.222729 |
| AL365205. | 97.88906 | -0.61419 | 0.224336 | -2.7378 | 0.006185 | 0.023265 | 0.653298 |
| ADAM11 | 305.5497 | -0.85796 | 0.313393 | -2.73764 | 0.006188 | 0.02327 | 0.551733 |
| MRGPRF | 246.0171 | -0.4893 | 0.178766 | -2.73711 | 0.006198 | 0.023289 | 0.71237 |
| HIST1H2A( | 271.0436 | -1.02714 | 0.375329 | -2.73663 | 0.006207 | 0.023318 | 0.490683 |
| LGI4 | 159.538 | -0.72196 | 0.263965 | -2.73505 | 0.006237 | 0.023414 | 0.606275 |
| ALKBH6 | 340.34 | -0.42604 | 0.155947 | -2.73196 | 0.006296 | 0.023608 | 0.744301 |
| PLAC9 | 68.81401 | -0.7457 | 0.27296 | -2.73191 | 0.006297 | 0.023608 | 0.596378 |
| AOC3 | 87.56437 | -0.79846 | 0.292302 | -2.73161 | 0.006303 | 0.023624 | 0.574964 |
| TSPAN10 | 92.09459 | -0.73853 | 0.270463 | -2.73061 | 0.006322 | 0.023681 | 0.599351 |
| LENG8 | 2199.931 | -0.49956 | 0.183106 | -2.72823 | 0.006368 | 0.023842 | 0.707325 |
| SETD4 | 562.4331 | -0.48994 | 0.179593 | -2.72805 | 0.006371 | 0.02385 | 0.712055 |
| C1S | 341.1442 | -0.51609 | 0.189212 | -2.72758 | 0.00638 | 0.023874 | 0.699265 |
| FAM110D | 43.07044 | -0.79028 | 0.289827 | -2.72674 | 0.006396 | 0.023913 | 0.578231 |
| MCOLN3 | 297.5038 | -0.43943 | 0.161267 | -2.72484 | 0.006433 | 0.024025 | 0.737428 |
| RYR3 | 72.10624 | -1.09387 | 0.401518 | -2.72435 | 0.006443 | 0.024049 | 0.468502 |
| SLC24A4 | 30.59947 | -1.12947 | 0.41459 | -2.7243 | 0.006444 | 0.024049 | 0.457084 |
| PTK6 | 30.00176 | -0.87959 | 0.322937 | -2.72371 | 0.006455 | 0.024087 | 0.543523 |
| NBPF25P | 32.4099 | -0.98111 | 0.360292 | -2.7231 | 0.006467 | 0.024121 | 0.50659 |
| SERPINA1( | 29.76813 | -0.96558 | 0.354646 | -2.72266 | 0.006476 | 0.024148 | 0.512072 |
| CABP1 | 106.8934 | -0.53339 | 0.195922 | -2.72246 | 0.00648 | 0.024157 | 0.690929 |
| COL9A1 | 425.3424 | -0.47608 | 0.174916 | -2.72177 | 0.006493 | 0.024191 | 0.718928 |
| NAPSA | 13.42775 | -1.33171 | 0.489289 | -2.72172 | 0.006494 | 0.024191 | 0.397298 |
| AC140479 | 7.256322 | -2.37603 | 0.873094 | -2.72139 | 0.006501 | 0.024205 | 0.192638 |
| CTRL | 31.42382 | -0.92073 | 0.338336 | -2.72133 | 0.006502 | 0.024205 | 0.528243 |
| DGKQ | 531.1484 | -0.47368 | 0.174063 | -2.72131 | 0.006502 | 0.024205 | 0.720126 |
| OR7E38P | 55.32382 | -0.65161 | 0.239496 | -2.72074 | 0.006514 | 0.024226 | 0.636571 |
| AHSA2P | 898.2486 | -0.52423 | 0.192867 | -2.71806 | 0.006566 | 0.024396 | 0.695333 |

| | | | | | | | |
|---|---|---|---|---|---|---|---|
| SLC26A10 | 19.1824 | -2.33743 | 0.860482 | -2.71643 | 0.006599 | 0.024501 | 0.197862 |
| HIST1H2BJ | 38.93399 | -1.70534 | 0.628009 | -2.71547 | 0.006618 | 0.024547 | 0.306649 |
| CCL27 | 10.79398 | -1.78946 | 0.659118 | -2.71492 | 0.006629 | 0.024576 | 0.289281 |
| TMEM94 | 1271.117 | -0.42089 | 0.155027 | -2.71495 | 0.006629 | 0.024576 | 0.746963 |
| KIR3DX1 | 11.01505 | -1.7783 | 0.655307 | -2.71369 | 0.006654 | 0.024646 | 0.291526 |
| PTK7 | 6814.799 | -0.41534 | 0.153136 | -2.71223 | 0.006683 | 0.024734 | 0.749843 |
| IMPDH1P8 | 11.97789 | -1.78286 | 0.657884 | -2.70999 | 0.006728 | 0.024864 | 0.290606 |
| TMEM79 | 69.64469 | -0.75807 | 0.280008 | -2.70731 | 0.006783 | 0.025017 | 0.591287 |
| PSMB9 | 10.81947 | -1.66388 | 0.614673 | -2.70694 | 0.006791 | 0.025035 | 0.315589 |
| PRICKLE1 | 1022.695 | -0.56274 | 0.208109 | -2.70405 | 0.00685 | 0.0252 | 0.677017 |
| ABHD15 | 95.21646 | -0.58786 | 0.217413 | -2.70389 | 0.006853 | 0.025206 | 0.665329 |
| PROCR | 659.8399 | -0.49199 | 0.182194 | -2.70039 | 0.006926 | 0.025399 | 0.711042 |
| SLC27A6 | 255.1169 | -0.43169 | 0.159862 | -2.70037 | 0.006926 | 0.025399 | 0.741395 |
| PDIA5 | 755.0262 | -0.44519 | 0.164997 | -2.69816 | 0.006972 | 0.02553 | 0.734489 |
| KLK6 | 93.31442 | -0.55689 | 0.206519 | -2.69657 | 0.007006 | 0.025641 | 0.679765 |
| B3GALT5 | 1754.058 | -0.41817 | 0.155085 | -2.69639 | 0.007009 | 0.025649 | 0.748373 |
| STEAP4 | 19.98257 | -1.13403 | 0.420602 | -2.69622 | 0.007013 | 0.025657 | 0.45564 |
| AL049629. | 13.17689 | -3.65555 | 1.357209 | -2.69343 | 0.007072 | 0.025846 | 0.079354 |
| C21orf62 | 65.80534 | -0.74901 | 0.278368 | -2.69071 | 0.00713 | 0.026041 | 0.595013 |
| CTU1 | 150.2727 | -0.43198 | 0.160614 | -2.68955 | 0.007155 | 0.026111 | 0.741244 |
| RDH13 | 169.8944 | -0.55639 | 0.206881 | -2.68945 | 0.007157 | 0.026111 | 0.679999 |
| SRPK3 | 28.91801 | -1.75885 | 0.653954 | -2.68956 | 0.007155 | 0.026111 | 0.295484 |
| IL11RA | 98.92978 | -0.80117 | 0.298022 | -2.68829 | 0.007182 | 0.026191 | 0.573883 |
| AC139491 | 18.86899 | -1.70036 | 0.63257 | -2.68802 | 0.007188 | 0.026207 | 0.307709 |
| OR6D1P | 106.8747 | -0.61984 | 0.230698 | -2.68682 | 0.007214 | 0.026296 | 0.650741 |
| LY75 | 51.28961 | -1.13979 | 0.424334 | -2.68606 | 0.00723 | 0.026333 | 0.453827 |
| SH3RF3 | 54.55249 | -0.91668 | 0.341451 | -2.68466 | 0.00726 | 0.026421 | 0.529727 |
| PIF1 | 1211.367 | -0.52689 | 0.196352 | -2.68341 | 0.007287 | 0.026498 | 0.694047 |
| DNAH1 | 60.28911 | -0.69315 | 0.258351 | -2.68297 | 0.007297 | 0.026517 | 0.618503 |
| WDR13 | 616.9307 | -0.49704 | 0.185409 | -2.68079 | 0.007345 | 0.026667 | 0.708557 |
| ZC3H12D | 21.91218 | -1.04863 | 0.391313 | -2.67977 | 0.007367 | 0.026731 | 0.483427 |
| RASGRF2 | 323.3669 | -0.54296 | 0.202663 | -2.67911 | 0.007382 | 0.026778 | 0.686363 |
| LHFPL5 | 119.4642 | -0.66554 | 0.248549 | -2.67772 | 0.007413 | 0.026862 | 0.630451 |
| CYBC1 | 1121.077 | -0.47537 | 0.177545 | -2.67747 | 0.007418 | 0.026871 | 0.719281 |
| PROM2 | 153.676 | -0.82825 | 0.309384 | -2.6771 | 0.007426 | 0.026895 | 0.563211 |
| MCRIP2 | 475.325 | -0.52171 | 0.19492 | -2.67656 | 0.007438 | 0.026912 | 0.696544 |
| SPSB3 | 587.4414 | -0.44746 | 0.167228 | -2.67575 | 0.007456 | 0.026954 | 0.733332 |
| REV3L | 398.8688 | -0.4668 | 0.174488 | -2.67526 | 0.007467 | 0.026986 | 0.723568 |
| SLC10A4 | 54.83041 | -0.7437 | 0.278011 | -2.67509 | 0.007471 | 0.026988 | 0.597204 |
| KLK10 | 132.2275 | -0.46055 | 0.172234 | -2.67396 | 0.007496 | 0.027057 | 0.72671 |
| B4GALNT4 | 2426.485 | -1.08201 | 0.404807 | -2.6729 | 0.00752 | 0.027125 | 0.47237 |
| LRFN2 | 34.87588 | -1.02284 | 0.382712 | -2.67261 | 0.007526 | 0.02713 | 0.492146 |
| PMP22 | 119.9901 | -0.49269 | 0.184352 | -2.67256 | 0.007528 | 0.02713 | 0.710699 |
| FAM78B | 70.67728 | -0.65435 | 0.244878 | -2.67215 | 0.007537 | 0.027157 | 0.635361 |

| | | | | | | | |
|---|---|---|---|---|---|---|---|
| SNX14 | 705.942 | -0.42659 | 0.159645 | -2.67209 | 0.007538 | 0.027157 | 0.74402 |
| SLC51A | 123.8924 | -0.72962 | 0.273171 | -2.67092 | 0.007564 | 0.027229 | 0.603063 |
| PHF1 | 43.68737 | -1.29229 | 0.484055 | -2.66971 | 0.007592 | 0.027287 | 0.408304 |
| HTR1A | 123.4872 | -0.79973 | 0.299639 | -2.66898 | 0.007608 | 0.027341 | 0.574456 |
| MC1R | 104.6814 | -0.68525 | 0.256791 | -2.66851 | 0.007619 | 0.027362 | 0.621898 |
| GCNA | 37.97985 | -0.88132 | 0.330486 | -2.66674 | 0.007659 | 0.027466 | 0.54287 |
| PDCD6IPP | 3.441224 | -4.59445 | 1.723087 | -2.6664 | 0.007667 | 0.027483 | 0.041394 |
| TMED9 | 3633.198 | -0.51912 | 0.194827 | -2.66454 | 0.007709 | 0.027618 | 0.697796 |
| CHRNA10 | 12.33839 | -1.72207 | 0.646531 | -2.66356 | 0.007732 | 0.027667 | 0.303112 |
| MDK | 6908.992 | -0.43031 | 0.161555 | -2.66352 | 0.007733 | 0.027667 | 0.742105 |
| TNFAIP2 | 664.8786 | -0.53208 | 0.199791 | -2.66317 | 0.007741 | 0.027685 | 0.691559 |
| SPG7 | 2618.101 | -0.49806 | 0.187102 | -2.66195 | 0.007769 | 0.027779 | 0.708059 |
| OR52A1 | 191.9129 | -0.47916 | 0.18004 | -2.6614 | 0.007782 | 0.027819 | 0.717396 |
| ATP8B4 | 17.90768 | -1.45209 | 0.54658 | -2.65669 | 0.007891 | 0.02815 | 0.365491 |
| TSGA13 | 18.5761 | -1.22156 | 0.459822 | -2.65658 | 0.007894 | 0.02815 | 0.42882 |
| AC019257 | 54.46421 | -2.76138 | 1.0399 | -2.65543 | 0.007921 | 0.028217 | 0.147483 |
| AGAP12P | 27.14326 | -1.26328 | 0.475907 | -2.65446 | 0.007944 | 0.028268 | 0.416597 |
| CHRNA7 | 264.7266 | -0.46382 | 0.174836 | -2.65287 | 0.007981 | 0.028367 | 0.725065 |
| HEXIM2 | 198.4408 | -0.47009 | 0.177212 | -2.65269 | 0.007985 | 0.028377 | 0.721921 |
| AVPR1B | 19.23337 | -1.05732 | 0.398687 | -2.652 | 0.008002 | 0.028429 | 0.480525 |
| CYSLTR1 | 144.9649 | -0.5616 | 0.211808 | -2.65144 | 0.008015 | 0.02847 | 0.677552 |
| MYH7B | 37.70269 | -0.82393 | 0.310793 | -2.65105 | 0.008024 | 0.02848 | 0.564902 |
| PIGG | 802.7356 | -0.44992 | 0.169754 | -2.65045 | 0.008038 | 0.028501 | 0.732082 |
| RPS20P22 | 78.29231 | -0.88423 | 0.333812 | -2.64888 | 0.008076 | 0.028622 | 0.541778 |
| NT5M | 235.1675 | -0.44742 | 0.169036 | -2.64692 | 0.008123 | 0.028741 | 0.733351 |
| METTL17 | 1517.238 | -0.42372 | 0.160127 | -2.64616 | 0.008141 | 0.028782 | 0.745498 |
| AC068134 | 20.19887 | -1.16248 | 0.439854 | -2.64287 | 0.008221 | 0.029027 | 0.446745 |
| RPS3AP54 | 3.56428 | -3.76757 | 1.425902 | -2.64223 | 0.008236 | 0.029076 | 0.073426 |
| TMEM173 | 30.29595 | -1.16698 | 0.441705 | -2.64198 | 0.008242 | 0.029086 | 0.445354 |
| KIFC3 | 168.1564 | -0.72483 | 0.274438 | -2.64114 | 0.008263 | 0.029134 | 0.605069 |
| TRIM17 | 30.20264 | -0.93114 | 0.352816 | -2.63916 | 0.008311 | 0.029263 | 0.524445 |
| SLC10A3 | 209.1612 | -0.51634 | 0.195662 | -2.63894 | 0.008317 | 0.029276 | 0.699143 |
| CR769776 | 7.091004 | -2.65969 | 1.008752 | -2.63662 | 0.008374 | 0.029465 | 0.158253 |
| AL603832. | 8.523876 | -3.20426 | 1.215631 | -2.63588 | 0.008392 | 0.029524 | 0.108498 |
| RNFT1 | 125.3238 | -0.52821 | 0.200481 | -2.63473 | 0.00842 | 0.029599 | 0.693414 |
| GJA3 | 43.06651 | -0.90913 | 0.345076 | -2.63457 | 0.008424 | 0.029607 | 0.532507 |
| AC021087 | 14.36176 | -2.22609 | 0.845233 | -2.6337 | 0.008446 | 0.029664 | 0.213737 |
| TMEM191A | 94.49365 | -0.55269 | 0.209844 | -2.63384 | 0.008443 | 0.029664 | 0.681746 |
| GJC2 | 23.9516 | -0.98134 | 0.373096 | -2.63027 | 0.008532 | 0.029924 | 0.506508 |
| TMIE | 25.96121 | -0.9607 | 0.365546 | -2.62813 | 0.008586 | 0.030094 | 0.513806 |
| TSPAN15 | 288.5646 | -0.4632 | 0.176362 | -2.62644 | 0.008628 | 0.030238 | 0.725374 |
| CROCCP2 | 929.433 | -0.47293 | 0.180136 | -2.6254 | 0.008655 | 0.030318 | 0.7205 |
| MT-CO3 | 157907.9 | -0.78103 | 0.297852 | -2.6222 | 0.008736 | 0.030561 | 0.581952 |
| CDH15 | 11.56386 | -1.36714 | 0.521448 | -2.62181 | 0.008746 | 0.03059 | 0.38766 |

| | | | | | | | |
|---|---|---|---|---|---|---|---|
| STS | 116.475 | -0.48479 | 0.185039 | -2.61995 | 0.008794 | 0.030731 | 0.7146 |
| CAPN11 | 93.80527 | -0.597 | 0.228049 | -2.61783 | 0.008849 | 0.030879 | 0.66113 |
| ERVW-1 | 150.2149 | -0.49332 | 0.188664 | -2.6148 | 0.008928 | 0.031117 | 0.710389 |
| SLC39A13 | 755.3851 | -0.57453 | 0.219801 | -2.61384 | 0.008953 | 0.031192 | 0.671507 |
| AL671277. | 3.10472 | -3.63841 | 1.392645 | -2.61259 | 0.008986 | 0.031268 | 0.080303 |
| ATG16L2 | 564.1195 | -0.66699 | 0.255647 | -2.60903 | 0.00908 | 0.031531 | 0.629819 |
| PTH1R | 42.04222 | -1.17903 | 0.451984 | -2.60857 | 0.009092 | 0.031561 | 0.441647 |
| SLC52A1 | 30.4549 | -0.94334 | 0.361893 | -2.60667 | 0.009143 | 0.031718 | 0.520028 |
| ARHGEF33 | 12.49835 | -3.35002 | 1.285208 | -2.60659 | 0.009145 | 0.031719 | 0.098072 |
| HERC2P3 | 127.5095 | -0.46212 | 0.177698 | -2.60058 | 0.009307 | 0.032222 | 0.725921 |
| RNF32 | 45.76992 | -0.81639 | 0.314257 | -2.59783 | 0.009381 | 0.032442 | 0.567862 |
| EPHB6 | 214.1341 | -0.75504 | 0.290705 | -2.59726 | 0.009397 | 0.032489 | 0.59253 |
| ACTG1P3 | 4.315784 | -2.8799 | 1.110209 | -2.59401 | 0.009486 | 0.032746 | 0.135851 |
| SOCS1 | 224.3445 | -0.44051 | 0.16982 | -2.59401 | 0.009486 | 0.032746 | 0.736872 |
| CLN3 | 573.885 | -0.51073 | 0.196983 | -2.59278 | 0.00952 | 0.032843 | 0.701865 |
| PRR16 | 73.28748 | -0.84017 | 0.324074 | -2.59253 | 0.009527 | 0.03286 | 0.558577 |
| NRK | 100.5492 | -0.50794 | 0.195935 | -2.59237 | 0.009532 | 0.032867 | 0.703228 |
| HHIP | 52.13038 | -1.53902 | 0.59386 | -2.59155 | 0.009554 | 0.032901 | 0.344119 |
| PSD4 | 77.50564 | -0.82264 | 0.317451 | -2.59139 | 0.009559 | 0.032907 | 0.565406 |
| STRC | 17.81276 | -1.34081 | 0.517416 | -2.59135 | 0.00956 | 0.032907 | 0.3948 |
| SYT3 | 470.5042 | -0.46078 | 0.177856 | -2.59077 | 0.009576 | 0.032956 | 0.726591 |
| ALS2CL | 222.6137 | -0.67403 | 0.260194 | -2.59049 | 0.009584 | 0.032977 | 0.626754 |
| AC010323 | 4.247768 | -5.64542 | 2.179793 | -2.58989 | 0.009601 | 0.033021 | 0.019978 |
| MOK | 83.65934 | -0.85526 | 0.330409 | -2.58848 | 0.00964 | 0.03315 | 0.552767 |
| JAM2 | 220.4957 | -0.46165 | 0.178365 | -2.58823 | 0.009647 | 0.033162 | 0.726156 |
| GNA15 | 16.03019 | -1.42506 | 0.550724 | -2.58761 | 0.009664 | 0.033194 | 0.372404 |
| SOCS2 | 1198.765 | -0.52013 | 0.201124 | -2.58612 | 0.009706 | 0.033317 | 0.697308 |
| RECQL5 | 870.3743 | -0.61595 | 0.2383 | -2.58477 | 0.009744 | 0.033422 | 0.6525 |
| SLC6A8 | 4961.796 | -0.4243 | 0.164316 | -2.5822 | 0.009817 | 0.033652 | 0.745202 |
| ABCA9 | 13.00486 | -1.46139 | 0.566051 | -2.58173 | 0.009831 | 0.033684 | 0.363142 |
| C10orf143 | 40.50273 | -0.72861 | 0.282348 | -2.58054 | 0.009865 | 0.033787 | 0.603485 |
| SHISAL2B | 71.08957 | -0.53677 | 0.208144 | -2.57883 | 0.009914 | 0.033921 | 0.689314 |
| PIWIL2 | 13.32806 | -1.70195 | 0.660012 | -2.57867 | 0.009918 | 0.033924 | 0.30737 |
| PCK1 | 7.2996 | -3.98974 | 1.547539 | -2.57812 | 0.009934 | 0.033964 | 0.062946 |
| SLC2A6 | 384.1177 | -0.57504 | 0.223109 | -2.57738 | 0.009955 | 0.034023 | 0.671269 |
| CD63 | 6973.303 | -0.42041 | 0.163314 | -2.57421 | 0.010047 | 0.034282 | 0.747214 |
| TMCO6 | 455.509 | -0.42875 | 0.166635 | -2.57299 | 0.010082 | 0.034389 | 0.742905 |
| FKBP6 | 13.11712 | -2.91007 | 1.131048 | -2.5729 | 0.010085 | 0.034392 | 0.13304 |
| CSAD | 129.3347 | -0.8346 | 0.324523 | -2.57178 | 0.010118 | 0.03449 | 0.560738 |
| TAS2R6P | 11.17324 | -1.44936 | 0.56404 | -2.5696 | 0.010182 | 0.034666 | 0.366184 |
| AGMO | 34.35116 | -1.34298 | 0.522673 | -2.56945 | 0.010186 | 0.034675 | 0.394206 |
| TAT | 10.50126 | -1.52611 | 0.594069 | -2.56891 | 0.010202 | 0.034715 | 0.347213 |
| PRRT2 | 545.8717 | -0.6297 | 0.245136 | -2.56878 | 0.010206 | 0.034721 | 0.646311 |
| INPP5E | 653.0751 | -0.47952 | 0.186751 | -2.56769 | 0.010238 | 0.03481 | 0.717217 |

| | | | | | | | |
|---|---|---|---|---|---|---|---|
| FES | 126.5705 | -0.7289 | 0.284012 | -2.56645 | 0.010275 | 0.034907 | 0.603363 |
| CICP27 | 106.933 | -0.67162 | 0.261928 | -2.56414 | 0.010343 | 0.035106 | 0.627802 |
| RAD51AP2 | 8.340411 | -1.81809 | 0.709334 | -2.5631 | 0.010374 | 0.03519 | 0.283596 |
| GRM1 | 34.66728 | -1.02994 | 0.402013 | -2.56195 | 0.010409 | 0.035286 | 0.489732 |
| SLC3A1 | 4.6077 | -2.54537 | 0.993681 | -2.56155 | 0.01042 | 0.035313 | 0.171304 |
| SNAPC4 | 800.9849 | -0.48803 | 0.190537 | -2.56134 | 0.010427 | 0.03532 | 0.712998 |
| HLA-H | 6.31823 | -1.88398 | 0.735897 | -2.56011 | 0.010464 | 0.035417 | 0.270936 |
| TCF24 | 17.05011 | -1.06272 | 0.415364 | -2.55854 | 0.010511 | 0.035564 | 0.478727 |
| KRT17P2 | 3.143578 | -3.17834 | 1.242429 | -2.55817 | 0.010523 | 0.035595 | 0.110465 |
| C21orf58 | 418.9238 | -0.48053 | 0.187957 | -2.55659 | 0.01057 | 0.035736 | 0.716714 |
| CRX | 61.84296 | -1.25107 | 0.489777 | -2.55437 | 0.010638 | 0.035922 | 0.420136 |
| EPSTI1 | 21.21041 | -1.34793 | 0.527693 | -2.55439 | 0.010637 | 0.035922 | 0.392854 |
| TSC22D3 | 2162.494 | -0.50191 | 0.196549 | -2.55363 | 0.010661 | 0.035992 | 0.70617 |
| ECM1 | 37.7348 | -1.04533 | 0.4096 | -2.55207 | 0.010708 | 0.036096 | 0.484534 |
| SCGB2B2 | 14.0068 | -2.52082 | 0.988277 | -2.55072 | 0.01075 | 0.036201 | 0.174244 |
| AC011511 | 38.3838 | -1.04798 | 0.411022 | -2.5497 | 0.010782 | 0.036298 | 0.483644 |
| MYT1L | 39.18526 | -0.92846 | 0.364178 | -2.54946 | 0.010789 | 0.036304 | 0.52542 |
| ITGB4 | 158.5706 | -1.18965 | 0.466955 | -2.54768 | 0.010844 | 0.036456 | 0.438409 |
| BAIAP3 | 173.4916 | -0.95404 | 0.374588 | -2.54691 | 0.010868 | 0.036493 | 0.516184 |
| FAM129A | 1161.77 | -0.60678 | 0.238244 | -2.5469 | 0.010868 | 0.036493 | 0.656659 |
| MYH3 | 78.74431 | -1.06538 | 0.418606 | -2.54506 | 0.010926 | 0.036631 | 0.477848 |
| RSKR | 364.5239 | -0.50183 | 0.197179 | -2.54504 | 0.010927 | 0.036631 | 0.706212 |
| FOXL2 | 160.2553 | -0.61381 | 0.241195 | -2.54488 | 0.010931 | 0.03664 | 0.653468 |
| ECEL1 | 485.0115 | -0.46324 | 0.182032 | -2.5448 | 0.010934 | 0.036641 | 0.725357 |
| EPS8L1 | 544.3383 | -0.53094 | 0.20865 | -2.54465 | 0.010939 | 0.036643 | 0.692103 |
| ZBTB47 | 190.8493 | -0.45348 | 0.178273 | -2.54374 | 0.010967 | 0.036725 | 0.730279 |
| AMBP | 37.26785 | -0.82867 | 0.325893 | -2.54278 | 0.010998 | 0.036804 | 0.563047 |
| AC006978 | 114.2323 | -0.41532 | 0.163405 | -2.54167 | 0.011032 | 0.036906 | 0.749852 |
| CCDC125 | 30.7817 | -1.11863 | 0.440479 | -2.53957 | 0.011099 | 0.037086 | 0.460532 |
| LCTL | 210.154 | -0.44494 | 0.1754 | -2.53674 | 0.011189 | 0.0373 | 0.734613 |
| CACNA1C | 59.98756 | -0.87116 | 0.343559 | -2.53569 | 0.011223 | 0.037398 | 0.546708 |
| TMPRSS3 | 18.7262 | -1.63129 | 0.643595 | -2.53466 | 0.011256 | 0.037479 | 0.322799 |
| HAPLN4 | 499.4413 | -0.42754 | 0.168823 | -2.5325 | 0.011325 | 0.037672 | 0.743527 |
| PIP5KL1 | 313.5979 | -0.59425 | 0.234658 | -2.53243 | 0.011328 | 0.037672 | 0.662387 |
| CLEC4A | 7.015359 | -2.0564 | 0.81398 | -2.52635 | 0.011526 | 0.03826 | 0.240416 |
| ZNF440 | 4.802959 | -2.19918 | 0.870742 | -2.52564 | 0.011549 | 0.038315 | 0.217761 |
| ZFYVE27 | 506.2937 | -0.51218 | 0.202805 | -2.52546 | 0.011555 | 0.038319 | 0.701163 |
| KCNMB3 | 50.45498 | -0.69596 | 0.275672 | -2.52459 | 0.011583 | 0.038385 | 0.617299 |
| ZNF236 | 603.2284 | -0.65508 | 0.259525 | -2.52414 | 0.011598 | 0.038419 | 0.635041 |
| TMEM163 | 196.7539 | -0.65297 | 0.258815 | -2.52293 | 0.011638 | 0.03853 | 0.635969 |
| ABHD14A- | 29.12077 | -1.49802 | 0.594272 | -2.52076 | 0.01171 | 0.038753 | 0.354039 |
| TNXB | 120.022 | -0.67462 | 0.267902 | -2.51816 | 0.011797 | 0.038973 | 0.626497 |
| LFNG | 1045.872 | -0.49909 | 0.198243 | -2.51757 | 0.011817 | 0.039031 | 0.707553 |
| WDR97 | 37.42702 | -1.24222 | 0.493563 | -2.51685 | 0.011841 | 0.039065 | 0.42272 |

| | | | | | | | |
|---|---|---|---|---|---|---|---|
| MORN3 | 40.1993 | -0.80903 | 0.321454 | -2.51678 | 0.011843 | 0.039066 | 0.570766 |
| CGB7 | 43.3921 | -1.39007 | 0.552399 | -2.51642 | 0.011855 | 0.039083 | 0.381547 |
| SP100 | 20.0676 | -1.20709 | 0.47974 | -2.51613 | 0.011865 | 0.0391 | 0.433142 |
| SLC16A8 | 35.47897 | -1.33412 | 0.530329 | -2.51565 | 0.011881 | 0.039146 | 0.396633 |
| AMYH0202 | 3.687969 | -5.16268 | 2.05239 | -2.51545 | 0.011888 | 0.039154 | 0.027918 |
| ARSA | 356.1155 | -0.58457 | 0.23239 | -2.51547 | 0.011887 | 0.039154 | 0.666848 |
| FGFR3 | 2628.44 | -0.60414 | 0.24037 | -2.51336 | 0.011959 | 0.039349 | 0.657865 |
| IGFL4 | 10.71564 | -1.8534 | 0.737484 | -2.51314 | 0.011966 | 0.039362 | 0.276739 |
| LPIN3 | 210.7583 | -0.78484 | 0.312322 | -2.5129 | 0.011974 | 0.039377 | 0.580418 |
| LPL | 331.3615 | -0.51697 | 0.205747 | -2.51265 | 0.011983 | 0.039397 | 0.698837 |
| DUSP8 | 43.69195 | -0.84522 | 0.336478 | -2.51197 | 0.012006 | 0.039444 | 0.556625 |
| ZSWIM3 | 101.3639 | -0.4298 | 0.171309 | -2.50891 | 0.01211 | 0.039756 | 0.742365 |
| ZNF705D | 4.737339 | -2.31184 | 0.921679 | -2.5083 | 0.012131 | 0.039811 | 0.201403 |
| ABHD14A | 90.91051 | -0.7411 | 0.295505 | -2.50789 | 0.012145 | 0.039833 | 0.598285 |
| CELF6 | 6.476124 | -3.81103 | 1.520559 | -2.50634 | 0.012199 | 0.039979 | 0.071247 |
| HSF4 | 21.87266 | -1.929 | 0.769739 | -2.50605 | 0.012209 | 0.039997 | 0.262611 |
| TGFA | 219.3194 | -0.54107 | 0.215946 | -2.50557 | 0.012225 | 0.040043 | 0.687262 |
| CNTNAP3E | 387.6906 | -0.50179 | 0.200423 | -2.50363 | 0.012293 | 0.040224 | 0.706232 |
| FAM129C | 100.9249 | -0.68476 | 0.273528 | -2.50343 | 0.0123 | 0.04024 | 0.62211 |
| KCNJ15 | 53.15165 | -0.66259 | 0.264802 | -2.50222 | 0.012342 | 0.040355 | 0.631742 |
| MROH6 | 78.5324 | -1.52428 | 0.609586 | -2.50052 | 0.012401 | 0.040495 | 0.347653 |
| CD8B | 33.41028 | -1.82094 | 0.728307 | -2.50023 | 0.012411 | 0.04052 | 0.283037 |
| AMDHD1 | 79.42496 | -0.72694 | 0.290871 | -2.49918 | 0.012448 | 0.040616 | 0.604184 |
| EPHA10 | 41.44035 | -0.88037 | 0.35227 | -2.49912 | 0.01245 | 0.040616 | 0.543229 |
| CYP4F35P | 6.634476 | -3.12229 | 1.249505 | -2.49882 | 0.012461 | 0.040643 | 0.114841 |
| C19orf44 | 145.7261 | -0.4704 | 0.188468 | -2.49589 | 0.012564 | 0.040935 | 0.721766 |
| LEFTY2 | 54.32979 | -0.68897 | 0.276087 | -2.49547 | 0.012579 | 0.040968 | 0.620298 |
| AC092143 | 3.875221 | -5.50719 | 2.207244 | -2.49505 | 0.012594 | 0.041 | 0.021987 |
| PPP2R3B | 559.8085 | -0.57135 | 0.229024 | -2.49473 | 0.012605 | 0.041022 | 0.672986 |
| TECR | 2588.116 | -0.4329 | 0.173564 | -2.49417 | 0.012625 | 0.041072 | 0.740773 |
| MKX | 97.14956 | -0.72497 | 0.290685 | -2.494 | 0.012631 | 0.041083 | 0.60501 |
| MT-ND4L | 17967.88 | -0.4335 | 0.173821 | -2.49392 | 0.012634 | 0.041085 | 0.740465 |
| MT-ND4 | 202940.3 | -0.83121 | 0.333426 | -2.49293 | 0.012669 | 0.041176 | 0.562058 |
| LYPD6B | 68.81785 | -0.66461 | 0.266645 | -2.49249 | 0.012685 | 0.041188 | 0.630859 |
| SAMD11 | 33.72388 | -1.21447 | 0.487284 | -2.49231 | 0.012691 | 0.041201 | 0.430933 |
| KLK13 | 44.54042 | -0.70518 | 0.28333 | -2.48891 | 0.012814 | 0.041559 | 0.613364 |
| ABO | 24.96348 | -1.18994 | 0.478702 | -2.48577 | 0.012927 | 0.041888 | 0.43832 |
| CDHR4 | 16.57184 | -1.26266 | 0.508216 | -2.4845 | 0.012973 | 0.042013 | 0.416774 |
| TMEM88B | 24.34473 | -1.16504 | 0.46932 | -2.4824 | 0.01305 | 0.042206 | 0.445952 |
| HIC1 | 13.896 | -1.35852 | 0.547494 | -2.48133 | 0.013089 | 0.042309 | 0.389983 |
| ACAN | 97.60763 | -0.96508 | 0.389046 | -2.48062 | 0.013115 | 0.042385 | 0.512251 |
| SHANK1 | 94.00525 | -0.79336 | 0.319848 | -2.48043 | 0.013122 | 0.042401 | 0.576998 |
| SMG1P7 | 149.9171 | -0.58976 | 0.237815 | -2.47993 | 0.013141 | 0.042436 | 0.664452 |
| CEMIP | 28.3905 | -1.71203 | 0.69064 | -2.4789 | 0.013179 | 0.042543 | 0.305231 |

| | | | | | | | |
|---|---|---|---|---|---|---|---|
| LMNTD2 | 40.11961 | -0.99405 | 0.401004 | -2.4789 | 0.013179 | 0.042543 | 0.502066 |
| TMEM145 | 106.2947 | -0.76119 | 0.307361 | -2.47653 | 0.013267 | 0.042763 | 0.590009 |
| ANKLE1 | 65.68384 | -0.93246 | 0.376545 | -2.47637 | 0.013273 | 0.042774 | 0.523963 |
| HCN1 | 124.8559 | -0.56626 | 0.228711 | -2.47587 | 0.013291 | 0.042817 | 0.675365 |
| AC068580 | 123.0731 | -0.44237 | 0.178734 | -2.47504 | 0.013322 | 0.042885 | 0.735922 |
| SMCO2 | 6.908945 | -1.71603 | 0.693399 | -2.47481 | 0.013331 | 0.042897 | 0.304386 |
| CTF1 | 27.56362 | -0.81967 | 0.331752 | -2.47073 | 0.013484 | 0.043276 | 0.566572 |
| ENPP7P1 | 12.23149 | -1.62358 | 0.657476 | -2.46942 | 0.013533 | 0.043411 | 0.324529 |
| AL390726. | 4.488754 | -2.3626 | 0.957189 | -2.46827 | 0.013577 | 0.043519 | 0.19444 |
| ALOX12P2 | 226.0197 | -0.41904 | 0.169786 | -2.46802 | 0.013586 | 0.043524 | 0.747924 |
| LINC0067: | 86.84038 | -0.51458 | 0.208632 | -2.46645 | 0.013646 | 0.043708 | 0.699997 |
| ALDH1L2 | 697.8061 | -0.47969 | 0.19451 | -2.46615 | 0.013657 | 0.043736 | 0.717131 |
| AC132812 | 21.18069 | -1.0173 | 0.413272 | -2.46156 | 0.013833 | 0.044233 | 0.494041 |
| TNNI3 | 123.7922 | -0.52463 | 0.213333 | -2.4592 | 0.013925 | 0.044484 | 0.695138 |
| NMUR1 | 22.60803 | -0.89145 | 0.362522 | -2.45903 | 0.013931 | 0.044497 | 0.539071 |
| SMIM31 | 3.67152 | -3.60667 | 1.467753 | -2.45727 | 0.014 | 0.044666 | 0.082089 |
| SZT2 | 762.3951 | -0.4675 | 0.190518 | -2.45384 | 0.014134 | 0.045036 | 0.723217 |
| SLC17A6 | 5.89088 | -2.442 | 0.995257 | -2.45364 | 0.014142 | 0.045053 | 0.184028 |
| SSUH2 | 6.189651 | -2.35565 | 0.960113 | -2.45351 | 0.014147 | 0.045061 | 0.19538 |
| POM121L9 | 13.88865 | -1.33788 | 0.545344 | -2.45327 | 0.014156 | 0.045074 | 0.395602 |
| REELD1 | 5.596539 | -2.53117 | 1.03191 | -2.45289 | 0.014171 | 0.045113 | 0.172999 |
| REPS2 | 169.5378 | -0.86265 | 0.351944 | -2.45109 | 0.014242 | 0.045255 | 0.549942 |
| C1QTNF1 | 35.58631 | -0.89736 | 0.366592 | -2.44783 | 0.014372 | 0.045583 | 0.536869 |
| EIF4E1B | 37.99105 | -0.96911 | 0.396347 | -2.44509 | 0.014482 | 0.04586 | 0.510823 |
| TNS2 | 222.6187 | -0.59321 | 0.242615 | -2.44505 | 0.014483 | 0.04586 | 0.662868 |
| CDC14C | 6.578941 | -2.07958 | 0.850645 | -2.44471 | 0.014497 | 0.045879 | 0.236583 |
| IKZF3 | 57.59565 | -0.67748 | 0.277122 | -2.4447 | 0.014497 | 0.045879 | 0.625256 |
| RABGGTA | 327.4222 | -0.51287 | 0.209785 | -2.44476 | 0.014495 | 0.045879 | 0.700825 |
| ADAMTS17 | 59.67951 | -0.79757 | 0.326592 | -2.44211 | 0.014602 | 0.046142 | 0.575315 |
| TAS2R4 | 21.2362 | -1.00786 | 0.412714 | -2.44202 | 0.014605 | 0.046145 | 0.497284 |
| ZNF418 | 260.8635 | -0.54927 | 0.225145 | -2.43963 | 0.014702 | 0.046386 | 0.683366 |
| MRPL45P2 | 47.69049 | -0.80142 | 0.328619 | -2.43876 | 0.014738 | 0.046462 | 0.573783 |
| PCDHGA1: | 24.07839 | -1.19717 | 0.490896 | -2.43875 | 0.014738 | 0.046462 | 0.436129 |
| QPCTL | 623.7212 | -0.51066 | 0.209572 | -2.4367 | 0.014822 | 0.046685 | 0.701899 |
| RASGRP4 | 80.68575 | -0.52783 | 0.216674 | -2.43603 | 0.014849 | 0.046762 | 0.693599 |
| TMEM121 | 38.69517 | -1.07352 | 0.440748 | -2.43568 | 0.014864 | 0.046791 | 0.475158 |
| AC008736 | 84.49703 | -0.53831 | 0.221204 | -2.43355 | 0.014952 | 0.047032 | 0.688577 |
| AKR1E2 | 42.6098 | -0.78755 | 0.323691 | -2.43304 | 0.014973 | 0.047073 | 0.579326 |
| SLC4A10 | 39.403 | -0.91735 | 0.377054 | -2.43295 | 0.014976 | 0.047076 | 0.529479 |
| MOGAT3 | 9.02475 | -2.18101 | 0.898103 | -2.42846 | 0.015163 | 0.047593 | 0.220521 |
| TUBGCP6 | 1203.979 | -0.49665 | 0.20477 | -2.42539 | 0.015292 | 0.047936 | 0.708752 |
| CATSPER4 | 17.92518 | -1.02657 | 0.42329 | -2.42523 | 0.015299 | 0.047949 | 0.490874 |
| OPRD1 | 129.4001 | -0.57875 | 0.238646 | -2.42514 | 0.015302 | 0.047952 | 0.669544 |
| BGN | 180.0912 | -0.43288 | 0.17858 | -2.42401 | 0.01535 | 0.048046 | 0.740781 |

| | | | | | | | |
|---|---|---|---|---|---|---|---|
| WNT11 | 9.223473 | -2.39377 | 0.988176 | -2.42241 | 0.015418 | 0.048235 | 0.190285 |
| STAC3 | 49.29849 | -1.02069 | 0.42152 | -2.42146 | 0.015458 | 0.048317 | 0.492879 |
| PLAAT1 | 85.63174 | -0.55524 | 0.229309 | -2.42138 | 0.015462 | 0.048319 | 0.680542 |
| ZNF611 | 207.7363 | -0.62921 | 0.260231 | -2.41788 | 0.015611 | 0.048743 | 0.646532 |
| SBDSP1 | 155.3587 | -0.45154 | 0.186768 | -2.41764 | 0.015621 | 0.048766 | 0.731263 |
| THBS3 | 245.5169 | -0.6374 | 0.263772 | -2.41646 | 0.015672 | 0.048915 | 0.642873 |
| NPIPA3 | 118.9731 | -0.87132 | 0.360867 | -2.41452 | 0.015756 | 0.049141 | 0.546645 |
| APCDD1L | 51.83548 | -0.74249 | 0.30764 | -2.41351 | 0.015799 | 0.049255 | 0.597705 |
| CCN6 | 8.287337 | -1.74403 | 0.722853 | -2.41271 | 0.015835 | 0.049351 | 0.298534 |
| ADAMTS10 | 206.7266 | -0.60545 | 0.250974 | -2.4124 | 0.015848 | 0.049378 | 0.657267 |
| CCDC159 | 48.5371 | -1.01917 | 0.422528 | -2.41208 | 0.015862 | 0.049403 | 0.493399 |
| DLL3 | 135.1117 | -0.43101 | 0.178689 | -2.41205 | 0.015863 | 0.049403 | 0.741744 |
| IKBKE | 319.774 | -0.48443 | 0.200908 | -2.41122 | 0.015899 | 0.04949 | 0.714778 |
| MT-ND2 | 74905.59 | -0.777 | 0.322463 | -2.40959 | 0.01597 | 0.049657 | 0.583577 |
| PCDHB3 | 50.50703 | -0.70417 | 0.292257 | -2.40942 | 0.015978 | 0.049671 | 0.613795 |

**Supplementary Table S4. K184E Homozygous vs WT**

**UP - p<.05, FC>1.5**

| GeneID | Base mear | log2(FC) | StdErr | Wald-Stats | P-value | P-adj | FC |
|---|---|---|---|---|---|---|---|
| POLI | 1382.463 | 0.585089 | 0.119843 | 4.88211 | 1.05E-06 | 6.88E-06 | 1.500132 |
| ZBTB49 | 126.039 | 0.585244 | 0.231539 | 2.527624 | 0.011484 | 0.029355 | 1.500292 |
| DCBLD2 | 2745.479 | 0.585427 | 0.11627 | 5.035075 | 4.78E-07 | 3.34E-06 | 1.500483 |
| RC3H2 | 1337.045 | 0.585429 | 0.127157 | 4.603989 | 4.14E-06 | 2.42E-05 | 1.500486 |
| WDR31 | 115.8434 | 0.585439 | 0.200387 | 2.921545 | 0.003483 | 0.01034 | 1.500496 |
| KCNT2 | 453.4513 | 0.585942 | 0.125888 | 4.654464 | 3.25E-06 | 1.93E-05 | 1.501018 |
| TCTEX1D2 | 266.1934 | 0.586086 | 0.161831 | 3.621602 | 0.000293 | 0.00115 | 1.501168 |
| TRIP11 | 905.4751 | 0.586335 | 0.12548 | 4.672744 | 2.97E-06 | 1.78E-05 | 1.501428 |
| MAPRE1 | 5243.621 | 0.586356 | 0.062152 | 9.434222 | 3.94E-21 | 1.70E-19 | 1.501449 |
| PDS5A | 3264.01 | 0.586606 | 0.056594 | 10.3652 | 3.57E-25 | 2.32E-23 | 1.501709 |
| RANBP6 | 803.5543 | 0.588168 | 0.098202 | 5.989348 | 2.11E-09 | 2.21E-08 | 1.503336 |
| ACBD7 | 747.5924 | 0.588436 | 0.112809 | 5.216225 | 1.83E-07 | 1.38E-06 | 1.503616 |
| GNAQ | 1836.096 | 0.588535 | 0.13949 | 4.219186 | 2.45E-05 | 0.000123 | 1.503719 |
| GSTCD | 857.4786 | 0.588728 | 0.119765 | 4.915694 | 8.85E-07 | 5.90E-06 | 1.503921 |
| EFTUD2 | 3756.173 | 0.589131 | 0.095703 | 6.155819 | 7.47E-10 | 8.46E-09 | 1.50434 |
| FAM167A | 222.0653 | 0.58964 | 0.195479 | 3.016378 | 0.002558 | 0.007901 | 1.504871 |
| TMEM231 | 406.0858 | 0.589993 | 0.119451 | 4.939222 | 7.84E-07 | 5.28E-06 | 1.50524 |
| SOX2 | 9122.749 | 0.590078 | 0.084126 | 7.014193 | 2.31E-12 | 3.83E-11 | 1.505328 |
| ZNF670 | 404.9734 | 0.59009 | 0.117304 | 5.030421 | 4.89E-07 | 3.41E-06 | 1.50534 |
| VBP1 | 1772.188 | 0.590313 | 0.091894 | 6.423878 | 1.33E-10 | 1.71E-09 | 1.505574 |
| SHISAL1 | 327.0295 | 0.590346 | 0.120511 | 4.898684 | 9.65E-07 | 6.35E-06 | 1.505607 |
| SAP30L | 500.808 | 0.590687 | 0.247474 | 2.386865 | 0.016993 | 0.041279 | 1.505963 |
| FRS2 | 744.0508 | 0.59095 | 0.139951 | 4.222553 | 2.42E-05 | 0.000122 | 1.506239 |
| SCCPDH | 932.0148 | 0.591012 | 0.088498 | 6.678277 | 2.42E-11 | 3.43E-10 | 1.506303 |
| BMI1 | 302.2315 | 0.591107 | 0.17054 | 3.466086 | 0.000528 | 0.001946 | 1.506402 |
| KIAA0513 | 244.9993 | 0.591677 | 0.217198 | 2.724141 | 0.006447 | 0.017796 | 1.506997 |
| CELF2 | 1296.971 | 0.591958 | 0.09324 | 6.348771 | 2.17E-10 | 2.70E-09 | 1.507291 |
| FAM53B | 518.2395 | 0.592159 | 0.103151 | 5.740692 | 9.43E-09 | 8.93E-08 | 1.507501 |
| NEURL1B | 476.917 | 0.592227 | 0.150548 | 3.9338 | 8.36E-05 | 0.000373 | 1.507572 |
| DOCK7 | 1151.68 | 0.592302 | 0.144171 | 4.10834 | 3.99E-05 | 0.000191 | 1.507651 |
| PRXL2A | 1679.66 | 0.59232 | 0.122931 | 4.818304 | 1.45E-06 | 9.25E-06 | 1.507669 |
| COG6 | 604.4048 | 0.592323 | 0.104242 | 5.682181 | 1.33E-08 | 1.23E-07 | 1.507672 |
| TWF1 | 714.9863 | 0.593308 | 0.195482 | 3.035104 | 0.002405 | 0.007475 | 1.508702 |
| LRIG3 | 405.0033 | 0.593942 | 0.117568 | 5.051921 | 4.37E-07 | 3.08E-06 | 1.509365 |
| ADAM19 | 973.3008 | 0.594144 | 0.102936 | 5.771986 | 7.83E-09 | 7.54E-08 | 1.509577 |
| SHROOM3 | 2011.623 | 0.594158 | 0.098243 | 6.047817 | 1.47E-09 | 1.58E-08 | 1.509591 |
| CMTM3 | 626.7556 | 0.594347 | 0.089907 | 6.610652 | 3.83E-11 | 5.31E-10 | 1.509789 |
| C1orf54 | 215.3932 | 0.59479 | 0.21295 | 2.793095 | 0.005221 | 0.014809 | 1.510252 |
| SNCAIP | 133.3595 | 0.595111 | 0.242918 | 2.449847 | 0.014292 | 0.03546 | 1.510589 |
| MPDZ | 1788.018 | 0.595274 | 0.090473 | 6.579582 | 4.72E-11 | 6.49E-10 | 1.510759 |
| ZNF710 | 911.2598 | 0.595446 | 0.146219 | 4.072281 | 4.66E-05 | 0.00022 | 1.510939 |

| | | | | | | | |
|---|---|---|---|---|---|---|---|
| RPS27L | 807.5789 | 0.595554 | 0.095815 | 6.215685 | 5.11E-10 | 5.95E-09 | 1.511052 |
| BUB1 | 4723.82 | 0.595803 | 0.078787 | 7.562212 | 3.96E-14 | 8.11E-13 | 1.511314 |
| SMCR8 | 1184.598 | 0.596234 | 0.076747 | 7.768829 | 7.92E-15 | 1.76E-13 | 1.511765 |
| SOS2 | 392.8116 | 0.596346 | 0.11923 | 5.001642 | 5.68E-07 | 3.91E-06 | 1.511883 |
| TMEM14A | 170.7086 | 0.596453 | 0.23428 | 2.545898 | 0.0109 | 0.028021 | 1.511995 |
| GTF2IP4 | 3210.67 | 0.596837 | 0.084389 | 7.072425 | 1.52E-12 | 2.58E-11 | 1.512397 |
| KCTD20 | 1282.208 | 0.597251 | 0.151295 | 3.947607 | 7.89E-05 | 0.000354 | 1.512831 |
| CARF | 235.4635 | 0.598598 | 0.225446 | 2.655175 | 0.007927 | 0.021241 | 1.514245 |
| EIF4EBP2 | 7359.998 | 0.598664 | 0.084501 | 7.084685 | 1.39E-12 | 2.38E-11 | 1.514313 |
| UBQLN4 | 1897.839 | 0.598754 | 0.0813 | 7.364737 | 1.77E-13 | 3.35E-12 | 1.514408 |
| AASDH | 423.5015 | 0.598988 | 0.127165 | 4.710307 | 2.47E-06 | 1.51E-05 | 1.514653 |
| AP3M1 | 1435.547 | 0.599025 | 0.092577 | 6.47059 | 9.76E-11 | 1.28E-09 | 1.514693 |
| USP37 | 850.3631 | 0.600608 | 0.167989 | 3.575294 | 0.00035 | 0.001351 | 1.516356 |
| WBP4 | 502.2923 | 0.600657 | 0.132298 | 4.540192 | 5.62E-06 | 3.20E-05 | 1.516407 |
| DMXL2 | 973.4435 | 0.601587 | 0.169488 | 3.549446 | 0.000386 | 0.001474 | 1.517385 |
| DPH3 | 719.158 | 0.601814 | 0.101965 | 5.90215 | 3.59E-09 | 3.62E-08 | 1.517623 |
| NANOS1 | 169.2827 | 0.601868 | 0.170743 | 3.524998 | 0.000423 | 0.001601 | 1.51768 |
| PTPN11 | 5014.166 | 0.602936 | 0.059954 | 10.05665 | 8.59E-24 | 4.75E-22 | 1.518804 |
| TSNAX | 979.2349 | 0.603067 | 0.090951 | 6.630669 | 3.34E-11 | 4.67E-10 | 1.518942 |
| DCAF1 | 1236.218 | 0.603156 | 0.120717 | 4.996447 | 5.84E-07 | 4.01E-06 | 1.519036 |
| CCDC138 | 671.6672 | 0.603648 | 0.115621 | 5.220927 | 1.78E-07 | 1.35E-06 | 1.519554 |
| SCML2 | 918.7233 | 0.604016 | 0.098501 | 6.132095 | 8.67E-10 | 9.69E-09 | 1.519942 |
| ZNF260 | 937.3914 | 0.604517 | 0.128027 | 4.721772 | 2.34E-06 | 1.44E-05 | 1.520469 |
| ARFIP1 | 853.8457 | 0.604625 | 0.102121 | 5.920678 | 3.21E-09 | 3.26E-08 | 1.520583 |
| SPRYD3 | 166.4544 | 0.605314 | 0.187695 | 3.224991 | 0.00126 | 0.004214 | 1.52131 |
| ZBTB43 | 457.4818 | 0.605354 | 0.145844 | 4.15069 | 3.31E-05 | 0.000162 | 1.521352 |
| PDK2 | 421.1251 | 0.605849 | 0.102802 | 5.893379 | 3.78E-09 | 3.80E-08 | 1.521874 |
| DYRK2 | 1076.117 | 0.606011 | 0.1007 | 6.018001 | 1.77E-09 | 1.88E-08 | 1.522045 |
| HECW2 | 230.0984 | 0.606776 | 0.208142 | 2.915202 | 0.003555 | 0.010527 | 1.522852 |
| RBBP5 | 815.6749 | 0.607015 | 0.111495 | 5.444321 | 5.20E-08 | 4.38E-07 | 1.523105 |
| ADD3 | 3137.585 | 0.607057 | 0.106294 | 5.711105 | 1.12E-08 | 1.05E-07 | 1.523148 |
| DYNC1I2 | 2768.82 | 0.607288 | 0.074885 | 8.109625 | 5.08E-16 | 1.31E-14 | 1.523393 |
| PTMS | 6864.833 | 0.607979 | 0.089165 | 6.818592 | 9.19E-12 | 1.40E-10 | 1.524123 |
| ZNF730 | 158.7003 | 0.608242 | 0.204538 | 2.973732 | 0.002942 | 0.008939 | 1.5244 |
| HMGXB4 | 1215.34 | 0.608587 | 0.096135 | 6.33055 | 2.44E-10 | 3.02E-09 | 1.524765 |
| POMK | 237.1913 | 0.60862 | 0.156066 | 3.899749 | 9.63E-05 | 0.000424 | 1.5248 |
| DNA2 | 1345.149 | 0.609213 | 0.087526 | 6.960373 | 3.39E-12 | 5.49E-11 | 1.525427 |
| NHLRC2 | 1053.98 | 0.609329 | 0.152228 | 4.002734 | 6.26E-05 | 0.000287 | 1.525549 |
| C1orf174 | 476.8953 | 0.609395 | 0.119108 | 5.116323 | 3.12E-07 | 2.26E-06 | 1.525619 |
| ARL15 | 316.5388 | 0.609839 | 0.130279 | 4.681021 | 2.85E-06 | 1.72E-05 | 1.526089 |
| MYO1B | 1860.793 | 0.609928 | 0.102856 | 5.929905 | 3.03E-09 | 3.10E-08 | 1.526183 |
| XRN1 | 610.3236 | 0.610018 | 0.115456 | 5.283545 | 1.27E-07 | 9.91E-07 | 1.526278 |
| BAZ2B | 1003.745 | 0.610286 | 0.113425 | 5.380514 | 7.43E-08 | 6.07E-07 | 1.526562 |
| HASPIN | 544.228 | 0.610389 | 0.102772 | 5.939253 | 2.86E-09 | 2.94E-08 | 1.526671 |

| | | | | | | | |
|---|---|---|---|---|---|---|---|
| ARID2 | 1858.968 | 0.610732 | 0.09019 | 6.771649 | 1.27E-11 | 1.89E-10 | 1.527034 |
| TPGS2 | 3142.723 | 0.610771 | 0.074578 | 8.189741 | 2.62E-16 | 6.97E-15 | 1.527075 |
| ARL6IP1 | 7522.697 | 0.61088 | 0.152783 | 3.998347 | 6.38E-05 | 0.000292 | 1.52719 |
| FAM89B | 304.3875 | 0.611075 | 0.131442 | 4.648998 | 3.34E-06 | 1.98E-05 | 1.527397 |
| PHACTR1 | 213.4738 | 0.611268 | 0.196577 | 3.109566 | 0.001874 | 0.006013 | 1.527602 |
| HSP90AA1 | 33436.8 | 0.61147 | 0.052216 | 11.71033 | 1.13E-31 | 1.25E-29 | 1.527815 |
| PFKM | 2406.213 | 0.611532 | 0.089609 | 6.824476 | 8.82E-12 | 1.35E-10 | 1.52788 |
| EFL1 | 978.3166 | 0.611544 | 0.095209 | 6.423203 | 1.33E-10 | 1.71E-09 | 1.527893 |
| ADGRL2 | 5355.907 | 0.611569 | 0.132889 | 4.60209 | 4.18E-06 | 2.44E-05 | 1.52792 |
| AHCTF1 | 2164.87 | 0.611762 | 0.073903 | 8.277861 | 1.25E-16 | 3.51E-15 | 1.528124 |
| PHOSPHO | 116.2569 | 0.611799 | 0.261142 | 2.342781 | 0.019141 | 0.045638 | 1.528164 |
| RASSF2 | 415.4736 | 0.612152 | 0.158995 | 3.850144 | 0.000118 | 0.000508 | 1.528537 |
| ACYP1 | 124.8681 | 0.612383 | 0.198479 | 3.085384 | 0.002033 | 0.006452 | 1.528783 |
| FAM172A | 334.1268 | 0.612624 | 0.165285 | 3.70647 | 0.00021 | 0.00085 | 1.529037 |
| MEST | 4445.735 | 0.612694 | 0.098915 | 6.194163 | 5.86E-10 | 6.76E-09 | 1.529112 |
| DEK | 8187.844 | 0.612774 | 0.063251 | 9.687983 | 3.39E-22 | 1.59E-20 | 1.529197 |
| CEP57L1 | 256.9772 | 0.612855 | 0.197791 | 3.098499 | 0.001945 | 0.006214 | 1.529283 |
| AP003108. | 142.1474 | 0.612942 | 0.236639 | 2.590204 | 0.009592 | 0.025091 | 1.529375 |
| HEATR5A | 794.727 | 0.613136 | 0.165307 | 3.709082 | 0.000208 | 0.000842 | 1.529581 |
| PDS5B | 1317.754 | 0.613329 | 0.133071 | 4.609029 | 4.05E-06 | 2.37E-05 | 1.529785 |
| SEL1L | 1149.145 | 0.614204 | 0.086985 | 7.061051 | 1.65E-12 | 2.78E-11 | 1.530713 |
| ZFP69B | 183.4247 | 0.614515 | 0.149262 | 4.117019 | 3.84E-05 | 0.000185 | 1.531043 |
| OTUB2 | 145.7482 | 0.614719 | 0.241541 | 2.544988 | 0.010928 | 0.028075 | 1.53126 |
| DERPC | 675.0474 | 0.614848 | 0.196158 | 3.134451 | 0.001722 | 0.005573 | 1.531396 |
| AGO3 | 671.4699 | 0.614978 | 0.190857 | 3.222196 | 0.001272 | 0.004252 | 1.531535 |
| ZNF585A | 210.684 | 0.615325 | 0.170379 | 3.611514 | 0.000304 | 0.001189 | 1.531903 |
| GKAP1 | 189.4721 | 0.615414 | 0.141773 | 4.340847 | 1.42E-05 | 7.41E-05 | 1.531997 |
| GINS3 | 337.3931 | 0.615434 | 0.112142 | 5.487989 | 4.07E-08 | 3.49E-07 | 1.532019 |
| HSDL2 | 532.3215 | 0.615483 | 0.139904 | 4.39931 | 1.09E-05 | 5.81E-05 | 1.53207 |
| TP53BP2 | 1743.452 | 0.615665 | 0.070104 | 8.782178 | 1.60E-18 | 5.40E-17 | 1.532264 |
| ATAD5 | 866.0323 | 0.61604 | 0.097098 | 6.344519 | 2.23E-10 | 2.77E-09 | 1.532662 |
| GPSM2 | 425.4337 | 0.616267 | 0.200947 | 3.066811 | 0.002164 | 0.006809 | 1.532904 |
| USP42 | 625.4958 | 0.616395 | 0.102731 | 6.000116 | 1.97E-09 | 2.08E-08 | 1.53304 |
| AKT3 | 654.0328 | 0.616469 | 0.146511 | 4.207667 | 2.58E-05 | 0.000129 | 1.533118 |
| CHUK | 566.5378 | 0.616499 | 0.097372 | 6.331389 | 2.43E-10 | 3.00E-09 | 1.53315 |
| FBXW2 | 1731.06 | 0.617058 | 0.117414 | 5.255392 | 1.48E-07 | 1.14E-06 | 1.533745 |
| KLHL13 | 807.4711 | 0.617483 | 0.09801 | 6.300226 | 2.97E-10 | 3.61E-09 | 1.534196 |
| POLA1 | 2785.675 | 0.617514 | 0.068238 | 9.049439 | 1.44E-19 | 5.36E-18 | 1.534229 |
| ID3 | 2105.15 | 0.617711 | 0.084393 | 7.319459 | 2.49E-13 | 4.61E-12 | 1.534438 |
| KIAA0355 | 1195.075 | 0.617776 | 0.100806 | 6.128383 | 8.88E-10 | 9.91E-09 | 1.534508 |
| CCDC85C | 1866.33 | 0.617998 | 0.089285 | 6.921599 | 4.47E-12 | 7.11E-11 | 1.534744 |
| MELK | 1543.801 | 0.618297 | 0.108584 | 5.694168 | 1.24E-08 | 1.15E-07 | 1.535062 |
| CHD6 | 2034.592 | 0.618357 | 0.096942 | 6.378617 | 1.79E-10 | 2.25E-09 | 1.535126 |
| IRX3 | 345.0396 | 0.618389 | 0.234046 | 2.642162 | 0.008238 | 0.021962 | 1.53516 |

| | | | | | | | |
|---|---|---|---|---|---|---|---|
| CBLB | 944.8293 | 0.618403 | 0.138195 | 4.474869 | 7.65E-06 | 4.24E-05 | 1.535175 |
| ARPP19 | 6617.581 | 0.618559 | 0.092826 | 6.663677 | 2.67E-11 | 3.77E-10 | 1.535341 |
| MSL3 | 444.6283 | 0.618573 | 0.15035 | 4.114226 | 3.88E-05 | 0.000187 | 1.535356 |
| YOD1 | 521.7483 | 0.618932 | 0.096456 | 6.416734 | 1.39E-10 | 1.78E-09 | 1.535738 |
| CENPV | 1937.524 | 0.619547 | 0.092463 | 6.700506 | 2.08E-11 | 2.98E-10 | 1.536393 |
| TAB3 | 560.5704 | 0.619699 | 0.161664 | 3.833258 | 0.000126 | 0.000538 | 1.536555 |
| DBR1 | 517.2795 | 0.620305 | 0.105369 | 5.88698 | 3.93E-09 | 3.94E-08 | 1.5372 |
| AC093752 | 220.5207 | 0.62056 | 0.138514 | 4.480138 | 7.46E-06 | 4.15E-05 | 1.537472 |
| PGM3 | 1401.52 | 0.620886 | 0.102258 | 6.071781 | 1.26E-09 | 1.38E-08 | 1.537819 |
| STMN1 | 9202.947 | 0.620994 | 0.075425 | 8.233284 | 1.82E-16 | 4.99E-15 | 1.537934 |
| 5-Mar | 517.6032 | 0.621008 | 0.150215 | 4.134122 | 3.56E-05 | 0.000173 | 1.537949 |
| FKBP5 | 2005.278 | 0.621098 | 0.12092 | 5.136448 | 2.80E-07 | 2.05E-06 | 1.538045 |
| SKP1 | 4441.245 | 0.621229 | 0.081388 | 7.632887 | 2.30E-14 | 4.84E-13 | 1.538184 |
| PPIP5K2 | 1232.028 | 0.621302 | 0.134658 | 4.613923 | 3.95E-06 | 2.32E-05 | 1.538263 |
| TFIP11 | 746.5609 | 0.621364 | 0.117176 | 5.302807 | 1.14E-07 | 9.01E-07 | 1.538329 |
| USP1 | 2274.877 | 0.621497 | 0.058733 | 10.58182 | 3.62E-26 | 2.55E-24 | 1.53847 |
| AS3MT | 645.0524 | 0.62161 | 0.097699 | 6.362486 | 1.99E-10 | 2.48E-09 | 1.538591 |
| EEF1AKMT | 483.3689 | 0.62234 | 0.131333 | 4.73865 | 2.15E-06 | 1.33E-05 | 1.53937 |
| TGFBRAP1 | 736.7363 | 0.622375 | 0.091259 | 6.819896 | 9.11E-12 | 1.39E-10 | 1.539407 |
| MAN1A2 | 848.9104 | 0.622698 | 0.122576 | 5.080118 | 3.77E-07 | 2.68E-06 | 1.539752 |
| KATNBL1 | 665.4624 | 0.622821 | 0.102424 | 6.080787 | 1.20E-09 | 1.31E-08 | 1.539883 |
| IFT52 | 978.134 | 0.623205 | 0.104827 | 5.945089 | 2.76E-09 | 2.84E-08 | 1.540294 |
| RNF182 | 299.6187 | 0.623446 | 0.154922 | 4.024255 | 5.72E-05 | 0.000265 | 1.54055 |
| ZNF106 | 2486.613 | 0.623717 | 0.132206 | 4.717772 | 2.38E-06 | 1.46E-05 | 1.54084 |
| MSH6 | 6176.926 | 0.623735 | 0.07625 | 8.180147 | 2.83E-16 | 7.50E-15 | 1.540859 |
| OSCP1 | 86.18563 | 0.624219 | 0.223598 | 2.791707 | 0.005243 | 0.014862 | 1.541377 |
| SLC5A3 | 885.9885 | 0.624344 | 0.149102 | 4.187371 | 2.82E-05 | 0.00014 | 1.54151 |
| ZNF445 | 1726.713 | 0.624819 | 0.09336 | 6.692611 | 2.19E-11 | 3.13E-10 | 1.542017 |
| KIRREL1 | 1883.23 | 0.624978 | 0.083397 | 7.49403 | 6.68E-14 | 1.33E-12 | 1.542187 |
| GTPBP8 | 427.6199 | 0.625343 | 0.150416 | 4.157427 | 3.22E-05 | 0.000158 | 1.542577 |
| TMEM35A | 243.2173 | 0.625538 | 0.239197 | 2.615158 | 0.008919 | 0.023568 | 1.542786 |
| FARP2 | 709.7158 | 0.625668 | 0.120047 | 5.211868 | 1.87E-07 | 1.41E-06 | 1.542925 |
| HARBI1 | 113.6216 | 0.625759 | 0.227657 | 2.748689 | 0.005983 | 0.016715 | 1.543022 |
| TUBA1B | 44637.15 | 0.626085 | 0.061911 | 10.11275 | 4.85E-24 | 2.82E-22 | 1.543372 |
| CNN3 | 5731.583 | 0.626279 | 0.061488 | 10.18539 | 2.30E-24 | 1.40E-22 | 1.543578 |
| XKR5 | 222.6093 | 0.626329 | 0.128526 | 4.87316 | 1.10E-06 | 7.17E-06 | 1.543632 |
| ZKSCAN8 | 1116.591 | 0.627038 | 0.124176 | 5.049587 | 4.43E-07 | 3.11E-06 | 1.544391 |
| PXMP4 | 113.034 | 0.627398 | 0.219203 | 2.862184 | 0.004207 | 0.012224 | 1.544777 |
| CA2 | 183.3572 | 0.627461 | 0.166514 | 3.768207 | 0.000164 | 0.000681 | 1.544844 |
| ZFP14 | 320.6385 | 0.627789 | 0.175869 | 3.569647 | 0.000357 | 0.001378 | 1.545196 |
| DDX47 | 1419.98 | 0.627833 | 0.080579 | 7.791537 | 6.62E-15 | 1.49E-13 | 1.545242 |
| L3MBTL3 | 418.341 | 0.628074 | 0.210538 | 2.98318 | 0.002853 | 0.008706 | 1.5455 |
| RNF11 | 1591.109 | 0.629016 | 0.128321 | 4.90191 | 9.49E-07 | 6.27E-06 | 1.54651 |
| ZNF629 | 962.0052 | 0.629187 | 0.100889 | 6.236437 | 4.48E-10 | 5.27E-09 | 1.546693 |

| | | | | | | | |
|---|---|---|---|---|---|---|---|
| GLOD4 | 1837.478 | 0.629205 | 0.073612 | 8.547613 | 1.26E-17 | 3.95E-16 | 1.546712 |
| TSPY26P | 416.3883 | 0.62922 | 0.141999 | 4.431154 | 9.37E-06 | 5.10E-05 | 1.546728 |
| TNFAIP8 | 421.4801 | 0.629338 | 0.159552 | 3.944398 | 8.00E-05 | 0.000359 | 1.546855 |
| CAMKMT | 335.2607 | 0.62991 | 0.117264 | 5.371701 | 7.80E-08 | 6.36E-07 | 1.547468 |
| GANC | 259.9015 | 0.630073 | 0.219887 | 2.865444 | 0.004164 | 0.01212 | 1.547643 |
| AHDC1 | 935.6461 | 0.630094 | 0.146217 | 4.309308 | 1.64E-05 | 8.45E-05 | 1.547665 |
| TUBB | 42647.48 | 0.630788 | 0.07788 | 8.099494 | 5.52E-16 | 1.41E-14 | 1.54841 |
| ID4 | 546.3349 | 0.631586 | 0.086867 | 7.270701 | 3.58E-13 | 6.48E-12 | 1.549267 |
| C9orf72 | 757.0223 | 0.631862 | 0.126776 | 4.984078 | 6.23E-07 | 4.26E-06 | 1.549564 |
| EEF1AKNM | 1322.396 | 0.632098 | 0.090309 | 6.999245 | 2.57E-12 | 4.23E-11 | 1.549817 |
| RNF185 | 973.1731 | 0.632134 | 0.086933 | 7.271551 | 3.55E-13 | 6.45E-12 | 1.549856 |
| MYLK | 902.6755 | 0.632386 | 0.158868 | 3.980578 | 6.87E-05 | 0.000312 | 1.550126 |
| WDCP | 833.2577 | 0.632494 | 0.119733 | 5.282527 | 1.27E-07 | 9.94E-07 | 1.550242 |
| SHPRH | 1212.441 | 0.632509 | 0.085653 | 7.384547 | 1.53E-13 | 2.90E-12 | 1.550259 |
| TMEM60 | 326.284 | 0.633322 | 0.136555 | 4.637866 | 3.52E-06 | 2.08E-05 | 1.551133 |
| NECAB2 | 722.0255 | 0.6335 | 0.148846 | 4.256067 | 2.08E-05 | 0.000106 | 1.551324 |
| KBTBD2 | 879.3462 | 0.633579 | 0.120379 | 5.263184 | 1.42E-07 | 1.10E-06 | 1.551409 |
| MFAP1 | 1130.399 | 0.633664 | 0.079123 | 8.008551 | 1.16E-15 | 2.84E-14 | 1.5515 |
| KLHL23 | 2190.985 | 0.633879 | 0.097776 | 6.482956 | 8.99E-11 | 1.19E-09 | 1.551732 |
| TRIL | 292.8912 | 0.634385 | 0.12656 | 5.012527 | 5.37E-07 | 3.71E-06 | 1.552276 |
| SLCO1A2 | 322.4703 | 0.634569 | 0.172433 | 3.680082 | 0.000233 | 0.000934 | 1.552474 |
| ZNF197 | 548.1193 | 0.634715 | 0.136296 | 4.656877 | 3.21E-06 | 1.91E-05 | 1.552631 |
| WWOX | 253.6682 | 0.635449 | 0.152249 | 4.173743 | 3.00E-05 | 0.000148 | 1.553421 |
| DNAL1 | 558.9756 | 0.635653 | 0.146797 | 4.330139 | 1.49E-05 | 7.74E-05 | 1.553641 |
| TBK1 | 793.8064 | 0.635729 | 0.118713 | 5.355172 | 8.55E-08 | 6.93E-07 | 1.553723 |
| ATL2 | 1955.655 | 0.636139 | 0.109372 | 5.816313 | 6.02E-09 | 5.85E-08 | 1.554164 |
| MTF1 | 365.8504 | 0.637248 | 0.190956 | 3.33715 | 0.000846 | 0.002965 | 1.555359 |
| TOP1 | 3811.29 | 0.637433 | 0.068357 | 9.325016 | 1.11E-20 | 4.61E-19 | 1.555559 |
| NEK2 | 771.366 | 0.63775 | 0.110192 | 5.787628 | 7.14E-09 | 6.91E-08 | 1.555901 |
| RPL21 | 10275.11 | 0.638085 | 0.05519 | 11.56156 | 6.45E-31 | 6.63E-29 | 1.556262 |
| DIP2C | 593.9139 | 0.638122 | 0.101146 | 6.308954 | 2.81E-10 | 3.43E-09 | 1.556302 |
| PGAP1 | 1817.772 | 0.638237 | 0.104478 | 6.108823 | 1.00E-09 | 1.11E-08 | 1.556426 |
| PCSK1 | 167.165 | 0.638308 | 0.190915 | 3.343418 | 0.000828 | 0.002905 | 1.556503 |
| SYDE2 | 222.9819 | 0.638349 | 0.171003 | 3.73297 | 0.000189 | 0.000774 | 1.556547 |
| TTC28 | 911.1709 | 0.638461 | 0.211338 | 3.021044 | 0.002519 | 0.007793 | 1.556668 |
| ASPM | 2326.971 | 0.638588 | 0.150546 | 4.241814 | 2.22E-05 | 0.000112 | 1.556805 |
| VPS26A | 1446.302 | 0.639061 | 0.109448 | 5.838955 | 5.25E-09 | 5.16E-08 | 1.557316 |
| PHF20L1 | 786.4233 | 0.639268 | 0.137569 | 4.646882 | 3.37E-06 | 2.00E-05 | 1.557539 |
| ARNT | 983.2044 | 0.639295 | 0.084509 | 7.564838 | 3.88E-14 | 7.96E-13 | 1.557568 |
| HNRNPA1I | 342.8281 | 0.639578 | 0.111943 | 5.713426 | 1.11E-08 | 1.04E-07 | 1.557874 |
| SRCAP | 2550.495 | 0.639661 | 0.088162 | 7.255556 | 4.00E-13 | 7.21E-12 | 1.557963 |
| TMSB15B | 382.6109 | 0.640006 | 0.143366 | 4.464131 | 8.04E-06 | 4.43E-05 | 1.558335 |
| SREK1 | 1702.021 | 0.640093 | 0.086852 | 7.369942 | 1.71E-13 | 3.23E-12 | 1.55843 |
| SESTD1 | 799.8 | 0.640242 | 0.140393 | 4.560342 | 5.11E-06 | 2.93E-05 | 1.55859 |

| | | | | | | | |
|---|---|---|---|---|---|---|---|
| HLCS | 864.4824 | 0.640362 | 0.134187 | 4.772161 | 1.82E-06 | 1.14E-05 | 1.55872 |
| NCAPD2 | 4775.875 | 0.640578 | 0.067314 | 9.516314 | 1.79E-21 | 7.91E-20 | 1.558954 |
| TUBGCP4 | 582.5077 | 0.640854 | 0.133803 | 4.789535 | 1.67E-06 | 1.06E-05 | 1.559252 |
| EML4 | 3431.162 | 0.641073 | 0.087615 | 7.316937 | 2.54E-13 | 4.70E-12 | 1.559489 |
| CWF19L2 | 512.9505 | 0.641205 | 0.112023 | 5.72389 | 1.04E-08 | 9.81E-08 | 1.559632 |
| STON2 | 287.3739 | 0.641276 | 0.174588 | 3.673085 | 0.00024 | 0.000957 | 1.559709 |
| PNMA8A | 1334.296 | 0.641633 | 0.1025 | 6.259811 | 3.85E-10 | 4.61E-09 | 1.560094 |
| RALGAPB | 2273.38 | 0.641781 | 0.105859 | 6.062601 | 1.34E-09 | 1.45E-08 | 1.560254 |
| EIF4E | 3152.415 | 0.642474 | 0.073087 | 8.790546 | 1.49E-18 | 5.05E-17 | 1.561004 |
| SFXN1 | 2202.173 | 0.643421 | 0.117425 | 5.479407 | 4.27E-08 | 3.65E-07 | 1.562029 |
| GPR162 | 275.9028 | 0.643641 | 0.209654 | 3.070013 | 0.00214 | 0.006754 | 1.562267 |
| PANK1 | 799.0744 | 0.644552 | 0.099263 | 6.4934 | 8.39E-11 | 1.12E-09 | 1.563254 |
| DYNC2LI1 | 255.3382 | 0.644697 | 0.127316 | 5.063742 | 4.11E-07 | 2.91E-06 | 1.56341 |
| MDM1 | 364.5437 | 0.64471 | 0.137272 | 4.696582 | 2.65E-06 | 1.60E-05 | 1.563425 |
| MVK | 221.0095 | 0.64491 | 0.20285 | 3.179247 | 0.001477 | 0.004861 | 1.563641 |
| DUSP11 | 387.6852 | 0.645752 | 0.131211 | 4.921462 | 8.59E-07 | 5.74E-06 | 1.564555 |
| PPA2 | 1002.515 | 0.645842 | 0.098761 | 6.539457 | 6.17E-11 | 8.40E-10 | 1.564652 |
| SENP5 | 674.2193 | 0.645947 | 0.125032 | 5.166254 | 2.39E-07 | 1.77E-06 | 1.564766 |
| ANP32E | 5409.57 | 0.646478 | 0.059658 | 10.8364 | 2.31E-27 | 1.74E-25 | 1.565342 |
| NSD1 | 5832.574 | 0.646676 | 0.091198 | 7.090871 | 1.33E-12 | 2.27E-11 | 1.565557 |
| RXRA | 1575.355 | 0.647656 | 0.071957 | 9.000659 | 2.24E-19 | 8.17E-18 | 1.566621 |
| NFRKB | 1185.609 | 0.647688 | 0.11359 | 5.701976 | 1.18E-08 | 1.10E-07 | 1.566655 |
| ETFRF1 | 134.5292 | 0.647916 | 0.184626 | 3.509339 | 0.000449 | 0.001688 | 1.566903 |
| TRIM33 | 2681.946 | 0.648142 | 0.117598 | 5.511498 | 3.56E-08 | 3.09E-07 | 1.567148 |
| LCORL | 543.4494 | 0.648319 | 0.159406 | 4.067103 | 4.76E-05 | 0.000225 | 1.567341 |
| ASXL2 | 1327.62 | 0.648744 | 0.105608 | 6.142932 | 8.10E-10 | 9.12E-09 | 1.567803 |
| SNX4 | 896.0555 | 0.649212 | 0.089537 | 7.250772 | 4.14E-13 | 7.46E-12 | 1.568311 |
| STRN3 | 1129.562 | 0.649527 | 0.143895 | 4.513886 | 6.37E-06 | 3.59E-05 | 1.568654 |
| H2AFZ | 9228.995 | 0.649889 | 0.078128 | 8.318237 | 8.93E-17 | 2.56E-15 | 1.569048 |
| SORCS1 | 367.2733 | 0.649908 | 0.243746 | 2.666334 | 0.007668 | 0.020634 | 1.569069 |
| CIP2A | 1061.35 | 0.649927 | 0.103735 | 6.265244 | 3.72E-10 | 4.46E-09 | 1.569088 |
| DICER1 | 2654.484 | 0.650006 | 0.092747 | 7.008374 | 2.41E-12 | 3.98E-11 | 1.569175 |
| VPS36 | 830.3403 | 0.650054 | 0.128074 | 5.075594 | 3.86E-07 | 2.74E-06 | 1.569227 |
| TAF5 | 532.3846 | 0.650183 | 0.110848 | 5.865522 | 4.48E-09 | 4.45E-08 | 1.569367 |
| KIF4A | 1811.011 | 0.650425 | 0.06698 | 9.71072 | 2.71E-22 | 1.29E-20 | 1.56963 |
| MRTFB | 828.0315 | 0.650581 | 0.156887 | 4.146808 | 3.37E-05 | 0.000164 | 1.569801 |
| HELZ | 1455.084 | 0.650878 | 0.089821 | 7.24636 | 4.28E-13 | 7.69E-12 | 1.570123 |
| FAM160B1 | 539.2929 | 0.651837 | 0.100879 | 6.461565 | 1.04E-10 | 1.35E-09 | 1.571167 |
| CTTNBP2N | 862.5824 | 0.651943 | 0.11957 | 5.45238 | 4.97E-08 | 4.20E-07 | 1.571282 |
| DEPDC1 | 1413.566 | 0.65195 | 0.122582 | 5.318483 | 1.05E-07 | 8.35E-07 | 1.571291 |
| POGK | 1572.18 | 0.65237 | 0.109208 | 5.973664 | 2.32E-09 | 2.41E-08 | 1.571748 |
| SEC24B | 939.4837 | 0.652433 | 0.096404 | 6.767666 | 1.31E-11 | 1.93E-10 | 1.571817 |
| DHX8 | 1413.277 | 0.654192 | 0.15173 | 4.311564 | 1.62E-05 | 8.39E-05 | 1.573735 |
| ZBTB21 | 773.1446 | 0.654331 | 0.148962 | 4.392611 | 1.12E-05 | 5.98E-05 | 1.573886 |

| | | | | | | | |
|---|---|---|---|---|---|---|---|
| ATMIN | 1916.666 | 0.654484 | 0.066599 | 9.827294 | 8.59E-23 | 4.33E-21 | 1.574053 |
| USP6NL | 1093.051 | 0.654539 | 0.09307 | 7.032786 | 2.02E-12 | 3.38E-11 | 1.574113 |
| MTM1 | 777.6163 | 0.654707 | 0.122579 | 5.341093 | 9.24E-08 | 7.45E-07 | 1.574296 |
| PIMREG | 1715.408 | 0.654813 | 0.088037 | 7.437889 | 1.02E-13 | 2.00E-12 | 1.574412 |
| CSRNP3 | 175.7283 | 0.655184 | 0.240476 | 2.724533 | 0.006439 | 0.017783 | 1.574817 |
| C11orf49 | 604.124 | 0.655609 | 0.126798 | 5.170494 | 2.33E-07 | 1.74E-06 | 1.575281 |
| IFT74 | 471.6918 | 0.655687 | 0.137887 | 4.755257 | 1.98E-06 | 1.23E-05 | 1.575366 |
| FNDC3A | 1423.418 | 0.65586 | 0.092761 | 7.070447 | 1.54E-12 | 2.61E-11 | 1.575555 |
| KIF2A | 1708.779 | 0.656105 | 0.136635 | 4.801883 | 1.57E-06 | 9.98E-06 | 1.575822 |
| ZBTB5 | 515.2861 | 0.656323 | 0.106857 | 6.14206 | 8.15E-10 | 9.16E-09 | 1.576061 |
| SMARCAD1 | 2141.54 | 0.656543 | 0.065943 | 9.956171 | 2.37E-23 | 1.27E-21 | 1.576301 |
| ETV1 | 1875.655 | 0.656886 | 0.126994 | 5.17257 | 2.31E-07 | 1.72E-06 | 1.576676 |
| NASP | 14397.25 | 0.657094 | 0.08132 | 8.080315 | 6.46E-16 | 1.64E-14 | 1.576903 |
| AREL1 | 1231.661 | 0.657651 | 0.111657 | 5.889921 | 3.86E-09 | 3.88E-08 | 1.577512 |
| TCHP | 380.1944 | 0.657903 | 0.138007 | 4.767164 | 1.87E-06 | 1.17E-05 | 1.577787 |
| SNAPC3 | 932.972 | 0.658673 | 0.094669 | 6.957644 | 3.46E-12 | 5.58E-11 | 1.57863 |
| STXBP4 | 413.9054 | 0.65914 | 0.200369 | 3.289631 | 0.001003 | 0.003448 | 1.579142 |
| AURKA | 1636.158 | 0.660297 | 0.122754 | 5.379024 | 7.49E-08 | 6.12E-07 | 1.580408 |
| USP49 | 1226.467 | 0.660386 | 0.117661 | 5.612602 | 1.99E-08 | 1.79E-07 | 1.580505 |
| ZNF438 | 60.70368 | 0.660442 | 0.273093 | 2.418374 | 0.01559 | 0.038245 | 1.580566 |
| THAP5 | 475.059 | 0.660555 | 0.145112 | 4.552051 | 5.31E-06 | 3.03E-05 | 1.580691 |
| RIT1 | 627.0185 | 0.66071 | 0.105569 | 6.258535 | 3.89E-10 | 4.64E-09 | 1.580861 |
| KIF2C | 2142.849 | 0.661058 | 0.074492 | 8.874231 | 7.04E-19 | 2.47E-17 | 1.581242 |
| AR | 323.1298 | 0.661116 | 0.190482 | 3.470751 | 0.000519 | 0.001918 | 1.581306 |
| PUS10 | 121.6141 | 0.66133 | 0.266497 | 2.481569 | 0.013081 | 0.032817 | 1.58154 |
| BZW1 | 6617.708 | 0.661438 | 0.093112 | 7.103688 | 1.21E-12 | 2.08E-11 | 1.581659 |
| RAB11FIP2 | 651.2314 | 0.661703 | 0.221596 | 2.986085 | 0.002826 | 0.00863 | 1.581949 |
| KPNA1 | 1355.642 | 0.662068 | 0.102405 | 6.46518 | 1.01E-10 | 1.33E-09 | 1.582349 |
| ZBTB40 | 738.7347 | 0.662256 | 0.116487 | 5.685236 | 1.31E-08 | 1.21E-07 | 1.582556 |
| HNRNPH2 | 2344.598 | 0.66244 | 0.090317 | 7.334597 | 2.22E-13 | 4.15E-12 | 1.582757 |
| KNL1 | 1237.774 | 0.662765 | 0.150998 | 4.389238 | 1.14E-05 | 6.06E-05 | 1.583113 |
| VAMP4 | 231.4836 | 0.662786 | 0.152777 | 4.338254 | 1.44E-05 | 7.49E-05 | 1.583136 |
| ARHGAP19 | 1965.887 | 0.663004 | 0.098646 | 6.721047 | 1.80E-11 | 2.62E-10 | 1.583376 |
| TNS3 | 903.3496 | 0.663547 | 0.084434 | 7.858767 | 3.88E-15 | 8.99E-14 | 1.583972 |
| 11-Sep | 5384.805 | 0.664332 | 0.087615 | 7.582389 | 3.39E-14 | 6.99E-13 | 1.584834 |
| METTL25 | 106.5282 | 0.665172 | 0.195671 | 3.399438 | 0.000675 | 0.002426 | 1.585758 |
| DHX32 | 773.2196 | 0.665999 | 0.107613 | 6.188844 | 6.06E-10 | 6.97E-09 | 1.586666 |
| CDC5L | 1814.504 | 0.666012 | 0.081246 | 8.197486 | 2.45E-16 | 6.58E-15 | 1.586681 |
| GPHN | 387.5008 | 0.666521 | 0.148978 | 4.473954 | 7.68E-06 | 4.26E-05 | 1.58724 |
| CACNB3 | 700.7503 | 0.667012 | 0.169945 | 3.924861 | 8.68E-05 | 0.000386 | 1.587781 |
| GTF3C3 | 1452.907 | 0.667262 | 0.123697 | 5.39433 | 6.88E-08 | 5.66E-07 | 1.588056 |
| ST13 | 8296.919 | 0.667276 | 0.072591 | 9.192301 | 3.85E-20 | 1.52E-18 | 1.588072 |
| PUM1 | 3508.103 | 0.667513 | 0.063488 | 10.51404 | 7.44E-26 | 5.03E-24 | 1.588332 |
| NSMCE2 | 312.2761 | 0.668461 | 0.111217 | 6.010452 | 1.85E-09 | 1.96E-08 | 1.589377 |

| | | | | | | | |
|---|---|---|---|---|---|---|---|
| ZNF83 | 493.5888 | 0.668574 | 0.143045 | 4.67386 | 2.96E-06 | 1.77E-05 | 1.589501 |
| PPP2R5E | 2072.535 | 0.668705 | 0.135578 | 4.932266 | 8.13E-07 | 5.45E-06 | 1.589645 |
| SNRK | 503.2845 | 0.669018 | 0.131391 | 5.091817 | 3.55E-07 | 2.53E-06 | 1.58999 |
| PCDH19 | 231.4977 | 0.669424 | 0.248537 | 2.693464 | 0.007071 | 0.019257 | 1.590438 |
| KLHL15 | 787.196 | 0.670135 | 0.117024 | 5.726465 | 1.03E-08 | 9.68E-08 | 1.591221 |
| SEZ6 | 725.4783 | 0.670472 | 0.276464 | 2.425173 | 0.015301 | 0.037611 | 1.591593 |
| UBE2W | 612.3684 | 0.670882 | 0.182653 | 3.672993 | 0.00024 | 0.000957 | 1.592046 |
| LRRN2 | 127.3724 | 0.671748 | 0.256949 | 2.614326 | 0.00894 | 0.023612 | 1.593002 |
| CDKN1B | 1196.19 | 0.672012 | 0.079561 | 8.446451 | 3.00E-17 | 9.06E-16 | 1.593293 |
| KBTBD4 | 297.128 | 0.672243 | 0.142772 | 4.708506 | 2.50E-06 | 1.52E-05 | 1.593548 |
| SOCS6 | 746.957 | 0.672439 | 0.108132 | 6.218698 | 5.01E-10 | 5.85E-09 | 1.593766 |
| TMSB10 | 5127.916 | 0.672452 | 0.165634 | 4.059855 | 4.91E-05 | 0.000231 | 1.593779 |
| SLC35G1 | 287.2674 | 0.672523 | 0.174811 | 3.847135 | 0.00012 | 0.000513 | 1.593858 |
| SERTAD2 | 272.1853 | 0.67281 | 0.152729 | 4.405264 | 1.06E-05 | 5.67E-05 | 1.594175 |
| TEKT2 | 55.27707 | 0.672835 | 0.28237 | 2.382812 | 0.017181 | 0.041691 | 1.594203 |
| HMGB1 | 12434.31 | 0.672841 | 0.072117 | 9.329789 | 1.06E-20 | 4.42E-19 | 1.594209 |
| GPATCH2 | 462.1621 | 0.672886 | 0.177657 | 3.787562 | 0.000152 | 0.000636 | 1.594259 |
| ERCC6 | 313.7491 | 0.67306 | 0.151667 | 4.437752 | 9.09E-06 | 4.96E-05 | 1.594451 |
| CENPC | 556.2026 | 0.673335 | 0.189489 | 3.553434 | 0.00038 | 0.001456 | 1.594755 |
| IFIT5 | 155.4805 | 0.673566 | 0.22832 | 2.950092 | 0.003177 | 0.009565 | 1.595011 |
| PNPLA8 | 628.8073 | 0.673576 | 0.114385 | 5.888663 | 3.89E-09 | 3.90E-08 | 1.595022 |
| LRRC4B | 1013.975 | 0.674917 | 0.18924 | 3.566457 | 0.000362 | 0.001393 | 1.596505 |
| ZNF823 | 64.19467 | 0.674974 | 0.246245 | 2.741071 | 0.006124 | 0.017058 | 1.596568 |
| PNRC2 | 2326.413 | 0.675428 | 0.085998 | 7.853959 | 4.03E-15 | 9.28E-14 | 1.59707 |
| MCM8 | 1429.838 | 0.675961 | 0.099729 | 6.777948 | 1.22E-11 | 1.81E-10 | 1.59766 |
| FBXO21 | 1573.055 | 0.676159 | 0.114383 | 5.911372 | 3.39E-09 | 3.44E-08 | 1.59788 |
| EFNA5 | 1143.812 | 0.676205 | 0.129394 | 5.225949 | 1.73E-07 | 1.32E-06 | 1.597931 |
| ZNF702P | 224.638 | 0.676243 | 0.216561 | 3.122644 | 0.001792 | 0.005777 | 1.597973 |
| BRD3OS | 552.5904 | 0.676787 | 0.11106 | 6.093872 | 1.10E-09 | 1.21E-08 | 1.598576 |
| ZNF17 | 112.7169 | 0.677815 | 0.279653 | 2.423771 | 0.01536 | 0.037742 | 1.599715 |
| OTUD3 | 583.3543 | 0.677918 | 0.12081 | 5.611421 | 2.01E-08 | 1.81E-07 | 1.599829 |
| KIAA1324L | 1586.23 | 0.678104 | 0.094781 | 7.154466 | 8.40E-13 | 1.46E-11 | 1.600036 |
| FAM229B | 175.4756 | 0.678269 | 0.204875 | 3.310652 | 0.000931 | 0.003226 | 1.600219 |
| C5orf24 | 727.6759 | 0.678741 | 0.144042 | 4.712107 | 2.45E-06 | 1.50E-05 | 1.600742 |
| CCDC117 | 1403.587 | 0.678843 | 0.079029 | 8.58982 | 8.71E-18 | 2.76E-16 | 1.600855 |
| LYRM2 | 502.6015 | 0.678986 | 0.127202 | 5.337877 | 9.40E-08 | 7.58E-07 | 1.601015 |
| IFT140 | 1246.611 | 0.679726 | 0.176153 | 3.858733 | 0.000114 | 0.000493 | 1.601835 |
| SENP1 | 921.8466 | 0.680944 | 0.128198 | 5.311668 | 1.09E-07 | 8.62E-07 | 1.603189 |
| RLF | 1082.369 | 0.68108 | 0.09512 | 7.1602 | 8.06E-13 | 1.40E-11 | 1.60334 |
| ARMC1 | 1127.203 | 0.681407 | 0.08484 | 8.031692 | 9.61E-16 | 2.38E-14 | 1.603703 |
| ARPIN | 580.3553 | 0.681703 | 0.14727 | 4.628929 | 3.68E-06 | 2.17E-05 | 1.604032 |
| SPOP | 628.9751 | 0.681713 | 0.140538 | 4.850721 | 1.23E-06 | 7.97E-06 | 1.604043 |
| DDIAS | 698.3615 | 0.682125 | 0.092234 | 7.395582 | 1.41E-13 | 2.70E-12 | 1.604501 |
| RBBP9 | 505.6839 | 0.682245 | 0.127024 | 5.371014 | 7.83E-08 | 6.38E-07 | 1.604635 |

| | | | | | | | |
|---|---|---|---|---|---|---|---|
| TBC1D5 | 1553.798 | 0.68304 | 0.179662 | 3.801807 | 0.000144 | 0.000604 | 1.605519 |
| DCTN6 | 468.8843 | 0.683286 | 0.123748 | 5.521585 | 3.36E-08 | 2.93E-07 | 1.605793 |
| STOM | 1179.832 | 0.683478 | 0.181871 | 3.758038 | 0.000171 | 0.000706 | 1.606007 |
| SHCBP1 | 465.6515 | 0.684776 | 0.135195 | 5.065105 | 4.08E-07 | 2.89E-06 | 1.607452 |
| MAP2K4 | 1034.288 | 0.685275 | 0.087737 | 7.810561 | 5.69E-15 | 1.30E-13 | 1.608009 |
| TCF4 | 2265.858 | 0.685969 | 0.173645 | 3.95041 | 7.80E-05 | 0.00035 | 1.608782 |
| DLGAP5 | 3476.184 | 0.686636 | 0.080432 | 8.536902 | 1.38E-17 | 4.31E-16 | 1.609526 |
| G3BP1 | 9831.18 | 0.686885 | 0.120245 | 5.712373 | 1.11E-08 | 1.04E-07 | 1.609804 |
| TUBD1 | 314.5805 | 0.686934 | 0.135412 | 5.072899 | 3.92E-07 | 2.78E-06 | 1.609858 |
| RTKN2 | 934.1995 | 0.686961 | 0.093019 | 7.385143 | 1.52E-13 | 2.89E-12 | 1.609889 |
| TMTC2 | 264.8524 | 0.688305 | 0.194783 | 3.533701 | 0.00041 | 0.001555 | 1.611389 |
| FOS | 163.1114 | 0.688333 | 0.254077 | 2.709151 | 0.006746 | 0.01849 | 1.61142 |
| NBPF9 | 370.9908 | 0.688613 | 0.112092 | 6.14331 | 8.08E-10 | 9.10E-09 | 1.611733 |
| SPICE1 | 434.8852 | 0.688622 | 0.14792 | 4.655373 | 3.23E-06 | 1.92E-05 | 1.611744 |
| PLCD3 | 841.9523 | 0.688713 | 0.202773 | 3.396475 | 0.000683 | 0.002449 | 1.611845 |
| CCDC174 | 399.1634 | 0.690043 | 0.230821 | 2.989522 | 0.002794 | 0.008551 | 1.613332 |
| ANGPTL2 | 415.3775 | 0.69018 | 0.216761 | 3.184055 | 0.001452 | 0.004785 | 1.613485 |
| ZNF143 | 504.493 | 0.690203 | 0.200621 | 3.440338 | 0.000581 | 0.002125 | 1.613511 |
| ZMAT3 | 446.761 | 0.690455 | 0.126026 | 5.478662 | 4.29E-08 | 3.67E-07 | 1.613793 |
| MAGI3 | 339.2666 | 0.691213 | 0.14697 | 4.703083 | 2.56E-06 | 1.56E-05 | 1.614641 |
| FAM161A | 789.8299 | 0.691246 | 0.106339 | 6.5004 | 8.01E-11 | 1.07E-09 | 1.614678 |
| MRVI1 | 229.5007 | 0.691851 | 0.158282 | 4.371006 | 1.24E-05 | 6.54E-05 | 1.615355 |
| LPAR4 | 263.1942 | 0.692603 | 0.202859 | 3.414202 | 0.00064 | 0.002313 | 1.616197 |
| MOB1A | 2437.125 | 0.69264 | 0.086387 | 8.017828 | 1.08E-15 | 2.64E-14 | 1.616238 |
| RIMS1 | 60.1195 | 0.693373 | 0.286915 | 2.416649 | 0.015664 | 0.038401 | 1.617059 |
| CCDC113 | 570.7075 | 0.693393 | 0.137827 | 5.030875 | 4.88E-07 | 3.41E-06 | 1.617082 |
| CAPN2 | 1584.758 | 0.693567 | 0.117579 | 5.898741 | 3.66E-09 | 3.69E-08 | 1.617277 |
| DHFR2 | 167.9224 | 0.693668 | 0.173588 | 3.996047 | 6.44E-05 | 0.000295 | 1.61739 |
| PGBD2 | 118.7749 | 0.69369 | 0.185866 | 3.732197 | 0.00019 | 0.000776 | 1.617415 |
| CBX2 | 3741.963 | 0.693881 | 0.071861 | 9.655865 | 4.64E-22 | 2.17E-20 | 1.61763 |
| AC010186 | 53.18602 | 0.694068 | 0.289331 | 2.398869 | 0.016446 | 0.04008 | 1.617839 |
| MAPK8 | 1392.043 | 0.694192 | 0.111451 | 6.228654 | 4.70E-10 | 5.51E-09 | 1.617978 |
| PDE12 | 494.202 | 0.694253 | 0.092666 | 7.491969 | 6.78E-14 | 1.35E-12 | 1.618047 |
| BTRC | 1002.876 | 0.69433 | 0.113313 | 6.127526 | 8.93E-10 | 9.95E-09 | 1.618133 |
| IQCD | 61.57211 | 0.694357 | 0.236865 | 2.93145 | 0.003374 | 0.010074 | 1.618163 |
| KIF20B | 1707.264 | 0.694783 | 0.091325 | 7.607803 | 2.79E-14 | 5.80E-13 | 1.618641 |
| RTL6 | 1275.323 | 0.694886 | 0.082458 | 8.427108 | 3.54E-17 | 1.06E-15 | 1.618756 |
| ZSCAN9 | 255.9537 | 0.695133 | 0.14935 | 4.654398 | 3.25E-06 | 1.93E-05 | 1.619034 |
| MSANTD3- | 826.1974 | 0.695414 | 0.228093 | 3.04882 | 0.002297 | 0.007181 | 1.61935 |
| PCNA | 4236.448 | 0.695606 | 0.094144 | 7.388717 | 1.48E-13 | 2.82E-12 | 1.619565 |
| TIA1 | 2548.999 | 0.69621 | 0.097221 | 7.161128 | 8.00E-13 | 1.40E-11 | 1.620242 |
| ABL2 | 1076.583 | 0.696608 | 0.106781 | 6.523686 | 6.86E-11 | 9.24E-10 | 1.62069 |
| PANK3 | 3681.19 | 0.69685 | 0.103692 | 6.720396 | 1.81E-11 | 2.63E-10 | 1.620962 |
| ATPSCKMT | 181.5323 | 0.696975 | 0.212464 | 3.280443 | 0.001036 | 0.003549 | 1.621102 |

| | | | | | | | |
|---|---|---|---|---|---|---|---|
| MAD2L1 | 2394.393 | 0.697455 | 0.08092 | 8.619118 | 6.75E-18 | 2.16E-16 | 1.621641 |
| NR1D2 | 963.9316 | 0.697719 | 0.084665 | 8.240978 | 1.71E-16 | 4.70E-15 | 1.621938 |
| AP1M1 | 943.7738 | 0.697996 | 0.109009 | 6.40309 | 1.52E-10 | 1.94E-09 | 1.622249 |
| PLK4 | 805.219 | 0.69808 | 0.142204 | 4.909009 | 9.15E-07 | 6.08E-06 | 1.622344 |
| BRIP1 | 956.4612 | 0.698081 | 0.103125 | 6.769286 | 1.29E-11 | 1.92E-10 | 1.622346 |
| LDAH | 687.406 | 0.698477 | 0.113834 | 6.135926 | 8.47E-10 | 9.48E-09 | 1.62279 |
| BCLAF1 | 5024.985 | 0.699264 | 0.08619 | 8.113036 | 4.94E-16 | 1.28E-14 | 1.623676 |
| NCOA2 | 1464.081 | 0.699472 | 0.10776 | 6.491042 | 8.52E-11 | 1.13E-09 | 1.623911 |
| FAM72C | 262.5301 | 0.699585 | 0.19988 | 3.500021 | 0.000465 | 0.00174 | 1.624038 |
| KDM5A | 1219.455 | 0.699615 | 0.129728 | 5.392935 | 6.93E-08 | 5.69E-07 | 1.624072 |
| C3orf14 | 171.8343 | 0.700339 | 0.181202 | 3.864957 | 0.000111 | 0.000482 | 1.624887 |
| LRP12 | 500.0649 | 0.700659 | 0.131908 | 5.311734 | 1.09E-07 | 8.62E-07 | 1.625247 |
| RNF157 | 670.2192 | 0.70076 | 0.146436 | 4.785447 | 1.71E-06 | 1.08E-05 | 1.625361 |
| CCNF | 1614.598 | 0.700826 | 0.066872 | 10.48012 | 1.07E-25 | 7.18E-24 | 1.625435 |
| KLF12 | 501.1514 | 0.700834 | 0.177984 | 3.937628 | 8.23E-05 | 0.000368 | 1.625444 |
| NIPSNAP3/ | 211.5298 | 0.701168 | 0.148635 | 4.717387 | 2.39E-06 | 1.46E-05 | 1.62582 |
| ZNF407 | 235.195 | 0.701571 | 0.146103 | 4.801882 | 1.57E-06 | 9.98E-06 | 1.626275 |
| UHRF1 | 2098.909 | 0.702105 | 0.104619 | 6.711077 | 1.93E-11 | 2.79E-10 | 1.626877 |
| GNAI1 | 541.2205 | 0.702487 | 0.153273 | 4.583248 | 4.58E-06 | 2.65E-05 | 1.627307 |
| UBE3D | 248.2735 | 0.702754 | 0.157293 | 4.467813 | 7.90E-06 | 4.37E-05 | 1.627609 |
| WASHC3 | 117.9594 | 0.702756 | 0.24183 | 2.905992 | 0.003661 | 0.010807 | 1.627611 |
| AAGAB | 1207.376 | 0.703083 | 0.121882 | 5.76854 | 8.00E-09 | 7.67E-08 | 1.62798 |
| GK5 | 491.5932 | 0.703358 | 0.19339 | 3.636997 | 0.000276 | 0.001087 | 1.628291 |
| ZNF432 | 221.7148 | 0.703371 | 0.188724 | 3.726985 | 0.000194 | 0.000791 | 1.628305 |
| EFCAB11 | 287.6786 | 0.703469 | 0.176173 | 3.993046 | 6.52E-05 | 0.000298 | 1.628415 |
| DCAF16 | 1521.98 | 0.703579 | 0.088124 | 7.984008 | 1.42E-15 | 3.44E-14 | 1.62854 |
| IKZF2 | 156.7305 | 0.703583 | 0.302228 | 2.327984 | 0.019913 | 0.047199 | 1.628544 |
| APC2 | 633.7068 | 0.70384 | 0.196308 | 3.585379 | 0.000337 | 0.001303 | 1.628834 |
| ZBED4 | 1634.408 | 0.704475 | 0.072754 | 9.683037 | 3.56E-22 | 1.67E-20 | 1.629552 |
| DNAJC24 | 533.0308 | 0.704528 | 0.159239 | 4.424338 | 9.67E-06 | 5.25E-05 | 1.629611 |
| KIF3B | 1053.479 | 0.70474 | 0.077997 | 9.03543 | 1.63E-19 | 6.07E-18 | 1.629851 |
| COPG2 | 2538.089 | 0.70517 | 0.091852 | 7.677264 | 1.63E-14 | 3.49E-13 | 1.630337 |
| ETV5 | 1021.938 | 0.705361 | 0.257641 | 2.737771 | 0.006186 | 0.017181 | 1.630553 |
| TUBB4B | 5581.135 | 0.705438 | 0.138458 | 5.094961 | 3.49E-07 | 2.49E-06 | 1.63064 |
| TMCC1 | 784.3483 | 0.705808 | 0.150147 | 4.700782 | 2.59E-06 | 1.57E-05 | 1.631058 |
| DENND5B | 627.8675 | 0.705992 | 0.122199 | 5.777406 | 7.59E-09 | 7.31E-08 | 1.631266 |
| CRIPT | 243.561 | 0.706125 | 0.136576 | 5.170206 | 2.34E-07 | 1.74E-06 | 1.631416 |
| SMC1A | 4800.536 | 0.706244 | 0.090936 | 7.76641 | 8.07E-15 | 1.79E-13 | 1.631551 |
| EPB41 | 4265.962 | 0.706552 | 0.062306 | 11.34003 | 8.31E-30 | 7.53E-28 | 1.6319 |
| CENPA | 341.5508 | 0.706564 | 0.130457 | 5.416088 | 6.09E-08 | 5.06E-07 | 1.631913 |
| AL049834. | 206.8619 | 0.70683 | 0.145433 | 4.860188 | 1.17E-06 | 7.62E-06 | 1.632214 |
| AC090114 | 56.32672 | 0.707949 | 0.279158 | 2.53601 | 0.011212 | 0.028738 | 1.63348 |
| RAD51B | 96.41888 | 0.708005 | 0.221661 | 3.194097 | 0.001403 | 0.004633 | 1.633544 |
| NETO2 | 697.4835 | 0.708086 | 0.1912 | 3.703371 | 0.000213 | 0.00086 | 1.633635 |

| | | | | | | | |
|---|---|---|---|---|---|---|---|
| CABLES2 | 474.3928 | 0.708403 | 0.092552 | 7.654083 | 1.95E-14 | 4.14E-13 | 1.633995 |
| SP3 | 3081.772 | 0.709097 | 0.136691 | 5.1876 | 2.13E-07 | 1.59E-06 | 1.63478 |
| ZSCAN12 | 348.7796 | 0.709244 | 0.157235 | 4.510724 | 6.46E-06 | 3.63E-05 | 1.634947 |
| CALCOCO | 472.191 | 0.709556 | 0.141386 | 5.018568 | 5.21E-07 | 3.61E-06 | 1.635301 |
| DCAF7 | 3335.616 | 0.709716 | 0.066987 | 10.5948 | 3.15E-26 | 2.25E-24 | 1.635482 |
| SESN1 | 645.8695 | 0.709981 | 0.099646 | 7.125046 | 1.04E-12 | 1.80E-11 | 1.635782 |
| C12orf29 | 301.5476 | 0.710084 | 0.148089 | 4.794978 | 1.63E-06 | 1.03E-05 | 1.635899 |
| VEZF1 | 1863.29 | 0.710359 | 0.106956 | 6.641573 | 3.10E-11 | 4.36E-10 | 1.636211 |
| KDM3B | 4196.216 | 0.711054 | 0.065202 | 10.90543 | 1.09E-27 | 8.40E-26 | 1.636999 |
| SWT1 | 225.3476 | 0.711774 | 0.150215 | 4.738376 | 2.15E-06 | 1.33E-05 | 1.637817 |
| GNB4 | 1902.416 | 0.712082 | 0.114264 | 6.231905 | 4.61E-10 | 5.41E-09 | 1.638166 |
| CLASP2 | 2069.721 | 0.712266 | 0.112145 | 6.351278 | 2.14E-10 | 2.66E-09 | 1.638375 |
| BCAP29 | 639.1114 | 0.712631 | 0.128253 | 5.556438 | 2.75E-08 | 2.43E-07 | 1.63879 |
| FRG1CP | 264.2758 | 0.712765 | 0.137664 | 5.177582 | 2.25E-07 | 1.68E-06 | 1.638942 |
| SALL1 | 4806.142 | 0.712825 | 0.095801 | 7.44071 | 1.00E-13 | 1.96E-12 | 1.63901 |
| MED13L | 2113.707 | 0.712852 | 0.070283 | 10.14265 | 3.57E-24 | 2.10E-22 | 1.639041 |
| GLCCI1 | 660.1484 | 0.713406 | 0.121903 | 5.852226 | 4.85E-09 | 4.80E-08 | 1.639671 |
| FRYL | 900.2935 | 0.713519 | 0.077249 | 9.236666 | 2.54E-20 | 1.02E-18 | 1.639799 |
| TNPO1 | 7856.197 | 0.713521 | 0.119414 | 5.975182 | 2.30E-09 | 2.39E-08 | 1.639801 |
| CCDC102I | 38.16175 | 0.713604 | 0.309735 | 2.303915 | 0.021227 | 0.049909 | 1.639896 |
| BTBD3 | 2127.217 | 0.713884 | 0.115378 | 6.187343 | 6.12E-10 | 7.02E-09 | 1.640214 |
| MAPK11 | 44.21528 | 0.716276 | 0.304439 | 2.352774 | 0.018634 | 0.044615 | 1.642935 |
| C1orf198 | 1964.483 | 0.717023 | 0.06897 | 10.39609 | 2.58E-25 | 1.70E-23 | 1.643786 |
| AMOTL1 | 1337.633 | 0.717806 | 0.083687 | 8.577282 | 9.71E-18 | 3.07E-16 | 1.644679 |
| INTS2 | 1075.945 | 0.718043 | 0.085121 | 8.435541 | 3.30E-17 | 9.90E-16 | 1.644949 |
| EFNB2 | 990.8936 | 0.718102 | 0.099291 | 7.232335 | 4.75E-13 | 8.49E-12 | 1.645017 |
| GSK3B | 1876.301 | 0.718133 | 0.086688 | 8.284076 | 1.19E-16 | 3.35E-15 | 1.645052 |
| RPL36A | 7211.166 | 0.71831 | 0.09029 | 7.955596 | 1.78E-15 | 4.26E-14 | 1.645253 |
| WDR3 | 1596.276 | 0.718861 | 0.182737 | 3.933864 | 8.36E-05 | 0.000373 | 1.645883 |
| OSBPL11 | 1045.398 | 0.718997 | 0.07624 | 9.43065 | 4.08E-21 | 1.75E-19 | 1.646037 |
| DHX40 | 1503.648 | 0.719371 | 0.095947 | 7.497563 | 6.50E-14 | 1.30E-12 | 1.646465 |
| DRAXIN | 1417.066 | 0.720067 | 0.183888 | 3.915781 | 9.01E-05 | 0.000399 | 1.647258 |
| EPS8 | 226.9511 | 0.72055 | 0.164316 | 4.385151 | 1.16E-05 | 6.17E-05 | 1.647811 |
| HNRNPR | 5877.508 | 0.720562 | 0.057065 | 12.62712 | 1.50E-36 | 2.25E-34 | 1.647823 |
| MED17 | 674.758 | 0.72068 | 0.091316 | 7.892124 | 2.97E-15 | 6.96E-14 | 1.647958 |
| KDM3A | 1184.051 | 0.721178 | 0.186999 | 3.856588 | 0.000115 | 0.000496 | 1.648528 |
| CCSER2 | 605.2828 | 0.721448 | 0.216026 | 3.339639 | 0.000839 | 0.002941 | 1.648836 |
| RBL1 | 594.5328 | 0.721875 | 0.129281 | 5.583765 | 2.35E-08 | 2.09E-07 | 1.649324 |
| PDCL | 586.586 | 0.722031 | 0.105277 | 6.8584 | 6.96E-12 | 1.07E-10 | 1.649502 |
| NTN1 | 187.9134 | 0.722163 | 0.220758 | 3.271284 | 0.001071 | 0.00365 | 1.649654 |
| FAM98B | 1796.777 | 0.722222 | 0.114156 | 6.326642 | 2.51E-10 | 3.09E-09 | 1.649721 |
| SKP2 | 2163.575 | 0.72228 | 0.065975 | 10.94773 | 6.81E-28 | 5.41E-26 | 1.649787 |
| STIL | 1017.713 | 0.722893 | 0.099197 | 7.287463 | 3.16E-13 | 5.78E-12 | 1.650488 |
| SHLD1 | 61.18562 | 0.723454 | 0.294067 | 2.460165 | 0.013887 | 0.034608 | 1.65113 |

| | | | | | | | |
|---|---|---|---|---|---|---|---|
| ARRDC4 | 1071.649 | 0.723614 | 0.219097 | 3.302706 | 0.000958 | 0.003306 | 1.651314 |
| ZNF136 | 204.3024 | 0.724151 | 0.147892 | 4.896488 | 9.76E-07 | 6.42E-06 | 1.651928 |
| DCK | 630.199 | 0.724467 | 0.095654 | 7.573811 | 3.62E-14 | 7.45E-13 | 1.65229 |
| TMPO | 6717.087 | 0.724509 | 0.073373 | 9.874357 | 5.38E-23 | 2.78E-21 | 1.652339 |
| PRKACB | 1001.101 | 0.724973 | 0.116987 | 6.197053 | 5.75E-10 | 6.64E-09 | 1.652869 |
| C14orf132 | 1197.08 | 0.725188 | 0.135919 | 5.335429 | 9.53E-08 | 7.67E-07 | 1.653116 |
| GPR153 | 819.1632 | 0.725287 | 0.133337 | 5.439513 | 5.34E-08 | 4.49E-07 | 1.65323 |
| PHC2 | 1194.599 | 0.725813 | 0.095208 | 7.623413 | 2.47E-14 | 5.18E-13 | 1.653833 |
| PPP1CB | 4816.653 | 0.726182 | 0.101167 | 7.178019 | 7.07E-13 | 1.24E-11 | 1.654255 |
| FLNC | 1402.497 | 0.726768 | 0.121579 | 5.977739 | 2.26E-09 | 2.36E-08 | 1.654927 |
| MIER3 | 1113.704 | 0.727024 | 0.080587 | 9.021579 | 1.85E-19 | 6.80E-18 | 1.655221 |
| TTC30B | 189.4568 | 0.727051 | 0.2081 | 3.493748 | 0.000476 | 0.001777 | 1.655252 |
| SLC26A2 | 728.1558 | 0.727189 | 0.146241 | 4.972534 | 6.61E-07 | 4.49E-06 | 1.65541 |
| OSBPL8 | 2411.654 | 0.727545 | 0.091566 | 7.945601 | 1.93E-15 | 4.60E-14 | 1.655819 |
| KMT2E | 1624.866 | 0.727829 | 0.099824 | 7.291104 | 3.07E-13 | 5.63E-12 | 1.656145 |
| TTC12 | 223.9661 | 0.728436 | 0.180208 | 4.042197 | 5.30E-05 | 0.000247 | 1.656842 |
| RO60 | 1102.68 | 0.728643 | 0.15437 | 4.720111 | 2.36E-06 | 1.45E-05 | 1.65708 |
| SNN | 1845.829 | 0.728649 | 0.100677 | 7.237477 | 4.57E-13 | 8.19E-12 | 1.657087 |
| MAP1B | 7779.937 | 0.728808 | 0.090108 | 8.088201 | 6.06E-16 | 1.54E-14 | 1.65727 |
| HOMER1 | 1239.09 | 0.729024 | 0.158985 | 4.585485 | 4.53E-06 | 2.63E-05 | 1.657517 |
| SPATA6 | 437.4682 | 0.729501 | 0.178614 | 4.084237 | 4.42E-05 | 0.00021 | 1.658065 |
| CEP76 | 400.1601 | 0.72987 | 0.151638 | 4.813246 | 1.48E-06 | 9.48E-06 | 1.65849 |
| EPM2AIP1 | 477.7642 | 0.730077 | 0.230538 | 3.166835 | 0.001541 | 0.00505 | 1.658728 |
| CRBN | 593.862 | 0.730309 | 0.14438 | 5.058227 | 4.23E-07 | 2.98E-06 | 1.658994 |
| ZNF484 | 238.2011 | 0.730658 | 0.177544 | 4.115358 | 3.87E-05 | 0.000186 | 1.659396 |
| POLK | 337.3835 | 0.73087 | 0.172429 | 4.238664 | 2.25E-05 | 0.000114 | 1.65964 |
| CASP6 | 482.3099 | 0.730957 | 0.171758 | 4.25574 | 2.08E-05 | 0.000106 | 1.659739 |
| EIF1AD | 442.2404 | 0.731029 | 0.120431 | 6.070116 | 1.28E-09 | 1.39E-08 | 1.659823 |
| WDR36 | 1283.397 | 0.731046 | 0.077089 | 9.483188 | 2.47E-21 | 1.07E-19 | 1.659842 |
| AP3M2 | 994.0553 | 0.731059 | 0.091708 | 7.971603 | 1.57E-15 | 3.78E-14 | 1.659857 |
| RAB8B | 1331.304 | 0.731542 | 0.112161 | 6.522273 | 6.92E-11 | 9.32E-10 | 1.660413 |
| WNK3 | 875.259 | 0.731644 | 0.073847 | 9.907593 | 3.86E-23 | 2.02E-21 | 1.660531 |
| EED | 666.7644 | 0.731824 | 0.091846 | 7.96792 | 1.61E-15 | 3.89E-14 | 1.660737 |
| AMOT | 729.5944 | 0.731951 | 0.207354 | 3.529951 | 0.000416 | 0.001575 | 1.660883 |
| ZNF117 | 560.1167 | 0.732532 | 0.154108 | 4.753358 | 2.00E-06 | 1.24E-05 | 1.661552 |
| CCNA2 | 3260.636 | 0.733322 | 0.077992 | 9.402491 | 5.33E-21 | 2.26E-19 | 1.662462 |
| MEIOC | 225.2202 | 0.734531 | 0.213952 | 3.433152 | 0.000597 | 0.002175 | 1.663857 |
| EXOC6B | 1588.11 | 0.73462 | 0.1176 | 6.246761 | 4.19E-10 | 4.97E-09 | 1.663959 |
| MAP1A | 291.922 | 0.734896 | 0.131101 | 5.605556 | 2.08E-08 | 1.86E-07 | 1.664277 |
| ZFX | 699.2691 | 0.734944 | 0.125662 | 5.848571 | 4.96E-09 | 4.89E-08 | 1.664333 |
| LRRC49 | 380.0911 | 0.735337 | 0.138279 | 5.317764 | 1.05E-07 | 8.37E-07 | 1.664787 |
| GAREM1 | 271.9746 | 0.735797 | 0.176389 | 4.171439 | 3.03E-05 | 0.000149 | 1.665318 |
| TCEAL7 | 526.5943 | 0.736248 | 0.098601 | 7.466976 | 8.21E-14 | 1.62E-12 | 1.665838 |
| PIK3C2B | 882.3452 | 0.736828 | 0.122374 | 6.021129 | 1.73E-09 | 1.85E-08 | 1.666507 |

| | | | | | | | |
|---|---|---|---|---|---|---|---|
| MED4 | 968.3579 | 0.736855 | 0.091572 | 8.046721 | 8.50E-16 | 2.12E-14 | 1.666539 |
| DAB1 | 338.1902 | 0.737416 | 0.129424 | 5.697673 | 1.21E-08 | 1.13E-07 | 1.667187 |
| SGO2 | 1043.536 | 0.73798 | 0.131467 | 5.613406 | 1.98E-08 | 1.79E-07 | 1.667838 |
| CHEK1 | 2495.333 | 0.738439 | 0.097773 | 7.552562 | 4.27E-14 | 8.69E-13 | 1.66837 |
| DENND2A | 497.7356 | 0.738527 | 0.136201 | 5.422344 | 5.88E-08 | 4.90E-07 | 1.668472 |
| GABPA | 1027.096 | 0.738967 | 0.105541 | 7.001702 | 2.53E-12 | 4.16E-11 | 1.668981 |
| PPP1R17 | 53.12118 | 0.739109 | 0.271464 | 2.722677 | 0.006476 | 0.017862 | 1.669144 |
| WDR70 | 840.5033 | 0.739193 | 0.15222 | 4.856079 | 1.20E-06 | 7.78E-06 | 1.669241 |
| CAND1 | 5877.679 | 0.739445 | 0.070585 | 10.476 | 1.11E-25 | 7.47E-24 | 1.669534 |
| MGA | 2235.848 | 0.739534 | 0.107381 | 6.887037 | 5.70E-12 | 8.89E-11 | 1.669637 |
| RALGPS2 | 1740.219 | 0.73965 | 0.070854 | 10.43905 | 1.64E-25 | 1.10E-23 | 1.66977 |
| SMC2 | 1621.266 | 0.74026 | 0.079469 | 9.315088 | 1.22E-20 | 5.06E-19 | 1.670477 |
| ZC4H2 | 1049.189 | 0.740439 | 0.176184 | 4.202652 | 2.64E-05 | 0.000132 | 1.670684 |
| TRDMT1 | 588.1941 | 0.741377 | 0.141501 | 5.239383 | 1.61E-07 | 1.23E-06 | 1.671771 |
| LXN | 95.79464 | 0.741426 | 0.216263 | 3.428346 | 0.000607 | 0.002209 | 1.671828 |
| NADK2 | 655.1878 | 0.741604 | 0.092279 | 8.036537 | 9.24E-16 | 2.29E-14 | 1.672033 |
| PIKFYVE | 1558.609 | 0.742514 | 0.096263 | 7.71341 | 1.22E-14 | 2.66E-13 | 1.673089 |
| NSD2 | 6130.462 | 0.742758 | 0.100791 | 7.369291 | 1.72E-13 | 3.24E-12 | 1.673372 |
| AC087632 | 381.5676 | 0.743086 | 0.1417 | 5.244089 | 1.57E-07 | 1.20E-06 | 1.673752 |
| SMARCA1 | 4937.624 | 0.743829 | 0.071259 | 10.43844 | 1.66E-25 | 1.10E-23 | 1.674614 |
| PARP11 | 320.9069 | 0.744199 | 0.147937 | 5.030506 | 4.89E-07 | 3.41E-06 | 1.675044 |
| CCDC77 | 636.2999 | 0.744954 | 0.102226 | 7.287333 | 3.16E-13 | 5.78E-12 | 1.675921 |
| USP30 | 271.7819 | 0.745552 | 0.143212 | 5.20594 | 1.93E-07 | 1.45E-06 | 1.676615 |
| FAM110B | 104.3793 | 0.746329 | 0.287185 | 2.598772 | 0.009356 | 0.024589 | 1.677518 |
| CSTF2T | 1505.602 | 0.746357 | 0.074845 | 9.972065 | 2.02E-23 | 1.08E-21 | 1.677552 |
| FIGNL2 | 1024.252 | 0.748067 | 0.105841 | 7.067858 | 1.57E-12 | 2.66E-11 | 1.679541 |
| PPM1D | 477.4587 | 0.748798 | 0.159591 | 4.691989 | 2.71E-06 | 1.64E-05 | 1.680392 |
| STK4 | 1512.663 | 0.749004 | 0.080758 | 9.274673 | 1.78E-20 | 7.28E-19 | 1.680632 |
| ARHGAP5 | 2221.69 | 0.749252 | 0.114745 | 6.529739 | 6.59E-11 | 8.90E-10 | 1.680921 |
| FBXO16 | 113.9919 | 0.74986 | 0.197442 | 3.79788 | 0.000146 | 0.000613 | 1.68163 |
| BRD8 | 1569.042 | 0.749898 | 0.118152 | 6.346894 | 2.20E-10 | 2.73E-09 | 1.681674 |
| GNAZ | 541.2691 | 0.750962 | 0.118328 | 6.346433 | 2.20E-10 | 2.73E-09 | 1.682915 |
| ZNF319 | 228.6696 | 0.751796 | 0.15775 | 4.765755 | 1.88E-06 | 1.17E-05 | 1.683888 |
| HP1BP3 | 3022.811 | 0.752012 | 0.09127 | 8.239458 | 1.73E-16 | 4.75E-15 | 1.68414 |
| CKS2 | 1740.49 | 0.752115 | 0.087539 | 8.59179 | 8.56E-18 | 2.72E-16 | 1.68426 |
| FAM114A2 | 415.06 | 0.752609 | 0.120115 | 6.265742 | 3.71E-10 | 4.45E-09 | 1.684837 |
| RASA2 | 966.298 | 0.752668 | 0.110752 | 6.795998 | 1.08E-11 | 1.62E-10 | 1.684906 |
| HMGB1P6 | 1727.714 | 0.752751 | 0.084911 | 8.865148 | 7.64E-19 | 2.66E-17 | 1.685003 |
| MPHOSPH | 1053.999 | 0.752928 | 0.137328 | 5.482687 | 4.19E-08 | 3.59E-07 | 1.685209 |
| MED31 | 55.46317 | 0.753568 | 0.313107 | 2.406744 | 0.016095 | 0.039332 | 1.685957 |
| ZFP37 | 207.6588 | 0.753568 | 0.180025 | 4.185913 | 2.84E-05 | 0.00014 | 1.685957 |
| MPP7 | 297.0417 | 0.754306 | 0.176131 | 4.282628 | 1.85E-05 | 9.47E-05 | 1.68682 |
| CARD8 | 518.2622 | 0.7547 | 0.109768 | 6.875399 | 6.18E-12 | 9.60E-11 | 1.687281 |
| NBN | 1355.996 | 0.754957 | 0.10708 | 7.050419 | 1.78E-12 | 2.99E-11 | 1.687581 |

| | | | | | | | |
|---|---|---|---|---|---|---|---|
| EFHD1 | 126.2824 | 0.75527 | 0.244396 | 3.090354 | 0.001999 | 0.006361 | 1.687947 |
| CCDC181 | 164.9415 | 0.755462 | 0.198095 | 3.813637 | 0.000137 | 0.000579 | 1.688172 |
| RGS12 | 499.6106 | 0.75595 | 0.190183 | 3.974863 | 7.04E-05 | 0.000319 | 1.688743 |
| PLEKHM3 | 409.9708 | 0.756114 | 0.183956 | 4.110303 | 3.95E-05 | 0.00019 | 1.688935 |
| PAPOLG | 813.7592 | 0.756363 | 0.15274 | 4.951968 | 7.35E-07 | 4.97E-06 | 1.689227 |
| WDR35 | 1198.493 | 0.75648 | 0.135594 | 5.578992 | 2.42E-08 | 2.15E-07 | 1.689363 |
| THAP12 | 1598.437 | 0.756521 | 0.085692 | 8.828413 | 1.06E-18 | 3.65E-17 | 1.689412 |
| BACH1 | 1072.65 | 0.756647 | 0.086095 | 8.788495 | 1.52E-18 | 5.13E-17 | 1.68956 |
| IPO5 | 18889.83 | 0.756814 | 0.055603 | 13.6111 | 3.44E-42 | 7.90E-40 | 1.689755 |
| CALM2 | 9418.813 | 0.75741 | 0.065042 | 11.64488 | 2.44E-31 | 2.65E-29 | 1.690454 |
| MIS18BP1 | 1810.26 | 0.758335 | 0.092118 | 8.232183 | 1.84E-16 | 5.03E-15 | 1.691538 |
| TAOK1 | 3654.503 | 0.758896 | 0.077965 | 9.733853 | 2.16E-22 | 1.03E-20 | 1.692195 |
| BORA | 462.6919 | 0.759041 | 0.116794 | 6.498988 | 8.09E-11 | 1.08E-09 | 1.692365 |
| RIPK4 | 172.0596 | 0.759543 | 0.167999 | 4.521122 | 6.15E-06 | 3.48E-05 | 1.692954 |
| ZNF512 | 1149.334 | 0.760019 | 0.118301 | 6.424456 | 1.32E-10 | 1.70E-09 | 1.693513 |
| APAF1 | 787.2872 | 0.760297 | 0.092415 | 8.227025 | 1.92E-16 | 5.22E-15 | 1.69384 |
| NFYA | 916.6165 | 0.760406 | 0.101988 | 7.455869 | 8.93E-14 | 1.76E-12 | 1.693967 |
| RAD54L2 | 1559.199 | 0.760692 | 0.094426 | 8.05592 | 7.89E-16 | 1.97E-14 | 1.694303 |
| ATF2 | 1136.952 | 0.760729 | 0.085887 | 8.857378 | 8.19E-19 | 2.85E-17 | 1.694347 |
| KLHL24 | 842.2899 | 0.760795 | 0.167496 | 4.542173 | 5.57E-06 | 3.17E-05 | 1.694424 |
| TTC33 | 459.8687 | 0.760989 | 0.175498 | 4.33617 | 1.45E-05 | 7.55E-05 | 1.694652 |
| PACRGL | 262.6516 | 0.76164 | 0.151557 | 5.025439 | 5.02E-07 | 3.49E-06 | 1.695417 |
| FERMT2 | 5748.636 | 0.761999 | 0.084448 | 9.023277 | 1.83E-19 | 6.72E-18 | 1.695838 |
| FAM83D | 1260.889 | 0.762519 | 0.079174 | 9.630971 | 5.92E-22 | 2.73E-20 | 1.69645 |
| BNIP3L | 2096.658 | 0.762548 | 0.08729 | 8.735828 | 2.42E-18 | 8.04E-17 | 1.696484 |
| FANCC | 556.1209 | 0.762669 | 0.176963 | 4.309769 | 1.63E-05 | 8.44E-05 | 1.696626 |
| ITPRID2 | 340.7335 | 0.763543 | 0.136661 | 5.58715 | 2.31E-08 | 2.06E-07 | 1.697655 |
| MSL2 | 1210.598 | 0.76359 | 0.167107 | 4.569479 | 4.89E-06 | 2.82E-05 | 1.69771 |
| ARMC2 | 47.98679 | 0.765827 | 0.306595 | 2.497846 | 0.012495 | 0.031556 | 1.700345 |
| DOP1B | 449.1538 | 0.765842 | 0.145901 | 5.249054 | 1.53E-07 | 1.18E-06 | 1.700362 |
| TPX2 | 4289.066 | 0.766898 | 0.080495 | 9.527287 | 1.61E-21 | 7.15E-20 | 1.701607 |
| LNPK | 446.9193 | 0.767146 | 0.113134 | 6.780837 | 1.19E-11 | 1.78E-10 | 1.7019 |
| EFHC1 | 652.6838 | 0.767279 | 0.209345 | 3.665145 | 0.000247 | 0.000983 | 1.702057 |
| PCLAF | 1381.343 | 0.767478 | 0.131639 | 5.83017 | 5.54E-09 | 5.42E-08 | 1.702292 |
| VSNL1 | 786.142 | 0.767527 | 0.168586 | 4.552724 | 5.30E-06 | 3.02E-05 | 1.70235 |
| ASAP2 | 868.5205 | 0.767857 | 0.143864 | 5.33738 | 9.43E-08 | 7.59E-07 | 1.702739 |
| GEMIN4 | 1479.111 | 0.76792 | 0.106907 | 7.183067 | 6.82E-13 | 1.20E-11 | 1.702813 |
| IFT81 | 631.3577 | 0.768003 | 0.127825 | 6.008263 | 1.88E-09 | 1.99E-08 | 1.702911 |
| LPIN2 | 1070.617 | 0.768326 | 0.084401 | 9.103311 | 8.76E-20 | 3.34E-18 | 1.703292 |
| KIF3A | 784.4255 | 0.768344 | 0.083184 | 9.23666 | 2.54E-20 | 1.02E-18 | 1.703313 |
| ZBTB12 | 545.4365 | 0.768797 | 0.153785 | 4.99916 | 5.76E-07 | 3.96E-06 | 1.703848 |
| GUSBP2 | 61.10832 | 0.768905 | 0.333467 | 2.305793 | 0.021122 | 0.049693 | 1.703976 |
| FBXO10 | 1081.396 | 0.769503 | 0.117678 | 6.539064 | 6.19E-11 | 8.42E-10 | 1.704682 |
| UBE2C | 1964.642 | 0.770706 | 0.094372 | 8.16672 | 3.17E-16 | 8.31E-15 | 1.706105 |

| | | | | | | | |
|---|---|---|---|---|---|---|---|
| SIN3A | 3434.315 | 0.77228 | 0.061113 | 12.63688 | 1.32E-36 | 2.01E-34 | 1.707966 |
| PAQR3 | 1239.174 | 0.772418 | 0.090781 | 8.50855 | 1.76E-17 | 5.46E-16 | 1.70813 |
| SULF1 | 276.9916 | 0.772848 | 0.326783 | 2.365021 | 0.018029 | 0.043377 | 1.708639 |
| FANCI | 3547.41 | 0.773119 | 0.089054 | 8.681438 | 3.91E-18 | 1.28E-16 | 1.70896 |
| SUV39H2 | 802.1765 | 0.773394 | 0.143399 | 5.393297 | 6.92E-08 | 5.68E-07 | 1.709286 |
| PDCD4 | 764.4413 | 0.774345 | 0.160302 | 4.830541 | 1.36E-06 | 8.73E-06 | 1.710414 |
| CETN2 | 581.4898 | 0.775214 | 0.093382 | 8.301573 | 1.03E-16 | 2.91E-15 | 1.711444 |
| RFX7 | 1263.023 | 0.775513 | 0.085127 | 9.110075 | 8.23E-20 | 3.15E-18 | 1.711798 |
| PRTFDC1 | 273.2029 | 0.775908 | 0.207328 | 3.742422 | 0.000182 | 0.000746 | 1.712267 |
| FUT8 | 1076.799 | 0.775997 | 0.142434 | 5.448112 | 5.09E-08 | 4.29E-07 | 1.712373 |
| COIL | 568.9072 | 0.776403 | 0.097777 | 7.940584 | 2.01E-15 | 4.78E-14 | 1.712855 |
| ZBTB26 | 325.2065 | 0.776477 | 0.163229 | 4.75698 | 1.97E-06 | 1.22E-05 | 1.712943 |
| ALDH1B1 | 2269.3 | 0.778702 | 0.079572 | 9.786094 | 1.29E-22 | 6.33E-21 | 1.715586 |
| DCAF12L2 | 52.90022 | 0.779885 | 0.31985 | 2.438283 | 0.014757 | 0.03642 | 1.716994 |
| ZNF25 | 47.31388 | 0.779976 | 0.313736 | 2.486087 | 0.012916 | 0.032468 | 1.717102 |
| CCDC30 | 103.62 | 0.780247 | 0.295977 | 2.636175 | 0.008385 | 0.022316 | 1.717424 |
| AC242843 | 88.33315 | 0.781554 | 0.289783 | 2.697037 | 0.006996 | 0.019083 | 1.718982 |
| NCOA3 | 1093.327 | 0.781808 | 0.094229 | 8.296907 | 1.07E-16 | 3.02E-15 | 1.719284 |
| EYA3 | 764.3163 | 0.782422 | 0.114482 | 6.834461 | 8.23E-12 | 1.26E-10 | 1.720016 |
| ARHGEF39 | 509.8153 | 0.782968 | 0.135086 | 5.79609 | 6.79E-09 | 6.58E-08 | 1.720667 |
| RASSF10 | 57.58706 | 0.784012 | 0.336113 | 2.332584 | 0.01967 | 0.046689 | 1.721913 |
| PCDHGB7 | 842.0116 | 0.784165 | 0.102428 | 7.655776 | 1.92E-14 | 4.10E-13 | 1.722095 |
| RBM12 | 3562.794 | 0.785937 | 0.061597 | 12.75931 | 2.77E-37 | 4.46E-35 | 1.724212 |
| DHFRP1 | 213.0804 | 0.786485 | 0.212109 | 3.70792 | 0.000209 | 0.000846 | 1.724866 |
| ANO2 | 76.86848 | 0.787508 | 0.280646 | 2.806059 | 0.005015 | 0.014302 | 1.726091 |
| CTNNA2 | 365.9291 | 0.788114 | 0.150654 | 5.231301 | 1.68E-07 | 1.28E-06 | 1.726816 |
| GTSE1 | 1141.556 | 0.788288 | 0.087044 | 9.056185 | 1.35E-19 | 5.06E-18 | 1.727024 |
| RNASEL | 59.43909 | 0.789899 | 0.249683 | 3.163605 | 0.001558 | 0.005101 | 1.728953 |
| NFIC | 143.7474 | 0.790505 | 0.212753 | 3.715606 | 0.000203 | 0.000823 | 1.72968 |
| RFK | 382.4598 | 0.790905 | 0.126682 | 6.243239 | 4.29E-10 | 5.07E-09 | 1.730159 |
| NREP | 4313.119 | 0.791165 | 0.098244 | 8.053093 | 8.07E-16 | 2.02E-14 | 1.730471 |
| VPS35 | 2567.187 | 0.791934 | 0.113647 | 6.96836 | 3.21E-12 | 5.21E-11 | 1.731394 |
| SLC35E2A | 687.1724 | 0.792424 | 0.140431 | 5.642807 | 1.67E-08 | 1.53E-07 | 1.731982 |
| TMEM106C | 462.6826 | 0.792977 | 0.156673 | 5.061338 | 4.16E-07 | 2.94E-06 | 1.732646 |
| SSH1 | 1099.097 | 0.793174 | 0.12655 | 6.267666 | 3.66E-10 | 4.40E-09 | 1.732883 |
| HIPK2 | 1722.719 | 0.793224 | 0.136186 | 5.824581 | 5.73E-09 | 5.59E-08 | 1.732942 |
| FBXO5 | 889.5289 | 0.793806 | 0.081174 | 9.779086 | 1.38E-22 | 6.75E-21 | 1.733642 |
| CCNY | 1796.422 | 0.793967 | 0.166841 | 4.758813 | 1.95E-06 | 1.21E-05 | 1.733836 |
| FRG1BP | 86.12343 | 0.794532 | 0.265088 | 2.99724 | 0.002724 | 0.008362 | 1.734515 |
| AC068547 | 130.9106 | 0.794621 | 0.216195 | 3.675492 | 0.000237 | 0.000949 | 1.734622 |
| ST3GAL6 | 163.474 | 0.794658 | 0.189752 | 4.187874 | 2.82E-05 | 0.000139 | 1.734666 |
| ERV3-1 | 158.5465 | 0.794955 | 0.319873 | 2.485223 | 0.012947 | 0.032538 | 1.735024 |
| GGCT | 1387.363 | 0.795994 | 0.108259 | 7.352692 | 1.94E-13 | 3.65E-12 | 1.736274 |
| RNF169 | 447.5365 | 0.797416 | 0.139814 | 5.703392 | 1.17E-08 | 1.10E-07 | 1.737985 |

| | | | | | | | |
|---|---|---|---|---|---|---|---|
| FBXO4 | 33.68558 | 0.797483 | 0.341771 | 2.333386 | 0.019628 | 0.046613 | 1.738066 |
| PAFAH2 | 412.8701 | 0.797521 | 0.135831 | 5.871434 | 4.32E-09 | 4.31E-08 | 1.738112 |
| RNF175 | 222.1207 | 0.798477 | 0.173096 | 4.612923 | 3.97E-06 | 2.33E-05 | 1.739264 |
| ZNF887P | 43.51449 | 0.799081 | 0.311257 | 2.567272 | 0.01025 | 0.026575 | 1.739993 |
| PRDM10 | 465.5661 | 0.799111 | 0.135399 | 5.901884 | 3.59E-09 | 3.63E-08 | 1.740029 |
| CCNB1 | 3615.042 | 0.799974 | 0.081741 | 9.786745 | 1.28E-22 | 6.31E-21 | 1.741069 |
| SMAD1 | 585.9899 | 0.800431 | 0.114029 | 7.019545 | 2.23E-12 | 3.70E-11 | 1.741622 |
| TTC30A | 268.2423 | 0.80091 | 0.152242 | 5.260766 | 1.43E-07 | 1.11E-06 | 1.7422 |
| ZNF214 | 131.8278 | 0.800914 | 0.280688 | 2.853401 | 0.004325 | 0.012525 | 1.742205 |
| PLCH1 | 1024.034 | 0.801256 | 0.103703 | 7.726455 | 1.11E-14 | 2.41E-13 | 1.742618 |
| TTF2 | 2354.775 | 0.801334 | 0.106302 | 7.538308 | 4.76E-14 | 9.64E-13 | 1.742712 |
| AC138866 | 100.6106 | 0.801438 | 0.230501 | 3.476947 | 0.000507 | 0.00188 | 1.742838 |
| NUTM2A | 49.02836 | 0.801598 | 0.270885 | 2.959187 | 0.003085 | 0.009323 | 1.743031 |
| CENPL | 257.2724 | 0.801723 | 0.14853 | 5.397727 | 6.75E-08 | 5.56E-07 | 1.743182 |
| MAML3 | 266.4875 | 0.801744 | 0.201815 | 3.972664 | 7.11E-05 | 0.000322 | 1.743207 |
| AC092835 | 76.58162 | 0.802348 | 0.230813 | 3.476178 | 0.000509 | 0.001884 | 1.743937 |
| SNTB2 | 623.9708 | 0.802728 | 0.17104 | 4.693212 | 2.69E-06 | 1.63E-05 | 1.744397 |
| FCHSD2 | 428.5512 | 0.802769 | 0.153424 | 5.23237 | 1.67E-07 | 1.28E-06 | 1.744446 |
| BABAM2 | 665.107 | 0.803213 | 0.129514 | 6.201756 | 5.58E-10 | 6.47E-09 | 1.744983 |
| SPRY2 | 893.8755 | 0.803451 | 0.196215 | 4.094751 | 4.23E-05 | 0.000202 | 1.745271 |
| LRRC20 | 653.9745 | 0.803702 | 0.099415 | 8.084329 | 6.25E-16 | 1.59E-14 | 1.745575 |
| CEP290 | 529.8513 | 0.804233 | 0.147097 | 5.46736 | 4.57E-08 | 3.88E-07 | 1.746217 |
| LZTFL1 | 321.0575 | 0.804651 | 0.226842 | 3.547189 | 0.000389 | 0.001485 | 1.746723 |
| TENM3 | 2740.557 | 0.805851 | 0.089223 | 9.031859 | 1.69E-19 | 6.24E-18 | 1.748177 |
| SMAD4 | 2639.637 | 0.80616 | 0.081318 | 9.913684 | 3.63E-23 | 1.91E-21 | 1.748551 |
| KAT6A | 1314.29 | 0.806406 | 0.09203 | 8.762472 | 1.91E-18 | 6.37E-17 | 1.748849 |
| TRANK1 | 670.6384 | 0.807759 | 0.161818 | 4.991761 | 5.98E-07 | 4.10E-06 | 1.75049 |
| TYMS | 2171.688 | 0.807795 | 0.078772 | 10.25489 | 1.12E-24 | 7.06E-23 | 1.750534 |
| ZBTB6 | 274.079 | 0.807851 | 0.13783 | 5.861203 | 4.60E-09 | 4.56E-08 | 1.750602 |
| WDR48 | 1840.278 | 0.807932 | 0.076502 | 10.56093 | 4.52E-26 | 3.11E-24 | 1.7507 |
| FBXO3 | 829.3659 | 0.808488 | 0.158853 | 5.089527 | 3.59E-07 | 2.56E-06 | 1.751376 |
| TCF12 | 2886.925 | 0.808646 | 0.133539 | 6.055525 | 1.40E-09 | 1.51E-08 | 1.751567 |
| CANX | 1298.955 | 0.808775 | 0.244593 | 3.30662 | 0.000944 | 0.003265 | 1.751724 |
| NOVA1 | 301.4731 | 0.809105 | 0.169013 | 4.787221 | 1.69E-06 | 1.07E-05 | 1.752124 |
| CRB2 | 969.3128 | 0.809334 | 0.144515 | 5.600343 | 2.14E-08 | 1.92E-07 | 1.752402 |
| PRDM2 | 661.9947 | 0.810124 | 0.110424 | 7.336494 | 2.19E-13 | 4.09E-12 | 1.753363 |
| FEZ1 | 2248.619 | 0.811144 | 0.111935 | 7.246539 | 4.28E-13 | 7.69E-12 | 1.754603 |
| UFC1 | 972.874 | 0.812201 | 0.07184 | 11.30573 | 1.23E-29 | 1.10E-27 | 1.755889 |
| IRX5 | 155.2648 | 0.812302 | 0.292742 | 2.774809 | 0.005523 | 0.015541 | 1.756012 |
| OSBPL6 | 566.5702 | 0.812564 | 0.117483 | 6.916442 | 4.63E-12 | 7.35E-11 | 1.75633 |
| ECT2 | 2860.573 | 0.81268 | 0.070956 | 11.45326 | 2.26E-30 | 2.19E-28 | 1.756472 |
| CASP8AP2 | 1198.678 | 0.813006 | 0.097032 | 8.378777 | 5.35E-17 | 1.57E-15 | 1.756869 |
| KATNAL1 | 411.8564 | 0.813338 | 0.170806 | 4.761763 | 1.92E-06 | 1.20E-05 | 1.757273 |
| GPRIN1 | 344.6296 | 0.81457 | 0.170606 | 4.774571 | 1.80E-06 | 1.13E-05 | 1.758773 |

| | | | | | | | |
|---|---|---|---|---|---|---|---|
| RP2 | 355.3484 | 0.814698 | 0.120894 | 6.738941 | 1.60E-11 | 2.33E-10 | 1.75893 |
| HSF2 | 635.9605 | 0.814982 | 0.169212 | 4.816332 | 1.46E-06 | 9.34E-06 | 1.759277 |
| TOX3 | 721.1716 | 0.815132 | 0.159714 | 5.103713 | 3.33E-07 | 2.40E-06 | 1.759459 |
| PLCL2 | 996.0367 | 0.815222 | 0.098252 | 8.297219 | 1.07E-16 | 3.02E-15 | 1.759569 |
| SH3PXD2E | 3272.985 | 0.815542 | 0.067261 | 12.12499 | 7.79E-34 | 1.03E-31 | 1.759959 |
| CDCA3 | 571.506 | 0.816886 | 0.121253 | 6.737011 | 1.62E-11 | 2.36E-10 | 1.761599 |
| RTF1 | 2511.92 | 0.817169 | 0.088329 | 9.251423 | 2.22E-20 | 8.95E-19 | 1.761945 |
| CENPQ | 562.99 | 0.817693 | 0.10576 | 7.731585 | 1.06E-14 | 2.32E-13 | 1.762585 |
| GEM | 86.69061 | 0.81797 | 0.254265 | 3.216999 | 0.001295 | 0.004318 | 1.762924 |
| STON1 | 699.7094 | 0.818849 | 0.105902 | 7.732121 | 1.06E-14 | 2.32E-13 | 1.763998 |
| CBX1 | 4274.326 | 0.81887 | 0.08314 | 9.849318 | 6.90E-23 | 3.52E-21 | 1.764023 |
| GAN | 202.0245 | 0.819019 | 0.151445 | 5.408017 | 6.37E-08 | 5.27E-07 | 1.764206 |
| GAS2L3 | 420.316 | 0.819053 | 0.166606 | 4.916121 | 8.83E-07 | 5.88E-06 | 1.764248 |
| SETBP1 | 366.0244 | 0.819459 | 0.258102 | 3.174943 | 0.001499 | 0.004926 | 1.764744 |
| CREBRF | 187.661 | 0.821043 | 0.255636 | 3.211768 | 0.001319 | 0.004387 | 1.766683 |
| FAM168A | 1012.754 | 0.821202 | 0.116636 | 7.040721 | 1.91E-12 | 3.20E-11 | 1.766878 |
| PATL1 | 1110.395 | 0.821233 | 0.093106 | 8.820396 | 1.14E-18 | 3.90E-17 | 1.766915 |
| TGFBR3 | 185.4484 | 0.822101 | 0.203984 | 4.030223 | 5.57E-05 | 0.000259 | 1.767979 |
| RSF1 | 2181.039 | 0.82301 | 0.100037 | 8.227017 | 1.92E-16 | 5.22E-15 | 1.769093 |
| DCLRE1A | 719.5514 | 0.823087 | 0.11082 | 7.427239 | 1.11E-13 | 2.15E-12 | 1.769187 |
| LINC0068( | 67.35246 | 0.824907 | 0.310682 | 2.655153 | 0.007927 | 0.021241 | 1.771421 |
| NIN | 759.4408 | 0.825167 | 0.193206 | 4.270918 | 1.95E-05 | 9.95E-05 | 1.771741 |
| HEATR6 | 733.0047 | 0.825921 | 0.154017 | 5.362535 | 8.21E-08 | 6.68E-07 | 1.772666 |
| CAMK1G | 64.57548 | 0.82611 | 0.29583 | 2.792512 | 0.00523 | 0.01483 | 1.772899 |
| RBM23 | 1183.251 | 0.826991 | 0.116248 | 7.114042 | 1.13E-12 | 1.94E-11 | 1.773982 |
| ZBTB37 | 620.6784 | 0.827502 | 0.148212 | 5.583224 | 2.36E-08 | 2.10E-07 | 1.77461 |
| CLSPN | 1235.283 | 0.828707 | 0.129868 | 6.381151 | 1.76E-10 | 2.22E-09 | 1.776093 |
| PEG10 | 5206.018 | 0.828866 | 0.090519 | 9.156838 | 5.34E-20 | 2.09E-18 | 1.776289 |
| CTNND2 | 640.3714 | 0.829036 | 0.113062 | 7.332588 | 2.26E-13 | 4.20E-12 | 1.776498 |
| ARRDC3 | 513.6501 | 0.830123 | 0.20346 | 4.080039 | 4.50E-05 | 0.000214 | 1.777837 |
| FOXO3B | 542.7542 | 0.830585 | 0.187704 | 4.424977 | 9.65E-06 | 5.24E-05 | 1.778406 |
| BCL9 | 2166.815 | 0.830756 | 0.080109 | 10.37027 | 3.39E-25 | 2.21E-23 | 1.778617 |
| APBA2 | 194.6408 | 0.831416 | 0.162235 | 5.124777 | 2.98E-07 | 2.17E-06 | 1.779431 |
| PLEKHA8 | 1806.311 | 0.831549 | 0.096343 | 8.631124 | 6.08E-18 | 1.95E-16 | 1.779595 |
| COL4A6 | 2017.392 | 0.831646 | 0.108093 | 7.693816 | 1.43E-14 | 3.08E-13 | 1.779715 |
| EMSY | 852.5435 | 0.832901 | 0.126523 | 6.583024 | 4.61E-11 | 6.36E-10 | 1.781263 |
| PKN2 | 1267.965 | 0.833368 | 0.133196 | 6.256704 | 3.93E-10 | 4.68E-09 | 1.781841 |
| PLCXD2 | 103.0034 | 0.83362 | 0.181501 | 4.59291 | 4.37E-06 | 2.54E-05 | 1.782151 |
| KPNA2 | 10025.77 | 0.833703 | 0.077154 | 10.80566 | 3.24E-27 | 2.42E-25 | 1.782254 |
| CAP2 | 477.9283 | 0.834375 | 0.107453 | 7.764993 | 8.16E-15 | 1.81E-13 | 1.783085 |
| MIB1 | 3214.437 | 0.834521 | 0.082963 | 10.05894 | 8.39E-24 | 4.66E-22 | 1.783265 |
| GNE | 479.0596 | 0.834785 | 0.136666 | 6.108225 | 1.01E-09 | 1.11E-08 | 1.783591 |
| FHL1 | 2380.104 | 0.834967 | 0.085736 | 9.738799 | 2.06E-22 | 9.91E-21 | 1.783816 |
| LRRC6 | 82.22954 | 0.835725 | 0.353688 | 2.362885 | 0.018133 | 0.043611 | 1.784753 |

| | | | | | | | |
|---|---|---|---|---|---|---|---|
| PIBF1 | 281.6081 | 0.836386 | 0.203054 | 4.119035 | 3.80E-05 | 0.000184 | 1.785571 |
| PPP2R3A | 620.6594 | 0.837192 | 0.149251 | 5.609287 | 2.03E-08 | 1.83E-07 | 1.786569 |
| CYB5R4 | 353.7144 | 0.837246 | 0.252807 | 3.311799 | 0.000927 | 0.003215 | 1.786636 |
| COLGALT2 | 244.6878 | 0.838043 | 0.179313 | 4.673626 | 2.96E-06 | 1.77E-05 | 1.787624 |
| EXOC5 | 3006.118 | 0.838495 | 0.121741 | 6.887545 | 5.68E-12 | 8.86E-11 | 1.788183 |
| TRIM17 | 42.76576 | 0.839111 | 0.314109 | 2.671402 | 0.007554 | 0.020369 | 1.788947 |
| PUM2 | 3517.288 | 0.840028 | 0.085394 | 9.837134 | 7.79E-23 | 3.93E-21 | 1.790085 |
| LIN52 | 368.8393 | 0.840257 | 0.129669 | 6.48 | 9.17E-11 | 1.21E-09 | 1.790369 |
| CBX5 | 11048.61 | 0.840734 | 0.082658 | 10.17119 | 2.67E-24 | 1.61E-22 | 1.790961 |
| STK17B | 216.8738 | 0.840848 | 0.22332 | 3.765214 | 0.000166 | 0.000688 | 1.791103 |
| CDON | 884.0377 | 0.84207 | 0.111636 | 7.542972 | 4.59E-14 | 9.31E-13 | 1.79262 |
| CDC27 | 1448.376 | 0.843346 | 0.171947 | 4.90468 | 9.36E-07 | 6.20E-06 | 1.794207 |
| ANKRD50 | 1359.537 | 0.84354 | 0.102492 | 8.230271 | 1.87E-16 | 5.10E-15 | 1.794448 |
| TP53I11 | 5400.442 | 0.844328 | 0.106929 | 7.896165 | 2.88E-15 | 6.75E-14 | 1.795428 |
| ARHGEF26 | 337.4824 | 0.845598 | 0.116478 | 7.259734 | 3.88E-13 | 7.01E-12 | 1.79701 |
| POLD3 | 1264.937 | 0.846466 | 0.084611 | 10.00417 | 1.46E-23 | 7.88E-22 | 1.798091 |
| HJURP | 1392.007 | 0.847224 | 0.097157 | 8.720127 | 2.78E-18 | 9.19E-17 | 1.799037 |
| BBS12 | 133.9697 | 0.847416 | 0.210929 | 4.017547 | 5.88E-05 | 0.000272 | 1.799275 |
| CEP55 | 576.6496 | 0.847728 | 0.093097 | 9.1059 | 8.56E-20 | 3.27E-18 | 1.799665 |
| LAMTOR3 | 622.9617 | 0.849129 | 0.103337 | 8.217113 | 2.08E-16 | 5.63E-15 | 1.801413 |
| DMRTA1 | 230.6915 | 0.850582 | 0.188483 | 4.512767 | 6.40E-06 | 3.60E-05 | 1.803228 |
| CPNE4 | 91.56083 | 0.852177 | 0.307921 | 2.767517 | 0.005649 | 0.015866 | 1.805223 |
| TCAF1 | 2131.31 | 0.853226 | 0.123026 | 6.93534 | 4.05E-12 | 6.48E-11 | 1.806536 |
| SPRY1 | 519.7821 | 0.853282 | 0.132053 | 6.461673 | 1.04E-10 | 1.35E-09 | 1.806606 |
| KIAA0232 | 1629.636 | 0.853913 | 0.080319 | 10.63149 | 2.13E-26 | 1.55E-24 | 1.807396 |
| ZNF658 | 177.5336 | 0.853969 | 0.179596 | 4.754947 | 1.98E-06 | 1.23E-05 | 1.807467 |
| ULK2 | 482.189 | 0.855038 | 0.105587 | 8.097929 | 5.59E-16 | 1.43E-14 | 1.808806 |
| UBTD2 | 1329.261 | 0.855247 | 0.078122 | 10.94754 | 6.83E-28 | 5.41E-26 | 1.809069 |
| ST8SIA2 | 124.417 | 0.856948 | 0.234667 | 3.65176 | 0.00026 | 0.001032 | 1.811203 |
| STAT1 | 1891.631 | 0.857531 | 0.073208 | 11.71366 | 1.08E-31 | 1.21E-29 | 1.811934 |
| SEMA5A | 626.9368 | 0.85765 | 0.089494 | 9.583335 | 9.40E-22 | 4.26E-20 | 1.812084 |
| SCHIP1 | 303.2069 | 0.858753 | 0.318171 | 2.699034 | 0.006954 | 0.018983 | 1.813471 |
| SNIP1 | 396.0456 | 0.859857 | 0.139207 | 6.176822 | 6.54E-10 | 7.46E-09 | 1.814858 |
| ERCC6L | 800.3024 | 0.859868 | 0.081155 | 10.59533 | 3.13E-26 | 2.25E-24 | 1.814872 |
| C15orf41 | 323.2363 | 0.860347 | 0.133473 | 6.445845 | 1.15E-10 | 1.49E-09 | 1.815475 |
| SPATA18 | 115.4444 | 0.862491 | 0.289894 | 2.975198 | 0.002928 | 0.008899 | 1.818175 |
| SOGA1 | 2401.023 | 0.863209 | 0.06825 | 12.64781 | 1.15E-36 | 1.76E-34 | 1.81908 |
| G2E3 | 1732.217 | 0.864029 | 0.106685 | 8.098881 | 5.55E-16 | 1.42E-14 | 1.820114 |
| BZW1P2 | 287.0366 | 0.865358 | 0.275367 | 3.142561 | 0.001675 | 0.00544 | 1.821792 |
| KIAA1958 | 1111.453 | 0.865446 | 0.115944 | 7.464324 | 8.37E-14 | 1.65E-12 | 1.821903 |
| C4orf46 | 1288.224 | 0.866878 | 0.094183 | 9.204146 | 3.44E-20 | 1.37E-18 | 1.823712 |
| DAB2 | 101.5815 | 0.866895 | 0.222393 | 3.898032 | 9.70E-05 | 0.000427 | 1.823734 |
| TIGD6 | 80.97201 | 0.867281 | 0.280515 | 3.091742 | 0.00199 | 0.006336 | 1.824222 |
| ZNF583 | 79.42155 | 0.868194 | 0.359745 | 2.413362 | 0.015806 | 0.038687 | 1.825377 |

| | | | | | | | |
|---|---|---|---|---|---|---|---|
| FPGT | 217.7349 | 0.868261 | 0.282119 | 3.077647 | 0.002086 | 0.006601 | 1.825462 |
| BCL2L11 | 638.2265 | 0.868858 | 0.157232 | 5.525959 | 3.28E-08 | 2.86E-07 | 1.826217 |
| AKAP13 | 1641.924 | 0.869338 | 0.084202 | 10.32444 | 5.46E-25 | 3.49E-23 | 1.826824 |
| SMAD6 | 176.5272 | 0.869641 | 0.233529 | 3.723907 | 0.000196 | 0.000799 | 1.827208 |
| C1QTNF6 | 216.3352 | 0.869905 | 0.248747 | 3.497146 | 0.00047 | 0.001756 | 1.827543 |
| C2orf72 | 69.42889 | 0.87029 | 0.3436 | 2.53286 | 0.011314 | 0.028975 | 1.828031 |
| POLR2J4 | 72.56254 | 0.870395 | 0.335452 | 2.594691 | 0.009468 | 0.024835 | 1.828163 |
| KBTBD7 | 645.7763 | 0.870535 | 0.086685 | 10.04246 | 9.92E-24 | 5.44E-22 | 1.82834 |
| RPS6KA5 | 163.3373 | 0.870787 | 0.184023 | 4.731936 | 2.22E-06 | 1.37E-05 | 1.82866 |
| ZYG11B | 1059.349 | 0.871632 | 0.1251 | 6.967478 | 3.23E-12 | 5.23E-11 | 1.829731 |
| RRM1 | 2983.292 | 0.871684 | 0.161306 | 5.403916 | 6.52E-08 | 5.39E-07 | 1.829798 |
| C6orf141 | 504.2002 | 0.871764 | 0.244554 | 3.564706 | 0.000364 | 0.001401 | 1.829899 |
| NCAPH | 1588.447 | 0.871909 | 0.068901 | 12.65459 | 1.06E-36 | 1.63E-34 | 1.830084 |
| LCOR | 1130.68 | 0.872684 | 0.114833 | 7.599574 | 2.97E-14 | 6.16E-13 | 1.831066 |
| SLAIN2 | 919.9439 | 0.873431 | 0.239754 | 3.643022 | 0.000269 | 0.001064 | 1.832014 |
| BCL7A | 677.1483 | 0.87413 | 0.100437 | 8.70326 | 3.22E-18 | 1.06E-16 | 1.832903 |
| METTL14 | 812.8678 | 0.874626 | 0.172023 | 5.084352 | 3.69E-07 | 2.63E-06 | 1.833533 |
| RASGRP1 | 181.7643 | 0.875027 | 0.203288 | 4.304366 | 1.67E-05 | 8.64E-05 | 1.834042 |
| STARD4 | 875.3619 | 0.875768 | 0.108891 | 8.042585 | 8.80E-16 | 2.19E-14 | 1.834985 |
| ELOVL2 | 83.24177 | 0.876815 | 0.216482 | 4.050282 | 5.12E-05 | 0.00024 | 1.836317 |
| AC008012 | 336.4707 | 0.877369 | 0.181159 | 4.843082 | 1.28E-06 | 8.26E-06 | 1.837022 |
| CENPE | 1666.156 | 0.877422 | 0.145262 | 6.040272 | 1.54E-09 | 1.65E-08 | 1.837089 |
| CDK14 | 571.7439 | 0.877663 | 0.152313 | 5.762226 | 8.30E-09 | 7.92E-08 | 1.837397 |
| RALGDS | 697.7425 | 0.878538 | 0.190908 | 4.601889 | 4.19E-06 | 2.44E-05 | 1.838512 |
| SP4 | 656.3444 | 0.879265 | 0.122488 | 7.178352 | 7.06E-13 | 1.24E-11 | 1.839438 |
| PEX2 | 540.8234 | 0.879599 | 0.171348 | 5.133409 | 2.85E-07 | 2.08E-06 | 1.839863 |
| RFX3 | 643.2513 | 0.880231 | 0.101505 | 8.671797 | 4.25E-18 | 1.38E-16 | 1.84067 |
| FAM222A | 43.66042 | 0.880647 | 0.332102 | 2.651735 | 0.008008 | 0.02143 | 1.841201 |
| MVB12B | 535.4254 | 0.88076 | 0.11369 | 7.747021 | 9.41E-15 | 2.08E-13 | 1.841345 |
| BTBD9 | 192.3044 | 0.88118 | 0.208428 | 4.227742 | 2.36E-05 | 0.000119 | 1.841882 |
| DDHD2 | 763.4689 | 0.882748 | 0.162548 | 5.430698 | 5.61E-08 | 4.70E-07 | 1.843884 |
| CDCA2 | 1187.697 | 0.883173 | 0.084728 | 10.42364 | 1.93E-25 | 1.28E-23 | 1.844427 |
| C21orf91 | 730.9333 | 0.883243 | 0.12361 | 7.145372 | 8.98E-13 | 1.56E-11 | 1.844517 |
| UBE2T | 799.0852 | 0.886459 | 0.087208 | 10.16493 | 2.84E-24 | 1.70E-22 | 1.848633 |
| DNAJC6 | 205.8671 | 0.887336 | 0.161006 | 5.511202 | 3.56E-08 | 3.09E-07 | 1.849758 |
| RBBP4 | 6215.155 | 0.888892 | 0.057347 | 15.50012 | 3.46E-54 | 1.77E-51 | 1.851753 |
| CDCA8 | 1596.866 | 0.889162 | 0.075048 | 11.84784 | 2.21E-32 | 2.57E-30 | 1.8521 |
| PM20D2 | 660.2413 | 0.889751 | 0.088632 | 10.03867 | 1.03E-23 | 5.62E-22 | 1.852857 |
| CEP152 | 517.5906 | 0.890796 | 0.123528 | 7.211266 | 5.54E-13 | 9.83E-12 | 1.854199 |
| TMEM123 | 3660.784 | 0.890864 | 0.10654 | 8.361767 | 6.18E-17 | 1.81E-15 | 1.854286 |
| RPGRIP1L | 430.6276 | 0.890926 | 0.158096 | 5.63534 | 1.75E-08 | 1.59E-07 | 1.854366 |
| DIP2B | 1251.569 | 0.891176 | 0.098096 | 9.084731 | 1.04E-19 | 3.93E-18 | 1.854687 |
| KIFC1 | 1932.507 | 0.892519 | 0.088563 | 10.07776 | 6.93E-24 | 3.89E-22 | 1.856414 |
| RAPH1 | 704.6481 | 0.892614 | 0.168201 | 5.306822 | 1.12E-07 | 8.83E-07 | 1.856537 |

| | | | | | | | |
|---|---|---|---|---|---|---|---|
| CAPN6 | 604.415 | 0.894023 | 0.11021 | 8.112 | 4.98E-16 | 1.29E-14 | 1.85835 |
| DPYSL3 | 26451.04 | 0.894636 | 0.087388 | 10.23747 | 1.35E-24 | 8.36E-23 | 1.85914 |
| IGF2 | 88.04126 | 0.894872 | 0.31711 | 2.821958 | 0.004773 | 0.013681 | 1.859445 |
| RNGTT | 880.144 | 0.895903 | 0.088743 | 10.09553 | 5.78E-24 | 3.30E-22 | 1.860774 |
| TIPARP | 140.1912 | 0.896252 | 0.196686 | 4.556766 | 5.19E-06 | 2.97E-05 | 1.861224 |
| ANKRD36C | 81.75254 | 0.897363 | 0.279519 | 3.210384 | 0.001326 | 0.004403 | 1.862658 |
| ARNT2 | 1143.629 | 0.897486 | 0.098963 | 9.068932 | 1.20E-19 | 4.52E-18 | 1.862817 |
| MAP7D2 | 405.8038 | 0.898618 | 0.14526 | 6.186262 | 6.16E-10 | 7.06E-09 | 1.864279 |
| NRCAM | 463.3686 | 0.900081 | 0.22272 | 4.041321 | 5.32E-05 | 0.000248 | 1.866171 |
| KIF24 | 320.034 | 0.900862 | 0.121922 | 7.388827 | 1.48E-13 | 2.82E-12 | 1.867181 |
| PARVA | 522.5056 | 0.900999 | 0.168524 | 5.346414 | 8.97E-08 | 7.25E-07 | 1.867359 |
| AC010615 | 41.93615 | 0.901197 | 0.291647 | 3.090028 | 0.002001 | 0.006366 | 1.867615 |
| NCAPG | 2321.613 | 0.901862 | 0.100977 | 8.931408 | 4.21E-19 | 1.50E-17 | 1.868476 |
| HMGB2 | 7172.839 | 0.902356 | 0.057198 | 15.77592 | 4.56E-56 | 2.39E-53 | 1.869116 |
| FRMD4B | 123.0505 | 0.903305 | 0.20642 | 4.376062 | 1.21E-05 | 6.40E-05 | 1.870346 |
| C3orf70 | 96.753 | 0.906936 | 0.204891 | 4.426432 | 9.58E-06 | 5.21E-05 | 1.87506 |
| VIM | 7294.227 | 0.906962 | 0.139877 | 6.483992 | 8.93E-11 | 1.18E-09 | 1.875093 |
| H2BFM | 70.99081 | 0.908081 | 0.281402 | 3.226993 | 0.001251 | 0.004188 | 1.876548 |
| FAM217B | 608.891 | 0.90883 | 0.155231 | 5.854691 | 4.78E-09 | 4.73E-08 | 1.877523 |
| RPS3AP26 | 39.94969 | 0.90924 | 0.341288 | 2.66414 | 0.007719 | 0.020743 | 1.878055 |
| SORBS2 | 1054.557 | 0.91029 | 0.193372 | 4.707462 | 2.51E-06 | 1.53E-05 | 1.879423 |
| AC139769 | 114.4035 | 0.910472 | 0.180037 | 5.057148 | 4.26E-07 | 3.00E-06 | 1.879661 |
| ZNF20 | 72.17987 | 0.911706 | 0.3256 | 2.800079 | 0.005109 | 0.014533 | 1.881269 |
| PRICKLE2 | 316.2263 | 0.912837 | 0.222675 | 4.099414 | 4.14E-05 | 0.000198 | 1.882744 |
| TUBA1A | 10126.08 | 0.913395 | 0.109282 | 8.358139 | 6.37E-17 | 1.86E-15 | 1.883473 |
| ODC1 | 6346.363 | 0.913603 | 0.145936 | 6.260315 | 3.84E-10 | 4.60E-09 | 1.883744 |
| PTPRO | 50.26149 | 0.913694 | 0.298679 | 3.059114 | 0.00222 | 0.00697 | 1.883863 |
| VAT1L | 2796.194 | 0.915885 | 0.100803 | 9.085882 | 1.03E-19 | 3.90E-18 | 1.886726 |
| FAM161B | 55.51864 | 0.916758 | 0.33565 | 2.731293 | 0.006309 | 0.017478 | 1.887868 |
| RPS10-NU | 276.6093 | 0.917319 | 0.218818 | 4.192156 | 2.76E-05 | 0.000137 | 1.888602 |
| CNTNAP3E | 570.4824 | 0.917809 | 0.171247 | 5.359546 | 8.34E-08 | 6.78E-07 | 1.889244 |
| SMAD5 | 2030.086 | 0.917818 | 0.142618 | 6.43548 | 1.23E-10 | 1.59E-09 | 1.889255 |
| TNNI3 | 189.8508 | 0.920326 | 0.183308 | 5.020646 | 5.15E-07 | 3.57E-06 | 1.892543 |
| NEDD1 | 936.0022 | 0.921437 | 0.131338 | 7.015769 | 2.29E-12 | 3.79E-11 | 1.894001 |
| FDXACB1 | 94.69925 | 0.92204 | 0.236935 | 3.891533 | 9.96E-05 | 0.000438 | 1.894792 |
| BRMS1L | 296.1285 | 0.9221 | 0.240379 | 3.836029 | 0.000125 | 0.000533 | 1.894872 |
| ARHGAP11 | 2546.438 | 0.922479 | 0.092417 | 9.981683 | 1.83E-23 | 9.85E-22 | 1.895369 |
| CNTNAP3 | 669.6346 | 0.923916 | 0.095828 | 9.641449 | 5.34E-22 | 2.48E-20 | 1.897259 |
| EGR1 | 1582.099 | 0.924127 | 0.080708 | 11.4502 | 2.35E-30 | 2.26E-28 | 1.897536 |
| TFCP2 | 1277.118 | 0.927226 | 0.139694 | 6.637536 | 3.19E-11 | 4.46E-10 | 1.901616 |
| FBXO33 | 359.4759 | 0.927465 | 0.167386 | 5.540865 | 3.01E-08 | 2.64E-07 | 1.901931 |
| ROR2 | 712.6987 | 0.927693 | 0.140321 | 6.611212 | 3.81E-11 | 5.30E-10 | 1.902232 |
| AC079594 | 30.29276 | 0.927883 | 0.390236 | 2.37775 | 0.017419 | 0.042179 | 1.902483 |
| ZBED6 | 482.9282 | 0.929102 | 0.27603 | 3.365951 | 0.000763 | 0.002701 | 1.90409 |

| | | | | | | | |
|---|---|---|---|---|---|---|---|
| ZFP30 | 357.0899 | 0.929934 | 0.205251 | 4.530706 | 5.88E-06 | 3.33E-05 | 1.905189 |
| ZC3H12B | 221.6157 | 0.93254 | 0.181727 | 5.13153 | 2.87E-07 | 2.10E-06 | 1.908633 |
| PIK3R3 | 556.0956 | 0.932851 | 0.090656 | 10.29001 | 7.82E-25 | 4.95E-23 | 1.909044 |
| FRG1 | 302.1664 | 0.933936 | 0.168867 | 5.530597 | 3.19E-08 | 2.79E-07 | 1.910482 |
| ZNF350 | 92.74949 | 0.934097 | 0.24201 | 3.859737 | 0.000114 | 0.000491 | 1.910694 |
| SESN3 | 3718.826 | 0.934105 | 0.127221 | 7.3424 | 2.10E-13 | 3.93E-12 | 1.910705 |
| COMMD3 | 76.82922 | 0.936127 | 0.263505 | 3.552596 | 0.000381 | 0.00146 | 1.913385 |
| DGKK | 30.14598 | 0.936949 | 0.363517 | 2.57746 | 0.009953 | 0.025907 | 1.914476 |
| ZNF567 | 318.8119 | 0.937838 | 0.156967 | 5.974747 | 2.30E-09 | 2.40E-08 | 1.915656 |
| AC092329 | 94.43761 | 0.938247 | 0.28891 | 3.247539 | 0.001164 | 0.003931 | 1.916198 |
| KIAA1586 | 483.4818 | 0.938341 | 0.123356 | 7.60678 | 2.81E-14 | 5.84E-13 | 1.916324 |
| ASPH | 3463.348 | 0.938624 | 0.14413 | 6.512344 | 7.40E-11 | 9.93E-10 | 1.9167 |
| ZBTB33 | 1663.021 | 0.938757 | 0.067412 | 13.92559 | 4.43E-44 | 1.15E-41 | 1.916876 |
| KIF14 | 1127.65 | 0.94033 | 0.117296 | 8.016725 | 1.09E-15 | 2.66E-14 | 1.918967 |
| FZD5 | 4834.136 | 0.940391 | 0.191867 | 4.901274 | 9.52E-07 | 6.29E-06 | 1.919048 |
| ARHGAP32 | 1033.771 | 0.940996 | 0.089073 | 10.56428 | 4.36E-26 | 3.03E-24 | 1.919853 |
| DCUN1D4 | 1508.163 | 0.941893 | 0.115061 | 8.186034 | 2.70E-16 | 7.17E-15 | 1.921047 |
| LZIC | 672.5962 | 0.942473 | 0.184013 | 5.121763 | 3.03E-07 | 2.20E-06 | 1.92182 |
| SARM1 | 295.781 | 0.942674 | 0.132025 | 7.140106 | 9.33E-13 | 1.62E-11 | 1.922088 |
| NEIL3 | 276.938 | 0.943293 | 0.139003 | 6.786132 | 1.15E-11 | 1.72E-10 | 1.922913 |
| WASF3 | 1910.786 | 0.943397 | 0.066728 | 14.13786 | 2.22E-45 | 6.37E-43 | 1.923051 |
| ZNF850 | 242.6534 | 0.94448 | 0.258863 | 3.648574 | 0.000264 | 0.001043 | 1.924495 |
| KIF18A | 836.9979 | 0.944685 | 0.085157 | 11.09344 | 1.35E-28 | 1.13E-26 | 1.924769 |
| CAMK1 | 309.8729 | 0.9452 | 0.14424 | 6.552948 | 5.64E-11 | 7.71E-10 | 1.925456 |
| LANCL1 | 2313.317 | 0.945855 | 0.075106 | 12.59354 | 2.29E-36 | 3.37E-34 | 1.92633 |
| HMGA2 | 5532.224 | 0.94651 | 0.053012 | 17.85447 | 2.67E-71 | 3.27E-68 | 1.927204 |
| SMYD2 | 135.7326 | 0.946809 | 0.189877 | 4.986425 | 6.15E-07 | 4.21E-06 | 1.927604 |
| KIF11 | 2583.796 | 0.946983 | 0.070848 | 13.36639 | 9.50E-41 | 1.92E-38 | 1.927836 |
| EVI5 | 286.8935 | 0.947484 | 0.277544 | 3.413812 | 0.000641 | 0.002316 | 1.928507 |
| NRXN3 | 116.2045 | 0.947861 | 0.273373 | 3.467283 | 0.000526 | 0.00194 | 1.92901 |
| SF3A3 | 3349.704 | 0.94841 | 0.07991 | 11.86848 | 1.73E-32 | 2.03E-30 | 1.929744 |
| INCENP | 1565.898 | 0.94858 | 0.129049 | 7.350527 | 1.97E-13 | 3.71E-12 | 1.929972 |
| ANKRD12 | 693.3369 | 0.948916 | 0.245711 | 3.861914 | 0.000113 | 0.000487 | 1.930421 |
| ZNF699 | 61.55157 | 0.948991 | 0.332413 | 2.854858 | 0.004306 | 0.012482 | 1.930522 |
| NME5 | 40.51567 | 0.949017 | 0.321748 | 2.94956 | 0.003182 | 0.009575 | 1.930556 |
| CD82 | 194.8635 | 0.949155 | 0.22277 | 4.260684 | 2.04E-05 | 0.000104 | 1.930741 |
| SUFU | 722.6972 | 0.950219 | 0.110664 | 8.586515 | 8.96E-18 | 2.84E-16 | 1.932166 |
| SGO1 | 543.5828 | 0.951538 | 0.116356 | 8.177805 | 2.89E-16 | 7.62E-15 | 1.933933 |
| TP53INP1 | 401.6392 | 0.951773 | 0.152315 | 6.248713 | 4.14E-10 | 4.91E-09 | 1.934248 |
| IL17RD | 2399.177 | 0.951782 | 0.114615 | 8.304161 | 1.01E-16 | 2.86E-15 | 1.93426 |
| PUS7L | 891.9835 | 0.95189 | 0.121139 | 7.857815 | 3.91E-15 | 9.04E-14 | 1.934405 |
| CASP7 | 147.3812 | 0.952181 | 0.233387 | 4.079836 | 4.51E-05 | 0.000214 | 1.934795 |
| ZDBF2 | 1403.476 | 0.952264 | 0.074508 | 12.78071 | 2.10E-37 | 3.42E-35 | 1.934906 |
| PALLD | 1633.983 | 0.95311 | 0.122671 | 7.769636 | 7.87E-15 | 1.75E-13 | 1.936041 |

| | | | | | | | |
|---|---|---|---|---|---|---|---|
| ANKMY2 | 376.4515 | 0.953573 | 0.110046 | 8.665218 | 4.51E-18 | 1.46E-16 | 1.936663 |
| DEPDC1B | 1691.188 | 0.954412 | 0.068998 | 13.83255 | 1.62E-43 | 4.08E-41 | 1.937789 |
| BOC | 706.8388 | 0.954549 | 0.165327 | 5.773699 | 7.76E-09 | 7.47E-08 | 1.937974 |
| MAATS1 | 161.7994 | 0.955032 | 0.35246 | 2.709621 | 0.006736 | 0.018467 | 1.938623 |
| ARMC4 | 238.0089 | 0.95618 | 0.170987 | 5.592118 | 2.24E-08 | 2.00E-07 | 1.940165 |
| SRGAP2 | 1200.566 | 0.957293 | 0.12654 | 7.565147 | 3.87E-14 | 7.95E-13 | 1.941663 |
| ZBTB41 | 276.1105 | 0.957878 | 0.139408 | 6.871033 | 6.37E-12 | 9.87E-11 | 1.942451 |
| CAB39L | 184.8369 | 0.958365 | 0.159789 | 5.997676 | 2.00E-09 | 2.11E-08 | 1.943107 |
| RTL5 | 206.7817 | 0.959563 | 0.293872 | 3.265243 | 0.001094 | 0.003719 | 1.94472 |
| SHC3 | 194.6616 | 0.960269 | 0.271649 | 3.534966 | 0.000408 | 0.001548 | 1.945673 |
| CKAP2 | 2767.405 | 0.960344 | 0.076157 | 12.61011 | 1.86E-36 | 2.75E-34 | 1.945773 |
| DNAJB1 | 1603.621 | 0.963165 | 0.063433 | 15.18403 | 4.51E-52 | 2.02E-49 | 1.949582 |
| NCAM1 | 337.088 | 0.966616 | 0.163202 | 5.922837 | 3.16E-09 | 3.22E-08 | 1.954252 |
| CEP350 | 1536.68 | 0.969853 | 0.161603 | 6.001444 | 1.96E-09 | 2.06E-08 | 1.958642 |
| LIMCH1 | 1019.528 | 0.969998 | 0.203437 | 4.768058 | 1.86E-06 | 1.16E-05 | 1.958838 |
| ZBTB10 | 984.4135 | 0.972323 | 0.15551 | 6.252484 | 4.04E-10 | 4.80E-09 | 1.961997 |
| DRP2 | 66.27852 | 0.972633 | 0.280051 | 3.473063 | 0.000515 | 0.001904 | 1.96242 |
| BCAT1 | 12867.36 | 0.978091 | 0.088001 | 11.11458 | 1.07E-28 | 8.98E-27 | 1.969858 |
| NF1 | 1834.02 | 0.97907 | 0.17966 | 5.449582 | 5.05E-08 | 4.26E-07 | 1.971194 |
| TTK | 1448.252 | 0.980351 | 0.104399 | 9.390463 | 5.97E-21 | 2.52E-19 | 1.972946 |
| TBL1X | 3449.749 | 0.980443 | 0.084813 | 11.56011 | 6.56E-31 | 6.70E-29 | 1.973072 |
| ST8SIA6 | 68.87202 | 0.981809 | 0.310181 | 3.16528 | 0.001549 | 0.005074 | 1.97494 |
| GRM3 | 93.775 | 0.982012 | 0.25275 | 3.885312 | 0.000102 | 0.000448 | 1.975218 |
| TNC | 193.6113 | 0.982344 | 0.296414 | 3.314099 | 0.000919 | 0.003194 | 1.975673 |
| GPC3 | 6160.491 | 0.982859 | 0.19103 | 5.145056 | 2.67E-07 | 1.97E-06 | 1.976378 |
| ARHGAP28 | 576.7122 | 0.986646 | 0.100859 | 9.782404 | 1.34E-22 | 6.55E-21 | 1.981573 |
| POLH | 198.631 | 0.988549 | 0.18401 | 5.372259 | 7.78E-08 | 6.35E-07 | 1.984188 |
| PBRM1 | 2061.476 | 0.989777 | 0.098462 | 10.0524 | 8.97E-24 | 4.93E-22 | 1.985878 |
| IFT88 | 192.9443 | 0.989954 | 0.159619 | 6.201982 | 5.58E-10 | 6.47E-09 | 1.986122 |
| CDK1 | 2037.848 | 0.990008 | 0.103409 | 9.573662 | 1.03E-21 | 4.64E-20 | 1.986195 |
| NUSAP1 | 2268.752 | 0.990832 | 0.110141 | 8.996002 | 2.34E-19 | 8.49E-18 | 1.98733 |
| ZSWIM6 | 529.4578 | 0.991196 | 0.098732 | 10.03925 | 1.02E-23 | 5.60E-22 | 1.987832 |
| KIAA1211 | 738.9343 | 0.991529 | 0.143421 | 6.913397 | 4.73E-12 | 7.49E-11 | 1.988292 |
| CTDSPL2 | 2004.432 | 0.991994 | 0.112435 | 8.822812 | 1.12E-18 | 3.83E-17 | 1.988932 |
| FBXO43 | 66.80669 | 0.99219 | 0.283689 | 3.497463 | 0.00047 | 0.001755 | 1.989202 |
| HMGN2 | 11060.96 | 0.993548 | 0.097694 | 10.17001 | 2.70E-24 | 1.62E-22 | 1.991076 |
| S1PR1 | 34.39175 | 0.99444 | 0.333492 | 2.981896 | 0.002865 | 0.008736 | 1.992307 |
| TGFBR1 | 2668.062 | 0.994846 | 0.1444 | 6.889515 | 5.60E-12 | 8.75E-11 | 1.992868 |
| ZNF618 | 1531.319 | 0.995526 | 0.08589 | 11.59068 | 4.59E-31 | 4.80E-29 | 1.993807 |
| VPS41 | 1263.707 | 0.996778 | 0.097001 | 10.27598 | 9.04E-25 | 5.69E-23 | 1.995538 |
| CEP97 | 516.2284 | 0.997472 | 0.12668 | 7.873928 | 3.44E-15 | 8.01E-14 | 1.996498 |
| ZNF429 | 45.23488 | 0.997511 | 0.421359 | 2.367366 | 0.017915 | 0.043173 | 1.996553 |
| TAF1A | 267.5222 | 0.998273 | 0.148215 | 6.735299 | 1.64E-11 | 2.38E-10 | 1.997607 |
| HES1 | 408.6842 | 1.001578 | 0.170073 | 5.889104 | 3.88E-09 | 3.90E-08 | 2.002188 |

| | | | | | | | |
|---|---|---|---|---|---|---|---|
| DDIT4L | 152.7214 | 1.002162 | 0.170641 | 5.872922 | 4.28E-09 | 4.27E-08 | 2.002999 |
| GTDC1 | 411.1634 | 1.002563 | 0.126858 | 7.90302 | 2.72E-15 | 6.41E-14 | 2.003556 |
| HAPLN1 | 45.37883 | 1.004732 | 0.377216 | 2.663545 | 0.007732 | 0.020773 | 2.006571 |
| ZNF225 | 105.2086 | 1.00596 | 0.268801 | 3.742392 | 0.000182 | 0.000746 | 2.008279 |
| ASXL1 | 1061.869 | 1.006943 | 0.159246 | 6.323182 | 2.56E-10 | 3.15E-09 | 2.009649 |
| GPATCH11 | 172.5745 | 1.007781 | 0.156263 | 6.449272 | 1.12E-10 | 1.46E-09 | 2.010816 |
| E2F7 | 876.8215 | 1.009987 | 0.088838 | 11.36886 | 5.98E-30 | 5.49E-28 | 2.013893 |
| TET2 | 179.9226 | 1.011784 | 0.144938 | 6.980824 | 2.93E-12 | 4.79E-11 | 2.016403 |
| FAM72D | 232.7651 | 1.015528 | 0.202015 | 5.026988 | 4.98E-07 | 3.46E-06 | 2.021643 |
| TEAD1 | 2731.259 | 1.016146 | 0.100887 | 10.07214 | 7.34E-24 | 4.11E-22 | 2.022509 |
| YAF2 | 92.80268 | 1.016231 | 0.313985 | 3.236555 | 0.00121 | 0.004063 | 2.022628 |
| OTX2 | 2722.103 | 1.016255 | 0.156539 | 6.49202 | 8.47E-11 | 1.13E-09 | 2.022662 |
| RGMA | 2400.482 | 1.025293 | 0.161223 | 6.359485 | 2.02E-10 | 2.53E-09 | 2.035373 |
| RCBTB2 | 444.5469 | 1.025832 | 0.103561 | 9.905629 | 3.93E-23 | 2.05E-21 | 2.036134 |
| CPS1 | 1444.081 | 1.028963 | 0.111064 | 9.264569 | 1.96E-20 | 7.96E-19 | 2.040556 |
| EML1 | 1443.98 | 1.030172 | 0.15357 | 6.708178 | 1.97E-11 | 2.84E-10 | 2.042268 |
| NBPF8 | 136.1091 | 1.031921 | 0.199696 | 5.167454 | 2.37E-07 | 1.76E-06 | 2.044746 |
| KLHL8 | 307.2336 | 1.035066 | 0.134917 | 7.671882 | 1.69E-14 | 3.63E-13 | 2.049207 |
| MAPK4 | 209.2081 | 1.036448 | 0.181964 | 5.695912 | 1.23E-08 | 1.14E-07 | 2.051172 |
| RNF165 | 169.6314 | 1.037635 | 0.18407 | 5.637188 | 1.73E-08 | 1.57E-07 | 2.05286 |
| CROT | 123.7751 | 1.038145 | 0.184555 | 5.625124 | 1.85E-08 | 1.68E-07 | 2.053585 |
| TMEM170E | 306.1395 | 1.03852 | 0.115395 | 8.999673 | 2.26E-19 | 8.22E-18 | 2.054119 |
| SLITRK1 | 43.25537 | 1.040299 | 0.374451 | 2.778197 | 0.005466 | 0.015401 | 2.056654 |
| RACGAP1 | 1559.181 | 1.042582 | 0.10621 | 9.81622 | 9.59E-23 | 4.80E-21 | 2.059911 |
| PBK | 252.7252 | 1.044223 | 0.148221 | 7.045051 | 1.85E-12 | 3.11E-11 | 2.062255 |
| KIF20A | 1953.883 | 1.045404 | 0.0905 | 11.5514 | 7.26E-31 | 7.37E-29 | 2.063944 |
| SLIT1 | 91.40262 | 1.046151 | 0.278801 | 3.752319 | 0.000175 | 0.000721 | 2.065013 |
| SRSF12 | 353.7634 | 1.047011 | 0.157473 | 6.648814 | 2.95E-11 | 4.16E-10 | 2.066244 |
| F8 | 140.8645 | 1.048878 | 0.198605 | 5.281215 | 1.28E-07 | 1.00E-06 | 2.06892 |
| RAB36 | 107.5826 | 1.050502 | 0.180552 | 5.818269 | 5.95E-09 | 5.79E-08 | 2.07125 |
| ID2 | 1137.845 | 1.05067 | 0.175277 | 5.994339 | 2.04E-09 | 2.15E-08 | 2.071491 |
| TENM4 | 729.166 | 1.05398 | 0.213209 | 4.943421 | 7.68E-07 | 5.18E-06 | 2.07625 |
| ERCC4 | 308.4632 | 1.055635 | 0.205033 | 5.148614 | 2.62E-07 | 1.93E-06 | 2.078633 |
| PHF6 | 4831.184 | 1.058461 | 0.064425 | 16.42936 | 1.18E-60 | 8.33E-58 | 2.082708 |
| NRIP1 | 593.4803 | 1.059394 | 0.201495 | 5.257668 | 1.46E-07 | 1.13E-06 | 2.084055 |
| TNRC6B | 1371.743 | 1.059573 | 0.122175 | 8.672592 | 4.22E-18 | 1.38E-16 | 2.084315 |
| CITED2 | 5144.457 | 1.05997 | 0.156135 | 6.788823 | 1.13E-11 | 1.70E-10 | 2.084888 |
| RPS6KA3 | 1633.959 | 1.061896 | 0.078232 | 13.57371 | 5.73E-42 | 1.30E-39 | 2.087673 |
| EEF1AKMT | 67.8184 | 1.062308 | 0.293807 | 3.615665 | 0.0003 | 0.001172 | 2.08827 |
| ATP2B1 | 1985.639 | 1.062854 | 0.12377 | 8.587349 | 8.90E-18 | 2.82E-16 | 2.089061 |
| CENPI | 443.6578 | 1.063921 | 0.156495 | 6.798449 | 1.06E-11 | 1.59E-10 | 2.090606 |
| PRKCE | 44.79731 | 1.064466 | 0.439286 | 2.423173 | 0.015386 | 0.037794 | 2.091395 |
| CREB1 | 1777.127 | 1.064795 | 0.152063 | 7.002315 | 2.52E-12 | 4.15E-11 | 2.091873 |
| LPAR6 | 40.63191 | 1.065408 | 0.398518 | 2.673423 | 0.007508 | 0.020263 | 2.092762 |

| | | | | | | | |
|---|---|---|---|---|---|---|---|
| ZNF285 | 73.21516 | 1.069725 | 0.316029 | 3.384891 | 0.000712 | 0.002544 | 2.099033 |
| MBNL2 | 58.36227 | 1.070555 | 0.445548 | 2.402783 | 0.016271 | 0.039724 | 2.100241 |
| B9D2 | 51.09423 | 1.070787 | 0.271764 | 3.94013 | 8.14E-05 | 0.000364 | 2.100578 |
| FZD2 | 1286.783 | 1.071401 | 0.227087 | 4.71801 | 2.38E-06 | 1.46E-05 | 2.101473 |
| TMEM169 | 131.811 | 1.075194 | 0.276436 | 3.889489 | 0.0001 | 0.000441 | 2.107006 |
| SULT1C4 | 83.18115 | 1.076252 | 0.281802 | 3.819175 | 0.000134 | 0.000567 | 2.108551 |
| TAGLN3 | 72.2271 | 1.07659 | 0.26162 | 4.115087 | 3.87E-05 | 0.000186 | 2.109045 |
| PTPRD | 1669.101 | 1.077026 | 0.09929 | 10.84728 | 2.05E-27 | 1.55E-25 | 2.109683 |
| RNF217 | 293.5766 | 1.077088 | 0.347964 | 3.095399 | 0.001965 | 0.006267 | 2.109773 |
| 8-Mar | 480.9016 | 1.078306 | 0.152266 | 7.081735 | 1.42E-12 | 2.42E-11 | 2.111555 |
| FANCB | 135.1816 | 1.082428 | 0.199797 | 5.417628 | 6.04E-08 | 5.02E-07 | 2.117597 |
| GRASP | 99.38228 | 1.088667 | 0.250737 | 4.341867 | 1.41E-05 | 7.38E-05 | 2.126775 |
| RSBN1 | 628.7482 | 1.088707 | 0.194906 | 5.585813 | 2.33E-08 | 2.07E-07 | 2.126833 |
| PAX8 | 82.29562 | 1.089962 | 0.346341 | 3.147074 | 0.001649 | 0.005364 | 2.128684 |
| SACS | 1876.372 | 1.09214 | 0.114064 | 9.574826 | 1.02E-21 | 4.61E-20 | 2.1319 |
| UBA6 | 1768.127 | 1.096222 | 0.371127 | 2.953763 | 0.003139 | 0.009465 | 2.13794 |
| CCDC160 | 359.7367 | 1.097455 | 0.132513 | 8.281897 | 1.21E-16 | 3.40E-15 | 2.139769 |
| C5 | 108.9974 | 1.097877 | 0.36321 | 3.022708 | 0.002505 | 0.007754 | 2.140395 |
| POU3F1 | 711.9604 | 1.098695 | 0.12842 | 8.555476 | 1.17E-17 | 3.69E-16 | 2.141609 |
| CADM1 | 982.8089 | 1.099758 | 0.214809 | 5.119689 | 3.06E-07 | 2.22E-06 | 2.143187 |
| CBFB | 915.195 | 1.100541 | 0.156846 | 7.016713 | 2.27E-12 | 3.77E-11 | 2.144352 |
| RAB9B | 39.86113 | 1.10473 | 0.369901 | 2.986555 | 0.002821 | 0.008618 | 2.150587 |
| KCNK10 | 142.9637 | 1.105291 | 0.22524 | 4.907173 | 9.24E-07 | 6.13E-06 | 2.151423 |
| NR6A1 | 8738.925 | 1.106342 | 0.094262 | 11.73683 | 8.25E-32 | 9.25E-30 | 2.152991 |
| ITGA4 | 136.8562 | 1.107465 | 0.276322 | 4.007882 | 6.13E-05 | 0.000282 | 2.154668 |
| LIX1L | 170.4346 | 1.107564 | 0.159182 | 6.95785 | 3.46E-12 | 5.58E-11 | 2.154816 |
| ZNF148 | 590.301 | 1.108906 | 0.184111 | 6.023021 | 1.71E-09 | 1.83E-08 | 2.15682 |
| FIGN | 907.5349 | 1.109827 | 0.150072 | 7.395289 | 1.41E-13 | 2.70E-12 | 2.158198 |
| YPEL1 | 462.065 | 1.111151 | 0.233927 | 4.750001 | 2.03E-06 | 1.26E-05 | 2.16018 |
| GALNT10 | 671.2332 | 1.115559 | 0.199458 | 5.592962 | 2.23E-08 | 2.00E-07 | 2.166789 |
| TOP2A | 9902.905 | 1.117748 | 0.074389 | 15.02563 | 4.99E-51 | 2.04E-48 | 2.17008 |
| TIAM1 | 1117.192 | 1.117809 | 0.111069 | 10.06408 | 7.96E-24 | 4.43E-22 | 2.170171 |
| TTC3 | 2060.609 | 1.120288 | 0.114938 | 9.746904 | 1.90E-22 | 9.20E-21 | 2.173904 |
| RBM12B | 1515.892 | 1.120818 | 0.152263 | 7.361062 | 1.82E-13 | 3.44E-12 | 2.174703 |
| BCORL1 | 1093.546 | 1.121911 | 0.123473 | 9.086316 | 1.02E-19 | 3.89E-18 | 2.17635 |
| PDIK1L | 138.8316 | 1.125054 | 0.207557 | 5.420457 | 5.94E-08 | 4.95E-07 | 2.181096 |
| EFNB1 | 1331.957 | 1.126644 | 0.177978 | 6.33023 | 2.45E-10 | 3.02E-09 | 2.183503 |
| MBNL3 | 505.3931 | 1.126859 | 0.12632 | 8.920661 | 4.64E-19 | 1.65E-17 | 2.183828 |
| EFCAB12 | 31.148 | 1.127107 | 0.352669 | 3.19593 | 0.001394 | 0.004607 | 2.184202 |
| ASPRV1 | 27.81336 | 1.127434 | 0.418189 | 2.695991 | 0.007018 | 0.019131 | 2.184699 |
| BMPR1B | 43.86986 | 1.133937 | 0.377936 | 3.000338 | 0.002697 | 0.00829 | 2.194568 |
| IDI1 | 1096.659 | 1.134735 | 0.194881 | 5.822695 | 5.79E-09 | 5.65E-08 | 2.195783 |
| BMT2 | 190.9598 | 1.13589 | 0.201249 | 5.644204 | 1.66E-08 | 1.52E-07 | 2.197541 |
| FZD3 | 2420.273 | 1.136664 | 0.12176 | 9.335304 | 1.01E-20 | 4.21E-19 | 2.198721 |

| | | | | | | | |
|---|---|---|---|---|---|---|---|
| ADAMTS6 | 47.10772 | 1.136989 | 0.397506 | 2.860303 | 0.004232 | 0.012289 | 2.199215 |
| LRTOMT | 135.3989 | 1.138754 | 0.283626 | 4.014986 | 5.94E-05 | 0.000275 | 2.201908 |
| CCDC167 | 138.03 | 1.139496 | 0.18103 | 6.294501 | 3.08E-10 | 3.74E-09 | 2.20304 |
| PMF1-BGL | 92.85145 | 1.141656 | 0.215804 | 5.29025 | 1.22E-07 | 9.58E-07 | 2.206341 |
| TCAF1P1 | 215.7704 | 1.142928 | 0.151248 | 7.556645 | 4.14E-14 | 8.44E-13 | 2.208288 |
| CFL2 | 512.2304 | 1.144319 | 0.16399 | 6.977985 | 2.99E-12 | 4.88E-11 | 2.210417 |
| ANKRD36E | 53.11708 | 1.144779 | 0.374447 | 3.05725 | 0.002234 | 0.007007 | 2.211123 |
| KIAA0895 | 137.3499 | 1.146078 | 0.256168 | 4.473932 | 7.68E-06 | 4.26E-05 | 2.213114 |
| EPHB3 | 105.9056 | 1.146832 | 0.358616 | 3.19794 | 0.001384 | 0.004579 | 2.214271 |
| CASTOR3 | 277.5634 | 1.151336 | 0.214858 | 5.358579 | 8.39E-08 | 6.82E-07 | 2.221194 |
| ZNF343 | 142.1441 | 1.158379 | 0.219723 | 5.27199 | 1.35E-07 | 1.05E-06 | 2.232066 |
| CCDC80 | 366.6669 | 1.158401 | 0.226477 | 5.114867 | 3.14E-07 | 2.28E-06 | 2.232099 |
| TULP4 | 884.713 | 1.161014 | 0.167713 | 6.922632 | 4.43E-12 | 7.06E-11 | 2.236145 |
| DYRK3 | 227.2127 | 1.162329 | 0.179833 | 6.463397 | 1.02E-10 | 1.34E-09 | 2.238185 |
| H2AFY2 | 1578.545 | 1.163388 | 0.118837 | 9.789772 | 1.25E-22 | 6.15E-21 | 2.239828 |
| APOLD1 | 44.93671 | 1.167817 | 0.459779 | 2.539953 | 0.011087 | 0.028443 | 2.246715 |
| ZSCAN23 | 80.1111 | 1.169687 | 0.310851 | 3.762851 | 0.000168 | 0.000693 | 2.249628 |
| ZNF77 | 35.70916 | 1.171075 | 0.398683 | 2.937359 | 0.00331 | 0.009912 | 2.251795 |
| AMPH | 380.8146 | 1.176427 | 0.102968 | 11.42514 | 3.13E-30 | 2.98E-28 | 2.260163 |
| RLN2 | 51.7104 | 1.178636 | 0.343314 | 3.433117 | 0.000597 | 0.002175 | 2.263627 |
| RFX2 | 130.1471 | 1.179866 | 0.269953 | 4.370638 | 1.24E-05 | 6.55E-05 | 2.265557 |
| DUSP4 | 609.4326 | 1.182157 | 0.120593 | 9.802867 | 1.09E-22 | 5.44E-21 | 2.269158 |
| TLE4 | 1391.037 | 1.185012 | 0.123016 | 9.632979 | 5.80E-22 | 2.69E-20 | 2.273653 |
| TBC1D19 | 108.4568 | 1.186488 | 0.242279 | 4.89719 | 9.72E-07 | 6.40E-06 | 2.27598 |
| ETV3L | 19.90632 | 1.187208 | 0.436636 | 2.718989 | 0.006548 | 0.01803 | 2.277117 |
| CYP26A1 | 1645.951 | 1.187405 | 0.375398 | 3.163057 | 0.001561 | 0.005109 | 2.277427 |
| DLL1 | 205.2444 | 1.190403 | 0.182613 | 6.518715 | 7.09E-11 | 9.54E-10 | 2.282165 |
| PCDHGC4 | 309.357 | 1.190621 | 0.162652 | 7.320055 | 2.48E-13 | 4.60E-12 | 2.282509 |
| ZNF608 | 1617.903 | 1.19378 | 0.104755 | 11.39588 | 4.38E-30 | 4.10E-28 | 2.287513 |
| FBN1 | 919.0218 | 1.193981 | 0.148697 | 8.029619 | 9.78E-16 | 2.41E-14 | 2.287832 |
| APPBP2 | 548.8092 | 1.19575 | 0.186597 | 6.408181 | 1.47E-10 | 1.87E-09 | 2.290638 |
| PTPN13 | 6925.02 | 1.196702 | 0.100559 | 11.90054 | 1.18E-32 | 1.42E-30 | 2.292151 |
| EFR3B | 774.5371 | 1.196824 | 0.115991 | 10.31823 | 5.83E-25 | 3.71E-23 | 2.292344 |
| ZSCAN20 | 112.9538 | 1.197349 | 0.203501 | 5.88376 | 4.01E-09 | 4.01E-08 | 2.293178 |
| GAD1 | 162.3221 | 1.19836 | 0.305125 | 3.927442 | 8.59E-05 | 0.000382 | 2.294787 |
| RTN4RL1 | 22.82095 | 1.198793 | 0.475755 | 2.519772 | 0.011743 | 0.029947 | 2.295476 |
| GLDN | 26.35879 | 1.199323 | 0.430452 | 2.786193 | 0.005333 | 0.015073 | 2.296319 |
| ZNF664 | 3058.609 | 1.200227 | 0.091662 | 13.09411 | 3.56E-39 | 6.35E-37 | 2.297758 |
| TNFRSF11 | 39.60461 | 1.200651 | 0.39951 | 3.00531 | 0.002653 | 0.008169 | 2.298434 |
| COMMD8 | 80.61942 | 1.20345 | 0.221801 | 5.42581 | 5.77E-08 | 4.82E-07 | 2.302897 |
| AC005943 | 99.76813 | 1.208597 | 0.432181 | 2.796505 | 0.005166 | 0.014674 | 2.311128 |
| RBM15 | 524.8658 | 1.210528 | 0.104274 | 11.60913 | 3.70E-31 | 3.91E-29 | 2.314223 |
| HMGCS1 | 3096.383 | 1.210873 | 0.194167 | 6.236243 | 4.48E-10 | 5.27E-09 | 2.314777 |
| PSD3 | 830.8413 | 1.212803 | 0.148205 | 8.183296 | 2.76E-16 | 7.32E-15 | 2.317876 |

| | | | | | | | |
|---|---|---|---|---|---|---|---|
| DMD | 959.9856 | 1.213731 | 0.114965 | 10.55744 | 4.69E-26 | 3.22E-24 | 2.319367 |
| PKNOX2 | 193.8319 | 1.215062 | 0.231674 | 5.2447 | 1.57E-07 | 1.20E-06 | 2.321508 |
| CKAP2L | 775.4162 | 1.216488 | 0.124821 | 9.745866 | 1.92E-22 | 9.27E-21 | 2.323804 |
| PLXNC1 | 105.0544 | 1.227122 | 0.366332 | 3.349756 | 0.000809 | 0.002845 | 2.340995 |
| ESCO2 | 462.4315 | 1.24066 | 0.278046 | 4.462066 | 8.12E-06 | 4.47E-05 | 2.363066 |
| KLHDC8A | 135.0876 | 1.24318 | 0.238681 | 5.208536 | 1.90E-07 | 1.44E-06 | 2.367197 |
| PIH1D2 | 22.87226 | 1.248239 | 0.382377 | 3.26442 | 0.001097 | 0.003728 | 2.375512 |
| MSRB3 | 74.55194 | 1.25164 | 0.244333 | 5.122689 | 3.01E-07 | 2.19E-06 | 2.381119 |
| ELP4 | 714.107 | 1.253405 | 0.143647 | 8.725576 | 2.65E-18 | 8.77E-17 | 2.384035 |
| IQCK | 141.3783 | 1.253624 | 0.21052 | 5.954903 | 2.60E-09 | 2.69E-08 | 2.384396 |
| SYT11 | 44.42463 | 1.254755 | 0.330655 | 3.794754 | 0.000148 | 0.00062 | 2.386267 |
| AFF3 | 72.13196 | 1.256326 | 0.419707 | 2.993341 | 0.002759 | 0.008455 | 2.388866 |
| NCOA6 | 747.3133 | 1.260728 | 0.089349 | 14.11023 | 3.29E-45 | 9.15E-43 | 2.396167 |
| SDK2 | 2236.037 | 1.262987 | 0.121571 | 10.38884 | 2.79E-25 | 1.83E-23 | 2.39992 |
| TMSB15A | 757.9308 | 1.270455 | 0.124631 | 10.19377 | 2.11E-24 | 1.29E-22 | 2.412377 |
| NEBL | 1534.039 | 1.274319 | 0.1578 | 8.075548 | 6.72E-16 | 1.69E-14 | 2.418846 |
| CXCR4 | 102.3341 | 1.274427 | 0.381303 | 3.342297 | 0.000831 | 0.002916 | 2.419028 |
| IRX1 | 383.5204 | 1.27535 | 0.246577 | 5.172215 | 2.31E-07 | 1.72E-06 | 2.420575 |
| MT1H | 49.46184 | 1.28095 | 0.325582 | 3.934336 | 8.34E-05 | 0.000372 | 2.42999 |
| BTG2 | 163.5536 | 1.285695 | 0.321046 | 4.004706 | 6.21E-05 | 0.000285 | 2.437995 |
| ZNF516 | 974.0916 | 1.288191 | 0.187321 | 6.876909 | 6.12E-12 | 9.50E-11 | 2.442216 |
| DNAJA1 | 3988.12 | 1.288346 | 0.100688 | 12.7954 | 1.74E-37 | 2.85E-35 | 2.442479 |
| IFIT1 | 226.4006 | 1.296214 | 0.215238 | 6.022245 | 1.72E-09 | 1.84E-08 | 2.455836 |
| NSUN3 | 244.5436 | 1.299252 | 0.207214 | 6.270087 | 3.61E-10 | 4.34E-09 | 2.461013 |
| CDH11 | 744.9283 | 1.300076 | 0.129671 | 10.02599 | 1.17E-23 | 6.33E-22 | 2.462419 |
| FAM89A | 25.58064 | 1.30216 | 0.439478 | 2.96297 | 0.003047 | 0.009224 | 2.465978 |
| AC091959 | 161.391 | 1.311532 | 0.310793 | 4.219956 | 2.44E-05 | 0.000123 | 2.48205 |
| FJX1 | 1391.984 | 1.312992 | 0.174725 | 7.514615 | 5.71E-14 | 1.15E-12 | 2.484563 |
| ZNF518B | 513.1885 | 1.319751 | 0.193775 | 6.810728 | 9.71E-12 | 1.47E-10 | 2.496231 |
| FBXL14 | 238.8383 | 1.319832 | 0.170691 | 7.73229 | 1.06E-14 | 2.32E-13 | 2.496371 |
| CNKSR2 | 732.2322 | 1.320882 | 0.09888 | 13.35838 | 1.06E-40 | 2.11E-38 | 2.498187 |
| AURKC | 96.34241 | 1.333425 | 0.20718 | 6.436065 | 1.23E-10 | 1.58E-09 | 2.520003 |
| HSPA8 | 24835.41 | 1.336617 | 0.189087 | 7.068784 | 1.56E-12 | 2.64E-11 | 2.525584 |
| HMGB1P5 | 102.1534 | 1.338247 | 0.308621 | 4.336216 | 1.45E-05 | 7.55E-05 | 2.528439 |
| GFRA1 | 188.8582 | 1.338545 | 0.436196 | 3.068674 | 0.00215 | 0.006778 | 2.528961 |
| PTN | 1966.56 | 1.339336 | 0.146632 | 9.133974 | 6.60E-20 | 2.55E-18 | 2.530349 |
| SCUBE1 | 421.6091 | 1.341671 | 0.212836 | 6.303789 | 2.90E-10 | 3.54E-09 | 2.534448 |
| MAP2K6 | 1353.416 | 1.34511 | 0.14 | 9.607948 | 7.40E-22 | 3.38E-20 | 2.540496 |
| SIPA1L2 | 842.8845 | 1.347251 | 0.109723 | 12.27866 | 1.18E-34 | 1.63E-32 | 2.544269 |
| TPBG | 1739.798 | 1.3516 | 0.133625 | 10.11489 | 4.75E-24 | 2.77E-22 | 2.55195 |
| HECTD2 | 698.1369 | 1.352581 | 0.100461 | 13.46369 | 2.56E-41 | 5.40E-39 | 2.553686 |
| LRRC4 | 613.5808 | 1.353584 | 0.113269 | 11.95022 | 6.48E-33 | 7.88E-31 | 2.555462 |
| HCFC2 | 61.27707 | 1.354844 | 0.334904 | 4.045472 | 5.22E-05 | 0.000244 | 2.557695 |
| BLACAT1 | 659.5276 | 1.36699 | 0.15186 | 9.001658 | 2.22E-19 | 8.11E-18 | 2.579319 |

| | | | | | | | |
|---|---|---|---|---|---|---|---|
| HEY1 | 46.99624 | 1.369087 | 0.34476 | 3.971132 | 7.15E-05 | 0.000323 | 2.583071 |
| DOCK10 | 26.98729 | 1.374771 | 0.389328 | 3.531138 | 0.000414 | 0.001569 | 2.593268 |
| STMN2 | 58.23516 | 1.377438 | 0.41571 | 3.313458 | 0.000921 | 0.003199 | 2.598065 |
| EPHA7 | 585.1665 | 1.378753 | 0.172635 | 7.98654 | 1.39E-15 | 3.37E-14 | 2.600435 |
| CCDC146 | 17.77592 | 1.384711 | 0.549554 | 2.519699 | 0.011746 | 0.029949 | 2.611196 |
| STOX1 | 188.3461 | 1.39215 | 0.149611 | 9.305135 | 1.34E-20 | 5.53E-19 | 2.624695 |
| FANK1 | 20.80871 | 1.399444 | 0.518056 | 2.701334 | 0.006906 | 0.018863 | 2.637998 |
| MAK | 18.40022 | 1.400484 | 0.488725 | 2.865589 | 0.004162 | 0.012116 | 2.639902 |
| MLLT3 | 222.7978 | 1.403588 | 0.208245 | 6.740089 | 1.58E-11 | 2.32E-10 | 2.645587 |
| TRIM61 | 121.3072 | 1.406506 | 0.232997 | 6.036592 | 1.57E-09 | 1.69E-08 | 2.650944 |
| SPON1 | 468.5828 | 1.409607 | 0.129157 | 10.91391 | 9.89E-28 | 7.70E-26 | 2.656647 |
| SCD5 | 98.62161 | 1.417647 | 0.252131 | 5.622654 | 1.88E-08 | 1.70E-07 | 2.671495 |
| PCDH17 | 32.12327 | 1.417878 | 0.363539 | 3.900212 | 9.61E-05 | 0.000424 | 2.671922 |
| TSHZ2 | 36.16395 | 1.424828 | 0.539895 | 2.639084 | 0.008313 | 0.022143 | 2.684824 |
| JCAD | 194.143 | 1.426345 | 0.186326 | 7.655121 | 1.93E-14 | 4.11E-13 | 2.687649 |
| AC118553 | 68.34788 | 1.428635 | 0.458875 | 3.113343 | 0.00185 | 0.005948 | 2.69192 |
| PAMR1 | 1036.156 | 1.429931 | 0.259777 | 5.504447 | 3.70E-08 | 3.20E-07 | 2.694337 |
| RIN2 | 45.36216 | 1.442087 | 0.501172 | 2.877429 | 0.004009 | 0.011711 | 2.717137 |
| CNTNAP2 | 1412.904 | 1.446089 | 0.106794 | 13.54092 | 8.97E-42 | 1.94E-39 | 2.724684 |
| GRAMD1C | 73.25079 | 1.449298 | 0.377906 | 3.835079 | 0.000126 | 0.000535 | 2.730752 |
| PCDHGB6 | 698.5512 | 1.452582 | 0.132719 | 10.94479 | 7.04E-28 | 5.55E-26 | 2.736975 |
| CCDC121 | 78.56738 | 1.453324 | 0.284189 | 5.113941 | 3.16E-07 | 2.29E-06 | 2.738383 |
| TRPM3 | 394.678 | 1.454309 | 0.142257 | 10.22312 | 1.56E-24 | 9.60E-23 | 2.740252 |
| USP3 | 493.1811 | 1.45436 | 0.122988 | 11.82527 | 2.89E-32 | 3.32E-30 | 2.74035 |
| CAMK2D | 795.5089 | 1.454978 | 0.123206 | 11.8093 | 3.49E-32 | 3.99E-30 | 2.741525 |
| NTF3 | 44.1841 | 1.45687 | 0.427984 | 3.404031 | 0.000664 | 0.002389 | 2.745122 |
| PKDREJ | 23.55169 | 1.463216 | 0.536718 | 2.726227 | 0.006406 | 0.017703 | 2.757222 |
| S1PR3 | 289.8539 | 1.464014 | 0.187316 | 7.815752 | 5.46E-15 | 1.24E-13 | 2.758749 |
| ENPP2 | 136.8087 | 1.465174 | 0.238097 | 6.153675 | 7.57E-10 | 8.56E-09 | 2.760967 |
| EXOC6 | 332.0651 | 1.466368 | 0.148562 | 9.870415 | 5.59E-23 | 2.87E-21 | 2.763255 |
| SOX11 | 3739.078 | 1.468259 | 0.112643 | 13.03462 | 7.78E-39 | 1.35E-36 | 2.766877 |
| LEMD1 | 31.50809 | 1.471025 | 0.363219 | 4.049962 | 5.12E-05 | 0.00024 | 2.772187 |
| TMEM88 | 72.83216 | 1.471438 | 0.309427 | 4.755369 | 1.98E-06 | 1.23E-05 | 2.772982 |
| GRK5 | 77.49503 | 1.479185 | 0.260248 | 5.683746 | 1.32E-08 | 1.22E-07 | 2.787913 |
| MAP6 | 182.2521 | 1.484549 | 0.16597 | 8.944669 | 3.73E-19 | 1.33E-17 | 2.798296 |
| GLUD2 | 96.71097 | 1.489029 | 0.243319 | 6.119666 | 9.38E-10 | 1.04E-08 | 2.807 |
| ZBBX | 50.34952 | 1.495589 | 0.454588 | 3.289988 | 0.001002 | 0.003444 | 2.819792 |
| RSPH4A | 15.31129 | 1.498146 | 0.492264 | 3.043381 | 0.002339 | 0.007298 | 2.824795 |
| SRR | 237.8986 | 1.49918 | 0.196243 | 7.639412 | 2.18E-14 | 4.63E-13 | 2.826819 |
| ZNF287 | 179.7732 | 1.50784 | 0.25443 | 5.926337 | 3.10E-09 | 3.16E-08 | 2.84384 |
| SEZ6L | 178.9439 | 1.512829 | 0.205745 | 7.352931 | 1.94E-13 | 3.65E-12 | 2.853691 |
| ST6GALNA | 340.8732 | 1.520594 | 0.156783 | 9.698722 | 3.05E-22 | 1.44E-20 | 2.869092 |
| NRGN | 18.49333 | 1.525331 | 0.500074 | 3.050211 | 0.002287 | 0.007156 | 2.878527 |
| LRRC55 | 151.4354 | 1.526198 | 0.172582 | 8.843346 | 9.29E-19 | 3.21E-17 | 2.880259 |

| | | | | | | | |
|---|---|---|---|---|---|---|---|
| SOAT1 | 607.4154 | 1.529894 | 0.18299 | 8.360533 | 6.24E-17 | 1.82E-15 | 2.887646 |
| DYDC2 | 49.08279 | 1.530835 | 0.398675 | 3.839803 | 0.000123 | 0.000526 | 2.88953 |
| SCUBE3 | 210.2849 | 1.532323 | 0.161127 | 9.510023 | 1.91E-21 | 8.36E-20 | 2.892513 |
| CEMIP2 | 2608.706 | 1.535709 | 0.327647 | 4.687087 | 2.77E-06 | 1.67E-05 | 2.89931 |
| GLIPR1 | 30.12163 | 1.549276 | 0.588449 | 2.632813 | 0.008468 | 0.022512 | 2.926702 |
| NAV1 | 3280.844 | 1.5502 | 0.128157 | 12.09613 | 1.11E-33 | 1.44E-31 | 2.928578 |
| NEDD9 | 103.3906 | 1.557335 | 0.370568 | 4.202563 | 2.64E-05 | 0.000132 | 2.943097 |
| SPA17 | 138.862 | 1.559634 | 0.304951 | 5.114369 | 3.15E-07 | 2.28E-06 | 2.947791 |
| SEMA3C | 41.74913 | 1.567498 | 0.408851 | 3.833911 | 0.000126 | 0.000537 | 2.963903 |
| ST8SIA4 | 16.49959 | 1.576527 | 0.492123 | 3.203522 | 0.001358 | 0.004501 | 2.98251 |
| MEGF9 | 94.6183 | 1.576556 | 0.211038 | 7.470492 | 7.99E-14 | 1.58E-12 | 2.982571 |
| CDH6 | 441.6482 | 1.589375 | 0.316227 | 5.026058 | 5.01E-07 | 3.48E-06 | 3.009189 |
| PAPPA | 284.8992 | 1.596207 | 0.278562 | 5.730167 | 1.00E-08 | 9.49E-08 | 3.023473 |
| NCALD | 130.2312 | 1.596271 | 0.284828 | 5.604329 | 2.09E-08 | 1.88E-07 | 3.023609 |
| MAPK10 | 827.0036 | 1.612266 | 0.13562 | 11.8881 | 1.36E-32 | 1.64E-30 | 3.057318 |
| IRF8 | 10.95748 | 1.612733 | 0.693468 | 2.325607 | 0.02004 | 0.047474 | 3.058307 |
| WDR63 | 8.593141 | 1.614589 | 0.630586 | 2.560457 | 0.010453 | 0.027045 | 3.062244 |
| CD37 | 40.19431 | 1.619223 | 0.528274 | 3.065121 | 0.002176 | 0.006843 | 3.072095 |
| EPHA3 | 37.29272 | 1.628623 | 0.401518 | 4.056165 | 4.99E-05 | 0.000234 | 3.092178 |
| PPP1R3C | 108.3642 | 1.637074 | 0.277339 | 5.902792 | 3.57E-09 | 3.61E-08 | 3.110344 |
| FHOD3 | 978.1571 | 1.637339 | 0.096471 | 16.97233 | 1.32E-64 | 1.05E-61 | 3.110916 |
| CLVS2 | 56.84805 | 1.644772 | 0.431131 | 3.815014 | 0.000136 | 0.000576 | 3.126985 |
| NRG1 | 200.7257 | 1.648577 | 0.227827 | 7.236101 | 4.62E-13 | 8.26E-12 | 3.135243 |
| RNF152 | 151.8081 | 1.649978 | 0.4105 | 4.019436 | 5.83E-05 | 0.00027 | 3.138289 |
| ARID5B | 9.631978 | 1.65914 | 0.706743 | 2.347588 | 0.018895 | 0.045125 | 3.158283 |
| HMGCLL1 | 23.44734 | 1.660297 | 0.43996 | 3.773742 | 0.000161 | 0.000668 | 3.160815 |
| CNTN2 | 396.3039 | 1.672577 | 0.180301 | 9.276592 | 1.75E-20 | 7.21E-19 | 3.187835 |
| ME3 | 120.7626 | 1.67523 | 0.291101 | 5.7548 | 8.67E-09 | 8.25E-08 | 3.193703 |
| GPR85 | 17.09633 | 1.675748 | 0.645751 | 2.595035 | 0.009458 | 0.024819 | 3.194849 |
| MID1 | 1157.531 | 1.682377 | 0.149456 | 11.25671 | 2.15E-29 | 1.87E-27 | 3.209563 |
| CBX8 | 63.8969 | 1.684783 | 0.292372 | 5.762469 | 8.29E-09 | 7.91E-08 | 3.21492 |
| MEX3B | 553.9781 | 1.687991 | 0.155078 | 10.8848 | 1.36E-27 | 1.04E-25 | 3.222077 |
| FRZB | 813.568 | 1.699196 | 0.272493 | 6.235734 | 4.50E-10 | 5.29E-09 | 3.2472 |
| GPR1 | 13.36857 | 1.705463 | 0.730073 | 2.336016 | 0.01949 | 0.046335 | 3.261336 |
| CNTNAP3F | 61.57824 | 1.709622 | 0.32191 | 5.310866 | 1.09E-07 | 8.65E-07 | 3.270752 |
| DMBX1 | 151.1057 | 1.712026 | 0.379313 | 4.51349 | 6.38E-06 | 3.59E-05 | 3.276206 |
| SNURF | 97.85319 | 1.714382 | 0.516816 | 3.317198 | 0.000909 | 0.003163 | 3.281561 |
| ACRV1 | 24.62031 | 1.722301 | 0.589309 | 2.922577 | 0.003471 | 0.010312 | 3.299622 |
| AC023055 | 26.74425 | 1.723555 | 0.427412 | 4.032535 | 5.52E-05 | 0.000257 | 3.302491 |
| EPHA4 | 1778.642 | 1.725622 | 0.34668 | 4.977567 | 6.44E-07 | 4.39E-06 | 3.307227 |
| RBM24 | 95.22231 | 1.72619 | 0.251748 | 6.856814 | 7.04E-12 | 1.08E-10 | 3.308529 |
| GAS1 | 365.2729 | 1.740253 | 0.19852 | 8.766132 | 1.85E-18 | 6.19E-17 | 3.340938 |
| TSPAN18 | 675.4608 | 1.744309 | 0.117009 | 14.90744 | 2.95E-50 | 1.15E-47 | 3.350343 |
| LONRF3 | 13.34952 | 1.747728 | 0.573028 | 3.049988 | 0.002289 | 0.007158 | 3.358294 |

| | | | | | | | |
|---|---|---|---|---|---|---|---|
| APBA1 | 73.39617 | 1.751302 | 0.421468 | 4.155243 | 3.25E-05 | 0.000159 | 3.366624 |
| LMO3 | 44.89808 | 1.751676 | 0.333385 | 5.254217 | 1.49E-07 | 1.15E-06 | 3.367495 |
| FST | 2946.496 | 1.752629 | 0.212257 | 8.257097 | 1.49E-16 | 4.14E-15 | 3.36972 |
| CXXC4 | 177.6608 | 1.756263 | 0.236328 | 7.431467 | 1.07E-13 | 2.09E-12 | 3.37822 |
| SOX9 | 776.164 | 1.757239 | 0.116154 | 15.12848 | 1.05E-51 | 4.49E-49 | 3.380504 |
| ANGPT1 | 49.40107 | 1.768315 | 0.323146 | 5.47218 | 4.45E-08 | 3.79E-07 | 3.406558 |
| ARHGAP11 | 18.64804 | 1.776775 | 0.493722 | 3.598736 | 0.00032 | 0.001244 | 3.426594 |
| AL035425. | 40.93955 | 1.778016 | 0.308177 | 5.769473 | 7.95E-09 | 7.64E-08 | 3.429543 |
| CCBE1 | 343.0104 | 1.778876 | 0.233048 | 7.633086 | 2.29E-14 | 4.84E-13 | 3.431586 |
| PDE10A | 136.0241 | 1.779175 | 0.295442 | 6.022072 | 1.72E-09 | 1.84E-08 | 3.432298 |
| KIF25 | 28.53173 | 1.782113 | 0.347238 | 5.132258 | 2.86E-07 | 2.10E-06 | 3.439294 |
| LPAR1 | 185.6537 | 1.788824 | 0.187944 | 9.517875 | 1.77E-21 | 7.81E-20 | 3.455331 |
| GLI3 | 1447.345 | 1.789194 | 0.12293 | 14.55454 | 5.47E-48 | 1.79E-45 | 3.456217 |
| MN1 | 725.3233 | 1.793124 | 0.122876 | 14.59291 | 3.12E-48 | 1.04E-45 | 3.465645 |
| FRMD3 | 63.10212 | 1.810232 | 0.351386 | 5.151697 | 2.58E-07 | 1.90E-06 | 3.506986 |
| RTL9 | 11.97796 | 1.810435 | 0.725081 | 2.496872 | 0.012529 | 0.031628 | 3.507481 |
| ZNF624 | 72.03542 | 1.823839 | 0.327863 | 5.562814 | 2.65E-08 | 2.34E-07 | 3.54022 |
| SNX18P7 | 9.31266 | 1.83195 | 0.713195 | 2.568651 | 0.01021 | 0.026488 | 3.560179 |
| ARL4C | 1747.866 | 1.838433 | 0.172648 | 10.64845 | 1.77E-26 | 1.29E-24 | 3.576214 |
| ZNF436 | 87.88061 | 1.839258 | 0.232368 | 7.915272 | 2.47E-15 | 5.84E-14 | 3.578259 |
| TNFRSF19 | 445.3561 | 1.844205 | 0.290548 | 6.347337 | 2.19E-10 | 2.72E-09 | 3.590551 |
| SOX21 | 2822.419 | 1.849603 | 0.187368 | 9.871492 | 5.53E-23 | 2.85E-21 | 3.604009 |
| FLRT3 | 298.2419 | 1.87853 | 0.504563 | 3.723082 | 0.000197 | 0.000801 | 3.677002 |
| SHISA2 | 1844.078 | 1.882075 | 0.227192 | 8.284072 | 1.19E-16 | 3.35E-15 | 3.686048 |
| EGR3 | 71.57142 | 1.890218 | 0.279065 | 6.773405 | 1.26E-11 | 1.87E-10 | 3.706912 |
| ZNF704 | 304.6004 | 1.894306 | 0.351056 | 5.396024 | 6.81E-08 | 5.61E-07 | 3.71743 |
| SOX6 | 22.90246 | 1.896891 | 0.598596 | 3.168903 | 0.00153 | 0.005019 | 3.724098 |
| CDH2 | 2980.707 | 1.899618 | 0.178377 | 10.64947 | 1.75E-26 | 1.28E-24 | 3.731143 |
| SNAI2 | 11.20239 | 1.903565 | 0.640502 | 2.971988 | 0.002959 | 0.008985 | 3.741367 |
| RPH3A | 12.66259 | 1.905876 | 0.696332 | 2.737023 | 0.0062 | 0.017205 | 3.747363 |
| ALDH1A2 | 82.07841 | 1.908951 | 0.5889 | 3.241556 | 0.001189 | 0.004003 | 3.75536 |
| RSPO1 | 64.2228 | 1.91397 | 0.370806 | 5.16165 | 2.45E-07 | 1.81E-06 | 3.768447 |
| FGF8 | 50.51445 | 1.91667 | 0.354961 | 5.399667 | 6.68E-08 | 5.51E-07 | 3.775505 |
| LRRN3 | 126.5861 | 1.920628 | 0.422525 | 4.545602 | 5.48E-06 | 3.12E-05 | 3.785879 |
| SORCS2 | 146.0915 | 1.932592 | 0.23931 | 8.075699 | 6.71E-16 | 1.69E-14 | 3.817404 |
| CLEC18B | 15.09679 | 1.932611 | 0.749768 | 2.577612 | 0.009949 | 0.025904 | 3.817454 |
| TMEM74 | 224.441 | 1.9422 | 0.16702 | 11.62852 | 2.95E-31 | 3.17E-29 | 3.842913 |
| RAI1 | 281.3928 | 1.945709 | 0.206308 | 9.431096 | 4.06E-21 | 1.74E-19 | 3.852272 |
| KAT2B | 76.7104 | 1.949502 | 0.31437 | 6.201293 | 5.60E-10 | 6.49E-09 | 3.862412 |
| CCNG2 | 723.316 | 1.957903 | 0.127027 | 15.41326 | 1.33E-53 | 6.45E-51 | 3.884967 |
| CNTNAP5 | 25.94425 | 1.997451 | 0.665242 | 3.002592 | 0.002677 | 0.008234 | 3.99294 |
| DPYSL5 | 2241.707 | 1.999073 | 0.134617 | 14.85011 | 6.95E-50 | 2.61E-47 | 3.997431 |
| ZNF521 | 1107.06 | 2.007887 | 0.166229 | 12.07904 | 1.36E-33 | 1.75E-31 | 4.021929 |
| SOX5 | 88.07528 | 2.011672 | 0.340018 | 5.916367 | 3.29E-09 | 3.34E-08 | 4.032494 |

| | | | | | | | |
|---|---|---|---|---|---|---|---|
| FEZF1 | 3895.397 | 2.013077 | 0.308081 | 6.53425 | 6.39E-11 | 8.66E-10 | 4.036423 |
| CCDC177 | 331.2259 | 2.033106 | 0.169087 | 12.02403 | 2.66E-33 | 3.30E-31 | 4.092849 |
| IRS4 | 293.3119 | 2.040472 | 0.146684 | 13.91062 | 5.46E-44 | 1.39E-41 | 4.113801 |
| NPAS3 | 19.63261 | 2.043752 | 0.549071 | 3.722201 | 0.000197 | 0.000803 | 4.123165 |
| ESRRB | 29.47946 | 2.045874 | 0.44773 | 4.569434 | 4.89E-06 | 2.82E-05 | 4.129234 |
| CYP7B1 | 52.45548 | 2.05237 | 0.45121 | 4.548593 | 5.40E-06 | 3.08E-05 | 4.147868 |
| SKIDA1 | 706.4293 | 2.073015 | 0.166034 | 12.48551 | 8.96E-36 | 1.30E-33 | 4.207652 |
| AC025165 | 29.1445 | 2.074114 | 0.834028 | 2.486863 | 0.012887 | 0.032424 | 4.210859 |
| PRSS23 | 563.6664 | 2.077745 | 0.134685 | 15.42672 | 1.08E-53 | 5.38E-51 | 4.221468 |
| SMIM32 | 8.73355 | 2.082669 | 0.762923 | 2.729854 | 0.006336 | 0.017546 | 4.235901 |
| BCO1 | 7.561295 | 2.086172 | 0.902026 | 2.312763 | 0.020736 | 0.04889 | 4.246198 |
| TBC1D9 | 123.1584 | 2.094812 | 0.22018 | 9.514072 | 1.83E-21 | 8.06E-20 | 4.271704 |
| AC093151 | 11.44694 | 2.111552 | 0.733988 | 2.876819 | 0.004017 | 0.01173 | 4.321559 |
| GUCY1B1 | 46.74708 | 2.112359 | 0.34252 | 6.167107 | 6.96E-10 | 7.91E-09 | 4.323978 |
| RPL23AP4! | 9.877929 | 2.114843 | 0.82389 | 2.5669 | 0.010261 | 0.0266 | 4.331427 |
| EFCC1 | 30.74821 | 2.122701 | 0.451155 | 4.705041 | 2.54E-06 | 1.54E-05 | 4.355084 |
| ZNF781 | 43.08278 | 2.13853 | 0.407186 | 5.251977 | 1.50E-07 | 1.16E-06 | 4.403133 |
| TSPAN11 | 331.7173 | 2.14802 | 0.280426 | 7.659843 | 1.86E-14 | 3.98E-13 | 4.432192 |
| DCLK2 | 59.20083 | 2.148941 | 0.488475 | 4.399281 | 1.09E-05 | 5.81E-05 | 4.435021 |
| CER1 | 31.72287 | 2.158635 | 0.498177 | 4.333073 | 1.47E-05 | 7.65E-05 | 4.464923 |
| SCUBE2 | 46.83978 | 2.192722 | 0.504007 | 4.350575 | 1.36E-05 | 7.12E-05 | 4.571672 |
| IER5L | 85.30864 | 2.219011 | 0.226112 | 9.813765 | 9.82E-23 | 4.91E-21 | 4.655743 |
| HESX1 | 452.3979 | 2.233973 | 0.196802 | 11.35135 | 7.30E-30 | 6.68E-28 | 4.704276 |
| FGF17 | 22.36656 | 2.247477 | 0.453051 | 4.960765 | 7.02E-07 | 4.76E-06 | 4.748517 |
| OLFM3 | 62.08888 | 2.247762 | 0.477758 | 4.704816 | 2.54E-06 | 1.55E-05 | 4.749454 |
| SPATA17 | 30.27167 | 2.255565 | 0.658392 | 3.42587 | 0.000613 | 0.002226 | 4.775214 |
| C9orf43 | 10.98602 | 2.260146 | 0.753721 | 2.998652 | 0.002712 | 0.008329 | 4.790399 |
| PYGO1 | 150.5597 | 2.261354 | 0.220867 | 10.23851 | 1.33E-24 | 8.30E-23 | 4.794412 |
| MYO3B | 95.01011 | 2.26328 | 0.222837 | 10.15668 | 3.09E-24 | 1.84E-22 | 4.800817 |
| IGFBP5 | 625.2919 | 2.271561 | 0.232909 | 9.753009 | 1.79E-22 | 8.68E-21 | 4.828452 |
| MPPED1 | 44.1581 | 2.272032 | 0.422402 | 5.378836 | 7.50E-08 | 6.12E-07 | 4.830031 |
| COL3A1 | 37.60855 | 2.27932 | 0.440699 | 5.172051 | 2.32E-07 | 1.72E-06 | 4.854492 |
| MAP2 | 1364.083 | 2.284626 | 0.12933 | 17.66515 | 7.78E-70 | 8.41E-67 | 4.872379 |
| MDGA1 | 68.52241 | 2.28739 | 0.465547 | 4.913339 | 8.95E-07 | 5.96E-06 | 4.881721 |
| VWF | 327.7134 | 2.296347 | 0.332397 | 6.908444 | 4.90E-12 | 7.72E-11 | 4.912123 |
| ZNF680 | 107.116 | 2.297492 | 0.26489 | 8.673375 | 4.19E-18 | 1.37E-16 | 4.916022 |
| ADAM12 | 62.11317 | 2.298861 | 0.40194 | 5.71942 | 1.07E-08 | 1.01E-07 | 4.920691 |
| GLRA4 | 24.41493 | 2.319721 | 0.975751 | 2.377371 | 0.017437 | 0.042211 | 4.992356 |
| NPY2R | 22.06554 | 2.327724 | 0.48094 | 4.839952 | 1.30E-06 | 8.38E-06 | 5.020129 |
| CNTFR | 405.7055 | 2.331071 | 0.14773 | 15.77922 | 4.33E-56 | 2.34E-53 | 5.031787 |
| WNT5B | 80.03519 | 2.339814 | 0.27895 | 8.387923 | 4.95E-17 | 1.46E-15 | 5.062372 |
| C6orf118 | 11.26589 | 2.340076 | 0.697841 | 3.353307 | 0.000799 | 0.002813 | 5.063295 |
| ASCL1 | 5.670581 | 2.348114 | 0.96651 | 2.429478 | 0.015121 | 0.037222 | 5.091581 |
| APLNR | 40.45242 | 2.351166 | 0.332151 | 7.078615 | 1.46E-12 | 2.47E-11 | 5.102365 |

| | | | | | | | |
|---|---|---|---|---|---|---|---|
| GBX2 | 86.68519 | 2.354194 | 0.289761 | 8.124604 | 4.49E-16 | 1.16E-14 | 5.113083 |
| MSX2 | 15.62445 | 2.36486 | 0.616875 | 3.833611 | 0.000126 | 0.000537 | 5.151027 |
| FZD1 | 201.5312 | 2.367073 | 0.215041 | 11.00753 | 3.51E-28 | 2.82E-26 | 5.158933 |
| RIMBP3B | 11.77142 | 2.378058 | 0.869258 | 2.735733 | 0.006224 | 0.017267 | 5.198367 |
| RHOU | 1352.206 | 2.379161 | 0.12048 | 19.74729 | 8.46E-87 | 2.22E-83 | 5.202341 |
| ANO3 | 31.27575 | 2.39329 | 0.592826 | 4.037086 | 5.41E-05 | 0.000252 | 5.25354 |
| UNC5C | 54.76507 | 2.401039 | 0.52709 | 4.55527 | 5.23E-06 | 2.99E-05 | 5.281832 |
| LRRC4C | 26.2793 | 2.420599 | 0.523927 | 4.620107 | 3.84E-06 | 2.25E-05 | 5.353934 |
| KIF26B | 37.52241 | 2.421012 | 0.475062 | 5.096203 | 3.47E-07 | 2.48E-06 | 5.355467 |
| RD3 | 7.618949 | 2.446027 | 0.871943 | 2.805258 | 0.005028 | 0.014333 | 5.449133 |
| MEI1 | 9.000612 | 2.457859 | 1.050757 | 2.339132 | 0.019329 | 0.046022 | 5.49401 |
| DKK1 | 31.27635 | 2.462239 | 0.479933 | 5.130377 | 2.89E-07 | 2.11E-06 | 5.510713 |
| FOXB2 | 17.64127 | 2.468067 | 0.584749 | 4.220729 | 2.44E-05 | 0.000122 | 5.533021 |
| LMO1 | 179.657 | 2.474011 | 0.617081 | 4.009218 | 6.09E-05 | 0.000281 | 5.555863 |
| SP8 | 516.9157 | 2.474524 | 0.129978 | 19.03803 | 8.26E-81 | 1.38E-77 | 5.557839 |
| LHX5 | 1690.508 | 2.482858 | 0.431573 | 5.753038 | 8.77E-09 | 8.33E-08 | 5.590039 |
| PLAGL1 | 97.22315 | 2.498437 | 0.236045 | 10.58457 | 3.51E-26 | 2.48E-24 | 5.650728 |
| WNT8B | 54.21681 | 2.528269 | 0.307985 | 8.209059 | 2.23E-16 | 6.01E-15 | 5.768793 |
| ATP10D | 22.72638 | 2.53493 | 0.646515 | 3.920915 | 8.82E-05 | 0.000392 | 5.795486 |
| NOS2 | 120.5263 | 2.547125 | 0.222646 | 11.44023 | 2.63E-30 | 2.52E-28 | 5.844685 |
| MECOM | 31.07577 | 2.550688 | 0.57099 | 4.46713 | 7.93E-06 | 4.38E-05 | 5.859137 |
| DCT | 12.77095 | 2.554656 | 0.744049 | 3.433451 | 0.000596 | 0.002174 | 5.875272 |
| FABP7 | 230.5176 | 2.577159 | 0.235493 | 10.94366 | 7.13E-28 | 5.60E-26 | 5.967635 |
| LRP2 | 1212.881 | 2.583673 | 0.433155 | 5.964783 | 2.45E-09 | 2.54E-08 | 5.994641 |
| FOXB1 | 570.4445 | 2.586073 | 0.283129 | 9.133892 | 6.61E-20 | 2.55E-18 | 6.004621 |
| AC005524 | 12.37616 | 2.597227 | 0.676824 | 3.837373 | 0.000124 | 0.000531 | 6.051223 |
| RGL1 | 147.8336 | 2.614657 | 0.216487 | 12.07765 | 1.39E-33 | 1.77E-31 | 6.124775 |
| PRTG | 2806.675 | 2.624413 | 0.195317 | 13.43668 | 3.69E-41 | 7.53E-39 | 6.166332 |
| FAT4 | 429.573 | 2.626969 | 0.20441 | 12.85149 | 8.44E-38 | 1.40E-35 | 6.177269 |
| KCNH5 | 36.83277 | 2.628326 | 0.522307 | 5.032145 | 4.85E-07 | 3.39E-06 | 6.183083 |
| WLS | 1031.267 | 2.630053 | 0.439854 | 5.97938 | 2.24E-09 | 2.34E-08 | 6.190489 |
| NKAIN3 | 9.595827 | 2.652208 | 0.866252 | 3.061703 | 0.002201 | 0.006917 | 6.286286 |
| FGF9 | 18.3483 | 2.656057 | 0.926976 | 2.865292 | 0.004166 | 0.012123 | 6.303081 |
| SP5 | 182.9619 | 2.68 | 0.303023 | 8.844223 | 9.22E-19 | 3.19E-17 | 6.408559 |
| MEIS2 | 28.52745 | 2.698241 | 0.745699 | 3.618407 | 0.000296 | 0.001162 | 6.490103 |
| DMRT3 | 6.969482 | 2.702222 | 0.959228 | 2.817081 | 0.004846 | 0.013864 | 6.508034 |
| CDH20 | 30.38697 | 2.707966 | 0.452573 | 5.983492 | 2.18E-09 | 2.28E-08 | 6.533997 |
| CDH10 | 19.61913 | 2.713913 | 0.646301 | 4.199146 | 2.68E-05 | 0.000133 | 6.560989 |
| COL2A1 | 570.7539 | 2.725295 | 0.208878 | 13.04731 | 6.58E-39 | 1.15E-36 | 6.612955 |
| CLEC19A | 9.940838 | 2.732973 | 0.816955 | 3.345316 | 0.000822 | 0.002888 | 6.648243 |
| ANXA1 | 56.37983 | 2.733391 | 0.316039 | 8.648892 | 5.20E-18 | 1.67E-16 | 6.650167 |
| TRH | 8.252503 | 2.735698 | 0.988339 | 2.767976 | 0.005641 | 0.015849 | 6.66081 |
| RELN | 12.32671 | 2.808552 | 0.689815 | 4.071455 | 4.67E-05 | 0.000221 | 7.005812 |
| LMX1A | 23.42152 | 2.863987 | 0.568604 | 5.036873 | 4.73E-07 | 3.31E-06 | 7.280246 |

| | | | | | | | |
|---|---|---|---|---|---|---|---|
| ILDR2 | 441.319 | 2.873971 | 0.501359 | 5.732361 | 9.90E-09 | 9.37E-08 | 7.330799 |
| LEF1 | 106.2409 | 2.937811 | 0.397046 | 7.399177 | 1.37E-13 | 2.64E-12 | 7.662479 |
| NPPB | 4.474483 | 2.940272 | 1.160868 | 2.532821 | 0.011315 | 0.028975 | 7.67556 |
| DCDC1 | 17.14961 | 2.942127 | 0.546478 | 5.383794 | 7.29E-08 | 5.97E-07 | 7.685437 |
| RGS4 | 5.11912 | 2.948646 | 1.238485 | 2.38085 | 0.017273 | 0.041881 | 7.72024 |
| EOMES | 20.58505 | 2.956592 | 0.610789 | 4.840611 | 1.29E-06 | 8.35E-06 | 7.762882 |
| GDF10 | 13.1369 | 2.971811 | 0.651365 | 4.562436 | 5.06E-06 | 2.90E-05 | 7.845202 |
| SMOC1 | 1013.275 | 2.985021 | 0.2623 | 11.38018 | 5.25E-30 | 4.87E-28 | 7.917367 |
| GASK1B | 7.682289 | 2.987844 | 0.982859 | 3.039953 | 0.002366 | 0.00737 | 7.932875 |
| HPSE2 | 49.81537 | 3.004747 | 0.364515 | 8.243146 | 1.68E-16 | 4.62E-15 | 8.026365 |
| BNIP3P11 | 3.948284 | 3.01319 | 1.202681 | 2.505395 | 0.012231 | 0.030994 | 8.073476 |
| ARHGAP24 | 8.786156 | 3.021382 | 0.877309 | 3.443921 | 0.000573 | 0.0021 | 8.119449 |
| LIX1 | 844.0082 | 3.032802 | 0.235066 | 12.9019 | 4.39E-38 | 7.47E-36 | 8.183978 |
| MYOM2 | 5.440304 | 3.03816 | 1.164124 | 2.609824 | 0.009059 | 0.02388 | 8.214429 |
| ADAMTS9 | 133.3471 | 3.084981 | 0.33296 | 9.265324 | 1.94E-20 | 7.93E-19 | 8.485389 |
| LGI1 | 220.5699 | 3.115528 | 0.363354 | 8.574352 | 9.96E-18 | 3.14E-16 | 8.66697 |
| MYO16 | 8.0503 | 3.1171 | 1.0354 | 3.010527 | 0.002608 | 0.008041 | 8.676423 |
| KIAA1755 | 6.688689 | 3.121076 | 0.899847 | 3.468451 | 0.000523 | 0.001933 | 8.700367 |
| EFCAB1 | 6.993381 | 3.163513 | 0.926686 | 3.413793 | 0.000641 | 0.002316 | 8.960091 |
| ELAVL4 | 22.18895 | 3.16382 | 0.598829 | 5.283345 | 1.27E-07 | 9.91E-07 | 8.961994 |
| FBLN7 | 100.0383 | 3.244437 | 0.326099 | 9.949243 | 2.54E-23 | 1.35E-21 | 9.477042 |
| RFX4 | 27.36703 | 3.26809 | 0.59272 | 5.513714 | 3.51E-08 | 3.05E-07 | 9.633699 |
| CCDC92 | 30.13567 | 3.278683 | 0.51695 | 6.342362 | 2.26E-10 | 2.80E-09 | 9.704696 |
| OTX1 | 281.3015 | 3.291859 | 0.228386 | 14.4136 | 4.25E-47 | 1.32E-44 | 9.793735 |
| NEUROG2 | 2.832689 | 3.320753 | 1.41203 | 2.351758 | 0.018685 | 0.044703 | 9.99186 |
| GREB1L | 4425.744 | 3.347608 | 0.365381 | 9.161966 | 5.10E-20 | 2.00E-18 | 10.17959 |
| AMER2 | 36.49688 | 3.382065 | 0.471818 | 7.168155 | 7.60E-13 | 1.33E-11 | 10.42565 |
| RLBP1 | 6.101038 | 3.394659 | 1.086938 | 3.123141 | 0.001789 | 0.005769 | 10.51706 |
| PRDM16 | 48.62301 | 3.405417 | 0.389912 | 8.733803 | 2.46E-18 | 8.17E-17 | 10.59577 |
| ZEB1 | 81.97893 | 3.427097 | 0.339066 | 10.10746 | 5.12E-24 | 2.96E-22 | 10.7562 |
| SRSF9P1 | 3.164645 | 3.44549 | 1.348052 | 2.555903 | 0.010591 | 0.027332 | 10.89421 |
| GLIS3 | 58.72014 | 3.463138 | 0.444798 | 7.785863 | 6.92E-15 | 1.56E-13 | 11.0283 |
| LGR5 | 20.80365 | 3.468082 | 0.617006 | 5.620827 | 1.90E-08 | 1.72E-07 | 11.06616 |
| CT75 | 20.09561 | 3.473333 | 0.890292 | 3.901341 | 9.57E-05 | 0.000422 | 11.1065 |
| GSC | 3.011403 | 3.476369 | 1.359893 | 2.556356 | 0.010577 | 0.0273 | 11.1299 |
| ZFHX4 | 560.5225 | 3.508804 | 0.166751 | 21.04221 | 2.70E-98 | 1.65E-94 | 11.38296 |
| NDP | 6.389262 | 3.514747 | 1.023665 | 3.433494 | 0.000596 | 0.002174 | 11.42995 |
| SFRP4 | 5.967097 | 3.541655 | 1.07625 | 3.290737 | 0.000999 | 0.003437 | 11.64513 |
| MESP1 | 4.545703 | 3.546075 | 1.17289 | 3.023366 | 0.0025 | 0.007739 | 11.68086 |
| VSX2 | 3.153016 | 3.54936 | 1.408026 | 2.520805 | 0.011709 | 0.029871 | 11.70749 |
| ZEB2 | 763.2854 | 3.563787 | 0.431324 | 8.262439 | 1.43E-16 | 3.97E-15 | 11.82515 |
| EBF2 | 3.16954 | 3.676581 | 1.437752 | 2.557173 | 0.010553 | 0.027251 | 12.78677 |
| MYT1 | 19.10954 | 3.68437 | 0.908903 | 4.053644 | 5.04E-05 | 0.000237 | 12.856 |
| AC138696 | 7.077395 | 3.689765 | 1.501756 | 2.456966 | 0.014012 | 0.034871 | 12.90416 |

| | | | | | | | |
|---|---|---|---|---|---|---|---|
| AL139011. | 10.56907 | 3.704208 | 1.449724 | 2.555112 | 0.010615 | 0.027386 | 13.034 |
| SIX3 | 449.8051 | 3.763371 | 0.288038 | 13.06555 | 5.18E-39 | 9.16E-37 | 13.57962 |
| ANKRD1 | 59.78972 | 3.774355 | 0.357204 | 10.56638 | 4.27E-26 | 2.97E-24 | 13.6834 |
| ENC1 | 2405.774 | 3.81839 | 0.195177 | 19.56373 | 3.15E-85 | 6.44E-82 | 14.1075 |
| WNT7B | 129.957 | 3.825479 | 0.301822 | 12.67462 | 8.18E-37 | 1.28E-34 | 14.17699 |
| PRRX1 | 34.86002 | 3.841225 | 0.630834 | 6.089123 | 1.14E-09 | 1.24E-08 | 14.33256 |
| SH2D1A | 2.308923 | 3.84638 | 1.650688 | 2.330168 | 0.019797 | 0.046961 | 14.38387 |
| ZBTB16 | 34.83882 | 3.954714 | 0.534605 | 7.397453 | 1.39E-13 | 2.67E-12 | 15.50556 |
| PAX3 | 38.79343 | 3.968194 | 0.479387 | 8.277643 | 1.26E-16 | 3.51E-15 | 15.65112 |
| GREM1 | 63.86567 | 3.973363 | 1.033763 | 3.843593 | 0.000121 | 0.000519 | 15.7073 |
| BAHCC1 | 243.7868 | 3.997951 | 0.297326 | 13.44637 | 3.23E-41 | 6.68E-39 | 15.97729 |
| DLK1 | 3167.606 | 4.115052 | 0.382978 | 10.74487 | 6.26E-27 | 4.64E-25 | 17.32823 |
| DACH1 | 136.5361 | 4.122966 | 0.336464 | 12.25381 | 1.60E-34 | 2.20E-32 | 17.42354 |
| FGF14 | 15.05063 | 4.174983 | 0.639266 | 6.5309 | 6.54E-11 | 8.83E-10 | 18.06322 |
| ARX | 8.830455 | 4.176072 | 0.952774 | 4.383067 | 1.17E-05 | 6.22E-05 | 18.07686 |
| DMRTC1B | 2.980253 | 4.205614 | 1.526427 | 2.755202 | 0.005866 | 0.016421 | 18.45083 |
| DBX2 | 4.82329 | 4.234707 | 1.272083 | 3.328955 | 0.000872 | 0.003048 | 18.82668 |
| ASXL3 | 17.45589 | 4.277894 | 0.705067 | 6.067356 | 1.30E-09 | 1.41E-08 | 19.39878 |
| GPR149 | 1.878629 | 4.323404 | 1.781105 | 2.427371 | 0.015209 | 0.037403 | 20.02048 |
| GABRR2 | 1.905388 | 4.341566 | 1.817905 | 2.388225 | 0.01693 | 0.041142 | 20.2741 |
| ZIC1 | 122.5251 | 4.351133 | 0.329356 | 13.21103 | 7.58E-40 | 1.42E-37 | 20.40899 |
| CYP4F22 | 1.960137 | 4.388774 | 1.627855 | 2.696047 | 0.007017 | 0.019131 | 20.94848 |
| RAX | 99.86741 | 4.441196 | 0.39243 | 11.31718 | 1.08E-29 | 9.67E-28 | 21.72367 |
| FAM181A | 2.174183 | 4.524284 | 1.652733 | 2.737456 | 0.006192 | 0.017195 | 23.01152 |
| IL1RAPL1 | 19.7962 | 4.535168 | 0.701293 | 6.466868 | 1.00E-10 | 1.31E-09 | 23.18578 |
| NR2F2 | 74.83602 | 4.570027 | 0.974063 | 4.691716 | 2.71E-06 | 1.64E-05 | 23.75282 |
| SPATA31A7 | 2.342281 | 4.6325 | 1.57963 | 2.93265 | 0.003361 | 0.010039 | 24.80399 |
| CNTNAP3C | 2.440387 | 4.699688 | 1.947285 | 2.413457 | 0.015802 | 0.038682 | 25.98645 |
| BMP5 | 14.21064 | 4.701185 | 0.870889 | 5.398143 | 6.73E-08 | 5.55E-07 | 26.01344 |
| NCKAP1L | 4.907219 | 4.733293 | 2.010708 | 2.354043 | 0.01857 | 0.044482 | 26.59886 |
| SOX1 | 102.8999 | 4.819818 | 0.374364 | 12.87468 | 6.25E-38 | 1.05E-35 | 28.24293 |
| DBX1 | 15.68984 | 4.861547 | 0.943993 | 5.149979 | 2.61E-07 | 1.92E-06 | 29.07176 |
| AL121594. | 19.61772 | 4.886069 | 1.070374 | 4.564825 | 5.00E-06 | 2.87E-05 | 29.57013 |
| SP9 | 2.762461 | 4.88748 | 1.511068 | 3.234455 | 0.001219 | 0.004091 | 29.59907 |
| ZNF503 | 73.952 | 4.991344 | 0.44173 | 11.29953 | 1.32E-29 | 1.17E-27 | 31.80857 |
| ZNF57 | 33.95075 | 5.02413 | 0.614798 | 8.172003 | 3.03E-16 | 7.99E-15 | 32.53972 |
| NTRK2 | 21.91673 | 5.02894 | 0.787286 | 6.387688 | 1.68E-10 | 2.13E-09 | 32.6484 |
| CEACAM21 | 7.146418 | 5.067027 | 1.429486 | 3.544651 | 0.000393 | 0.001497 | 33.52179 |
| NDUFA6 | 16.65983 | 5.086779 | 0.924911 | 5.499747 | 3.80E-08 | 3.28E-07 | 33.98389 |
| INSM2 | 3.285763 | 5.13506 | 1.433048 | 3.583313 | 0.000339 | 0.001313 | 35.14044 |
| POU3F3 | 13.14625 | 5.140057 | 0.962542 | 5.340084 | 9.29E-08 | 7.49E-07 | 35.26235 |
| RXRG | 5.74023 | 5.149861 | 1.251505 | 4.114935 | 3.87E-05 | 0.000186 | 35.50279 |
| LHX2 | 95.41669 | 5.178912 | 0.441006 | 11.7434 | 7.63E-32 | 8.66E-30 | 36.22494 |
| NEUROD4 | 3.498019 | 5.227217 | 1.425701 | 3.666419 | 0.000246 | 0.000979 | 37.45838 |

| | | | | | | | |
|---|---|---|---|---|---|---|---|
| ARHGAP15 | 34.12676 | 5.438219 | 0.74667 | 7.283296 | 3.26E-13 | 5.94E-12 | 43.35777 |
| SIX6 | 17.44485 | 5.454038 | 0.958827 | 5.688237 | 1.28E-08 | 1.19E-07 | 43.8358 |
| EN2 | 4.406856 | 5.557628 | 1.335868 | 4.160314 | 3.18E-05 | 0.000156 | 47.09912 |
| GAD2 | 7.395144 | 5.597603 | 1.217794 | 4.596512 | 4.30E-06 | 2.50E-05 | 48.42242 |
| EYA4 | 7.5807 | 5.66668 | 1.218038 | 4.652302 | 3.28E-06 | 1.95E-05 | 50.79731 |
| POU3F2 | 74.18 | 5.713994 | 0.500194 | 11.42356 | 3.19E-30 | 3.02E-28 | 52.49083 |
| TCIM | 5.052107 | 5.753238 | 1.286351 | 4.472524 | 7.73E-06 | 4.28E-05 | 53.9383 |
| PAX2 | 24.06664 | 5.784949 | 0.924366 | 6.258289 | 3.89E-10 | 4.64E-09 | 55.13701 |
| MAB21L2 | 28.5177 | 5.82875 | 0.796797 | 7.31523 | 2.57E-13 | 4.75E-12 | 56.83667 |
| LHX9 | 5.659422 | 5.916815 | 1.400394 | 4.225108 | 2.39E-05 | 0.00012 | 60.41416 |
| RSPO3 | 42.91921 | 6.172966 | 0.736027 | 8.386878 | 4.99E-17 | 1.47E-15 | 72.15194 |
| FOXG1 | 117.4301 | 6.183575 | 0.467234 | 13.23443 | 5.55E-40 | 1.06E-37 | 72.68444 |
| DMRTA2 | 11.78784 | 6.240142 | 1.178193 | 5.296367 | 1.18E-07 | 9.30E-07 | 75.59097 |
| NDST4 | 7.175034 | 6.261942 | 1.281996 | 4.884525 | 1.04E-06 | 6.80E-06 | 76.74187 |
| GUCY1A1 | 7.407168 | 6.293625 | 1.301707 | 4.834901 | 1.33E-06 | 8.56E-06 | 78.44582 |
| INMT-MIND | 7.798514 | 6.380284 | 1.719697 | 3.71012 | 0.000207 | 0.000839 | 83.30228 |
| DIPK1C | 8.772039 | 6.527019 | 1.247028 | 5.234062 | 1.66E-07 | 1.27E-06 | 92.2207 |
| PAX6 | 342.687 | 6.572143 | 1.202752 | 5.464252 | 4.65E-08 | 3.94E-07 | 95.15071 |
| TBR1 | 10.35166 | 6.691278 | 1.374792 | 4.867121 | 1.13E-06 | 7.38E-06 | 103.3417 |
| VAX1 | 12.83276 | 7.094164 | 1.140719 | 6.21903 | 5.00E-10 | 5.84E-09 | 136.6332 |
| FEZF2 | 52.34324 | 7.132259 | 0.879148 | 8.112691 | 4.95E-16 | 1.28E-14 | 140.2891 |
| RTL1 | 40.69649 | 7.333916 | 1.002499 | 7.315635 | 2.56E-13 | 4.74E-12 | 161.3351 |
| EMX2 | 61.71359 | 8.028519 | 0.978884 | 8.201707 | 2.37E-16 | 6.37E-15 | 261.1109 |
| ZIC4 | 27.83192 | 8.213984 | 1.081387 | 7.595787 | 3.06E-14 | 6.33E-13 | 296.931 |
| NR2F1 | 52.08224 | 8.489733 | 1.051182 | 8.076365 | 6.67E-16 | 1.69E-14 | 359.4714 |
| RSC1A1 | 35.52025 | 8.568184 | 1.078052 | 7.947842 | 1.90E-15 | 4.52E-14 | 379.5601 |
| HES5 | 54.13193 | 9.175656 | 1.053142 | 8.712646 | 2.97E-18 | 9.78E-17 | 578.2927 |
| MMRN1 | 56.36379 | 9.234375 | 1.048052 | 8.810986 | 1.24E-18 | 4.23E-17 | 602.3153 |
| AL035078. | 178.9692 | 10.90147 | 1.0349 | 10.53385 | 6.03E-26 | 4.12E-24 | 1912.805 |

**DOWN - p<.05, FC<.75**

| GeneID | Base mear | log2(FC) | StdErr | Wald-Stats | P-value | P-adj | FC |
|---|---|---|---|---|---|---|---|
| ZNF544 | 678.4107 | -5.50244 | 0.170029 | -32.3618 | 9.45E-230 | 1.74E-225 | 0.02206 |
| HHLA1 | 15570 | -2.21798 | 0.092092 | -24.0844 | 3.64E-128 | 3.35E-124 | 0.214942 |
| SERPINB9 | 2671.655 | -2.13354 | 0.101831 | -20.9518 | 1.81E-97 | 8.31E-94 | 0.227897 |
| RAB17 | 1096.015 | -2.71132 | 0.131052 | -20.6889 | 4.36E-95 | 1.60E-91 | 0.15269 |
| CYP2S1 | 6035.196 | -1.73661 | 0.086162 | -20.1551 | 2.43E-90 | 7.44E-87 | 0.300075 |
| HAPLN3 | 1543.625 | -2.32079 | 0.118116 | -19.6484 | 5.96E-86 | 1.37E-82 | 0.200158 |
| LRRC61 | 363.3605 | -2.63182 | 0.137322 | -19.1653 | 7.22E-82 | 1.33E-78 | 0.161341 |
| BCAN | 349.8487 | -2.87641 | 0.153906 | -18.6894 | 6.04E-78 | 9.25E-75 | 0.136181 |
| CLN8 | 2213.385 | -2.14243 | 0.117095 | -18.2966 | 8.82E-75 | 1.25E-71 | 0.226498 |
| GAL | 673.7821 | -2.0265 | 0.112568 | -18.0024 | 1.87E-72 | 2.45E-69 | 0.24545 |
| IDO1 | 526.1102 | -2.49848 | 0.140215 | -17.8189 | 5.04E-71 | 5.79E-68 | 0.176963 |
| BICDL2 | 431.0824 | -2.50415 | 0.144235 | -17.3616 | 1.61E-67 | 1.65E-64 | 0.176268 |
| PRSS8 | 3048.82 | -1.28545 | 0.074332 | -17.2935 | 5.27E-67 | 5.10E-64 | 0.410242 |
| CDH1 | 7332.422 | -1.38475 | 0.080151 | -17.2768 | 7.04E-67 | 6.47E-64 | 0.382957 |
| CDH3 | 3365.481 | -1.89569 | 0.109802 | -17.2647 | 8.68E-67 | 7.60E-64 | 0.268745 |
| CHGA | 1438.494 | -2.26763 | 0.133491 | -16.9871 | 1.02E-64 | 8.55E-62 | 0.207671 |
| NFKB2 | 1312.964 | -1.66795 | 0.098335 | -16.9619 | 1.57E-64 | 1.20E-61 | 0.3147 |
| ECEL1P1 | 471.0597 | -2.87201 | 0.170568 | -16.8379 | 1.29E-63 | 9.47E-61 | 0.136597 |
| PIM2 | 2478.72 | -1.62167 | 0.098777 | -16.4174 | 1.43E-60 | 9.48E-58 | 0.324959 |
| TNFRSF12/ | 1162.777 | -1.65167 | 0.100607 | -16.4171 | 1.44E-60 | 9.48E-58 | 0.318271 |
| LRRK1 | 66194.95 | -1.85417 | 0.113586 | -16.3239 | 6.67E-60 | 4.23E-57 | 0.276593 |
| GPC4 | 12684.04 | -1.156 | 0.072511 | -15.9423 | 3.22E-57 | 1.97E-54 | 0.448756 |
| MMP25 | 1240.854 | -1.48566 | 0.093599 | -15.8725 | 9.82E-57 | 5.66E-54 | 0.357085 |
| PLPP2 | 590.1901 | -1.73648 | 0.109403 | -15.8723 | 9.85E-57 | 5.66E-54 | 0.300101 |
| B3GNT7 | 1622.402 | -1.57284 | 0.099649 | -15.7838 | 4.02E-56 | 2.24E-53 | 0.336145 |
| PDLIM1 | 2217.975 | -0.94187 | 0.061657 | -15.2761 | 1.10E-52 | 5.20E-50 | 0.520557 |
| NMRK2 | 1232.109 | -1.08502 | 0.071131 | -15.254 | 1.55E-52 | 7.12E-50 | 0.471385 |
| COL23A1 | 255.7575 | -2.07454 | 0.136975 | -15.1454 | 8.12E-52 | 3.56E-49 | 0.237411 |
| C13orf42 | 1951.915 | -1.67309 | 0.110928 | -15.0826 | 2.11E-51 | 8.80E-49 | 0.313582 |
| TNFRSF8 | 381.1334 | -2.06201 | 0.137831 | -14.9605 | 1.33E-50 | 5.32E-48 | 0.239482 |
| GRM4 | 892.9114 | -1.52322 | 0.102392 | -14.8763 | 4.70E-50 | 1.80E-47 | 0.347908 |
| USP9X | 23078.85 | -0.85902 | 0.057868 | -14.8444 | 7.56E-50 | 2.78E-47 | 0.551328 |
| SPINT1 | 3194.075 | -1.29749 | 0.087505 | -14.8276 | 9.72E-50 | 3.50E-47 | 0.406834 |
| COL6A1 | 3114.506 | -2.0194 | 0.137282 | -14.7099 | 5.57E-49 | 1.97E-46 | 0.246661 |
| FAM20A | 1035.039 | -2.62097 | 0.17826 | -14.7031 | 6.16E-49 | 2.14E-46 | 0.162559 |
| L1CAM | 975.9409 | -1.67876 | 0.114888 | -14.6121 | 2.35E-48 | 8.00E-46 | 0.312351 |
| LAMC3 | 466.8789 | -3.00138 | 0.207066 | -14.4948 | 1.31E-47 | 4.22E-45 | 0.12488 |
| COL6A2 | 1935.146 | -1.89837 | 0.131498 | -14.4365 | 3.05E-47 | 9.66E-45 | 0.268247 |
| VCAN | 7004.752 | -1.09069 | 0.076089 | -14.3344 | 1.33E-46 | 4.09E-44 | 0.469538 |
| PKP3 | 213.7895 | -2.93689 | 0.205468 | -14.2936 | 2.40E-46 | 7.22E-44 | 0.13059 |
| ESPN | 335.8288 | -1.5873 | 0.11124 | -14.2691 | 3.41E-46 | 1.01E-43 | 0.332794 |
| MMP24 | 1140.368 | -1.35624 | 0.09548 | -14.2044 | 8.60E-46 | 2.51E-43 | 0.390599 |

| | | | | | | | |
|---|---|---|---|---|---|---|---|
| FXYD5 | 1418.954 | -1.23061 | 0.087211 | -14.1107 | 3.26E-45 | 9.15E-43 | 0.426137 |
| TENT5B | 1829.607 | -1.12677 | 0.080008 | -14.0832 | 4.82E-45 | 1.32E-42 | 0.45794 |
| CRYBG1 | 535.5703 | -1.62942 | 0.11585 | -14.0649 | 6.24E-45 | 1.69E-42 | 0.323219 |
| RARRES2 | 3256.3 | -1.74719 | 0.124705 | -14.0106 | 1.34E-44 | 3.58E-42 | 0.297882 |
| PHC1 | 12691.15 | -1.11577 | 0.079759 | -13.9894 | 1.81E-44 | 4.75E-42 | 0.461444 |
| LAD1 | 476.8818 | -1.48585 | 0.107673 | -13.7996 | 2.56E-43 | 6.37E-41 | 0.357039 |
| EZR | 6356.415 | -0.91242 | 0.066287 | -13.7648 | 4.15E-43 | 1.02E-40 | 0.531292 |
| TMEM42 | 109.2671 | -4.05544 | 0.294651 | -13.7635 | 4.22E-43 | 1.02E-40 | 0.060144 |
| GRIN2D | 381.1855 | -2.11177 | 0.153511 | -13.7565 | 4.65E-43 | 1.11E-40 | 0.231362 |
| SEMA4A | 391.2433 | -1.83275 | 0.133372 | -13.7416 | 5.72E-43 | 1.35E-40 | 0.280729 |
| TFPI | 709.53 | -1.46229 | 0.106951 | -13.6725 | 1.48E-42 | 3.45E-40 | 0.362917 |
| ABHD12B | 934.1934 | -1.82826 | 0.134739 | -13.5689 | 6.12E-42 | 1.37E-39 | 0.281604 |
| GADD45G | 193.5349 | -2.97217 | 0.219073 | -13.5671 | 6.28E-42 | 1.39E-39 | 0.127435 |
| ITPR3 | 2391.819 | -1.0609 | 0.078283 | -13.5521 | 7.70E-42 | 1.69E-39 | 0.479333 |
| FAM189A2 | 307.2073 | -2.29032 | 0.169341 | -13.5249 | 1.11E-41 | 2.38E-39 | 0.20443 |
| IL27RA | 532.067 | -1.52136 | 0.113092 | -13.4524 | 2.98E-41 | 6.22E-39 | 0.348357 |
| CLDN7 | 1305.933 | -1.93703 | 0.145211 | -13.3394 | 1.37E-40 | 2.70E-38 | 0.261154 |
| EPHX1 | 879.9262 | -1.65555 | 0.12417 | -13.333 | 1.49E-40 | 2.91E-38 | 0.317416 |
| SMIM10L2 | 739.0909 | -1.38842 | 0.104674 | -13.2643 | 3.73E-40 | 7.21E-38 | 0.381982 |
| TUBA4A | 557.8691 | -1.61475 | 0.122182 | -13.216 | 7.09E-40 | 1.34E-37 | 0.32652 |
| OGDHL | 707.2678 | -1.19749 | 0.090892 | -13.1748 | 1.23E-39 | 2.27E-37 | 0.436033 |
| TMEM200E | 214.7939 | -1.99411 | 0.151725 | -13.143 | 1.87E-39 | 3.43E-37 | 0.251023 |
| GPR176 | 3597.047 | -1.03889 | 0.079084 | -13.1365 | 2.03E-39 | 3.70E-37 | 0.486702 |
| GLB1L3 | 593.8344 | -2.06925 | 0.157712 | -13.1204 | 2.52E-39 | 4.53E-37 | 0.238283 |
| CALB1 | 1465.98 | -1.77949 | 0.137491 | -12.9426 | 2.59E-38 | 4.45E-36 | 0.291287 |
| SCGB3A2 | 1842.892 | -1.78114 | 0.138446 | -12.8653 | 7.06E-38 | 1.18E-35 | 0.290953 |
| MFGE8 | 19848.71 | -1.56876 | 0.12311 | -12.7428 | 3.42E-37 | 5.47E-35 | 0.337098 |
| AC007326 | 2772.936 | -1.84396 | 0.144841 | -12.7309 | 3.98E-37 | 6.31E-35 | 0.278557 |
| CPXM2 | 398.4183 | -1.73313 | 0.136777 | -12.6713 | 8.53E-37 | 1.33E-34 | 0.300798 |
| CCN1 | 1193.539 | -1.1695 | 0.092649 | -12.6229 | 1.58E-36 | 2.36E-34 | 0.444574 |
| ITM2C | 9122.456 | -0.94459 | 0.075355 | -12.5353 | 4.79E-36 | 6.98E-34 | 0.519576 |
| SLC38A5 | 700.899 | -1.60814 | 0.128892 | -12.4767 | 1.00E-35 | 1.44E-33 | 0.32802 |
| ATP1A3 | 613.2251 | -1.50598 | 0.121244 | -12.421 | 2.01E-35 | 2.86E-33 | 0.352091 |
| TDGF1 | 5845.027 | -1.77755 | 0.14354 | -12.3836 | 3.20E-35 | 4.53E-33 | 0.291678 |
| FOXH1 | 1974.336 | -1.48887 | 0.120759 | -12.3293 | 6.30E-35 | 8.84E-33 | 0.356291 |
| RPL39L | 1376.864 | -1.91924 | 0.156001 | -12.3028 | 8.75E-35 | 1.22E-32 | 0.264394 |
| EEF1A2 | 706.1993 | -1.49392 | 0.12206 | -12.2392 | 1.92E-34 | 2.61E-32 | 0.355047 |
| ADAMTS8 | 246.565 | -2.02363 | 0.166036 | -12.1879 | 3.60E-34 | 4.87E-32 | 0.245938 |
| AP1M2 | 1672.923 | -1.68455 | 0.138332 | -12.1776 | 4.09E-34 | 5.49E-32 | 0.311099 |
| EPCAM | 4963.593 | -1.28197 | 0.105369 | -12.1665 | 4.69E-34 | 6.24E-32 | 0.411234 |
| SLC7A3 | 4773.524 | -0.86622 | 0.071491 | -12.1165 | 8.63E-34 | 1.13E-31 | 0.548581 |
| SLC1A5 | 12685.3 | -0.85906 | 0.071058 | -12.0895 | 1.20E-33 | 1.55E-31 | 0.551312 |
| EPHX3 | 272.8688 | -1.64907 | 0.136759 | -12.0582 | 1.76E-33 | 2.23E-31 | 0.318846 |
| CNTN1 | 329.443 | -2.85192 | 0.236628 | -12.0523 | 1.89E-33 | 2.37E-31 | 0.138512 |

| | | | | | | | |
|---|---|---|---|---|---|---|---|
| AC073111 | 482.567 | -1.21857 | 0.101112 | -12.0517 | 1.90E-33 | 2.38E-31 | 0.42971 |
| AP002956. | 655.0298 | -1.47586 | 0.122764 | -12.0219 | 2.73E-33 | 3.36E-31 | 0.35952 |
| ZNF718 | 121.0173 | -7.44303 | 0.621376 | -11.9783 | 4.62E-33 | 5.66E-31 | 0.005747 |
| TRNP1 | 628.492 | -1.45374 | 0.122305 | -11.8861 | 1.40E-32 | 1.67E-30 | 0.365075 |
| IFITM1 | 2328.606 | -1.37109 | 0.115359 | -11.8854 | 1.41E-32 | 1.67E-30 | 0.386599 |
| RELL2 | 429.155 | -1.44342 | 0.121761 | -11.8546 | 2.04E-32 | 2.39E-30 | 0.367695 |
| KLHDC7A | 1033.103 | -1.14075 | 0.096324 | -11.8428 | 2.34E-32 | 2.71E-30 | 0.453524 |
| SLC30A1 | 730.1538 | -2.08685 | 0.177777 | -11.7386 | 8.09E-32 | 9.12E-30 | 0.235395 |
| CXCL5 | 360.6726 | -2.00045 | 0.170956 | -11.7015 | 1.25E-31 | 1.38E-29 | 0.249922 |
| USP43 | 167.6477 | -1.8399 | 0.157958 | -11.6481 | 2.35E-31 | 2.57E-29 | 0.279341 |
| TMEM63A | 726.9652 | -1.52186 | 0.130722 | -11.642 | 2.52E-31 | 2.73E-29 | 0.348236 |
| PCOLCE | 699.5104 | -1.57058 | 0.135271 | -11.6106 | 3.64E-31 | 3.89E-29 | 0.336674 |
| CHPF | 1391.262 | -1.20302 | 0.103626 | -11.6093 | 3.70E-31 | 3.91E-29 | 0.434364 |
| ANO9 | 880.7377 | -2.88818 | 0.248949 | -11.6015 | 4.05E-31 | 4.25E-29 | 0.135074 |
| GABRP | 1251.825 | -1.62809 | 0.140513 | -11.5868 | 4.81E-31 | 4.99E-29 | 0.323517 |
| SCARF2 | 117.0053 | -2.25402 | 0.194719 | -11.5758 | 5.47E-31 | 5.65E-29 | 0.209639 |
| PDZD4 | 1902.921 | -1.58936 | 0.137697 | -11.5424 | 8.07E-31 | 8.15E-29 | 0.33232 |
| ZDHHC22 | 494.047 | -1.93464 | 0.168104 | -11.5086 | 1.19E-30 | 1.20E-28 | 0.261587 |
| FBXL16 | 290.0759 | -1.54765 | 0.134674 | -11.4918 | 1.45E-30 | 1.45E-28 | 0.342068 |
| RRBP1 | 3004.223 | -0.91763 | 0.079857 | -11.491 | 1.46E-30 | 1.45E-28 | 0.529377 |
| C9orf64 | 157.7848 | -7.91773 | 0.689589 | -11.4818 | 1.63E-30 | 1.61E-28 | 0.004135 |
| ATP2A3 | 748.8405 | -1.68178 | 0.146648 | -11.4681 | 1.91E-30 | 1.87E-28 | 0.311699 |
| ANXA2 | 2370.973 | -0.85001 | 0.074141 | -11.4648 | 1.98E-30 | 1.94E-28 | 0.554781 |
| MAD2L2 | 3578.697 | -0.8899 | 0.077637 | -11.4624 | 2.04E-30 | 1.98E-28 | 0.53965 |
| COL6A3 | 223.3986 | -1.84221 | 0.161644 | -11.3968 | 4.34E-30 | 4.09E-28 | 0.278894 |
| SCNN1A | 2255.135 | -1.56941 | 0.137719 | -11.3957 | 4.39E-30 | 4.10E-28 | 0.336946 |
| ST14 | 851.6385 | -1.43409 | 0.126143 | -11.3688 | 5.98E-30 | 5.49E-28 | 0.370079 |
| ACE | 102.1044 | -2.8027 | 0.247142 | -11.3404 | 8.27E-30 | 7.53E-28 | 0.143319 |
| EPHA1 | 3177.08 | -1.48595 | 0.131103 | -11.3342 | 8.88E-30 | 8.00E-28 | 0.357014 |
| OLFM2 | 1984.083 | -1.0778 | 0.095423 | -11.295 | 1.39E-29 | 1.23E-27 | 0.473751 |
| STC2 | 6314.83 | -0.74795 | 0.06631 | -11.2797 | 1.65E-29 | 1.45E-27 | 0.595447 |
| SLC43A1 | 1116.702 | -1.48235 | 0.131551 | -11.2682 | 1.88E-29 | 1.65E-27 | 0.357907 |
| CNMD | 2216.541 | -1.85793 | 0.165769 | -11.2079 | 3.73E-29 | 3.23E-27 | 0.275873 |
| IGSF8 | 1363.932 | -1.53114 | 0.137102 | -11.1679 | 5.86E-29 | 5.05E-27 | 0.346003 |
| KCND1 | 333.4374 | -1.711 | 0.153393 | -11.1544 | 6.82E-29 | 5.86E-27 | 0.305447 |
| ZNF677 | 77.5047 | -6.54318 | 0.587418 | -11.1389 | 8.11E-29 | 6.94E-27 | 0.010723 |
| FUT1 | 604.0841 | -0.94264 | 0.08473 | -11.1252 | 9.46E-29 | 8.05E-27 | 0.520281 |
| ZBED6CL | 177.6412 | -2.91028 | 0.261754 | -11.1184 | 1.02E-28 | 8.65E-27 | 0.133021 |
| PLPP1 | 989.1083 | -1.2957 | 0.116882 | -11.0856 | 1.47E-28 | 1.23E-26 | 0.407339 |
| TSPYL5 | 246.074 | -11.4115 | 1.030071 | -11.0784 | 1.60E-28 | 1.33E-26 | 0.000367 |
| KIAA1522 | 2777.785 | -0.81905 | 0.073996 | -11.0688 | 1.78E-28 | 1.47E-26 | 0.566814 |
| CCN2 | 1192.988 | -0.93318 | 0.084391 | -11.0578 | 2.01E-28 | 1.65E-26 | 0.523704 |
| ADM | 1045.134 | -1.58866 | 0.143839 | -11.0447 | 2.32E-28 | 1.90E-26 | 0.332481 |
| CD74 | 1135.463 | -1.33431 | 0.120857 | -11.0404 | 2.44E-28 | 1.98E-26 | 0.396581 |

| | | | | | | | |
|---|---|---|---|---|---|---|---|
| ADRA2A | 309.9062 | -1.59569 | 0.144706 | -11.0271 | 2.83E-28 | 2.29E-26 | 0.330863 |
| SPINK5 | 166.9238 | -4.14969 | 0.376825 | -11.0123 | 3.34E-28 | 2.69E-26 | 0.05634 |
| FOXO1 | 1047.687 | -1.25865 | 0.114733 | -10.9703 | 5.31E-28 | 4.24E-26 | 0.417934 |
| RAP1GAP2 | 972.681 | -1.13399 | 0.103857 | -10.9188 | 9.37E-28 | 7.33E-26 | 0.455653 |
| MATK | 358.3627 | -1.43319 | 0.131422 | -10.9052 | 1.09E-27 | 8.40E-26 | 0.370311 |
| SSH3 | 551.5755 | -1.19777 | 0.109917 | -10.8971 | 1.19E-27 | 9.15E-26 | 0.435947 |
| ABCB6 | 783.1053 | -1.87511 | 0.172097 | -10.8956 | 1.21E-27 | 9.26E-26 | 0.272606 |
| FOXD3 | 242.6102 | -2.25374 | 0.20712 | -10.8813 | 1.41E-27 | 1.07E-25 | 0.209681 |
| SLC3A2 | 11530.26 | -0.73801 | 0.067829 | -10.8804 | 1.43E-27 | 1.08E-25 | 0.599566 |
| TMEM30B | 481.9094 | -1.21401 | 0.112503 | -10.791 | 3.80E-27 | 2.83E-25 | 0.431068 |
| FLT1 | 1990.257 | -1.56242 | 0.146224 | -10.6851 | 1.20E-26 | 8.83E-25 | 0.338584 |
| SMPDL3B | 1113.89 | -1.76502 | 0.16539 | -10.6718 | 1.38E-26 | 1.01E-24 | 0.294223 |
| ZNF296 | 150.6173 | -1.6689 | 0.157049 | -10.6266 | 2.24E-26 | 1.62E-24 | 0.314493 |
| PALM3 | 608.1817 | -1.35327 | 0.127669 | -10.5999 | 2.98E-26 | 2.15E-24 | 0.391403 |
| GSTP1 | 10628.04 | -0.55182 | 0.05209 | -10.5937 | 3.19E-26 | 2.27E-24 | 0.682158 |
| CDCP1 | 568.7899 | -1.16402 | 0.10995 | -10.5868 | 3.43E-26 | 2.44E-24 | 0.446268 |
| LIMD2 | 1181.915 | -0.82943 | 0.078432 | -10.5752 | 3.88E-26 | 2.72E-24 | 0.56275 |
| DHRS3 | 393.3709 | -2.13822 | 0.202356 | -10.5666 | 4.25E-26 | 2.97E-24 | 0.227159 |
| ACAP1 | 395.7677 | -1.7637 | 0.166995 | -10.5614 | 4.50E-26 | 3.11E-24 | 0.294491 |
| TFE3 | 2296.984 | -1.54309 | 0.146514 | -10.532 | 6.15E-26 | 4.19E-24 | 0.343149 |
| APOBEC3E | 461.1286 | -1.47813 | 0.140576 | -10.5149 | 7.38E-26 | 5.00E-24 | 0.358953 |
| PAPSS2 | 562.0929 | -1.00968 | 0.096562 | -10.4564 | 1.37E-25 | 9.16E-24 | 0.496656 |
| KCNK5 | 1094.13 | -1.11075 | 0.107047 | -10.3763 | 3.18E-25 | 2.08E-23 | 0.463053 |
| ALPK3 | 1011.738 | -1.61585 | 0.155907 | -10.3642 | 3.61E-25 | 2.34E-23 | 0.326273 |
| CYBA | 2209.801 | -1.58189 | 0.152834 | -10.3504 | 4.17E-25 | 2.69E-23 | 0.334043 |
| C19orf66 | 439.837 | -1.33177 | 0.128713 | -10.3468 | 4.33E-25 | 2.78E-23 | 0.39728 |
| SCG3 | 946.9921 | -0.98464 | 0.095189 | -10.344 | 4.45E-25 | 2.85E-23 | 0.505351 |
| ZSCAN10 | 3412.371 | -1.05327 | 0.102482 | -10.2777 | 8.89E-25 | 5.61E-23 | 0.481874 |
| RASGRP2 | 340.8884 | -2.18143 | 0.212803 | -10.2509 | 1.17E-24 | 7.33E-23 | 0.220458 |
| AC092299 | 120.5165 | -3.86546 | 0.377593 | -10.2371 | 1.35E-24 | 8.37E-23 | 0.068609 |
| SLC44A3-A | 658.5046 | -1.43902 | 0.140728 | -10.2256 | 1.52E-24 | 9.40E-23 | 0.368818 |
| SLC22A31 | 157.1973 | -2.06312 | 0.202178 | -10.2045 | 1.89E-24 | 1.16E-22 | 0.239298 |
| APELA | 1547.165 | -1.52676 | 0.149627 | -10.2037 | 1.91E-24 | 1.16E-22 | 0.347056 |
| GSTT2B | 71.53446 | -4.20349 | 0.413105 | -10.1754 | 2.55E-24 | 1.54E-22 | 0.054278 |
| HSD11B2 | 313.6299 | -1.5203 | 0.149477 | -10.1708 | 2.68E-24 | 1.61E-22 | 0.348613 |
| ADAMTSL4 | 207.1469 | -2.46328 | 0.242733 | -10.1481 | 3.38E-24 | 2.00E-22 | 0.181334 |
| INHBE | 2039.608 | -1.51218 | 0.149065 | -10.1444 | 3.51E-24 | 2.07E-22 | 0.350581 |
| PYCARD | 354.4406 | -1.33333 | 0.13175 | -10.1202 | 4.50E-24 | 2.64E-22 | 0.396852 |
| COL1A1 | 6749.423 | -1.3922 | 0.137614 | -10.1167 | 4.66E-24 | 2.73E-22 | 0.380983 |
| MAPK13 | 426.7715 | -1.69988 | 0.168151 | -10.1093 | 5.03E-24 | 2.91E-22 | 0.307812 |
| SCARB1 | 856.5506 | -1.23614 | 0.122377 | -10.1011 | 5.46E-24 | 3.15E-22 | 0.424508 |
| STRA6 | 345.9426 | -1.22282 | 0.121079 | -10.0993 | 5.56E-24 | 3.20E-22 | 0.428445 |
| CPZ | 743.193 | -1.67537 | 0.165928 | -10.097 | 5.69E-24 | 3.26E-22 | 0.313085 |
| CBSL | 414.3765 | -2.18159 | 0.216223 | -10.0895 | 6.15E-24 | 3.50E-22 | 0.220433 |

| | | | | | | | |
|---|---|---|---|---|---|---|---|
| ITM2A | 278.244 | -1.51166 | 0.149865 | -10.0868 | 6.32E-24 | 3.59E-22 | 0.350708 |
| FLNB | 7508.133 | -1.84889 | 0.18339 | -10.0817 | 6.66E-24 | 3.76E-22 | 0.277607 |
| KRT19 | 2439.717 | -0.7123 | 0.070661 | -10.0805 | 6.74E-24 | 3.80E-22 | 0.610347 |
| FAM241A | 1360.641 | -0.88747 | 0.088262 | -10.0549 | 8.74E-24 | 4.82E-22 | 0.540562 |
| PCDHA10 | 238.527 | -2.4273 | 0.241873 | -10.0354 | 1.07E-23 | 5.79E-22 | 0.185913 |
| THY1 | 4729.259 | -1.39586 | 0.139198 | -10.0279 | 1.15E-23 | 6.23E-22 | 0.380017 |
| STMN3 | 1136.725 | -1.04295 | 0.104841 | -9.94799 | 2.57E-23 | 1.37E-21 | 0.485333 |
| RHBDL1 | 165.3637 | -2.45006 | 0.246565 | -9.93676 | 2.88E-23 | 1.53E-21 | 0.183003 |
| CAPN1 | 3469.792 | -0.73268 | 0.073843 | -9.9221 | 3.34E-23 | 1.76E-21 | 0.601784 |
| ECHDC2 | 274.2536 | -1.43635 | 0.144904 | -9.91238 | 3.68E-23 | 1.93E-21 | 0.369501 |
| SH2D3A | 282.3657 | -2.53127 | 0.255753 | -9.89735 | 4.27E-23 | 2.22E-21 | 0.172986 |
| PPIC | 198.5701 | -1.84244 | 0.186521 | -9.87791 | 5.19E-23 | 2.69E-21 | 0.27885 |
| HSPB8 | 391.948 | -1.70312 | 0.172668 | -9.86359 | 5.99E-23 | 3.07E-21 | 0.307121 |
| PFN1 | 18424.26 | -0.63973 | 0.064955 | -9.84894 | 6.93E-23 | 3.53E-21 | 0.641832 |
| MTRNR2L8 | 29423.6 | -1.55809 | 0.158268 | -9.84463 | 7.23E-23 | 3.67E-21 | 0.339601 |
| IGFBP4 | 2336.153 | -0.92959 | 0.094599 | -9.82667 | 8.64E-23 | 4.34E-21 | 0.525008 |
| HAGHL | 390.0762 | -1.83147 | 0.186718 | -9.80878 | 1.03E-22 | 5.14E-21 | 0.280978 |
| TMEM151E | 351.8928 | -1.28347 | 0.130953 | -9.80096 | 1.12E-22 | 5.52E-21 | 0.410806 |
| CUZD1 | 365.5338 | -1.57988 | 0.161384 | -9.7896 | 1.25E-22 | 6.15E-21 | 0.33451 |
| RASGRF2 | 230.3457 | -2.25733 | 0.23084 | -9.7788 | 1.39E-22 | 6.75E-21 | 0.209158 |
| OBSL1 | 3140.593 | -1.17 | 0.120271 | -9.72802 | 2.29E-22 | 1.09E-20 | 0.444422 |
| CH25H | 92.47537 | -4.27902 | 0.439951 | -9.72613 | 2.33E-22 | 1.11E-20 | 0.051509 |
| RGL3 | 832.1394 | -2.07192 | 0.213412 | -9.70857 | 2.77E-22 | 1.31E-20 | 0.237842 |
| MFSD2A | 247.0836 | -1.30121 | 0.134078 | -9.70484 | 2.88E-22 | 1.36E-20 | 0.405786 |
| ACP5 | 102.8957 | -2.2724 | 0.234804 | -9.67788 | 3.74E-22 | 1.75E-20 | 0.206985 |
| CAVIN3 | 433.7213 | -1.01451 | 0.105221 | -9.64165 | 5.33E-22 | 2.48E-20 | 0.494997 |
| PFKP | 2488.32 | -0.78638 | 0.081693 | -9.62611 | 6.20E-22 | 2.86E-20 | 0.579796 |
| S100A10 | 469.7648 | -1.32465 | 0.137685 | -9.62087 | 6.53E-22 | 3.00E-20 | 0.399247 |
| FAM24B | 57.65304 | -4.78313 | 0.497761 | -9.60929 | 7.30E-22 | 3.35E-20 | 0.036319 |
| LAMC2 | 462.8058 | -1.3791 | 0.143611 | -9.60303 | 7.76E-22 | 3.54E-20 | 0.38446 |
| SCG2 | 172.4519 | -1.84375 | 0.192257 | -9.59001 | 8.81E-22 | 4.01E-20 | 0.278597 |
| MT1X | 170.768 | -3.29927 | 0.344286 | -9.58296 | 9.43E-22 | 4.27E-20 | 0.101583 |
| KREMEN2 | 517.6155 | -2.05559 | 0.214692 | -9.57461 | 1.02E-21 | 4.61E-20 | 0.24055 |
| CGNL1 | 2252.147 | -1.08529 | 0.113365 | -9.57338 | 1.03E-21 | 4.64E-20 | 0.471298 |
| LEFTY1 | 70.81171 | -3.01351 | 0.314996 | -9.56683 | 1.10E-21 | 4.93E-20 | 0.123835 |
| UTF1 | 76.42497 | -2.38468 | 0.249427 | -9.56062 | 1.17E-21 | 5.22E-20 | 0.191487 |
| NPR1 | 387.3516 | -1.14383 | 0.119648 | -9.55995 | 1.18E-21 | 5.23E-20 | 0.452556 |
| ECEL1P2 | 271.2223 | -2.17424 | 0.228799 | -9.50285 | 2.04E-21 | 8.94E-20 | 0.221558 |
| TMEM8A | 2891.604 | -0.91336 | 0.096204 | -9.49402 | 2.22E-21 | 9.70E-20 | 0.530947 |
| FAM83F | 478.4103 | -1.18903 | 0.125553 | -9.47028 | 2.79E-21 | 1.21E-19 | 0.438599 |
| ITGB5 | 3349.013 | -0.83867 | 0.088683 | -9.45689 | 3.17E-21 | 1.38E-19 | 0.55916 |
| TDRP | 1209.12 | -0.83938 | 0.088837 | -9.44857 | 3.43E-21 | 1.49E-19 | 0.558884 |
| SLC17A9 | 374.2348 | -2.02766 | 0.214914 | -9.43476 | 3.92E-21 | 1.69E-19 | 0.245253 |
| SYT2 | 240.8388 | -1.72744 | 0.18336 | -9.42099 | 4.47E-21 | 1.91E-19 | 0.301988 |

| | | | | | | | |
|---|---|---|---|---|---|---|---|
| FBXO2 | 924.5771 | -1.05662 | 0.112185 | -9.4185 | 4.58E-21 | 1.95E-19 | 0.480758 |
| MUC3A | 432.3077 | -1.25472 | 0.133382 | -9.40699 | 5.11E-21 | 2.17E-19 | 0.419074 |
| CDS1 | 463.1009 | -1.0996 | 0.117067 | -9.39293 | 5.84E-21 | 2.47E-19 | 0.466645 |
| SMOC2 | 220.7636 | -1.93736 | 0.206579 | -9.37827 | 6.71E-21 | 2.82E-19 | 0.261094 |
| NADSYN1 | 1574.528 | -1.01997 | 0.109182 | -9.34193 | 9.46E-21 | 3.97E-19 | 0.493128 |
| PYGM | 244.5811 | -1.67825 | 0.179702 | -9.33907 | 9.72E-21 | 4.07E-19 | 0.312461 |
| LGI4 | 107.3702 | -2.53617 | 0.272304 | -9.31372 | 1.23E-20 | 5.11E-19 | 0.1724 |
| PCDH1 | 5545.053 | -0.94387 | 0.101758 | -9.27561 | 1.77E-20 | 7.26E-19 | 0.519838 |
| MLXIPL | 484.7073 | -2.17227 | 0.234218 | -9.27457 | 1.78E-20 | 7.28E-19 | 0.221861 |
| COMTD1 | 254.7799 | -1.67161 | 0.180566 | -9.25763 | 2.09E-20 | 8.48E-19 | 0.313903 |
| KLKB1 | 108.109 | -1.97338 | 0.213171 | -9.25729 | 2.10E-20 | 8.49E-19 | 0.254655 |
| CXCL12 | 1010.891 | -0.90364 | 0.097681 | -9.25097 | 2.22E-20 | 8.97E-19 | 0.534535 |
| FAM220CF | 122.8307 | -2.03724 | 0.220433 | -9.24202 | 2.42E-20 | 9.73E-19 | 0.243629 |
| UBALD1 | 503.7611 | -0.895 | 0.09688 | -9.23823 | 2.51E-20 | 1.01E-18 | 0.537746 |
| RBCK1 | 1157.351 | -1.00173 | 0.108719 | -9.21397 | 3.14E-20 | 1.25E-18 | 0.499401 |
| MTRNR2L6 | 9499.456 | -1.50237 | 0.163315 | -9.19921 | 3.61E-20 | 1.43E-18 | 0.352974 |
| EME2 | 974.262 | -1.91362 | 0.208047 | -9.19803 | 3.65E-20 | 1.44E-18 | 0.265426 |
| CARD11 | 722.6431 | -1.45783 | 0.158696 | -9.18629 | 4.07E-20 | 1.60E-18 | 0.36404 |
| CR2 | 135.1071 | -1.66434 | 0.181193 | -9.18546 | 4.10E-20 | 1.61E-18 | 0.315489 |
| PLOD3 | 1192.612 | -1.11521 | 0.121762 | -9.15892 | 5.24E-20 | 2.05E-18 | 0.461625 |
| SLC9A5 | 311.7466 | -1.83316 | 0.200272 | -9.15336 | 5.52E-20 | 2.15E-18 | 0.280649 |
| SNPH | 285.0121 | -1.40839 | 0.154052 | -9.14226 | 6.12E-20 | 2.38E-18 | 0.376733 |
| AASS | 8150.05 | -1.05572 | 0.115529 | -9.13817 | 6.35E-20 | 2.46E-18 | 0.481057 |
| SYT4 | 188.7981 | -2.52936 | 0.276929 | -9.13361 | 6.63E-20 | 2.55E-18 | 0.173215 |
| CREB3L1 | 521.9908 | -1.03527 | 0.113542 | -9.11797 | 7.65E-20 | 2.94E-18 | 0.487924 |
| PCSK1N | 126.0164 | -2.18937 | 0.240141 | -9.11703 | 7.72E-20 | 2.96E-18 | 0.219247 |
| TFEB | 97.92857 | -2.38692 | 0.262254 | -9.10153 | 8.91E-20 | 3.39E-18 | 0.191191 |
| AP3B2 | 413.0328 | -1.35865 | 0.149634 | -9.07985 | 1.09E-19 | 4.10E-18 | 0.389947 |
| SLC4A11 | 429.3357 | -1.26738 | 0.139651 | -9.07535 | 1.13E-19 | 4.27E-18 | 0.415414 |
| MTRNR2L1 | 147163.8 | -1.44331 | 0.159304 | -9.06006 | 1.30E-19 | 4.89E-18 | 0.367723 |
| PIGO | 968.0416 | -0.83374 | 0.092082 | -9.05423 | 1.38E-19 | 5.14E-18 | 0.561075 |
| ARHGAP4( | 78.51293 | -2.05843 | 0.227485 | -9.04867 | 1.45E-19 | 5.38E-18 | 0.240077 |
| AC073585 | 53.31214 | -4.3621 | 0.482816 | -9.0347 | 1.64E-19 | 6.09E-18 | 0.048627 |
| LAYN | 102.7342 | -2.27157 | 0.251532 | -9.03092 | 1.70E-19 | 6.28E-18 | 0.207105 |
| TLE2 | 216.1025 | -2.26142 | 0.250651 | -9.0222 | 1.84E-19 | 6.78E-18 | 0.208566 |
| MRAP2 | 119.648 | -2.13548 | 0.236805 | -9.01789 | 1.92E-19 | 7.02E-18 | 0.227592 |
| PGGHG | 2488.278 | -1.93486 | 0.214744 | -9.01007 | 2.06E-19 | 7.52E-18 | 0.261548 |
| C19orf33 | 52.56988 | -2.85564 | 0.318019 | -8.97945 | 2.72E-19 | 9.84E-18 | 0.138155 |
| SDC4 | 1429.987 | -0.88937 | 0.099098 | -8.97471 | 2.84E-19 | 1.03E-17 | 0.539849 |
| ERBB3 | 1130.108 | -1.0465 | 0.116827 | -8.95767 | 3.32E-19 | 1.19E-17 | 0.484141 |
| DENND4B | 2047.562 | -1.29928 | 0.145095 | -8.95466 | 3.41E-19 | 1.22E-17 | 0.40633 |
| FMNL1 | 319.1402 | -1.2654 | 0.141469 | -8.94474 | 3.73E-19 | 1.33E-17 | 0.415983 |
| ERVW-1 | 125.8749 | -1.88283 | 0.211004 | -8.92319 | 4.53E-19 | 1.61E-17 | 0.271152 |
| LDHD | 201.7231 | -1.81037 | 0.203051 | -8.91583 | 4.84E-19 | 1.72E-17 | 0.285118 |

| | | | | | | | |
|---|---|---|---|---|---|---|---|
| TMEM125 | 350.7679 | -1.76889 | 0.19858 | -8.90766 | 5.21E-19 | 1.85E-17 | 0.293435 |
| PPL | 1344.371 | -0.99268 | 0.111502 | -8.90273 | 5.45E-19 | 1.93E-17 | 0.502545 |
| EMC7 | 866.6007 | -0.7955 | 0.089527 | -8.88554 | 6.36E-19 | 2.24E-17 | 0.576145 |
| TUFM | 5226.292 | -0.58359 | 0.065686 | -8.88457 | 6.42E-19 | 2.25E-17 | 0.667301 |
| TDGF1P3 | 90.80173 | -2.07468 | 0.233632 | -8.88012 | 6.68E-19 | 2.34E-17 | 0.237388 |
| ID1 | 874.7519 | -1.01091 | 0.113989 | -8.8685 | 7.41E-19 | 2.59E-17 | 0.496232 |
| SPOCK3 | 36.63132 | -3.76433 | 0.425409 | -8.84873 | 8.85E-19 | 3.07E-17 | 0.073591 |
| SIGIRR | 622.5489 | -2.01597 | 0.2281 | -8.83808 | 9.74E-19 | 3.36E-17 | 0.247248 |
| DCLK1 | 1199.828 | -0.77837 | 0.088228 | -8.82227 | 1.12E-18 | 3.85E-17 | 0.583023 |
| ACOT2 | 212.5513 | -1.3283 | 0.150628 | -8.81845 | 1.16E-18 | 3.96E-17 | 0.398237 |
| SLC2A3 | 18494.69 | -0.75135 | 0.085408 | -8.79717 | 1.40E-18 | 4.78E-17 | 0.594049 |
| GRIP2 | 144.7815 | -1.79933 | 0.204849 | -8.78371 | 1.58E-18 | 5.33E-17 | 0.287307 |
| PRODH | 1084.297 | -1.42486 | 0.162276 | -8.78043 | 1.63E-18 | 5.47E-17 | 0.372456 |
| FURIN | 1249.456 | -0.63619 | 0.072546 | -8.76946 | 1.80E-18 | 6.02E-17 | 0.643411 |
| CAMKV | 888.0161 | -1.08962 | 0.124302 | -8.76588 | 1.85E-18 | 6.19E-17 | 0.469886 |
| B3GALT5 | 1312.744 | -1.39383 | 0.15928 | -8.7508 | 2.12E-18 | 7.05E-17 | 0.380554 |
| EDEM2 | 589.0146 | -0.9935 | 0.114177 | -8.70138 | 3.28E-18 | 1.08E-16 | 0.502259 |
| TMPRSS2 | 541.7386 | -0.91888 | 0.105946 | -8.67306 | 4.21E-18 | 1.37E-16 | 0.528921 |
| LZTS3 | 1100.93 | -1.1375 | 0.131165 | -8.67226 | 4.24E-18 | 1.38E-16 | 0.454546 |
| TSPAN4 | 1362.421 | -1.12564 | 0.130065 | -8.65441 | 4.95E-18 | 1.60E-16 | 0.4583 |
| AGPAT2 | 463.4512 | -1.24384 | 0.14375 | -8.6528 | 5.03E-18 | 1.62E-16 | 0.422248 |
| TLE5 | 2645.409 | -0.77633 | 0.089861 | -8.63923 | 5.66E-18 | 1.82E-16 | 0.58385 |
| HID1 | 236.2085 | -1.38452 | 0.16071 | -8.61502 | 6.99E-18 | 2.23E-16 | 0.383018 |
| TWF2 | 957.0173 | -0.71519 | 0.083126 | -8.60364 | 7.72E-18 | 2.46E-16 | 0.609125 |
| NEDD4 | 868.0541 | -0.73417 | 0.085921 | -8.54469 | 1.29E-17 | 4.04E-16 | 0.601165 |
| SLC29A1 | 4927.219 | -0.70978 | 0.083072 | -8.54408 | 1.30E-17 | 4.06E-16 | 0.611415 |
| RPS6KL1 | 351.8485 | -1.78652 | 0.20934 | -8.53403 | 1.41E-17 | 4.41E-16 | 0.289871 |
| AC104389 | 507.3379 | -1.15212 | 0.135036 | -8.53196 | 1.44E-17 | 4.48E-16 | 0.449963 |
| SLC15A3 | 429.7624 | -1.09289 | 0.128096 | -8.53181 | 1.44E-17 | 4.48E-16 | 0.46882 |
| NANOG | 1108.402 | -1.88459 | 0.220943 | -8.52974 | 1.47E-17 | 4.55E-16 | 0.270821 |
| CD9 | 2266.685 | -1.40371 | 0.165432 | -8.48513 | 2.15E-17 | 6.67E-16 | 0.377956 |
| SPARC | 6924.099 | -0.70583 | 0.083247 | -8.47881 | 2.28E-17 | 7.03E-16 | 0.613089 |
| C1orf210 | 153.2295 | -2.10097 | 0.247936 | -8.47382 | 2.37E-17 | 7.31E-16 | 0.233102 |
| HSPG2 | 4769.901 | -1.44557 | 0.170635 | -8.47175 | 2.42E-17 | 7.43E-16 | 0.367146 |
| CRB3 | 237.7771 | -1.28268 | 0.151545 | -8.46397 | 2.58E-17 | 7.91E-16 | 0.411032 |
| IFITM2 | 1555.619 | -0.79955 | 0.094464 | -8.46408 | 2.58E-17 | 7.91E-16 | 0.574528 |
| MFSD3 | 987.4603 | -1.09747 | 0.129665 | -8.4639 | 2.59E-17 | 7.91E-16 | 0.467335 |
| SPTBN1 | 7148.497 | -1.15418 | 0.136377 | -8.46312 | 2.60E-17 | 7.95E-16 | 0.449323 |
| RBPMS2 | 2869.35 | -1.11066 | 0.131253 | -8.46202 | 2.63E-17 | 8.01E-16 | 0.463082 |
| AC007325 | 2918.771 | -1.12277 | 0.132719 | -8.45973 | 2.68E-17 | 8.15E-16 | 0.459212 |
| SYP | 284.9462 | -1.69293 | 0.200295 | -8.45217 | 2.86E-17 | 8.69E-16 | 0.309299 |
| ICOSLG | 254.4866 | -1.16649 | 0.138043 | -8.45023 | 2.91E-17 | 8.82E-16 | 0.445503 |
| COL18A1 | 9146.437 | -1.24101 | 0.146885 | -8.44888 | 2.94E-17 | 8.91E-16 | 0.423075 |
| HTR7 | 853.7182 | -1.44712 | 0.171312 | -8.44728 | 2.98E-17 | 9.01E-16 | 0.366753 |

| | | | | | | | |
|---|---|---|---|---|---|---|---|
| IFI30 | 576.2912 | -1.54926 | 0.183484 | -8.44358 | 3.08E-17 | 9.27E-16 | 0.341686 |
| CLUH | 2347.275 | -0.66434 | 0.078693 | -8.44212 | 3.12E-17 | 9.37E-16 | 0.630978 |
| FOXI2 | 40.09876 | -3.40963 | 0.404663 | -8.42586 | 3.58E-17 | 1.07E-15 | 0.094102 |
| IGSF1 | 1861.347 | -1.06932 | 0.126932 | -8.4243 | 3.63E-17 | 1.08E-15 | 0.476545 |
| IL6R | 85.04679 | -2.02515 | 0.24039 | -8.42443 | 3.62E-17 | 1.08E-15 | 0.245679 |
| SYNGR1 | 508.8035 | -1.25605 | 0.149172 | -8.42016 | 3.76E-17 | 1.12E-15 | 0.418688 |
| FLII | 5219.684 | -0.76129 | 0.090514 | -8.41082 | 4.07E-17 | 1.21E-15 | 0.589967 |
| RASD2 | 100.0507 | -1.73423 | 0.206209 | -8.41008 | 4.10E-17 | 1.22E-15 | 0.300569 |
| GRB7 | 572.1066 | -1.06163 | 0.12636 | -8.40161 | 4.40E-17 | 1.31E-15 | 0.479091 |
| FHOD1 | 991.2641 | -1.14842 | 0.136734 | -8.3989 | 4.51E-17 | 1.33E-15 | 0.451119 |
| ICAM1 | 290.886 | -1.15343 | 0.13755 | -8.38551 | 5.05E-17 | 1.49E-15 | 0.449555 |
| TCF7L1 | 3288.072 | -0.64996 | 0.0776 | -8.37566 | 5.49E-17 | 1.61E-15 | 0.6373 |
| TMEM151A | 39.01862 | -3.3029 | 0.395246 | -8.35656 | 6.46E-17 | 1.88E-15 | 0.101328 |
| SLC39A11 | 296.7753 | -1.03302 | 0.123701 | -8.3509 | 6.77E-17 | 1.97E-15 | 0.488687 |
| EPHA8 | 175.5392 | -1.66695 | 0.199709 | -8.3469 | 7.01E-17 | 2.03E-15 | 0.314919 |
| CTSC | 7580.687 | -0.81454 | 0.097722 | -8.33528 | 7.73E-17 | 2.23E-15 | 0.568589 |
| KDR | 2354.81 | -0.93664 | 0.112392 | -8.33367 | 7.84E-17 | 2.26E-15 | 0.522448 |
| OLFM1 | 1348.648 | -0.80174 | 0.096236 | -8.33094 | 8.02E-17 | 2.31E-15 | 0.573659 |
| PRSS12 | 182.059 | -1.6375 | 0.196639 | -8.32743 | 8.26E-17 | 2.38E-15 | 0.321413 |
| MTRNR2L1 | 7511.781 | -1.59999 | 0.192222 | -8.32368 | 8.53E-17 | 2.45E-15 | 0.329879 |
| MLPH | 167.9171 | -1.56002 | 0.187454 | -8.32217 | 8.64E-17 | 2.48E-15 | 0.339146 |
| NRK | 67.29396 | -2.49576 | 0.300209 | -8.31342 | 9.30E-17 | 2.66E-15 | 0.177297 |
| SEMA4B | 1624.245 | -0.79161 | 0.095224 | -8.31311 | 9.32E-17 | 2.66E-15 | 0.5777 |
| RASL11B | 567.8716 | -1.28413 | 0.154504 | -8.31134 | 9.46E-17 | 2.70E-15 | 0.410618 |
| BCAM | 2276.001 | -1.20266 | 0.144759 | -8.30803 | 9.73E-17 | 2.77E-15 | 0.434474 |
| ADGRB2 | 972.2227 | -1.34758 | 0.162463 | -8.29467 | 1.09E-16 | 3.07E-15 | 0.392952 |
| LBH | 441.9851 | -1.20481 | 0.145424 | -8.28486 | 1.18E-16 | 3.33E-15 | 0.433825 |
| MAFK | 295.9415 | -1.0215 | 0.123521 | -8.26984 | 1.34E-16 | 3.75E-15 | 0.492604 |
| FLNA | 50888 | -1.44092 | 0.174244 | -8.26956 | 1.34E-16 | 3.75E-15 | 0.368332 |
| LSR | 4023.291 | -0.8267 | 0.100085 | -8.25997 | 1.46E-16 | 4.05E-15 | 0.563817 |
| SLC1A3 | 240.708 | -1.52972 | 0.185257 | -8.25729 | 1.49E-16 | 4.14E-15 | 0.346344 |
| RHBDF1 | 711.4561 | -1.54215 | 0.186785 | -8.25631 | 1.50E-16 | 4.16E-15 | 0.343373 |
| MMP17 | 691.5437 | -0.79362 | 0.096239 | -8.24632 | 1.63E-16 | 4.51E-15 | 0.576896 |
| MGAT4B | 2075.295 | -0.89732 | 0.108846 | -8.24393 | 1.67E-16 | 4.60E-15 | 0.536883 |
| NME3 | 916.6825 | -1.15959 | 0.140807 | -8.23535 | 1.79E-16 | 4.91E-15 | 0.447638 |
| SMARCA2 | 320.1388 | -1.97176 | 0.239638 | -8.22807 | 1.90E-16 | 5.19E-15 | 0.254942 |
| RIPOR3 | 78.86267 | -2.24053 | 0.272437 | -8.22401 | 1.97E-16 | 5.34E-15 | 0.211609 |
| HPDL | 278.0678 | -1.24095 | 0.150914 | -8.22287 | 1.99E-16 | 5.39E-15 | 0.423095 |
| TPTEP1 | 243.5667 | -1.24721 | 0.151707 | -8.22118 | 2.02E-16 | 5.45E-15 | 0.421261 |
| ACTN3 | 2291.285 | -1.51971 | 0.185057 | -8.21215 | 2.17E-16 | 5.86E-15 | 0.348756 |
| SLC27A6 | 199.0448 | -1.16159 | 0.141576 | -8.20471 | 2.31E-16 | 6.22E-15 | 0.447021 |
| TSPAN7 | 389.9866 | -1.06026 | 0.129329 | -8.19818 | 2.44E-16 | 6.55E-15 | 0.479545 |
| PWWP2B | 296.3469 | -1.06871 | 0.130433 | -8.1935 | 2.54E-16 | 6.79E-15 | 0.476746 |
| ALS2CL | 168.6658 | -2.5724 | 0.314057 | -8.19085 | 2.59E-16 | 6.93E-15 | 0.168125 |

| | | | | | | | |
|---|---|---|---|---|---|---|---|
| KCNH2 | 401.8555 | -0.83555 | 0.102034 | -8.1889 | 2.64E-16 | 7.01E-15 | 0.560371 |
| CAPN12 | 106.587 | -2.05164 | 0.250715 | -8.18316 | 2.76E-16 | 7.32E-15 | 0.24121 |
| CD177 | 64.38121 | -2.19954 | 0.268937 | -8.17864 | 2.87E-16 | 7.58E-15 | 0.217707 |
| B3GALT1 | 403.1931 | -0.95093 | 0.116386 | -8.17045 | 3.07E-16 | 8.08E-15 | 0.5173 |
| NMNAT3 | 2347.319 | -0.67853 | 0.083062 | -8.16898 | 3.11E-16 | 8.17E-15 | 0.6248 |
| ECE1 | 1983.156 | -0.63721 | 0.078125 | -8.15627 | 3.46E-16 | 9.03E-15 | 0.642956 |
| ZFP36 | 116.0668 | -1.60262 | 0.196666 | -8.14891 | 3.67E-16 | 9.59E-15 | 0.329279 |
| NOMO2 | 2554.753 | -0.94295 | 0.115737 | -8.1474 | 3.72E-16 | 9.69E-15 | 0.520167 |
| AC003112 | 168.0456 | -1.61307 | 0.198239 | -8.13699 | 4.05E-16 | 1.05E-14 | 0.326902 |
| DUSP8 | 40.05722 | -4.02756 | 0.495347 | -8.1308 | 4.26E-16 | 1.11E-14 | 0.061317 |
| PPIAP46 | 152.1415 | -1.54212 | 0.189761 | -8.12665 | 4.41E-16 | 1.15E-14 | 0.34338 |
| MAP7 | 1634.875 | -0.85031 | 0.104913 | -8.10492 | 5.28E-16 | 1.36E-14 | 0.554664 |
| ZYX | 3955.87 | -0.66939 | 0.08259 | -8.10502 | 5.27E-16 | 1.36E-14 | 0.628772 |
| IL34 | 66.11051 | -2.49883 | 0.308863 | -8.09042 | 5.95E-16 | 1.52E-14 | 0.17692 |
| TRIML2 | 424.7984 | -1.40406 | 0.173722 | -8.08225 | 6.36E-16 | 1.62E-14 | 0.377864 |
| SEZ6L2 | 2198.286 | -0.80967 | 0.100196 | -8.08083 | 6.43E-16 | 1.63E-14 | 0.570513 |
| ARHGAP4 | 909.2369 | -1.2959 | 0.160408 | -8.07878 | 6.54E-16 | 1.66E-14 | 0.407281 |
| HMCN2 | 952.6346 | -1.91033 | 0.236543 | -8.07606 | 6.69E-16 | 1.69E-14 | 0.266031 |
| EVA1B | 170.0223 | -1.52608 | 0.189006 | -8.07428 | 6.79E-16 | 1.71E-14 | 0.347218 |
| RNF128 | 62.37807 | -2.22044 | 0.275372 | -8.06342 | 7.42E-16 | 1.86E-14 | 0.214576 |
| DPF1 | 180.5889 | -1.5947 | 0.197851 | -8.06014 | 7.62E-16 | 1.91E-14 | 0.33109 |
| BRINP1 | 278.8581 | -1.13763 | 0.141169 | -8.05863 | 7.72E-16 | 1.93E-14 | 0.454506 |
| SCNN1G | 68.44051 | -2.76686 | 0.34397 | -8.04389 | 8.70E-16 | 2.17E-14 | 0.146924 |
| GCHFR | 312.3564 | -1.04932 | 0.130548 | -8.03784 | 9.14E-16 | 2.27E-14 | 0.483195 |
| LRP1 | 9391.434 | -0.78703 | 0.097951 | -8.0349 | 9.37E-16 | 2.32E-14 | 0.579538 |
| MST1R | 130.5672 | -1.87272 | 0.233202 | -8.03047 | 9.71E-16 | 2.40E-14 | 0.273058 |
| SERPINI1 | 199.085 | -1.23381 | 0.153663 | -8.02932 | 9.80E-16 | 2.41E-14 | 0.425194 |
| ARHGEF1 | 1179.139 | -0.77857 | 0.096975 | -8.02857 | 9.86E-16 | 2.43E-14 | 0.582944 |
| DNER | 169.0085 | -1.27372 | 0.158741 | -8.0239 | 1.02E-15 | 2.52E-14 | 0.413591 |
| 6-Sep | 1322.892 | -0.74334 | 0.092664 | -8.02188 | 1.04E-15 | 2.56E-14 | 0.597354 |
| FGF11 | 729.1559 | -0.76387 | 0.095511 | -7.99775 | 1.27E-15 | 3.09E-14 | 0.588915 |
| HCN1 | 83.50071 | -2.26857 | 0.28405 | -7.98652 | 1.39E-15 | 3.37E-14 | 0.207535 |
| VWDE | 338.7234 | -1.31787 | 0.165232 | -7.97586 | 1.51E-15 | 3.67E-14 | 0.401126 |
| C1orf94 | 238.0663 | -1.51241 | 0.189658 | -7.97439 | 1.53E-15 | 3.70E-14 | 0.350526 |
| KCNK6 | 646.5227 | -1.52771 | 0.191573 | -7.97454 | 1.53E-15 | 3.70E-14 | 0.346829 |
| UBR4 | 11486.09 | -0.87243 | 0.109481 | -7.96871 | 1.60E-15 | 3.87E-14 | 0.546228 |
| PMEPA1 | 267.1395 | -1.32821 | 0.166744 | -7.96557 | 1.64E-15 | 3.96E-14 | 0.398262 |
| ADGRG2 | 356.8538 | -1.15437 | 0.14495 | -7.96389 | 1.67E-15 | 4.01E-14 | 0.449264 |
| SLC15A1 | 45.52646 | -2.90514 | 0.365008 | -7.95913 | 1.73E-15 | 4.16E-14 | 0.133495 |
| LARGE2 | 2587.644 | -0.83272 | 0.104645 | -7.95757 | 1.75E-15 | 4.20E-14 | 0.561471 |
| TSHZ3 | 179.5302 | -1.75279 | 0.220269 | -7.95751 | 1.76E-15 | 4.20E-14 | 0.296727 |
| GPAA1 | 1539.533 | -0.88552 | 0.111354 | -7.95226 | 1.83E-15 | 4.37E-14 | 0.541293 |
| PABPC1L | 1397.714 | -0.70836 | 0.089286 | -7.93366 | 2.13E-15 | 5.05E-14 | 0.612014 |
| MRPS34 | 1889.753 | -0.55517 | 0.070042 | -7.92629 | 2.26E-15 | 5.35E-14 | 0.680575 |

| | | | | | | | |
|---|---|---|---|---|---|---|---|
| ELMO3 | 254.9385 | -1.97602 | 0.249732 | -7.91254 | 2.52E-15 | 5.96E-14 | 0.254191 |
| CSPG4 | 327.5657 | -1.23178 | 0.155852 | -7.90352 | 2.71E-15 | 6.40E-14 | 0.425792 |
| CDC42BP( | 729.4943 | -1.46749 | 0.18579 | -7.89863 | 2.82E-15 | 6.64E-14 | 0.361612 |
| ITIH5 | 106.2489 | -1.79565 | 0.227357 | -7.8979 | 2.84E-15 | 6.67E-14 | 0.288043 |
| COL8A2 | 62.6742 | -2.1928 | 0.278031 | -7.88691 | 3.10E-15 | 7.25E-14 | 0.218726 |
| HPN | 699.8816 | -1.23333 | 0.156567 | -7.87734 | 3.34E-15 | 7.82E-14 | 0.425335 |
| ERVMER34 | 353.603 | -1.07903 | 0.137 | -7.87615 | 3.38E-15 | 7.88E-14 | 0.473347 |
| WNT4 | 244.902 | -2.17657 | 0.276526 | -7.87113 | 3.51E-15 | 8.18E-14 | 0.221201 |
| SLC22A20 | 183.6657 | -1.6347 | 0.207848 | -7.86488 | 3.69E-15 | 8.57E-14 | 0.322037 |
| LRP10 | 537.4464 | -1.04961 | 0.133568 | -7.85819 | 3.90E-15 | 9.02E-14 | 0.4831 |
| JPH4 | 1711.971 | -0.75942 | 0.096658 | -7.85674 | 3.94E-15 | 9.10E-14 | 0.590735 |
| ZNF512B | 1388.717 | -0.70716 | 0.090015 | -7.85603 | 3.96E-15 | 9.14E-14 | 0.612525 |
| CGN | 1895.315 | -1.03598 | 0.132102 | -7.84228 | 4.42E-15 | 1.02E-13 | 0.487683 |
| SLC37A1 | 426.2555 | -1.26469 | 0.161442 | -7.83369 | 4.74E-15 | 1.09E-13 | 0.416189 |
| MAPK8IP2 | 384.1001 | -1.61592 | 0.206446 | -7.82731 | 4.98E-15 | 1.14E-13 | 0.326258 |
| LCK | 902.3448 | -1.36282 | 0.174185 | -7.82399 | 5.12E-15 | 1.17E-13 | 0.388822 |
| FKBP11 | 486.3461 | -1.29375 | 0.165458 | -7.81923 | 5.31E-15 | 1.21E-13 | 0.407888 |
| CASP10 | 262.3197 | -1.47899 | 0.189197 | -7.81719 | 5.40E-15 | 1.23E-13 | 0.35874 |
| HRH2 | 169.865 | -1.33072 | 0.170253 | -7.81614 | 5.45E-15 | 1.24E-13 | 0.39757 |
| ADCK2 | 376.4684 | -0.93431 | 0.119654 | -7.80844 | 5.79E-15 | 1.32E-13 | 0.523293 |
| KRTCAP3 | 347.9019 | -1.06173 | 0.136 | -7.80681 | 5.87E-15 | 1.33E-13 | 0.479059 |
| SLC9A1 | 869.9407 | -0.78447 | 0.100485 | -7.80688 | 5.86E-15 | 1.33E-13 | 0.580563 |
| CLDN4 | 932.6038 | -1.33713 | 0.171414 | -7.8006 | 6.16E-15 | 1.39E-13 | 0.395807 |
| SH3BP1 | 646.5304 | -0.98021 | 0.125688 | -7.79876 | 6.25E-15 | 1.41E-13 | 0.506907 |
| SUSD2 | 108.9036 | -1.50719 | 0.193516 | -7.78845 | 6.78E-15 | 1.53E-13 | 0.351797 |
| ESRP2 | 878.3223 | -0.95746 | 0.123047 | -7.78123 | 7.18E-15 | 1.61E-13 | 0.514964 |
| DDRGK1 | 792.5328 | -0.73367 | 0.094332 | -7.77753 | 7.40E-15 | 1.66E-13 | 0.601372 |
| SYT7 | 789.9467 | -0.7572 | 0.097367 | -7.77668 | 7.45E-15 | 1.67E-13 | 0.591645 |
| NPIPP1 | 427.5375 | -1.4386 | 0.185127 | -7.7709 | 7.79E-15 | 1.74E-13 | 0.368925 |
| SLC25A39 | 3669.716 | -0.50485 | 0.064969 | -7.77057 | 7.81E-15 | 1.74E-13 | 0.704734 |
| UGP2 | 12183.37 | -0.61702 | 0.079413 | -7.76971 | 7.87E-15 | 1.75E-13 | 0.652017 |
| PCSK4 | 213.2966 | -1.39317 | 0.179662 | -7.75441 | 8.88E-15 | 1.96E-13 | 0.380726 |
| GALNT3 | 454.1853 | -1.56762 | 0.202408 | -7.74482 | 9.57E-15 | 2.11E-13 | 0.337365 |
| SLC7A5 | 15141.75 | -0.653 | 0.084342 | -7.74234 | 9.76E-15 | 2.15E-13 | 0.635955 |
| CTSH | 446.9135 | -1.32004 | 0.17066 | -7.73494 | 1.03E-14 | 2.27E-13 | 0.400523 |
| CKB | 6105.922 | -0.63537 | 0.0822 | -7.72957 | 1.08E-14 | 2.36E-13 | 0.643777 |
| RAB13 | 3714.807 | -1.23599 | 0.160011 | -7.7244 | 1.12E-14 | 2.45E-13 | 0.424551 |
| ZMAT1 | 697.5953 | -0.74927 | 0.097005 | -7.72408 | 1.13E-14 | 2.45E-13 | 0.594902 |
| ELL2 | 589.9044 | -1.25574 | 0.16267 | -7.71957 | 1.17E-14 | 2.54E-13 | 0.418777 |
| FBLL1 | 248.1305 | -1.30984 | 0.169767 | -7.71552 | 1.20E-14 | 2.62E-13 | 0.403365 |
| SYNGR2 | 971.2545 | -0.8967 | 0.116287 | -7.71108 | 1.25E-14 | 2.70E-13 | 0.537114 |
| ATP1B2 | 508.6472 | -1.27413 | 0.165359 | -7.70524 | 1.31E-14 | 2.83E-13 | 0.413475 |
| CCDC88B | 539.9842 | -1.87249 | 0.243109 | -7.70224 | 1.34E-14 | 2.89E-13 | 0.273102 |
| CCDC120 | 494.5031 | -0.93966 | 0.122274 | -7.68488 | 1.53E-14 | 3.30E-13 | 0.521355 |

| | | | | | | | |
|---|---|---|---|---|---|---|---|
| TRIP10 | 1473.441 | -0.7973 | 0.103833 | -7.67865 | 1.61E-14 | 3.46E-13 | 0.575426 |
| SYNGR3 | 349.0064 | -1.22772 | 0.159922 | -7.677 | 1.63E-14 | 3.49E-13 | 0.426991 |
| EDIL3 | 1498.088 | -0.84147 | 0.109767 | -7.66597 | 1.77E-14 | 3.80E-13 | 0.558075 |
| AC012435 | 546.9856 | -0.85739 | 0.11197 | -7.6573 | 1.90E-14 | 4.05E-13 | 0.551951 |
| RAC3 | 1162.549 | -0.707 | 0.092379 | -7.6532 | 1.96E-14 | 4.17E-13 | 0.612594 |
| SRRM3 | 336.7742 | -0.93221 | 0.122005 | -7.64073 | 2.16E-14 | 4.58E-13 | 0.524057 |
| TMEM205 | 933.1896 | -0.84667 | 0.110833 | -7.63915 | 2.19E-14 | 4.63E-13 | 0.556068 |
| TOR4A | 286.9138 | -1.3249 | 0.173622 | -7.63095 | 2.33E-14 | 4.91E-13 | 0.399177 |
| NAGLU | 844.9698 | -0.91009 | 0.119326 | -7.62689 | 2.40E-14 | 5.05E-13 | 0.532153 |
| BNC1 | 75.00841 | -4.2169 | 0.55298 | -7.62578 | 2.43E-14 | 5.09E-13 | 0.053776 |
| GPR37 | 145.7659 | -1.52363 | 0.199878 | -7.62281 | 2.48E-14 | 5.20E-13 | 0.34781 |
| KDF1 | 289.5536 | -0.98697 | 0.129543 | -7.61885 | 2.56E-14 | 5.35E-13 | 0.504535 |
| IL17RC | 301.3333 | -0.97222 | 0.127675 | -7.61483 | 2.64E-14 | 5.51E-13 | 0.509721 |
| TEX264 | 286.9628 | -1.21488 | 0.159625 | -7.61089 | 2.72E-14 | 5.67E-13 | 0.430808 |
| ICAM5 | 94.46758 | -2.39704 | 0.315192 | -7.60501 | 2.85E-14 | 5.92E-13 | 0.189854 |
| YIPF2 | 885.7013 | -0.87082 | 0.114714 | -7.59126 | 3.17E-14 | 6.55E-13 | 0.546835 |
| SAMD4A | 321.4466 | -1.1611 | 0.152981 | -7.58984 | 3.20E-14 | 6.61E-13 | 0.447172 |
| PLEC | 5554.83 | -1.22141 | 0.160979 | -7.58741 | 3.26E-14 | 6.73E-13 | 0.428862 |
| TERF1 | 7486.132 | -0.81518 | 0.107667 | -7.57129 | 3.70E-14 | 7.59E-13 | 0.568339 |
| VRTN | 3924.892 | -0.73228 | 0.096836 | -7.56213 | 3.97E-14 | 8.11E-13 | 0.601951 |
| PACSIN3 | 814.8168 | -0.69312 | 0.091754 | -7.55406 | 4.22E-14 | 8.60E-13 | 0.618515 |
| SAT2 | 870.0741 | -0.60154 | 0.079672 | -7.55021 | 4.35E-14 | 8.83E-13 | 0.659051 |
| SLC18A2 | 146.0809 | -1.41076 | 0.186903 | -7.54805 | 4.42E-14 | 8.97E-13 | 0.376114 |
| CCDC78 | 224.0652 | -2.13865 | 0.283442 | -7.54528 | 4.51E-14 | 9.16E-13 | 0.227093 |
| SFRP5 | 186.8839 | -1.72168 | 0.228436 | -7.53685 | 4.81E-14 | 9.73E-13 | 0.303194 |
| ILDR1 | 60.96914 | -2.37151 | 0.315104 | -7.52612 | 5.23E-14 | 1.05E-12 | 0.193243 |
| APOBEC3( | 877.771 | -0.84745 | 0.112765 | -7.51518 | 5.68E-14 | 1.15E-12 | 0.555767 |
| MTRNR2L1 | 392.222 | -1.71938 | 0.228911 | -7.51111 | 5.86E-14 | 1.18E-12 | 0.30368 |
| NPC1 | 1241.846 | -0.6597 | 0.087897 | -7.50543 | 6.12E-14 | 1.23E-12 | 0.633008 |
| PPP1R14B | 3425.291 | -0.55581 | 0.074061 | -7.50473 | 6.16E-14 | 1.23E-12 | 0.680277 |
| UGT8 | 627.999 | -0.86765 | 0.115967 | -7.48189 | 7.33E-14 | 1.46E-12 | 0.548037 |
| PARP12 | 304.0441 | -1.05904 | 0.141627 | -7.47766 | 7.57E-14 | 1.50E-12 | 0.479951 |
| SPEG | 747.0236 | -0.82163 | 0.109887 | -7.47707 | 7.60E-14 | 1.51E-12 | 0.565802 |
| ORAI1 | 478.8631 | -0.71786 | 0.096176 | -7.46394 | 8.40E-14 | 1.66E-12 | 0.608001 |
| HOOK2 | 1165.95 | -1.44949 | 0.194315 | -7.45948 | 8.69E-14 | 1.71E-12 | 0.36615 |
| THBS2 | 527.1149 | -0.99384 | 0.133326 | -7.45418 | 9.04E-14 | 1.78E-12 | 0.50214 |
| GPX3 | 770.141 | -0.70473 | 0.094543 | -7.45403 | 9.05E-14 | 1.78E-12 | 0.613558 |
| PARP15 | 348.2254 | -0.97476 | 0.130801 | -7.45225 | 9.18E-14 | 1.80E-12 | 0.508823 |
| ELOVL1 | 491.5385 | -0.93892 | 0.126142 | -7.44339 | 9.81E-14 | 1.92E-12 | 0.521623 |
| TNFSF11 | 166.4231 | -2.00133 | 0.269076 | -7.43777 | 1.02E-13 | 2.00E-12 | 0.24977 |
| PLSCR3 | 560.9861 | -0.71789 | 0.096576 | -7.43336 | 1.06E-13 | 2.06E-12 | 0.607988 |
| ACSL6 | 97.30536 | -2.12962 | 0.286639 | -7.42961 | 1.09E-13 | 2.12E-12 | 0.228519 |
| MDN1 | 5161.961 | -1.06604 | 0.143551 | -7.4262 | 1.12E-13 | 2.17E-12 | 0.477629 |
| CILP2 | 211.917 | -1.30039 | 0.175232 | -7.42099 | 1.16E-13 | 2.25E-12 | 0.406016 |

| | | | | | | | |
|---|---|---|---|---|---|---|---|
| AEBP1 | 1169.975 | -1.01508 | 0.136889 | -7.41532 | 1.21E-13 | 2.35E-12 | 0.494802 |
| IGFBP3 | 762.6106 | -1.12576 | 0.151851 | -7.41357 | 1.23E-13 | 2.38E-12 | 0.458261 |
| SLC52A3 | 140.2258 | -1.22836 | 0.16585 | -7.40648 | 1.30E-13 | 2.50E-12 | 0.426802 |
| FLOT1 | 155.7146 | -25.0436 | 3.383632 | -7.4014 | 1.35E-13 | 2.60E-12 | 2.89E-08 |
| ZNF700 | 26.10317 | -8.17711 | 1.105638 | -7.39583 | 1.41E-13 | 2.70E-12 | 0.003455 |
| SEMA3B | 333.0873 | -1.73222 | 0.234312 | -7.39278 | 1.44E-13 | 2.75E-12 | 0.300989 |
| NDRG2 | 418.67 | -0.93108 | 0.125971 | -7.39116 | 1.46E-13 | 2.78E-12 | 0.524467 |
| TMC6 | 1331.426 | -1.22986 | 0.166401 | -7.39094 | 1.46E-13 | 2.79E-12 | 0.426359 |
| NDUFAF7 | 1040.2 | -0.78836 | 0.106697 | -7.38876 | 1.48E-13 | 2.82E-12 | 0.579001 |
| EMD | 1752.213 | -0.5752 | 0.077888 | -7.38503 | 1.52E-13 | 2.89E-12 | 0.671192 |
| PDZRN3 | 521.0711 | -0.7576 | 0.102668 | -7.37917 | 1.59E-13 | 3.02E-12 | 0.591478 |
| CD40 | 128.3894 | -2.00882 | 0.272615 | -7.3687 | 1.72E-13 | 3.25E-12 | 0.248476 |
| RAB37 | 171.2203 | -1.58171 | 0.215448 | -7.3415 | 2.11E-13 | 3.96E-12 | 0.334085 |
| SHISA5 | 4256.145 | -0.57223 | 0.077951 | -7.34089 | 2.12E-13 | 3.97E-12 | 0.672577 |
| IFITM3 | 2511.175 | -0.68603 | 0.093478 | -7.33895 | 2.15E-13 | 4.03E-12 | 0.621561 |
| CC2D1A | 812.8529 | -0.9677 | 0.131871 | -7.3382 | 2.16E-13 | 4.04E-12 | 0.511321 |
| MST1L | 108.9648 | -2.72079 | 0.371095 | -7.33179 | 2.27E-13 | 4.22E-12 | 0.151691 |
| CTSF | 73.90929 | -2.68058 | 0.365733 | -7.32933 | 2.31E-13 | 4.30E-12 | 0.155979 |
| BAMBI | 196.1094 | -1.27491 | 0.17396 | -7.32877 | 2.32E-13 | 4.31E-12 | 0.413251 |
| PCDHGB4 | 72.25719 | -1.81694 | 0.248438 | -7.31345 | 2.60E-13 | 4.80E-12 | 0.283822 |
| MAMDC4 | 401.9636 | -1.28385 | 0.175721 | -7.3062 | 2.75E-13 | 5.06E-12 | 0.410698 |
| SEMG1 | 36.08876 | -3.54891 | 0.485952 | -7.30301 | 2.81E-13 | 5.18E-12 | 0.085442 |
| RTN2 | 287.5426 | -1.21369 | 0.166234 | -7.30107 | 2.85E-13 | 5.24E-12 | 0.431165 |
| TMEM198 | 195.9508 | -0.97279 | 0.133239 | -7.30109 | 2.85E-13 | 5.24E-12 | 0.50952 |
| MMP9 | 71.91638 | -1.91455 | 0.262396 | -7.2964 | 2.96E-13 | 5.42E-12 | 0.265255 |
| PLTP | 3799.36 | -0.7162 | 0.098267 | -7.28825 | 3.14E-13 | 5.75E-12 | 0.6087 |
| NKAP | 842.4263 | -0.78937 | 0.108478 | -7.27679 | 3.42E-13 | 6.23E-12 | 0.578597 |
| DHCR24 | 8257.096 | -0.82419 | 0.11329 | -7.27504 | 3.46E-13 | 6.30E-12 | 0.564798 |
| AC092070 | 25.3398 | -4.60423 | 0.633156 | -7.27189 | 3.55E-13 | 6.44E-12 | 0.041114 |
| FOXL2NB | 313.04 | -2.14477 | 0.295245 | -7.26435 | 3.75E-13 | 6.79E-12 | 0.226131 |
| CARMIL2 | 957.0571 | -1.13548 | 0.156351 | -7.26238 | 3.80E-13 | 6.88E-12 | 0.455183 |
| KLK13 | 29.97216 | -3.65472 | 0.503529 | -7.25822 | 3.92E-13 | 7.08E-12 | 0.0794 |
| ADAM15 | 1124.96 | -0.8949 | 0.123579 | -7.24155 | 4.44E-13 | 7.95E-12 | 0.537783 |
| AC019257 | 46.45728 | -3.9364 | 0.544302 | -7.23201 | 4.76E-13 | 8.50E-12 | 0.065317 |
| TRIM46 | 539.3049 | -1.15995 | 0.160512 | -7.22655 | 4.95E-13 | 8.84E-12 | 0.447528 |
| KRT7 | 212.5462 | -1.24099 | 0.171774 | -7.22454 | 5.03E-13 | 8.96E-12 | 0.423082 |
| BEST2 | 76.13613 | -1.80008 | 0.24929 | -7.22083 | 5.17E-13 | 9.20E-12 | 0.287158 |
| HTR1D | 201.1999 | -1.19422 | 0.165401 | -7.22018 | 5.19E-13 | 9.24E-12 | 0.437021 |
| NEIL1 | 198.1167 | -1.51166 | 0.209437 | -7.21775 | 5.29E-13 | 9.39E-12 | 0.350707 |
| MYH14 | 624.7623 | -0.81329 | 0.112992 | -7.19778 | 6.12E-13 | 1.08E-11 | 0.569082 |
| ARHGEF16 | 570.3417 | -0.90638 | 0.125975 | -7.19493 | 6.25E-13 | 1.10E-11 | 0.533522 |
| PPIB | 4652.605 | -0.92192 | 0.128167 | -7.19311 | 6.33E-13 | 1.12E-11 | 0.527807 |
| DPP4 | 147.914 | -1.59011 | 0.22114 | -7.19052 | 6.45E-13 | 1.14E-11 | 0.332146 |
| ZNF665 | 111.2744 | -2.50824 | 0.349043 | -7.18606 | 6.67E-13 | 1.18E-11 | 0.175769 |

| | | | | | | | |
|---|---|---|---|---|---|---|---|
| UAP1L1 | 406.5751 | -0.95491 | 0.132952 | -7.18239 | 6.85E-13 | 1.20E-11 | 0.515874 |
| PXDNL | 28.04549 | -3.76032 | 0.524045 | -7.17557 | 7.20E-13 | 1.26E-11 | 0.073795 |
| SLC16A1 | 15444.22 | -0.59495 | 0.083033 | -7.1652 | 7.77E-13 | 1.36E-11 | 0.662068 |
| SLITRK2 | 98.79347 | -2.12508 | 0.29658 | -7.16526 | 7.76E-13 | 1.36E-11 | 0.229239 |
| RILPL2 | 560.4399 | -0.70521 | 0.098483 | -7.16071 | 8.03E-13 | 1.40E-11 | 0.613353 |
| CD24 | 33859.17 | -0.53156 | 0.074248 | -7.15927 | 8.11E-13 | 1.41E-11 | 0.691806 |
| REEP5 | 1400.876 | -0.76244 | 0.106505 | -7.15875 | 8.14E-13 | 1.42E-11 | 0.589498 |
| KIRREL2 | 112.2967 | -1.40123 | 0.196311 | -7.1378 | 9.48E-13 | 1.64E-11 | 0.378607 |
| EXOC3L2 | 50.42678 | -2.28292 | 0.320552 | -7.12185 | 1.06E-12 | 1.84E-11 | 0.205481 |
| PPP1R14A | 241.8217 | -1.15255 | 0.161861 | -7.12061 | 1.07E-12 | 1.85E-11 | 0.449829 |
| COL9A3 | 1709.691 | -1.74658 | 0.245306 | -7.11998 | 1.08E-12 | 1.86E-11 | 0.298008 |
| PLAU | 817.8839 | -0.57316 | 0.080567 | -7.11412 | 1.13E-12 | 1.94E-11 | 0.672141 |
| NFE2L3 | 2064.984 | -1.10308 | 0.155125 | -7.1109 | 1.15E-12 | 1.98E-11 | 0.465522 |
| SLC52A2 | 1314.823 | -0.85517 | 0.120492 | -7.09735 | 1.27E-12 | 2.17E-11 | 0.552799 |
| EML2 | 409.8087 | -0.99444 | 0.140199 | -7.0931 | 1.31E-12 | 2.24E-11 | 0.501929 |
| EIF5A | 13667.93 | -0.50689 | 0.071584 | -7.08101 | 1.43E-12 | 2.44E-11 | 0.703738 |
| VASH2 | 1176.156 | -0.78818 | 0.111335 | -7.0794 | 1.45E-12 | 2.46E-11 | 0.579072 |
| RRAS | 252.905 | -1.10328 | 0.15596 | -7.07412 | 1.50E-12 | 2.55E-11 | 0.465457 |
| FGD5 | 572.809 | -1.30988 | 0.185352 | -7.06698 | 1.58E-12 | 2.67E-11 | 0.403355 |
| RTEL1-TNF | 323.8687 | -1.29006 | 0.182628 | -7.06382 | 1.62E-12 | 2.73E-11 | 0.408935 |
| SPOCK2 | 125.7035 | -2.06378 | 0.292364 | -7.05894 | 1.68E-12 | 2.82E-11 | 0.239189 |
| PLCB2 | 160.0308 | -1.59713 | 0.226409 | -7.05418 | 1.74E-12 | 2.92E-11 | 0.330533 |
| M1AP | 34.07578 | -7.7639 | 1.103563 | -7.0353 | 1.99E-12 | 3.32E-11 | 0.004601 |
| MORC4 | 577.4524 | -0.93981 | 0.133648 | -7.032 | 2.04E-12 | 3.40E-11 | 0.5213 |
| NPEPL1 | 1270.055 | -1.05772 | 0.15054 | -7.02621 | 2.12E-12 | 3.53E-11 | 0.48039 |
| QSOX1 | 2106.237 | -0.94884 | 0.135303 | -7.0127 | 2.34E-12 | 3.87E-11 | 0.518049 |
| C6orf132 | 1248.993 | -0.92013 | 0.131288 | -7.00846 | 2.41E-12 | 3.98E-11 | 0.528461 |
| ATP2C2 | 137.0939 | -2.24474 | 0.320471 | -7.00449 | 2.48E-12 | 4.09E-11 | 0.210993 |
| IFNGR1 | 837.6591 | -0.53289 | 0.076125 | -7.00015 | 2.56E-12 | 4.20E-11 | 0.691171 |
| CRACR2B | 310.5965 | -1.36073 | 0.194467 | -6.99726 | 2.61E-12 | 4.28E-11 | 0.389385 |
| SSC5D | 161.7485 | -1.54679 | 0.221113 | -6.99547 | 2.64E-12 | 4.33E-11 | 0.342272 |
| NR2F6 | 2074.231 | -0.46759 | 0.066874 | -6.99214 | 2.71E-12 | 4.43E-11 | 0.723172 |
| DUS3L | 534.8517 | -0.89867 | 0.128666 | -6.98457 | 2.86E-12 | 4.67E-11 | 0.53638 |
| MRS2 | 1650.849 | -0.77888 | 0.111554 | -6.98207 | 2.91E-12 | 4.75E-11 | 0.582819 |
| PRICKLE1 | 758.7219 | -1.51713 | 0.21743 | -6.97757 | 3.00E-12 | 4.89E-11 | 0.349379 |
| CHCHD10 | 618.7613 | -1.11993 | 0.160733 | -6.96765 | 3.22E-12 | 5.23E-11 | 0.460116 |
| PLD3 | 5824.176 | -0.69893 | 0.100331 | -6.96625 | 3.25E-12 | 5.28E-11 | 0.616027 |
| PLCB3 | 3031.488 | -0.65096 | 0.093477 | -6.9638 | 3.31E-12 | 5.36E-11 | 0.636858 |
| CERS4 | 1513.277 | -0.89049 | 0.12791 | -6.96182 | 3.36E-12 | 5.43E-11 | 0.539432 |
| CHST8 | 211.3441 | -1.18357 | 0.170074 | -6.95915 | 3.42E-12 | 5.53E-11 | 0.440261 |
| SLC18B1 | 516.0927 | -0.67763 | 0.097466 | -6.95252 | 3.59E-12 | 5.77E-11 | 0.625191 |
| MTCO1P12 | 1218.42 | -1.54594 | 0.222407 | -6.95093 | 3.63E-12 | 5.83E-11 | 0.342473 |
| GNA14 | 154.1929 | -1.95516 | 0.281347 | -6.94926 | 3.67E-12 | 5.90E-11 | 0.257893 |
| TAZ | 706.1045 | -0.75552 | 0.108745 | -6.94763 | 3.71E-12 | 5.96E-11 | 0.592334 |

| | | | | | | | |
|---|---|---|---|---|---|---|---|
| JAM2 | 202.887 | -1.21412 | 0.174854 | -6.94362 | 3.82E-12 | 6.13E-11 | 0.431035 |
| TAF1D | 2352.509 | -0.53411 | 0.076978 | -6.93855 | 3.96E-12 | 6.35E-11 | 0.690583 |
| AC100868 | 44.90584 | -2.60254 | 0.375219 | -6.93606 | 4.03E-12 | 6.45E-11 | 0.164648 |
| MAPK8IP1 | 1446.784 | -0.59943 | 0.086432 | -6.93531 | 4.05E-12 | 6.48E-11 | 0.660014 |
| FBXO41 | 674.8734 | -0.87143 | 0.125663 | -6.93469 | 4.07E-12 | 6.50E-11 | 0.546604 |
| TMED3 | 1117.776 | -0.67037 | 0.096709 | -6.93183 | 4.15E-12 | 6.63E-11 | 0.628346 |
| POU5F1 | 2695.533 | -0.94828 | 0.136969 | -6.92328 | 4.41E-12 | 7.03E-11 | 0.518252 |
| LAMA5 | 9326.581 | -2.42053 | 0.349796 | -6.91982 | 4.52E-12 | 7.19E-11 | 0.186788 |
| ASAP3 | 903.821 | -0.74177 | 0.107229 | -6.91761 | 4.59E-12 | 7.30E-11 | 0.598006 |
| PRAG1 | 1549.241 | -0.55551 | 0.080318 | -6.91638 | 4.63E-12 | 7.35E-11 | 0.680418 |
| NAT14 | 918.9863 | -0.95277 | 0.137831 | -6.91261 | 4.76E-12 | 7.53E-11 | 0.516638 |
| TACR3 | 194.1535 | -1.16348 | 0.16832 | -6.91232 | 4.77E-12 | 7.53E-11 | 0.446434 |
| TSC22D4 | 507.8803 | -0.78938 | 0.114206 | -6.91193 | 4.78E-12 | 7.55E-11 | 0.578592 |
| MYBBP1A | 2812.768 | -0.66417 | 0.096184 | -6.90522 | 5.01E-12 | 7.89E-11 | 0.631051 |
| APH1B | 843.6539 | -0.87426 | 0.126624 | -6.90436 | 5.04E-12 | 7.93E-11 | 0.545534 |
| PCDHB15 | 131.28 | -1.39027 | 0.201549 | -6.89796 | 5.28E-12 | 8.28E-11 | 0.381492 |
| SEC14L4 | 34.39486 | -2.79902 | 0.405845 | -6.89678 | 5.32E-12 | 8.34E-11 | 0.143684 |
| NQO2 | 1055.814 | -0.55937 | 0.08117 | -6.89137 | 5.53E-12 | 8.65E-11 | 0.678597 |
| SULT4A1 | 364.8977 | -0.84466 | 0.12258 | -6.8907 | 5.55E-12 | 8.68E-11 | 0.556842 |
| UNC79 | 161.9105 | -1.12493 | 0.163374 | -6.88558 | 5.76E-12 | 8.97E-11 | 0.458526 |
| TLCD1 | 386.0722 | -0.92956 | 0.135073 | -6.8819 | 5.91E-12 | 9.20E-11 | 0.525018 |
| ISOC2 | 817.8747 | -0.63012 | 0.0916 | -6.87907 | 6.02E-12 | 9.38E-11 | 0.646121 |
| HCN4 | 819.8215 | -0.89538 | 0.130189 | -6.87752 | 6.09E-12 | 9.47E-11 | 0.537607 |
| COL21A1 | 110.8997 | -1.81134 | 0.263577 | -6.87214 | 6.32E-12 | 9.80E-11 | 0.284926 |
| HRH3 | 111.4093 | -1.41278 | 0.205727 | -6.86727 | 6.54E-12 | 1.01E-10 | 0.375586 |
| RELB | 351.2751 | -0.86245 | 0.125659 | -6.86344 | 6.72E-12 | 1.04E-10 | 0.550017 |
| ACAP3 | 1239.217 | -1.71736 | 0.250467 | -6.85661 | 7.05E-12 | 1.09E-10 | 0.304106 |
| PRX | 345.1733 | -0.90371 | 0.131814 | -6.85593 | 7.08E-12 | 1.09E-10 | 0.534511 |
| FADS3 | 1208.548 | -1.09362 | 0.159664 | -6.84955 | 7.41E-12 | 1.14E-10 | 0.468583 |
| CEP170B | 1549.868 | -0.6608 | 0.096517 | -6.84644 | 7.57E-12 | 1.16E-10 | 0.632527 |
| TMEM130 | 179.4905 | -1.71415 | 0.25053 | -6.8421 | 7.80E-12 | 1.20E-10 | 0.304782 |
| UCKL1 | 683.0257 | -1.08615 | 0.159032 | -6.82974 | 8.51E-12 | 1.30E-10 | 0.471018 |
| CD8B | 36.75634 | -4.97366 | 0.729082 | -6.8218 | 8.99E-12 | 1.37E-10 | 0.031826 |
| EGFL7 | 1163.591 | -1.16131 | 0.170322 | -6.81832 | 9.21E-12 | 1.40E-10 | 0.447107 |
| FRAS1 | 4190.727 | -0.50348 | 0.073894 | -6.81354 | 9.52E-12 | 1.44E-10 | 0.705403 |
| TMEM35B | 310.5747 | -0.89918 | 0.132078 | -6.80795 | 9.90E-12 | 1.50E-10 | 0.536191 |
| KLRG2 | 369.5038 | -0.91624 | 0.134646 | -6.8048 | 1.01E-11 | 1.53E-10 | 0.529889 |
| SLC39A1 | 5864.372 | -0.69887 | 0.102739 | -6.80238 | 1.03E-11 | 1.55E-10 | 0.616053 |
| EFNA1 | 104.3564 | -1.42746 | 0.21003 | -6.79646 | 1.07E-11 | 1.62E-10 | 0.371785 |
| TCN2 | 1527.22 | -0.76288 | 0.112282 | -6.79434 | 1.09E-11 | 1.64E-10 | 0.589319 |
| PMAIP1 | 1866.391 | -1.03561 | 0.152548 | -6.78872 | 1.13E-11 | 1.70E-10 | 0.487811 |
| PRNP | 884.5773 | -0.7417 | 0.109262 | -6.78826 | 1.13E-11 | 1.70E-10 | 0.598035 |
| PHC1P1 | 1549.046 | -0.95491 | 0.140714 | -6.78614 | 1.15E-11 | 1.72E-10 | 0.515875 |
| MARVELD3 | 688.3198 | -0.92279 | 0.136011 | -6.78469 | 1.16E-11 | 1.74E-10 | 0.527486 |

| | | | | | | | |
|---|---|---|---|---|---|---|---|
| GPR160 | 324.4879 | -0.9866 | 0.145455 | -6.7828 | 1.18E-11 | 1.76E-10 | 0.504667 |
| NPIPB5 | 2514.463 | -0.88082 | 0.129888 | -6.78139 | 1.19E-11 | 1.78E-10 | 0.543058 |
| P2RY11 | 201.2013 | -0.91375 | 0.134784 | -6.77936 | 1.21E-11 | 1.80E-10 | 0.530803 |
| MCRIP2 | 383.5796 | -1.32553 | 0.195663 | -6.77457 | 1.25E-11 | 1.85E-10 | 0.399003 |
| GTF2IP20 | 333.2549 | -1.07333 | 0.15847 | -6.77309 | 1.26E-11 | 1.87E-10 | 0.475221 |
| TMEM161A | 1105.584 | -0.75949 | 0.112181 | -6.77023 | 1.29E-11 | 1.90E-10 | 0.590704 |
| TRMT61A | 401.3091 | -0.8443 | 0.124736 | -6.76865 | 1.30E-11 | 1.92E-10 | 0.556983 |
| PRKCG | 208.9274 | -1.40681 | 0.208282 | -6.75436 | 1.43E-11 | 2.11E-10 | 0.377144 |
| CYB5R1 | 308.7431 | -0.98149 | 0.145319 | -6.75405 | 1.44E-11 | 2.12E-10 | 0.506456 |
| CACNA1F | 63.36933 | -2.13494 | 0.316148 | -6.75298 | 1.45E-11 | 2.13E-10 | 0.227677 |
| SAP25 | 84.8124 | -2.65096 | 0.392568 | -6.75287 | 1.45E-11 | 2.13E-10 | 0.159214 |
| PKLR | 173.4607 | -1.39437 | 0.206645 | -6.74764 | 1.50E-11 | 2.21E-10 | 0.380411 |
| COX6B2 | 95.12625 | -2.11736 | 0.313825 | -6.74694 | 1.51E-11 | 2.21E-10 | 0.230468 |
| FBLIM1 | 1345.068 | -0.86912 | 0.128844 | -6.74553 | 1.52E-11 | 2.23E-10 | 0.54748 |
| TPST2 | 1180.434 | -0.58395 | 0.086656 | -6.73875 | 1.60E-11 | 2.33E-10 | 0.667133 |
| PDPN | 1963.402 | -1.17365 | 0.174418 | -6.72896 | 1.71E-11 | 2.49E-10 | 0.443298 |
| CACNA1I | 86.55289 | -1.81037 | 0.269174 | -6.72566 | 1.75E-11 | 2.54E-10 | 0.285118 |
| C16orf58 | 1251.118 | -0.53631 | 0.079769 | -6.72333 | 1.78E-11 | 2.58E-10 | 0.689532 |
| HRC | 85.46958 | -1.72593 | 0.257015 | -6.71529 | 1.88E-11 | 2.72E-10 | 0.302303 |
| SPATA20 | 1072.179 | -0.66849 | 0.099586 | -6.71267 | 1.91E-11 | 2.76E-10 | 0.629167 |
| ANKRD33E | 527.3957 | -0.75967 | 0.113173 | -6.71248 | 1.91E-11 | 2.76E-10 | 0.590631 |
| FAM86B1 | 1010.303 | -0.68428 | 0.10197 | -6.71053 | 1.94E-11 | 2.80E-10 | 0.622318 |
| ZNF836 | 105.3218 | -1.68028 | 0.250541 | -6.70663 | 1.99E-11 | 2.87E-10 | 0.312021 |
| FGF19 | 99.82711 | -1.71375 | 0.255539 | -6.7064 | 1.99E-11 | 2.87E-10 | 0.304867 |
| LAMP1 | 5838.818 | -0.63155 | 0.094183 | -6.70557 | 2.01E-11 | 2.88E-10 | 0.645482 |
| P2RX2 | 35.48494 | -3.39974 | 0.507001 | -6.7056 | 2.01E-11 | 2.88E-10 | 0.094749 |
| CYGB | 109.7015 | -1.34271 | 0.200403 | -6.70006 | 2.08E-11 | 2.98E-10 | 0.39428 |
| SPINT2 | 5491.654 | -0.93483 | 0.139561 | -6.69834 | 2.11E-11 | 3.02E-10 | 0.523106 |
| BUD23 | 1622.058 | -0.47155 | 0.07043 | -6.69532 | 2.15E-11 | 3.08E-10 | 0.72119 |
| DSG2 | 7313.755 | -0.43454 | 0.06491 | -6.69455 | 2.16E-11 | 3.09E-10 | 0.739928 |
| FAM83E | 59.15085 | -2.28773 | 0.341836 | -6.69248 | 2.19E-11 | 3.13E-10 | 0.204798 |
| MSL3P1 | 245.9523 | -0.89916 | 0.134407 | -6.68981 | 2.23E-11 | 3.19E-10 | 0.536199 |
| CTSD | 6139.623 | -0.91881 | 0.137403 | -6.68695 | 2.28E-11 | 3.25E-10 | 0.528945 |
| TGFA | 162.1671 | -1.7117 | 0.256032 | -6.68548 | 2.30E-11 | 3.28E-10 | 0.305301 |
| AIFM2 | 152.7087 | -1.20352 | 0.180098 | -6.68261 | 2.35E-11 | 3.34E-10 | 0.434213 |
| AC140479 | 29.2008 | -2.69319 | 0.403096 | -6.68125 | 2.37E-11 | 3.37E-10 | 0.154622 |
| SLC16A5 | 335.6185 | -0.74487 | 0.111535 | -6.67832 | 2.42E-11 | 3.43E-10 | 0.596722 |
| TNFRSF1A | 1633.436 | -0.51642 | 0.077331 | -6.67807 | 2.42E-11 | 3.43E-10 | 0.699102 |
| GRPR | 237.3797 | -1.06099 | 0.158953 | -6.67488 | 2.47E-11 | 3.50E-10 | 0.479304 |
| ALPG | 52.49022 | -2.55024 | 0.382137 | -6.67362 | 2.50E-11 | 3.53E-10 | 0.170727 |
| CHST4 | 183.8513 | -1.30566 | 0.195758 | -6.66979 | 2.56E-11 | 3.62E-10 | 0.404535 |
| ARTN | 109.7523 | -2.35802 | 0.353673 | -6.66724 | 2.61E-11 | 3.68E-10 | 0.195058 |
| PHLDA1 | 1918.839 | -1.02918 | 0.154719 | -6.65191 | 2.89E-11 | 4.07E-10 | 0.489989 |
| KCNF1 | 39.49115 | -2.44526 | 0.367779 | -6.64873 | 2.96E-11 | 4.16E-10 | 0.183613 |

| | | | | | | | |
|---|---|---|---|---|---|---|---|
| TMX2 | 2045.399 | -0.49983 | 0.075192 | -6.64745 | 2.98E-11 | 4.19E-10 | 0.707188 |
| NPW | 70.4619 | -1.8966 | 0.2856 | -6.64077 | 3.12E-11 | 4.37E-10 | 0.268575 |
| SHD | 44.8663 | -2.96979 | 0.447436 | -6.63735 | 3.19E-11 | 4.47E-10 | 0.127645 |
| FXYD7 | 98.35166 | -1.39123 | 0.209839 | -6.62999 | 3.36E-11 | 4.69E-10 | 0.381239 |
| DLGAP3 | 213.975 | -1.63233 | 0.246363 | -6.62571 | 3.46E-11 | 4.82E-10 | 0.322566 |
| ADCY2 | 776.0734 | -1.07924 | 0.162947 | -6.62323 | 3.51E-11 | 4.89E-10 | 0.473279 |
| RYR1 | 1178.415 | -1.3545 | 0.20459 | -6.62058 | 3.58E-11 | 4.98E-10 | 0.39107 |
| IGFBP6 | 75.02554 | -1.94728 | 0.294266 | -6.61741 | 3.66E-11 | 5.08E-10 | 0.259305 |
| GRIPAP1 | 1425.173 | -0.76491 | 0.115705 | -6.61086 | 3.82E-11 | 5.30E-10 | 0.588491 |
| HPSE | 80.48266 | -1.95926 | 0.296452 | -6.60903 | 3.87E-11 | 5.36E-10 | 0.25716 |
| VAV1 | 43.26868 | -2.34663 | 0.355835 | -6.59472 | 4.26E-11 | 5.90E-10 | 0.196605 |
| SYPL2 | 58.26729 | -2.23593 | 0.339414 | -6.58762 | 4.47E-11 | 6.18E-10 | 0.212284 |
| CHST13 | 64.15989 | -2.00497 | 0.304546 | -6.58347 | 4.60E-11 | 6.35E-10 | 0.24914 |
| KIF5C | 3201.313 | -0.71706 | 0.109025 | -6.57703 | 4.80E-11 | 6.60E-10 | 0.608335 |
| SLC49A3 | 109.752 | -1.34135 | 0.20399 | -6.57556 | 4.85E-11 | 6.66E-10 | 0.394651 |
| PDCL3P4 | 26.90628 | -3.13681 | 0.477209 | -6.57326 | 4.92E-11 | 6.76E-10 | 0.113691 |
| CYB561 | 893.1013 | -0.67031 | 0.102187 | -6.55964 | 5.39E-11 | 7.39E-10 | 0.628372 |
| TLCD2 | 120.1555 | -1.15574 | 0.17622 | -6.5585 | 5.44E-11 | 7.44E-10 | 0.448835 |
| E2F1 | 901.6339 | -0.51355 | 0.078336 | -6.55576 | 5.54E-11 | 7.58E-10 | 0.700497 |
| RUNDC3A | 101.5701 | -1.48294 | 0.22632 | -6.55241 | 5.66E-11 | 7.74E-10 | 0.357759 |
| ESYT2 | 2129.579 | -0.60992 | 0.093136 | -6.5487 | 5.80E-11 | 7.93E-10 | 0.655232 |
| VWA1 | 1143.28 | -1.0896 | 0.166524 | -6.54322 | 6.02E-11 | 8.21E-10 | 0.46989 |
| PLEKHJ1 | 2176.642 | -0.49704 | 0.075971 | -6.54253 | 6.05E-11 | 8.25E-10 | 0.708558 |
| HK1 | 4943.194 | -0.82543 | 0.126184 | -6.5415 | 6.09E-11 | 8.30E-10 | 0.564313 |
| H2AFJ | 241.9722 | -1.17646 | 0.179935 | -6.53823 | 6.23E-11 | 8.46E-10 | 0.442436 |
| AC139256 | 196.8794 | -1.13217 | 0.17319 | -6.53713 | 6.27E-11 | 8.51E-10 | 0.456229 |
| HLA-DOA | 543.4576 | -0.83804 | 0.128198 | -6.53709 | 6.27E-11 | 8.51E-10 | 0.559403 |
| SLC35E3 | 494.6651 | -1.20829 | 0.184887 | -6.53527 | 6.35E-11 | 8.61E-10 | 0.432781 |
| EPS8L2 | 600.823 | -1.2509 | 0.191504 | -6.53197 | 6.49E-11 | 8.78E-10 | 0.420186 |
| ANK3 | 1573.259 | -0.57621 | 0.088257 | -6.52869 | 6.63E-11 | 8.95E-10 | 0.670725 |
| KIAA0319L | 2645.752 | -0.64519 | 0.098993 | -6.51746 | 7.15E-11 | 9.61E-10 | 0.639411 |
| LRIG2 | 1459.219 | -0.65094 | 0.099926 | -6.51418 | 7.31E-11 | 9.82E-10 | 0.636866 |
| RFNG | 813.2742 | -0.93322 | 0.143264 | -6.51396 | 7.32E-11 | 9.83E-10 | 0.523689 |
| LUZP2 | 102.2007 | -1.64595 | 0.252824 | -6.51025 | 7.50E-11 | 1.01E-09 | 0.319536 |
| ACSS1 | 95.19269 | -1.44671 | 0.222248 | -6.50941 | 7.54E-11 | 1.01E-09 | 0.366858 |
| PLPPR4 | 110.4917 | -2.18373 | 0.33589 | -6.50131 | 7.96E-11 | 1.06E-09 | 0.220106 |
| GET4 | 1998.089 | -0.47404 | 0.072921 | -6.50066 | 8.00E-11 | 1.07E-09 | 0.719947 |
| MMP24OS | 476.4713 | -0.61282 | 0.094397 | -6.492 | 8.47E-11 | 1.13E-09 | 0.653915 |
| PSD | 182.6667 | -1.83559 | 0.282794 | -6.49089 | 8.53E-11 | 1.13E-09 | 0.280178 |
| HIST1H2A( | 197.8055 | -2.58557 | 0.398475 | -6.48865 | 8.66E-11 | 1.15E-09 | 0.166597 |
| ERBB2 | 7495.176 | -0.56932 | 0.087766 | -6.48681 | 8.77E-11 | 1.16E-09 | 0.673935 |
| STAC2 | 77.74142 | -1.3611 | 0.21002 | -6.48083 | 9.12E-11 | 1.20E-09 | 0.389285 |
| ZNF204P | 322.1583 | -0.83071 | 0.128201 | -6.47977 | 9.19E-11 | 1.21E-09 | 0.562252 |
| SOCS3 | 268.8406 | -1.04394 | 0.161137 | -6.47856 | 9.26E-11 | 1.22E-09 | 0.485002 |

| | | | | | | | |
|---|---|---|---|---|---|---|---|
| GTF2IP13 | 302.1089 | -0.92498 | 0.142832 | -6.476 | 9.42E-11 | 1.24E-09 | 0.526688 |
| CSRP1 | 20.15634 | -4.59773 | 0.71017 | -6.47412 | 9.54E-11 | 1.25E-09 | 0.0413 |
| NECTIN2 | 4607.073 | -0.91658 | 0.141751 | -6.46612 | 1.01E-10 | 1.32E-09 | 0.529764 |
| ANK1 | 159.0763 | -1.57479 | 0.243669 | -6.46283 | 1.03E-10 | 1.35E-09 | 0.335692 |
| CYB5R2 | 415.9592 | -0.67338 | 0.104226 | -6.46077 | 1.04E-10 | 1.36E-09 | 0.627035 |
| OVOL2 | 115.1563 | -1.51955 | 0.235212 | -6.46032 | 1.04E-10 | 1.36E-09 | 0.348796 |
| OVOL1 | 57.50956 | -2.30347 | 0.356578 | -6.45993 | 1.05E-10 | 1.37E-09 | 0.202575 |
| APOE | 22580.33 | -1.06337 | 0.164642 | -6.45868 | 1.06E-10 | 1.38E-09 | 0.478514 |
| CEBPZ | 4943.98 | -0.52203 | 0.080831 | -6.45838 | 1.06E-10 | 1.38E-09 | 0.696389 |
| NPIPB12 | 1353.856 | -1.04109 | 0.161315 | -6.4538 | 1.09E-10 | 1.42E-09 | 0.485959 |
| C16orf54 | 97.00101 | -2.06813 | 0.320522 | -6.45237 | 1.10E-10 | 1.43E-09 | 0.238469 |
| DDOST | 7107.426 | -0.63689 | 0.098721 | -6.45143 | 1.11E-10 | 1.44E-09 | 0.643098 |
| COL27A1 | 864.9921 | -0.92931 | 0.144094 | -6.4493 | 1.12E-10 | 1.46E-09 | 0.525111 |
| C1S | 227.7144 | -1.56878 | 0.243298 | -6.448 | 1.13E-10 | 1.47E-09 | 0.337092 |
| HTR2C | 161.4524 | -1.05354 | 0.163453 | -6.44552 | 1.15E-10 | 1.49E-09 | 0.481785 |
| KLK10 | 102.0572 | -1.31401 | 0.203893 | -6.44462 | 1.16E-10 | 1.50E-09 | 0.402201 |
| ZNF44 | 20.29264 | -5.03873 | 0.782442 | -6.43975 | 1.20E-10 | 1.55E-09 | 0.030422 |
| IL17C | 19.73677 | -4.24915 | 0.660159 | -6.43655 | 1.22E-10 | 1.58E-09 | 0.052587 |
| G6PC3 | 2654.552 | -0.70044 | 0.109006 | -6.42567 | 1.31E-10 | 1.69E-09 | 0.615385 |
| ZSCAN2 | 337.0921 | -0.90518 | 0.140966 | -6.42123 | 1.35E-10 | 1.73E-09 | 0.533967 |
| LMAN2 | 2658.328 | -0.69001 | 0.107479 | -6.41997 | 1.36E-10 | 1.75E-09 | 0.619848 |
| LMNA | 1030.647 | -0.79007 | 0.123076 | -6.41938 | 1.37E-10 | 1.75E-09 | 0.578316 |
| PHETA2 | 23.43029 | -3.53504 | 0.551033 | -6.4153 | 1.41E-10 | 1.80E-09 | 0.086268 |
| SPATS2L | 1533.623 | -1.59416 | 0.248689 | -6.41024 | 1.45E-10 | 1.85E-09 | 0.331215 |
| TUBG2 | 394.4769 | -0.96017 | 0.149832 | -6.40829 | 1.47E-10 | 1.87E-09 | 0.513998 |
| VIL1 | 457.1746 | -0.92381 | 0.144437 | -6.39594 | 1.60E-10 | 2.02E-09 | 0.527116 |
| CCL26 | 39.52674 | -3.53334 | 0.553123 | -6.38798 | 1.68E-10 | 2.13E-09 | 0.086369 |
| ADAMTS14 | 94.81098 | -1.79227 | 0.280655 | -6.38602 | 1.70E-10 | 2.15E-09 | 0.288718 |
| ACAN | 73.20069 | -2.26054 | 0.35453 | -6.37617 | 1.82E-10 | 2.29E-09 | 0.208693 |
| TIMP4 | 143.3957 | -1.41841 | 0.222518 | -6.37437 | 1.84E-10 | 2.31E-09 | 0.374124 |
| TKT | 14510.26 | -0.44374 | 0.069706 | -6.36585 | 1.94E-10 | 2.44E-09 | 0.735228 |
| SERPINH1 | 11885.79 | -0.60179 | 0.094572 | -6.36329 | 1.97E-10 | 2.47E-09 | 0.658935 |
| CYP27A1 | 354.483 | -0.95664 | 0.150382 | -6.36137 | 2.00E-10 | 2.50E-09 | 0.515256 |
| LAPTM4B | 11247.46 | -0.44535 | 0.070037 | -6.35883 | 2.03E-10 | 2.54E-09 | 0.734405 |
| LRP8 | 2126.769 | -0.62512 | 0.098356 | -6.35568 | 2.08E-10 | 2.59E-09 | 0.648366 |
| CYC1 | 2897.292 | -0.44411 | 0.069942 | -6.3497 | 2.16E-10 | 2.68E-09 | 0.735038 |
| TMCO4 | 84.01161 | -1.8541 | 0.291994 | -6.34979 | 2.16E-10 | 2.68E-09 | 0.276605 |
| CBLC | 244.3094 | -1.28751 | 0.20277 | -6.34959 | 2.16E-10 | 2.68E-09 | 0.409658 |
| TRIM14 | 404.5505 | -0.84269 | 0.133006 | -6.33568 | 2.36E-10 | 2.92E-09 | 0.557605 |
| SLC45A3 | 460.3122 | -0.90579 | 0.14304 | -6.33245 | 2.41E-10 | 2.98E-09 | 0.53374 |
| TESMIN | 555.442 | -0.98676 | 0.155874 | -6.33046 | 2.44E-10 | 3.02E-09 | 0.504611 |
| PACS2 | 1882.955 | -0.73724 | 0.11651 | -6.32771 | 2.49E-10 | 3.07E-09 | 0.599885 |
| SLC38A9 | 973.8333 | -0.57649 | 0.091152 | -6.32448 | 2.54E-10 | 3.13E-09 | 0.670595 |
| PROCR | 504.5754 | -1.22117 | 0.1932 | -6.32074 | 2.60E-10 | 3.20E-09 | 0.428936 |

| | | | | | | | |
|---|---|---|---|---|---|---|---|
| PIK3R6 | 52.17312 | -1.91649 | 0.303255 | -6.31975 | 2.62E-10 | 3.22E-09 | 0.264897 |
| EXOC3L1 | 30.62496 | -2.62321 | 0.415412 | -6.31471 | 2.71E-10 | 3.32E-09 | 0.162306 |
| CDC42EP1 | 600.1613 | -1.23977 | 0.196343 | -6.31432 | 2.71E-10 | 3.33E-09 | 0.42344 |
| SPIB | 161.7542 | -2.55826 | 0.40533 | -6.31155 | 2.76E-10 | 3.38E-09 | 0.16978 |
| HCN2 | 84.56931 | -1.37903 | 0.218514 | -6.31096 | 2.77E-10 | 3.39E-09 | 0.384477 |
| INSYN1 | 151.8134 | -1.71136 | 0.271218 | -6.30992 | 2.79E-10 | 3.41E-09 | 0.305372 |
| CYP2B6 | 34.7977 | -2.38644 | 0.378269 | -6.30883 | 2.81E-10 | 3.43E-09 | 0.191254 |
| GDF15 | 163.9056 | -1.88204 | 0.298328 | -6.30862 | 2.82E-10 | 3.43E-09 | 0.2713 |
| ST8SIA5 | 65.61274 | -3.04876 | 0.483787 | -6.30188 | 2.94E-10 | 3.58E-09 | 0.120845 |
| ACADVL | 3821.035 | -1.15943 | 0.184036 | -6.29999 | 2.98E-10 | 3.62E-09 | 0.447691 |
| SLC35F2 | 765.8172 | -0.64965 | 0.103168 | -6.29701 | 3.03E-10 | 3.68E-09 | 0.637436 |
| ABCC9 | 267.3464 | -0.80302 | 0.127652 | -6.29069 | 3.16E-10 | 3.83E-09 | 0.573148 |
| AC005840 | 244.6708 | -0.90932 | 0.144603 | -6.28836 | 3.21E-10 | 3.88E-09 | 0.532437 |
| RIMS4 | 1473.047 | -0.55742 | 0.0887 | -6.28436 | 3.29E-10 | 3.98E-09 | 0.679515 |
| TSGA10IP | 97.62063 | -1.3813 | 0.219972 | -6.27943 | 3.40E-10 | 4.10E-09 | 0.383873 |
| TFCP2L1 | 137.0921 | -2.24858 | 0.358225 | -6.277 | 3.45E-10 | 4.16E-09 | 0.210431 |
| ATP13A2 | 2919.657 | -0.85831 | 0.136804 | -6.27403 | 3.52E-10 | 4.23E-09 | 0.551598 |
| VN1R51P | 28.17762 | -3.00287 | 0.479064 | -6.26821 | 3.65E-10 | 4.39E-09 | 0.124752 |
| CYP2U1 | 569.9142 | -0.63617 | 0.101551 | -6.26453 | 3.74E-10 | 4.48E-09 | 0.643418 |
| KCNK1 | 420.8884 | -0.86821 | 0.13868 | -6.26051 | 3.84E-10 | 4.59E-09 | 0.547827 |
| ALDH1A3 | 140.0639 | -1.35261 | 0.216082 | -6.2597 | 3.86E-10 | 4.61E-09 | 0.391584 |
| S100A11 | 945.2757 | -0.76937 | 0.122928 | -6.25873 | 3.88E-10 | 4.64E-09 | 0.586672 |
| AC010507 | 55.30743 | -2.37929 | 0.380357 | -6.2554 | 3.96E-10 | 4.72E-09 | 0.192205 |
| RAB15 | 1398.256 | -0.58911 | 0.094226 | -6.25211 | 4.05E-10 | 4.81E-09 | 0.664751 |
| KIAA0040 | 395.8396 | -1.22384 | 0.195843 | -6.24908 | 4.13E-10 | 4.90E-09 | 0.428142 |
| FKBP10 | 5891.995 | -0.81854 | 0.131048 | -6.24616 | 4.21E-10 | 4.98E-09 | 0.567014 |
| EBI3 | 24.80655 | -3.39786 | 0.544156 | -6.24428 | 4.26E-10 | 5.04E-09 | 0.094873 |
| NCLN | 2915.296 | -0.71031 | 0.113803 | -6.24156 | 4.33E-10 | 5.12E-09 | 0.61119 |
| HSD17B12 | 2761.064 | -0.48906 | 0.078386 | -6.23911 | 4.40E-10 | 5.19E-09 | 0.712489 |
| SCNN1D | 183.6364 | -1.53264 | 0.245691 | -6.23806 | 4.43E-10 | 5.22E-09 | 0.345645 |
| YJEFN3 | 474.6551 | -1.58371 | 0.253944 | -6.23644 | 4.48E-10 | 5.27E-09 | 0.333624 |
| DGKZ | 2591.013 | -0.60966 | 0.097775 | -6.23534 | 4.51E-10 | 5.30E-09 | 0.655351 |
| KIFC2 | 619.1996 | -1.4804 | 0.237505 | -6.23312 | 4.57E-10 | 5.37E-09 | 0.35839 |
| AC004156 | 1131.85 | -0.56288 | 0.090378 | -6.22805 | 4.72E-10 | 5.53E-09 | 0.67695 |
| GSE1 | 3566.671 | -0.54733 | 0.087916 | -6.22563 | 4.80E-10 | 5.61E-09 | 0.684285 |
| ACHE | 79.57448 | -1.59301 | 0.255896 | -6.22521 | 4.81E-10 | 5.62E-09 | 0.33148 |
| NRP1 | 166.361 | -1.21746 | 0.195662 | -6.22223 | 4.90E-10 | 5.73E-09 | 0.43004 |
| TMEM82 | 57.05532 | -1.72232 | 0.277093 | -6.21567 | 5.11E-10 | 5.95E-09 | 0.303062 |
| SLC39A14 | 7134.657 | -0.67206 | 0.108205 | -6.21096 | 5.27E-10 | 6.13E-09 | 0.627611 |
| PDLIM2 | 389.0085 | -0.90057 | 0.145104 | -6.20637 | 5.42E-10 | 6.30E-09 | 0.535674 |
| FUCA2 | 2101.431 | -0.53345 | 0.085977 | -6.20454 | 5.49E-10 | 6.37E-09 | 0.690903 |
| SH3BP2 | 1128.235 | -0.51588 | 0.083176 | -6.20231 | 5.56E-10 | 6.46E-09 | 0.699365 |
| NPIPB11 | 163.0707 | -1.38374 | 0.223191 | -6.19981 | 5.65E-10 | 6.54E-09 | 0.383224 |
| RGS11 | 68.85815 | -2.06147 | 0.332539 | -6.1992 | 5.68E-10 | 6.56E-09 | 0.239571 |

| | | | | | | | |
|---|---|---|---|---|---|---|---|
| CDIP1 | 787.091 | -0.60014 | 0.096923 | -6.19195 | 5.94E-10 | 6.84E-09 | 0.659688 |
| MT-ATP8 | 13250.68 | -1.06726 | 0.17236 | -6.19204 | 5.94E-10 | 6.84E-09 | 0.477224 |
| ADAM8 | 98.89513 | -1.58815 | 0.256623 | -6.18866 | 6.07E-10 | 6.97E-09 | 0.332597 |
| CACNA1C | 50.20475 | -2.15754 | 0.348685 | -6.18764 | 6.11E-10 | 7.02E-09 | 0.224139 |
| UNC5A | 572.5461 | -0.95797 | 0.154829 | -6.1873 | 6.12E-10 | 7.02E-09 | 0.51478 |
| FCHSD1 | 588.8438 | -1.14711 | 0.185438 | -6.18596 | 6.17E-10 | 7.07E-09 | 0.451528 |
| RAB24 | 357.2769 | -0.76818 | 0.124207 | -6.1847 | 6.22E-10 | 7.12E-09 | 0.587156 |
| STK10 | 500.3602 | -0.92988 | 0.150458 | -6.18032 | 6.40E-10 | 7.32E-09 | 0.524903 |
| NLGN4X | 2675.205 | -0.56718 | 0.091788 | -6.17926 | 6.44E-10 | 7.36E-09 | 0.674934 |
| BICD1 | 933.8451 | -0.51104 | 0.082715 | -6.17832 | 6.48E-10 | 7.40E-09 | 0.701718 |
| TRIP6 | 2154.812 | -0.53984 | 0.087488 | -6.17046 | 6.81E-10 | 7.76E-09 | 0.687847 |
| TES | 672.4076 | -1.10977 | 0.179934 | -6.16765 | 6.93E-10 | 7.89E-09 | 0.463369 |
| TM4SF18 | 102.6481 | -1.31323 | 0.21294 | -6.16711 | 6.96E-10 | 7.91E-09 | 0.40242 |
| TPRA1 | 485.4564 | -0.61323 | 0.099449 | -6.1663 | 6.99E-10 | 7.95E-09 | 0.653731 |
| PILRB | 1375.114 | -1.17434 | 0.190497 | -6.16459 | 7.07E-10 | 8.03E-09 | 0.443088 |
| ANO1 | 276.9839 | -1.19283 | 0.193599 | -6.16136 | 7.21E-10 | 8.18E-09 | 0.437443 |
| SLC7A10 | 77.15881 | -1.83559 | 0.298001 | -6.15966 | 7.29E-10 | 8.26E-09 | 0.280178 |
| SRPX | 280.7676 | -0.83447 | 0.135572 | -6.1552 | 7.50E-10 | 8.49E-09 | 0.560788 |
| TRPC6 | 53.51909 | -2.58536 | 0.42008 | -6.15445 | 7.53E-10 | 8.52E-09 | 0.16662 |
| ADGRD1 | 83.51238 | -2.43689 | 0.396257 | -6.14976 | 7.76E-10 | 8.77E-09 | 0.184682 |
| RFLNB | 487.8518 | -1.05693 | 0.17189 | -6.14889 | 7.80E-10 | 8.81E-09 | 0.480653 |
| PRKCH | 152.1401 | -1.59039 | 0.258777 | -6.14578 | 7.96E-10 | 8.98E-09 | 0.332082 |
| TMEM26 | 59.34057 | -2.16181 | 0.351786 | -6.14523 | 7.98E-10 | 9.00E-09 | 0.223476 |
| TPCN2 | 515.881 | -0.79687 | 0.129734 | -6.14229 | 8.13E-10 | 9.15E-09 | 0.575598 |
| LIPA | 1159.51 | -0.62373 | 0.101553 | -6.14185 | 8.16E-10 | 9.16E-09 | 0.648993 |
| DOCK6 | 1310.43 | -0.98639 | 0.160653 | -6.13986 | 8.26E-10 | 9.27E-09 | 0.50474 |
| ACOT8 | 403.3398 | -0.62446 | 0.101744 | -6.13762 | 8.38E-10 | 9.39E-09 | 0.648661 |
| SCAP | 3467.56 | -0.7068 | 0.115175 | -6.13676 | 8.42E-10 | 9.44E-09 | 0.612677 |
| RNF167 | 2663.695 | -0.56512 | 0.092136 | -6.13351 | 8.60E-10 | 9.62E-09 | 0.6759 |
| ST6GAL1 | 2655.577 | -0.50942 | 0.083054 | -6.13361 | 8.59E-10 | 9.62E-09 | 0.702505 |
| PIEZO1 | 2777.12 | -1.3757 | 0.224348 | -6.13196 | 8.68E-10 | 9.69E-09 | 0.385367 |
| TINAGL1 | 106.6891 | -1.66107 | 0.271285 | -6.12298 | 9.18E-10 | 1.02E-08 | 0.316204 |
| KCNK17 | 59.58266 | -1.88174 | 0.307396 | -6.12154 | 9.27E-10 | 1.03E-08 | 0.271357 |
| PNPLA2 | 1130.58 | -1.06399 | 0.173857 | -6.11994 | 9.36E-10 | 1.04E-08 | 0.478307 |
| MMP19 | 137.8808 | -1.3535 | 0.22124 | -6.11778 | 9.49E-10 | 1.05E-08 | 0.391341 |
| PDGFB | 113.0823 | -2.25441 | 0.368577 | -6.11653 | 9.56E-10 | 1.06E-08 | 0.209582 |
| ESRP1 | 2776.946 | -0.70169 | 0.114771 | -6.11381 | 9.73E-10 | 1.08E-08 | 0.614852 |
| AL669831. | 2478.831 | -0.85345 | 0.139653 | -6.1112 | 9.89E-10 | 1.09E-08 | 0.55346 |
| SLC22A23 | 802.621 | -0.78507 | 0.128491 | -6.1099 | 9.97E-10 | 1.10E-08 | 0.580325 |
| TUBB4A | 460.8983 | -0.7532 | 0.123296 | -6.1089 | 1.00E-09 | 1.11E-08 | 0.593284 |
| ABHD12 | 1622.558 | -0.57296 | 0.093797 | -6.10848 | 1.01E-09 | 1.11E-08 | 0.672237 |
| MYC | 774.5877 | -1.07877 | 0.176623 | -6.10777 | 1.01E-09 | 1.11E-08 | 0.473431 |
| TMEM74B | 160.8183 | -1.8989 | 0.311052 | -6.10476 | 1.03E-09 | 1.13E-08 | 0.268148 |
| KIF12 | 51.43096 | -2.28333 | 0.374048 | -6.10437 | 1.03E-09 | 1.14E-08 | 0.205423 |

| | | | | | | | |
|---|---|---|---|---|---|---|---|
| PDIA5 | 596.4953 | -1.04667 | 0.171776 | -6.0932 | 1.11E-09 | 1.21E-08 | 0.484086 |
| ENOSF1 | 626.7115 | -0.56255 | 0.092513 | -6.08075 | 1.20E-09 | 1.31E-08 | 0.677105 |
| EPB41L4B | 605.5411 | -0.86787 | 0.142748 | -6.0797 | 1.20E-09 | 1.31E-08 | 0.547957 |
| LCP1 | 60.61008 | -1.87184 | 0.307891 | -6.07954 | 1.21E-09 | 1.31E-08 | 0.273225 |
| NUDT16L1 | 753.6078 | -0.5952 | 0.097909 | -6.07911 | 1.21E-09 | 1.32E-08 | 0.661953 |
| AACS | 985.002 | -0.49411 | 0.081299 | -6.07765 | 1.22E-09 | 1.33E-08 | 0.71 |
| TMTC1 | 808.9405 | -0.53311 | 0.087845 | -6.06871 | 1.29E-09 | 1.40E-08 | 0.691065 |
| ITGA3 | 863.0702 | -1.03128 | 0.169946 | -6.06829 | 1.29E-09 | 1.40E-08 | 0.489275 |
| DLL3 | 107.4019 | -1.22072 | 0.201248 | -6.06574 | 1.31E-09 | 1.42E-08 | 0.429069 |
| DOCK9 | 466.655 | -1.20765 | 0.199091 | -6.06581 | 1.31E-09 | 1.42E-08 | 0.432974 |
| OSBPL10 | 914.6618 | -0.96737 | 0.159477 | -6.06588 | 1.31E-09 | 1.42E-08 | 0.511437 |
| ARFRP1 | 558.119 | -0.65493 | 0.108035 | -6.06219 | 1.34E-09 | 1.45E-08 | 0.635105 |
| SGSM2 | 1842.006 | -1.27175 | 0.209871 | -6.05969 | 1.36E-09 | 1.47E-08 | 0.414156 |
| PCDHGA1( | 94.03901 | -1.37752 | 0.227786 | -6.04743 | 1.47E-09 | 1.58E-08 | 0.384879 |
| IFRD2 | 1209.793 | -0.44983 | 0.074485 | -6.03914 | 1.55E-09 | 1.66E-08 | 0.732131 |
| ADRA2B | 217.363 | -0.87436 | 0.144831 | -6.03712 | 1.57E-09 | 1.68E-08 | 0.545496 |
| JAKMIP3 | 79.56911 | -1.72674 | 0.286186 | -6.03362 | 1.60E-09 | 1.72E-08 | 0.302134 |
| ADAP1 | 158.5259 | -2.12594 | 0.352407 | -6.03262 | 1.61E-09 | 1.73E-08 | 0.229102 |
| AIFM1 | 1469.007 | -0.49065 | 0.081492 | -6.02082 | 1.74E-09 | 1.85E-08 | 0.711705 |
| ETS1 | 346.7656 | -0.96561 | 0.160462 | -6.01769 | 1.77E-09 | 1.88E-08 | 0.512062 |
| PLEKHO2 | 700.7805 | -0.54245 | 0.090167 | -6.01605 | 1.79E-09 | 1.90E-08 | 0.686605 |
| SGPP1 | 404.0385 | -0.63098 | 0.104923 | -6.01372 | 1.81E-09 | 1.93E-08 | 0.645739 |
| C19orf24 | 402.5126 | -0.77222 | 0.128554 | -6.00697 | 1.89E-09 | 2.00E-08 | 0.585516 |
| RAB20 | 184.7531 | -1.12715 | 0.187677 | -6.00579 | 1.90E-09 | 2.02E-08 | 0.457819 |
| ANXA11 | 1168.647 | -0.94195 | 0.156876 | -6.00442 | 1.92E-09 | 2.03E-08 | 0.520528 |
| BCAP31 | 2635.122 | -0.59199 | 0.098595 | -6.0043 | 1.92E-09 | 2.03E-08 | 0.663427 |
| ATXN2L | 6609.647 | -0.49122 | 0.08183 | -6.00299 | 1.94E-09 | 2.05E-08 | 0.711421 |
| CDHR1 | 187.9619 | -1.22505 | 0.204072 | -6.00302 | 1.94E-09 | 2.05E-08 | 0.427783 |
| CASR | 237.6419 | -1.05122 | 0.175156 | -6.00164 | 1.95E-09 | 2.06E-08 | 0.482559 |
| SLC27A2 | 220.5937 | -1.15499 | 0.192605 | -5.99665 | 2.01E-09 | 2.12E-08 | 0.44907 |
| CCND3 | 444.2513 | -0.74838 | 0.124873 | -5.99311 | 2.06E-09 | 2.16E-08 | 0.595272 |
| FGF21 | 50.57239 | -1.78331 | 0.297743 | -5.98941 | 2.11E-09 | 2.21E-08 | 0.290517 |
| CTHRC1 | 10.97436 | -6.92572 | 1.156715 | -5.9874 | 2.13E-09 | 2.24E-08 | 0.008225 |
| SOCS1 | 171.6823 | -1.27892 | 0.213718 | -5.98418 | 2.17E-09 | 2.28E-08 | 0.412102 |
| MEF2B | 287.1265 | -1.50746 | 0.251949 | -5.98317 | 2.19E-09 | 2.29E-08 | 0.351731 |
| GRTP1 | 732.9861 | -0.53609 | 0.089666 | -5.97877 | 2.25E-09 | 2.35E-08 | 0.689636 |
| SYK | 84.41838 | -1.46 | 0.244367 | -5.97464 | 2.31E-09 | 2.40E-08 | 0.363492 |
| MFSD10 | 2544.119 | -1.12848 | 0.189029 | -5.96986 | 2.37E-09 | 2.47E-08 | 0.457398 |
| DOK3 | 103.1539 | -1.32679 | 0.222339 | -5.96744 | 2.41E-09 | 2.50E-08 | 0.398653 |
| ZMIZ2 | 2108.45 | -0.77572 | 0.130037 | -5.96539 | 2.44E-09 | 2.53E-08 | 0.584097 |
| MYORG | 195.2387 | -1.13801 | 0.190789 | -5.96478 | 2.45E-09 | 2.54E-08 | 0.454385 |
| CEBPD | 71.96938 | -1.87434 | 0.31445 | -5.96069 | 2.51E-09 | 2.60E-08 | 0.272752 |
| MRPL4 | 1322.121 | -0.52247 | 0.087689 | -5.95821 | 2.55E-09 | 2.64E-08 | 0.696179 |
| CDC42EP2 | 158.8606 | -1.23814 | 0.207939 | -5.95434 | 2.61E-09 | 2.70E-08 | 0.423919 |

| | | | | | | | |
|---|---|---|---|---|---|---|---|
| COL12A1 | 152.3969 | -1.41725 | 0.23814 | -5.95132 | 2.66E-09 | 2.75E-08 | 0.374427 |
| UBE2S | 3377.322 | -0.50873 | 0.085486 | -5.95103 | 2.66E-09 | 2.75E-08 | 0.702841 |
| HCK | 46.53558 | -2.26878 | 0.381291 | -5.95026 | 2.68E-09 | 2.76E-08 | 0.207505 |
| PCK2 | 7333.662 | -0.5318 | 0.08939 | -5.94927 | 2.69E-09 | 2.78E-08 | 0.691689 |
| SIGMAR1 | 2234.749 | -0.42486 | 0.071421 | -5.94873 | 2.70E-09 | 2.78E-08 | 0.744909 |
| MFSD6L | 53.82203 | -2.17052 | 0.36492 | -5.94794 | 2.72E-09 | 2.80E-08 | 0.22213 |
| PPM1J | 297.8692 | -1.00983 | 0.169821 | -5.94646 | 2.74E-09 | 2.82E-08 | 0.496603 |
| MIGA2 | 1008.437 | -1.01246 | 0.170415 | -5.94118 | 2.83E-09 | 2.91E-08 | 0.495699 |
| TAP1 | 264.4532 | -1.31692 | 0.221677 | -5.94072 | 2.84E-09 | 2.92E-08 | 0.401391 |
| GCNT2 | 910.6128 | -0.64081 | 0.107927 | -5.93739 | 2.90E-09 | 2.97E-08 | 0.641355 |
| CTSA | 1076.633 | -0.92208 | 0.155358 | -5.93519 | 2.94E-09 | 3.01E-08 | 0.527747 |
| PANX2 | 165.9141 | -1.17115 | 0.197358 | -5.93416 | 2.95E-09 | 3.03E-08 | 0.444066 |
| CD151 | 1870.425 | -0.87877 | 0.148112 | -5.93316 | 2.97E-09 | 3.04E-08 | 0.543831 |
| FCER2 | 31.7036 | -2.062 | 0.347709 | -5.93025 | 3.02E-09 | 3.10E-08 | 0.239484 |
| GRK2 | 1936.406 | -0.46905 | 0.079109 | -5.92914 | 3.05E-09 | 3.11E-08 | 0.722442 |
| NDUFV1 | 1906.882 | -0.42569 | 0.071856 | -5.92427 | 3.14E-09 | 3.20E-08 | 0.74448 |
| SLC30A3 | 203.638 | -1.32263 | 0.223257 | -5.92425 | 3.14E-09 | 3.20E-08 | 0.399805 |
| COASY | 1610.476 | -0.50493 | 0.08524 | -5.92367 | 3.15E-09 | 3.21E-08 | 0.704692 |
| ETFB | 659.3228 | -0.57314 | 0.096782 | -5.92196 | 3.18E-09 | 3.24E-08 | 0.672154 |
| TIMM13 | 1854.25 | -0.49296 | 0.083251 | -5.92139 | 3.19E-09 | 3.25E-08 | 0.710565 |
| INAFM2 | 581.436 | -0.55693 | 0.09407 | -5.92034 | 3.21E-09 | 3.26E-08 | 0.679748 |
| TUNAR | 1448.393 | -0.49597 | 0.083837 | -5.91585 | 3.30E-09 | 3.35E-08 | 0.709085 |
| GPR78 | 51.28066 | -2.25123 | 0.380779 | -5.91216 | 3.38E-09 | 3.42E-08 | 0.210045 |
| GRIN3B | 72.75044 | -1.74949 | 0.296006 | -5.91031 | 3.41E-09 | 3.46E-08 | 0.297407 |
| SH3YL1 | 437.8361 | -0.79801 | 0.135032 | -5.90977 | 3.43E-09 | 3.47E-08 | 0.575142 |
| TMEM163 | 146.8752 | -1.77403 | 0.30035 | -5.90655 | 3.49E-09 | 3.53E-08 | 0.29239 |
| SLC24A3 | 162.1171 | -1.15226 | 0.195251 | -5.90143 | 3.60E-09 | 3.64E-08 | 0.44992 |
| PLA2R1 | 65.35388 | -1.62473 | 0.275359 | -5.9004 | 3.63E-09 | 3.66E-08 | 0.324271 |
| ENG | 94.18012 | -1.88836 | 0.32012 | -5.89892 | 3.66E-09 | 3.69E-08 | 0.270114 |
| ALDOC | 680.6116 | -0.76123 | 0.129056 | -5.89841 | 3.67E-09 | 3.69E-08 | 0.589994 |
| MT-ND4L | 14800.91 | -1.10341 | 0.187217 | -5.89378 | 3.77E-09 | 3.80E-08 | 0.465414 |
| SBSPON | 103.7554 | -1.53181 | 0.260095 | -5.88942 | 3.88E-09 | 3.89E-08 | 0.345844 |
| SLC4A2 | 2242.035 | -1.16847 | 0.198458 | -5.88774 | 3.92E-09 | 3.92E-08 | 0.444893 |
| PGM1 | 1390.798 | -0.55262 | 0.09387 | -5.88713 | 3.93E-09 | 3.94E-08 | 0.681779 |
| MTA1 | 4684.534 | -0.66224 | 0.112498 | -5.88668 | 3.94E-09 | 3.94E-08 | 0.631896 |
| DAGLB | 796.7127 | -0.582 | 0.099111 | -5.87222 | 4.30E-09 | 4.29E-08 | 0.668037 |
| AC010507 | 42.07876 | -2.78397 | 0.474222 | -5.87061 | 4.34E-09 | 4.33E-08 | 0.145191 |
| TMEM191E | 31.16041 | -3.15142 | 0.536821 | -5.87052 | 4.34E-09 | 4.33E-08 | 0.112546 |
| SLC50A1 | 775.3483 | -0.54251 | 0.092437 | -5.86895 | 4.39E-09 | 4.36E-08 | 0.686577 |
| SLC51A | 85.12374 | -1.82461 | 0.310931 | -5.86821 | 4.41E-09 | 4.38E-08 | 0.282318 |
| SYTL1 | 495.3381 | -0.73172 | 0.124696 | -5.86799 | 4.41E-09 | 4.38E-08 | 0.602187 |
| CPVL | 1091.701 | -0.73981 | 0.126133 | -5.8653 | 4.48E-09 | 4.45E-08 | 0.598819 |
| JUP | 3355.626 | -0.41512 | 0.070788 | -5.86435 | 4.51E-09 | 4.47E-08 | 0.749955 |
| B2M | 1588.705 | -0.71056 | 0.121239 | -5.86083 | 4.61E-09 | 4.56E-08 | 0.611083 |

| | | | | | | | |
|---|---|---|---|---|---|---|---|
| CLDN3 | 642.5483 | -1.12728 | 0.192384 | -5.85949 | 4.64E-09 | 4.60E-08 | 0.457779 |
| COL26A1 | 2558.636 | -0.54867 | 0.09365 | -5.85877 | 4.66E-09 | 4.62E-08 | 0.683649 |
| LINC0114! | 277.443 | -0.79712 | 0.136233 | -5.85114 | 4.88E-09 | 4.82E-08 | 0.575498 |
| NPIPB4 | 324.9651 | -0.88707 | 0.151673 | -5.84855 | 4.96E-09 | 4.89E-08 | 0.540713 |
| SLC43A3 | 671.3744 | -0.63763 | 0.109027 | -5.84835 | 4.96E-09 | 4.90E-08 | 0.642769 |
| MET | 334.9828 | -1.00717 | 0.172286 | -5.84589 | 5.04E-09 | 4.97E-08 | 0.497523 |
| GDAP1L1 | 213.6183 | -1.21236 | 0.207407 | -5.84531 | 5.06E-09 | 4.98E-08 | 0.431563 |
| STAP2 | 213.9072 | -0.89716 | 0.15357 | -5.84203 | 5.16E-09 | 5.08E-08 | 0.536943 |
| YIF1A | 1028.666 | -0.56868 | 0.097375 | -5.84015 | 5.22E-09 | 5.13E-08 | 0.674233 |
| GADD45B | 76.0762 | -1.40464 | 0.240523 | -5.83993 | 5.22E-09 | 5.14E-08 | 0.377713 |
| JAZF1 | 140.8213 | -1.53247 | 0.262528 | -5.83735 | 5.30E-09 | 5.21E-08 | 0.345686 |
| RYR2 | 111.0617 | -1.19191 | 0.204297 | -5.83421 | 5.40E-09 | 5.30E-08 | 0.437723 |
| SLCO4A1 | 224.9373 | -0.93088 | 0.15957 | -5.83368 | 5.42E-09 | 5.32E-08 | 0.524537 |
| CBS | 10718.23 | -0.58011 | 0.099476 | -5.83172 | 5.49E-09 | 5.38E-08 | 0.668911 |
| ATP6V0A1 | 1449.099 | -0.62458 | 0.107214 | -5.82553 | 5.69E-09 | 5.57E-08 | 0.648609 |
| ADRM1 | 2452.754 | -0.50845 | 0.087294 | -5.82464 | 5.72E-09 | 5.59E-08 | 0.702975 |
| XYLT2 | 1136.065 | -0.76702 | 0.1317 | -5.82401 | 5.75E-09 | 5.61E-08 | 0.587629 |
| TBC1D8B | 1448.956 | -0.55033 | 0.09458 | -5.8186 | 5.93E-09 | 5.78E-08 | 0.682866 |
| STAT5A | 164.7431 | -0.86933 | 0.149481 | -5.81567 | 6.04E-09 | 5.87E-08 | 0.5474 |
| NGFR | 458.9766 | -0.7344 | 0.126336 | -5.81307 | 6.13E-09 | 5.96E-08 | 0.601068 |
| HTRA1 | 428.0534 | -1.04942 | 0.180545 | -5.8125 | 6.15E-09 | 5.98E-08 | 0.483164 |
| YIF1B | 1389.154 | -0.64885 | 0.111668 | -5.81056 | 6.23E-09 | 6.05E-08 | 0.637788 |
| SULT1B1 | 504.377 | -0.8623 | 0.148794 | -5.79529 | 6.82E-09 | 6.61E-08 | 0.550073 |
| MINK1 | 2020.669 | -0.69752 | 0.120414 | -5.79267 | 6.93E-09 | 6.71E-08 | 0.616631 |
| ECEL1 | 362.8755 | -1.25777 | 0.217197 | -5.79091 | 7.00E-09 | 6.78E-08 | 0.41819 |
| NDRG4 | 569.5301 | -0.65417 | 0.113061 | -5.78596 | 7.21E-09 | 6.97E-08 | 0.635443 |
| TRMT9B | 39.64166 | -1.85453 | 0.320658 | -5.78351 | 7.32E-09 | 7.07E-08 | 0.276523 |
| CD320 | 1188.645 | -1.02326 | 0.177008 | -5.78085 | 7.43E-09 | 7.17E-08 | 0.492005 |
| HIST3H2A | 109.6164 | -1.51296 | 0.261859 | -5.77775 | 7.57E-09 | 7.30E-08 | 0.350393 |
| PCDHB4 | 41.35952 | -2.21008 | 0.382952 | -5.77118 | 7.87E-09 | 7.57E-08 | 0.216122 |
| ENTPD8 | 27.40224 | -3.01129 | 0.522182 | -5.76675 | 8.08E-09 | 7.74E-08 | 0.124025 |
| CBR1 | 1062.391 | -0.49289 | 0.085518 | -5.76362 | 8.23E-09 | 7.87E-08 | 0.710599 |
| ZNF879 | 13.28701 | -7.18231 | 1.246147 | -5.76361 | 8.23E-09 | 7.87E-08 | 0.006885 |
| SNCG | 113.7077 | -1.10863 | 0.192445 | -5.76076 | 8.37E-09 | 7.99E-08 | 0.463734 |
| EOGT | 357.8565 | -1.11205 | 0.193067 | -5.75991 | 8.42E-09 | 8.02E-08 | 0.462637 |
| CXCL6 | 66.60834 | -1.40169 | 0.243471 | -5.75712 | 8.56E-09 | 8.14E-08 | 0.378486 |
| INHBA | 22.66093 | -3.51738 | 0.61228 | -5.74472 | 9.21E-09 | 8.73E-08 | 0.08733 |
| COL16A1 | 473.4288 | -1.65018 | 0.287459 | -5.74057 | 9.44E-09 | 8.93E-08 | 0.318601 |
| ATP13A1 | 1934.878 | -0.81741 | 0.142747 | -5.72626 | 1.03E-08 | 9.69E-08 | 0.56746 |
| PRKAR1B | 949.3425 | -0.56621 | 0.098917 | -5.7241 | 1.04E-08 | 9.80E-08 | 0.675388 |
| ROBO3 | 396.0193 | -1.15819 | 0.202425 | -5.7216 | 1.06E-08 | 9.94E-08 | 0.448073 |
| AIRE | 15.12815 | -6.688 | 1.169522 | -5.71857 | 1.07E-08 | 1.01E-07 | 0.009699 |
| AL732372. | 985.9878 | -0.52814 | 0.092384 | -5.71681 | 1.09E-08 | 1.02E-07 | 0.693448 |
| RPS9 | 11393.79 | -1.11941 | 0.195903 | -5.71411 | 1.10E-08 | 1.03E-07 | 0.460281 |

| | | | | | | | |
|---|---|---|---|---|---|---|---|
| HLTF | 980.6578 | -0.78162 | 0.136841 | -5.71186 | 1.12E-08 | 1.05E-07 | 0.581714 |
| GRAMD1A | 2459.193 | -0.65989 | 0.115644 | -5.70624 | 1.15E-08 | 1.08E-07 | 0.632926 |
| P4HA2 | 291.4382 | -1.34575 | 0.235942 | -5.70375 | 1.17E-08 | 1.10E-07 | 0.393449 |
| FASTK | 1161.536 | -0.90736 | 0.159096 | -5.70323 | 1.18E-08 | 1.10E-07 | 0.533159 |
| HDAC7 | 1960.487 | -0.70009 | 0.122802 | -5.70096 | 1.19E-08 | 1.11E-07 | 0.615534 |
| SH2D3C | 171.5889 | -1.2273 | 0.215323 | -5.69983 | 1.20E-08 | 1.12E-07 | 0.427115 |
| KCNN1 | 239.0731 | -1.0883 | 0.190988 | -5.69826 | 1.21E-08 | 1.13E-07 | 0.470315 |
| RIMKLB | 1906.725 | -0.53807 | 0.094426 | -5.69831 | 1.21E-08 | 1.13E-07 | 0.688693 |
| ENO3 | 322.119 | -1.06812 | 0.18747 | -5.69757 | 1.22E-08 | 1.13E-07 | 0.476939 |
| PHF21B | 758.9812 | -0.80589 | 0.141485 | -5.6959 | 1.23E-08 | 1.14E-07 | 0.572011 |
| MST1P2 | 37.35044 | -1.89487 | 0.332748 | -5.69462 | 1.24E-08 | 1.15E-07 | 0.268897 |
| UNC13A | 1625.271 | -0.63605 | 0.111813 | -5.68848 | 1.28E-08 | 1.19E-07 | 0.643474 |
| STX10 | 533.7479 | -0.61877 | 0.108915 | -5.68123 | 1.34E-08 | 1.24E-07 | 0.651224 |
| CEND1 | 100.1025 | -1.7284 | 0.304383 | -5.67835 | 1.36E-08 | 1.25E-07 | 0.301787 |
| CPT1A | 204.8043 | -1.67219 | 0.294549 | -5.67712 | 1.37E-08 | 1.26E-07 | 0.313777 |
| RNF144B | 112.9489 | -1.43616 | 0.253352 | -5.66864 | 1.44E-08 | 1.32E-07 | 0.369549 |
| ADAMTS10 | 206.6222 | -1.11305 | 0.196367 | -5.6682 | 1.44E-08 | 1.33E-07 | 0.462315 |
| TPR | 3479.573 | -0.6415 | 0.113209 | -5.66655 | 1.46E-08 | 1.34E-07 | 0.641044 |
| P2RX5 | 113.3112 | -1.42766 | 0.251966 | -5.66607 | 1.46E-08 | 1.34E-07 | 0.371734 |
| COL20A1 | 34.98082 | -2.98602 | 0.528139 | -5.65386 | 1.57E-08 | 1.44E-07 | 0.126217 |
| SH3D21 | 348.8018 | -0.88467 | 0.156672 | -5.64666 | 1.64E-08 | 1.50E-07 | 0.54161 |
| FLAD1 | 1457.726 | -0.42052 | 0.074477 | -5.64637 | 1.64E-08 | 1.50E-07 | 0.747153 |
| PKD1 | 3159.915 | -1.12869 | 0.199932 | -5.64537 | 1.65E-08 | 1.51E-07 | 0.457331 |
| FGFR4 | 1504.177 | -0.91583 | 0.162374 | -5.64021 | 1.70E-08 | 1.55E-07 | 0.530041 |
| LTB4R | 541.2985 | -1.46792 | 0.26039 | -5.63739 | 1.73E-08 | 1.57E-07 | 0.361503 |
| ARHGAP22 | 203.3284 | -1.01144 | 0.179463 | -5.63592 | 1.74E-08 | 1.58E-07 | 0.496051 |
| MYBPC2 | 140.0833 | -1.18595 | 0.210506 | -5.63383 | 1.76E-08 | 1.60E-07 | 0.439534 |
| JARID2 | 5457.944 | -0.55077 | 0.097786 | -5.63245 | 1.78E-08 | 1.61E-07 | 0.682654 |
| TM6SF1 | 25.04264 | -2.84175 | 0.504696 | -5.63062 | 1.80E-08 | 1.63E-07 | 0.139491 |
| FAM110A | 158.3898 | -0.86794 | 0.154158 | -5.63021 | 1.80E-08 | 1.63E-07 | 0.547928 |
| MATN4 | 23.0744 | -2.50021 | 0.444183 | -5.62877 | 1.81E-08 | 1.64E-07 | 0.176752 |
| ZRSR2 | 436.0086 | -0.6139 | 0.109098 | -5.62707 | 1.83E-08 | 1.66E-07 | 0.653428 |
| GSN | 783.8706 | -0.62034 | 0.110508 | -5.61355 | 1.98E-08 | 1.79E-07 | 0.650516 |
| DOC2A | 178.5666 | -1.62543 | 0.289599 | -5.61268 | 1.99E-08 | 1.79E-07 | 0.324114 |
| SUMF2 | 2290.782 | -0.64383 | 0.114827 | -5.60691 | 2.06E-08 | 1.85E-07 | 0.640013 |
| OR52A1 | 153.3392 | -1.12044 | 0.200056 | -5.60065 | 2.14E-08 | 1.92E-07 | 0.459952 |
| NETO1 | 152.8511 | -1.26717 | 0.226463 | -5.59548 | 2.20E-08 | 1.97E-07 | 0.415474 |
| CCDC69 | 317.5435 | -1.43485 | 0.256611 | -5.59153 | 2.25E-08 | 2.01E-07 | 0.369885 |
| TMEM145 | 80.98787 | -2.25205 | 0.402884 | -5.58984 | 2.27E-08 | 2.03E-07 | 0.209925 |
| HTR3A | 165.0421 | -1.22321 | 0.219001 | -5.58542 | 2.33E-08 | 2.08E-07 | 0.428328 |
| TGFB1 | 206.3055 | -0.84576 | 0.151497 | -5.58268 | 2.37E-08 | 2.10E-07 | 0.556417 |
| CHRNA7 | 236.1948 | -0.95499 | 0.171221 | -5.57756 | 2.44E-08 | 2.16E-07 | 0.515844 |
| FBXL6 | 614.7967 | -0.96914 | 0.17384 | -5.57486 | 2.48E-08 | 2.20E-07 | 0.510812 |
| ACY1 | 612.9065 | -0.61734 | 0.110786 | -5.57236 | 2.51E-08 | 2.22E-07 | 0.651872 |

| | | | | | | | |
|---|---|---|---|---|---|---|---|
| AMH | 494.8855 | -1.69852 | 0.304829 | -5.57203 | 2.52E-08 | 2.23E-07 | 0.308103 |
| CCDC169 | 317.4067 | -1.06821 | 0.192003 | -5.56352 | 2.64E-08 | 2.34E-07 | 0.476909 |
| SREBF1 | 1936.297 | -0.88331 | 0.158801 | -5.56237 | 2.66E-08 | 2.35E-07 | 0.542123 |
| TEAD4 | 820.8494 | -1.07927 | 0.194248 | -5.55613 | 2.76E-08 | 2.43E-07 | 0.47327 |
| ENTPD2 | 59.92435 | -1.98171 | 0.356873 | -5.55299 | 2.81E-08 | 2.47E-07 | 0.253189 |
| MEGF8 | 3231.41 | -0.50228 | 0.09049 | -5.55071 | 2.85E-08 | 2.50E-07 | 0.705989 |
| WDR74 | 1343.093 | -0.58981 | 0.106275 | -5.54984 | 2.86E-08 | 2.51E-07 | 0.66443 |
| IL20RB | 104.2189 | -2.2214 | 0.400344 | -5.54873 | 2.88E-08 | 2.53E-07 | 0.214433 |
| MYO15B | 340.7382 | -1.35435 | 0.244081 | -5.54876 | 2.88E-08 | 2.53E-07 | 0.391112 |
| TXNRD2 | 455.575 | -0.651 | 0.117369 | -5.54662 | 2.91E-08 | 2.56E-07 | 0.636839 |
| GFPT2 | 2138.81 | -0.48972 | 0.088325 | -5.54455 | 2.95E-08 | 2.59E-07 | 0.712161 |
| ADAMTSL2 | 101.6236 | -1.02391 | 0.184843 | -5.53937 | 3.04E-08 | 2.66E-07 | 0.49178 |
| DCUN1D5 | 1335.597 | -0.48081 | 0.086835 | -5.53705 | 3.08E-08 | 2.69E-07 | 0.716577 |
| CLASRP | 1043.833 | -1.0337 | 0.186693 | -5.53691 | 3.08E-08 | 2.70E-07 | 0.488456 |
| N4BP3 | 1201.475 | -0.8264 | 0.149642 | -5.52252 | 3.34E-08 | 2.92E-07 | 0.563935 |
| APBB3 | 317.0799 | -1.0693 | 0.193639 | -5.52215 | 3.35E-08 | 2.92E-07 | 0.47655 |
| GPRIN2 | 269.6384 | -0.89091 | 0.161337 | -5.52205 | 3.35E-08 | 2.92E-07 | 0.539274 |
| CHST9 | 156.4187 | -0.97561 | 0.176711 | -5.5209 | 3.37E-08 | 2.94E-07 | 0.508526 |
| ATAD3C | 130.8403 | -1.96496 | 0.355975 | -5.51995 | 3.39E-08 | 2.95E-07 | 0.256146 |
| MYLIP | 394.7518 | -0.6956 | 0.126015 | -5.52001 | 3.39E-08 | 2.95E-07 | 0.617452 |
| SPP1 | 540.4043 | -0.69425 | 0.125774 | -5.5198 | 3.39E-08 | 2.95E-07 | 0.618031 |
| SLC2A8 | 499.568 | -0.75293 | 0.136445 | -5.5182 | 3.42E-08 | 2.98E-07 | 0.593397 |
| SLC16A2 | 2912.33 | -0.44565 | 0.080783 | -5.51661 | 3.46E-08 | 3.00E-07 | 0.734254 |
| POR | 2497.52 | -0.79613 | 0.144431 | -5.51215 | 3.54E-08 | 3.08E-07 | 0.575894 |
| COL25A1 | 122.0988 | -1.64834 | 0.299139 | -5.5103 | 3.58E-08 | 3.10E-07 | 0.319006 |
| CLCNKB | 76.96944 | -1.43787 | 0.26096 | -5.50991 | 3.59E-08 | 3.11E-07 | 0.369112 |
| ZDHHC24 | 471.2016 | -0.93315 | 0.16942 | -5.50793 | 3.63E-08 | 3.14E-07 | 0.523712 |
| NPIPB3 | 1483.13 | -1.24204 | 0.225668 | -5.50384 | 3.72E-08 | 3.21E-07 | 0.422774 |
| HS3ST5 | 126.1241 | -1.24329 | 0.225928 | -5.50301 | 3.73E-08 | 3.22E-07 | 0.422409 |
| GJC3 | 31.5191 | -2.24305 | 0.407663 | -5.50222 | 3.75E-08 | 3.24E-07 | 0.211239 |
| DENND1C | 231.2502 | -1.00283 | 0.182346 | -5.49961 | 3.81E-08 | 3.28E-07 | 0.499019 |
| MYO5B | 536.3183 | -0.66604 | 0.12112 | -5.49902 | 3.82E-08 | 3.29E-07 | 0.630233 |
| ASGR1 | 123.647 | -1.12336 | 0.204305 | -5.49844 | 3.83E-08 | 3.30E-07 | 0.459023 |
| MEGF11 | 118.3176 | -1.49295 | 0.271983 | -5.48914 | 4.04E-08 | 3.47E-07 | 0.355285 |
| LAT | 87.79027 | -1.83686 | 0.334651 | -5.48887 | 4.05E-08 | 3.48E-07 | 0.279931 |
| MPG | 531.5874 | -0.52597 | 0.09584 | -5.48801 | 4.06E-08 | 3.49E-07 | 0.694493 |
| DPEP1 | 92.53645 | -1.60896 | 0.293324 | -5.48526 | 4.13E-08 | 3.54E-07 | 0.327835 |
| ANKRD24 | 262.729 | -1.00649 | 0.183631 | -5.48107 | 4.23E-08 | 3.62E-07 | 0.497754 |
| SLC1A1 | 251.5038 | -1.05813 | 0.193201 | -5.47683 | 4.33E-08 | 3.70E-07 | 0.480255 |
| BTN2A1 | 408.7725 | -0.60243 | 0.110012 | -5.47609 | 4.35E-08 | 3.71E-07 | 0.658642 |
| L1TD1 | 7515.833 | -0.79519 | 0.145252 | -5.47453 | 4.39E-08 | 3.74E-07 | 0.576268 |
| MEDAG | 74.99693 | -1.25396 | 0.22918 | -5.47149 | 4.46E-08 | 3.80E-07 | 0.419297 |
| SLC35A2 | 560.6983 | -0.77914 | 0.142403 | -5.47135 | 4.47E-08 | 3.80E-07 | 0.582715 |
| SOCS2 | 923.1632 | -1.20283 | 0.219941 | -5.46887 | 4.53E-08 | 3.85E-07 | 0.434422 |

| | | | | | | | |
|---|---|---|---|---|---|---|---|
| STK11IP | 548.4236 | -1.00498 | 0.183762 | -5.4689 | 4.53E-08 | 3.85E-07 | 0.498278 |
| HERC2 | 2020.647 | -0.76371 | 0.139752 | -5.46478 | 4.63E-08 | 3.93E-07 | 0.588979 |
| NAGS | 83.69269 | -1.30923 | 0.239574 | -5.46481 | 4.63E-08 | 3.93E-07 | 0.403537 |
| PLA2G6 | 410.9942 | -0.9459 | 0.173178 | -5.46201 | 4.71E-08 | 3.99E-07 | 0.519106 |
| GRN | 4101.109 | -0.7846 | 0.143665 | -5.46133 | 4.73E-08 | 4.00E-07 | 0.580511 |
| NDUFS7 | 1146.974 | -0.57423 | 0.105189 | -5.45905 | 4.79E-08 | 4.05E-07 | 0.671645 |
| CHPT1 | 351.8777 | -0.62831 | 0.115098 | -5.45888 | 4.79E-08 | 4.05E-07 | 0.646936 |
| ANKRD18C | 176.8269 | -0.91065 | 0.167259 | -5.44456 | 5.19E-08 | 4.38E-07 | 0.531944 |
| ATP12A | 44.36791 | -1.71503 | 0.314995 | -5.44463 | 5.19E-08 | 4.38E-07 | 0.304597 |
| BGN | 143.4855 | -1.06717 | 0.196038 | -5.44366 | 5.22E-08 | 4.39E-07 | 0.477256 |
| TMEM59 | 1771.466 | -0.41931 | 0.077084 | -5.43964 | 5.34E-08 | 4.49E-07 | 0.747782 |
| PLPP6 | 431.2802 | -0.55978 | 0.10299 | -5.43529 | 5.47E-08 | 4.59E-07 | 0.678404 |
| UVSSA | 1455.172 | -1.07597 | 0.197967 | -5.43507 | 5.48E-08 | 4.59E-07 | 0.474354 |
| USH1C | 38.45553 | -2.41717 | 0.444867 | -5.43348 | 5.53E-08 | 4.63E-07 | 0.187222 |
| BAIAP2L2 | 114.9883 | -1.37725 | 0.253728 | -5.42806 | 5.70E-08 | 4.76E-07 | 0.384951 |
| MYOZ3 | 369.7701 | -0.88883 | 0.163756 | -5.42775 | 5.71E-08 | 4.77E-07 | 0.540054 |
| VIPR1 | 172.9004 | -1.26756 | 0.233557 | -5.42719 | 5.72E-08 | 4.78E-07 | 0.415362 |
| AC008397 | 41.16333 | -2.53139 | 0.466482 | -5.42655 | 5.75E-08 | 4.80E-07 | 0.172973 |
| PRELID3A | 146.6395 | -1.00352 | 0.184988 | -5.42476 | 5.80E-08 | 4.84E-07 | 0.498783 |
| NPFFR2 | 81.28433 | -1.47921 | 0.272773 | -5.42287 | 5.86E-08 | 4.89E-07 | 0.358685 |
| CPNE7 | 503.9576 | -1.20261 | 0.221957 | -5.41819 | 6.02E-08 | 5.01E-07 | 0.43449 |
| PODXL | 41396.47 | -0.46598 | 0.086055 | -5.41494 | 6.13E-08 | 5.09E-07 | 0.723977 |
| GAA | 2712.728 | -0.93154 | 0.172035 | -5.41483 | 6.13E-08 | 5.09E-07 | 0.524299 |
| P2RX1 | 22.28554 | -2.67254 | 0.49365 | -5.41385 | 6.17E-08 | 5.12E-07 | 0.15685 |
| NFKBIE | 358.4941 | -0.7668 | 0.14169 | -5.41183 | 6.24E-08 | 5.17E-07 | 0.587718 |
| TSTA3 | 970.3548 | -0.54839 | 0.101353 | -5.41067 | 6.28E-08 | 5.20E-07 | 0.683784 |
| CYP3A5 | 155.5626 | -0.87744 | 0.162281 | -5.40691 | 6.41E-08 | 5.30E-07 | 0.544333 |
| GPR50 | 151.729 | -0.91418 | 0.169146 | -5.4047 | 6.49E-08 | 5.37E-07 | 0.530645 |
| SLC22A17 | 1249.02 | -0.83383 | 0.154416 | -5.3999 | 6.67E-08 | 5.51E-07 | 0.561038 |
| JUN | 416.4747 | -1.01546 | 0.188161 | -5.39676 | 6.79E-08 | 5.59E-07 | 0.494671 |
| FAM157A | 890.6517 | -1.07668 | 0.199529 | -5.3961 | 6.81E-08 | 5.61E-07 | 0.47412 |
| BSPRY | 198.0291 | -0.82336 | 0.152709 | -5.39173 | 6.98E-08 | 5.73E-07 | 0.565123 |
| DEPP1 | 74.81031 | -1.4541 | 0.269751 | -5.39051 | 7.03E-08 | 5.76E-07 | 0.364983 |
| TSPO | 229.1727 | -0.73351 | 0.136084 | -5.39015 | 7.04E-08 | 5.77E-07 | 0.601439 |
| PPP1R16B | 266.4492 | -0.939 | 0.174429 | -5.38329 | 7.31E-08 | 5.98E-07 | 0.521593 |
| TMEM141 | 730.2919 | -0.62983 | 0.117041 | -5.38126 | 7.40E-08 | 6.05E-07 | 0.646253 |
| FAM20C | 549.2043 | -0.58178 | 0.108135 | -5.38014 | 7.44E-08 | 6.08E-07 | 0.668138 |
| LY75 | 48.98584 | -2.3909 | 0.445205 | -5.37034 | 7.86E-08 | 6.40E-07 | 0.190663 |
| HSPB1 | 1304.577 | -0.53076 | 0.09909 | -5.35635 | 8.49E-08 | 6.90E-07 | 0.692189 |
| C9orf135 | 107.0543 | -1.1775 | 0.21984 | -5.35617 | 8.50E-08 | 6.90E-07 | 0.442117 |
| HDAC10 | 396.5128 | -1.4506 | 0.270872 | -5.3553 | 8.54E-08 | 6.93E-07 | 0.365869 |
| AGPAT3 | 370.3542 | -0.99813 | 0.186406 | -5.35462 | 8.57E-08 | 6.95E-07 | 0.500648 |
| STX4 | 419.8238 | -0.57159 | 0.10678 | -5.35297 | 8.65E-08 | 7.01E-07 | 0.672875 |
| DPP7 | 578.8587 | -0.92147 | 0.172159 | -5.35242 | 8.68E-08 | 7.03E-07 | 0.527971 |

| | | | | | | | |
|---|---|---|---|---|---|---|---|
| F2RL1 | 1118.285 | -0.45427 | 0.084885 | -5.35167 | 8.71E-08 | 7.06E-07 | 0.729877 |
| MT-ND3 | 12419.48 | -1.04979 | 0.196303 | -5.34783 | 8.90E-08 | 7.20E-07 | 0.483037 |
| ACAA1 | 408.5616 | -0.6515 | 0.121856 | -5.34646 | 8.97E-08 | 7.25E-07 | 0.636618 |
| TLE6 | 34.0333 | -2.24638 | 0.420365 | -5.34388 | 9.10E-08 | 7.35E-07 | 0.210752 |
| WASH5P | 1559.786 | -0.57706 | 0.108062 | -5.34012 | 9.29E-08 | 7.49E-07 | 0.670327 |
| TNFAIP2 | 535.7148 | -1.20134 | 0.225075 | -5.33752 | 9.42E-08 | 7.59E-07 | 0.434871 |
| CAPRIN2 | 1385.265 | -0.89203 | 0.16719 | -5.33542 | 9.53E-08 | 7.67E-07 | 0.538855 |
| SPTAN1 | 8569.634 | -1.59928 | 0.299752 | -5.33536 | 9.54E-08 | 7.67E-07 | 0.330041 |
| ANTXR1 | 1158.443 | -0.56957 | 0.106803 | -5.3329 | 9.67E-08 | 7.76E-07 | 0.673816 |
| LOXL3 | 511.5714 | -0.75591 | 0.141783 | -5.33148 | 9.74E-08 | 7.81E-07 | 0.592172 |
| RPS19BP1 | 969.3722 | -0.46675 | 0.0877 | -5.32214 | 1.03E-07 | 8.21E-07 | 0.723591 |
| AC242376 | 124.7799 | -1.39139 | 0.261682 | -5.31709 | 1.05E-07 | 8.40E-07 | 0.381198 |
| MUC16 | 18.8303 | -3.34666 | 0.629609 | -5.31545 | 1.06E-07 | 8.47E-07 | 0.0983 |
| AC114267 | 73.61584 | -1.28397 | 0.241588 | -5.3147 | 1.07E-07 | 8.50E-07 | 0.410664 |
| LTBP3 | 306.6432 | -1.22001 | 0.229622 | -5.31314 | 1.08E-07 | 8.57E-07 | 0.429279 |
| LIME1 | 180.5489 | -1.40435 | 0.264414 | -5.31116 | 1.09E-07 | 8.64E-07 | 0.377789 |
| TELO2 | 1780.688 | -0.88106 | 0.165934 | -5.30971 | 1.10E-07 | 8.71E-07 | 0.542969 |
| PACSIN1 | 333.4509 | -0.67873 | 0.127935 | -5.30524 | 1.13E-07 | 8.91E-07 | 0.624717 |
| MUC1 | 330.2358 | -1.59778 | 0.301261 | -5.30363 | 1.14E-07 | 8.98E-07 | 0.330386 |
| UBR5 | 5358.352 | -0.63932 | 0.120565 | -5.30266 | 1.14E-07 | 9.02E-07 | 0.642017 |
| AFAP1L1 | 106.501 | -1.4311 | 0.26991 | -5.30216 | 1.14E-07 | 9.04E-07 | 0.370847 |
| TMEM212 | 151.4762 | -0.84163 | 0.158813 | -5.29948 | 1.16E-07 | 9.16E-07 | 0.558014 |
| SSBP4 | 626.6962 | -0.60138 | 0.113507 | -5.29821 | 1.17E-07 | 9.21E-07 | 0.659122 |
| RAB34 | 2118.967 | -0.46558 | 0.087933 | -5.29464 | 1.19E-07 | 9.38E-07 | 0.724182 |
| SLC12A9 | 982.5184 | -0.78157 | 0.147661 | -5.293 | 1.20E-07 | 9.45E-07 | 0.581733 |
| ACSL1 | 66.88592 | -1.62007 | 0.306109 | -5.29247 | 1.21E-07 | 9.47E-07 | 0.325319 |
| RHOT2 | 3357.934 | -0.94781 | 0.179154 | -5.29046 | 1.22E-07 | 9.57E-07 | 0.518419 |
| ZNF75D | 373.4196 | -0.80299 | 0.151802 | -5.28972 | 1.23E-07 | 9.60E-07 | 0.573159 |
| MBOAT1 | 125.6622 | -1.14984 | 0.217445 | -5.28798 | 1.24E-07 | 9.69E-07 | 0.450674 |
| AFAP1L2 | 179.6876 | -1.03855 | 0.196408 | -5.28768 | 1.24E-07 | 9.70E-07 | 0.486818 |
| PLPPR2 | 231.9934 | -0.92187 | 0.174351 | -5.28744 | 1.24E-07 | 9.71E-07 | 0.527824 |
| AC243919 | 678.743 | -4.09745 | 0.775127 | -5.28617 | 1.25E-07 | 9.77E-07 | 0.058418 |
| CLDN1 | 219.565 | -0.88134 | 0.166788 | -5.28417 | 1.26E-07 | 9.88E-07 | 0.542864 |
| CHGB | 138.248 | -1.03962 | 0.197003 | -5.27717 | 1.31E-07 | 1.02E-06 | 0.486456 |
| SORBS3 | 1146.824 | -0.83938 | 0.159209 | -5.27222 | 1.35E-07 | 1.05E-06 | 0.558883 |
| DBNDD1 | 1122.167 | -0.4467 | 0.084737 | -5.27167 | 1.35E-07 | 1.05E-06 | 0.733717 |
| HIF3A | 986.7533 | -0.70177 | 0.133212 | -5.26808 | 1.38E-07 | 1.07E-06 | 0.614816 |
| ZNF787 | 1135.892 | -0.59305 | 0.112636 | -5.26521 | 1.40E-07 | 1.08E-06 | 0.66294 |
| HERC1 | 1706.187 | -0.70804 | 0.134568 | -5.26155 | 1.43E-07 | 1.11E-06 | 0.612153 |
| LAMB3 | 63.88533 | -1.67284 | 0.318365 | -5.25446 | 1.48E-07 | 1.15E-06 | 0.313636 |
| USHBP1 | 161.5896 | -1.06228 | 0.202166 | -5.2545 | 1.48E-07 | 1.15E-06 | 0.478874 |
| FAM207A | 586.5499 | -0.6047 | 0.115085 | -5.25438 | 1.49E-07 | 1.15E-06 | 0.657608 |
| NFASC | 390.3757 | -0.72381 | 0.137798 | -5.25267 | 1.50E-07 | 1.16E-06 | 0.605497 |
| HLA-V | 20.75746 | -2.79814 | 0.532732 | -5.25243 | 1.50E-07 | 1.16E-06 | 0.143773 |

| | | | | | | | |
|---|---|---|---|---|---|---|---|
| LZTS1 | 363.473 | -0.74777 | 0.142523 | -5.2467 | 1.55E-07 | 1.19E-06 | 0.595522 |
| ADAMTSL3 | 436.4618 | -0.64195 | 0.122409 | -5.24436 | 1.57E-07 | 1.20E-06 | 0.640844 |
| TCF7L2 | 1600.243 | -0.51591 | 0.098412 | -5.24234 | 1.59E-07 | 1.22E-06 | 0.699352 |
| MOB3C | 142.6334 | -1.03257 | 0.197121 | -5.23823 | 1.62E-07 | 1.24E-06 | 0.488839 |
| GDPD2 | 867.686 | -1.00551 | 0.191976 | -5.23769 | 1.63E-07 | 1.24E-06 | 0.498094 |
| RDH11 | 2762.52 | -0.52813 | 0.100863 | -5.23615 | 1.64E-07 | 1.25E-06 | 0.693452 |
| SHB | 299.994 | -0.75012 | 0.143279 | -5.2354 | 1.65E-07 | 1.26E-06 | 0.594554 |
| PLAAT5 | 283.096 | -1.21283 | 0.232142 | -5.22451 | 1.75E-07 | 1.33E-06 | 0.431422 |
| SLC16A10 | 227.3984 | -1.35966 | 0.260415 | -5.22113 | 1.78E-07 | 1.35E-06 | 0.389674 |
| CLDN10 | 434.0785 | -0.84065 | 0.161022 | -5.22073 | 1.78E-07 | 1.35E-06 | 0.558391 |
| RCN3 | 136.2398 | -1.66676 | 0.319261 | -5.22067 | 1.78E-07 | 1.35E-06 | 0.314961 |
| LEMD2 | 1893.399 | -0.6118 | 0.117193 | -5.22047 | 1.78E-07 | 1.36E-06 | 0.65438 |
| CSMD2 | 688.2454 | -0.64918 | 0.124388 | -5.219 | 1.80E-07 | 1.37E-06 | 0.637643 |
| DNM1 | 1337.308 | -0.95098 | 0.182269 | -5.21746 | 1.81E-07 | 1.38E-06 | 0.51728 |
| PROCA1 | 378.3988 | -0.8203 | 0.157267 | -5.21594 | 1.83E-07 | 1.38E-06 | 0.566325 |
| BX255925. | 579.6091 | -1.03073 | 0.197738 | -5.21261 | 1.86E-07 | 1.41E-06 | 0.489463 |
| ELF3 | 176.14 | -1.06524 | 0.204522 | -5.20846 | 1.90E-07 | 1.44E-06 | 0.477892 |
| RHBDF2 | 592.9668 | -0.92534 | 0.177685 | -5.20774 | 1.91E-07 | 1.44E-06 | 0.526557 |
| PNMA3 | 280.6629 | -0.80955 | 0.155495 | -5.20628 | 1.93E-07 | 1.45E-06 | 0.570559 |
| CHRNB1 | 510.2994 | -0.56956 | 0.109423 | -5.2051 | 1.94E-07 | 1.46E-06 | 0.673825 |
| UNC93B1 | 426.5681 | -0.84893 | 0.163189 | -5.20213 | 1.97E-07 | 1.48E-06 | 0.555195 |
| ASIC3 | 88.35369 | -2.46292 | 0.473517 | -5.20134 | 1.98E-07 | 1.49E-06 | 0.181379 |
| ARHGAP9 | 114.5845 | -1.467 | 0.282094 | -5.2004 | 1.99E-07 | 1.49E-06 | 0.361734 |
| MX2 | 137.1845 | -1.00986 | 0.194234 | -5.19916 | 2.00E-07 | 1.50E-06 | 0.496596 |
| INTS11 | 2791.344 | -0.65938 | 0.12695 | -5.19401 | 2.06E-07 | 1.54E-06 | 0.63315 |
| P4HB | 10133.19 | -0.59967 | 0.115497 | -5.19213 | 2.08E-07 | 1.56E-06 | 0.659903 |
| AC139768 | 222.5474 | -1.01499 | 0.195532 | -5.19089 | 2.09E-07 | 1.57E-06 | 0.494833 |
| ESYT3 | 148.9638 | -1.10637 | 0.213176 | -5.18996 | 2.10E-07 | 1.57E-06 | 0.46446 |
| CCDC24 | 222.8572 | -0.98783 | 0.190407 | -5.18798 | 2.13E-07 | 1.59E-06 | 0.504236 |
| MCOLN2 | 70.15782 | -1.41924 | 0.273669 | -5.18599 | 2.15E-07 | 1.60E-06 | 0.373909 |
| SPIN4 | 1029.701 | -0.48437 | 0.093494 | -5.18074 | 2.21E-07 | 1.65E-06 | 0.71481 |
| WIPF3 | 509.2564 | -0.61801 | 0.119289 | -5.1808 | 2.21E-07 | 1.65E-06 | 0.651569 |
| ALDH1L1 | 43.96707 | -2.10172 | 0.406244 | -5.17353 | 2.30E-07 | 1.71E-06 | 0.232981 |
| B4GALT4 | 1144.213 | -0.49221 | 0.095146 | -5.17323 | 2.30E-07 | 1.71E-06 | 0.710935 |
| PLPPR5 | 117.5938 | -1.18425 | 0.228946 | -5.17261 | 2.31E-07 | 1.72E-06 | 0.440053 |
| PORCN | 319.0846 | -0.75011 | 0.145152 | -5.16778 | 2.37E-07 | 1.76E-06 | 0.594558 |
| COL1A2 | 4528.117 | -0.68024 | 0.131664 | -5.16648 | 2.39E-07 | 1.77E-06 | 0.624062 |
| EMC3 | 558.2458 | -0.74496 | 0.14424 | -5.16468 | 2.41E-07 | 1.78E-06 | 0.596686 |
| CDK11A | 713.3758 | -1.07463 | 0.208142 | -5.16297 | 2.43E-07 | 1.80E-06 | 0.474792 |
| AIG1 | 568.5279 | -0.5242 | 0.101581 | -5.16038 | 2.46E-07 | 1.82E-06 | 0.695345 |
| NMNAT1P3 | 13.6621 | -3.87452 | 0.750945 | -5.15952 | 2.48E-07 | 1.83E-06 | 0.06818 |
| WDR90 | 1374.64 | -1.18692 | 0.230127 | -5.15767 | 2.50E-07 | 1.85E-06 | 0.43924 |
| GAS6 | 1604.497 | -0.95528 | 0.185238 | -5.15703 | 2.51E-07 | 1.85E-06 | 0.515743 |
| SQSTM1 | 3109.2 | -0.73029 | 0.141642 | -5.1559 | 2.52E-07 | 1.86E-06 | 0.602782 |

| | | | | | | | |
|---|---|---|---|---|---|---|---|
| LRCH4 | 791.3191 | -0.55191 | 0.107064 | -5.15495 | 2.54E-07 | 1.87E-06 | 0.682116 |
| TNS2 | 182.5325 | -1.49299 | 0.289676 | -5.154 | 2.55E-07 | 1.88E-06 | 0.355276 |
| CACNA2D1 | 2164.229 | -0.57026 | 0.110797 | -5.14693 | 2.65E-07 | 1.95E-06 | 0.673493 |
| TOR3A | 939.8523 | -0.54028 | 0.105109 | -5.1402 | 2.74E-07 | 2.02E-06 | 0.687636 |
| AC005726 | 358.6195 | -0.99789 | 0.194255 | -5.13701 | 2.79E-07 | 2.05E-06 | 0.500731 |
| TMEM64 | 2941.781 | -0.43478 | 0.08469 | -5.13383 | 2.84E-07 | 2.08E-06 | 0.739804 |
| UCMA | 31.06703 | -1.96446 | 0.382894 | -5.13057 | 2.89E-07 | 2.11E-06 | 0.256235 |
| CFAP54 | 136.7436 | -0.88573 | 0.17266 | -5.12989 | 2.90E-07 | 2.12E-06 | 0.541215 |
| MED14 | 3360.376 | -0.44355 | 0.086474 | -5.12923 | 2.91E-07 | 2.13E-06 | 0.735324 |
| TNFRSF25 | 321.5665 | -1.48022 | 0.288606 | -5.12888 | 2.91E-07 | 2.13E-06 | 0.358433 |
| TRMT2A | 727.4307 | -0.69119 | 0.134784 | -5.12809 | 2.93E-07 | 2.14E-06 | 0.619345 |
| MTRNR2L1 | 51.08629 | -1.79469 | 0.350107 | -5.12612 | 2.96E-07 | 2.15E-06 | 0.288234 |
| SAMD12 | 671.9322 | -0.57771 | 0.1127 | -5.12612 | 2.96E-07 | 2.15E-06 | 0.670025 |
| GLS2 | 340.4132 | -0.85881 | 0.167613 | -5.12377 | 2.99E-07 | 2.18E-06 | 0.551408 |
| GLT8D2 | 90.16134 | -1.04447 | 0.20398 | -5.12046 | 3.05E-07 | 2.21E-06 | 0.484823 |
| CACNB1 | 318.9854 | -1.09669 | 0.214479 | -5.11325 | 3.17E-07 | 2.29E-06 | 0.467589 |
| AL732372. | 351.3578 | -1.03947 | 0.203395 | -5.11059 | 3.21E-07 | 2.32E-06 | 0.486507 |
| VSIG10 | 1231.661 | -0.82952 | 0.162318 | -5.1105 | 3.21E-07 | 2.32E-06 | 0.562715 |
| LPL | 255.4209 | -1.15998 | 0.227 | -5.11006 | 3.22E-07 | 2.33E-06 | 0.447518 |
| ELF4 | 73.87646 | -1.39588 | 0.2732 | -5.10936 | 3.23E-07 | 2.33E-06 | 0.380013 |
| PPP1R1B | 220.0129 | -0.9439 | 0.184791 | -5.10796 | 3.26E-07 | 2.35E-06 | 0.519825 |
| ARRB1 | 1058.876 | -0.75279 | 0.147449 | -5.10542 | 3.30E-07 | 2.38E-06 | 0.593455 |
| GRIN1 | 58.53212 | -2.61287 | 0.511949 | -5.10376 | 3.33E-07 | 2.40E-06 | 0.163474 |
| CHST15 | 274.0662 | -1.02492 | 0.200834 | -5.1033 | 3.34E-07 | 2.40E-06 | 0.491438 |
| ZDHHC23 | 333.0786 | -0.83179 | 0.163034 | -5.10193 | 3.36E-07 | 2.42E-06 | 0.561833 |
| SRRM2 | 19729.18 | -0.90054 | 0.176517 | -5.10173 | 3.37E-07 | 2.42E-06 | 0.535686 |
| BEST1 | 176.2752 | -1.41717 | 0.277791 | -5.10159 | 3.37E-07 | 2.42E-06 | 0.374445 |
| PKD1P6 | 372.0093 | -0.9501 | 0.186245 | -5.10133 | 3.37E-07 | 2.42E-06 | 0.517597 |
| ALOX12P2 | 185.2554 | -0.89576 | 0.175659 | -5.09944 | 3.41E-07 | 2.44E-06 | 0.537464 |
| NOXO1 | 11.75441 | -6.06463 | 1.189841 | -5.09701 | 3.45E-07 | 2.47E-06 | 0.014941 |
| ALKAL2 | 42.8463 | -2.0877 | 0.409613 | -5.09678 | 3.45E-07 | 2.47E-06 | 0.235255 |
| TMEM184A | 63.0002 | -3.01951 | 0.592651 | -5.09493 | 3.49E-07 | 2.49E-06 | 0.123321 |
| DDX24 | 535.7837 | -1.08578 | 0.213256 | -5.09144 | 3.55E-07 | 2.54E-06 | 0.471138 |
| YBX2 | 717.1477 | -0.82242 | 0.161604 | -5.08913 | 3.60E-07 | 2.57E-06 | 0.565491 |
| MIB2 | 1232.34 | -1.02628 | 0.201845 | -5.08448 | 3.69E-07 | 2.63E-06 | 0.490976 |
| PAK6 | 240.3981 | -1.34039 | 0.26362 | -5.08456 | 3.68E-07 | 2.63E-06 | 0.394914 |
| PPP1R12C | 1318.984 | -0.85172 | 0.167517 | -5.0844 | 3.69E-07 | 2.63E-06 | 0.554123 |
| 1-Mar | 830.5188 | -0.56072 | 0.110289 | -5.0841 | 3.69E-07 | 2.63E-06 | 0.677964 |
| KCNQ4 | 133.1505 | -0.98204 | 0.193164 | -5.08397 | 3.70E-07 | 2.63E-06 | 0.506263 |
| SNCB | 101.2786 | -1.19818 | 0.235727 | -5.0829 | 3.72E-07 | 2.64E-06 | 0.435826 |
| ALG1 | 1128.254 | -0.57743 | 0.113701 | -5.07847 | 3.80E-07 | 2.70E-06 | 0.670158 |
| TMEM259 | 4546.789 | -0.86084 | 0.169679 | -5.07337 | 3.91E-07 | 2.77E-06 | 0.55063 |
| SLC13A5 | 68.90415 | -1.52569 | 0.300865 | -5.07101 | 3.96E-07 | 2.80E-06 | 0.347314 |
| PDIA3 | 11726.39 | -0.49248 | 0.097128 | -5.07043 | 3.97E-07 | 2.81E-06 | 0.710801 |

| | | | | | | | |
|---|---|---|---|---|---|---|---|
| RASSF7 | 496.8572 | -0.88498 | 0.174624 | -5.06795 | 4.02E-07 | 2.85E-06 | 0.541493 |
| ADAMTS13 | 401.9021 | -1.4324 | 0.2829 | -5.06327 | 4.12E-07 | 2.91E-06 | 0.370514 |
| MPP1 | 100.7909 | -1.69265 | 0.334379 | -5.06206 | 4.15E-07 | 2.93E-06 | 0.309358 |
| RTKN | 919.7591 | -0.51485 | 0.101774 | -5.05878 | 4.22E-07 | 2.98E-06 | 0.699866 |
| MFSD6 | 122.9748 | -1.1481 | 0.22704 | -5.05679 | 4.26E-07 | 3.00E-06 | 0.45122 |
| DBNDD2 | 560.8091 | -0.68756 | 0.135979 | -5.05636 | 4.27E-07 | 3.01E-06 | 0.620903 |
| S1PR5 | 55.00301 | -1.46778 | 0.290481 | -5.05293 | 4.35E-07 | 3.06E-06 | 0.361539 |
| OGFOD2 | 361.3141 | -0.88056 | 0.174286 | -5.0524 | 4.36E-07 | 3.07E-06 | 0.543156 |
| FAM241B | 240.5455 | -0.72052 | 0.142675 | -5.05006 | 4.42E-07 | 3.11E-06 | 0.606879 |
| KIFC3 | 140.8549 | -1.53541 | 0.304107 | -5.04893 | 4.44E-07 | 3.12E-06 | 0.344981 |
| PLEKHF1 | 126.9485 | -0.93949 | 0.186104 | -5.04819 | 4.46E-07 | 3.13E-06 | 0.521418 |
| AMFR | 1093.113 | -0.43092 | 0.085381 | -5.04697 | 4.49E-07 | 3.15E-06 | 0.74179 |
| VSIG1 | 46.86759 | -1.56988 | 0.31154 | -5.03909 | 4.68E-07 | 3.28E-06 | 0.336837 |
| CAMSAP3 | 983.8666 | -1.041 | 0.206727 | -5.0356 | 4.76E-07 | 3.34E-06 | 0.485992 |
| BSCL2 | 618.6437 | -1.02741 | 0.20408 | -5.03434 | 4.79E-07 | 3.35E-06 | 0.49059 |
| NSUN5P1 | 1710.082 | -1.04687 | 0.207999 | -5.03306 | 4.83E-07 | 3.38E-06 | 0.484017 |
| PVR | 701.3102 | -0.71655 | 0.142415 | -5.03146 | 4.87E-07 | 3.40E-06 | 0.608549 |
| TMEM43 | 1442.154 | -0.46026 | 0.091485 | -5.03099 | 4.88E-07 | 3.40E-06 | 0.726856 |
| P3H1 | 1387.609 | -0.65644 | 0.130522 | -5.02939 | 4.92E-07 | 3.43E-06 | 0.63444 |
| NCCRP1 | 67.96255 | -1.25879 | 0.250361 | -5.02789 | 4.96E-07 | 3.45E-06 | 0.417895 |
| PCNX2 | 1714.832 | -0.65606 | 0.130487 | -5.02776 | 4.96E-07 | 3.45E-06 | 0.63461 |
| ZNRF3 | 684.8626 | -0.5319 | 0.105808 | -5.02702 | 4.98E-07 | 3.46E-06 | 0.691644 |
| LMCD1 | 115.8737 | -1.4702 | 0.292531 | -5.02581 | 5.01E-07 | 3.48E-06 | 0.360932 |
| GALNT6 | 334.8358 | -0.91619 | 0.182376 | -5.02366 | 5.07E-07 | 3.52E-06 | 0.529906 |
| PIGQ | 1304.978 | -0.85475 | 0.170203 | -5.02198 | 5.11E-07 | 3.55E-06 | 0.55296 |
| AL353625. | 56.75602 | -1.64751 | 0.328304 | -5.01825 | 5.21E-07 | 3.61E-06 | 0.31919 |
| WFDC2 | 310.1979 | -0.85435 | 0.170321 | -5.01616 | 5.27E-07 | 3.65E-06 | 0.553113 |
| BCL11B | 50.2594 | -1.55738 | 0.310663 | -5.01308 | 5.36E-07 | 3.71E-06 | 0.339767 |
| TESPA1 | 23.37092 | -2.85633 | 0.569817 | -5.01272 | 5.37E-07 | 3.71E-06 | 0.138089 |
| MST1 | 192.4773 | -1.41882 | 0.283237 | -5.0093 | 5.46E-07 | 3.77E-06 | 0.374018 |
| CCDC151 | 45.68428 | -1.59363 | 0.31837 | -5.00559 | 5.57E-07 | 3.84E-06 | 0.331337 |
| TBC1D8 | 418.4143 | -0.77763 | 0.155427 | -5.00317 | 5.64E-07 | 3.89E-06 | 0.583326 |
| SLC25A1 | 1976.309 | -0.41537 | 0.083115 | -4.9975 | 5.81E-07 | 3.99E-06 | 0.749828 |
| SPTBN5 | 87.84252 | -2.07739 | 0.415709 | -4.99721 | 5.82E-07 | 3.99E-06 | 0.236943 |
| SHISA9 | 724.4357 | -0.7388 | 0.148013 | -4.99143 | 5.99E-07 | 4.11E-06 | 0.599239 |
| MICAL1 | 2689.979 | -0.97668 | 0.195817 | -4.98771 | 6.11E-07 | 4.18E-06 | 0.508148 |
| JMJD1C | 3371.097 | -0.5909 | 0.118563 | -4.98383 | 6.23E-07 | 4.26E-06 | 0.663929 |
| CAMTA2 | 341.0431 | -0.78734 | 0.158012 | -4.98282 | 6.27E-07 | 4.28E-06 | 0.57941 |
| SH3GL3 | 158.236 | -0.83955 | 0.168507 | -4.98229 | 6.28E-07 | 4.29E-06 | 0.558817 |
| MYADM | 1771.403 | -0.61012 | 0.122523 | -4.97962 | 6.37E-07 | 4.35E-06 | 0.655142 |
| BCAR1 | 883.2291 | -0.73624 | 0.147878 | -4.97867 | 6.40E-07 | 4.37E-06 | 0.600304 |
| SNAI3 | 72.41351 | -1.3051 | 0.262202 | -4.97747 | 6.44E-07 | 4.39E-06 | 0.404692 |
| CCDC84 | 358.1979 | -0.69751 | 0.14018 | -4.9758 | 6.50E-07 | 4.43E-06 | 0.616635 |
| KRT18 | 4146.717 | -0.64718 | 0.130072 | -4.97555 | 6.51E-07 | 4.43E-06 | 0.638528 |

| | | | | | | | |
|---|---|---|---|---|---|---|---|
| SLC16A12 | 106.7796 | -1.10533 | 0.222156 | -4.97547 | 6.51E-07 | 4.43E-06 | 0.464796 |
| VASP | 2379.541 | -0.65016 | 0.130722 | -4.97359 | 6.57E-07 | 4.47E-06 | 0.637211 |
| PRDM14 | 919.6411 | -0.4557 | 0.091644 | -4.97246 | 6.61E-07 | 4.50E-06 | 0.729158 |
| R3HDM4 | 1308.456 | -0.47619 | 0.095815 | -4.96991 | 6.70E-07 | 4.55E-06 | 0.718873 |
| SOWAHD | 104.3391 | -1.03395 | 0.208424 | -4.96081 | 7.02E-07 | 4.76E-06 | 0.488371 |
| KCNK12 | 209.1928 | -1.32353 | 0.266837 | -4.96008 | 7.05E-07 | 4.78E-06 | 0.399555 |
| AKIRIN1 | 6003.973 | -0.53019 | 0.107004 | -4.95481 | 7.24E-07 | 4.90E-06 | 0.692465 |
| PHKG2 | 778.3523 | -0.46734 | 0.094371 | -4.95214 | 7.34E-07 | 4.97E-06 | 0.723298 |
| PTPN6 | 420.3565 | -0.90762 | 0.183303 | -4.95147 | 7.37E-07 | 4.98E-06 | 0.533064 |
| RGS5 | 483.3621 | -0.66102 | 0.13352 | -4.95071 | 7.39E-07 | 5.00E-06 | 0.63243 |
| RAD52 | 225.1711 | -0.78405 | 0.158431 | -4.94888 | 7.46E-07 | 5.04E-06 | 0.580733 |
| MPPE1 | 208.8004 | -0.99024 | 0.200122 | -4.94818 | 7.49E-07 | 5.06E-06 | 0.503394 |
| RFT1 | 981.5394 | -0.51664 | 0.104413 | -4.94799 | 7.50E-07 | 5.06E-06 | 0.699 |
| ADORA2A | 124.3477 | -1.21713 | 0.246283 | -4.94201 | 7.73E-07 | 5.21E-06 | 0.430137 |
| MTND1P23 | 68.28186 | -2.91502 | 0.589864 | -4.94184 | 7.74E-07 | 5.22E-06 | 0.132584 |
| GALT | 469.2938 | -0.53981 | 0.10931 | -4.93832 | 7.88E-07 | 5.30E-06 | 0.687862 |
| ACOT1 | 68.60778 | -1.41377 | 0.286361 | -4.93702 | 7.93E-07 | 5.33E-06 | 0.375329 |
| RASEF | 663.0346 | -0.53611 | 0.108594 | -4.9368 | 7.94E-07 | 5.34E-06 | 0.68963 |
| CNN1 | 296.0657 | -0.77515 | 0.157119 | -4.93353 | 8.08E-07 | 5.42E-06 | 0.584327 |
| MMP23B | 212.6512 | -1.25078 | 0.253821 | -4.92779 | 8.32E-07 | 5.57E-06 | 0.420222 |
| CARD10 | 474.8388 | -0.56413 | 0.114493 | -4.9272 | 8.34E-07 | 5.59E-06 | 0.676363 |
| SDCCAG8 | 110.8502 | -1.45021 | 0.294362 | -4.92662 | 8.37E-07 | 5.60E-06 | 0.365968 |
| MX1 | 201.6254 | -1.00982 | 0.205075 | -4.92414 | 8.47E-07 | 5.67E-06 | 0.496609 |
| EML3 | 662.8347 | -1.39102 | 0.282645 | -4.92142 | 8.59E-07 | 5.74E-06 | 0.381296 |
| AGAP7P | 163.4171 | -1.0667 | 0.216775 | -4.92076 | 8.62E-07 | 5.75E-06 | 0.477411 |
| REEP1 | 38.19828 | -1.53514 | 0.312096 | -4.9188 | 8.71E-07 | 5.81E-06 | 0.345046 |
| PLEKHA2 | 66.43818 | -1.47141 | 0.299146 | -4.91869 | 8.71E-07 | 5.81E-06 | 0.360631 |
| IQGAP2 | 1786.883 | -1.08108 | 0.219944 | -4.91525 | 8.87E-07 | 5.91E-06 | 0.472675 |
| OSMR | 108.3473 | -1.79663 | 0.36557 | -4.9146 | 8.90E-07 | 5.92E-06 | 0.287846 |
| RTN4RL2 | 652.7132 | -0.52643 | 0.107189 | -4.91127 | 9.05E-07 | 6.02E-06 | 0.694269 |
| NAP1L3 | 1767.556 | -0.44485 | 0.090599 | -4.91012 | 9.10E-07 | 6.05E-06 | 0.73466 |
| LPCAT4 | 955.1395 | -0.72126 | 0.14696 | -4.90785 | 9.21E-07 | 6.11E-06 | 0.606569 |
| WFIKKN2 | 72.552 | -1.79551 | 0.365844 | -4.90785 | 9.21E-07 | 6.11E-06 | 0.288071 |
| RASIP1 | 171.8537 | -1.3044 | 0.265809 | -4.90729 | 9.23E-07 | 6.12E-06 | 0.404889 |
| PPIF | 1491.396 | -0.43404 | 0.088497 | -4.90462 | 9.36E-07 | 6.20E-06 | 0.740185 |
| TP53I13 | 818.1521 | -0.80726 | 0.164617 | -4.90387 | 9.40E-07 | 6.21E-06 | 0.571467 |
| KCNG1 | 628.0872 | -0.55483 | 0.113157 | -4.90322 | 9.43E-07 | 6.23E-06 | 0.680736 |
| AC116562 | 216.2084 | -1.01556 | 0.207151 | -4.90251 | 9.46E-07 | 6.25E-06 | 0.494637 |
| SLC27A4 | 742.5304 | -0.72028 | 0.147017 | -4.89927 | 9.62E-07 | 6.34E-06 | 0.606981 |
| SEMA3D | 98.94835 | -1.30404 | 0.26643 | -4.89449 | 9.86E-07 | 6.48E-06 | 0.404992 |
| DLG4 | 1356.591 | -0.54472 | 0.111312 | -4.89357 | 9.90E-07 | 6.51E-06 | 0.685527 |
| GPR137 | 531.4397 | -0.97034 | 0.198394 | -4.89098 | 1.00E-06 | 6.59E-06 | 0.510385 |
| ZGLP1 | 99.59753 | -1.41007 | 0.288445 | -4.88851 | 1.02E-06 | 6.67E-06 | 0.376294 |
| MAMDC2 | 301.236 | -0.61947 | 0.126867 | -4.88285 | 1.05E-06 | 6.86E-06 | 0.650908 |

| | | | | | | | |
|---|---|---|---|---|---|---|---|
| CABP1 | 89.55229 | -1.1412 | 0.233804 | -4.88102 | 1.06E-06 | 6.91E-06 | 0.453383 |
| ADCYAP1F | 478.0293 | -0.92379 | 0.189297 | -4.88013 | 1.06E-06 | 6.94E-06 | 0.527122 |
| PLP2 | 106.1692 | -0.9236 | 0.189483 | -4.87431 | 1.09E-06 | 7.13E-06 | 0.527192 |
| PRR22 | 50.52401 | -1.92611 | 0.395382 | -4.8715 | 1.11E-06 | 7.23E-06 | 0.263139 |
| SNX22 | 154.8466 | -0.9914 | 0.203666 | -4.86776 | 1.13E-06 | 7.36E-06 | 0.50299 |
| TRIM54 | 90.27475 | -1.8857 | 0.387424 | -4.86727 | 1.13E-06 | 7.38E-06 | 0.270612 |
| PTPRF | 9847.776 | -0.44081 | 0.090636 | -4.86355 | 1.15E-06 | 7.51E-06 | 0.736719 |
| NLRP3P1 | 11.86398 | -4.94976 | 1.018381 | -4.86042 | 1.17E-06 | 7.62E-06 | 0.032357 |
| ETNK2 | 378.8832 | -0.7112 | 0.146347 | -4.85965 | 1.18E-06 | 7.64E-06 | 0.610813 |
| PXN | 2155.033 | -0.50665 | 0.104325 | -4.85643 | 1.20E-06 | 7.76E-06 | 0.703856 |
| BAK1 | 1014.559 | -0.44958 | 0.092616 | -4.85422 | 1.21E-06 | 7.85E-06 | 0.732257 |
| BDP1 | 280.2426 | -0.81669 | 0.16831 | -4.85229 | 1.22E-06 | 7.92E-06 | 0.567743 |
| SORBS1 | 766.219 | -0.74084 | 0.152775 | -4.84921 | 1.24E-06 | 8.03E-06 | 0.598392 |
| AC080038 | 1458.222 | -0.7676 | 0.158323 | -4.84834 | 1.24E-06 | 8.06E-06 | 0.587393 |
| LLGL2 | 1492.34 | -0.7375 | 0.152169 | -4.84659 | 1.26E-06 | 8.12E-06 | 0.599777 |
| PIPOX | 1221.254 | -0.55219 | 0.114046 | -4.84181 | 1.29E-06 | 8.31E-06 | 0.681985 |
| CENPT | 1275.285 | -0.73772 | 0.152437 | -4.83951 | 1.30E-06 | 8.39E-06 | 0.599686 |
| DNASE2 | 1512.519 | -0.51072 | 0.105548 | -4.8388 | 1.31E-06 | 8.42E-06 | 0.70187 |
| CORO1A | 176.2816 | -0.81323 | 0.168072 | -4.83858 | 1.31E-06 | 8.42E-06 | 0.569107 |
| TNK1 | 245.3257 | -0.84821 | 0.175305 | -4.83849 | 1.31E-06 | 8.42E-06 | 0.555473 |
| KCNQ1 | 117.3268 | -1.01511 | 0.209815 | -4.83812 | 1.31E-06 | 8.43E-06 | 0.494791 |
| TCF15 | 81.47633 | -1.11951 | 0.231401 | -4.83797 | 1.31E-06 | 8.44E-06 | 0.46025 |
| DOC2GP | 82.43487 | -1.44627 | 0.298989 | -4.8372 | 1.32E-06 | 8.46E-06 | 0.366968 |
| CYP2R1 | 188.1987 | -0.81154 | 0.167925 | -4.83278 | 1.35E-06 | 8.64E-06 | 0.569772 |
| RASAL1 | 139.0226 | -0.84618 | 0.175123 | -4.8319 | 1.35E-06 | 8.68E-06 | 0.556256 |
| ATP1B3 | 3872.237 | -0.66034 | 0.136676 | -4.83146 | 1.36E-06 | 8.69E-06 | 0.632727 |
| SEMA3E | 127.9664 | -1.3284 | 0.275141 | -4.82809 | 1.38E-06 | 8.83E-06 | 0.398208 |
| SLC39A4 | 19.57434 | -2.65665 | 0.550473 | -4.82612 | 1.39E-06 | 8.92E-06 | 0.158587 |
| IL11 | 51.18857 | -1.67381 | 0.346844 | -4.82583 | 1.39E-06 | 8.93E-06 | 0.313425 |
| MTND2P28 | 808.4076 | -1.0519 | 0.218092 | -4.82318 | 1.41E-06 | 9.04E-06 | 0.482334 |
| TMBIM1 | 930.4078 | -0.4749 | 0.09847 | -4.82283 | 1.42E-06 | 9.06E-06 | 0.719516 |
| CAPG | 716.5695 | -0.64314 | 0.133369 | -4.82228 | 1.42E-06 | 9.08E-06 | 0.640316 |
| PUSL1 | 518.766 | -0.55257 | 0.114616 | -4.82101 | 1.43E-06 | 9.13E-06 | 0.681807 |
| MALL | 333.5601 | -0.793 | 0.164543 | -4.8194 | 1.44E-06 | 9.20E-06 | 0.577144 |
| ITPA | 710.2736 | -0.43882 | 0.091202 | -4.81149 | 1.50E-06 | 9.55E-06 | 0.737738 |
| POMC | 45.55033 | -1.57911 | 0.328265 | -4.81048 | 1.51E-06 | 9.60E-06 | 0.334688 |
| CXCL16 | 73.38962 | -1.38334 | 0.287776 | -4.807 | 1.53E-06 | 9.75E-06 | 0.38333 |
| KIF1C | 4550.001 | -0.86999 | 0.180985 | -4.80697 | 1.53E-06 | 9.75E-06 | 0.54715 |
| GABRA3 | 162.9896 | -0.84824 | 0.176499 | -4.80592 | 1.54E-06 | 9.80E-06 | 0.555462 |
| TP53INP2 | 131.8134 | -1.61569 | 0.336732 | -4.79816 | 1.60E-06 | 1.02E-05 | 0.326308 |
| ADM5 | 102.7272 | -1.43961 | 0.300202 | -4.79547 | 1.62E-06 | 1.03E-05 | 0.368667 |
| OS9 | 2946.404 | -0.61203 | 0.127632 | -4.79524 | 1.62E-06 | 1.03E-05 | 0.654277 |
| SLFN13 | 401.4368 | -0.87296 | 0.182087 | -4.79419 | 1.63E-06 | 1.03E-05 | 0.546025 |
| AVPR1B | 14.38002 | -2.55665 | 0.533348 | -4.79359 | 1.64E-06 | 1.04E-05 | 0.16997 |

| | | | | | | | |
|---|---|---|---|---|---|---|---|
| KCNS1 | 53.32652 | -1.37578 | 0.287332 | -4.78813 | 1.68E-06 | 1.06E-05 | 0.385344 |
| ADAM33 | 267.9807 | -1.37626 | 0.287739 | -4.783 | 1.73E-06 | 1.09E-05 | 0.385217 |
| PIK3CD | 797.2748 | -0.76257 | 0.159437 | -4.78291 | 1.73E-06 | 1.09E-05 | 0.589445 |
| TSC2 | 2611.929 | -0.86887 | 0.181761 | -4.7803 | 1.75E-06 | 1.10E-05 | 0.547574 |
| ATXN7L1 | 506.1767 | -0.73433 | 0.15366 | -4.77894 | 1.76E-06 | 1.11E-05 | 0.601097 |
| HCFC1R1 | 838.0237 | -0.48143 | 0.100777 | -4.77717 | 1.78E-06 | 1.12E-05 | 0.716267 |
| CHST1 | 46.19748 | -1.49726 | 0.313634 | -4.77391 | 1.81E-06 | 1.13E-05 | 0.354226 |
| CRMP1 | 2231.616 | -0.43882 | 0.091956 | -4.77209 | 1.82E-06 | 1.14E-05 | 0.737736 |
| SIT1 | 68.18926 | -1.3075 | 0.274068 | -4.77073 | 1.84E-06 | 1.15E-05 | 0.40402 |
| STEAP3 | 692.0944 | -0.5929 | 0.124359 | -4.76764 | 1.86E-06 | 1.17E-05 | 0.663008 |
| TMEM238 | 44.46257 | -1.42957 | 0.299889 | -4.76699 | 1.87E-06 | 1.17E-05 | 0.371242 |
| RHOQ | 1507.076 | -0.58274 | 0.122248 | -4.76684 | 1.87E-06 | 1.17E-05 | 0.667696 |
| PAQR6 | 448.2198 | -1.30885 | 0.274679 | -4.76503 | 1.89E-06 | 1.18E-05 | 0.403641 |
| EDN1 | 51.55411 | -1.92212 | 0.403406 | -4.76472 | 1.89E-06 | 1.18E-05 | 0.263867 |
| PRKCSH | 6732.062 | -0.62775 | 0.131769 | -4.76405 | 1.90E-06 | 1.18E-05 | 0.647183 |
| CBX7 | 285.8956 | -0.82076 | 0.17235 | -4.76218 | 1.92E-06 | 1.19E-05 | 0.566142 |
| ATP8B1 | 48.35177 | -1.73272 | 0.364191 | -4.75771 | 1.96E-06 | 1.22E-05 | 0.300885 |
| KEL | 39.59001 | -2.07444 | 0.436033 | -4.75754 | 1.96E-06 | 1.22E-05 | 0.237427 |
| HIVEP2 | 318.792 | -1.58495 | 0.333408 | -4.75379 | 2.00E-06 | 1.24E-05 | 0.333336 |
| NPIPB15 | 94.52136 | -1.19884 | 0.252511 | -4.7477 | 2.06E-06 | 1.28E-05 | 0.435624 |
| BCL11A | 360.2766 | -1.2683 | 0.26726 | -4.74558 | 2.08E-06 | 1.29E-05 | 0.415148 |
| GNG4 | 753.3103 | -0.62617 | 0.132038 | -4.74236 | 2.11E-06 | 1.31E-05 | 0.647894 |
| AC005077 | 88.89331 | -0.90286 | 0.190413 | -4.74158 | 2.12E-06 | 1.31E-05 | 0.534826 |
| GNAO1 | 1247.448 | -0.50786 | 0.107136 | -4.74035 | 2.13E-06 | 1.32E-05 | 0.703263 |
| RHBG | 22.29194 | -2.76279 | 0.582974 | -4.73913 | 2.15E-06 | 1.33E-05 | 0.147339 |
| SLC13A3 | 537.8332 | -0.57281 | 0.120877 | -4.73874 | 2.15E-06 | 1.33E-05 | 0.672308 |
| CCDC17 | 31.65985 | -1.88881 | 0.398766 | -4.73664 | 2.17E-06 | 1.34E-05 | 0.27003 |
| PIGT | 1945.688 | -0.5851 | 0.123534 | -4.73633 | 2.18E-06 | 1.34E-05 | 0.666605 |
| FOXJ1 | 57.65438 | -1.6547 | 0.34956 | -4.73367 | 2.21E-06 | 1.36E-05 | 0.317603 |
| AC124319 | 2541.025 | -0.63604 | 0.134458 | -4.73045 | 2.24E-06 | 1.38E-05 | 0.643475 |
| RHPN1 | 893.9903 | -1.25492 | 0.265527 | -4.72615 | 2.29E-06 | 1.41E-05 | 0.419016 |
| MAP3K12 | 670.8967 | -1.01167 | 0.214076 | -4.72574 | 2.29E-06 | 1.41E-05 | 0.495972 |
| CPT1B | 854.4716 | -1.19258 | 0.252381 | -4.72533 | 2.30E-06 | 1.41E-05 | 0.437518 |
| IL32 | 30.58901 | -1.77915 | 0.376544 | -4.72496 | 2.30E-06 | 1.41E-05 | 0.291355 |
| RNF13 | 896.8815 | -0.5861 | 0.124114 | -4.72229 | 2.33E-06 | 1.43E-05 | 0.666141 |
| PLAAT3 | 1743.3 | -0.5911 | 0.125185 | -4.72182 | 2.34E-06 | 1.44E-05 | 0.663836 |
| VANGL1 | 737.0936 | -0.61969 | 0.13127 | -4.72077 | 2.35E-06 | 1.44E-05 | 0.650809 |
| GALNT18 | 199.3136 | -0.83227 | 0.176318 | -4.72032 | 2.35E-06 | 1.44E-05 | 0.561643 |
| CACNA2D: | 63.56882 | -1.3904 | 0.294654 | -4.71875 | 2.37E-06 | 1.45E-05 | 0.381459 |
| LGALS3 | 117.7235 | -0.94854 | 0.201092 | -4.71695 | 2.39E-06 | 1.46E-05 | 0.518157 |
| GIPR | 19.42679 | -2.83354 | 0.600786 | -4.71638 | 2.40E-06 | 1.47E-05 | 0.140288 |
| MAN2B1 | 2131.203 | -0.64562 | 0.136942 | -4.71454 | 2.42E-06 | 1.48E-05 | 0.63922 |
| FSIP2 | 443.8765 | -0.80425 | 0.170614 | -4.71384 | 2.43E-06 | 1.49E-05 | 0.572662 |
| ITGA7 | 2909.839 | -1.01011 | 0.214432 | -4.71064 | 2.47E-06 | 1.51E-05 | 0.496507 |

| | | | | | | | |
|---|---|---|---|---|---|---|---|
| TMEM165 | 1823.527 | -0.57135 | 0.121362 | -4.70783 | 2.50E-06 | 1.53E-05 | 0.672987 |
| GPR108 | 975.0863 | -0.57542 | 0.122268 | -4.70621 | 2.52E-06 | 1.54E-05 | 0.671091 |
| ADAMTS15 | 91.17565 | -1.20985 | 0.2571 | -4.70575 | 2.53E-06 | 1.54E-05 | 0.432313 |
| RDH16 | 52.28152 | -1.36008 | 0.289286 | -4.70151 | 2.58E-06 | 1.57E-05 | 0.38956 |
| KLC3 | 89.82686 | -1.08798 | 0.231423 | -4.70126 | 2.59E-06 | 1.57E-05 | 0.47042 |
| KLHL17 | 598.5741 | -1.15975 | 0.246752 | -4.70008 | 2.60E-06 | 1.58E-05 | 0.447589 |
| SLC4A3 | 300.429 | -0.96093 | 0.204474 | -4.69952 | 2.61E-06 | 1.58E-05 | 0.513725 |
| WASH6P | 918.0573 | -0.70801 | 0.150656 | -4.6995 | 2.61E-06 | 1.58E-05 | 0.612165 |
| NPIPB7 | 49.46432 | -1.67372 | 0.356522 | -4.69458 | 2.67E-06 | 1.62E-05 | 0.313444 |
| SUPT5H | 3507.709 | -0.53593 | 0.114162 | -4.69444 | 2.67E-06 | 1.62E-05 | 0.689716 |
| TSEN54 | 675.9727 | -0.58837 | 0.125337 | -4.69433 | 2.67E-06 | 1.62E-05 | 0.665092 |
| TCIRG1 | 466.9562 | -1.49393 | 0.318279 | -4.69377 | 2.68E-06 | 1.62E-05 | 0.355045 |
| ENPP1 | 826.6333 | -0.80749 | 0.172089 | -4.69228 | 2.70E-06 | 1.63E-05 | 0.571375 |
| GABRA5 | 274.6457 | -0.81247 | 0.173174 | -4.69167 | 2.71E-06 | 1.64E-05 | 0.569405 |
| RAPGEF3 | 91.68643 | -1.42484 | 0.303817 | -4.6898 | 2.73E-06 | 1.65E-05 | 0.37246 |
| KDELR1 | 4806.297 | -0.57705 | 0.123114 | -4.68716 | 2.77E-06 | 1.67E-05 | 0.670331 |
| UNKL | 600.5658 | -0.77148 | 0.164662 | -4.68523 | 2.80E-06 | 1.68E-05 | 0.585816 |
| SH2B3 | 1236.655 | -0.54883 | 0.117193 | -4.68314 | 2.83E-06 | 1.70E-05 | 0.683574 |
| TMEM221 | 122.305 | -0.88003 | 0.18803 | -4.68027 | 2.87E-06 | 1.72E-05 | 0.543357 |
| BMP6 | 78.40355 | -1.29082 | 0.27582 | -4.67992 | 2.87E-06 | 1.72E-05 | 0.408719 |
| VAMP8 | 217.0446 | -0.68619 | 0.146648 | -4.67916 | 2.88E-06 | 1.73E-05 | 0.621493 |
| CCDC114 | 68.08453 | -1.44202 | 0.3085 | -4.67429 | 2.95E-06 | 1.77E-05 | 0.368052 |
| ABCD4 | 1589.265 | -0.59308 | 0.126933 | -4.67239 | 2.98E-06 | 1.78E-05 | 0.662926 |
| TFPI2 | 87.06201 | -1.23047 | 0.263605 | -4.66787 | 3.04E-06 | 1.82E-05 | 0.426178 |
| MTRNR2L3 | 47.22142 | -1.76171 | 0.377565 | -4.66598 | 3.07E-06 | 1.83E-05 | 0.294898 |
| CSRNP1 | 212.2815 | -0.71447 | 0.153184 | -4.66413 | 3.10E-06 | 1.85E-05 | 0.60943 |
| GPS2 | 854.8699 | -0.54465 | 0.11678 | -4.6639 | 3.10E-06 | 1.85E-05 | 0.685557 |
| PTGES2 | 1260.802 | -0.46696 | 0.100162 | -4.66203 | 3.13E-06 | 1.87E-05 | 0.723488 |
| MAGI2 | 365.4285 | -0.61449 | 0.131891 | -4.65904 | 3.18E-06 | 1.89E-05 | 0.653162 |
| EPB41L3 | 634.063 | -0.72021 | 0.15463 | -4.65763 | 3.20E-06 | 1.91E-05 | 0.60701 |
| TMEM86B | 94.5554 | -1.31177 | 0.281688 | -4.65681 | 3.21E-06 | 1.91E-05 | 0.402827 |
| ITGB4 | 132.3804 | -2.18391 | 0.469279 | -4.65375 | 3.26E-06 | 1.94E-05 | 0.220079 |
| XKR7 | 285.8815 | -1.12782 | 0.242393 | -4.65286 | 3.27E-06 | 1.94E-05 | 0.457606 |
| CHTF18 | 2631.373 | -1.0942 | 0.235421 | -4.64786 | 3.35E-06 | 1.99E-05 | 0.468395 |
| PRCP | 2274.879 | -0.59698 | 0.128478 | -4.64659 | 3.37E-06 | 2.00E-05 | 0.661134 |
| TDRD5 | 24.9247 | -1.95716 | 0.421571 | -4.64254 | 3.44E-06 | 2.04E-05 | 0.257534 |
| TRPM4 | 316.5796 | -0.88417 | 0.190462 | -4.64226 | 3.45E-06 | 2.04E-05 | 0.541798 |
| GMPPA | 534.4139 | -0.4774 | 0.102911 | -4.63894 | 3.50E-06 | 2.07E-05 | 0.718272 |
| TIAF1 | 61.73075 | -1.7506 | 0.377378 | -4.63885 | 3.50E-06 | 2.07E-05 | 0.297178 |
| WDR18 | 1182.661 | -0.46942 | 0.101202 | -4.63844 | 3.51E-06 | 2.07E-05 | 0.722254 |
| DOCK4 | 528.7259 | -0.60543 | 0.130749 | -4.63052 | 3.65E-06 | 2.15E-05 | 0.657273 |
| IRF2BPL | 2492.477 | -0.75228 | 0.162501 | -4.62938 | 3.67E-06 | 2.16E-05 | 0.593666 |
| SIN3B | 2250.635 | -0.60829 | 0.131435 | -4.6281 | 3.69E-06 | 2.17E-05 | 0.655972 |
| ANKDD1A | 185.433 | -1.16365 | 0.251487 | -4.62707 | 3.71E-06 | 2.18E-05 | 0.446382 |

| | | | | | | | |
|---|---|---|---|---|---|---|---|
| GPRC5C | 603.2222 | -0.74833 | 0.161757 | -4.62625 | 3.72E-06 | 2.19E-05 | 0.595293 |
| MBP | 232.5397 | -0.99931 | 0.216029 | -4.62579 | 3.73E-06 | 2.20E-05 | 0.500241 |
| NDUFAF1 | 240.4427 | -0.67065 | 0.144994 | -4.62537 | 3.74E-06 | 2.20E-05 | 0.628222 |
| CASP8 | 169.6811 | -1.15891 | 0.250798 | -4.6209 | 3.82E-06 | 2.25E-05 | 0.44785 |
| ABHD8 | 417.9417 | -0.63303 | 0.13704 | -4.61933 | 3.85E-06 | 2.26E-05 | 0.64482 |
| BACE2 | 298.3541 | -0.84288 | 0.182487 | -4.61884 | 3.86E-06 | 2.27E-05 | 0.55753 |
| GPRC5B | 3391.953 | -0.53296 | 0.115403 | -4.61826 | 3.87E-06 | 2.27E-05 | 0.691136 |
| OR1D2 | 76.09969 | -1.12209 | 0.24316 | -4.6146 | 3.94E-06 | 2.31E-05 | 0.459429 |
| LASP1 | 4575.82 | -0.54891 | 0.118956 | -4.61441 | 3.94E-06 | 2.31E-05 | 0.683537 |
| APOBEC3F | 147.0027 | -0.87638 | 0.19 | -4.61254 | 3.98E-06 | 2.33E-05 | 0.544731 |
| BEX5 | 138.5111 | -1.04661 | 0.226905 | -4.61256 | 3.98E-06 | 2.33E-05 | 0.484103 |
| RFLNA | 72.20764 | -1.09682 | 0.237816 | -4.61204 | 3.99E-06 | 2.33E-05 | 0.467547 |
| RASGRP4 | 59.48393 | -1.42082 | 0.308133 | -4.61105 | 4.01E-06 | 2.35E-05 | 0.373501 |
| GGT7 | 824.9794 | -0.80214 | 0.174089 | -4.60765 | 4.07E-06 | 2.38E-05 | 0.573498 |
| PLXNB2 | 3647.948 | -0.75924 | 0.164791 | -4.6073 | 4.08E-06 | 2.38E-05 | 0.590806 |
| ILVBL | 1670.824 | -0.64602 | 0.140297 | -4.60468 | 4.13E-06 | 2.41E-05 | 0.63904 |
| DZANK1 | 203.4895 | -0.76674 | 0.166543 | -4.60384 | 4.15E-06 | 2.42E-05 | 0.587745 |
| STARD10 | 361.4887 | -0.69987 | 0.152224 | -4.59763 | 4.27E-06 | 2.49E-05 | 0.615629 |
| CXCR2 | 85.81423 | -1.95798 | 0.426198 | -4.59408 | 4.35E-06 | 2.53E-05 | 0.257388 |
| ZBTB46 | 629.921 | -0.52404 | 0.114125 | -4.59179 | 4.39E-06 | 2.55E-05 | 0.695423 |
| SHC2 | 563.6063 | -0.87151 | 0.190214 | -4.5817 | 4.61E-06 | 2.67E-05 | 0.546576 |
| ABCG2 | 43.05961 | -2.7808 | 0.607114 | -4.58035 | 4.64E-06 | 2.69E-05 | 0.145511 |
| ITGA9 | 119.6569 | -1.30616 | 0.285297 | -4.57826 | 4.69E-06 | 2.71E-05 | 0.404396 |
| PPP1R35 | 592.9568 | -0.61059 | 0.133367 | -4.57823 | 4.69E-06 | 2.71E-05 | 0.654931 |
| ATP6V0B | 1618.341 | -0.68739 | 0.150408 | -4.57018 | 4.87E-06 | 2.81E-05 | 0.620976 |
| STEAP2 | 544.6333 | -0.84402 | 0.184729 | -4.56894 | 4.90E-06 | 2.82E-05 | 0.557091 |
| ZNF229 | 9.690777 | -6.72711 | 1.472478 | -4.56856 | 4.91E-06 | 2.83E-05 | 0.009439 |
| KCNH6 | 52.99678 | -1.16141 | 0.254224 | -4.56846 | 4.91E-06 | 2.83E-05 | 0.447074 |
| RBM47 | 1169.788 | -0.80918 | 0.177157 | -4.56759 | 4.93E-06 | 2.84E-05 | 0.570706 |
| PNPLA5 | 98.90458 | -0.99942 | 0.218865 | -4.56637 | 4.96E-06 | 2.85E-05 | 0.500202 |
| ALKBH6 | 294.863 | -0.72457 | 0.158715 | -4.5652 | 4.99E-06 | 2.87E-05 | 0.605179 |
| AMN | 18.50133 | -3.98832 | 0.874711 | -4.55959 | 5.13E-06 | 2.94E-05 | 0.063008 |
| PLIN4 | 106.9913 | -0.85901 | 0.188474 | -4.55771 | 5.17E-06 | 2.96E-05 | 0.551331 |
| MAN2C1 | 1385.437 | -0.67055 | 0.147161 | -4.55656 | 5.20E-06 | 2.97E-05 | 0.628268 |
| QTRT1 | 1110.163 | -0.60001 | 0.131696 | -4.55602 | 5.21E-06 | 2.98E-05 | 0.65975 |
| LMF2 | 1274.38 | -0.72781 | 0.159825 | -4.55378 | 5.27E-06 | 3.01E-05 | 0.603822 |
| POLG2 | 473.0016 | -0.66337 | 0.145729 | -4.5521 | 5.31E-06 | 3.03E-05 | 0.6314 |
| MMP14 | 2084.507 | -0.50267 | 0.110527 | -4.54796 | 5.42E-06 | 3.09E-05 | 0.705797 |
| PLAC9 | 60.04511 | -1.22482 | 0.269752 | -4.54052 | 5.61E-06 | 3.19E-05 | 0.427852 |
| REEP4 | 784.6951 | -0.56848 | 0.125258 | -4.53851 | 5.67E-06 | 3.22E-05 | 0.674325 |
| NSUN5P2 | 623.246 | -1.07421 | 0.23682 | -4.53597 | 5.73E-06 | 3.25E-05 | 0.474932 |
| TACC2 | 1842.246 | -0.45522 | 0.100358 | -4.53598 | 5.73E-06 | 3.25E-05 | 0.729397 |
| KMT5A | 878.8601 | -0.52147 | 0.114969 | -4.53569 | 5.74E-06 | 3.26E-05 | 0.696664 |
| SH3RF3 | 43.13982 | -1.76186 | 0.38889 | -4.53049 | 5.88E-06 | 3.34E-05 | 0.294867 |

| | | | | | | | |
|---|---|---|---|---|---|---|---|
| GUCY2C | 24.64195 | -2.34823 | 0.518673 | -4.52738 | 5.97E-06 | 3.38E-05 | 0.196387 |
| NT5C | 785.7513 | -0.55094 | 0.121694 | -4.5273 | 5.97E-06 | 3.38E-05 | 0.682573 |
| RABGAP1L | 2380.768 | -0.51944 | 0.114768 | -4.52603 | 6.01E-06 | 3.40E-05 | 0.697641 |
| ELOVL7 | 282.347 | -0.59683 | 0.131903 | -4.52477 | 6.05E-06 | 3.42E-05 | 0.661204 |
| KLF9 | 38.87975 | -1.63285 | 0.361078 | -4.52216 | 6.12E-06 | 3.46E-05 | 0.32245 |
| IGF2R | 6658.562 | -0.59942 | 0.132572 | -4.52145 | 6.14E-06 | 3.47E-05 | 0.660021 |
| DVL1 | 2000.444 | -0.55833 | 0.123489 | -4.5213 | 6.15E-06 | 3.47E-05 | 0.679087 |
| ARHGEF10 | 1205.569 | -0.51526 | 0.11399 | -4.52019 | 6.18E-06 | 3.49E-05 | 0.699669 |
| LRRC2 | 106.3708 | -2.08519 | 0.461442 | -4.51887 | 6.22E-06 | 3.51E-05 | 0.235665 |
| CYTL1 | 38.4152 | -1.54736 | 0.342442 | -4.51862 | 6.22E-06 | 3.51E-05 | 0.342135 |
| SEMA3F | 1088.917 | -0.8441 | 0.186893 | -4.5165 | 6.29E-06 | 3.54E-05 | 0.557057 |
| TMEM129 | 1255.306 | -0.48672 | 0.107765 | -4.51647 | 6.29E-06 | 3.54E-05 | 0.713646 |
| CAPN5 | 479.8415 | -0.52636 | 0.116685 | -4.51095 | 6.45E-06 | 3.63E-05 | 0.694304 |
| DNAJC1 | 467.5756 | -0.44428 | 0.098567 | -4.50734 | 6.56E-06 | 3.69E-05 | 0.734952 |
| CLCNKA | 21.89949 | -2.21919 | 0.492376 | -4.5071 | 6.57E-06 | 3.69E-05 | 0.214762 |
| STUB1 | 1232.568 | -0.42328 | 0.093934 | -4.50607 | 6.60E-06 | 3.71E-05 | 0.74573 |
| NADK | 1853.089 | -0.50183 | 0.111383 | -4.50542 | 6.62E-06 | 3.72E-05 | 0.706212 |
| GRINA | 2154.649 | -0.63436 | 0.140812 | -4.50505 | 6.64E-06 | 3.72E-05 | 0.644225 |
| PLOD1 | 1826.1 | -0.58144 | 0.129137 | -4.5025 | 6.72E-06 | 3.77E-05 | 0.668297 |
| RNF166 | 620.8372 | -0.57396 | 0.127499 | -4.50164 | 6.74E-06 | 3.78E-05 | 0.671773 |
| NPDC1 | 1010.625 | -1.06835 | 0.237443 | -4.49942 | 6.81E-06 | 3.81E-05 | 0.476863 |
| PCDHA13 | 17.99597 | -2.1904 | 0.486922 | -4.49847 | 6.84E-06 | 3.83E-05 | 0.21909 |
| WDR27 | 1034.146 | -0.71307 | 0.158774 | -4.49112 | 7.09E-06 | 3.95E-05 | 0.610019 |
| TEC | 55.58 | -1.50181 | 0.334636 | -4.48789 | 7.19E-06 | 4.01E-05 | 0.35311 |
| SLC9A8 | 473.3976 | -0.52666 | 0.117369 | -4.4872 | 7.22E-06 | 4.02E-05 | 0.694161 |
| MACO1 | 460.0096 | -0.59732 | 0.133153 | -4.48598 | 7.26E-06 | 4.04E-05 | 0.660981 |
| PNMA6A | 163.2542 | -0.6996 | 0.155955 | -4.48587 | 7.26E-06 | 4.05E-05 | 0.615745 |
| BNC2 | 570.9504 | -0.98555 | 0.219709 | -4.48573 | 7.27E-06 | 4.05E-05 | 0.505031 |
| EPS8L1 | 435.7173 | -0.90192 | 0.201136 | -4.48414 | 7.32E-06 | 4.07E-05 | 0.535174 |
| CYP11A1 | 109.362 | -0.93306 | 0.208203 | -4.48146 | 7.41E-06 | 4.13E-05 | 0.523748 |
| CYB5A | 765.735 | -0.63718 | 0.142258 | -4.47904 | 7.50E-06 | 4.17E-05 | 0.642968 |
| PLEKHH1 | 1019.065 | -0.58434 | 0.130492 | -4.47797 | 7.54E-06 | 4.19E-05 | 0.666956 |
| IGLON5 | 1395.91 | -0.49792 | 0.111196 | -4.47784 | 7.54E-06 | 4.19E-05 | 0.708127 |
| SOD3 | 149.9502 | -0.8927 | 0.199402 | -4.47691 | 7.57E-06 | 4.21E-05 | 0.538604 |
| ARHGAP8 | 414.1475 | -0.60669 | 0.135564 | -4.47532 | 7.63E-06 | 4.23E-05 | 0.6567 |
| MTATP6P1 | 10225.04 | -1.30149 | 0.290892 | -4.47414 | 7.67E-06 | 4.25E-05 | 0.405706 |
| HEXB | 1473.061 | -0.49939 | 0.111679 | -4.4717 | 7.76E-06 | 4.29E-05 | 0.707404 |
| BCL3 | 38.21989 | -1.75704 | 0.392946 | -4.47145 | 7.77E-06 | 4.30E-05 | 0.295855 |
| SHOX | 27.59495 | -2.079 | 0.465031 | -4.47066 | 7.80E-06 | 4.31E-05 | 0.236679 |
| DIAPH1 | 4062.783 | -0.74554 | 0.16682 | -4.46911 | 7.85E-06 | 4.34E-05 | 0.596446 |
| KIAA0895L | 684.9786 | -1.08769 | 0.243456 | -4.4677 | 7.91E-06 | 4.37E-05 | 0.470515 |
| PTPRN | 102.7095 | -1.88541 | 0.422149 | -4.46621 | 7.96E-06 | 4.39E-05 | 0.270668 |
| PLBD1 | 315.3729 | -0.63705 | 0.14267 | -4.46524 | 8.00E-06 | 4.41E-05 | 0.643024 |
| KCNJ6 | 35.85948 | -2.5114 | 0.56315 | -4.45956 | 8.21E-06 | 4.52E-05 | 0.175385 |

| | | | | | | | |
|---|---|---|---|---|---|---|---|
| LGMN | 1751.361 | -0.57429 | 0.128784 | -4.45933 | 8.22E-06 | 4.53E-05 | 0.671616 |
| ZNF488 | 88.83619 | -0.96751 | 0.216991 | -4.45874 | 8.24E-06 | 4.54E-05 | 0.511389 |
| CHMP1B2I | 136.6014 | -0.88156 | 0.197735 | -4.45827 | 8.26E-06 | 4.55E-05 | 0.542781 |
| CLTCL1 | 623.1005 | -0.72966 | 0.163761 | -4.45565 | 8.36E-06 | 4.60E-05 | 0.603046 |
| SLC5A2 | 32.29807 | -2.02124 | 0.453661 | -4.45539 | 8.37E-06 | 4.60E-05 | 0.246346 |
| COL7A1 | 631.1208 | -1.76197 | 0.395574 | -4.45421 | 8.42E-06 | 4.63E-05 | 0.294845 |
| ST6GALNA | 253.6154 | -0.64282 | 0.144319 | -4.45414 | 8.42E-06 | 4.63E-05 | 0.640461 |
| LHPP | 451.8755 | -0.55212 | 0.123973 | -4.45357 | 8.45E-06 | 4.64E-05 | 0.682016 |
| CFP | 30.486 | -2.72462 | 0.61228 | -4.44996 | 8.59E-06 | 4.71E-05 | 0.151289 |
| MMP15 | 1822.423 | -0.62276 | 0.140014 | -4.44782 | 8.67E-06 | 4.75E-05 | 0.649428 |
| C16orf89 | 58.50933 | -1.1714 | 0.263553 | -4.44463 | 8.80E-06 | 4.82E-05 | 0.443991 |
| SELENOO | 462.7269 | -0.84234 | 0.189541 | -4.44411 | 8.83E-06 | 4.83E-05 | 0.557738 |
| PDIA4 | 6027.779 | -0.47333 | 0.106571 | -4.44151 | 8.93E-06 | 4.88E-05 | 0.720298 |
| NLRP12 | 62.04023 | -1.20103 | 0.27048 | -4.44036 | 8.98E-06 | 4.91E-05 | 0.434965 |
| ARHGAP33 | 480.4554 | -0.92141 | 0.207546 | -4.43953 | 9.02E-06 | 4.92E-05 | 0.527993 |
| MAPK12 | 600.9957 | -0.67211 | 0.151407 | -4.43909 | 9.03E-06 | 4.93E-05 | 0.627589 |
| ADCY8 | 218.1751 | -1.38828 | 0.312837 | -4.43771 | 9.09E-06 | 4.96E-05 | 0.382019 |
| GRIN2C | 11.4157 | -2.79112 | 0.629218 | -4.43586 | 9.17E-06 | 5.00E-05 | 0.144473 |
| AC243772 | 19.11424 | -2.32369 | 0.523953 | -4.43492 | 9.21E-06 | 5.02E-05 | 0.199756 |
| MUS81 | 839.2814 | -0.74024 | 0.167022 | -4.432 | 9.34E-06 | 5.08E-05 | 0.598639 |
| SPSB2 | 318.0568 | -0.77792 | 0.175682 | -4.42797 | 9.51E-06 | 5.17E-05 | 0.583209 |
| SVOPL | 75.56412 | -1.54541 | 0.349161 | -4.42606 | 9.60E-06 | 5.22E-05 | 0.342599 |
| AC005954 | 378.3386 | -0.7327 | 0.165559 | -4.42564 | 9.62E-06 | 5.22E-05 | 0.601775 |
| CORO6 | 123.4939 | -1.54648 | 0.34959 | -4.42369 | 9.70E-06 | 5.27E-05 | 0.342345 |
| CCM2L | 271.1727 | -0.52327 | 0.118306 | -4.42301 | 9.73E-06 | 5.28E-05 | 0.695794 |
| AUP1 | 1704.582 | -0.64561 | 0.145975 | -4.42277 | 9.74E-06 | 5.28E-05 | 0.639221 |
| GMDS | 515.6417 | -0.52615 | 0.118973 | -4.42243 | 9.76E-06 | 5.29E-05 | 0.694405 |
| KCNN4 | 18.22587 | -2.33714 | 0.528578 | -4.42157 | 9.80E-06 | 5.31E-05 | 0.197902 |
| GRID2 | 508.242 | -0.63075 | 0.142689 | -4.42043 | 9.85E-06 | 5.33E-05 | 0.645843 |
| BRSK2 | 578.9884 | -0.57845 | 0.130879 | -4.4197 | 9.88E-06 | 5.35E-05 | 0.669685 |
| FOXL2 | 132.6446 | -1.12524 | 0.25461 | -4.41947 | 9.89E-06 | 5.35E-05 | 0.458426 |
| MEGF6 | 1044.899 | -1.0643 | 0.240828 | -4.41935 | 9.90E-06 | 5.35E-05 | 0.478204 |
| SDHAP3 | 256.6251 | -1.07099 | 0.242359 | -4.41901 | 9.92E-06 | 5.36E-05 | 0.475993 |
| STAMBPL1 | 159.3489 | -0.9316 | 0.210819 | -4.41896 | 9.92E-06 | 5.36E-05 | 0.524277 |
| GALNT14 | 170.7662 | -1.04863 | 0.237312 | -4.41876 | 9.93E-06 | 5.36E-05 | 0.483428 |
| FN1 | 1695.689 | -0.56222 | 0.127287 | -4.41698 | 1.00E-05 | 5.40E-05 | 0.677257 |
| MAFA | 15.65319 | -2.93566 | 0.664748 | -4.4162 | 1.00E-05 | 5.41E-05 | 0.130701 |
| CLCN7 | 2821.861 | -0.81072 | 0.183593 | -4.41587 | 1.01E-05 | 5.42E-05 | 0.570097 |
| RPN1 | 5501.653 | -0.43248 | 0.098101 | -4.40849 | 1.04E-05 | 5.60E-05 | 0.740989 |
| PRRT4 | 23.46976 | -1.79616 | 0.407455 | -4.40824 | 1.04E-05 | 5.60E-05 | 0.28794 |
| OPTN | 788.7832 | -0.61632 | 0.139826 | -4.40775 | 1.04E-05 | 5.61E-05 | 0.652334 |
| L3MBTL4 | 66.06222 | -1.3413 | 0.304369 | -4.40681 | 1.05E-05 | 5.64E-05 | 0.394666 |
| CHKA | 1054.516 | -0.7092 | 0.160991 | -4.40521 | 1.06E-05 | 5.67E-05 | 0.61166 |
| GAK | 1601.577 | -0.60301 | 0.136889 | -4.40514 | 1.06E-05 | 5.67E-05 | 0.658377 |

| | | | | | | | |
|---|---|---|---|---|---|---|---|
| PTH1R | 38.89877 | -2.00892 | 0.45615 | -4.40407 | 1.06E-05 | 5.70E-05 | 0.24846 |
| AC068587 | 37.75859 | -1.4165 | 0.321677 | -4.40348 | 1.07E-05 | 5.71E-05 | 0.37462 |
| CRYM | 118.6618 | -0.94204 | 0.213937 | -4.40334 | 1.07E-05 | 5.72E-05 | 0.520496 |
| DPPA4 | 10320.12 | -0.62839 | 0.142724 | -4.40286 | 1.07E-05 | 5.73E-05 | 0.646896 |
| LHX4 | 212.8019 | -0.83715 | 0.190227 | -4.40082 | 1.08E-05 | 5.78E-05 | 0.559747 |
| THEM6 | 135.8253 | -0.83152 | 0.188966 | -4.40039 | 1.08E-05 | 5.79E-05 | 0.561936 |
| IDH3A | 468.9973 | -0.7533 | 0.171228 | -4.3994 | 1.09E-05 | 5.81E-05 | 0.593244 |
| KCNQ5 | 20.90154 | -3.53037 | 0.802664 | -4.39832 | 1.09E-05 | 5.84E-05 | 0.086547 |
| ZNF135 | 8.757334 | -4.49021 | 1.020972 | -4.39798 | 1.09E-05 | 5.84E-05 | 0.044495 |
| PKN3 | 581.5029 | -0.5933 | 0.134959 | -4.39616 | 1.10E-05 | 5.89E-05 | 0.662823 |
| APCDD1L | 36.53269 | -2.32759 | 0.529528 | -4.39561 | 1.10E-05 | 5.90E-05 | 0.199216 |
| AC138969 | 2206.903 | -0.89484 | 0.203739 | -4.39209 | 1.12E-05 | 5.99E-05 | 0.537807 |
| PDZD7 | 35.80601 | -2.05675 | 0.46838 | -4.39121 | 1.13E-05 | 6.01E-05 | 0.240357 |
| NRBP2 | 472.5307 | -0.9458 | 0.215406 | -4.39076 | 1.13E-05 | 6.02E-05 | 0.519143 |
| METTL26 | 687.0772 | -0.55329 | 0.126062 | -4.38905 | 1.14E-05 | 6.07E-05 | 0.681463 |
| AC005747 | 206.1516 | -1.00055 | 0.228065 | -4.38714 | 1.15E-05 | 6.12E-05 | 0.499808 |
| SLC7A8 | 5106.644 | -0.46086 | 0.1051 | -4.38495 | 1.16E-05 | 6.18E-05 | 0.726554 |
| ICA1 | 450.0812 | -0.82268 | 0.187661 | -4.38386 | 1.17E-05 | 6.20E-05 | 0.565391 |
| HS3ST3B1 | 253.6905 | -0.71412 | 0.162948 | -4.38251 | 1.17E-05 | 6.23E-05 | 0.609578 |
| EHD2 | 680.607 | -0.51557 | 0.117691 | -4.38075 | 1.18E-05 | 6.28E-05 | 0.699515 |
| SLC30A2 | 14.68091 | -3.5571 | 0.812435 | -4.37831 | 1.20E-05 | 6.34E-05 | 0.084959 |
| ICAM4 | 16.70556 | -2.75732 | 0.629891 | -4.37745 | 1.20E-05 | 6.36E-05 | 0.147899 |
| AKR1C3 | 29.86237 | -2.22254 | 0.50777 | -4.37707 | 1.20E-05 | 6.37E-05 | 0.214264 |
| PKMYT1 | 1229.522 | -0.7292 | 0.16663 | -4.37619 | 1.21E-05 | 6.40E-05 | 0.603237 |
| LPCAT3 | 1227.698 | -0.47088 | 0.107677 | -4.37309 | 1.22E-05 | 6.48E-05 | 0.721523 |
| MADCAM1 | 27.75192 | -2.08341 | 0.476599 | -4.37141 | 1.23E-05 | 6.53E-05 | 0.235956 |
| TRIM69 | 122.554 | -1.4093 | 0.322559 | -4.36912 | 1.25E-05 | 6.59E-05 | 0.376494 |
| LCAT | 103.4352 | -1.40988 | 0.322711 | -4.36887 | 1.25E-05 | 6.60E-05 | 0.376343 |
| PAH | 126.4043 | -1.17339 | 0.268657 | -4.36763 | 1.26E-05 | 6.63E-05 | 0.443377 |
| PCSK9 | 520.0189 | -1.22102 | 0.279712 | -4.3653 | 1.27E-05 | 6.70E-05 | 0.428978 |
| TMEM54 | 178.1212 | -0.89872 | 0.205913 | -4.36455 | 1.27E-05 | 6.72E-05 | 0.536364 |
| FOXO4 | 322.1675 | -0.67985 | 0.155773 | -4.36434 | 1.28E-05 | 6.72E-05 | 0.624231 |
| TSPAN15 | 255.4964 | -0.72391 | 0.16594 | -4.36248 | 1.29E-05 | 6.77E-05 | 0.605455 |
| CPNE9 | 67.45941 | -1.37037 | 0.31419 | -4.3616 | 1.29E-05 | 6.79E-05 | 0.386792 |
| NFE2L1 | 11604.58 | -0.45863 | 0.105205 | -4.35937 | 1.30E-05 | 6.86E-05 | 0.727679 |
| FTLP14 | 13.2251 | -3.40393 | 0.781499 | -4.35564 | 1.33E-05 | 6.97E-05 | 0.094475 |
| P3H2 | 2019.863 | -0.90127 | 0.206922 | -4.35561 | 1.33E-05 | 6.97E-05 | 0.535415 |
| DDX51 | 827.0425 | -1.0146 | 0.232945 | -4.35552 | 1.33E-05 | 6.97E-05 | 0.494967 |
| PMM2 | 922.9422 | -0.47717 | 0.109577 | -4.35466 | 1.33E-05 | 7.00E-05 | 0.718385 |
| FERMT1 | 403.0436 | -0.55216 | 0.126828 | -4.35361 | 1.34E-05 | 7.03E-05 | 0.681998 |
| TPH2 | 111.8718 | -0.92491 | 0.212558 | -4.35134 | 1.35E-05 | 7.10E-05 | 0.526712 |
| SLC19A1 | 887.263 | -0.59409 | 0.13663 | -4.34818 | 1.37E-05 | 7.19E-05 | 0.662461 |
| CCS | 764.4694 | -0.5441 | 0.12517 | -4.34693 | 1.38E-05 | 7.23E-05 | 0.685818 |
| VAMP5 | 10.47337 | -3.09293 | 0.71177 | -4.34541 | 1.39E-05 | 7.28E-05 | 0.117202 |

| | | | | | | | |
|---|---|---|---|---|---|---|---|
| C6orf223 | 43.17463 | -1.54242 | 0.354982 | -4.34506 | 1.39E-05 | 7.29E-05 | 0.34331 |
| ZSWIM9 | 613.5966 | -0.73764 | 0.169917 | -4.34116 | 1.42E-05 | 7.40E-05 | 0.599721 |
| NID1 | 1306.869 | -0.4788 | 0.11031 | -4.34052 | 1.42E-05 | 7.42E-05 | 0.717574 |
| ANKS3 | 509.7081 | -0.61704 | 0.142253 | -4.33759 | 1.44E-05 | 7.51E-05 | 0.652009 |
| NBEAL2 | 1961.309 | -1.50548 | 0.347191 | -4.33618 | 1.45E-05 | 7.55E-05 | 0.352212 |
| DAPK3 | 1318.417 | -0.43265 | 0.099802 | -4.33514 | 1.46E-05 | 7.58E-05 | 0.740897 |
| LRP5L | 105.8591 | -1.12446 | 0.259651 | -4.33067 | 1.49E-05 | 7.73E-05 | 0.458673 |
| ANKRD34E | 73.125 | -1.32965 | 0.307052 | -4.33038 | 1.49E-05 | 7.73E-05 | 0.397864 |
| NPIPB14P | 112.3334 | -1.07137 | 0.247406 | -4.33039 | 1.49E-05 | 7.73E-05 | 0.475868 |
| FGF2 | 3142.436 | -0.87807 | 0.202789 | -4.32998 | 1.49E-05 | 7.74E-05 | 0.544094 |
| C2orf40 | 30.43353 | -1.46618 | 0.338794 | -4.32765 | 1.51E-05 | 7.82E-05 | 0.361939 |
| CRYBB2P1 | 329.5082 | -0.48373 | 0.111998 | -4.3191 | 1.57E-05 | 8.12E-05 | 0.715125 |
| FAM110C | 183.931 | -1.13307 | 0.262681 | -4.31348 | 1.61E-05 | 8.32E-05 | 0.455945 |
| CEBPZOS | 436.9205 | -0.71009 | 0.164714 | -4.31107 | 1.62E-05 | 8.40E-05 | 0.611281 |
| TPRN | 480.5697 | -0.75997 | 0.176338 | -4.30971 | 1.63E-05 | 8.44E-05 | 0.59051 |
| NMUR1 | 16.86731 | -2.23069 | 0.51832 | -4.3037 | 1.68E-05 | 8.67E-05 | 0.213057 |
| AGAP9 | 169.5508 | -1.03415 | 0.24038 | -4.30214 | 1.69E-05 | 8.72E-05 | 0.488304 |
| MGAT5B | 308.3119 | -0.66616 | 0.15488 | -4.30112 | 1.70E-05 | 8.76E-05 | 0.630183 |
| PROM1 | 1138.769 | -0.52135 | 0.121253 | -4.29973 | 1.71E-05 | 8.81E-05 | 0.696718 |
| NAALADL1 | 68.42619 | -1.50631 | 0.350383 | -4.29903 | 1.72E-05 | 8.83E-05 | 0.35201 |
| QPCT | 214.6179 | -0.68841 | 0.160328 | -4.29374 | 1.76E-05 | 9.03E-05 | 0.620538 |
| NFKBIB | 740.9342 | -0.52254 | 0.121768 | -4.29127 | 1.78E-05 | 9.13E-05 | 0.696145 |
| GEMIN7 | 480.4742 | -0.65627 | 0.153022 | -4.28876 | 1.80E-05 | 9.23E-05 | 0.634515 |
| RNA5-8SP( | 68.56381 | -1.77924 | 0.415447 | -4.28271 | 1.85E-05 | 9.47E-05 | 0.291336 |
| MYL5 | 175.7187 | -0.90012 | 0.210305 | -4.28008 | 1.87E-05 | 9.58E-05 | 0.535842 |
| PAQR7 | 186.6869 | -0.82634 | 0.193375 | -4.27325 | 1.93E-05 | 9.86E-05 | 0.563958 |
| CYP46A1 | 70.96487 | -1.37229 | 0.321174 | -4.27273 | 1.93E-05 | 9.88E-05 | 0.386278 |
| CYSLTR1 | 122.6976 | -0.96235 | 0.225275 | -4.27188 | 1.94E-05 | 9.91E-05 | 0.513221 |
| KIF1A | 16785.68 | -0.41802 | 0.097874 | -4.27094 | 1.95E-05 | 9.95E-05 | 0.748453 |
| MACF1 | 8270.117 | -0.66833 | 0.156486 | -4.27083 | 1.95E-05 | 9.95E-05 | 0.629236 |
| ASPHD1 | 198.9095 | -0.67536 | 0.158155 | -4.27022 | 1.95E-05 | 9.98E-05 | 0.626177 |
| CAPN15 | 1904.521 | -0.612 | 0.143345 | -4.26944 | 1.96E-05 | 0.0001 | 0.654288 |
| DEPTOR | 69.90022 | -1.44806 | 0.339522 | -4.265 | 2.00E-05 | 0.000102 | 0.366514 |
| SULT1A1 | 101.7195 | -0.82054 | 0.19248 | -4.26297 | 2.02E-05 | 0.000103 | 0.566231 |
| TFR2 | 99.26461 | -1.19308 | 0.280139 | -4.2589 | 2.05E-05 | 0.000105 | 0.437367 |
| DHRS7 | 601.3994 | -0.57795 | 0.135779 | -4.25657 | 2.08E-05 | 0.000106 | 0.669914 |
| RUBCNL | 84.37902 | -0.97317 | 0.228647 | -4.25621 | 2.08E-05 | 0.000106 | 0.509386 |
| KIAA1614 | 130.3889 | -0.83804 | 0.196919 | -4.25574 | 2.08E-05 | 0.000106 | 0.559404 |
| CARMIL3 | 392.5128 | -1.08312 | 0.254513 | -4.25564 | 2.08E-05 | 0.000106 | 0.472008 |
| ATP6V0C | 3652.431 | -0.55373 | 0.130153 | -4.25442 | 2.10E-05 | 0.000106 | 0.681258 |
| GDF3 | 23.17519 | -2.14485 | 0.504332 | -4.25284 | 2.11E-05 | 0.000107 | 0.226119 |
| SLC26A6 | 1182.87 | -0.92142 | 0.216735 | -4.25135 | 2.12E-05 | 0.000108 | 0.527991 |
| EPHB6 | 152.1127 | -1.79132 | 0.421523 | -4.24963 | 2.14E-05 | 0.000109 | 0.288908 |
| NSG1 | 418.7613 | -0.66413 | 0.156467 | -4.24455 | 2.19E-05 | 0.000111 | 0.631068 |

| | | | | | | | |
|---|---|---|---|---|---|---|---|
| CKMT1A | 574.6093 | -0.61111 | 0.144187 | -4.23831 | 2.25E-05 | 0.000114 | 0.654694 |
| SPPL2B | 1762.298 | -0.63512 | 0.149911 | -4.23663 | 2.27E-05 | 0.000115 | 0.643889 |
| IQGAP1 | 5589.21 | -0.94281 | 0.222606 | -4.23535 | 2.28E-05 | 0.000115 | 0.520218 |
| RUNX3 | 17.23942 | -4.63318 | 1.094001 | -4.23508 | 2.28E-05 | 0.000116 | 0.040297 |
| CORO2A | 733.1282 | -0.60727 | 0.143394 | -4.23498 | 2.29E-05 | 0.000116 | 0.656437 |
| PRDX4 | 4214.768 | -0.57515 | 0.135914 | -4.23173 | 2.32E-05 | 0.000117 | 0.671216 |
| ITLN2 | 34.77484 | -1.40315 | 0.331681 | -4.23041 | 2.33E-05 | 0.000118 | 0.378104 |
| TAC1 | 218.7331 | -0.71264 | 0.16853 | -4.2286 | 2.35E-05 | 0.000119 | 0.610201 |
| NLRP10 | 15.59081 | -3.76037 | 0.889519 | -4.22742 | 2.36E-05 | 0.000119 | 0.073793 |
| DUSP26 | 41.13144 | -1.77712 | 0.42059 | -4.22531 | 2.39E-05 | 0.00012 | 0.291765 |
| TCTA | 309.6239 | -0.72395 | 0.171474 | -4.22194 | 2.42E-05 | 0.000122 | 0.605435 |
| SCPEP1 | 953.9962 | -0.67426 | 0.159707 | -4.22184 | 2.42E-05 | 0.000122 | 0.626654 |
| INF2 | 1376.434 | -0.7092 | 0.168096 | -4.21899 | 2.45E-05 | 0.000123 | 0.611661 |
| HHIP | 51.29842 | -2.44329 | 0.579224 | -4.21822 | 2.46E-05 | 0.000124 | 0.183863 |
| TMEM266 | 116.6499 | -0.97889 | 0.232229 | -4.21521 | 2.50E-05 | 0.000125 | 0.507369 |
| MRGPRF | 210.1863 | -0.81358 | 0.193026 | -4.21488 | 2.50E-05 | 0.000125 | 0.568967 |
| CBARP | 663.5152 | -0.63115 | 0.149751 | -4.21466 | 2.50E-05 | 0.000125 | 0.645662 |
| NTNG1 | 26.24257 | -2.58967 | 0.61454 | -4.21399 | 2.51E-05 | 0.000126 | 0.166124 |
| APOBEC3( | 59.49065 | -1.163 | 0.276017 | -4.21352 | 2.51E-05 | 0.000126 | 0.446582 |
| AMBP | 29.51097 | -1.58276 | 0.375766 | -4.2121 | 2.53E-05 | 0.000127 | 0.333842 |
| AGAP6 | 381.2873 | -0.66943 | 0.158957 | -4.21139 | 2.54E-05 | 0.000127 | 0.628755 |
| VWA2 | 99.81893 | -0.89868 | 0.213421 | -4.21082 | 2.54E-05 | 0.000127 | 0.536378 |
| GOLGA2P5 | 346.5495 | -0.79814 | 0.189645 | -4.20862 | 2.57E-05 | 0.000128 | 0.575089 |
| PIGL | 321.6742 | -0.56925 | 0.135281 | -4.20787 | 2.58E-05 | 0.000129 | 0.673969 |
| LGALS12 | 16.57211 | -2.07871 | 0.494016 | -4.20778 | 2.58E-05 | 0.000129 | 0.236726 |
| FAAH | 244.022 | -1.25185 | 0.297622 | -4.20616 | 2.60E-05 | 0.00013 | 0.41991 |
| EMC9 | 385.7509 | -0.48347 | 0.114956 | -4.20572 | 2.60E-05 | 0.00013 | 0.715254 |
| POFUT2 | 1145.192 | -0.58998 | 0.140308 | -4.20491 | 2.61E-05 | 0.00013 | 0.664351 |
| AJAP1 | 34.59192 | -2.59709 | 0.618093 | -4.20177 | 2.65E-05 | 0.000132 | 0.165272 |
| B3GNT9 | 233.0996 | -0.54482 | 0.129725 | -4.1998 | 2.67E-05 | 0.000133 | 0.685478 |
| GNB3 | 198.9308 | -1.34121 | 0.319419 | -4.1989 | 2.68E-05 | 0.000133 | 0.39469 |
| CA9 | 20.39368 | -2.03739 | 0.485336 | -4.19789 | 2.69E-05 | 0.000134 | 0.243605 |
| C19orf71 | 20.76846 | -2.37683 | 0.566357 | -4.1967 | 2.71E-05 | 0.000135 | 0.192532 |
| MT-CYB | 73717.29 | -1.35221 | 0.322216 | -4.1966 | 2.71E-05 | 0.000135 | 0.391691 |
| TMEM161E | 400.5317 | -0.54909 | 0.13086 | -4.19599 | 2.72E-05 | 0.000135 | 0.683451 |
| COLCA2 | 44.60385 | -1.27792 | 0.304798 | -4.19269 | 2.76E-05 | 0.000137 | 0.412389 |
| FAM78B | 56.43261 | -1.03713 | 0.247526 | -4.19 | 2.79E-05 | 0.000138 | 0.487295 |
| ZNF598 | 1293.939 | -0.68239 | 0.162881 | -4.18952 | 2.80E-05 | 0.000138 | 0.623131 |
| ZMYND15 | 12.16572 | -2.57073 | 0.615019 | -4.17991 | 2.92E-05 | 0.000144 | 0.16832 |
| DTNA | 621.5267 | -0.65487 | 0.156709 | -4.17888 | 2.93E-05 | 0.000145 | 0.635134 |
| PON3 | 4.333241 | -5.57716 | 1.335243 | -4.17688 | 2.96E-05 | 0.000146 | 0.020946 |
| SCN1B | 126.6063 | -0.72792 | 0.174303 | -4.17619 | 2.96E-05 | 0.000146 | 0.603774 |
| TMEM206 | 459.979 | -0.60168 | 0.144119 | -4.17489 | 2.98E-05 | 0.000147 | 0.658986 |
| TMIE | 19.5966 | -2.15155 | 0.51535 | -4.17494 | 2.98E-05 | 0.000147 | 0.22507 |

| | | | | | | | |
|---|---|---|---|---|---|---|---|
| 9-Mar | 223.2026 | -0.78555 | 0.188176 | -4.17456 | 2.99E-05 | 0.000147 | 0.580129 |
| LRFN4 | 1006.871 | -0.65112 | 0.156045 | -4.17265 | 3.01E-05 | 0.000148 | 0.636786 |
| INTS1 | 4558.428 | -0.60137 | 0.144349 | -4.16608 | 3.10E-05 | 0.000152 | 0.659127 |
| AL133216. | 227.5995 | -0.6232 | 0.149926 | -4.15674 | 3.23E-05 | 0.000158 | 0.649228 |
| TRPT1 | 392.8441 | -0.49177 | 0.118318 | -4.1564 | 3.23E-05 | 0.000159 | 0.71115 |
| REX1BD | 810.7838 | -0.5277 | 0.126994 | -4.15535 | 3.25E-05 | 0.000159 | 0.693658 |
| SLC46A3 | 20.81557 | -2.02261 | 0.486911 | -4.15397 | 3.27E-05 | 0.00016 | 0.246112 |
| MAP3K14 | 330.9889 | -0.61843 | 0.148923 | -4.15269 | 3.29E-05 | 0.000161 | 0.651378 |
| HIST1H4J | 147.3376 | -0.6144 | 0.147959 | -4.1525 | 3.29E-05 | 0.000161 | 0.653202 |
| CLDN6 | 10953.71 | -0.79287 | 0.191046 | -4.15018 | 3.32E-05 | 0.000162 | 0.577194 |
| FBXO44 | 659.2402 | -0.79365 | 0.191236 | -4.15011 | 3.32E-05 | 0.000162 | 0.576883 |
| A2ML1 | 127.4131 | -1.62015 | 0.390397 | -4.15002 | 3.32E-05 | 0.000162 | 0.325301 |
| SRRM4 | 106.6669 | -1.76457 | 0.425267 | -4.14933 | 3.33E-05 | 0.000163 | 0.294313 |
| FAM95C | 348.2311 | -0.77159 | 0.185957 | -4.14926 | 3.34E-05 | 0.000163 | 0.585773 |
| IL17RB | 194.4831 | -0.69529 | 0.167586 | -4.14887 | 3.34E-05 | 0.000163 | 0.617584 |
| IL9R | 19.31436 | -2.4625 | 0.593762 | -4.14729 | 3.36E-05 | 0.000164 | 0.181432 |
| GABARAPL | 1728.624 | -0.49849 | 0.120221 | -4.14646 | 3.38E-05 | 0.000165 | 0.707847 |
| TMPRSS3 | 16.8477 | -2.51524 | 0.606939 | -4.14414 | 3.41E-05 | 0.000166 | 0.174919 |
| TRPC4 | 564.8113 | -0.65499 | 0.158118 | -4.14241 | 3.44E-05 | 0.000167 | 0.635079 |
| HSPA12B | 139.507 | -0.7914 | 0.191253 | -4.13794 | 3.50E-05 | 0.00017 | 0.577785 |
| INHBB | 12.99065 | -2.70881 | 0.654649 | -4.1378 | 3.51E-05 | 0.00017 | 0.152957 |
| SLC2A4RG | 925.3066 | -0.58596 | 0.141612 | -4.13776 | 3.51E-05 | 0.00017 | 0.666208 |
| SPNS1 | 1978.443 | -0.54498 | 0.131711 | -4.13774 | 3.51E-05 | 0.00017 | 0.685399 |
| HS3ST4 | 61.08858 | -1.00305 | 0.242495 | -4.13636 | 3.53E-05 | 0.000171 | 0.498946 |
| HLA-G | 25.99234 | -3.17738 | 0.768728 | -4.1333 | 3.58E-05 | 0.000173 | 0.110538 |
| TERT | 323.8709 | -0.96477 | 0.233469 | -4.13231 | 3.59E-05 | 0.000174 | 0.512362 |
| HPCAL4 | 50.50841 | -1.40082 | 0.339073 | -4.13132 | 3.61E-05 | 0.000175 | 0.378714 |
| AL096814. | 20.10691 | -2.37615 | 0.575378 | -4.12971 | 3.63E-05 | 0.000176 | 0.192623 |
| CNIH4 | 2106.202 | -0.51952 | 0.125828 | -4.12884 | 3.65E-05 | 0.000176 | 0.697603 |
| MT-ND4 | 169460.3 | -1.3465 | 0.326541 | -4.12353 | 3.73E-05 | 0.00018 | 0.393245 |
| SPR | 84.00497 | -0.95928 | 0.232806 | -4.12053 | 3.78E-05 | 0.000182 | 0.514312 |
| ACSS2 | 308.6949 | -0.57307 | 0.1391 | -4.1198 | 3.79E-05 | 0.000183 | 0.672187 |
| FAM50B | 30.86435 | -1.76439 | 0.42858 | -4.11682 | 3.84E-05 | 0.000185 | 0.294352 |
| RAB25 | 500.658 | -0.59661 | 0.144963 | -4.1156 | 3.86E-05 | 0.000186 | 0.661306 |
| DGKA | 375.9732 | -0.60942 | 0.148091 | -4.11515 | 3.87E-05 | 0.000186 | 0.655461 |
| MT-ND1 | 52151.71 | -1.26309 | 0.307126 | -4.11263 | 3.91E-05 | 0.000188 | 0.416649 |
| MUTYH | 1064.024 | -0.45935 | 0.111719 | -4.11164 | 3.93E-05 | 0.000189 | 0.727314 |
| TMEM132A | 2291.889 | -0.81579 | 0.198437 | -4.11107 | 3.94E-05 | 0.000189 | 0.568098 |
| VCX | 4.357755 | -5.58176 | 1.358204 | -4.10967 | 3.96E-05 | 0.00019 | 0.02088 |
| KANK3 | 85.28237 | -1.07374 | 0.261328 | -4.10879 | 3.98E-05 | 0.000191 | 0.475085 |
| 4-Sep | 176.3758 | -0.85255 | 0.207746 | -4.10382 | 4.06E-05 | 0.000195 | 0.553804 |
| PTPRU | 1904.299 | -0.56115 | 0.1368 | -4.10197 | 4.10E-05 | 0.000196 | 0.677763 |
| LARP1B | 661.1511 | -0.46145 | 0.112511 | -4.10136 | 4.11E-05 | 0.000197 | 0.726257 |
| MAP3K5 | 74.92416 | -0.97941 | 0.238829 | -4.10089 | 4.12E-05 | 0.000197 | 0.507186 |

| | | | | | | | |
|---|---|---|---|---|---|---|---|
| MOV10 | 1081.412 | -0.66649 | 0.162595 | -4.09909 | 4.15E-05 | 0.000198 | 0.630038 |
| EVPL | 16.48013 | -2.74624 | 0.670083 | -4.09837 | 4.16E-05 | 0.000199 | 0.149038 |
| MTRNR2L4 | 55.56409 | -1.3678 | 0.333744 | -4.09834 | 4.16E-05 | 0.000199 | 0.387482 |
| AC018638 | 139.0283 | -0.91412 | 0.223103 | -4.09732 | 4.18E-05 | 0.0002 | 0.530667 |
| ADRA1D | 30.92482 | -1.48118 | 0.361731 | -4.0947 | 4.23E-05 | 0.000202 | 0.358196 |
| LETM2 | 75.54611 | -1.16333 | 0.284195 | -4.0934 | 4.25E-05 | 0.000203 | 0.446482 |
| SLC35G2 | 133.0772 | -0.80137 | 0.195915 | -4.0904 | 4.31E-05 | 0.000205 | 0.573804 |
| MNT | 602.0166 | -0.43373 | 0.106044 | -4.09011 | 4.31E-05 | 0.000205 | 0.740344 |
| SLCO4C1 | 209.9677 | -0.80834 | 0.197799 | -4.0867 | 4.38E-05 | 0.000208 | 0.571037 |
| GPT | 15.11244 | -2.80181 | 0.685705 | -4.08603 | 4.39E-05 | 0.000209 | 0.143408 |
| FER1L4 | 164.0149 | -1.73191 | 0.424023 | -4.08447 | 4.42E-05 | 0.00021 | 0.301054 |
| HEY2 | 107.1492 | -1.20758 | 0.295702 | -4.08377 | 4.43E-05 | 0.000211 | 0.432995 |
| GFAP | 42.52952 | -1.61757 | 0.396112 | -4.08362 | 4.43E-05 | 0.000211 | 0.325883 |
| KCP | 179.7244 | -0.85077 | 0.208377 | -4.08283 | 4.45E-05 | 0.000211 | 0.554489 |
| CHRNA3 | 142.7596 | -0.85038 | 0.208507 | -4.07841 | 4.53E-05 | 0.000215 | 0.55464 |
| RAB43 | 340.6908 | -0.90569 | 0.222116 | -4.07756 | 4.55E-05 | 0.000216 | 0.533776 |
| METTL7A | 174.4407 | -1.36796 | 0.335601 | -4.07615 | 4.58E-05 | 0.000217 | 0.387439 |
| SLC9A3 | 360.6525 | -1.03672 | 0.254337 | -4.07615 | 4.58E-05 | 0.000217 | 0.487435 |
| PILRA | 61.04689 | -1.21944 | 0.299198 | -4.0757 | 4.59E-05 | 0.000217 | 0.429449 |
| NSUN5 | 1160.242 | -0.42439 | 0.104146 | -4.07495 | 4.60E-05 | 0.000218 | 0.745154 |
| PPP1R13B | 190.7314 | -1.02767 | 0.252245 | -4.0741 | 4.62E-05 | 0.000219 | 0.4905 |
| IGSF21 | 200.6122 | -0.87955 | 0.215901 | -4.07385 | 4.62E-05 | 0.000219 | 0.543537 |
| ZNF860 | 50.94657 | -1.37456 | 0.337718 | -4.07015 | 4.70E-05 | 0.000222 | 0.385669 |
| SMPD5 | 34.4363 | -1.5667 | 0.384936 | -4.07002 | 4.70E-05 | 0.000222 | 0.33758 |
| BICDL1 | 411.7237 | -0.64517 | 0.158556 | -4.06902 | 4.72E-05 | 0.000223 | 0.639418 |
| IKBKG | 388.0983 | -0.60299 | 0.1482 | -4.06876 | 4.73E-05 | 0.000223 | 0.658388 |
| AL627309. | 177.3911 | -1.54145 | 0.37899 | -4.06726 | 4.76E-05 | 0.000224 | 0.34354 |
| TM7SF2 | 947.9225 | -0.53536 | 0.131634 | -4.06704 | 4.76E-05 | 0.000225 | 0.689986 |
| KLK1 | 18.97299 | -2.28988 | 0.563199 | -4.06584 | 4.79E-05 | 0.000226 | 0.204493 |
| U2AF1 | 549.2629 | -0.76276 | 0.187633 | -4.06518 | 4.80E-05 | 0.000226 | 0.589367 |
| JAG2 | 415.5792 | -1.04384 | 0.256791 | -4.06496 | 4.80E-05 | 0.000226 | 0.485034 |
| WASH3P | 686.8443 | -0.5424 | 0.133509 | -4.0626 | 4.85E-05 | 0.000228 | 0.68663 |
| QPCTL | 519.1316 | -0.87032 | 0.214277 | -4.06168 | 4.87E-05 | 0.000229 | 0.547024 |
| MOXD1 | 243.0417 | -0.67791 | 0.167005 | -4.05924 | 4.92E-05 | 0.000231 | 0.625069 |
| CX3CL1 | 96.50155 | -0.84588 | 0.208586 | -4.05529 | 5.01E-05 | 0.000235 | 0.556373 |
| MATN2 | 430.8117 | -0.55351 | 0.136559 | -4.05324 | 5.05E-05 | 0.000237 | 0.681362 |
| RIPOR1 | 563.0174 | -0.46597 | 0.114978 | -4.05271 | 5.06E-05 | 0.000237 | 0.723983 |
| 2-Mar | 183.4897 | -0.86227 | 0.212775 | -4.05251 | 5.07E-05 | 0.000238 | 0.550085 |
| JPH2 | 54.58229 | -1.04586 | 0.258128 | -4.05172 | 5.08E-05 | 0.000238 | 0.484356 |
| DNAJC4 | 330.0298 | -0.68682 | 0.169576 | -4.05021 | 5.12E-05 | 0.00024 | 0.621223 |
| F11R | 5540.917 | -0.51199 | 0.126419 | -4.04993 | 5.12E-05 | 0.00024 | 0.701255 |
| KMT5C | 867.626 | -0.78981 | 0.195031 | -4.04964 | 5.13E-05 | 0.00024 | 0.578422 |
| ST7 | 318.5324 | -0.46338 | 0.114425 | -4.04966 | 5.13E-05 | 0.00024 | 0.725283 |
| CRTC2 | 535.3704 | -0.78796 | 0.194801 | -4.04497 | 5.23E-05 | 0.000244 | 0.579161 |

| | | | | | | | |
|---|---|---|---|---|---|---|---|
| ADCK5 | 122.5709 | -0.70941 | 0.175464 | -4.04302 | 5.28E-05 | 0.000246 | 0.611572 |
| GIGYF1 | 3913.719 | -0.67516 | 0.167034 | -4.04203 | 5.30E-05 | 0.000247 | 0.626263 |
| SLC22A3 | 35.98384 | -1.73405 | 0.429042 | -4.04169 | 5.31E-05 | 0.000248 | 0.300606 |
| ACOT11 | 200.8192 | -0.7459 | 0.184722 | -4.03796 | 5.39E-05 | 0.000251 | 0.596296 |
| RASSF4 | 379.5285 | -0.87935 | 0.217898 | -4.03561 | 5.45E-05 | 0.000253 | 0.543612 |
| LAMB2 | 2835.722 | -1.46514 | 0.363363 | -4.03216 | 5.53E-05 | 0.000257 | 0.3622 |
| ULK1 | 2111.725 | -0.47793 | 0.11855 | -4.03149 | 5.54E-05 | 0.000258 | 0.718005 |
| OPRD1 | 120.8688 | -0.70037 | 0.173849 | -4.02861 | 5.61E-05 | 0.000261 | 0.615414 |
| IQSEC3 | 70.76065 | -2.60306 | 0.646631 | -4.02557 | 5.68E-05 | 0.000264 | 0.164589 |
| SFXN3 | 34.38353 | -1.94855 | 0.48409 | -4.02518 | 5.69E-05 | 0.000264 | 0.259077 |
| SLC25A18 | 40.09375 | -1.33425 | 0.331692 | -4.02257 | 5.76E-05 | 0.000267 | 0.396597 |
| UPP1 | 1292.506 | -0.66499 | 0.165319 | -4.02244 | 5.76E-05 | 0.000267 | 0.630695 |
| NUCB1 | 3165.935 | -0.6256 | 0.155634 | -4.01969 | 5.83E-05 | 0.00027 | 0.648149 |
| PDIA2 | 25.90003 | -2.36435 | 0.588244 | -4.01935 | 5.84E-05 | 0.00027 | 0.194204 |
| SKIL | 1127.556 | -0.7382 | 0.183703 | -4.01845 | 5.86E-05 | 0.000271 | 0.599486 |
| THSD7B | 27.91548 | -2.6131 | 0.650494 | -4.0171 | 5.89E-05 | 0.000272 | 0.163448 |
| CTIF | 572.3146 | -0.83342 | 0.207496 | -4.01657 | 5.91E-05 | 0.000273 | 0.561196 |
| ZBTB7B | 930.8665 | -0.76038 | 0.189485 | -4.01288 | 6.00E-05 | 0.000277 | 0.590341 |
| NPIPA3 | 101.6866 | -1.20635 | 0.300731 | -4.0114 | 6.04E-05 | 0.000279 | 0.433363 |
| TIMP1 | 775.7126 | -0.54039 | 0.134748 | -4.01034 | 6.06E-05 | 0.00028 | 0.687587 |
| RABEP2 | 253.3126 | -0.56234 | 0.140227 | -4.01022 | 6.07E-05 | 0.00028 | 0.677203 |
| TMEM51 | 325.8126 | -0.67103 | 0.167344 | -4.00988 | 6.07E-05 | 0.00028 | 0.628059 |
| NUDT22 | 360.6733 | -0.7604 | 0.189635 | -4.00979 | 6.08E-05 | 0.00028 | 0.590334 |
| GALNT17 | 1522.274 | -0.49708 | 0.123974 | -4.00952 | 6.08E-05 | 0.00028 | 0.708542 |
| HSF4 | 14.95653 | -3.55329 | 0.886466 | -4.00838 | 6.11E-05 | 0.000282 | 0.085183 |
| PPP1R36 | 16.159 | -2.53681 | 0.632972 | -4.00777 | 6.13E-05 | 0.000282 | 0.172324 |
| PTPRN2 | 23.37364 | -1.69455 | 0.422837 | -4.00757 | 6.13E-05 | 0.000282 | 0.308951 |
| HM13 | 1997.734 | -0.4332 | 0.108141 | -4.00588 | 6.18E-05 | 0.000284 | 0.740617 |
| BBC3 | 552.4801 | -0.52829 | 0.131882 | -4.0058 | 6.18E-05 | 0.000284 | 0.693374 |
| CLU | 13469.01 | -0.66241 | 0.165365 | -4.00576 | 6.18E-05 | 0.000284 | 0.631821 |
| GABBR1 | 1383.64 | -0.70369 | 0.175725 | -4.00451 | 6.21E-05 | 0.000285 | 0.613999 |
| NDOR1 | 605.9945 | -0.44867 | 0.112045 | -4.00437 | 6.22E-05 | 0.000285 | 0.732718 |
| PTGER4 | 19.93559 | -2.5072 | 0.626363 | -4.0028 | 6.26E-05 | 0.000287 | 0.175896 |
| SEMA6B | 2665.564 | -0.42363 | 0.105834 | -4.00278 | 6.26E-05 | 0.000287 | 0.745547 |
| ANAPC7 | 1591.953 | -0.45304 | 0.113207 | -4.00184 | 6.29E-05 | 0.000288 | 0.730503 |
| ARAP1 | 2008.762 | -0.57474 | 0.143811 | -3.9965 | 6.43E-05 | 0.000294 | 0.671406 |
| FGF1 | 28.08542 | -1.71615 | 0.42941 | -3.99653 | 6.43E-05 | 0.000294 | 0.30436 |
| TMEM158 | 376.1015 | -1.01271 | 0.253544 | -3.99421 | 6.49E-05 | 0.000297 | 0.495616 |
| MLKL | 39.51077 | -1.33051 | 0.333213 | -3.99299 | 6.52E-05 | 0.000298 | 0.397626 |
| OSBP2 | 187.9082 | -0.5471 | 0.137037 | -3.99236 | 6.54E-05 | 0.000299 | 0.684394 |
| CCDC85B | 564.6584 | -0.46697 | 0.116997 | -3.99132 | 6.57E-05 | 0.0003 | 0.723482 |
| RNF212 | 19.13531 | -2.11989 | 0.531145 | -3.99116 | 6.57E-05 | 0.0003 | 0.230065 |
| DST | 4589.698 | -0.74016 | 0.185472 | -3.99067 | 6.59E-05 | 0.0003 | 0.598673 |
| ITM2B | 3471.443 | -0.52611 | 0.131836 | -3.99066 | 6.59E-05 | 0.0003 | 0.694424 |

| | | | | | | | |
|---|---|---|---|---|---|---|---|
| MYO7A | 98.15464 | -1.18561 | 0.297172 | -3.98965 | 6.62E-05 | 0.000302 | 0.439638 |
| DIRAS1 | 677.3857 | -0.4191 | 0.10508 | -3.98842 | 6.65E-05 | 0.000303 | 0.74789 |
| MKX | 84.07499 | -1.36668 | 0.342837 | -3.98637 | 6.71E-05 | 0.000305 | 0.387783 |
| MYO18A | 1674.245 | -0.53132 | 0.133293 | -3.98612 | 6.72E-05 | 0.000306 | 0.691921 |
| SEPHS1 | 10415.61 | -0.55314 | 0.138911 | -3.98195 | 6.84E-05 | 0.000311 | 0.681536 |
| COL5A3 | 81.34173 | -1.10764 | 0.27825 | -3.98074 | 6.87E-05 | 0.000312 | 0.464052 |
| PQLC2L | 5.364936 | -5.01329 | 1.259754 | -3.97958 | 6.90E-05 | 0.000313 | 0.030963 |
| DRAM1 | 244.8137 | -0.62702 | 0.157615 | -3.97816 | 6.95E-05 | 0.000315 | 0.647514 |
| VEGFB | 375.212 | -0.68398 | 0.171949 | -3.9778 | 6.96E-05 | 0.000315 | 0.622446 |
| CACNG8 | 259.9792 | -0.82614 | 0.207806 | -3.97554 | 7.02E-05 | 0.000318 | 0.564037 |
| ZNF593 | 270.0327 | -0.56347 | 0.141734 | -3.97556 | 7.02E-05 | 0.000318 | 0.676672 |
| FBF1 | 412.1853 | -0.58582 | 0.147468 | -3.97254 | 7.11E-05 | 0.000322 | 0.66627 |
| CELSR3 | 2000.536 | -0.55381 | 0.139479 | -3.97054 | 7.17E-05 | 0.000324 | 0.681221 |
| ADCY9 | 347.3369 | -0.88346 | 0.222525 | -3.97017 | 7.18E-05 | 0.000324 | 0.542064 |
| NKX1-2 | 534.1872 | -0.96282 | 0.242554 | -3.96952 | 7.20E-05 | 0.000325 | 0.513052 |
| RTBDN | 40.62997 | -1.56642 | 0.394763 | -3.968 | 7.25E-05 | 0.000327 | 0.337645 |
| C1QTNF12 | 60.57417 | -1.81588 | 0.457797 | -3.96655 | 7.29E-05 | 0.000329 | 0.284031 |
| FAR2P1 | 81.48716 | -1.53743 | 0.387816 | -3.96433 | 7.36E-05 | 0.000332 | 0.344498 |
| CALR | 16155.11 | -0.47946 | 0.121145 | -3.95775 | 7.57E-05 | 0.000341 | 0.717245 |
| PCDH8 | 81.77105 | -1.54656 | 0.391042 | -3.95496 | 7.65E-05 | 0.000344 | 0.342326 |
| EMC10 | 3126.906 | -0.42707 | 0.108007 | -3.95409 | 7.68E-05 | 0.000345 | 0.743772 |
| TTC9 | 630.4065 | -0.58721 | 0.148525 | -3.9536 | 7.70E-05 | 0.000346 | 0.66563 |
| PRORSD1F | 54.34236 | -1.31572 | 0.332837 | -3.95306 | 7.72E-05 | 0.000347 | 0.401724 |
| GOLGA6A | 41.8511 | -2.15249 | 0.544666 | -3.95195 | 7.75E-05 | 0.000348 | 0.224924 |
| C1R | 313.7636 | -0.83482 | 0.211466 | -3.94777 | 7.89E-05 | 0.000354 | 0.560654 |
| CDC25B | 842.5491 | -0.43884 | 0.111161 | -3.94776 | 7.89E-05 | 0.000354 | 0.73773 |
| DISP2 | 111.592 | -2.04161 | 0.517504 | -3.94512 | 7.98E-05 | 0.000358 | 0.242892 |
| AGRN | 7453.575 | -0.75213 | 0.190721 | -3.94363 | 8.03E-05 | 0.000359 | 0.593725 |
| SLC37A2 | 148.4975 | -0.90579 | 0.229887 | -3.94014 | 8.14E-05 | 0.000364 | 0.533741 |
| DUSP5 | 473.6781 | -0.57841 | 0.146832 | -3.93928 | 8.17E-05 | 0.000365 | 0.6697 |
| TMEM38A | 197.2377 | -0.75576 | 0.191908 | -3.93813 | 8.21E-05 | 0.000367 | 0.592235 |
| PPP2R2B | 798.8787 | -0.45663 | 0.116166 | -3.93087 | 8.46E-05 | 0.000377 | 0.728684 |
| VENTX | 113.4622 | -0.94188 | 0.239608 | -3.93091 | 8.46E-05 | 0.000377 | 0.520555 |
| AL590426. | 5.417238 | -5.13815 | 1.307615 | -3.92941 | 8.52E-05 | 0.000379 | 0.028396 |
| EBF4 | 123.0646 | -1.16881 | 0.297455 | -3.92939 | 8.52E-05 | 0.000379 | 0.444787 |
| DLL4 | 17.30821 | -2.14722 | 0.546465 | -3.92929 | 8.52E-05 | 0.000379 | 0.225747 |
| AC074141 | 39.27191 | -1.32806 | 0.338003 | -3.92914 | 8.53E-05 | 0.000379 | 0.398303 |
| RNF123 | 1017.527 | -0.5383 | 0.137201 | -3.92342 | 8.73E-05 | 0.000388 | 0.688582 |
| MANEAL | 189.7637 | -0.788 | 0.200878 | -3.92279 | 8.75E-05 | 0.000389 | 0.579145 |
| HLA-E | 1094.233 | -0.67071 | 0.171072 | -3.92065 | 8.83E-05 | 0.000392 | 0.628196 |
| MYDGF | 835.3713 | -0.54032 | 0.137817 | -3.92058 | 8.83E-05 | 0.000392 | 0.687617 |
| ALG12 | 670.2402 | -0.67817 | 0.173087 | -3.91809 | 8.93E-05 | 0.000396 | 0.624957 |
| NPHS1 | 38.60558 | -1.43014 | 0.365074 | -3.91741 | 8.95E-05 | 0.000397 | 0.371094 |
| DHRS7B | 441.2335 | -0.57328 | 0.146346 | -3.91728 | 8.96E-05 | 0.000397 | 0.672088 |

| | | | | | | | |
|---|---|---|---|---|---|---|---|
| CD83 | 155.5972 | -0.62004 | 0.158351 | -3.91559 | 9.02E-05 | 0.000399 | 0.650653 |
| PUS1 | 982.4468 | -0.47519 | 0.121383 | -3.91477 | 9.05E-05 | 0.000401 | 0.719373 |
| FAM124B | 15.51824 | -2.16259 | 0.552464 | -3.91445 | 9.06E-05 | 0.000401 | 0.223355 |
| RAB12 | 748.0356 | -0.43193 | 0.110342 | -3.91443 | 9.06E-05 | 0.000401 | 0.741272 |
| KLHL33 | 30.28451 | -1.39897 | 0.357527 | -3.91292 | 9.12E-05 | 0.000403 | 0.379199 |
| AL671277. | 11.99043 | -3.65343 | 0.93402 | -3.91152 | 9.17E-05 | 0.000405 | 0.079471 |
| ADGRB1 | 399.0639 | -0.94693 | 0.242179 | -3.91005 | 9.23E-05 | 0.000408 | 0.518735 |
| MDGA2 | 32.16939 | -1.88147 | 0.481355 | -3.9087 | 9.28E-05 | 0.00041 | 0.271407 |
| GAS2L1 | 118.3376 | -0.73271 | 0.187523 | -3.90733 | 9.33E-05 | 0.000412 | 0.601771 |
| SARS2 | 865.8561 | -0.59908 | 0.153342 | -3.90682 | 9.35E-05 | 0.000413 | 0.660174 |
| AHNAK | 1254.169 | -0.97376 | 0.249308 | -3.90584 | 9.39E-05 | 0.000414 | 0.509179 |
| LRG1 | 17.0749 | -1.85304 | 0.475224 | -3.89929 | 9.65E-05 | 0.000425 | 0.276809 |
| RPLP0P2 | 20.57666 | -1.69815 | 0.435618 | -3.89824 | 9.69E-05 | 0.000427 | 0.308182 |
| CD72 | 58.74233 | -1.12275 | 0.288037 | -3.89792 | 9.70E-05 | 0.000427 | 0.459219 |
| SLCO2A1 | 122.116 | -1.77175 | 0.45463 | -3.89711 | 9.73E-05 | 0.000428 | 0.292854 |
| JAK3 | 131.0724 | -0.75919 | 0.194824 | -3.89678 | 9.75E-05 | 0.000429 | 0.59083 |
| C3orf52 | 247.6548 | -0.47655 | 0.122435 | -3.8923 | 9.93E-05 | 0.000436 | 0.718693 |
| PTGFRN | 1037.26 | -0.71941 | 0.184849 | -3.89189 | 9.95E-05 | 0.000437 | 0.607345 |
| INO80D | 835.4108 | -0.51083 | 0.131313 | -3.89014 | 0.0001 | 0.00044 | 0.701821 |
| ETV4 | 1896.605 | -0.77827 | 0.200163 | -3.88818 | 0.000101 | 0.000443 | 0.583065 |
| GSAP | 102.6309 | -1.07318 | 0.276102 | -3.8869 | 0.000102 | 0.000445 | 0.475269 |
| NPIPA1 | 1898.149 | -0.72076 | 0.185479 | -3.88595 | 0.000102 | 0.000447 | 0.606777 |
| FABP3 | 177.3026 | -0.66693 | 0.171706 | -3.88411 | 0.000103 | 0.00045 | 0.629847 |
| PCNX3 | 1655.197 | -0.57546 | 0.148187 | -3.88332 | 0.000103 | 0.000451 | 0.671073 |
| CAPN11 | 74.99877 | -1.05473 | 0.271717 | -3.88172 | 0.000104 | 0.000454 | 0.481387 |
| SELENOM | 197.5779 | -0.69772 | 0.179761 | -3.88139 | 0.000104 | 0.000454 | 0.616544 |
| OXCT2 | 41.96213 | -1.3577 | 0.350195 | -3.87697 | 0.000106 | 0.000462 | 0.390205 |
| CASTOR1 | 155.4216 | -0.79159 | 0.204185 | -3.87681 | 0.000106 | 0.000462 | 0.577709 |
| MEF2D | 547.9378 | -0.66368 | 0.171235 | -3.87587 | 0.000106 | 0.000464 | 0.631264 |
| ACP7 | 10.52538 | -2.74076 | 0.70743 | -3.87425 | 0.000107 | 0.000466 | 0.149606 |
| PRR16 | 50.81972 | -1.71505 | 0.442702 | -3.87404 | 0.000107 | 0.000467 | 0.304593 |
| CA7 | 24.94015 | -1.6579 | 0.427975 | -3.87382 | 0.000107 | 0.000467 | 0.316901 |
| KCNE3 | 51.41402 | -1.51249 | 0.390631 | -3.87191 | 0.000108 | 0.00047 | 0.350507 |
| C3orf80 | 27.4541 | -1.94739 | 0.503022 | -3.87139 | 0.000108 | 0.000471 | 0.259284 |
| SLC25A19 | 227.2524 | -0.55954 | 0.144555 | -3.87077 | 0.000108 | 0.000472 | 0.67852 |
| SELENOT | 1400.721 | -0.46906 | 0.121233 | -3.86911 | 0.000109 | 0.000475 | 0.722433 |
| TYK2 | 590.945 | -0.92181 | 0.23826 | -3.86892 | 0.000109 | 0.000475 | 0.527846 |
| SH2B1 | 1051.532 | -0.67024 | 0.173483 | -3.86341 | 0.000112 | 0.000485 | 0.628404 |
| CD109 | 315.4716 | -0.4548 | 0.117735 | -3.86287 | 0.000112 | 0.000486 | 0.729614 |
| MFAP4 | 193.9254 | -0.75742 | 0.196102 | -3.86235 | 0.000112 | 0.000487 | 0.591555 |
| PLXNA3 | 895.9859 | -0.76077 | 0.197065 | -3.86051 | 0.000113 | 0.00049 | 0.59018 |
| NRROS | 94.62893 | -0.86908 | 0.225152 | -3.85996 | 0.000113 | 0.000491 | 0.547496 |
| TSPEAR | 52.04111 | -1.31365 | 0.340325 | -3.85998 | 0.000113 | 0.000491 | 0.402302 |
| RHOXF1P1 | 6.783685 | -3.59637 | 0.931906 | -3.85915 | 0.000114 | 0.000492 | 0.082677 |

| | | | | | | | |
|---|---|---|---|---|---|---|---|
| ALPP | 15.43378 | -2.62562 | 0.680446 | -3.85867 | 0.000114 | 0.000493 | 0.162035 |
| KLF16 | 731.3802 | -0.42989 | 0.111411 | -3.85861 | 0.000114 | 0.000493 | 0.742317 |
| METRNL | 243.4356 | -0.62897 | 0.163007 | -3.85854 | 0.000114 | 0.000493 | 0.646639 |
| PGPEP1 | 894.2733 | -0.75198 | 0.194893 | -3.85845 | 0.000114 | 0.000493 | 0.593786 |
| NAAA | 38.90423 | -1.6886 | 0.437756 | -3.85739 | 0.000115 | 0.000495 | 0.310228 |
| AVPI1 | 176.341 | -0.58964 | 0.152884 | -3.85676 | 0.000115 | 0.000496 | 0.66451 |
| LRP5 | 1504.408 | -0.48859 | 0.126681 | -3.85682 | 0.000115 | 0.000496 | 0.712724 |
| NPR2 | 405.0014 | -0.74537 | 0.193278 | -3.85645 | 0.000115 | 0.000496 | 0.596516 |
| ABCG1 | 69.24314 | -1.12895 | 0.29276 | -3.85624 | 0.000115 | 0.000496 | 0.457248 |
| TNKS1BP1 | 4962.131 | -0.51067 | 0.13246 | -3.85531 | 0.000116 | 0.000498 | 0.701894 |
| ART5 | 148.9799 | -0.84684 | 0.219857 | -3.85177 | 0.000117 | 0.000505 | 0.556001 |
| TEX14 | 20.92266 | -1.58025 | 0.410348 | -3.85101 | 0.000118 | 0.000506 | 0.334424 |
| RHOV | 75.55233 | -0.87499 | 0.22725 | -3.85034 | 0.000118 | 0.000507 | 0.545259 |
| AC118754 | 28.87115 | -1.55391 | 0.403588 | -3.85025 | 0.000118 | 0.000508 | 0.340585 |
| CD63 | 6222.286 | -0.59889 | 0.155578 | -3.84944 | 0.000118 | 0.000509 | 0.660263 |
| FBLN2 | 596.2319 | -0.46625 | 0.121131 | -3.84917 | 0.000119 | 0.000509 | 0.723841 |
| HTR1A | 108.2493 | -1.06687 | 0.277285 | -3.84756 | 0.000119 | 0.000512 | 0.477354 |
| DBN1 | 10134.14 | -0.65295 | 0.169767 | -3.84617 | 0.00012 | 0.000515 | 0.635977 |
| MROH1 | 729.1078 | -0.81513 | 0.211997 | -3.84503 | 0.000121 | 0.000517 | 0.568356 |
| SYT12 | 111.5611 | -1.0693 | 0.278148 | -3.84435 | 0.000121 | 0.000518 | 0.476551 |
| CTSV | 2287.647 | -0.59811 | 0.155585 | -3.84429 | 0.000121 | 0.000518 | 0.660617 |
| KMT2B | 2021.64 | -0.64067 | 0.166693 | -3.84343 | 0.000121 | 0.000519 | 0.641414 |
| KLHL34 | 9.223803 | -3.03597 | 0.789986 | -3.84307 | 0.000122 | 0.00052 | 0.121922 |
| DNHD1 | 398.0509 | -0.94501 | 0.245929 | -3.84262 | 0.000122 | 0.000521 | 0.519426 |
| CACNG6 | 43.50968 | -1.29025 | 0.335859 | -3.84163 | 0.000122 | 0.000522 | 0.408882 |
| GTPBP3 | 866.0779 | -0.53123 | 0.138338 | -3.84005 | 0.000123 | 0.000525 | 0.691966 |
| SSTR1 | 24.21998 | -1.55163 | 0.404127 | -3.83948 | 0.000123 | 0.000526 | 0.341124 |
| PRRG3 | 187.1384 | -1.49999 | 0.391044 | -3.83587 | 0.000125 | 0.000533 | 0.353556 |
| MT1F | 53.01561 | -1.21432 | 0.316716 | -3.83411 | 0.000126 | 0.000537 | 0.430975 |
| PDE4B | 75.92372 | -0.91679 | 0.239223 | -3.83236 | 0.000127 | 0.00054 | 0.529687 |
| MROH6 | 73.84641 | -2.49907 | 0.652172 | -3.83192 | 0.000127 | 0.000541 | 0.176891 |
| RND1 | 383.13 | -0.46024 | 0.120121 | -3.83153 | 0.000127 | 0.000541 | 0.726863 |
| TSPAN9 | 815.8442 | -0.59629 | 0.15567 | -3.8305 | 0.000128 | 0.000543 | 0.661451 |
| DGKQ | 482.9479 | -0.66726 | 0.174364 | -3.82683 | 0.00013 | 0.000551 | 0.629701 |
| NMU | 154.0925 | -0.89506 | 0.234037 | -3.82446 | 0.000131 | 0.000556 | 0.537723 |
| JADE2 | 1035.81 | -0.62478 | 0.16339 | -3.82389 | 0.000131 | 0.000557 | 0.648517 |
| PI4KAP1 | 675.3436 | -0.69661 | 0.182187 | -3.82361 | 0.000132 | 0.000558 | 0.61702 |
| RNF112 | 77.18338 | -1.02755 | 0.268891 | -3.82143 | 0.000133 | 0.000562 | 0.490544 |
| DYSF | 185.7522 | -1.0487 | 0.274479 | -3.82071 | 0.000133 | 0.000564 | 0.483402 |
| FSD1 | 1625.129 | -0.45392 | 0.118827 | -3.82001 | 0.000133 | 0.000565 | 0.730056 |
| SH2D5 | 246.9876 | -0.76876 | 0.201293 | -3.81908 | 0.000134 | 0.000567 | 0.586923 |
| ERP44 | 862.2016 | -0.41882 | 0.109676 | -3.81869 | 0.000134 | 0.000568 | 0.748036 |
| TMEM102 | 191.9001 | -0.72963 | 0.191166 | -3.81677 | 0.000135 | 0.000572 | 0.603057 |
| CLUHP3 | 369.1762 | -0.44901 | 0.117658 | -3.81622 | 0.000136 | 0.000573 | 0.732547 |

| | | | | | | | |
|---|---|---|---|---|---|---|---|
| LYPD6B | 52.06371 | -1.41164 | 0.370013 | -3.8151 | 0.000136 | 0.000576 | 0.375884 |
| FP565260. | 45.54537 | -1.10056 | 0.288576 | -3.81374 | 0.000137 | 0.000579 | 0.466337 |
| CARD6 | 23.78565 | -1.43468 | 0.376231 | -3.81329 | 0.000137 | 0.000579 | 0.36993 |
| LAMA3 | 46.33314 | -2.68405 | 0.70405 | -3.8123 | 0.000138 | 0.000581 | 0.155604 |
| FOXA3 | 508.7687 | -0.51229 | 0.134406 | -3.81151 | 0.000138 | 0.000583 | 0.701108 |
| TMEM190 | 7.655328 | -2.95262 | 0.774664 | -3.81149 | 0.000138 | 0.000583 | 0.129173 |
| LIMA1 | 1295.35 | -0.43579 | 0.114352 | -3.81094 | 0.000138 | 0.000584 | 0.73929 |
| AC010422 | 25.34359 | -3.21131 | 0.843035 | -3.80922 | 0.000139 | 0.000588 | 0.107969 |
| LGALS3BP | 1919.858 | -0.59604 | 0.156499 | -3.80857 | 0.00014 | 0.000589 | 0.661569 |
| ABHD14B | 168.4267 | -0.76469 | 0.200895 | -3.80641 | 0.000141 | 0.000594 | 0.58858 |
| ELOVL4 | 440.7536 | -0.43702 | 0.11492 | -3.80276 | 0.000143 | 0.000602 | 0.738661 |
| NANOGP1 | 24.71565 | -2.26717 | 0.596771 | -3.79906 | 0.000145 | 0.00061 | 0.207737 |
| SPHK2 | 861.4184 | -0.44372 | 0.1168 | -3.79899 | 0.000145 | 0.00061 | 0.735236 |
| ARHGDIG | 34.22224 | -1.45444 | 0.383147 | -3.79605 | 0.000147 | 0.000617 | 0.364896 |
| PDGFRL | 103.4977 | -0.93071 | 0.24519 | -3.79586 | 0.000147 | 0.000617 | 0.5246 |
| EMX1 | 44.05488 | -1.55113 | 0.408775 | -3.79458 | 0.000148 | 0.00062 | 0.341243 |
| SLC7A14 | 42.43607 | -1.75369 | 0.462208 | -3.79416 | 0.000148 | 0.000621 | 0.296542 |
| GSDMB | 252.6338 | -0.98184 | 0.258812 | -3.79364 | 0.000148 | 0.000622 | 0.506334 |
| KLK14 | 16.4445 | -2.5133 | 0.662728 | -3.79236 | 0.000149 | 0.000625 | 0.175154 |
| RSAD1 | 663.4514 | -0.41817 | 0.110305 | -3.79107 | 0.00015 | 0.000628 | 0.748371 |
| CD55 | 578.5272 | -0.4846 | 0.127863 | -3.78998 | 0.000151 | 0.00063 | 0.714695 |
| SLC52A1 | 25.49888 | -1.62807 | 0.429747 | -3.78843 | 0.000152 | 0.000634 | 0.323521 |
| ENPP7P1 | 9.436415 | -3.56789 | 0.942034 | -3.78743 | 0.000152 | 0.000636 | 0.084325 |
| ZBTB7A | 471.6499 | -0.46738 | 0.123408 | -3.7873 | 0.000152 | 0.000636 | 0.723275 |
| GRIN2B | 52.75137 | -1.48792 | 0.393027 | -3.7858 | 0.000153 | 0.00064 | 0.356526 |
| SLC45A1 | 55.89904 | -0.99392 | 0.26267 | -3.78391 | 0.000154 | 0.000644 | 0.502112 |
| PNPLA6 | 2178.924 | -0.71673 | 0.189568 | -3.78088 | 0.000156 | 0.000651 | 0.608474 |
| RINL | 202.7607 | -0.80767 | 0.213673 | -3.77996 | 0.000157 | 0.000654 | 0.571302 |
| XKR6 | 168.5156 | -0.80076 | 0.211872 | -3.77943 | 0.000157 | 0.000655 | 0.574048 |
| PNMA5 | 23.8391 | -1.96613 | 0.52069 | -3.776 | 0.000159 | 0.000663 | 0.255939 |
| ASPG | 36.73857 | -1.80358 | 0.477737 | -3.77525 | 0.00016 | 0.000665 | 0.286464 |
| ARHGAP23 | 1194.857 | -0.59155 | 0.156707 | -3.77486 | 0.00016 | 0.000666 | 0.66363 |
| PLBD2 | 1931.656 | -0.41711 | 0.110499 | -3.77476 | 0.00016 | 0.000666 | 0.748926 |
| ARFGAP1 | 2481.548 | -0.71127 | 0.188437 | -3.77457 | 0.00016 | 0.000666 | 0.610783 |
| ELAVL2 | 300.1815 | -0.63932 | 0.169374 | -3.77458 | 0.00016 | 0.000666 | 0.642017 |
| SIRT7 | 332.8814 | -0.74683 | 0.197871 | -3.77432 | 0.00016 | 0.000667 | 0.595913 |
| PDE8A | 257.6605 | -0.9011 | 0.238776 | -3.77385 | 0.000161 | 0.000668 | 0.535478 |
| KDM4C | 397.0742 | -0.61815 | 0.163845 | -3.77277 | 0.000161 | 0.00067 | 0.651507 |
| OXLD1 | 475.3958 | -0.44408 | 0.117718 | -3.77239 | 0.000162 | 0.000671 | 0.735054 |
| ORAI3 | 295.384 | -0.49471 | 0.131144 | -3.77227 | 0.000162 | 0.000671 | 0.709703 |
| ACCS | 197.2847 | -0.58056 | 0.153924 | -3.77173 | 0.000162 | 0.000672 | 0.668705 |
| SLC38A3 | 84.48669 | -1.05535 | 0.279872 | -3.77084 | 0.000163 | 0.000675 | 0.48118 |
| ZNF442 | 36.5533 | -2.37777 | 0.630672 | -3.77022 | 0.000163 | 0.000676 | 0.192407 |
| ADCY3 | 1015.947 | -0.57071 | 0.151449 | -3.76835 | 0.000164 | 0.000681 | 0.673284 |

| | | | | | | | |
|---|---|---|---|---|---|---|---|
| CNTNAP1 | 1075.573 | -0.51296 | 0.13614 | -3.76791 | 0.000165 | 0.000682 | 0.700782 |
| CHD5 | 129.6749 | -1.49711 | 0.397344 | -3.7678 | 0.000165 | 0.000682 | 0.354262 |
| ABCG4 | 58.81372 | -1.0553 | 0.280091 | -3.76771 | 0.000165 | 0.000682 | 0.481197 |
| AC087651 | 19.4415 | -1.66245 | 0.441293 | -3.76722 | 0.000165 | 0.000683 | 0.315903 |
| MOCOS | 658.6521 | -0.83569 | 0.221909 | -3.76592 | 0.000166 | 0.000686 | 0.560314 |
| SCT | 14.16622 | -2.99534 | 0.795635 | -3.76471 | 0.000167 | 0.000689 | 0.125405 |
| CLDN19 | 320.421 | -0.59792 | 0.158872 | -3.76356 | 0.000168 | 0.000692 | 0.660704 |
| ZNF749 | 330.4984 | -0.54425 | 0.144853 | -3.75725 | 0.000172 | 0.000708 | 0.685748 |
| PDCD1 | 34.35876 | -2.27797 | 0.606307 | -3.75713 | 0.000172 | 0.000708 | 0.206187 |
| OR1A1 | 75.94913 | -0.9467 | 0.252055 | -3.7559 | 0.000173 | 0.000712 | 0.518819 |
| TNK2 | 902.7173 | -0.66701 | 0.177646 | -3.7547 | 0.000174 | 0.000715 | 0.629811 |
| MAGED4B | 4076.176 | -0.73489 | 0.195797 | -3.75333 | 0.000175 | 0.000718 | 0.600864 |
| PTCHD1 | 168.6802 | -0.85024 | 0.226691 | -3.75068 | 0.000176 | 0.000725 | 0.554691 |
| PAQR4 | 988.7385 | -0.44351 | 0.118264 | -3.7502 | 0.000177 | 0.000726 | 0.735342 |
| OTOG | 90.76663 | -3.63829 | 0.970405 | -3.74925 | 0.000177 | 0.000729 | 0.080309 |
| RCHY1 | 889.1632 | -0.57348 | 0.15299 | -3.74846 | 0.000178 | 0.000731 | 0.671994 |
| PLPP3 | 1085.677 | -0.49577 | 0.132322 | -3.7467 | 0.000179 | 0.000736 | 0.709183 |
| MRI1 | 532.0682 | -0.66313 | 0.177126 | -3.74383 | 0.000181 | 0.000743 | 0.631507 |
| FAM3A | 465.6881 | -0.57379 | 0.153305 | -3.74279 | 0.000182 | 0.000746 | 0.671851 |
| SH3BP5 | 345.738 | -0.53471 | 0.142927 | -3.74114 | 0.000183 | 0.00075 | 0.690297 |
| SCARF1 | 39.85329 | -2.22094 | 0.594505 | -3.73578 | 0.000187 | 0.000765 | 0.214502 |
| LRRFIP1 | 1285.45 | -0.61102 | 0.163733 | -3.73182 | 0.00019 | 0.000777 | 0.654732 |
| PCDH7 | 286.9375 | -0.87458 | 0.23438 | -3.73146 | 0.00019 | 0.000778 | 0.545413 |
| FES | 113.3334 | -1.08374 | 0.290491 | -3.73071 | 0.000191 | 0.00078 | 0.471805 |
| AARSD1 | 709.5768 | -0.42897 | 0.115041 | -3.72888 | 0.000192 | 0.000785 | 0.74279 |
| EMILIN2 | 221.5677 | -0.55131 | 0.14791 | -3.72733 | 0.000194 | 0.00079 | 0.682401 |
| TBC1D3L | 498.0843 | -0.83797 | 0.224884 | -3.72621 | 0.000194 | 0.000792 | 0.559431 |
| SFT2D1 | 610.6899 | -0.50117 | 0.13452 | -3.72563 | 0.000195 | 0.000794 | 0.706534 |
| PDLIM3 | 183.3334 | -0.65952 | 0.177091 | -3.72421 | 0.000196 | 0.000798 | 0.633088 |
| CALML6 | 18.6529 | -1.99871 | 0.536829 | -3.72318 | 0.000197 | 0.000801 | 0.250223 |
| PPP1R13L | 629.9786 | -0.57689 | 0.154987 | -3.7222 | 0.000197 | 0.000803 | 0.670406 |
| FCRL5 | 40.87048 | -1.65389 | 0.444693 | -3.71917 | 0.0002 | 0.000812 | 0.317782 |
| NMB | 112.6798 | -0.85461 | 0.229881 | -3.7176 | 0.000201 | 0.000817 | 0.553017 |
| MERTK | 211.1207 | -0.55632 | 0.149666 | -3.71706 | 0.000202 | 0.000818 | 0.680035 |
| TRABD2A | 213.2035 | -0.75316 | 0.202707 | -3.71548 | 0.000203 | 0.000823 | 0.593305 |
| TMEM191A | 82.20292 | -0.82734 | 0.222707 | -3.71495 | 0.000203 | 0.000825 | 0.563566 |
| NFKBID | 106.957 | -1.31348 | 0.353584 | -3.71476 | 0.000203 | 0.000825 | 0.402349 |
| AC116533 | 166.8522 | -10.8504 | 2.924496 | -3.71018 | 0.000207 | 0.000839 | 0.000542 |
| SATL1 | 64.5299 | -1.05187 | 0.283615 | -3.70881 | 0.000208 | 0.000843 | 0.482342 |
| EPOR | 348.8603 | -0.74347 | 0.200586 | -3.70647 | 0.00021 | 0.00085 | 0.597303 |
| PGRMC1 | 4881.651 | -0.43476 | 0.11735 | -3.70481 | 0.000212 | 0.000855 | 0.739816 |
| NT5C3A | 575.1128 | -0.51493 | 0.138999 | -3.70453 | 0.000212 | 0.000856 | 0.699829 |
| PLXNB1 | 4300.357 | -1.19438 | 0.322545 | -3.703 | 0.000213 | 0.000861 | 0.436973 |
| NTN5 | 21.36689 | -2.75374 | 0.743909 | -3.70172 | 0.000214 | 0.000865 | 0.148266 |

| | | | | | | | |
|---|---|---|---|---|---|---|---|
| ATP6AP1 | 2027.537 | -0.51561 | 0.139305 | -3.70126 | 0.000215 | 0.000866 | 0.699499 |
| SMAD7 | 228.6641 | -0.93284 | 0.252123 | -3.69992 | 0.000216 | 0.00087 | 0.523827 |
| CD99 | 1906.042 | -0.62395 | 0.16865 | -3.69967 | 0.000216 | 0.000871 | 0.648892 |
| SSR4 | 1490.183 | -0.51824 | 0.1401 | -3.69907 | 0.000216 | 0.000872 | 0.698222 |
| RMND5B | 1627.325 | -0.55587 | 0.150351 | -3.69716 | 0.000218 | 0.000879 | 0.680245 |
| MAT1A | 194.3993 | -0.57548 | 0.155664 | -3.69696 | 0.000218 | 0.000879 | 0.671062 |
| TNFSF13 | 96.82564 | -0.95062 | 0.257175 | -3.69639 | 0.000219 | 0.000881 | 0.517411 |
| STK32C | 356.6629 | -0.44332 | 0.119985 | -3.69484 | 0.00022 | 0.000886 | 0.735438 |
| SLC38A10 | 1384.149 | -0.66846 | 0.181018 | -3.6928 | 0.000222 | 0.000892 | 0.629176 |
| ANXA3 | 528.479 | -0.47872 | 0.12965 | -3.69236 | 0.000222 | 0.000893 | 0.717616 |
| DGAT1 | 581.4899 | -0.69516 | 0.188321 | -3.69138 | 0.000223 | 0.000896 | 0.617639 |
| TPO | 16.30874 | -2.5034 | 0.678343 | -3.69047 | 0.000224 | 0.000899 | 0.17636 |
| LINC0051[illegible] | 21.36708 | -1.60726 | 0.435823 | -3.68788 | 0.000226 | 0.000907 | 0.32822 |
| AC107918 | 65.45937 | -0.80649 | 0.218871 | -3.68478 | 0.000229 | 0.000918 | 0.571771 |
| ADRA2C | 490.3678 | -0.46885 | 0.127274 | -3.68375 | 0.00023 | 0.000921 | 0.722542 |
| CRIM1 | 629.7425 | -0.70424 | 0.191176 | -3.68372 | 0.00023 | 0.000921 | 0.613766 |
| NOCT | 257.8563 | -0.58673 | 0.159462 | -3.67944 | 0.000234 | 0.000936 | 0.66585 |
| AC015813 | 376.2532 | -1.24547 | 0.338532 | -3.67904 | 0.000234 | 0.000937 | 0.42177 |
| ARMC5 | 295.6954 | -0.48327 | 0.131438 | -3.67681 | 0.000236 | 0.000944 | 0.715354 |
| NMI | 45.45411 | -1.0995 | 0.299044 | -3.6767 | 0.000236 | 0.000944 | 0.46668 |
| TMEM37 | 216.8015 | -0.74648 | 0.203133 | -3.67484 | 0.000238 | 0.000951 | 0.596056 |
| PRCD | 17.34903 | -2.40423 | 0.654862 | -3.67135 | 0.000241 | 0.000963 | 0.18891 |
| MSC | 21.09943 | -1.92203 | 0.523579 | -3.67095 | 0.000242 | 0.000964 | 0.263882 |
| ABI3 | 54.4936 | -1.1171 | 0.304486 | -3.66883 | 0.000244 | 0.000971 | 0.461018 |
| LRFN2 | 27.91622 | -1.84328 | 0.502539 | -3.66794 | 0.000245 | 0.000974 | 0.278687 |
| DTX3L | 234.2776 | -0.62337 | 0.169989 | -3.66713 | 0.000245 | 0.000977 | 0.649152 |
| MPZL3 | 114.463 | -0.98511 | 0.268645 | -3.66698 | 0.000245 | 0.000977 | 0.505186 |
| OGFR | 607.3264 | -0.56962 | 0.155351 | -3.66664 | 0.000246 | 0.000978 | 0.673796 |
| VCX3A | 5.460166 | -5.92237 | 1.616047 | -3.66473 | 0.000248 | 0.000984 | 0.016489 |
| KCTD11 | 242.923 | -0.53215 | 0.145242 | -3.66387 | 0.000248 | 0.000987 | 0.691524 |
| FP565260. | 265.6723 | -0.53609 | 0.146385 | -3.66218 | 0.00025 | 0.000994 | 0.689639 |
| CLTB | 493.3992 | -0.4272 | 0.116685 | -3.66116 | 0.000251 | 0.000998 | 0.743703 |
| ICAM3 | 186.0956 | -0.64592 | 0.176456 | -3.6605 | 0.000252 | 0.001 | 0.639086 |
| NOMO3 | 62.22686 | -1.3262 | 0.362309 | -3.66042 | 0.000252 | 0.001 | 0.398816 |
| MT-CO3 | 137724.9 | -1.06577 | 0.291424 | -3.6571 | 0.000255 | 0.001012 | 0.477719 |
| KDELR3 | 139.3768 | -0.75038 | 0.20521 | -3.65665 | 0.000256 | 0.001014 | 0.594447 |
| ERFE | 76.15156 | -2.07869 | 0.568743 | -3.65488 | 0.000257 | 0.00102 | 0.236729 |
| STUM | 25.85922 | -1.52106 | 0.41618 | -3.6548 | 0.000257 | 0.00102 | 0.348431 |
| AKR1E2 | 38.20719 | -1.30305 | 0.356655 | -3.65352 | 0.000259 | 0.001025 | 0.405269 |
| MVP | 108.3103 | -0.97033 | 0.26569 | -3.65212 | 0.00026 | 0.00103 | 0.510388 |
| HYAL1 | 57.11934 | -1.22136 | 0.334631 | -3.64986 | 0.000262 | 0.001039 | 0.428879 |
| RIPK2 | 615.3855 | -0.4353 | 0.119268 | -3.64982 | 0.000262 | 0.001039 | 0.739537 |
| ODF3L2 | 10.52045 | -2.75165 | 0.754088 | -3.64898 | 0.000263 | 0.001042 | 0.148481 |
| IRF1 | 146.9997 | -0.80511 | 0.220689 | -3.64816 | 0.000264 | 0.001045 | 0.572319 |

| | | | | | | | |
|---|---|---|---|---|---|---|---|
| ABCA5 | 87.89925 | -1.28918 | 0.353692 | -3.64493 | 0.000267 | 0.001058 | 0.409183 |
| TRPM6 | 49.06229 | -1.20031 | 0.32933 | -3.64469 | 0.000268 | 0.001058 | 0.435183 |
| ABCD1 | 147.7366 | -0.5785 | 0.158728 | -3.64463 | 0.000268 | 0.001058 | 0.669658 |
| ACRBP | 48.49196 | -1.2623 | 0.346461 | -3.64342 | 0.000269 | 0.001063 | 0.416878 |
| TRIM7 | 12.81311 | -2.50001 | 0.686211 | -3.6432 | 0.000269 | 0.001064 | 0.176776 |
| SPTBN2 | 3206.052 | -1.25995 | 0.345996 | -3.64151 | 0.000271 | 0.00107 | 0.417558 |
| PPOX | 555.3458 | -0.66814 | 0.183515 | -3.6408 | 0.000272 | 0.001072 | 0.629317 |
| GLI4 | 234.2815 | -0.72524 | 0.199217 | -3.64045 | 0.000272 | 0.001074 | 0.604896 |
| IL15RA | 13.22161 | -2.26928 | 0.623791 | -3.63789 | 0.000275 | 0.001084 | 0.207433 |
| RBFOX3 | 315.0382 | -0.92328 | 0.253854 | -3.63704 | 0.000276 | 0.001087 | 0.52731 |
| SOHLH2 | 37.55413 | -1.32125 | 0.363419 | -3.63561 | 0.000277 | 0.001093 | 0.400188 |
| ZNF692 | 638.5525 | -0.79744 | 0.219637 | -3.6307 | 0.000283 | 0.001113 | 0.57537 |
| NEDD4L | 1666.409 | -0.41882 | 0.115421 | -3.62867 | 0.000285 | 0.001121 | 0.748034 |
| CRYZL2P | 9.741243 | -2.94763 | 0.812442 | -3.62811 | 0.000285 | 0.001123 | 0.129621 |
| COL13A1 | 304.8012 | -1.14427 | 0.315551 | -3.62627 | 0.000288 | 0.001131 | 0.452417 |
| CSDC2 | 45.04399 | -0.9889 | 0.272771 | -3.62539 | 0.000289 | 0.001135 | 0.503861 |
| ADAMTS2 | 269.9125 | -0.75013 | 0.206969 | -3.62433 | 0.00029 | 0.001138 | 0.594552 |
| SUMF1 | 294.564 | -0.52575 | 0.145062 | -3.62428 | 0.00029 | 0.001138 | 0.6946 |
| ALOXE3 | 13.28719 | -2.7202 | 0.750871 | -3.62272 | 0.000292 | 0.001145 | 0.151753 |
| BTN2A2 | 425.9425 | -0.52619 | 0.145286 | -3.62175 | 0.000293 | 0.001149 | 0.694386 |
| CD8A | 15.04236 | -2.29527 | 0.633799 | -3.62144 | 0.000293 | 0.00115 | 0.20373 |
| ECSCR | 30.00959 | -1.56244 | 0.431549 | -3.62054 | 0.000294 | 0.001154 | 0.338578 |
| GRIA3 | 120.2774 | -0.91256 | 0.252075 | -3.62019 | 0.000294 | 0.001155 | 0.531242 |
| TMEM25 | 314.4118 | -0.62902 | 0.173762 | -3.61998 | 0.000295 | 0.001156 | 0.646617 |
| CDX4 | 37.60913 | -1.08 | 0.298375 | -3.61961 | 0.000295 | 0.001157 | 0.473028 |
| A1BG | 130.2664 | -0.81964 | 0.226502 | -3.61868 | 0.000296 | 0.001161 | 0.566585 |
| MTLN | 261.1652 | -0.54381 | 0.15029 | -3.61839 | 0.000296 | 0.001162 | 0.685957 |
| AGAP3 | 1195.944 | -0.42211 | 0.116734 | -3.61601 | 0.000299 | 0.001171 | 0.746332 |
| PCDHGA6 | 41.94841 | -1.3526 | 0.374078 | -3.61582 | 0.000299 | 0.001172 | 0.391586 |
| CHST7 | 321.2452 | -0.45806 | 0.126707 | -3.61509 | 0.0003 | 0.001174 | 0.727966 |
| NAP1L6 | 9.370522 | -3.09969 | 0.857615 | -3.61431 | 0.000301 | 0.001177 | 0.116654 |
| RTP1 | 3.416038 | -5.24149 | 1.450221 | -3.61427 | 0.000301 | 0.001177 | 0.026434 |
| TMEM265 | 97.2006 | -0.81799 | 0.226392 | -3.61314 | 0.000303 | 0.001182 | 0.567233 |
| CD164L2 | 40.25529 | -1.50651 | 0.417216 | -3.61087 | 0.000305 | 0.001192 | 0.351961 |
| CABLES1 | 593.8669 | -0.43672 | 0.121042 | -3.60801 | 0.000309 | 0.001204 | 0.738811 |
| PLXNB3 | 75.30427 | -1.53938 | 0.426799 | -3.60679 | 0.00031 | 0.00121 | 0.344034 |
| SLC2A6 | 340.8193 | -0.8273 | 0.229407 | -3.60627 | 0.000311 | 0.001212 | 0.563582 |
| CIDECP1 | 150.2461 | -0.76596 | 0.212407 | -3.60609 | 0.000311 | 0.001212 | 0.588063 |
| CAPS | 114.0502 | -1.02837 | 0.285229 | -3.60542 | 0.000312 | 0.001215 | 0.490263 |
| VSTM2A | 23.54888 | -2.38024 | 0.660282 | -3.60488 | 0.000312 | 0.001217 | 0.192078 |
| ITPR2 | 586.5713 | -0.71654 | 0.198782 | -3.60466 | 0.000313 | 0.001218 | 0.608554 |
| NRN1L | 46.62097 | -1.45532 | 0.403886 | -3.6033 | 0.000314 | 0.001224 | 0.364674 |
| PGAP3 | 571.2234 | -0.52868 | 0.146934 | -3.59805 | 0.000321 | 0.001247 | 0.693191 |
| SCRIB | 3467.601 | -0.69048 | 0.191912 | -3.59792 | 0.000321 | 0.001247 | 0.619646 |

| | | | | | | | |
|---|---|---|---|---|---|---|---|
| TEK | 587.7211 | -0.50403 | 0.140127 | -3.59696 | 0.000322 | 0.001251 | 0.705134 |
| GALE | 386.2094 | -0.46668 | 0.129777 | -3.59603 | 0.000323 | 0.001255 | 0.723626 |
| PGA4 | 36.43537 | -1.66934 | 0.464355 | -3.59497 | 0.000324 | 0.00126 | 0.314396 |
| CBWD5 | 642.9267 | -0.4656 | 0.129539 | -3.59428 | 0.000325 | 0.001263 | 0.724169 |
| CD44 | 174.1803 | -1.05844 | 0.294548 | -3.59342 | 0.000326 | 0.001267 | 0.480153 |
| TPCN1 | 993.9744 | -0.53809 | 0.149956 | -3.58829 | 0.000333 | 0.00129 | 0.688683 |
| HIST1H3H | 32.0771 | -1.3713 | 0.382519 | -3.58491 | 0.000337 | 0.001305 | 0.386544 |
| CEACAM1¦ | 103.3568 | -1.04513 | 0.291697 | -3.58295 | 0.00034 | 0.001315 | 0.4846 |
| IFI6 | 137.9931 | -0.91782 | 0.256832 | -3.5736 | 0.000352 | 0.00136 | 0.52931 |
| VN1R1 | 29.40891 | -1.47549 | 0.413011 | -3.57253 | 0.000354 | 0.001364 | 0.35961 |
| GUSB | 1474.161 | -0.46117 | 0.12913 | -3.57138 | 0.000355 | 0.001369 | 0.726397 |
| IGFBP2 | 9098.603 | -1.0804 | 0.302796 | -3.56809 | 0.00036 | 0.001385 | 0.472897 |
| CDA | 60.33405 | -0.9163 | 0.256832 | -3.5677 | 0.00036 | 0.001387 | 0.529867 |
| OR7E122P | 55.34678 | -1.03915 | 0.291413 | -3.56588 | 0.000363 | 0.001395 | 0.486615 |
| RAB3A | 111.746 | -0.83125 | 0.233119 | -3.56577 | 0.000363 | 0.001396 | 0.562042 |
| NFKBIA | 412.9582 | -0.42898 | 0.120378 | -3.56357 | 0.000366 | 0.001406 | 0.742789 |
| AC092865 | 18.69592 | -1.71665 | 0.48195 | -3.56188 | 0.000368 | 0.001414 | 0.304255 |
| RASA3 | 637.0516 | -0.47038 | 0.132114 | -3.56041 | 0.00037 | 0.001421 | 0.721774 |
| MID2 | 62.91729 | -0.90991 | 0.255769 | -3.55755 | 0.000374 | 0.001435 | 0.532218 |
| PERM1 | 9.8861 | -3.1827 | 0.894696 | -3.5573 | 0.000375 | 0.001436 | 0.110131 |
| TMEM121E | 358.8818 | -0.50041 | 0.14067 | -3.55733 | 0.000375 | 0.001436 | 0.706906 |
| APOBEC3D | 83.46134 | -1.02967 | 0.289632 | -3.55508 | 0.000378 | 0.001447 | 0.489824 |
| PPP1R37 | 783.7077 | -0.48466 | 0.136404 | -3.55314 | 0.000381 | 0.001458 | 0.714665 |
| SCGB2B2 | 12.75821 | -3.32208 | 0.935198 | -3.55228 | 0.000382 | 0.001461 | 0.099989 |
| AACSP1 | 14.90324 | -1.80922 | 0.509426 | -3.5515 | 0.000383 | 0.001465 | 0.285345 |
| NEURL4 | 1325.174 | -0.69785 | 0.196493 | -3.55152 | 0.000383 | 0.001465 | 0.616491 |
| CASP9 | 356.5447 | -0.4846 | 0.136466 | -3.55103 | 0.000384 | 0.001467 | 0.714697 |
| DEGS1 | 1368.556 | -0.47751 | 0.134511 | -3.54995 | 0.000385 | 0.001472 | 0.718217 |
| MAP11 | 322.8061 | -0.61914 | 0.174437 | -3.54939 | 0.000386 | 0.001474 | 0.651057 |
| C20orf96 | 361.4629 | -0.41824 | 0.117844 | -3.54912 | 0.000387 | 0.001476 | 0.748335 |
| MAL2 | 3733.02 | -0.43669 | 0.123046 | -3.549 | 0.000387 | 0.001476 | 0.738828 |
| TEN1-CDK | 119.4172 | -1.19174 | 0.335854 | -3.54838 | 0.000388 | 0.001479 | 0.437775 |
| LIMS2 | 78.62052 | -0.89577 | 0.252621 | -3.5459 | 0.000391 | 0.001491 | 0.537461 |
| LMNTD2 | 34.41154 | -1.37623 | 0.38812 | -3.54589 | 0.000391 | 0.001491 | 0.385224 |
| RHBDD3 | 1142.689 | -0.54272 | 0.153068 | -3.54561 | 0.000392 | 0.001492 | 0.686475 |
| EIF4HP2 | 23.08931 | -1.71344 | 0.483342 | -3.54498 | 0.000393 | 0.001495 | 0.304933 |
| SYNJ2 | 452.1083 | -0.51259 | 0.144697 | -3.54252 | 0.000396 | 0.001507 | 0.700962 |
| TMUB1 | 513.502 | -0.57826 | 0.163265 | -3.54187 | 0.000397 | 0.001511 | 0.669769 |
| PHF19 | 596.0062 | -0.55416 | 0.15674 | -3.53557 | 0.000407 | 0.001545 | 0.681051 |
| CACNA1H | 2238.357 | -0.54174 | 0.153267 | -3.53463 | 0.000408 | 0.00155 | 0.68694 |
| PLOD2 | 604.4568 | -0.86092 | 0.243612 | -3.53396 | 0.000409 | 0.001554 | 0.550603 |
| SVIL-AS1 | 17.23559 | -2.47687 | 0.700995 | -3.53336 | 0.00041 | 0.001556 | 0.179634 |
| ZDHHC11 | 453.0612 | -0.72155 | 0.204319 | -3.53148 | 0.000413 | 0.001567 | 0.606446 |
| KLF15 | 399.193 | -0.46462 | 0.131576 | -3.53123 | 0.000414 | 0.001568 | 0.72466 |

| | | | | | | | |
|---|---|---|---|---|---|---|---|
| KLHL29 | 116.7316 | -0.77079 | 0.218384 | -3.52953 | 0.000416 | 0.001576 | 0.586095 |
| OR2A20P | 161.0509 | -1.13151 | 0.320579 | -3.52958 | 0.000416 | 0.001576 | 0.456438 |
| PEX5L | 107.5403 | -1.70649 | 0.483848 | -3.52692 | 0.00042 | 0.00159 | 0.306404 |
| ENTPD3 | 11.51084 | -2.57572 | 0.730944 | -3.52383 | 0.000425 | 0.001607 | 0.167737 |
| SORCS3 | 34.87691 | -1.20483 | 0.341961 | -3.5233 | 0.000426 | 0.00161 | 0.43382 |
| ATF3 | 925.1005 | -0.73384 | 0.208308 | -3.52288 | 0.000427 | 0.001612 | 0.6013 |
| MME | 266.5897 | -1.09049 | 0.30973 | -3.52078 | 0.00043 | 0.001624 | 0.469601 |
| GAS8 | 287.0746 | -0.71934 | 0.204395 | -3.51936 | 0.000433 | 0.001632 | 0.607376 |
| SPTSSB | 157.1819 | -1.28327 | 0.364739 | -3.51833 | 0.000434 | 0.001637 | 0.410863 |
| TPSD1 | 6.556821 | -4.73721 | 1.346449 | -3.5183 | 0.000434 | 0.001637 | 0.037494 |
| UCP1 | 15.23879 | -2.32305 | 0.660457 | -3.51734 | 0.000436 | 0.001643 | 0.199844 |
| IGSF5 | 48.47122 | -1.03417 | 0.294088 | -3.51654 | 0.000437 | 0.001647 | 0.488296 |
| OFD1 | 615.8803 | -0.46213 | 0.131428 | -3.51625 | 0.000438 | 0.001649 | 0.725911 |
| ATAD3B | 1314.869 | -0.53594 | 0.152444 | -3.51567 | 0.000439 | 0.001652 | 0.689708 |
| INHA | 13.80549 | -2.38672 | 0.679133 | -3.51436 | 0.000441 | 0.001659 | 0.191216 |
| FAM71F1 | 134.4249 | -0.8551 | 0.243432 | -3.51268 | 0.000444 | 0.001669 | 0.552827 |
| RASSF1 | 312.7279 | -0.42794 | 0.121898 | -3.51063 | 0.000447 | 0.001681 | 0.743322 |
| CECR7 | 195.0775 | -0.58107 | 0.165573 | -3.50941 | 0.000449 | 0.001688 | 0.66847 |
| TMEM182 | 233.9566 | -0.48941 | 0.139459 | -3.50932 | 0.000449 | 0.001688 | 0.712318 |
| TMEM86A | 77.53472 | -1.27112 | 0.362287 | -3.5086 | 0.00045 | 0.001692 | 0.414337 |
| SERPINE1 | 199.8607 | -0.62579 | 0.178363 | -3.50851 | 0.000451 | 0.001692 | 0.648066 |
| PLEKHH3 | 681.5703 | -0.51322 | 0.146336 | -3.50713 | 0.000453 | 0.001701 | 0.700657 |
| SERINC2 | 803.7976 | -0.48914 | 0.139488 | -3.50671 | 0.000454 | 0.001703 | 0.712448 |
| CYP4V2 | 160.6087 | -0.72319 | 0.206236 | -3.50661 | 0.000454 | 0.001703 | 0.605756 |
| MYMX | 45.40968 | -1.07662 | 0.307065 | -3.50616 | 0.000455 | 0.001706 | 0.474138 |
| AC068946 | 43.00696 | -1.53056 | 0.436587 | -3.50574 | 0.000455 | 0.001708 | 0.346143 |
| ATG16L2 | 546.7143 | -0.82648 | 0.235758 | -3.50562 | 0.000456 | 0.001709 | 0.563903 |
| AC027763 | 14.60626 | -2.04169 | 0.582465 | -3.50525 | 0.000456 | 0.00171 | 0.242879 |
| OR2A9P | 210.3138 | -0.54168 | 0.154566 | -3.50453 | 0.000457 | 0.001714 | 0.68697 |
| ALG9 | 673.2824 | -0.47966 | 0.136914 | -3.50337 | 0.000459 | 0.001721 | 0.717146 |
| SOX15 | 55.88452 | -0.89942 | 0.256731 | -3.50335 | 0.000459 | 0.001721 | 0.536102 |
| CATSPER4 | 14.74178 | -1.75411 | 0.500721 | -3.50317 | 0.00046 | 0.001722 | 0.296455 |
| CNKSR1 | 413.574 | -0.76226 | 0.217596 | -3.50307 | 0.00046 | 0.001722 | 0.589574 |
| C10orf95 | 43.29634 | -1.11074 | 0.317217 | -3.5015 | 0.000463 | 0.001731 | 0.463058 |
| MFSD11 | 369.1356 | -0.64765 | 0.185052 | -3.49983 | 0.000466 | 0.001741 | 0.63832 |
| ARSD | 332.9311 | -0.68283 | 0.195166 | -3.49871 | 0.000468 | 0.001747 | 0.622942 |
| MICB | 139.5223 | -0.67286 | 0.192349 | -3.49812 | 0.000469 | 0.001751 | 0.627263 |
| OPLAH | 973.3314 | -0.61205 | 0.175004 | -3.49737 | 0.00047 | 0.001755 | 0.654264 |
| EHBP1L1 | 351.1468 | -1.13027 | 0.323539 | -3.49345 | 0.000477 | 0.001778 | 0.456831 |
| ARHGEF15 | 40.97899 | -1.28167 | 0.36696 | -3.49266 | 0.000478 | 0.001783 | 0.41132 |
| MT-CO2 | 137315.1 | -1.14263 | 0.327504 | -3.48891 | 0.000485 | 0.001807 | 0.452933 |
| MMRN2 | 47.43715 | -1.13059 | 0.324189 | -3.48743 | 0.000488 | 0.001816 | 0.45673 |
| KLK7 | 11.43845 | -2.17447 | 0.624341 | -3.48282 | 0.000496 | 0.001845 | 0.221523 |
| PDE2A | 189.5139 | -0.82373 | 0.236546 | -3.48232 | 0.000497 | 0.001848 | 0.56498 |

| | | | | | | | |
|---|---|---|---|---|---|---|---|
| CLEC4G | 6.406233 | -3.2469 | 0.932496 | -3.48194 | 0.000498 | 0.00185 | 0.105339 |
| DGAT2 | 333.4247 | -0.68622 | 0.197124 | -3.48115 | 0.000499 | 0.001855 | 0.621481 |
| ZNF589 | 2023.173 | -0.44179 | 0.126964 | -3.47967 | 0.000502 | 0.001864 | 0.736219 |
| GOLGA6L9 | 494.0892 | -0.63624 | 0.182907 | -3.47847 | 0.000504 | 0.001871 | 0.64339 |
| SLC2A10 | 502.3274 | -0.42223 | 0.121405 | -3.4779 | 0.000505 | 0.001874 | 0.746268 |
| NEURL2 | 50.82207 | -0.99973 | 0.287534 | -3.47691 | 0.000507 | 0.00188 | 0.500094 |
| GALNT9 | 25.4039 | -1.56221 | 0.449396 | -3.47623 | 0.000509 | 0.001884 | 0.338633 |
| PTPMT1 | 744.0072 | -0.48293 | 0.139057 | -3.47288 | 0.000515 | 0.001905 | 0.715524 |
| CD70 | 17.86189 | -1.77506 | 0.511388 | -3.47106 | 0.000518 | 0.001916 | 0.292183 |
| CHRM1 | 15.79767 | -2.37863 | 0.685262 | -3.47113 | 0.000518 | 0.001916 | 0.192291 |
| HCLS1 | 16.56065 | -1.8292 | 0.52723 | -3.46945 | 0.000522 | 0.001927 | 0.281421 |
| PBX2 | 286.8666 | -0.6949 | 0.200357 | -3.46831 | 0.000524 | 0.001934 | 0.617752 |
| AC006254 | 45.77171 | -1.2381 | 0.357345 | -3.46471 | 0.000531 | 0.001955 | 0.423931 |
| RRN3P3 | 96.09266 | -0.74307 | 0.214706 | -3.46089 | 0.000538 | 0.001981 | 0.597466 |
| OR5H6 | 3.132728 | -5.10782 | 1.477366 | -3.45738 | 0.000545 | 0.002005 | 0.029 |
| PLA2G12A | 687.5249 | -0.54862 | 0.158742 | -3.45604 | 0.000548 | 0.002015 | 0.683674 |
| CUL9 | 557.9678 | -0.84111 | 0.243425 | -3.45532 | 0.00055 | 0.002019 | 0.558213 |
| PPT2-EGFL | 18.15837 | -2.54245 | 0.735801 | -3.45536 | 0.00055 | 0.002019 | 0.17165 |
| RPRML | 27.94211 | -1.43694 | 0.41595 | -3.45459 | 0.000551 | 0.002024 | 0.36935 |
| MT-ATP6 | 123207.4 | -1.1277 | 0.326514 | -3.45376 | 0.000553 | 0.00203 | 0.457645 |
| MT-ND2 | 64993.3 | -1.09197 | 0.31665 | -3.44851 | 0.000564 | 0.002067 | 0.469121 |
| NIPAL3 | 347.3166 | -0.50406 | 0.146213 | -3.44747 | 0.000566 | 0.002074 | 0.705118 |
| GPR26 | 6.797982 | -3.05944 | 0.887882 | -3.44577 | 0.000569 | 0.002087 | 0.119954 |
| SERPINB6 | 550.5263 | -0.51601 | 0.149797 | -3.44474 | 0.000572 | 0.002094 | 0.699302 |
| GYG1 | 694.5362 | -0.42056 | 0.122115 | -3.44402 | 0.000573 | 0.002099 | 0.747132 |
| PSMG4 | 411.4777 | -0.43216 | 0.12556 | -3.44184 | 0.000578 | 0.002114 | 0.741152 |
| DRD5P2 | 8.083217 | -3.30586 | 0.960981 | -3.44009 | 0.000582 | 0.002127 | 0.10112 |
| ZNF883 | 103.2663 | -0.7213 | 0.209702 | -3.43963 | 0.000583 | 0.00213 | 0.606552 |
| AF228730. | 183.6921 | -0.71453 | 0.207754 | -3.43929 | 0.000583 | 0.002132 | 0.609405 |
| HEXA | 1434.394 | -0.44187 | 0.128547 | -3.43741 | 0.000587 | 0.002146 | 0.73618 |
| LAMA2 | 136.0097 | -1.02022 | 0.29696 | -3.43555 | 0.000591 | 0.002159 | 0.493041 |
| LYSMD2 | 38.8182 | -1.10533 | 0.32173 | -3.43559 | 0.000591 | 0.002159 | 0.464795 |
| MBOAT7 | 1056.562 | -0.61718 | 0.179717 | -3.43417 | 0.000594 | 0.002169 | 0.651944 |
| G0S2 | 203.4281 | -1.14153 | 0.332544 | -3.43273 | 0.000598 | 0.002178 | 0.453277 |
| MCOLN3 | 262.62 | -0.63505 | 0.185013 | -3.43248 | 0.000598 | 0.002179 | 0.643917 |
| C11orf80 | 666.5101 | -0.54911 | 0.159988 | -3.4322 | 0.000599 | 0.002181 | 0.683441 |
| VEPH1 | 186.9654 | -0.8362 | 0.243778 | -3.43019 | 0.000603 | 0.002196 | 0.560115 |
| HLA-F | 59.44934 | -2.11342 | 0.616204 | -3.42973 | 0.000604 | 0.002199 | 0.231099 |
| TDH | 24.15149 | -1.49359 | 0.43564 | -3.42849 | 0.000607 | 0.002208 | 0.355128 |
| MGAT4C | 120.5237 | -0.88536 | 0.258277 | -3.42797 | 0.000608 | 0.002211 | 0.541351 |
| PRR5 | 327.1169 | -0.53138 | 0.155134 | -3.42531 | 0.000614 | 0.00223 | 0.691892 |
| TBC1D24 | 598.5317 | -0.42359 | 0.123684 | -3.42479 | 0.000615 | 0.002233 | 0.745566 |
| CRELD2 | 486.6524 | -0.61633 | 0.180071 | -3.42272 | 0.00062 | 0.00225 | 0.652327 |
| TCL1A | 3.176407 | -5.14545 | 1.503583 | -3.42213 | 0.000621 | 0.002255 | 0.028253 |

| | | | | | | | |
|---|---|---|---|---|---|---|---|
| CNR1 | 36.2675 | -1.24867 | 0.365126 | -3.41982 | 0.000627 | 0.002272 | 0.420837 |
| TGM1 | 38.84174 | -1.57986 | 0.462267 | -3.41763 | 0.000632 | 0.002289 | 0.334515 |
| TMEM198E | 409.4074 | -0.49972 | 0.146232 | -3.41729 | 0.000632 | 0.002291 | 0.707246 |
| ZNF767P | 297.6388 | -0.46946 | 0.137445 | -3.41563 | 0.000636 | 0.002304 | 0.722233 |
| PCDHGA1: | 22.24843 | -1.71645 | 0.502666 | -3.4147 | 0.000639 | 0.00231 | 0.304296 |
| AC093668 | 43.33477 | -2.5088 | 0.735052 | -3.41309 | 0.000642 | 0.002321 | 0.175701 |
| GFRA3 | 49.28541 | -1.10314 | 0.323242 | -3.41273 | 0.000643 | 0.002323 | 0.465503 |
| AC008977 | 479.4526 | -0.49218 | 0.144237 | -3.41226 | 0.000644 | 0.002326 | 0.710952 |
| GALR2 | 9.142589 | -2.38336 | 0.69865 | -3.41138 | 0.000646 | 0.002333 | 0.191663 |
| TREML2 | 7.13744 | -3.14831 | 0.923379 | -3.40955 | 0.000651 | 0.002348 | 0.112789 |
| ANKRD30E | 6.038005 | -2.95002 | 0.865257 | -3.40941 | 0.000651 | 0.002348 | 0.129407 |
| SPOCK1 | 244.5072 | -0.58662 | 0.172093 | -3.40872 | 0.000653 | 0.002353 | 0.665902 |
| HIST1H2BJ | 26.25658 | -2.16467 | 0.635318 | -3.40722 | 0.000656 | 0.002365 | 0.223033 |
| GSTT2 | 10.38486 | -3.12695 | 0.91801 | -3.40623 | 0.000659 | 0.002373 | 0.11447 |
| RAMP2 | 207.0121 | -0.62746 | 0.184286 | -3.40481 | 0.000662 | 0.002384 | 0.647316 |
| GNA15 | 13.32962 | -2.22472 | 0.653418 | -3.40474 | 0.000662 | 0.002384 | 0.21394 |
| PLPPR3 | 1625.525 | -0.69492 | 0.204106 | -3.4047 | 0.000662 | 0.002384 | 0.617745 |
| TNFRSF11/ | 240.1467 | -1.05836 | 0.311052 | -3.40253 | 0.000668 | 0.002401 | 0.480176 |
| GABRB3 | 3113.948 | -0.44828 | 0.131767 | -3.40204 | 0.000669 | 0.002405 | 0.732918 |
| CSF1 | 220.3571 | -0.66547 | 0.195719 | -3.40015 | 0.000674 | 0.00242 | 0.630482 |
| HS3ST2 | 22.77024 | -1.74019 | 0.512123 | -3.39798 | 0.000679 | 0.002438 | 0.299331 |
| NAPB | 442.4337 | -0.46239 | 0.136102 | -3.39737 | 0.00068 | 0.002443 | 0.725784 |
| CTRL | 28.45482 | -1.4703 | 0.432842 | -3.39686 | 0.000682 | 0.002447 | 0.360907 |
| NR5A2 | 32.26178 | -1.26375 | 0.372042 | -3.39679 | 0.000682 | 0.002447 | 0.416461 |
| RETSAT | 523.4362 | -0.45908 | 0.135194 | -3.3957 | 0.000685 | 0.002455 | 0.727451 |
| ZNF365 | 19.22798 | -1.51079 | 0.444954 | -3.39537 | 0.000685 | 0.002458 | 0.35092 |
| CYB561A3 | 470.3143 | -0.49721 | 0.146568 | -3.39237 | 0.000693 | 0.002483 | 0.708473 |
| CKMT1B | 348.0831 | -0.60496 | 0.178397 | -3.39107 | 0.000696 | 0.002494 | 0.657492 |
| GABRD | 34.41404 | -1.39039 | 0.41007 | -3.3906 | 0.000697 | 0.002498 | 0.381463 |
| CYP4X1 | 76.86213 | -0.94575 | 0.279017 | -3.38957 | 0.0007 | 0.002506 | 0.519161 |
| HACD1 | 227.5477 | -0.63929 | 0.18861 | -3.38946 | 0.0007 | 0.002507 | 0.64203 |
| SIAE | 143.7079 | -0.69866 | 0.206224 | -3.38788 | 0.000704 | 0.00252 | 0.616143 |
| BSG | 19559.15 | -0.52516 | 0.155016 | -3.3878 | 0.000705 | 0.002521 | 0.69488 |
| AC010761 | 424.1698 | -0.69055 | 0.20391 | -3.38655 | 0.000708 | 0.002532 | 0.619617 |
| PLEKHG6 | 572.9893 | -0.41822 | 0.123512 | -3.38604 | 0.000709 | 0.002535 | 0.748349 |
| SLC29A2 | 625.2466 | -0.62694 | 0.185192 | -3.38535 | 0.000711 | 0.002541 | 0.64755 |
| LSMEM1 | 19.21808 | -1.99867 | 0.59041 | -3.38522 | 0.000711 | 0.002542 | 0.250231 |
| VWA5B1 | 26.5291 | -1.47452 | 0.435573 | -3.38523 | 0.000711 | 0.002542 | 0.359854 |
| TMEM114 | 54.19019 | -1.16277 | 0.343531 | -3.38477 | 0.000712 | 0.002544 | 0.446653 |
| PSMB8 | 39.38682 | -2.84345 | 0.840183 | -3.38433 | 0.000714 | 0.002547 | 0.139327 |
| NPIPA7 | 98.74911 | -1.41541 | 0.418352 | -3.3833 | 0.000716 | 0.002556 | 0.374904 |
| CAMK2N2 | 197.6965 | -0.60215 | 0.178094 | -3.38106 | 0.000722 | 0.002573 | 0.658773 |
| CD248 | 63.13704 | -1.14545 | 0.338851 | -3.38038 | 0.000724 | 0.002579 | 0.45205 |
| CLIP2 | 1325.241 | -0.42711 | 0.12641 | -3.37878 | 0.000728 | 0.002593 | 0.743748 |

| | | | | | | | |
|---|---|---|---|---|---|---|---|
| LRCH1 | 214.7041 | -0.53813 | 0.159341 | -3.37725 | 0.000732 | 0.002605 | 0.688661 |
| OR5H8 | 2.889319 | -5.00193 | 1.481113 | -3.37714 | 0.000732 | 0.002605 | 0.031208 |
| WDR86 | 108.6267 | -0.86021 | 0.254746 | -3.37674 | 0.000734 | 0.002608 | 0.550873 |
| PARP6 | 656.2173 | -0.52387 | 0.155215 | -3.37514 | 0.000738 | 0.002622 | 0.695503 |
| ZNF93 | 5.803384 | -3.67895 | 1.090023 | -3.37511 | 0.000738 | 0.002622 | 0.078078 |
| ATP10B | 27.70377 | -1.53592 | 0.455096 | -3.37493 | 0.000738 | 0.002623 | 0.34486 |
| IER2 | 2735.403 | -0.42469 | 0.125888 | -3.37355 | 0.000742 | 0.002635 | 0.745 |
| PRPF40B | 759.2236 | -0.61598 | 0.1827 | -3.37156 | 0.000747 | 0.002652 | 0.652485 |
| SGK3 | 161.1031 | -0.62976 | 0.186855 | -3.37032 | 0.000751 | 0.002663 | 0.646283 |
| C17orf58 | 378.0462 | -0.54014 | 0.160383 | -3.36779 | 0.000758 | 0.002686 | 0.687706 |
| CD81 | 4884.692 | -0.53482 | 0.158849 | -3.36684 | 0.00076 | 0.002695 | 0.690245 |
| ADGRE5 | 60.31894 | -1.29094 | 0.383472 | -3.36646 | 0.000761 | 0.002697 | 0.408684 |
| SFXN5 | 699.7057 | -0.45595 | 0.135443 | -3.3664 | 0.000762 | 0.002697 | 0.729028 |
| ANKZF1 | 1206.42 | -0.49197 | 0.146155 | -3.36607 | 0.000762 | 0.0027 | 0.711054 |
| HECW1 | 65.4811 | -1.68378 | 0.500404 | -3.36484 | 0.000766 | 0.002711 | 0.311266 |
| ATXN7L2 | 222.1341 | -0.53609 | 0.159326 | -3.36476 | 0.000766 | 0.002711 | 0.689635 |
| TMCO6 | 430.5083 | -0.5338 | 0.158734 | -3.36283 | 0.000771 | 0.002728 | 0.690734 |
| ALOX15B | 11.5331 | -2.04843 | 0.609351 | -3.36166 | 0.000775 | 0.002739 | 0.241747 |
| PRRX2 | 66.11606 | -0.9112 | 0.271114 | -3.36094 | 0.000777 | 0.002745 | 0.531743 |
| SLC22A5 | 178.2438 | -0.68062 | 0.202549 | -3.3603 | 0.000779 | 0.002751 | 0.623895 |
| PRRG4 | 94.12151 | -0.64031 | 0.190636 | -3.3588 | 0.000783 | 0.002763 | 0.641576 |
| ULK4P3 | 33.22427 | -1.58556 | 0.472239 | -3.35755 | 0.000786 | 0.002774 | 0.333195 |
| CLEC2L | 13.16928 | -2.11606 | 0.630339 | -3.35701 | 0.000788 | 0.002779 | 0.230677 |
| WDR46 | 96.92551 | -10.0667 | 2.999423 | -3.3562 | 0.00079 | 0.002786 | 0.000932 |
| ZNF530 | 136.7622 | -0.75015 | 0.223569 | -3.35535 | 0.000793 | 0.002795 | 0.59454 |
| INSR | 1277.56 | -0.56613 | 0.168795 | -3.35398 | 0.000797 | 0.002807 | 0.675424 |
| VSTM4 | 130.089 | -0.65943 | 0.19673 | -3.35195 | 0.000802 | 0.002825 | 0.633129 |
| CDKN1C | 330.6013 | -0.5897 | 0.176039 | -3.3498 | 0.000809 | 0.002845 | 0.664483 |
| DUSP13 | 31.12044 | -1.53757 | 0.459125 | -3.34892 | 0.000811 | 0.002852 | 0.344465 |
| AC106865 | 64.20758 | -0.82245 | 0.245727 | -3.34699 | 0.000817 | 0.002871 | 0.565482 |
| IKBKE | 271.2014 | -0.90314 | 0.270014 | -3.34479 | 0.000823 | 0.002893 | 0.534722 |
| SNX14 | 703.0852 | -0.52996 | 0.158581 | -3.34188 | 0.000832 | 0.00292 | 0.692575 |
| CALCRL | 71.67879 | -0.89435 | 0.267647 | -3.34155 | 0.000833 | 0.002922 | 0.537988 |
| PRRG2 | 64.44074 | -1.2347 | 0.36962 | -3.34046 | 0.000836 | 0.002932 | 0.42493 |
| AKAP8L | 2143.391 | -0.61986 | 0.185621 | -3.33939 | 0.00084 | 0.002943 | 0.650734 |
| SLC5A7 | 2.760421 | -4.93159 | 1.476879 | -3.3392 | 0.00084 | 0.002944 | 0.032767 |
| REEP2 | 1126.222 | -0.472 | 0.141422 | -3.33757 | 0.000845 | 0.002961 | 0.720962 |
| TRIM58 | 22.42354 | -1.52389 | 0.456897 | -3.33531 | 0.000852 | 0.002983 | 0.347746 |
| GALK1 | 500.1113 | -0.47744 | 0.143294 | -3.33192 | 0.000863 | 0.003018 | 0.718249 |
| PAN2 | 980.8844 | -0.66474 | 0.199592 | -3.3305 | 0.000867 | 0.003032 | 0.630801 |
| REXO2 | 905.353 | -0.43496 | 0.130616 | -3.33005 | 0.000868 | 0.003037 | 0.739716 |
| C16orf74 | 80.40585 | -0.80818 | 0.242838 | -3.32804 | 0.000875 | 0.003058 | 0.571103 |
| SLC2A5 | 7.29779 | -2.49974 | 0.751373 | -3.3269 | 0.000878 | 0.00307 | 0.176808 |
| NGB | 5.758623 | -3.46993 | 1.043082 | -3.32661 | 0.000879 | 0.003072 | 0.09025 |

| | | | | | | | |
|---|---|---|---|---|---|---|---|
| STYK1 | 44.42527 | -1.38436 | 0.416143 | -3.32663 | 0.000879 | 0.003072 | 0.38306 |
| AL669831. | 48.55939 | -1.05972 | 0.318799 | -3.32409 | 0.000887 | 0.003095 | 0.479726 |
| CPSF1 | 3785.205 | -0.5889 | 0.177182 | -3.32368 | 0.000888 | 0.003099 | 0.664852 |
| TRAC | 12.71828 | -2.68445 | 0.807699 | -3.32358 | 0.000889 | 0.0031 | 0.155561 |
| AMZ1 | 34.16554 | -1.91289 | 0.575981 | -3.32109 | 0.000897 | 0.003126 | 0.26556 |
| SOX3 | 263.87 | -0.44738 | 0.134845 | -3.31775 | 0.000907 | 0.00316 | 0.733372 |
| GZMM | 3.09892 | -5.10305 | 1.538213 | -3.31752 | 0.000908 | 0.003161 | 0.029096 |
| FCGRT | 634.0326 | -0.59528 | 0.179474 | -3.31681 | 0.000911 | 0.003166 | 0.661915 |
| IAH1 | 135.8878 | -0.85601 | 0.258191 | -3.31542 | 0.000915 | 0.003181 | 0.552478 |
| TMEM56 | 123.5447 | -1.23911 | 0.37375 | -3.31535 | 0.000915 | 0.003181 | 0.423633 |
| EVC2 | 328.7882 | -0.41837 | 0.126247 | -3.31392 | 0.00092 | 0.003195 | 0.748268 |
| SLC16A13 | 103.241 | -0.67133 | 0.202605 | -3.3135 | 0.000921 | 0.003199 | 0.627927 |
| MBTPS2 | 576.4431 | -0.44134 | 0.133219 | -3.31287 | 0.000923 | 0.003205 | 0.736451 |
| IL21R | 30.78692 | -1.18274 | 0.357078 | -3.31227 | 0.000925 | 0.00321 | 0.440515 |
| AC063952 | 22.25285 | -1.37143 | 0.414416 | -3.30932 | 0.000935 | 0.003239 | 0.386507 |
| MFSD4A | 20.81315 | -1.90911 | 0.576896 | -3.30927 | 0.000935 | 0.003239 | 0.266257 |
| TLN1 | 4021.158 | -0.58049 | 0.175472 | -3.30815 | 0.000939 | 0.003251 | 0.668738 |
| EHHADH | 113.8514 | -1.1698 | 0.353618 | -3.30807 | 0.000939 | 0.003251 | 0.444484 |
| AKAP1 | 1782.38 | -0.48381 | 0.146283 | -3.30739 | 0.000942 | 0.003258 | 0.715085 |
| POLRMT | 2686.924 | -0.43231 | 0.130715 | -3.3073 | 0.000942 | 0.003258 | 0.741072 |
| SHISA3 | 566.8098 | -0.50708 | 0.153415 | -3.30527 | 0.000949 | 0.00328 | 0.703646 |
| MT-ND5 | 69358.71 | -1.00993 | 0.305603 | -3.30471 | 0.000951 | 0.003286 | 0.496571 |
| SH2D2A | 117.3327 | -0.69752 | 0.211106 | -3.30412 | 0.000953 | 0.003291 | 0.616632 |
| KLK6 | 72.6117 | -1.14283 | 0.345947 | -3.30347 | 0.000955 | 0.003298 | 0.452871 |
| PCDHA7 | 21.41136 | -1.93685 | 0.586759 | -3.30092 | 0.000964 | 0.003325 | 0.261187 |
| PPM1B | 3474.714 | -0.46668 | 0.141407 | -3.30026 | 0.000966 | 0.003332 | 0.723629 |
| AC105233 | 12.75857 | -2.19111 | 0.664011 | -3.2998 | 0.000968 | 0.003337 | 0.218984 |
| NLRC5 | 81.97369 | -1.25941 | 0.381703 | -3.29944 | 0.000969 | 0.00334 | 0.417715 |
| SYT17 | 129.3773 | -1.03595 | 0.314058 | -3.29858 | 0.000972 | 0.003349 | 0.487696 |
| EMP3 | 182.3573 | -0.57891 | 0.17554 | -3.29788 | 0.000974 | 0.003357 | 0.669469 |
| NT5M | 220.5728 | -0.59659 | 0.180904 | -3.29783 | 0.000974 | 0.003357 | 0.661316 |
| TAF1C | 1970.147 | -0.50166 | 0.152153 | -3.2971 | 0.000977 | 0.003365 | 0.706292 |
| SLC25A16 | 250.7348 | -1.09302 | 0.331532 | -3.29689 | 0.000978 | 0.003367 | 0.468778 |
| DPEP3 | 3.108957 | -5.08176 | 1.541922 | -3.29573 | 0.000982 | 0.00338 | 0.029528 |
| SCRT2 | 11.07985 | -2.09721 | 0.636365 | -3.29561 | 0.000982 | 0.003381 | 0.23371 |
| IL4R | 657.3113 | -0.47944 | 0.145515 | -3.29476 | 0.000985 | 0.00339 | 0.717258 |
| CLSTN3 | 1065.435 | -0.48604 | 0.147674 | -3.29128 | 0.000997 | 0.003431 | 0.713984 |
| SUN2 | 453.1407 | -0.50247 | 0.152788 | -3.28869 | 0.001007 | 0.003458 | 0.705896 |
| MED10 | 495.2014 | -0.46824 | 0.142392 | -3.28838 | 0.001008 | 0.003462 | 0.722847 |
| PIDD1 | 829.7202 | -0.84203 | 0.256208 | -3.2865 | 0.001014 | 0.003484 | 0.557859 |
| TNFSF9 | 27.43717 | -1.44135 | 0.438613 | -3.28615 | 0.001016 | 0.003487 | 0.368223 |
| DPM3 | 218.8809 | -0.61516 | 0.187206 | -3.28603 | 0.001016 | 0.003488 | 0.652855 |
| SRPK3 | 28.50316 | -1.99737 | 0.608437 | -3.28279 | 0.001028 | 0.003524 | 0.250456 |
| TMEM179E | 391.1017 | -0.51271 | 0.156227 | -3.28185 | 0.001031 | 0.003534 | 0.700902 |

| | | | | | | | |
|---|---|---|---|---|---|---|---|
| IKZF3 | 48.83759 | -1.11849 | 0.340823 | -3.28173 | 0.001032 | 0.003534 | 0.460577 |
| VAV3 | 200.9214 | -0.75111 | 0.228939 | -3.28081 | 0.001035 | 0.003545 | 0.594148 |
| PLA2G4B | 225.0047 | -1.19946 | 0.365673 | -3.28015 | 0.001038 | 0.003551 | 0.435438 |
| SLC39A8 | 248.9488 | -0.51408 | 0.156747 | -3.27967 | 0.001039 | 0.003555 | 0.70024 |
| PPP2R3B | 512.6212 | -0.73175 | 0.223156 | -3.2791 | 0.001041 | 0.00356 | 0.602173 |
| VAMP2 | 385.1126 | -0.43191 | 0.131782 | -3.27747 | 0.001047 | 0.00358 | 0.741279 |
| ACSF2 | 330.9056 | -0.8777 | 0.268075 | -3.2741 | 0.00106 | 0.003618 | 0.544234 |
| ITGAX | 19.81631 | -1.47928 | 0.451858 | -3.27378 | 0.001061 | 0.003621 | 0.358667 |
| ZNF697 | 631.4073 | -0.51024 | 0.155996 | -3.27087 | 0.001072 | 0.003655 | 0.702104 |
| STAC | 164.9486 | -0.6395 | 0.1956 | -3.26942 | 0.001078 | 0.00367 | 0.641937 |
| TSNARE1 | 370.2132 | -0.69383 | 0.212225 | -3.26931 | 0.001078 | 0.003671 | 0.618211 |
| REC8 | 40.39019 | -1.735 | 0.530996 | -3.26744 | 0.001085 | 0.003694 | 0.300409 |
| TRAF3IP2 | 258.0101 | -0.57528 | 0.176093 | -3.26688 | 0.001087 | 0.0037 | 0.671158 |
| LIPH | 17.271 | -1.64091 | 0.502595 | -3.26488 | 0.001095 | 0.003723 | 0.320654 |
| AC007546 | 76.33244 | -0.87911 | 0.269302 | -3.26441 | 0.001097 | 0.003728 | 0.543702 |
| GFOD1 | 218.6347 | -0.67685 | 0.207373 | -3.26394 | 0.001099 | 0.003731 | 0.625529 |
| TJP2 | 3070.343 | -0.47023 | 0.144079 | -3.26372 | 0.0011 | 0.003734 | 0.721848 |
| CCDC3 | 42.84758 | -1.15954 | 0.355333 | -3.26327 | 0.001101 | 0.003739 | 0.447654 |
| KCND2 | 220.7649 | -0.69083 | 0.211851 | -3.26092 | 0.001111 | 0.003767 | 0.619498 |
| IL17RE | 62.58472 | -0.91734 | 0.281361 | -3.26038 | 0.001113 | 0.003774 | 0.529483 |
| FGFR3 | 2387.631 | -0.76707 | 0.235283 | -3.26019 | 0.001113 | 0.003775 | 0.587611 |
| AC073585 | 11.61153 | -2.88978 | 0.886589 | -3.25943 | 0.001116 | 0.003784 | 0.134925 |
| ABCA2 | 1086.819 | -0.69888 | 0.214482 | -3.25848 | 0.00112 | 0.003796 | 0.616048 |
| CIB1 | 393.4999 | -0.44421 | 0.136387 | -3.25697 | 0.001126 | 0.003815 | 0.734987 |
| ACP6 | 630.0423 | -0.46203 | 0.142052 | -3.25257 | 0.001144 | 0.003868 | 0.725962 |
| PLK3 | 144.4279 | -0.73871 | 0.227121 | -3.25247 | 0.001144 | 0.003869 | 0.599277 |
| TRIM72 | 110.3163 | -1.19821 | 0.368929 | -3.2478 | 0.001163 | 0.003928 | 0.435816 |
| BRICD5 | 127.3908 | -1.31672 | 0.405535 | -3.24687 | 0.001167 | 0.003938 | 0.401447 |
| SULT1A2 | 12.49703 | -2.03514 | 0.62701 | -3.24578 | 0.001171 | 0.003951 | 0.243985 |
| ABCA7 | 918.816 | -1.40637 | 0.433671 | -3.24293 | 0.001183 | 0.003986 | 0.377261 |
| DOCK2 | 30.53987 | -4.59564 | 1.417124 | -3.24293 | 0.001183 | 0.003986 | 0.04136 |
| AQP5 | 18.93584 | -1.41976 | 0.438105 | -3.24069 | 0.001192 | 0.004014 | 0.373774 |
| FAS | 279.0702 | -0.50256 | 0.155095 | -3.2403 | 0.001194 | 0.004019 | 0.705855 |
| SLC6A16 | 25.30786 | -1.81209 | 0.559615 | -3.2381 | 0.001203 | 0.004045 | 0.284778 |
| GRID2IP | 44.15329 | -0.92168 | 0.284643 | -3.23804 | 0.001204 | 0.004045 | 0.527892 |
| PPARA | 262.1389 | -0.70594 | 0.218031 | -3.23778 | 0.001205 | 0.004048 | 0.613045 |
| PAWR | 4776.79 | -0.52366 | 0.162065 | -3.23118 | 0.001233 | 0.004135 | 0.695604 |
| FAM228B | 193.9291 | -0.58941 | 0.182464 | -3.23025 | 0.001237 | 0.004147 | 0.664616 |
| AL162739. | 5.182388 | -3.68298 | 1.140315 | -3.22979 | 0.001239 | 0.004153 | 0.07786 |
| RTEL1 | 768.5872 | -0.75454 | 0.233788 | -3.22743 | 0.001249 | 0.004182 | 0.592737 |
| CPT1C | 1297.957 | -0.5454 | 0.169057 | -3.22614 | 0.001255 | 0.004199 | 0.6852 |
| C1QL1 | 123.8078 | -0.65547 | 0.20338 | -3.22289 | 0.001269 | 0.004243 | 0.634867 |
| TEPSIN | 433.3785 | -0.5919 | 0.183676 | -3.2225 | 0.001271 | 0.004248 | 0.66347 |
| DHX58 | 43.94081 | -1.22929 | 0.381748 | -3.22017 | 0.001281 | 0.004278 | 0.426527 |

| | | | | | | | |
|---|---|---|---|---|---|---|---|
| MDFIC | 311.3665 | -0.49988 | 0.155234 | -3.22018 | 0.001281 | 0.004278 | 0.707165 |
| MT-ND6 | 15887.53 | -1.20943 | 0.375667 | -3.21942 | 0.001285 | 0.004288 | 0.432439 |
| NOTO | 4.747913 | -4.19378 | 1.302954 | -3.21867 | 0.001288 | 0.004297 | 0.054644 |
| WNT10B | 93.70642 | -0.66052 | 0.205225 | -3.2185 | 0.001289 | 0.004298 | 0.632652 |
| CHAC2 | 146.4659 | -0.61486 | 0.191057 | -3.21819 | 0.00129 | 0.004302 | 0.652995 |
| LTB4R2 | 154.1567 | -0.62274 | 0.193525 | -3.2179 | 0.001291 | 0.004306 | 0.649435 |
| GALR1 | 7.929941 | -2.65024 | 0.824036 | -3.21616 | 0.001299 | 0.00433 | 0.159294 |
| NAPRT | 816.5456 | -1.32009 | 0.410526 | -3.2156 | 0.001302 | 0.004337 | 0.400511 |
| ABHD14A- | 24.59157 | -1.88216 | 0.585468 | -3.2148 | 0.001305 | 0.004348 | 0.271276 |
| SLC25A29 | 3078.593 | -0.48109 | 0.149769 | -3.21222 | 0.001317 | 0.004382 | 0.716437 |
| OR9H1P | 19.87072 | -1.35253 | 0.421146 | -3.21155 | 0.00132 | 0.004389 | 0.391605 |
| TMEM268 | 598.4153 | -0.491 | 0.15289 | -3.21145 | 0.001321 | 0.00439 | 0.711533 |
| MOGS | 1738.733 | -0.51785 | 0.161267 | -3.21113 | 0.001322 | 0.004394 | 0.698412 |
| ACADS | 189.5229 | -0.57382 | 0.178715 | -3.21081 | 0.001324 | 0.004398 | 0.671835 |
| TRMU | 1021.623 | -0.48683 | 0.151635 | -3.21051 | 0.001325 | 0.004402 | 0.713593 |
| CST3 | 574.8121 | -0.45751 | 0.142534 | -3.20981 | 0.001328 | 0.00441 | 0.728244 |
| MPZL2 | 39.53962 | -1.04895 | 0.32704 | -3.20742 | 0.001339 | 0.004446 | 0.483318 |
| SCNN1B | 6.456091 | -3.31442 | 1.033623 | -3.20661 | 0.001343 | 0.004457 | 0.100522 |
| FAM162B | 23.66715 | -1.45191 | 0.452835 | -3.20627 | 0.001345 | 0.004461 | 0.365537 |
| AC012368 | 3.503268 | -4.50848 | 1.406661 | -3.20509 | 0.00135 | 0.004478 | 0.043935 |
| RNF31 | 1135.98 | -0.58644 | 0.183264 | -3.2 | 0.001374 | 0.004551 | 0.665982 |
| MUC19 | 29.44204 | -3.17979 | 0.993905 | -3.19929 | 0.001378 | 0.00456 | 0.110354 |
| AGFG2 | 160.724 | -0.74597 | 0.233373 | -3.19649 | 0.001391 | 0.004602 | 0.596266 |
| N4BP2L1 | 77.00127 | -0.82084 | 0.25682 | -3.19618 | 0.001393 | 0.004604 | 0.566112 |
| HTRA3 | 157.9028 | -0.62865 | 0.196731 | -3.19548 | 0.001396 | 0.004613 | 0.646781 |
| TMPRSS6 | 5.992629 | -2.82582 | 0.884328 | -3.19544 | 0.001396 | 0.004613 | 0.141041 |
| AC139491 | 17.09019 | -2.48667 | 0.778587 | -3.19382 | 0.001404 | 0.004636 | 0.178418 |
| MICALL2 | 327.0116 | -1.11864 | 0.350367 | -3.19277 | 0.001409 | 0.004652 | 0.460526 |
| CD36 | 13.97837 | -2.47602 | 0.775704 | -3.19197 | 0.001413 | 0.004664 | 0.17974 |
| SLC26A11 | 535.0421 | -0.5609 | 0.175744 | -3.19157 | 0.001415 | 0.00467 | 0.67788 |
| GTF2IRD1P | 2.766814 | -4.92998 | 1.544765 | -3.19141 | 0.001416 | 0.004672 | 0.032804 |
| TOR2A | 378.6409 | -0.47391 | 0.148543 | -3.19042 | 0.001421 | 0.004687 | 0.720009 |
| HMOX1 | 459.1419 | -0.42583 | 0.133518 | -3.18934 | 0.001426 | 0.004704 | 0.744409 |
| ERICH5 | 27.375 | -1.36037 | 0.426557 | -3.18919 | 0.001427 | 0.004705 | 0.389482 |
| OPTC | 82.39483 | -0.87488 | 0.274473 | -3.18751 | 0.001435 | 0.004732 | 0.545298 |
| CXCL14 | 22.40805 | -1.45157 | 0.45565 | -3.18572 | 0.001444 | 0.00476 | 0.365623 |
| TMED9 | 3293.535 | -0.60776 | 0.190867 | -3.18419 | 0.001452 | 0.004784 | 0.656216 |
| PHLDA3 | 408.1088 | -0.4658 | 0.146315 | -3.1835 | 0.001455 | 0.004793 | 0.724072 |
| PCSK2 | 49.3736 | -1.25947 | 0.396212 | -3.17878 | 0.001479 | 0.004868 | 0.417698 |
| NXNL2 | 14.8406 | -1.85258 | 0.583031 | -3.17751 | 0.001485 | 0.004888 | 0.276896 |
| PPM1K | 271.652 | -0.59901 | 0.188578 | -3.17644 | 0.001491 | 0.004903 | 0.660209 |
| ITGAM | 45.74812 | -1.89114 | 0.595928 | -3.17343 | 0.001507 | 0.00495 | 0.269595 |
| CCDC154 | 34.86887 | -1.81848 | 0.573123 | -3.17294 | 0.001509 | 0.004957 | 0.283519 |
| SDF4 | 1949.387 | -0.48369 | 0.152465 | -3.17244 | 0.001512 | 0.004965 | 0.715148 |

| | | | | | | | |
|---|---|---|---|---|---|---|---|
| PPP1R16A | 658.7131 | -0.6541 | 0.206233 | -3.17165 | 0.001516 | 0.004977 | 0.635473 |
| CCNA1 | 20.78378 | -1.42943 | 0.451074 | -3.16894 | 0.00153 | 0.005019 | 0.371279 |
| CSF2RA | 56.04068 | -0.88875 | 0.280632 | -3.16698 | 0.00154 | 0.005048 | 0.54008 |
| TEX22 | 25.07292 | -1.5176 | 0.479398 | -3.16564 | 0.001547 | 0.005069 | 0.349265 |
| APLN | 177.6567 | -0.97622 | 0.30885 | -3.16082 | 0.001573 | 0.005143 | 0.508311 |
| SLC16A6 | 19.26689 | -1.48522 | 0.469959 | -3.16032 | 0.001576 | 0.00515 | 0.357194 |
| TTC22 | 11.50158 | -2.62244 | 0.829902 | -3.15993 | 0.001578 | 0.005156 | 0.162393 |
| TOGARAM2 | 107.6093 | -0.63014 | 0.199444 | -3.1595 | 0.00158 | 0.005162 | 0.646112 |
| PRADC1 | 135.4312 | -0.62031 | 0.196497 | -3.15687 | 0.001595 | 0.005206 | 0.650529 |
| AC139530 | 16.54691 | -2.24743 | 0.712657 | -3.15359 | 0.001613 | 0.005261 | 0.210599 |
| TVP23C | 136.9638 | -0.77626 | 0.246209 | -3.15283 | 0.001617 | 0.005274 | 0.58388 |
| AC087289 | 112.1888 | -1.12657 | 0.357411 | -3.15205 | 0.001621 | 0.005287 | 0.458002 |
| FAM98C | 174.2756 | -0.65403 | 0.207593 | -3.15055 | 0.00163 | 0.005311 | 0.635501 |
| PLEKHG5 | 699.6087 | -0.50328 | 0.159778 | -3.14986 | 0.001633 | 0.005323 | 0.705502 |
| PLCXD3 | 6.462895 | -2.97526 | 0.944731 | -3.14932 | 0.001636 | 0.005331 | 0.127162 |
| RANBP17 | 275.5523 | -0.65426 | 0.207755 | -3.14921 | 0.001637 | 0.005332 | 0.635399 |
| AC011448 | 74.75277 | -1.12959 | 0.358873 | -3.14759 | 0.001646 | 0.005357 | 0.457047 |
| PRSS50 | 73.42112 | -1.21897 | 0.387293 | -3.14742 | 0.001647 | 0.005359 | 0.429588 |
| KCNJ5 | 10.22901 | -1.85371 | 0.589137 | -3.14648 | 0.001652 | 0.005374 | 0.276681 |
| AIFM3 | 52.58546 | -0.91886 | 0.292038 | -3.14638 | 0.001653 | 0.005375 | 0.528925 |
| RAMP1 | 10.08495 | -2.46552 | 0.783966 | -3.14493 | 0.001661 | 0.005399 | 0.181053 |
| TRIM56 | 1119.479 | -0.45148 | 0.143605 | -3.14392 | 0.001667 | 0.005417 | 0.731291 |
| FBN3 | 2021.065 | -0.72787 | 0.231607 | -3.14269 | 0.001674 | 0.005439 | 0.603795 |
| PPP1R15A | 516.1609 | -0.44061 | 0.140246 | -3.14168 | 0.00168 | 0.005454 | 0.736823 |
| FAM193B | 1365.453 | -0.67972 | 0.216364 | -3.14157 | 0.00168 | 0.005455 | 0.624285 |
| BNIPL | 62.57034 | -1.21162 | 0.385933 | -3.13946 | 0.001693 | 0.005492 | 0.431783 |
| KCNJ4 | 76.37455 | -0.76569 | 0.243933 | -3.13895 | 0.001696 | 0.0055 | 0.58817 |
| LTO1 | 374.5772 | -0.50041 | 0.159471 | -3.1379 | 0.001702 | 0.005518 | 0.706908 |
| POMT1 | 1102.789 | -1.11755 | 0.356207 | -3.13735 | 0.001705 | 0.005527 | 0.460877 |
| CLPTM1 | 2100.328 | -0.41809 | 0.133285 | -3.13679 | 0.001708 | 0.005536 | 0.748417 |
| PITPNC1 | 595.9708 | -0.56724 | 0.180885 | -3.13592 | 0.001713 | 0.00555 | 0.674907 |
| GLI1 | 316.5888 | -0.69985 | 0.223177 | -3.13583 | 0.001714 | 0.005551 | 0.615638 |
| C17orf107 | 20.61661 | -1.34368 | 0.428518 | -3.13565 | 0.001715 | 0.005553 | 0.394013 |
| MAN1C1 | 406.8426 | -0.54029 | 0.172375 | -3.13439 | 0.001722 | 0.005573 | 0.687632 |
| SULT1A3 | 302.9457 | -1.77886 | 0.567783 | -3.13299 | 0.00173 | 0.005598 | 0.291414 |
| SOX7 | 5.303936 | -5.86277 | 1.871924 | -3.13195 | 0.001736 | 0.005617 | 0.017184 |
| ABCC10 | 759.6977 | -0.61342 | 0.196224 | -3.12612 | 0.001771 | 0.005718 | 0.653645 |
| POMT2 | 852.8103 | -0.63738 | 0.203931 | -3.12548 | 0.001775 | 0.005728 | 0.642879 |
| RCCD1 | 618.5589 | -0.47224 | 0.15109 | -3.12554 | 0.001775 | 0.005728 | 0.720845 |
| AL390719. | 48.17905 | -1.11024 | 0.355296 | -3.12482 | 0.001779 | 0.00574 | 0.463218 |
| OSTF1 | 186.245 | -0.61887 | 0.19838 | -3.11963 | 0.001811 | 0.005832 | 0.65118 |
| AL627309. | 8.598134 | -2.26666 | 0.726877 | -3.11836 | 0.001819 | 0.005857 | 0.20781 |
| CFAP300 | 5.063176 | -4.66736 | 1.49707 | -3.11766 | 0.001823 | 0.005869 | 0.039353 |
| AMDHD1 | 80.72528 | -0.84816 | 0.27207 | -3.11743 | 0.001824 | 0.005873 | 0.555493 |

| | | | | | | | |
|---|---|---|---|---|---|---|---|
| PI4KAP2 | 483.5475 | -0.65788 | 0.21113 | -3.11598 | 0.001833 | 0.0059 | 0.63381 |
| DHX34 | 827.9515 | -0.48164 | 0.15469 | -3.11356 | 0.001848 | 0.005947 | 0.716165 |
| GOLGA6L5 | 65.18048 | -1.35864 | 0.436404 | -3.11326 | 0.00185 | 0.005948 | 0.38995 |
| OCEL1 | 171.7665 | -0.53666 | 0.172429 | -3.11237 | 0.001856 | 0.005963 | 0.689364 |
| CD79B | 26.41556 | -1.19396 | 0.38366 | -3.11203 | 0.001858 | 0.005969 | 0.437101 |
| CLIC6 | 221.8539 | -0.68585 | 0.220398 | -3.11189 | 0.001859 | 0.005971 | 0.621638 |
| OBSCN | 2595.847 | -0.66798 | 0.214663 | -3.11174 | 0.00186 | 0.005973 | 0.629389 |
| NAP1L5 | 98.76354 | -0.71743 | 0.230602 | -3.1111 | 0.001864 | 0.005985 | 0.608182 |
| ACTN2 | 53.5932 | -1.58252 | 0.509072 | -3.10863 | 0.00188 | 0.00603 | 0.333898 |
| MYT1L | 35.06829 | -1.24436 | 0.400336 | -3.10829 | 0.001882 | 0.006035 | 0.422095 |
| TMEM115 | 626.5007 | -0.42242 | 0.135949 | -3.10717 | 0.001889 | 0.006056 | 0.746174 |
| TBC1D2 | 92.86371 | -0.72518 | 0.233404 | -3.10696 | 0.00189 | 0.006059 | 0.604923 |
| SLC25A34 | 33.92891 | -1.71488 | 0.551982 | -3.10677 | 0.001891 | 0.00606 | 0.304628 |
| PDE6A | 39.50244 | -0.94782 | 0.305163 | -3.10595 | 0.001897 | 0.006073 | 0.518415 |
| MBD6 | 887.0206 | -0.58059 | 0.18706 | -3.10378 | 0.001911 | 0.006114 | 0.668689 |
| POSTN | 12.58157 | -1.80582 | 0.582122 | -3.10213 | 0.001921 | 0.006145 | 0.286018 |
| ASPSCR1 | 887.1156 | -0.52489 | 0.169328 | -3.09984 | 0.001936 | 0.006188 | 0.695013 |
| LAG3 | 62.18316 | -1.0358 | 0.334352 | -3.09794 | 0.001949 | 0.006222 | 0.487745 |
| CCN6 | 6.991654 | -2.60908 | 0.842507 | -3.09681 | 0.001956 | 0.006242 | 0.163903 |
| IL11RA | 99.84922 | -0.61977 | 0.200145 | -3.09658 | 0.001958 | 0.006244 | 0.650777 |
| SLC12A4 | 808.6831 | -0.4741 | 0.153176 | -3.0951 | 0.001967 | 0.006272 | 0.719918 |
| SYT3 | 412.2651 | -0.62004 | 0.200409 | -3.09386 | 0.001976 | 0.006294 | 0.650655 |
| ARHGEF19 | 1118.568 | -0.46942 | 0.15187 | -3.09091 | 0.001995 | 0.006352 | 0.722257 |
| NPIPA8 | 13.83832 | -2.49824 | 0.808473 | -3.09007 | 0.002001 | 0.006366 | 0.176993 |
| GRHL2 | 599.0212 | -0.70975 | 0.229707 | -3.0898 | 0.002003 | 0.00637 | 0.611427 |
| NECTIN4 | 4.950137 | -3.61376 | 1.169927 | -3.08887 | 0.002009 | 0.006386 | 0.081687 |
| ARSE | 55.38615 | -1.1164 | 0.361456 | -3.08863 | 0.002011 | 0.00639 | 0.461242 |
| HEXD | 630.9213 | -0.60262 | 0.195206 | -3.08708 | 0.002021 | 0.006422 | 0.658559 |
| LRP2BP | 58.27745 | -1.13364 | 0.36725 | -3.08684 | 0.002023 | 0.006426 | 0.455764 |
| GADL1 | 25.04962 | -1.06253 | 0.34424 | -3.08659 | 0.002025 | 0.00643 | 0.478792 |
| AP000781. | 9.303348 | -2.47518 | 0.802032 | -3.08614 | 0.002028 | 0.006439 | 0.179844 |
| STIM2 | 539.9183 | -0.46393 | 0.150342 | -3.08585 | 0.00203 | 0.006444 | 0.725007 |
| ASTL | 9.156644 | -2.83204 | 0.917905 | -3.08533 | 0.002033 | 0.006452 | 0.140434 |
| CEACAM1 | 40.31272 | -1.35667 | 0.439779 | -3.08489 | 0.002036 | 0.00646 | 0.390483 |
| CMTM7 | 425.6947 | -0.47148 | 0.15286 | -3.08443 | 0.002039 | 0.006469 | 0.721222 |
| RCE1 | 664.2243 | -0.46738 | 0.151565 | -3.08366 | 0.002045 | 0.006484 | 0.723279 |
| ARHGAP30 | 10.80966 | -1.70144 | 0.55178 | -3.08356 | 0.002045 | 0.006485 | 0.307478 |
| ADCY4 | 22.6192 | -1.66205 | 0.539039 | -3.08335 | 0.002047 | 0.006488 | 0.31599 |
| SCAND2P | 198.0898 | -0.76133 | 0.24699 | -3.08244 | 0.002053 | 0.006506 | 0.589951 |
| TMEM178A | 166.7817 | -0.70653 | 0.22927 | -3.08166 | 0.002058 | 0.006521 | 0.612791 |
| BTN2A3P | 73.98527 | -0.79585 | 0.258322 | -3.08085 | 0.002064 | 0.006536 | 0.576003 |
| PYDC1 | 28.7945 | -1.57325 | 0.510872 | -3.07953 | 0.002073 | 0.006564 | 0.336051 |
| PROKR2 | 15.18035 | -1.69585 | 0.55075 | -3.07918 | 0.002076 | 0.00657 | 0.308672 |
| PMP22 | 109.5867 | -0.60391 | 0.196206 | -3.07791 | 0.002085 | 0.006596 | 0.65797 |

| | | | | | | | |
|---|---|---|---|---|---|---|---|
| FGF18 | 21.44508 | -1.79731 | 0.584532 | -3.07479 | 0.002107 | 0.006661 | 0.28771 |
| IL9RP3 | 14.2848 | -2.21918 | 0.721781 | -3.07459 | 0.002108 | 0.006665 | 0.214764 |
| SELENOP | 368.5347 | -0.6204 | 0.201794 | -3.07444 | 0.002109 | 0.006667 | 0.650489 |
| AC022400 | 195.6041 | -0.93381 | 0.303797 | -3.07378 | 0.002114 | 0.006678 | 0.523475 |
| SERPINB1 | 338.5417 | -0.48683 | 0.158394 | -3.07353 | 0.002115 | 0.006683 | 0.713592 |
| F12 | 57.35914 | -1.21911 | 0.396859 | -3.0719 | 0.002127 | 0.006716 | 0.429548 |
| SPATA2L | 386.5347 | -0.4573 | 0.148904 | -3.07113 | 0.002133 | 0.00673 | 0.728346 |
| DIPK1A | 152.2695 | -0.68645 | 0.223621 | -3.06969 | 0.002143 | 0.00676 | 0.621382 |
| COL4A4 | 31.41301 | -1.42116 | 0.463099 | -3.06879 | 0.002149 | 0.006778 | 0.373413 |
| C8orf58 | 287.8563 | -0.47034 | 0.153331 | -3.0675 | 0.002159 | 0.006802 | 0.721793 |
| KCNJ10 | 28.84658 | -1.5614 | 0.509026 | -3.06743 | 0.002159 | 0.006802 | 0.338822 |
| SGSM3 | 1613.807 | -0.69723 | 0.227303 | -3.06742 | 0.002159 | 0.006802 | 0.616754 |
| SLITRK3 | 51.87225 | -1.23931 | 0.404061 | -3.06714 | 0.002161 | 0.006804 | 0.423575 |
| TUBB8P1 | 2.513366 | -4.78488 | 1.560059 | -3.06711 | 0.002161 | 0.006804 | 0.036275 |
| KCNB2 | 14.58224 | -1.6712 | 0.54492 | -3.06687 | 0.002163 | 0.006809 | 0.313992 |
| DOK2 | 6.247787 | -3.78403 | 1.235153 | -3.06362 | 0.002187 | 0.006876 | 0.072593 |
| PTAFR | 67.12638 | -0.83082 | 0.271283 | -3.06255 | 0.002195 | 0.0069 | 0.562211 |
| CITED4 | 178.9142 | -0.53831 | 0.175781 | -3.06238 | 0.002196 | 0.006902 | 0.688578 |
| GOLGA6D | 24.64455 | -1.31794 | 0.43071 | -3.05993 | 0.002214 | 0.006953 | 0.401106 |
| SCN3B | 36.61319 | -1.29363 | 0.422766 | -3.05991 | 0.002214 | 0.006953 | 0.407924 |
| CLDN9 | 17.97738 | -1.36696 | 0.446777 | -3.05961 | 0.002216 | 0.006959 | 0.387707 |
| AC105339 | 3.582966 | -4.54965 | 1.48739 | -3.05881 | 0.002222 | 0.006975 | 0.042699 |
| AC099489 | 317.7289 | -0.63832 | 0.208925 | -3.05526 | 0.002249 | 0.007048 | 0.642461 |
| PNMT | 33.05005 | -1.31337 | 0.430107 | -3.05358 | 0.002261 | 0.007085 | 0.402381 |
| TRIM62 | 261.7205 | -0.86434 | 0.28306 | -3.05355 | 0.002262 | 0.007085 | 0.549299 |
| ZFYVE19 | 246.5235 | -0.45021 | 0.147451 | -3.05326 | 0.002264 | 0.00709 | 0.731938 |
| EXOC3L4 | 4.44776 | -3.75104 | 1.22908 | -3.05191 | 0.002274 | 0.007121 | 0.074272 |
| PCDHA8 | 19.00229 | -2.14886 | 0.704232 | -3.05136 | 0.002278 | 0.007133 | 0.22549 |
| LRFN1 | 193.6951 | -0.54072 | 0.177258 | -3.05047 | 0.002285 | 0.007151 | 0.687428 |
| DDR2 | 244.68 | -0.83945 | 0.275254 | -3.04973 | 0.00229 | 0.007163 | 0.558857 |
| GATD1 | 1345.28 | -0.48034 | 0.157509 | -3.0496 | 0.002291 | 0.007164 | 0.716809 |
| PRRG1 | 235.481 | -0.63795 | 0.20927 | -3.04846 | 0.0023 | 0.007187 | 0.642625 |
| TBC1D10A | 162.9835 | -0.44438 | 0.145789 | -3.04808 | 0.002303 | 0.007193 | 0.734902 |
| UBXN8 | 213.3041 | -0.43877 | 0.143981 | -3.04743 | 0.002308 | 0.007205 | 0.737763 |
| STXBP2 | 1253.3 | -0.45512 | 0.149519 | -3.04391 | 0.002335 | 0.007288 | 0.729448 |
| TAT | 9.104581 | -2.06958 | 0.679924 | -3.04385 | 0.002336 | 0.007288 | 0.238228 |
| FOXD1 | 23.33151 | -1.2014 | 0.394764 | -3.04334 | 0.00234 | 0.007298 | 0.434853 |
| LRRTM4 | 163.6097 | -0.4651 | 0.152884 | -3.04215 | 0.002349 | 0.007323 | 0.724422 |
| MYO1A | 6.307694 | -2.95974 | 0.973171 | -3.04134 | 0.002355 | 0.007339 | 0.128537 |
| C1orf116 | 68.98521 | -0.7904 | 0.25991 | -3.04106 | 0.002357 | 0.007344 | 0.578182 |
| NKD2 | 164.4149 | -0.61884 | 0.203603 | -3.03946 | 0.00237 | 0.00738 | 0.651193 |
| LPCAT1 | 3590.077 | -0.4999 | 0.164554 | -3.03788 | 0.002382 | 0.007414 | 0.707158 |
| SP140L | 36.52313 | -1.23156 | 0.405421 | -3.03773 | 0.002384 | 0.007416 | 0.425856 |
| MAPK8IP3 | 1626.351 | -0.44631 | 0.146952 | -3.0371 | 0.002389 | 0.007431 | 0.733919 |

| | | | | | | | |
|---|---|---|---|---|---|---|---|
| GPR82 | 13.33918 | -1.59709 | 0.525937 | -3.03666 | 0.002392 | 0.00744 | 0.330542 |
| PATL2 | 15.2326 | -2.09234 | 0.68918 | -3.03598 | 0.002398 | 0.007456 | 0.2345 |
| NACAD | 214.4901 | -0.65628 | 0.216213 | -3.03533 | 0.002403 | 0.007471 | 0.634514 |
| PPARGC1E | 389.9361 | -0.61649 | 0.20323 | -3.03348 | 0.002418 | 0.007514 | 0.652254 |
| TAPBP | 423.8059 | -0.67743 | 0.223386 | -3.03256 | 0.002425 | 0.007536 | 0.625277 |
| TTC39C | 278.9699 | -0.5082 | 0.167587 | -3.03247 | 0.002426 | 0.007537 | 0.703097 |
| FA2H | 33.31988 | -1.42922 | 0.471335 | -3.03228 | 0.002427 | 0.00754 | 0.371332 |
| AC100821 | 11.70465 | -2.2769 | 0.751248 | -3.03083 | 0.002439 | 0.007569 | 0.20634 |
| DSG3 | 13.82059 | -1.6165 | 0.533351 | -3.03083 | 0.002439 | 0.007569 | 0.326126 |
| ELL3 | 102.7865 | -0.7768 | 0.256301 | -3.03081 | 0.002439 | 0.007569 | 0.58366 |
| AOC2 | 73.65416 | -0.64447 | 0.2128 | -3.0285 | 0.002458 | 0.00762 | 0.63973 |
| PSMB10 | 181.1423 | -0.4522 | 0.149414 | -3.02651 | 0.002474 | 0.007667 | 0.730925 |
| TMEM80 | 199.0728 | -0.50331 | 0.166332 | -3.02591 | 0.002479 | 0.00768 | 0.705489 |
| FRK | 190.8375 | -0.4166 | 0.137706 | -3.02531 | 0.002484 | 0.007693 | 0.749186 |
| CKMT2 | 19.43991 | -1.59322 | 0.527101 | -3.02262 | 0.002506 | 0.007755 | 0.33143 |
| LRRC8E | 98.95807 | -0.85494 | 0.28289 | -3.02217 | 0.00251 | 0.007765 | 0.552888 |
| ABCB9 | 133.0621 | -0.82533 | 0.273218 | -3.02077 | 0.002521 | 0.007799 | 0.564354 |
| RNASET2 | 398.552 | -0.72705 | 0.240768 | -3.01971 | 0.00253 | 0.007822 | 0.604138 |
| MFSD13A | 235.5838 | -0.50507 | 0.167323 | -3.0185 | 0.00254 | 0.007851 | 0.704628 |
| AP000346. | 84.59489 | -0.96129 | 0.318515 | -3.01804 | 0.002544 | 0.007861 | 0.513596 |
| GAL3ST1 | 209.812 | -0.48104 | 0.159456 | -3.01675 | 0.002555 | 0.007894 | 0.716462 |
| MYH7B | 33.9359 | -1.00928 | 0.334576 | -3.0166 | 0.002556 | 0.007896 | 0.496793 |
| ST3GAL4 | 240.4005 | -0.42904 | 0.142249 | -3.01614 | 0.00256 | 0.007905 | 0.742754 |
| LINC0152 | 25.73108 | -1.45358 | 0.482139 | -3.01485 | 0.002571 | 0.007936 | 0.365115 |
| CR1 | 8.793904 | -2.57445 | 0.854173 | -3.01397 | 0.002579 | 0.007958 | 0.167886 |
| NLGN2 | 3689.273 | -0.58365 | 0.193655 | -3.01389 | 0.002579 | 0.007959 | 0.667271 |
| PRR5-ARH | 24.03367 | -1.69968 | 0.56401 | -3.01356 | 0.002582 | 0.007966 | 0.307855 |
| MVD | 916.8956 | -0.46505 | 0.154406 | -3.01186 | 0.002597 | 0.00801 | 0.724446 |
| RRNAD1 | 492.7829 | -0.59267 | 0.196845 | -3.01084 | 0.002605 | 0.008035 | 0.663115 |
| STRIT1 | 3.331352 | -4.42027 | 1.469706 | -3.00759 | 0.002633 | 0.008113 | 0.046705 |
| PTPN18 | 329.1264 | -0.43795 | 0.145715 | -3.0055 | 0.002651 | 0.008165 | 0.738184 |
| C15orf53 | 13.25753 | -2.42889 | 0.809093 | -3.00199 | 0.002682 | 0.008248 | 0.185709 |
| CDH15 | 8.486239 | -2.51198 | 0.836983 | -3.00123 | 0.002689 | 0.008267 | 0.175315 |
| MISP3 | 33.22325 | -1.21934 | 0.406513 | -2.99951 | 0.002704 | 0.008311 | 0.429479 |
| NDUFA3 | 252.8586 | -0.42844 | 0.142853 | -2.9992 | 0.002707 | 0.008315 | 0.743062 |
| MUSTN1 | 11.60006 | -2.14311 | 0.715003 | -2.99734 | 0.002723 | 0.008361 | 0.226392 |
| SLC48A1 | 341.4335 | -0.43974 | 0.14671 | -2.99734 | 0.002723 | 0.008361 | 0.737267 |
| RAB3IL1 | 253.0273 | -0.42922 | 0.143226 | -2.99679 | 0.002728 | 0.008371 | 0.742665 |
| SP140 | 8.207097 | -2.75986 | 0.921194 | -2.99596 | 0.002736 | 0.008393 | 0.147638 |
| ADCY5 | 315.6643 | -1.67285 | 0.558389 | -2.99585 | 0.002737 | 0.008395 | 0.313633 |
| EBP | 1196.714 | -0.49244 | 0.164381 | -2.99575 | 0.002738 | 0.008396 | 0.71082 |
| LENG8 | 2075.987 | -0.54321 | 0.18137 | -2.99502 | 0.002744 | 0.008413 | 0.686243 |
| GPR39 | 21.35468 | -1.32586 | 0.442889 | -2.99366 | 0.002756 | 0.008449 | 0.398911 |
| ANKRD22 | 8.107825 | -2.47821 | 0.827885 | -2.99342 | 0.002759 | 0.008455 | 0.179468 |

| | | | | | | | |
|---|---|---|---|---|---|---|---|
| KREMEN1 | 177.7943 | -1.01659 | 0.339718 | -2.99246 | 0.002767 | 0.008479 | 0.494283 |
| MCOLN1 | 406.3732 | -0.62553 | 0.209229 | -2.98967 | 0.002793 | 0.008551 | 0.648183 |
| LMTK3 | 368.9041 | -0.62074 | 0.207637 | -2.98957 | 0.002794 | 0.008551 | 0.650335 |
| ZFYVE28 | 71.3068 | -1.25221 | 0.418904 | -2.98925 | 0.002797 | 0.008556 | 0.419805 |
| FLVCR2 | 273.066 | -0.66219 | 0.221559 | -2.98878 | 0.002801 | 0.008564 | 0.631918 |
| NDUFAF5 | 210.3126 | -0.48575 | 0.162682 | -2.98587 | 0.002828 | 0.008635 | 0.714127 |
| FAM219A | 450.1679 | -0.5003 | 0.167574 | -2.98553 | 0.002831 | 0.008643 | 0.706962 |
| SLC25A20 | 119.3995 | -0.56379 | 0.188932 | -2.98407 | 0.002844 | 0.008683 | 0.676524 |
| TJP3 | 524.6284 | -0.49187 | 0.164943 | -2.98205 | 0.002863 | 0.008733 | 0.711105 |
| LHFPL5 | 109.905 | -0.73773 | 0.247423 | -2.98168 | 0.002867 | 0.008739 | 0.599681 |
| CEBPB | 1343.095 | -0.53155 | 0.178363 | -2.98013 | 0.002881 | 0.008782 | 0.691813 |
| GCK | 43.40912 | -0.90809 | 0.304747 | -2.97981 | 0.002884 | 0.008788 | 0.532891 |
| WNT5A | 427.3379 | -0.47234 | 0.15852 | -2.97971 | 0.002885 | 0.00879 | 0.720792 |
| KAT2A | 1378.292 | -0.55821 | 0.187348 | -2.97955 | 0.002887 | 0.008791 | 0.679143 |
| SPTB | 213.6977 | -0.53679 | 0.180178 | -2.97922 | 0.00289 | 0.008796 | 0.689303 |
| SCN9A | 136.6175 | -0.64778 | 0.217507 | -2.97822 | 0.002899 | 0.008821 | 0.63826 |
| SLC25A45 | 21.90076 | -1.51545 | 0.508864 | -2.97811 | 0.0029 | 0.008823 | 0.349788 |
| GCNA | 31.18582 | -1.10731 | 0.371868 | -2.97771 | 0.002904 | 0.008833 | 0.464157 |
| IRF6 | 295.0574 | -0.5469 | 0.183716 | -2.97685 | 0.002912 | 0.008854 | 0.684491 |
| CLDN15 | 111.1135 | -0.58244 | 0.195777 | -2.97501 | 0.00293 | 0.008903 | 0.667834 |
| FAM110D | 39.20301 | -0.93901 | 0.31609 | -2.97072 | 0.002971 | 0.009017 | 0.521589 |
| NOXA1 | 71.4323 | -0.95281 | 0.320864 | -2.96951 | 0.002983 | 0.009051 | 0.516626 |
| ANPEP | 80.97868 | -1.07525 | 0.362193 | -2.96873 | 0.00299 | 0.009069 | 0.474588 |
| SLC15A2 | 178.1625 | -1.11578 | 0.375839 | -2.96877 | 0.00299 | 0.009069 | 0.461443 |
| SLC44A3 | 60.29568 | -0.85768 | 0.288937 | -2.96841 | 0.002993 | 0.009077 | 0.551839 |
| MCF2 | 33.26447 | -1.17411 | 0.395623 | -2.96776 | 0.003 | 0.009093 | 0.443156 |
| MOGAT3 | 6.627291 | -2.78265 | 0.937638 | -2.96772 | 0.003 | 0.009093 | 0.145324 |
| PLA2G2A | 24.65933 | -1.33604 | 0.450343 | -2.96672 | 0.00301 | 0.009121 | 0.396107 |
| DENND2C | 163.1465 | -0.58337 | 0.196652 | -2.96652 | 0.003012 | 0.009126 | 0.667403 |
| MIIP | 1221.022 | -0.42465 | 0.143238 | -2.96461 | 0.003031 | 0.009179 | 0.745022 |
| MFSD12 | 1080.116 | -0.45645 | 0.153981 | -2.96433 | 0.003033 | 0.009186 | 0.728777 |
| WIPI1 | 332.5269 | -0.44146 | 0.148996 | -2.96287 | 0.003048 | 0.009225 | 0.736392 |
| SLC16A11 | 8.141519 | -2.76562 | 0.933908 | -2.96135 | 0.003063 | 0.009267 | 0.14705 |
| C1QL4 | 53.89348 | -0.95162 | 0.321394 | -2.96092 | 0.003067 | 0.009278 | 0.517051 |
| DUSP2 | 130.0299 | -0.56724 | 0.191587 | -2.96073 | 0.003069 | 0.009282 | 0.674907 |
| ELN | 239.0839 | -0.70752 | 0.239035 | -2.95989 | 0.003077 | 0.009304 | 0.612374 |
| SLC24A2 | 79.43576 | -0.90774 | 0.306852 | -2.95822 | 0.003094 | 0.009346 | 0.53302 |
| OXER1 | 45.97105 | -0.85831 | 0.290507 | -2.95453 | 0.003131 | 0.009447 | 0.551597 |
| ANKRD54 | 282.3461 | -0.56739 | 0.192052 | -2.95438 | 0.003133 | 0.00945 | 0.674835 |
| E4F1 | 807.6179 | -0.49051 | 0.16615 | -2.95222 | 0.003155 | 0.009509 | 0.711772 |
| TMEM229E | 17.59589 | -1.96608 | 0.666701 | -2.94897 | 0.003188 | 0.009591 | 0.255948 |
| TMEM88B | 20.7973 | -1.58929 | 0.53935 | -2.94667 | 0.003212 | 0.009656 | 0.332335 |
| TMEM120E | 971.2359 | -0.59458 | 0.201957 | -2.94411 | 0.003239 | 0.009725 | 0.662235 |
| DDX18P1 | 3.295076 | -4.41132 | 1.498391 | -2.94404 | 0.00324 | 0.009726 | 0.046996 |

| | | | | | | | |
|---|---|---|---|---|---|---|---|
| C12orf73 | 220.7136 | -0.51679 | 0.175581 | -2.94332 | 0.003247 | 0.009745 | 0.698925 |
| PARM1 | 114.3634 | -0.66718 | 0.226721 | -2.94274 | 0.003253 | 0.00976 | 0.629736 |
| ROPN1B | 5.860457 | -2.82139 | 0.959084 | -2.94175 | 0.003264 | 0.00979 | 0.141475 |
| MACROD1 | 368.8788 | -0.44884 | 0.152639 | -2.94053 | 0.003277 | 0.009822 | 0.732632 |
| DEF6 | 147.3254 | -0.64211 | 0.218446 | -2.93947 | 0.003288 | 0.009853 | 0.640773 |
| NRXN1 | 405.2131 | -0.61308 | 0.208749 | -2.93694 | 0.003315 | 0.00992 | 0.653798 |
| ATP8B3 | 89.5229 | -0.75155 | 0.255982 | -2.93594 | 0.003325 | 0.009944 | 0.593966 |
| BTBD11 | 150.7863 | -0.56362 | 0.191997 | -2.93558 | 0.003329 | 0.009954 | 0.6766 |
| AKAP14 | 6.756727 | -3.33871 | 1.137719 | -2.93456 | 0.00334 | 0.009984 | 0.098844 |
| PODN | 13.77574 | -1.59873 | 0.545711 | -2.92962 | 0.003394 | 0.010121 | 0.330168 |
| CBWD4P | 15.95044 | -2.22609 | 0.759904 | -2.92944 | 0.003396 | 0.010125 | 0.213737 |
| DOK6 | 23.04528 | -2.23638 | 0.763659 | -2.9285 | 0.003406 | 0.010152 | 0.212218 |
| RBM41 | 478.7975 | -0.53825 | 0.183816 | -2.92819 | 0.003409 | 0.010161 | 0.688605 |
| CGB7 | 36.60502 | -1.77568 | 0.606718 | -2.9267 | 0.003426 | 0.010203 | 0.292056 |
| UPK1A | 43.18687 | -1.10315 | 0.377017 | -2.92599 | 0.003434 | 0.010221 | 0.4655 |
| SEC63 | 3274.435 | -0.46591 | 0.159332 | -2.92416 | 0.003454 | 0.010273 | 0.724013 |
| CDC14C | 5.527403 | -3.33297 | 1.140071 | -2.92348 | 0.003461 | 0.010291 | 0.099237 |
| SERPINF2 | 28.99461 | -1.01649 | 0.347753 | -2.92301 | 0.003467 | 0.010305 | 0.494319 |
| XG | 17.08734 | -1.62036 | 0.554368 | -2.92289 | 0.003468 | 0.010307 | 0.325255 |
| BCL6B | 110.4621 | -0.909 | 0.311021 | -2.92264 | 0.003471 | 0.010312 | 0.532553 |
| GOLT1A | 64.23617 | -0.97599 | 0.333975 | -2.92234 | 0.003474 | 0.010317 | 0.508392 |
| OSGIN1 | 24.08497 | -1.27722 | 0.437207 | -2.92132 | 0.003486 | 0.010344 | 0.41259 |
| CCDC92B | 46.79582 | -0.95381 | 0.326552 | -2.92085 | 0.003491 | 0.010358 | 0.516268 |
| FLVCR1 | 901.3693 | -0.65053 | 0.22273 | -2.92073 | 0.003492 | 0.010359 | 0.637044 |
| GDAP1 | 1046.212 | -0.52868 | 0.181069 | -2.91977 | 0.003503 | 0.010385 | 0.693188 |
| SYNE4 | 333.4809 | -0.42038 | 0.144011 | -2.91907 | 0.003511 | 0.010407 | 0.747229 |
| ABO | 22.09493 | -1.52985 | 0.524626 | -2.91608 | 0.003545 | 0.010501 | 0.346313 |
| NT5E | 29.91303 | -1.83392 | 0.629387 | -2.91382 | 0.00357 | 0.010568 | 0.280501 |
| SDR16C5 | 4.040054 | -3.92894 | 1.348821 | -2.91287 | 0.003581 | 0.010599 | 0.065656 |
| POM121L9 | 12.42399 | -1.56263 | 0.536592 | -2.91214 | 0.00359 | 0.01062 | 0.338532 |
| CD1D | 34.63655 | -0.95315 | 0.327464 | -2.9107 | 0.003606 | 0.010661 | 0.516504 |
| GPR68 | 32.63325 | -1.08661 | 0.373514 | -2.90916 | 0.003624 | 0.010707 | 0.470867 |
| PRR15 | 115.6147 | -0.63882 | 0.219844 | -2.90581 | 0.003663 | 0.01081 | 0.642237 |
| ATF7-NPFF | 24.61873 | -1.24185 | 0.427474 | -2.90509 | 0.003671 | 0.010829 | 0.422829 |
| MYO15A | 73.13293 | -1.22277 | 0.420973 | -2.90463 | 0.003677 | 0.010842 | 0.428459 |
| CERS5 | 1137.038 | -0.51845 | 0.178507 | -2.90439 | 0.00368 | 0.010848 | 0.698119 |
| PCDHB5 | 16.35466 | -1.51041 | 0.520105 | -2.90405 | 0.003684 | 0.010858 | 0.351011 |
| BFSP1 | 41.63579 | -1.05998 | 0.365254 | -2.90203 | 0.003708 | 0.010924 | 0.479639 |
| GJA3 | 38.79207 | -1.08869 | 0.375245 | -2.90128 | 0.003716 | 0.010948 | 0.470187 |
| C19orf47 | 348.6087 | -0.50098 | 0.17268 | -2.90118 | 0.003718 | 0.01095 | 0.706629 |
| LGALS4 | 6.765615 | -2.34996 | 0.810272 | -2.90021 | 0.003729 | 0.01098 | 0.196152 |
| TCP10L2 | 2.256652 | -4.6083 | 1.591078 | -2.89634 | 0.003775 | 0.011109 | 0.040998 |
| PKD1P1 | 201.9782 | -0.81463 | 0.281405 | -2.89486 | 0.003793 | 0.011156 | 0.568555 |
| EGFL6 | 146.1535 | -0.7582 | 0.262009 | -2.89379 | 0.003806 | 0.011191 | 0.591234 |

| | | | | | | | |
|---|---|---|---|---|---|---|---|
| CCDC36 | 7.126812 | -2.24223 | 0.775819 | -2.89015 | 0.003851 | 0.011311 | 0.211359 |
| PLA2G4C | 26.43783 | -1.28995 | 0.446587 | -2.88846 | 0.003871 | 0.011364 | 0.408966 |
| MT-CO1 | 425937.6 | -0.89991 | 0.311574 | -2.88827 | 0.003874 | 0.011369 | 0.535921 |
| PLAAT1 | 76.64106 | -0.61039 | 0.211513 | -2.88581 | 0.003904 | 0.011451 | 0.655021 |
| AL451007. | 2.199892 | -4.60846 | 1.597049 | -2.88561 | 0.003907 | 0.011457 | 0.040993 |
| AC239798 | 5.215335 | -3.29565 | 1.142325 | -2.88504 | 0.003914 | 0.011472 | 0.101838 |
| FANCA | 1778.158 | -0.6479 | 0.224587 | -2.88486 | 0.003916 | 0.011475 | 0.638208 |
| PIF1 | 1098.023 | -0.55616 | 0.192834 | -2.88414 | 0.003925 | 0.011499 | 0.68011 |
| PTK6 | 26.76409 | -1.08525 | 0.376405 | -2.88321 | 0.003936 | 0.011528 | 0.471309 |
| MAP3K13 | 209.4301 | -0.67431 | 0.233892 | -2.88302 | 0.003939 | 0.011533 | 0.626631 |
| PTK7 | 6377.541 | -0.4283 | 0.14857 | -2.88281 | 0.003941 | 0.011539 | 0.743138 |
| FAM186A | 5.722793 | -3.14031 | 1.089345 | -2.88275 | 0.003942 | 0.011539 | 0.113416 |
| RNASE1 | 41.13441 | -0.87121 | 0.302423 | -2.88077 | 0.003967 | 0.011606 | 0.546689 |
| CD300A | 45.29936 | -1.78772 | 0.620688 | -2.88022 | 0.003974 | 0.011623 | 0.28963 |
| NOTUM | 21.87237 | -1.47876 | 0.513659 | -2.87887 | 0.003991 | 0.011665 | 0.358797 |
| SETD4 | 534.4063 | -0.53392 | 0.185535 | -2.87775 | 0.004005 | 0.011703 | 0.690674 |
| GOLGA8K | 13.40561 | -2.0722 | 0.720109 | -2.87762 | 0.004007 | 0.011706 | 0.237796 |
| AP004833. | 2.79208 | -4.17269 | 1.451501 | -2.87474 | 0.004044 | 0.011806 | 0.055449 |
| SLC2A14 | 104.4741 | -0.71136 | 0.247672 | -2.87219 | 0.004076 | 0.011896 | 0.610743 |
| CD209 | 4.48006 | -3.57709 | 1.245816 | -2.87129 | 0.004088 | 0.011925 | 0.083789 |
| SLC12A8 | 293.1675 | -0.60965 | 0.21238 | -2.87057 | 0.004097 | 0.011947 | 0.655355 |
| NFIB | 102.4866 | -0.72644 | 0.253159 | -2.8695 | 0.004111 | 0.011982 | 0.604394 |
| KLK8 | 17.04879 | -1.61477 | 0.562837 | -2.86899 | 0.004118 | 0.012 | 0.326516 |
| POU2F3 | 129.0641 | -0.75982 | 0.26497 | -2.86758 | 0.004136 | 0.012048 | 0.590569 |
| HIP1R | 470.2182 | -0.695 | 0.242375 | -2.86745 | 0.004138 | 0.012051 | 0.61771 |
| PRSS27 | 91.05016 | -0.74697 | 0.260801 | -2.86412 | 0.004182 | 0.01216 | 0.595855 |
| TFPT | 71.73915 | -0.99719 | 0.348268 | -2.86329 | 0.004193 | 0.012187 | 0.500974 |
| EEF2K | 85.60221 | -1.37668 | 0.480864 | -2.86293 | 0.004197 | 0.012197 | 0.385103 |
| GUSBP5 | 16.66563 | -1.78988 | 0.625531 | -2.86137 | 0.004218 | 0.012251 | 0.289197 |
| DMXL1 | 1397.017 | -0.42859 | 0.14983 | -2.86054 | 0.004229 | 0.012281 | 0.742985 |
| MROH5 | 7.830853 | -2.7659 | 0.967366 | -2.8592 | 0.004247 | 0.012327 | 0.147022 |
| NME8 | 3.854336 | -3.33753 | 1.168166 | -2.85707 | 0.004276 | 0.012405 | 0.098925 |
| TSPAN32 | 14.28055 | -2.4773 | 0.867333 | -2.85623 | 0.004287 | 0.012433 | 0.17958 |
| SHROOM1 | 152.397 | -0.73407 | 0.25706 | -2.85564 | 0.004295 | 0.012453 | 0.601205 |
| RAC2 | 31.5498 | -0.95598 | 0.334892 | -2.8546 | 0.004309 | 0.01249 | 0.51549 |
| CHAC1 | 2379.903 | -0.4253 | 0.148999 | -2.85437 | 0.004312 | 0.012497 | 0.744684 |
| ZC3H12D | 19.68783 | -1.27799 | 0.44778 | -2.85406 | 0.004316 | 0.012507 | 0.412369 |
| C19orf38 | 6.861484 | -2.16504 | 0.758597 | -2.85401 | 0.004317 | 0.012507 | 0.222975 |
| YPEL3 | 223.214 | -0.75325 | 0.263964 | -2.85361 | 0.004322 | 0.01252 | 0.593265 |
| AL137013. | 9.750822 | -2.43573 | 0.854211 | -2.85144 | 0.004352 | 0.012595 | 0.18483 |
| C1orf127 | 19.68139 | -2.51579 | 0.882585 | -2.85048 | 0.004365 | 0.012624 | 0.174853 |
| FGF22 | 22.78486 | -1.74995 | 0.613961 | -2.85026 | 0.004368 | 0.01263 | 0.297313 |
| SLC19A3 | 87.47629 | -0.99104 | 0.347974 | -2.84802 | 0.004399 | 0.012705 | 0.503116 |
| ITGA1 | 195.1123 | -0.60191 | 0.211355 | -2.84785 | 0.004402 | 0.01271 | 0.658882 |

| | | | | | | | |
|---|---|---|---|---|---|---|---|
| STAT4 | 16.78828 | -2.37201 | 0.833411 | -2.84615 | 0.004425 | 0.012772 | 0.193176 |
| SMCO2 | 5.342527 | -2.60321 | 0.915035 | -2.84493 | 0.004442 | 0.012819 | 0.164572 |
| EXD3 | 208.9123 | -0.68648 | 0.241816 | -2.83884 | 0.004528 | 0.013055 | 0.621369 |
| TULP1 | 28.58807 | -1.12412 | 0.39634 | -2.83625 | 0.004565 | 0.013151 | 0.458782 |
| C9orf85 | 216.8138 | -0.47281 | 0.166709 | -2.83617 | 0.004566 | 0.013153 | 0.720558 |
| NINL | 2165.718 | -0.43912 | 0.15505 | -2.83213 | 0.004624 | 0.013299 | 0.737583 |
| CPSF1P1 | 135.5813 | -0.7316 | 0.258333 | -2.83202 | 0.004625 | 0.013302 | 0.602234 |
| AC008736 | 68.09103 | -1.19023 | 0.420358 | -2.83147 | 0.004633 | 0.013323 | 0.438233 |
| CACNA1G | 211.2073 | -0.49829 | 0.176026 | -2.8308 | 0.004643 | 0.013346 | 0.707943 |
| IFNLR1 | 292.4811 | -0.4954 | 0.175018 | -2.83053 | 0.004647 | 0.013355 | 0.709367 |
| ADAD2 | 12.50628 | -3.01316 | 1.065343 | -2.82835 | 0.004679 | 0.013445 | 0.123865 |
| AC124068 | 23.32596 | -2.00453 | 0.708911 | -2.82762 | 0.00469 | 0.013467 | 0.249216 |
| CAPN10 | 789.1459 | -0.62121 | 0.21969 | -2.82765 | 0.004689 | 0.013467 | 0.650127 |
| FRMD1 | 20.55328 | -1.39053 | 0.491835 | -2.82724 | 0.004695 | 0.013481 | 0.381423 |
| PDE3A | 283.0125 | -0.49356 | 0.174677 | -2.82557 | 0.00472 | 0.013547 | 0.710269 |
| CD53 | 12.44568 | -2.08203 | 0.736993 | -2.82504 | 0.004728 | 0.013565 | 0.236182 |
| CGREF1 | 69.27681 | -0.84217 | 0.298111 | -2.82502 | 0.004728 | 0.013565 | 0.557804 |
| PCYT2 | 487.6523 | -0.59854 | 0.211897 | -2.82469 | 0.004733 | 0.013572 | 0.66042 |
| USP18 | 108.4514 | -0.81268 | 0.28796 | -2.82219 | 0.00477 | 0.013674 | 0.569324 |
| AL445423. | 124.0207 | -0.71525 | 0.253547 | -2.82096 | 0.004788 | 0.01372 | 0.609101 |
| PCDHGA5 | 15.09792 | -1.54895 | 0.549404 | -2.81932 | 0.004813 | 0.013779 | 0.34176 |
| PPP2R5B | 188.5074 | -0.85942 | 0.304834 | -2.81932 | 0.004813 | 0.013779 | 0.551173 |
| IL22RA1 | 14.95204 | -1.61176 | 0.572145 | -2.81704 | 0.004847 | 0.013864 | 0.327199 |
| VWA7 | 28.54445 | -1.878 | 0.666702 | -2.81685 | 0.00485 | 0.013871 | 0.272061 |
| SGSH | 402.1596 | -0.59788 | 0.212276 | -2.81653 | 0.004855 | 0.01388 | 0.660724 |
| SLC22A4 | 14.85493 | -1.58051 | 0.561414 | -2.81522 | 0.004874 | 0.01393 | 0.334365 |
| TBXA2R | 49.93516 | -0.8038 | 0.285565 | -2.81479 | 0.004881 | 0.013945 | 0.572837 |
| RFPL2 | 10.62604 | -1.75996 | 0.625884 | -2.81196 | 0.004924 | 0.014064 | 0.295256 |
| SSC4D | 510.6764 | -0.52795 | 0.18778 | -2.81153 | 0.004931 | 0.014078 | 0.69354 |
| CTAGE12P | 4.894449 | -4.19852 | 1.494342 | -2.80961 | 0.00496 | 0.01416 | 0.054465 |
| AGAP5 | 135.8143 | -0.68133 | 0.242514 | -2.80946 | 0.004963 | 0.014165 | 0.623588 |
| VDAC1P13 | 3.821575 | -4.61409 | 1.643103 | -2.80816 | 0.004983 | 0.014215 | 0.040834 |
| ZNF251 | 430.3948 | -0.69265 | 0.24672 | -2.80743 | 0.004994 | 0.014243 | 0.618717 |
| CEL | 72.26864 | -0.97724 | 0.348363 | -2.80524 | 0.005028 | 0.014333 | 0.50795 |
| AC104304 | 8.264115 | -2.42388 | 0.864108 | -2.80507 | 0.005031 | 0.014335 | 0.186354 |
| TGFBR3L | 56.76883 | -1.16741 | 0.416186 | -2.80501 | 0.005031 | 0.014335 | 0.445221 |
| AL355987. | 4.122247 | -3.97738 | 1.418068 | -2.80479 | 0.005035 | 0.014342 | 0.063488 |
| GNPTG | 671.7045 | -0.45319 | 0.161717 | -2.80234 | 0.005073 | 0.014445 | 0.730428 |
| WASHC1 | 277.6173 | -0.43006 | 0.153515 | -2.80141 | 0.005088 | 0.014482 | 0.742232 |
| SLC22A2 | 3.692632 | -3.82086 | 1.364738 | -2.79971 | 0.005115 | 0.014548 | 0.070763 |
| SYNPO | 205.4352 | -0.52501 | 0.187607 | -2.79846 | 0.005135 | 0.014602 | 0.694954 |
| FAM90A1 | 13.00037 | -1.61051 | 0.575559 | -2.79816 | 0.005139 | 0.014613 | 0.327484 |
| OR2B6 | 3.254568 | -5.18787 | 1.854098 | -2.79806 | 0.005141 | 0.014615 | 0.027434 |
| AP5Z1 | 742.9141 | -0.45994 | 0.164401 | -2.79765 | 0.005148 | 0.014629 | 0.727018 |

| | | | | | | | |
|---|---|---|---|---|---|---|---|
| PGA5 | 38.1646 | -1.42021 | 0.50777 | -2.79695 | 0.005159 | 0.014659 | 0.373658 |
| BDH1 | 347.5984 | -0.51193 | 0.183131 | -2.79542 | 0.005183 | 0.014719 | 0.701285 |
| TMEM92 | 273.6046 | -0.45776 | 0.163792 | -2.79474 | 0.005194 | 0.014747 | 0.728118 |
| OR6J1 | 30.80527 | -1.06812 | 0.382331 | -2.79371 | 0.005211 | 0.014786 | 0.476939 |
| CREG2 | 49.63157 | -1.39242 | 0.498449 | -2.79351 | 0.005214 | 0.014793 | 0.380926 |
| TMEM199 | 523.3408 | -0.46239 | 0.165585 | -2.79249 | 0.00523 | 0.01483 | 0.725781 |
| WNK4 | 25.71103 | -1.0929 | 0.391438 | -2.79202 | 0.005238 | 0.01485 | 0.468817 |
| PPM1N | 111.6767 | -0.57869 | 0.207323 | -2.79123 | 0.005251 | 0.014881 | 0.669573 |
| TLR6 | 13.12176 | -2.31923 | 0.831099 | -2.79056 | 0.005262 | 0.014908 | 0.200375 |
| C1orf159 | 371.6633 | -0.51783 | 0.185596 | -2.7901 | 0.005269 | 0.014924 | 0.698421 |
| TRPM2 | 6.53792 | -2.58623 | 0.926927 | -2.79011 | 0.005269 | 0.014924 | 0.16652 |
| UPRT | 394.5133 | -0.4366 | 0.15649 | -2.78994 | 0.005272 | 0.014929 | 0.738876 |
| C1orf56 | 110.2535 | -0.77983 | 0.279569 | -2.78941 | 0.00528 | 0.014951 | 0.582435 |
| CACNA1D | 50.37799 | -1.00506 | 0.360489 | -2.78805 | 0.005303 | 0.015 | 0.498248 |
| SOWAHB | 18.90104 | -1.32891 | 0.476761 | -2.78737 | 0.005314 | 0.015025 | 0.398069 |
| TRPV2 | 39.34143 | -1.42332 | 0.511064 | -2.78501 | 0.005353 | 0.015123 | 0.372853 |
| SUSD3 | 35.21856 | -1.22622 | 0.440409 | -2.78427 | 0.005365 | 0.015153 | 0.427436 |
| GMIP | 287.4476 | -0.48209 | 0.17318 | -2.78376 | 0.005373 | 0.015174 | 0.715939 |
| CDHR4 | 15.27483 | -1.42793 | 0.512976 | -2.78363 | 0.005375 | 0.015178 | 0.371662 |
| GRIK2 | 83.10497 | -1.02012 | 0.36649 | -2.7835 | 0.005378 | 0.015182 | 0.493074 |
| SH3GL1P3 | 9.270918 | -1.97624 | 0.710563 | -2.78124 | 0.005415 | 0.015279 | 0.254151 |
| CHRFAM7/ | 117.2172 | -1.08807 | 0.391301 | -2.78065 | 0.005425 | 0.015302 | 0.47039 |
| NFATC4 | 1557.682 | -0.58191 | 0.209277 | -2.78059 | 0.005426 | 0.015302 | 0.668077 |
| NHLH2 | 38.40688 | -0.91563 | 0.329316 | -2.78039 | 0.005429 | 0.015309 | 0.530113 |
| CA10 | 8.134271 | -2.11351 | 0.76034 | -2.77969 | 0.005441 | 0.01534 | 0.231085 |
| AC020915 | 2.061729 | -4.52603 | 1.629075 | -2.77828 | 0.005465 | 0.015399 | 0.043404 |
| 12-Sep | 3.252699 | -3.8256 | 1.377632 | -2.77694 | 0.005487 | 0.015457 | 0.070531 |
| CLPTM1L | 1600.805 | -0.49932 | 0.179823 | -2.77672 | 0.005491 | 0.015465 | 0.707442 |
| NTSR1 | 19.0077 | -1.26649 | 0.456169 | -2.77637 | 0.005497 | 0.015479 | 0.415668 |
| STAG3 | 242.2175 | -0.56658 | 0.204154 | -2.77526 | 0.005516 | 0.015526 | 0.675216 |
| MS4A4E | 172.7311 | -0.50418 | 0.181677 | -2.77513 | 0.005518 | 0.015528 | 0.705063 |
| AGBL2 | 63.74657 | -0.92848 | 0.335046 | -2.7712 | 0.005585 | 0.015698 | 0.525412 |
| ITK | 7.435134 | -2.30016 | 0.831044 | -2.7678 | 0.005644 | 0.015855 | 0.20304 |
| INPPL1 | 3200.538 | -0.44061 | 0.159252 | -2.76672 | 0.005662 | 0.015903 | 0.736825 |
| SMIM5 | 13.08672 | -1.57289 | 0.568916 | -2.76472 | 0.005697 | 0.015986 | 0.336133 |
| RIPK3 | 5.910096 | -2.38046 | 0.861316 | -2.76374 | 0.005714 | 0.016029 | 0.192048 |
| NKAIN4 | 200.5184 | -0.55985 | 0.202863 | -2.75972 | 0.005785 | 0.01622 | 0.678375 |
| B4GALNT1 | 32.8049 | -1.08137 | 0.391863 | -2.75955 | 0.005788 | 0.016226 | 0.472581 |
| CNTD2 | 2.329776 | -4.66951 | 1.692281 | -2.7593 | 0.005793 | 0.016236 | 0.039295 |
| ITGB1BP2 | 24.33357 | -1.15236 | 0.417639 | -2.75922 | 0.005794 | 0.016238 | 0.44989 |
| VDR | 7.706724 | -2.20111 | 0.797777 | -2.75905 | 0.005797 | 0.016244 | 0.217471 |
| CYP2F2P | 5.802091 | -2.52553 | 0.91541 | -2.75891 | 0.005799 | 0.016248 | 0.173675 |
| RASSF6 | 5.380035 | -3.64729 | 1.322835 | -2.75718 | 0.00583 | 0.016331 | 0.07981 |
| IFIH1 | 93.30823 | -0.70485 | 0.255867 | -2.75475 | 0.005874 | 0.016441 | 0.613507 |

| | | | | | | | |
|---|---|---|---|---|---|---|---|
| PKD1L2 | 125.022 | -0.58844 | 0.213782 | -2.75251 | 0.005914 | 0.016547 | 0.665063 |
| PLEKHN1 | 17.58304 | -1.50826 | 0.548195 | -2.75132 | 0.005936 | 0.016596 | 0.351535 |
| TLR2 | 32.3505 | -1.37516 | 0.499825 | -2.75127 | 0.005936 | 0.016596 | 0.385511 |
| AL136295. | 56.51924 | -1.58469 | 0.576451 | -2.74904 | 0.005977 | 0.016702 | 0.333397 |
| ST8SIA3 | 245.1531 | -0.73919 | 0.268929 | -2.74863 | 0.005984 | 0.016716 | 0.599077 |
| POLR3G | 2451.62 | -0.48611 | 0.17689 | -2.74811 | 0.005994 | 0.016739 | 0.713946 |
| MAP3K10 | 530.9213 | -0.48261 | 0.175667 | -2.74731 | 0.006009 | 0.016778 | 0.715682 |
| SHISAL2A | 12.26142 | -1.99903 | 0.727779 | -2.74675 | 0.006019 | 0.016804 | 0.250169 |
| GPR158 | 22.13714 | -1.42683 | 0.519627 | -2.74588 | 0.006035 | 0.016846 | 0.371947 |
| TPPP | 72.34085 | -0.78272 | 0.28511 | -2.74533 | 0.006045 | 0.016867 | 0.58127 |
| TM6SF2 | 129.1482 | -0.61123 | 0.222668 | -2.74501 | 0.006051 | 0.016878 | 0.65464 |
| L3MBTL1 | 656.5702 | -0.41653 | 0.151797 | -2.74397 | 0.00607 | 0.016929 | 0.749227 |
| AL445238. | 3.701251 | -3.15038 | 1.148255 | -2.74362 | 0.006077 | 0.016944 | 0.112627 |
| PLD2 | 1024.818 | -0.42455 | 0.1549 | -2.74078 | 0.006129 | 0.017071 | 0.745072 |
| IGSF10 | 55.37045 | -1.23422 | 0.450366 | -2.74048 | 0.006135 | 0.017082 | 0.425073 |
| VILL | 169.0438 | -0.44515 | 0.162459 | -2.74006 | 0.006143 | 0.017098 | 0.734509 |
| SHISA4 | 150.8437 | -0.56658 | 0.20683 | -2.73935 | 0.006156 | 0.017122 | 0.675215 |
| SLC46A1 | 247.4032 | -0.5748 | 0.209865 | -2.73891 | 0.006164 | 0.017142 | 0.671379 |
| ALOX15 | 16.27378 | -1.47628 | 0.539057 | -2.73864 | 0.006169 | 0.017147 | 0.359414 |
| CHRNA4 | 77.62085 | -1.39995 | 0.511187 | -2.73862 | 0.00617 | 0.017147 | 0.378943 |
| STX1B | 309.3369 | -0.57288 | 0.209193 | -2.73851 | 0.006172 | 0.01715 | 0.672274 |
| GTF2A1L | 3.111737 | -4.27161 | 1.559971 | -2.73826 | 0.006177 | 0.017161 | 0.051775 |
| AQP3 | 38.4623 | -0.92765 | 0.339224 | -2.73462 | 0.006245 | 0.017323 | 0.525714 |
| ADGRA2 | 57.69921 | -0.81999 | 0.299985 | -2.73344 | 0.006268 | 0.01738 | 0.566445 |
| FAM91A2P | 15.27445 | -1.58695 | 0.580662 | -2.73301 | 0.006276 | 0.0174 | 0.332873 |
| PTGER2 | 15.47687 | -1.53818 | 0.562859 | -2.73281 | 0.00628 | 0.017406 | 0.344319 |
| COBLL1 | 120.5075 | -0.65341 | 0.239129 | -2.73247 | 0.006286 | 0.017421 | 0.635774 |
| ATG4D | 670.5326 | -0.42967 | 0.157299 | -2.73152 | 0.006304 | 0.017468 | 0.742434 |
| BAIAP3 | 147.345 | -1.26933 | 0.46489 | -2.73038 | 0.006326 | 0.017521 | 0.414854 |
| TNNT2 | 71.43785 | -0.74688 | 0.273759 | -2.72824 | 0.006367 | 0.017621 | 0.595891 |
| NXPH4 | 58.01305 | -0.79266 | 0.290571 | -2.72793 | 0.006373 | 0.017636 | 0.57728 |
| REEP6 | 374.446 | -0.46588 | 0.170818 | -2.72732 | 0.006385 | 0.017663 | 0.724032 |
| AGA | 360.0402 | -0.49383 | 0.181083 | -2.72711 | 0.006389 | 0.01767 | 0.710135 |
| GLMP | 1203.768 | -0.49323 | 0.180916 | -2.72627 | 0.006406 | 0.017703 | 0.710435 |
| OR2AG2 | 12.81483 | -1.74586 | 0.640841 | -2.72433 | 0.006443 | 0.017789 | 0.298156 |
| RUNX1T1 | 224.0264 | -0.8357 | 0.306821 | -2.72375 | 0.006454 | 0.017809 | 0.56031 |
| HERC6 | 131.7467 | -0.65716 | 0.241386 | -2.72244 | 0.00648 | 0.017869 | 0.634126 |
| KCNC4 | 343.614 | -0.52104 | 0.191438 | -2.72174 | 0.006494 | 0.017905 | 0.696868 |
| MNS1 | 48.81655 | -0.87582 | 0.322074 | -2.71932 | 0.006542 | 0.01802 | 0.544943 |
| ASB16 | 54.34286 | -0.83949 | 0.30875 | -2.71899 | 0.006548 | 0.01803 | 0.558842 |
| USP9Y | 3.095978 | -4.14185 | 1.524179 | -2.71743 | 0.006579 | 0.018104 | 0.056647 |
| GNG3 | 70.58942 | -0.66183 | 0.243598 | -2.71688 | 0.00659 | 0.01813 | 0.632078 |
| CRLF1 | 44.51941 | -1.07554 | 0.395927 | -2.71652 | 0.006597 | 0.018143 | 0.474492 |
| SLC47A1 | 85.55044 | -0.7575 | 0.278901 | -2.71601 | 0.006607 | 0.018164 | 0.591521 |

| | | | | | | | |
|---|---|---|---|---|---|---|---|
| GALNT4 | 132.0694 | -0.86161 | 0.317659 | -2.71238 | 0.00668 | 0.018344 | 0.550337 |
| KCNK13 | 4.688856 | -3.09379 | 1.140732 | -2.71211 | 0.006686 | 0.018352 | 0.117132 |
| KCNJ15 | 49.39122 | -0.69817 | 0.257518 | -2.71114 | 0.006705 | 0.018399 | 0.616355 |
| CYSRT1 | 27.31808 | -1.2935 | 0.477201 | -2.7106 | 0.006716 | 0.018423 | 0.40796 |
| ZNF876P | 2.698283 | -4.21947 | 1.557642 | -2.70889 | 0.006751 | 0.018499 | 0.05368 |
| HS3ST3A1 | 219.9004 | -0.67707 | 0.250269 | -2.70538 | 0.006823 | 0.018674 | 0.625433 |
| KLHL31 | 13.00344 | -1.5687 | 0.580221 | -2.70363 | 0.006859 | 0.018764 | 0.337111 |
| GTF3C1 | 4639.116 | -0.44404 | 0.164262 | -2.70327 | 0.006866 | 0.018781 | 0.735071 |
| IRX6 | 25.53976 | -1.14661 | 0.424269 | -2.70254 | 0.006881 | 0.018817 | 0.451687 |
| 11-Mar | 27.12902 | -1.43663 | 0.531578 | -2.70258 | 0.00688 | 0.018817 | 0.369429 |
| CLUL1 | 15.26612 | -2.30758 | 0.853974 | -2.70216 | 0.006889 | 0.018833 | 0.202 |
| TCF24 | 13.70436 | -1.48703 | 0.550417 | -2.70163 | 0.0069 | 0.018854 | 0.356747 |
| FBXO6 | 40.2494 | -0.88089 | 0.326218 | -2.70031 | 0.006928 | 0.018918 | 0.543033 |
| PROSER2 | 235.8469 | -0.42766 | 0.158418 | -2.69957 | 0.006943 | 0.018958 | 0.743467 |
| PROM2 | 120.6665 | -1.18847 | 0.440488 | -2.69808 | 0.006974 | 0.019034 | 0.438768 |
| CDK10 | 1232.081 | -0.45247 | 0.167727 | -2.69764 | 0.006983 | 0.019054 | 0.730792 |
| BHMT | 21.73206 | -1.30013 | 0.482408 | -2.69508 | 0.007037 | 0.019178 | 0.40609 |
| MIOX | 11.39547 | -1.5822 | 0.587403 | -2.69355 | 0.00707 | 0.019255 | 0.333972 |
| BMP8A | 83.29876 | -0.59016 | 0.21913 | -2.69319 | 0.007077 | 0.01927 | 0.66427 |
| PLA2G3 | 253.5679 | -0.65789 | 0.244309 | -2.69285 | 0.007085 | 0.019287 | 0.633806 |
| OR6D1P | 98.5398 | -0.67953 | 0.252374 | -2.69257 | 0.00709 | 0.0193 | 0.624367 |
| SFTPD | 10.98615 | -1.87574 | 0.696741 | -2.69217 | 0.007099 | 0.01932 | 0.272486 |
| AL355877. | 29.92842 | -1.01824 | 0.378805 | -2.68802 | 0.007188 | 0.019553 | 0.493719 |
| XCR1 | 2.552712 | -4.0075 | 1.490932 | -2.68792 | 0.00719 | 0.019557 | 0.062176 |
| MTCO2P12 | 23.94034 | -1.34142 | 0.499141 | -2.68745 | 0.0072 | 0.019575 | 0.394633 |
| CTF1 | 24.79525 | -0.97289 | 0.362127 | -2.68661 | 0.007218 | 0.019622 | 0.509483 |
| ZNF425 | 268.0719 | -0.70435 | 0.262276 | -2.68552 | 0.007242 | 0.019677 | 0.61372 |
| NODAL | 23.56226 | -1.08729 | 0.404944 | -2.68504 | 0.007252 | 0.019699 | 0.470644 |
| SPRY4 | 2387.226 | -0.5245 | 0.195795 | -2.67884 | 0.007388 | 0.020021 | 0.695198 |
| TNS4 | 10.40703 | -2.26987 | 0.847526 | -2.67823 | 0.007401 | 0.020048 | 0.207349 |
| DNAH17 | 59.88327 | -1.40138 | 0.523286 | -2.67803 | 0.007406 | 0.020054 | 0.378568 |
| VWA5B2 | 19.69044 | -1.56472 | 0.584296 | -2.67795 | 0.007407 | 0.020056 | 0.338044 |
| REELD1 | 5.48374 | -2.87137 | 1.072327 | -2.6777 | 0.007413 | 0.020068 | 0.136656 |
| FSTL4 | 101.911 | -0.62306 | 0.232793 | -2.67644 | 0.007441 | 0.020129 | 0.649294 |
| CDRT4 | 278.0994 | -0.94132 | 0.351853 | -2.67533 | 0.007466 | 0.02019 | 0.520755 |
| TUBB8 | 19.54396 | -1.57246 | 0.587849 | -2.67493 | 0.007474 | 0.020205 | 0.336236 |
| STAC3 | 41.72156 | -1.18718 | 0.443849 | -2.67473 | 0.007479 | 0.020211 | 0.439161 |
| ZNF579 | 540.6963 | -0.48897 | 0.182822 | -2.67455 | 0.007483 | 0.020217 | 0.712535 |
| KISS1R | 41.72641 | -1.33535 | 0.499303 | -2.67444 | 0.007485 | 0.02022 | 0.396295 |
| PLD1 | 40.08015 | -1.01965 | 0.381329 | -2.67395 | 0.007496 | 0.020244 | 0.493235 |
| GSTM3P1 | 15.43433 | -1.39265 | 0.52084 | -2.67385 | 0.007499 | 0.020246 | 0.380865 |
| IL23A | 18.95013 | -1.60208 | 0.599247 | -2.67349 | 0.007507 | 0.020262 | 0.329402 |
| MC1R | 96.51368 | -0.75206 | 0.281475 | -2.67185 | 0.007543 | 0.020346 | 0.593755 |
| KIR3DX1 | 9.968906 | -1.94821 | 0.72929 | -2.67138 | 0.007554 | 0.020369 | 0.259138 |

| | | | | | | | |
|---|---|---|---|---|---|---|---|
| CALCR | 11.42473 | -1.6101 | 0.602787 | -2.67109 | 0.00756 | 0.02038 | 0.327576 |
| EAF2 | 65.42371 | -0.70815 | 0.265181 | -2.67045 | 0.007575 | 0.020416 | 0.612103 |
| AC092072 | 101.6822 | -0.53876 | 0.201899 | -2.66847 | 0.00762 | 0.020522 | 0.688362 |
| FKBP2 | 626.0395 | -0.45733 | 0.171398 | -2.66825 | 0.007625 | 0.020533 | 0.728332 |
| OR2B11 | 27.96181 | -0.88853 | 0.333027 | -2.66803 | 0.00763 | 0.020537 | 0.540166 |
| FKBP6 | 12.58216 | -2.92 | 1.095175 | -2.66624 | 0.00767 | 0.020634 | 0.132127 |
| TCHH | 20.3996 | -1.08271 | 0.406108 | -2.66607 | 0.007674 | 0.020642 | 0.472141 |
| CERCAM | 109.7507 | -0.70928 | 0.266151 | -2.66495 | 0.0077 | 0.020706 | 0.611626 |
| ANO8 | 910.3746 | -0.61688 | 0.23153 | -2.66438 | 0.007713 | 0.020734 | 0.652079 |
| ABCB11 | 36.73416 | -0.86569 | 0.325007 | -2.6636 | 0.007731 | 0.020773 | 0.548785 |
| AC112198 | 2.010574 | -4.48226 | 1.683444 | -2.66255 | 0.007755 | 0.020828 | 0.044741 |
| DPPA2 | 21.70513 | -1.14867 | 0.431601 | -2.66142 | 0.007781 | 0.020887 | 0.45104 |
| HSPB2 | 5.745076 | -2.23246 | 0.83922 | -2.66016 | 0.00781 | 0.020956 | 0.212795 |
| AC003665 | 204.9141 | -0.6853 | 0.257855 | -2.6577 | 0.007868 | 0.021097 | 0.621877 |
| TEKT3 | 11.44567 | -2.11621 | 0.796538 | -2.65676 | 0.00789 | 0.021153 | 0.230653 |
| FAM214A | 208.523 | -0.4806 | 0.180902 | -2.6567 | 0.007891 | 0.021153 | 0.716678 |
| LCN10 | 49.25556 | -0.93923 | 0.353555 | -2.65653 | 0.007895 | 0.021161 | 0.521511 |
| TIAM2 | 304.5875 | -0.53023 | 0.199807 | -2.6537 | 0.007961 | 0.021324 | 0.692445 |
| ITGA2B | 287.5441 | -0.81894 | 0.308793 | -2.65206 | 0.008 | 0.021415 | 0.56686 |
| ZNF157 | 33.15335 | -0.98189 | 0.370629 | -2.64925 | 0.008067 | 0.021572 | 0.506316 |
| GPC5 | 7.253882 | -2.40278 | 0.907561 | -2.64752 | 0.008109 | 0.021673 | 0.189099 |
| ZDHHC19 | 13.04221 | -2.23393 | 0.843795 | -2.64748 | 0.008109 | 0.021673 | 0.212578 |
| SSPO | 67.91862 | -1.23744 | 0.467494 | -2.64696 | 0.008122 | 0.021699 | 0.424125 |
| DNAJA1P3 | 4.335199 | -2.5882 | 0.978135 | -2.64606 | 0.008144 | 0.021739 | 0.166293 |
| KLF2P1 | 12.58332 | -1.38284 | 0.522601 | -2.64607 | 0.008143 | 0.021739 | 0.383463 |
| ZNF391 | 398.0229 | -0.59206 | 0.223842 | -2.64501 | 0.008169 | 0.021803 | 0.663393 |
| CCT6P3 | 214.7672 | -0.42155 | 0.15939 | -2.64479 | 0.008174 | 0.021814 | 0.74662 |
| CNGA1 | 14.0398 | -1.57417 | 0.595237 | -2.6446 | 0.008179 | 0.021823 | 0.335837 |
| FBXL15 | 232.7432 | -0.47005 | 0.177913 | -2.64202 | 0.008241 | 0.021965 | 0.72194 |
| SUSD5 | 157.1835 | -0.59156 | 0.22399 | -2.64103 | 0.008265 | 0.022023 | 0.663624 |
| PNPLA7 | 59.74314 | -0.85172 | 0.32253 | -2.64074 | 0.008273 | 0.022038 | 0.554125 |
| CROCC | 2130.513 | -0.50034 | 0.189625 | -2.63857 | 0.008326 | 0.022173 | 0.706941 |
| CLEC4GP1 | 2.807442 | -4.94549 | 1.876062 | -2.6361 | 0.008386 | 0.022316 | 0.032453 |
| SLC34A2 | 236.8809 | -0.81677 | 0.309839 | -2.63612 | 0.008386 | 0.022316 | 0.567711 |
| RLIMP1 | 2.590981 | -4.03664 | 1.532078 | -2.63474 | 0.00842 | 0.022395 | 0.060933 |
| C19orf73 | 26.42996 | -0.92652 | 0.351925 | -2.63273 | 0.00847 | 0.022512 | 0.526124 |
| GPRC5A | 33.63728 | -1.91435 | 0.727212 | -2.63246 | 0.008477 | 0.022527 | 0.265291 |
| DCBLD1 | 376.5778 | -0.47786 | 0.181646 | -2.6307 | 0.008521 | 0.022637 | 0.718044 |
| FAM114A1 | 194.4767 | -0.73059 | 0.277847 | -2.62947 | 0.008552 | 0.02271 | 0.602657 |
| TRPM5 | 5.015647 | -2.77087 | 1.053756 | -2.62952 | 0.008551 | 0.02271 | 0.146516 |
| MAB21L3 | 84.89864 | -0.62851 | 0.23912 | -2.62841 | 0.008578 | 0.022777 | 0.646846 |
| SLC2A11 | 319.2721 | -0.5795 | 0.220644 | -2.62641 | 0.008629 | 0.022905 | 0.669194 |
| WASH2P | 367.2553 | -0.44714 | 0.170301 | -2.62557 | 0.00865 | 0.022948 | 0.733497 |
| DLG2 | 68.85003 | -1.27385 | 0.485307 | -2.62482 | 0.008669 | 0.022986 | 0.413556 |

| | | | | | | | |
|---|---|---|---|---|---|---|---|
| AC135721 | 8.856712 | -1.8812 | 0.717174 | -2.62308 | 0.008714 | 0.023094 | 0.271457 |
| HAPLN2 | 10.56461 | -1.66269 | 0.633871 | -2.62307 | 0.008714 | 0.023094 | 0.31585 |
| CD99P1 | 149.5419 | -0.43672 | 0.166516 | -2.62268 | 0.008724 | 0.023114 | 0.738813 |
| TTYH2 | 169.6894 | -0.74076 | 0.282455 | -2.62256 | 0.008727 | 0.023119 | 0.598426 |
| TENM2 | 72.51068 | -1.35753 | 0.518422 | -2.61859 | 0.008829 | 0.023373 | 0.390249 |
| DLEU7 | 76.9909 | -0.61474 | 0.23478 | -2.61837 | 0.008835 | 0.023385 | 0.653047 |
| HNMT | 112.7944 | -0.7189 | 0.274635 | -2.61766 | 0.008853 | 0.023423 | 0.60756 |
| PARP10 | 148.5029 | -1.05586 | 0.403421 | -2.61727 | 0.008864 | 0.023446 | 0.48101 |
| PTRH1 | 114.1126 | -0.50566 | 0.193214 | -2.61712 | 0.008868 | 0.023453 | 0.704337 |
| PCED1B | 55.1237 | -0.74984 | 0.286885 | -2.61372 | 0.008956 | 0.02365 | 0.59467 |
| STAB1 | 18.14159 | -1.72937 | 0.66169 | -2.61357 | 0.00896 | 0.023658 | 0.301584 |
| CFLAR | 446.5076 | -0.60582 | 0.231832 | -2.6132 | 0.00897 | 0.02368 | 0.657096 |
| SHISAL2B | 65.47159 | -0.68008 | 0.260381 | -2.61187 | 0.009005 | 0.023765 | 0.624131 |
| ZFYVE27 | 478.0169 | -0.54295 | 0.207882 | -2.61181 | 0.009006 | 0.023765 | 0.686367 |
| DCDC2 | 43.45489 | -0.9192 | 0.351955 | -2.6117 | 0.009009 | 0.023767 | 0.528802 |
| TMSB4XP4 | 22.24525 | -1.20842 | 0.462928 | -2.61039 | 0.009044 | 0.023851 | 0.432742 |
| TNFSF12 | 110.5621 | -0.49891 | 0.191142 | -2.61014 | 0.009051 | 0.023865 | 0.707643 |
| ANKRD13E | 472.0074 | -0.4975 | 0.190626 | -2.60984 | 0.009059 | 0.02388 | 0.708332 |
| OLFML3 | 2519.062 | -0.52159 | 0.199982 | -2.6082 | 0.009102 | 0.023984 | 0.696602 |
| LRRC75B | 332.2712 | -0.53265 | 0.204375 | -2.60623 | 0.009155 | 0.024112 | 0.691285 |
| TLL1 | 15.96689 | -2.16934 | 0.832938 | -2.60444 | 0.009202 | 0.024224 | 0.222313 |
| COL6A4P2 | 21.01315 | -1.80591 | 0.693996 | -2.60219 | 0.009263 | 0.024373 | 0.286001 |
| CDHR3 | 73.47357 | -0.8777 | 0.337854 | -2.59786 | 0.009381 | 0.024643 | 0.544235 |
| TMEM173 | 27.4008 | -1.02906 | 0.396454 | -2.59566 | 0.009441 | 0.024788 | 0.490028 |
| LEXM | 2.546046 | -4.81956 | 1.857044 | -2.59529 | 0.009451 | 0.024808 | 0.035413 |
| PAPLN | 2041.425 | -0.59294 | 0.228495 | -2.59499 | 0.009459 | 0.024819 | 0.66299 |
| RECQL4 | 2109.581 | -0.8951 | 0.344925 | -2.59504 | 0.009458 | 0.024819 | 0.537712 |
| OVGP1 | 28.64713 | -1.36146 | 0.524715 | -2.59466 | 0.009468 | 0.024835 | 0.389189 |
| HGS | 1819.224 | -0.42067 | 0.162224 | -2.59314 | 0.00951 | 0.024931 | 0.747078 |
| CYP3A43 | 2.311306 | -3.92823 | 1.515678 | -2.59173 | 0.00955 | 0.025006 | 0.065688 |
| TMPRSS9 | 21.48447 | -1.2353 | 0.476739 | -2.59114 | 0.009566 | 0.025041 | 0.424755 |
| ITGA10 | 14.48387 | -1.45844 | 0.563327 | -2.58898 | 0.009626 | 0.025166 | 0.363885 |
| PSORS1C1 | 1.929493 | -4.40281 | 1.700914 | -2.5885 | 0.00964 | 0.025194 | 0.047274 |
| AGMAT | 162.3343 | -0.52452 | 0.202742 | -2.58712 | 0.009678 | 0.025274 | 0.695191 |
| ENPP7P8 | 18.22128 | -1.16199 | 0.449276 | -2.58637 | 0.009699 | 0.025318 | 0.446895 |
| MTCO2P2 | 7.144467 | -2.33561 | 0.90333 | -2.58556 | 0.009722 | 0.025374 | 0.198112 |
| AL445670. | 37.05382 | -0.80618 | 0.311846 | -2.58518 | 0.009733 | 0.025398 | 0.571895 |
| BRSK1 | 817.4588 | -0.51912 | 0.200913 | -2.58383 | 0.009771 | 0.025494 | 0.697795 |
| ST7L | 278.8861 | -0.4418 | 0.171107 | -2.58202 | 0.009822 | 0.02561 | 0.736215 |
| NPPC | 11.3355 | -1.57132 | 0.60874 | -2.58126 | 0.009844 | 0.025663 | 0.336501 |
| C1QL3 | 48.34721 | -0.7138 | 0.27658 | -2.5808 | 0.009857 | 0.025693 | 0.609713 |
| CRHBP | 4.027137 | -3.40381 | 1.319036 | -2.58053 | 0.009865 | 0.02571 | 0.094483 |
| PABPC1L2 | 23.87811 | -1.2382 | 0.479885 | -2.5802 | 0.009874 | 0.025731 | 0.423901 |
| TPM3P9 | 233.3884 | -0.44742 | 0.173517 | -2.57852 | 0.009922 | 0.025841 | 0.733354 |

| | | | | | | | |
|---|---|---|---|---|---|---|---|
| KIAA0319 | 91.14832 | -0.50843 | 0.197293 | -2.57703 | 0.009965 | 0.025935 | 0.702988 |
| SLC16A8 | 33.91795 | -1.28285 | 0.49845 | -2.57369 | 0.010062 | 0.026172 | 0.410982 |
| AC093827 | 94.71823 | -0.60588 | 0.235435 | -2.57346 | 0.010069 | 0.026185 | 0.657069 |
| CRISPLD2 | 58.4063 | -0.87829 | 0.341319 | -2.57321 | 0.010076 | 0.026197 | 0.544014 |
| SYN1 | 177.0487 | -0.5729 | 0.222637 | -2.57323 | 0.010076 | 0.026197 | 0.672266 |
| GALR3 | 14.42381 | -1.37469 | 0.534555 | -2.57166 | 0.010121 | 0.026292 | 0.385635 |
| KRTAP12-1 | 1.883621 | -4.36498 | 1.698008 | -2.57065 | 0.010151 | 0.026362 | 0.04853 |
| NTF4 | 39.62344 | -1.11862 | 0.435211 | -2.57029 | 0.010161 | 0.026385 | 0.460534 |
| HSBP1L1 | 19.60574 | -1.18691 | 0.461935 | -2.56944 | 0.010186 | 0.026443 | 0.439242 |
| NR3C1 | 165.6385 | -0.79557 | 0.309621 | -2.56949 | 0.010185 | 0.026443 | 0.576117 |
| ANXA9 | 81.50626 | -0.54519 | 0.212282 | -2.56826 | 0.010221 | 0.026511 | 0.685299 |
| ARSJ | 89.1506 | -0.70395 | 0.274094 | -2.56826 | 0.010221 | 0.026511 | 0.61389 |
| INHBC | 33.32273 | -0.86205 | 0.335751 | -2.56752 | 0.010243 | 0.02656 | 0.550171 |
| PRSS45P | 8.571173 | -1.96411 | 0.765267 | -2.56657 | 0.010271 | 0.026621 | 0.256297 |
| BTK | 17.68922 | -1.46591 | 0.571226 | -2.56625 | 0.01028 | 0.026642 | 0.362007 |
| COL5A1 | 711.6056 | -0.69394 | 0.270477 | -2.56563 | 0.010299 | 0.026678 | 0.618162 |
| TCF7 | 259.9683 | -0.44856 | 0.175014 | -2.56297 | 0.010378 | 0.026872 | 0.732776 |
| PCDHAC1 | 134.721 | -0.52993 | 0.20687 | -2.56166 | 0.010417 | 0.026958 | 0.692588 |
| FOSL1 | 83.82041 | -0.61409 | 0.239854 | -2.56028 | 0.010459 | 0.027054 | 0.65334 |
| CCDC61 | 193.1749 | -0.49415 | 0.193014 | -2.5602 | 0.010461 | 0.027057 | 0.709978 |
| SMIM4 | 164.2137 | -0.55434 | 0.21655 | -2.55987 | 0.010471 | 0.027079 | 0.680969 |
| EFEMP2 | 266.7421 | -0.49588 | 0.193728 | -2.55968 | 0.010477 | 0.02709 | 0.709128 |
| AL391825. | 4.138513 | -2.59451 | 1.014069 | -2.55851 | 0.010512 | 0.027162 | 0.165567 |
| CCL5 | 28.19475 | -0.91813 | 0.358922 | -2.55803 | 0.010527 | 0.027196 | 0.529194 |
| DCST2 | 27.89707 | -1.56899 | 0.613524 | -2.55735 | 0.010547 | 0.027241 | 0.337043 |
| SMG1P7 | 139.074 | -0.66895 | 0.261605 | -2.55711 | 0.010554 | 0.027252 | 0.628963 |
| PPM1M | 167.8695 | -0.51839 | 0.202763 | -2.55663 | 0.010569 | 0.027286 | 0.698151 |
| GPR83 | 15.99172 | -1.26566 | 0.495063 | -2.55656 | 0.010571 | 0.027288 | 0.41591 |
| GOLGA8H | 18.86541 | -1.17938 | 0.461621 | -2.55488 | 0.010623 | 0.027399 | 0.44154 |
| CALHM2 | 120.1453 | -0.43892 | 0.17186 | -2.55395 | 0.010651 | 0.027459 | 0.737685 |
| BNIP1 | 172.2789 | -0.49286 | 0.193011 | -2.55352 | 0.010664 | 0.027489 | 0.710616 |
| FABP6 | 17.39286 | -1.38441 | 0.542472 | -2.55203 | 0.01071 | 0.027591 | 0.383047 |
| FAM167B | 26.61918 | -1.06058 | 0.415624 | -2.55177 | 0.010718 | 0.027608 | 0.47944 |
| NKX2-4 | 4.032187 | -2.85583 | 1.121019 | -2.54753 | 0.010849 | 0.027918 | 0.138137 |
| MXRA5Y | 7.07193 | -2.08402 | 0.818255 | -2.54691 | 0.010868 | 0.027956 | 0.235857 |
| SLC22A15 | 141.9975 | -0.60075 | 0.235891 | -2.54674 | 0.010873 | 0.027966 | 0.659409 |
| GOLGA6L1 | 42.96056 | -1.02973 | 0.404394 | -2.54636 | 0.010885 | 0.027988 | 0.489801 |
| TLR5 | 83.90388 | -0.84016 | 0.330069 | -2.54542 | 0.010915 | 0.028052 | 0.558581 |
| SLC4A9 | 7.66474 | -1.86178 | 0.731561 | -2.54494 | 0.01093 | 0.028075 | 0.275136 |
| KLK3 | 2.547135 | -3.99505 | 1.570895 | -2.54317 | 0.010985 | 0.02821 | 0.062715 |
| FAR2P3 | 123.6503 | -0.67214 | 0.264483 | -2.54132 | 0.011043 | 0.02834 | 0.627576 |
| ZC3H12A | 109.6012 | -0.48483 | 0.190917 | -2.53949 | 0.011101 | 0.028472 | 0.71458 |
| GABRA1 | 12.27758 | -1.8131 | 0.714947 | -2.536 | 0.011213 | 0.028738 | 0.284578 |
| FLT3 | 53.07463 | -0.69864 | 0.275941 | -2.53186 | 0.011346 | 0.029044 | 0.616151 |

| | | | | | | | |
|---|---|---|---|---|---|---|---|
| PTPRCAP | 19.35632 | -2.51269 | 0.992606 | -2.53141 | 0.011361 | 0.029076 | 0.175229 |
| SPDYE2B | 4.127041 | -5.51157 | 2.177784 | -2.53082 | 0.01138 | 0.029113 | 0.021921 |
| RIPOR2 | 208.7655 | -0.42923 | 0.169717 | -2.5291 | 0.011435 | 0.029247 | 0.742658 |
| IDUA | 95.25167 | -0.81995 | 0.324255 | -2.52871 | 0.011448 | 0.029272 | 0.566462 |
| AZGP1 | 8.205157 | -2.79219 | 1.104874 | -2.52716 | 0.011499 | 0.029381 | 0.144367 |
| GLA | 608.5983 | -0.46063 | 0.182271 | -2.52717 | 0.011499 | 0.029381 | 0.726669 |
| PEAR1 | 17.39944 | -1.7229 | 0.681736 | -2.52723 | 0.011497 | 0.029381 | 0.302938 |
| LYG1 | 14.99828 | -1.48986 | 0.590311 | -2.52385 | 0.011608 | 0.029647 | 0.356048 |
| RPL26P30 | 5.476977 | -2.33679 | 0.926686 | -2.52166 | 0.01168 | 0.029807 | 0.19795 |
| TAS2R4 | 19.63018 | -1.09085 | 0.432707 | -2.521 | 0.011702 | 0.029859 | 0.469483 |
| CCR10 | 41.60045 | -0.88235 | 0.350074 | -2.52046 | 0.01172 | 0.029897 | 0.542484 |
| PYY | 20.31309 | -1.0424 | 0.413723 | -2.51955 | 0.01175 | 0.029957 | 0.48552 |
| CNDP1 | 12.02859 | -2.4677 | 0.979942 | -2.51821 | 0.011795 | 0.03005 | 0.180779 |
| CEP131 | 731.3411 | -0.59166 | 0.235207 | -2.5155 | 0.011887 | 0.030245 | 0.663578 |
| SERPINE3 | 9.07412 | -2.04932 | 0.814662 | -2.51554 | 0.011885 | 0.030245 | 0.241598 |
| FIBIN | 37.20405 | -1.03038 | 0.409833 | -2.51415 | 0.011932 | 0.030347 | 0.48958 |
| GLRX2 | 336.9751 | -0.49193 | 0.195666 | -2.51412 | 0.011933 | 0.030347 | 0.711075 |
| APOL1 | 11.55769 | -1.81174 | 0.720959 | -2.51296 | 0.011972 | 0.030436 | 0.284846 |
| SAMSN1 | 6.144745 | -2.92757 | 1.164961 | -2.51302 | 0.01197 | 0.030436 | 0.131436 |
| ACSBG1 | 56.43195 | -0.963 | 0.383475 | -2.51124 | 0.012031 | 0.030562 | 0.51299 |
| LEAP2 | 34.5503 | -0.95157 | 0.379081 | -2.51021 | 0.012066 | 0.030643 | 0.517068 |
| NFAM1 | 36.2123 | -0.83974 | 0.334544 | -2.51011 | 0.012069 | 0.030647 | 0.558744 |
| BHLHE40 | 209.6864 | -0.57528 | 0.229274 | -2.50914 | 0.012102 | 0.030727 | 0.671155 |
| PNKP | 1037.033 | -0.44696 | 0.178189 | -2.50834 | 0.01213 | 0.030793 | 0.733588 |
| AC016582 | 13.00554 | -2.68045 | 1.068913 | -2.50764 | 0.012154 | 0.030834 | 0.155993 |
| PRSS22 | 5.462491 | -2.47689 | 0.987745 | -2.50762 | 0.012155 | 0.030834 | 0.179631 |
| PROZ | 5.448932 | -2.35897 | 0.940837 | -2.50731 | 0.012165 | 0.030856 | 0.19493 |
| TTLL10 | 19.57704 | -1.77026 | 0.706053 | -2.50726 | 0.012167 | 0.030857 | 0.293156 |
| ATP6V0A4 | 21.96041 | -1.08859 | 0.434205 | -2.5071 | 0.012173 | 0.030867 | 0.47022 |
| SPSB3 | 544.8096 | -0.45299 | 0.180703 | -2.50684 | 0.012182 | 0.030878 | 0.730526 |
| ZDHHC11I | 24.8398 | -1.35351 | 0.540173 | -2.5057 | 0.012221 | 0.030972 | 0.391339 |
| HSPA1B | 92.5662 | -1.17362 | 0.468507 | -2.50502 | 0.012244 | 0.031023 | 0.443308 |
| SBF1 | 3643.488 | -0.47065 | 0.187931 | -2.50438 | 0.012266 | 0.031074 | 0.721639 |
| DDIT3 | 521.0394 | -0.47773 | 0.190911 | -2.50237 | 0.012337 | 0.031235 | 0.718107 |
| C12orf50 | 185.6585 | -0.5451 | 0.2179 | -2.5016 | 0.012363 | 0.031294 | 0.685344 |
| ZNF398 | 1083.624 | -0.49873 | 0.199373 | -2.50147 | 0.012368 | 0.031301 | 0.707731 |
| SLC10A3 | 195.7419 | -0.55176 | 0.22065 | -2.50061 | 0.012398 | 0.031369 | 0.682188 |
| WNT9B | 8.348672 | -1.83958 | 0.735671 | -2.50055 | 0.0124 | 0.031369 | 0.279402 |
| ADGRD2 | 14.51428 | -1.76687 | 0.706736 | -2.50004 | 0.012418 | 0.031407 | 0.293845 |
| SERPINA1( | 24.9634 | -1.09778 | 0.439187 | -2.49957 | 0.012434 | 0.031439 | 0.467235 |
| AHNAK2 | 172.2712 | -0.83163 | 0.332744 | -2.49931 | 0.012443 | 0.031454 | 0.561894 |
| AL031315. | 30.39585 | -0.91235 | 0.365074 | -2.49908 | 0.012451 | 0.031469 | 0.531319 |
| LYPD3 | 34.33353 | -0.86469 | 0.346069 | -2.49861 | 0.012468 | 0.031503 | 0.549163 |
| CARNS1 | 60.97756 | -0.99326 | 0.397643 | -2.49786 | 0.012495 | 0.031556 | 0.502343 |

| | | | | | | | |
|---|---|---|---|---|---|---|---|
| LRRN4CL | 3.603024 | -3.76843 | 1.510017 | -2.49562 | 0.012574 | 0.03173 | 0.073382 |
| AANAT | 7.121246 | -1.96677 | 0.78858 | -2.49407 | 0.012629 | 0.031856 | 0.255825 |
| CCDC172 | 41.90654 | -1.55745 | 0.624476 | -2.494 | 0.012631 | 0.031858 | 0.339752 |
| PRLR | 101.8599 | -0.92496 | 0.37098 | -2.49328 | 0.012657 | 0.031919 | 0.526697 |
| CRYM-AS1 | 6.818536 | -2.12315 | 0.851679 | -2.4929 | 0.012671 | 0.031948 | 0.229546 |
| PRSS53 | 160.3611 | -1.55984 | 0.626467 | -2.4899 | 0.012778 | 0.032184 | 0.339188 |
| DUSP15 | 62.03898 | -0.59128 | 0.237529 | -2.48932 | 0.012799 | 0.032228 | 0.663752 |
| EPB41L4A | 231.4003 | -0.49053 | 0.197226 | -2.48713 | 0.012878 | 0.032408 | 0.711764 |
| MYLK3 | 51.23307 | -0.63659 | 0.255979 | -2.48689 | 0.012886 | 0.032424 | 0.643231 |
| CCDC162I | 124.4062 | -0.59944 | 0.241069 | -2.48661 | 0.012897 | 0.032434 | 0.660008 |
| ZMAT4 | 144.5439 | -0.78493 | 0.315656 | -2.48665 | 0.012895 | 0.032434 | 0.580381 |
| CYP2C9 | 3.405863 | -3.18838 | 1.282361 | -2.48634 | 0.012907 | 0.03245 | 0.109699 |
| PITPNM1 | 998.0233 | -0.45238 | 0.181945 | -2.48634 | 0.012906 | 0.03245 | 0.730837 |
| WWTR1 | 490.9379 | -0.41863 | 0.168492 | -2.4846 | 0.01297 | 0.032587 | 0.748132 |
| AC092821 | 168.8755 | -0.72758 | 0.292861 | -2.48438 | 0.012978 | 0.032602 | 0.603916 |
| TUBB1 | 6.129421 | -2.56946 | 1.034318 | -2.4842 | 0.012984 | 0.032605 | 0.168468 |
| C11orf52 | 15.65101 | -1.22918 | 0.494868 | -2.48386 | 0.012997 | 0.032632 | 0.426559 |
| SNX31 | 17.17771 | -1.41565 | 0.570151 | -2.48293 | 0.013031 | 0.032713 | 0.374842 |
| AC007068 | 18.93727 | -1.78677 | 0.719803 | -2.48231 | 0.013053 | 0.032761 | 0.289819 |
| BORCS8 | 232.0695 | -0.46142 | 0.185891 | -2.48224 | 0.013056 | 0.032763 | 0.726269 |
| ESPNP | 102.7203 | -0.74732 | 0.301151 | -2.48155 | 0.013081 | 0.032817 | 0.595708 |
| ATP6V1FN | 40.6543 | -0.89597 | 0.361294 | -2.47988 | 0.013143 | 0.032949 | 0.537387 |
| ZBTB48 | 406.556 | -0.59911 | 0.241623 | -2.47953 | 0.013156 | 0.032977 | 0.66016 |
| ENKD1 | 254.1387 | -0.68025 | 0.274508 | -2.47806 | 0.01321 | 0.033104 | 0.624059 |
| TAS2R6P | 10.43495 | -1.43705 | 0.580109 | -2.47721 | 0.013241 | 0.033178 | 0.369321 |
| CGRRF1 | 406.3644 | -0.43335 | 0.175039 | -2.47575 | 0.013295 | 0.033278 | 0.740539 |
| PCK1 | 6.294825 | -4.32497 | 1.747226 | -2.47534 | 0.013311 | 0.033312 | 0.049895 |
| GOLGA6L4 | 60.11053 | -0.94404 | 0.381544 | -2.47427 | 0.013351 | 0.033402 | 0.519774 |
| STIMATE-M | 125.4772 | -0.55057 | 0.222743 | -2.47178 | 0.013444 | 0.033609 | 0.68275 |
| DHRS2 | 229.9799 | -1.51459 | 0.613027 | -2.47067 | 0.013486 | 0.033699 | 0.349996 |
| IPPK | 330.3846 | -0.4368 | 0.177344 | -2.46301 | 0.013778 | 0.034377 | 0.738772 |
| AC008770 | 2.295628 | -4.67007 | 1.896343 | -2.46267 | 0.013791 | 0.034404 | 0.03928 |
| SLC2A1 | 8321.419 | -0.48995 | 0.198976 | -2.46237 | 0.013802 | 0.034429 | 0.712048 |
| SLC27A1 | 490.1029 | -0.44726 | 0.181713 | -2.46135 | 0.013842 | 0.034513 | 0.733435 |
| CHMP4C | 125.2386 | -0.76456 | 0.310748 | -2.4604 | 0.013878 | 0.034595 | 0.588631 |
| SLC26A10 | 24.62394 | -2.61948 | 1.064787 | -2.46009 | 0.01389 | 0.03461 | 0.162727 |
| POU5F1P3 | 4.561543 | -2.40286 | 0.976954 | -2.45954 | 0.013911 | 0.034645 | 0.189089 |
| GJB5 | 13.80482 | -1.36288 | 0.554418 | -2.45822 | 0.013963 | 0.034763 | 0.388805 |
| KCNK15 | 22.49372 | -1.07694 | 0.438227 | -2.4575 | 0.013991 | 0.034824 | 0.474033 |
| AC068279 | 20.03283 | -1.34413 | 0.547734 | -2.45399 | 0.014128 | 0.035141 | 0.39389 |
| GFY | 29.6951 | -0.9229 | 0.376126 | -2.4537 | 0.014139 | 0.03516 | 0.527448 |
| PQLC2 | 472.9165 | -0.42754 | 0.174285 | -2.45311 | 0.014163 | 0.035209 | 0.743529 |
| SLC25A48 | 23.07206 | -1.23871 | 0.5053 | -2.45144 | 0.014228 | 0.035353 | 0.42375 |
| CALCA | 87.96179 | -0.76511 | 0.312162 | -2.45098 | 0.014247 | 0.035393 | 0.588411 |

| | | | | | | | |
|---|---|---|---|---|---|---|---|
| SLC26A5 | 3.734038 | -3.45574 | 1.410178 | -2.45057 | 0.014263 | 0.035414 | 0.091142 |
| PTER | 230.8832 | -0.50177 | 0.204778 | -2.45033 | 0.014273 | 0.035424 | 0.706238 |
| KCNJ8 | 7.053927 | -2.27046 | 0.926803 | -2.44977 | 0.014295 | 0.03546 | 0.207264 |
| RHCG | 22.80753 | -1.00824 | 0.411561 | -2.44979 | 0.014294 | 0.03546 | 0.497152 |
| AGAP12P | 25.94457 | -1.15139 | 0.470119 | -2.44913 | 0.01432 | 0.035511 | 0.450193 |
| CROCCP3 | 219.6084 | -0.52514 | 0.214421 | -2.44911 | 0.014321 | 0.035511 | 0.694891 |
| GCSAM | 27.8346 | -1.59558 | 0.652296 | -2.44609 | 0.014441 | 0.035766 | 0.33089 |
| CHRM3 | 222.6933 | -0.54239 | 0.221831 | -2.44505 | 0.014483 | 0.03586 | 0.686633 |
| FSD2 | 42.19807 | -0.71318 | 0.291726 | -2.44469 | 0.014498 | 0.035886 | 0.609975 |
| BTNL8 | 12.36497 | -1.4775 | 0.604423 | -2.44448 | 0.014506 | 0.035897 | 0.35911 |
| PRDM6 | 5.356062 | -3.11218 | 1.27314 | -2.44449 | 0.014506 | 0.035897 | 0.115649 |
| NDUFB4P1 | 3.487224 | -4.22074 | 1.727697 | -2.44298 | 0.014566 | 0.036034 | 0.053633 |
| PGAM1P8 | 10.2315 | -1.92105 | 0.786544 | -2.44239 | 0.01459 | 0.036086 | 0.264063 |
| NEB | 66.08629 | -1.00889 | 0.41329 | -2.44113 | 0.014641 | 0.036203 | 0.496927 |
| ISG20 | 42.55664 | -0.74505 | 0.305304 | -2.44035 | 0.014673 | 0.036262 | 0.596648 |
| SLC22A1 | 4.767713 | -2.40405 | 0.985285 | -2.43995 | 0.014689 | 0.036287 | 0.188934 |
| KLK11 | 3.489262 | -3.27569 | 1.343237 | -2.43865 | 0.014742 | 0.036388 | 0.103257 |
| MAMSTR | 34.88376 | -0.77922 | 0.319634 | -2.43784 | 0.014776 | 0.036461 | 0.582684 |
| ELMSAN1 | 372.2973 | -0.57048 | 0.234089 | -2.43704 | 0.014808 | 0.036531 | 0.67339 |
| ANKRD13D | 460.0908 | -0.5437 | 0.223153 | -2.43643 | 0.014833 | 0.036588 | 0.686011 |
| HLF | 13.8535 | -2.36203 | 0.969745 | -2.43573 | 0.014862 | 0.036644 | 0.194517 |
| OXCT2P1 | 25.27549 | -1.20996 | 0.497046 | -2.4343 | 0.014921 | 0.036774 | 0.432281 |
| DDX11 | 2857.037 | -0.42814 | 0.175897 | -2.43403 | 0.014932 | 0.036797 | 0.74322 |
| CCER2 | 13.83227 | -1.29198 | 0.531202 | -2.43219 | 0.015008 | 0.036969 | 0.408389 |
| TRAF1 | 116.3626 | -0.58554 | 0.240759 | -2.43205 | 0.015014 | 0.036979 | 0.666401 |
| FAM240A | 2.46256 | -3.94514 | 1.622974 | -2.43081 | 0.015065 | 0.037096 | 0.064923 |
| KCNN2 | 555.8544 | -0.53303 | 0.219511 | -2.42828 | 0.015171 | 0.03733 | 0.6911 |
| TCTE3 | 49.02936 | -0.70235 | 0.289336 | -2.42746 | 0.015205 | 0.0374 | 0.61457 |
| DOCK5 | 375.6338 | -0.45074 | 0.185692 | -2.42733 | 0.01521 | 0.037403 | 0.73167 |
| NPM2 | 28.50111 | -1.29964 | 0.536027 | -2.42458 | 0.015326 | 0.037663 | 0.406227 |
| PGF | 81.38986 | -0.53192 | 0.219548 | -2.42278 | 0.015402 | 0.037825 | 0.691635 |
| ALPI | 3.514896 | -3.69161 | 1.524972 | -2.42077 | 0.015488 | 0.038024 | 0.077396 |
| CALHM1 | 21.17138 | -0.99139 | 0.410032 | -2.41783 | 0.015613 | 0.038297 | 0.502994 |
| SLC9A2 | 6.163496 | -3.23474 | 1.338104 | -2.4174 | 0.015632 | 0.038336 | 0.10623 |
| ABCA11P | 138.3344 | -0.45095 | 0.186584 | -2.41689 | 0.015654 | 0.03838 | 0.731559 |
| CRYBG2 | 423.9144 | -0.41846 | 0.173166 | -2.41651 | 0.01567 | 0.038411 | 0.748224 |
| CDH26 | 21.26003 | -1.18948 | 0.492482 | -2.41527 | 0.015724 | 0.038521 | 0.438462 |
| NWD1 | 6.114031 | -2.17557 | 0.901049 | -2.41449 | 0.015757 | 0.038583 | 0.221354 |
| DGKB | 31.97589 | -1.21248 | 0.502606 | -2.41239 | 0.015848 | 0.038785 | 0.431526 |
| RASD1 | 39.47251 | -0.85152 | 0.353062 | -2.41183 | 0.015873 | 0.03884 | 0.554199 |
| AC244258 | 2.684368 | -4.21821 | 1.750923 | -2.40913 | 0.01599 | 0.039107 | 0.053727 |
| LGALS7 | 2.315974 | -4.6312 | 1.922618 | -2.4088 | 0.016005 | 0.039137 | 0.040352 |
| TNFAIP6 | 1.903522 | -4.36893 | 1.814334 | -2.40801 | 0.01604 | 0.039212 | 0.048397 |
| RRP7BP | 365.69 | -0.43568 | 0.180992 | -2.40719 | 0.016076 | 0.039289 | 0.739344 |

| | | | | | | | |
|---|---|---|---|---|---|---|---|
| CCDC188 | 3.191339 | -3.56569 | 1.481702 | -2.40648 | 0.016107 | 0.039355 | 0.084454 |
| SLC35E4 | 121.7416 | -0.64264 | 0.267136 | -2.40566 | 0.016143 | 0.039434 | 0.640541 |
| CALR3 | 9.509396 | -1.77715 | 0.738846 | -2.4053 | 0.016159 | 0.039466 | 0.291759 |
| TNFRSF13( | 79.24528 | -0.72579 | 0.30175 | -2.40526 | 0.016161 | 0.039466 | 0.604667 |
| MTND5P11 | 4.676539 | -3.08238 | 1.283436 | -2.40167 | 0.016321 | 0.039835 | 0.118062 |
| TRDN | 1.813854 | -4.27736 | 1.781045 | -2.4016 | 0.016323 | 0.039836 | 0.051569 |
| NUDT13 | 51.14514 | -0.66592 | 0.277444 | -2.40019 | 0.016387 | 0.039964 | 0.630287 |
| WFDC1 | 15.38896 | -1.78365 | 0.743115 | -2.40023 | 0.016385 | 0.039964 | 0.290447 |
| AC004890 | 81.15574 | -0.62778 | 0.261784 | -2.39809 | 0.016481 | 0.040153 | 0.647171 |
| EIF4E1B | 25.98366 | -1.21022 | 0.505246 | -2.39531 | 0.016606 | 0.040414 | 0.432203 |
| MED15P9 | 32.10501 | -1.22376 | 0.510892 | -2.39534 | 0.016605 | 0.040414 | 0.428165 |
| HIST1H2A( | 60.30989 | -0.59438 | 0.248231 | -2.39448 | 0.016644 | 0.040495 | 0.662327 |
| PRR35 | 5.770172 | -2.33815 | 0.978902 | -2.38854 | 0.016915 | 0.041112 | 0.197764 |
| REV3L | 419.4656 | -0.48827 | 0.204502 | -2.38761 | 0.016958 | 0.041205 | 0.712878 |
| EFNA3 | 327.6743 | -0.48951 | 0.205085 | -2.38686 | 0.016993 | 0.041279 | 0.712268 |
| AKR7A2P1 | 6.550762 | -2.49635 | 1.046289 | -2.3859 | 0.017037 | 0.04138 | 0.177225 |
| LPXN | 52.96063 | -0.78975 | 0.33114 | -2.38494 | 0.017082 | 0.041478 | 0.578444 |
| NUPR1 | 63.83755 | -1.39012 | 0.58287 | -2.38496 | 0.017081 | 0.041478 | 0.381532 |
| ABLIM2 | 38.47799 | -0.86555 | 0.363002 | -2.38442 | 0.017106 | 0.041531 | 0.548838 |
| FBLN5 | 23.42375 | -1.38609 | 0.581396 | -2.38408 | 0.017122 | 0.041565 | 0.3826 |
| ARRDC1 | 491.2776 | -0.47009 | 0.197262 | -2.3831 | 0.017168 | 0.04167 | 0.721918 |
| LOX | 44.89288 | -1.08292 | 0.454434 | -2.38301 | 0.017172 | 0.041674 | 0.472072 |
| RNF208 | 86.61272 | -0.62388 | 0.262039 | -2.38086 | 0.017272 | 0.041881 | 0.648924 |
| 5-Sep | 138.1609 | -0.5033 | 0.211408 | -2.3807 | 0.01728 | 0.041892 | 0.705492 |
| TAS2R5 | 32.00001 | -1.15477 | 0.485067 | -2.38064 | 0.017283 | 0.041894 | 0.449138 |
| ABCC3 | 16.43997 | -1.34729 | 0.565961 | -2.38054 | 0.017287 | 0.041899 | 0.393029 |
| HAP1 | 157.8847 | -0.48469 | 0.203713 | -2.37928 | 0.017347 | 0.042027 | 0.714651 |
| ADGRE2 | 60.4296 | -0.99209 | 0.417073 | -2.3787 | 0.017374 | 0.042079 | 0.502748 |
| DACT3 | 65.28125 | -0.65569 | 0.275834 | -2.3771 | 0.017449 | 0.042231 | 0.634774 |
| CCL27 | 10.3352 | -1.625 | 0.684126 | -2.37529 | 0.017535 | 0.04241 | 0.32421 |
| CXCL1 | 1.63181 | -4.18588 | 1.762521 | -2.37494 | 0.017552 | 0.042439 | 0.054945 |
| HPD | 13.86255 | -1.42014 | 0.598166 | -2.37415 | 0.017589 | 0.042508 | 0.373677 |
| ZNF440 | 4.006675 | -2.60969 | 1.0993 | -2.37396 | 0.017599 | 0.042525 | 0.163834 |
| AMPD2 | 768.4834 | -0.50347 | 0.212139 | -2.37329 | 0.01763 | 0.042585 | 0.705409 |
| AL731733. | 8.269975 | -1.65669 | 0.699232 | -2.3693 | 0.017822 | 0.042968 | 0.317166 |
| MAPK15 | 134.0158 | -0.92878 | 0.392044 | -2.36906 | 0.017833 | 0.042991 | 0.525304 |
| C11orf71 | 143.8106 | -0.51736 | 0.21855 | -2.36725 | 0.017921 | 0.043173 | 0.698648 |
| 1-Mar | 145.3635 | -0.47857 | 0.20216 | -2.36728 | 0.017919 | 0.043173 | 0.717689 |
| PDGFRB | 277.8033 | -0.43551 | 0.184023 | -2.36658 | 0.017953 | 0.043246 | 0.739435 |
| NAT1 | 61.06861 | -0.88074 | 0.372286 | -2.36575 | 0.017993 | 0.04332 | 0.54309 |
| SYNDIG1L | 45.17061 | -0.99367 | 0.420006 | -2.36585 | 0.017989 | 0.04332 | 0.502199 |
| MYOCD | 70.4946 | -1.0608 | 0.448507 | -2.36518 | 0.018021 | 0.043375 | 0.479366 |
| TIGIT | 3.608281 | -3.92012 | 1.659855 | -2.36173 | 0.01819 | 0.043736 | 0.066058 |
| LY6H | 6.991627 | -2.5728 | 1.08949 | -2.36147 | 0.018203 | 0.04376 | 0.168078 |

| | | | | | | | |
|---|---|---|---|---|---|---|---|
| D2HGDH | 465.8505 | -0.51427 | 0.217876 | -2.36037 | 0.018257 | 0.043867 | 0.700148 |
| TRIM53AP | 3.687916 | -3.14373 | 1.332295 | -2.35963 | 0.018293 | 0.043931 | 0.113147 |
| C11orf96 | 21.1911 | -1.00517 | 0.426156 | -2.35868 | 0.01834 | 0.044033 | 0.498213 |
| ATP23 | 102.142 | -0.57007 | 0.241748 | -2.35811 | 0.018368 | 0.044089 | 0.673585 |
| PHF1 | 41.93216 | -1.03557 | 0.439207 | -2.35782 | 0.018383 | 0.044118 | 0.487823 |
| LRRC56 | 47.26793 | -0.67768 | 0.28743 | -2.35773 | 0.018387 | 0.044123 | 0.625169 |
| IL20RA | 44.85929 | -0.93253 | 0.395701 | -2.35666 | 0.01844 | 0.044233 | 0.523939 |
| LRRC25 | 4.126138 | -2.74389 | 1.164653 | -2.35597 | 0.018474 | 0.044303 | 0.149282 |
| SRMS | 5.663825 | -2.03234 | 0.862898 | -2.35525 | 0.01851 | 0.044384 | 0.244459 |
| ATP2B4 | 2435.43 | -0.45104 | 0.191578 | -2.35434 | 0.018556 | 0.044458 | 0.731516 |
| GAL3ST4 | 169.6487 | -0.57731 | 0.245375 | -2.35276 | 0.018635 | 0.044615 | 0.670213 |
| KCTD8 | 77.73703 | -0.87244 | 0.370931 | -2.35202 | 0.018672 | 0.044683 | 0.546223 |
| PRELP | 39.96867 | -0.75526 | 0.321218 | -2.35124 | 0.018711 | 0.044754 | 0.592439 |
| DECR2 | 226.1483 | -0.60401 | 0.257014 | -2.3501 | 0.018768 | 0.044873 | 0.657923 |
| CDK3 | 127.8871 | -0.44755 | 0.190449 | -2.34999 | 0.018774 | 0.044881 | 0.733286 |
| PNCK | 310.5046 | -0.63422 | 0.270019 | -2.3488 | 0.018834 | 0.045007 | 0.644289 |
| RUFY4 | 18.42544 | -1.57819 | 0.672139 | -2.34801 | 0.018874 | 0.045088 | 0.334902 |
| COL4A5 | 2307.142 | -0.44205 | 0.188509 | -2.34499 | 0.019027 | 0.045387 | 0.736087 |
| SLC38A6 | 156.2199 | -0.53482 | 0.22823 | -2.34334 | 0.019112 | 0.045577 | 0.690245 |
| MAG | 4.71657 | -3.19285 | 1.363969 | -2.34085 | 0.01924 | 0.045834 | 0.109359 |
| ATP10A | 87.04837 | -0.92226 | 0.394018 | -2.34066 | 0.01925 | 0.045852 | 0.527681 |
| MYRF | 413.6518 | -0.4609 | 0.196943 | -2.34026 | 0.01927 | 0.045894 | 0.726534 |
| OLIG3 | 24.66677 | -1.04586 | 0.447209 | -2.33863 | 0.019354 | 0.046063 | 0.484357 |
| ECHDC3 | 5.511873 | -2.10342 | 0.899792 | -2.33768 | 0.019404 | 0.046165 | 0.232706 |
| PRSS40A | 10.84235 | -1.43994 | 0.616209 | -2.33678 | 0.019451 | 0.046264 | 0.368582 |
| GALNT13 | 229.7593 | -0.5447 | 0.233226 | -2.33549 | 0.019518 | 0.046394 | 0.685535 |
| MYCBPAP | 43.35644 | -0.79266 | 0.339409 | -2.33541 | 0.019522 | 0.046398 | 0.577279 |
| CHRNA10 | 13.98733 | -1.45241 | 0.622235 | -2.33418 | 0.019586 | 0.046538 | 0.36541 |
| NRG2 | 15.90919 | -1.30421 | 0.558932 | -2.33341 | 0.019627 | 0.046613 | 0.404942 |
| GJB3 | 11.82308 | -1.59748 | 0.685174 | -2.33149 | 0.019727 | 0.046814 | 0.330454 |
| MYH2 | 18.05368 | -2.24622 | 0.963785 | -2.33063 | 0.019773 | 0.04691 | 0.210775 |
| GTF2H2C | 179.0706 | -0.43536 | 0.186846 | -2.33007 | 0.019803 | 0.046968 | 0.739507 |
| TTLL2 | 14.48518 | -1.43711 | 0.616899 | -2.32957 | 0.019829 | 0.047017 | 0.369306 |
| KCNH7 | 8.362042 | -1.83381 | 0.787651 | -2.3282 | 0.019902 | 0.047178 | 0.280523 |
| TGFB1I1 | 87.02882 | -0.90145 | 0.38725 | -2.32782 | 0.019922 | 0.047214 | 0.53535 |
| SLC6A10P | 39.18764 | -1.18478 | 0.50898 | -2.32776 | 0.019925 | 0.047215 | 0.43989 |
| ENPP7P11 | 2.424532 | -4.74006 | 2.037863 | -2.326 | 0.020019 | 0.047431 | 0.03742 |
| STX16-NPE | 71.62018 | -0.94525 | 0.406547 | -2.32508 | 0.020068 | 0.047536 | 0.519339 |
| TCTEX1D1 | 29.68669 | -1.09595 | 0.471417 | -2.3248 | 0.020083 | 0.047564 | 0.467828 |
| UNC5CL | 66.8934 | -0.87545 | 0.376616 | -2.32451 | 0.020098 | 0.047595 | 0.545084 |
| E2F3P2 | 28.73304 | -0.94317 | 0.405964 | -2.3233 | 0.020163 | 0.047731 | 0.520087 |
| SLC25A35 | 92.83109 | -0.6152 | 0.26479 | -2.32334 | 0.020161 | 0.047731 | 0.65284 |
| TTC39A | 166.9135 | -0.47707 | 0.205435 | -2.32226 | 0.020219 | 0.047844 | 0.718434 |
| B3GNT4 | 27.91697 | -1.03948 | 0.447763 | -2.3215 | 0.02026 | 0.047922 | 0.486502 |

| | | | | | | | |
|---|---|---|---|---|---|---|---|
| PRAP1 | 4.404576 | -2.73185 | 1.177441 | -2.32016 | 0.020332 | 0.048053 | 0.150533 |
| TMEM63C | 131.9858 | -0.80686 | 0.347764 | -2.32014 | 0.020333 | 0.048053 | 0.571624 |
| IRS3P | 13.65816 | -1.38641 | 0.598067 | -2.31815 | 0.020441 | 0.048283 | 0.382516 |
| EPHA10 | 36.72162 | -0.87504 | 0.377557 | -2.31764 | 0.020469 | 0.048342 | 0.545239 |
| FLRT2 | 322.3784 | -0.67467 | 0.291172 | -2.31707 | 0.0205 | 0.048409 | 0.626477 |
| STEAP4 | 19.37717 | -1.00714 | 0.43502 | -2.31515 | 0.020605 | 0.048626 | 0.497533 |
| ABCC6 | 22.2663 | -1.36357 | 0.589326 | -2.31378 | 0.02068 | 0.048786 | 0.388618 |
| ETV7 | 23.75614 | -0.94731 | 0.409468 | -2.31351 | 0.020695 | 0.048806 | 0.518599 |
| MYRIP | 51.75026 | -0.82604 | 0.357541 | -2.31034 | 0.020869 | 0.049173 | 0.564075 |
| SAMD10 | 107.9742 | -0.52511 | 0.227337 | -2.30984 | 0.020897 | 0.049232 | 0.694904 |
| PSKH2 | 6.104288 | -2.13937 | 0.92635 | -2.30947 | 0.020918 | 0.049275 | 0.226978 |
| POLD4 | 166.1001 | -0.64788 | 0.280563 | -2.30923 | 0.020931 | 0.0493 | 0.638215 |
| MGST2 | 153.9358 | -0.48319 | 0.209311 | -2.30849 | 0.020972 | 0.049384 | 0.715393 |
